# Effects of an acute bout of exercise on cognitive function in adults with cognitive impairment: A systematic review with meta-analysis of randomised controlled trials

**DOI:** 10.1101/2024.10.04.24314881

**Authors:** Charlotte L Scott, Mia Morgan, George A Kelley, Samuel R Nyman

## Abstract

**Objective:** Examine the effects of an acute bout of physical exercise on cognitive function in adults with cognitive impairment (CI).

**Design:** Systematic review with meta-analysis of randomised controlled trials (RCTs) that investigated the effects of a single exercise session on cognitive function. Dual data abstraction, risk of bias assessment (RoB2) and strength of evidence assessment (GRADE) were conducted. Results were pooled using the inverse variance heterogeneity (IVhet) model or synthesised narratively.

**Data sources:** Six databases from inception to July 2024: (1) PubMed, (2) SportDiscus, (3) PsychINFO, (4) Cochrane Central, (5) PEDro, (6) Embase.

**Eligibility criteria:** RCT’s of acute exercise with male/female participants, aged 18+, and physician diagnosed or self-reported CI.

**Results:** 15 studies (8 parallel group, 7 crossover) representing 500 participants were included, 10 in the meta-analysis. Acute exercise significantly improved executive function (“moderate” evidence, 9 studies, 330 participants, *g*=0.33, small effect, 95%CI [0.07, 0.59], *p*=0.01, *I²*=26.77, 95%CI [00.00, 65.82]. However, this was only for high risk of bias/crossover studies. Improvements in direction of benefit but non-significant for reaction time (“very low” evidence, 7 studies, 271 participants, *g*=0.17, small effect 95%CI [-0.20, 0.54], *p*=0.36, *I²*=50.18, 95%CI [00.00, 78.88]. Improvements in memory, but mixed evidence for attention, information processing and motor memory noted via narrative assessment.

**Conclusion:** In a healthy population, acute exercise improves cognition. For adults with CI, we found evidence of improvement in executive function, reaction time and memory. However, the review was limited by the inclusion of studies primarily with “high”/“some concerns” for bias and “very low”/“low” strength of evidence.

**Summary box:**

- Chronic exercise interventions have demonstrated significant improvements in cognition for healthy and cognitively impaired adults (1,2)
- Acute exercise improves cognitive function in healthy adults (3–5) but evidence for the beneficial effects of acute exercise in *cognitively impaired* adults is uncertain.
- Our meta-analysis of 10 randomised controlled trials found that acute exercise improves executive function (small effect, moderate strength evidence) in adults with cognitive impairment.
- It may also lead to improvements in reaction time (small effect, very low strength evidence) and memory (low strength evidence).

## Introduction

Mild Cognitive Impairment (MCI) does not have a uniform definition, but typically refers to the transitional state between normal aging and the onset of dementia (6). Internationally, MCI has a prevalence of 3-37% among adults (7,8). Key characteristics associated with MCI include objective and self-reported cognitive impairment, with a notable decline in cognition and minor functional difficulties (9,10). While some with MCI do not progress to dementia, or even revert back to their prior level of cognition (11), conversion rates from MCI to dementia have been found to vary between 15%-87% (12). Dementia diagnosis is characterised by a chronic, acquired loss of more than two cognitive abilities caused by brain disease or injury (13).

Given the ongoing challenges in the development of effective, disease curing drugs (14) and the increased prevalence of cognitive impairment that will inevitably come with an aging population, it is of great importance that the therapeutic effects of non-pharmacological strategies such as exercise are explored. Exercise has been widely established as being beneficial for brain health and cognitive functioning (15,16), and even a single bout of acute exercise has been shown to improve cognitive function in adults (3–5).

The majority of existing reviews in this area have focused on the effects of acute exercise on cognitive function in healthy populations (3–5,17–22). Of the reviews exploring the effects of exercise in cognitively impaired populations, these have primarily focused on chronic exercise interventions where participants take part in multiple sessions over weeks or months (23–27). However, establishing the effects of an *acute* bout of exercise is also important for cognitively impaired populations who may stand to gain the most from the strategic use of exercise prior to completing daily activities that require executive function (e.g., driving) (28). To the best of the authors’ knowledge, only four reviews have explored the effects of acute exercise in cognitively impaired populations (4,29–31). However, these reviews are limited by strict inclusion criteria, i.e., aerobic exercise and crossover designs only (29), an exclusive focus on memory function, (4) were conducted over a decade ago (30), or exclusively assessed participants aged 60+ (31). Thus, upon consultation of the decision framework for updating systematic reviews, it is clear that an additional review of this topic is now justified (32).

To address these shortcomings, the present review broadens the scope of previous reviews to explore adults of all ages, all cognitive function domains, and additionally, investigate populations where MCI typically co-occurs (e.g., Down Syndrome, Multiple Sclerosis, Cancer, Schizophrenia, Parkinson’s) (33–38). More specifically, the primary objective was to examine the effects of an acute bout of physical exercise on cognitive functioning (both global and specific cognitive function domains) in adults with cognitive impairment. A secondary objective was to explore the impact of potential covariates on cognitive impairment (e.g., exercise intensity/duration/mode, severity of cognitive impairments, age).

## Methods

### Overview

The reporting of this study followed the guidelines from the 2020 Preferred Reporting Items for Systematic Reviews and Meta-Analyses (PRISMA) statement (39). The protocol for this review was registered in PROSPERO (#CRD42021232726) in January 2021.

### Equity, diversity, and inclusion statement

This review benefitted from a diverse authorship ranging from established academics to an early career researcher and PhD student. In addition, the author team included both genders (two females, two males) and perspectives from multiple disciplines, i.e., biostatistics, exercise and physical activity, and psychology.

### Eligibility criteria

The inclusion criteria for this systematic review based on the PICOS framework (40) were as follows: (1) Population – males and females aged 18+ years with self-reported or confirmed cognitive impairment (e.g., Mini Mental State Exam (MMSE) less than 24 (41), Montreal Cognitive Assessment (MoCA) less than 26 (42), Quick-Mild Cognitive Impairment (Q-MCI) less than 62 (43) and/or a confirmed clinical diagnosis known to affect cognitive function (e.g., Dementia, Parkinson’s, Stroke); (2) Intervention – studies using an individual acute exposure to an exercise session of any type, intensity, and duration; (3) Comparison - studies using a passive control (e.g., watching a video) or active control (e.g., stretching exercise that is not expected to produce cognitive benefits); (4) Outcomes – changes (i.e., comparing before and after exercise) in global cognitive functioning (e.g., improvement in MMSE) or specific cognitive function domains (e.g., memory); (5) Study Design - Randomised controlled trials (RCTs) with at least one intervention arm and one control arm (parallel groups or crossover) given that non-randomised trials tend to overestimate the effects of healthcare interventions (44,45).

### Information sources

Six electronic databases and registers were searched from inception to 26th July 2024: (1) PubMed, (2) SportDiscus (EBSCOhost), (3) PsychINFO (EBSCOhost), (4) Cochrane Central, (5) PEDro and (6) Embase. In addition to electronic searches, cross-referencing was conducted by examining the reference lists of previous review articles in this area, as well as each included study. Legacy RefWorks was used to store references.

### Search strategy

Search strategies specific to each database were developed by all authors. The search strategy and results for all databases searched are shown in Supplementary file 1. All database searches and article retrieval were conducted by CS.

### Study records

#### Study selection

To minimise selection bias, dual screening of all abstracts was conducted. CS and four undergraduate research assistants screened the abstracts independent of each other. The team was given thorough training (i.e., established κ >0.70 on a large test screening set) before screening abstracts. On completion of abstract screening, inter-rater agreement prior to correcting discrepant items was κ = 0.69. Any discrepancies were resolved by discussion. If agreement could not be reached, CS served as an arbitrator. When selecting the final number of studies to be included in the review, the overall precision of searches was computed by dividing the number of included studies by the total number of studies screened after removing duplicates. The number needed to read (NNR) was then calculated as the reciprocal of the precision.

#### Data abstraction

Microsoft Excel (version 16.66.1) was used to hold up to 247 items per study. The major categories of variables coded included (1) study characteristics (author, journal, year of publication, etc), (2) participant characteristics (age, gender, MMSE score, etc), (3) intervention characteristics (mode, intensity, duration, etc), and (4) data for primary and secondary outcomes (sample sizes, baseline and post-exercise cognitive test means/SDs, etc). To avoid data abstraction bias, CS and MM independently coded all studies. Based on Cohen’s kappa, initial agreement was 0.80. Disagreements were then discussed until mutual agreement was achieved. If agreement could not be reached, GK and SN provided a recommendation. Extracted data from included studies can be found in Supplementary file 2.

### Outcomes and prioritisation

The a priori primary outcomes in this study were global changes in cognitive function and specific cognitive function domains. Missing data for specific cognitive function domain outcomes were requested via electronic mail by CS.

### Risk-of-bias assessment in individual studies

The risk of bias for each study included in the meta-analysis was assessed using the Cochrane Risk of Bias instrument for RCTs (RoB2) (46). Briefly, this instrument assesses risk of bias in five distinct domains, details of which are described elsewhere (45). A further domain was added specifically for cross-over trials with respect to assessing bias arising from period and carryover effects. Assessment for risk of bias was limited to the primary outcomes of interest. Based on signalling questions, each domain was assessed as either ‘low risk’, ‘high risk’, or ‘some concerns’ (45, 46, 47). For the domain ‘blinding of participants and personnel’, all studies were classified as having a high risk of bias given the difficulty researchers face when blinding participants to group assignment in exercise intervention protocols. No trial was excluded based on risk-of-bias results. CS and MM independently assessed risk of bias for all studies. Any disagreements in the items coded were discussed until mutual agreement was reached. Inter-rater agreement prior to resolving disagreements was κ=0.61.

### Strength of Evidence

Confidence in the cumulative evidence was independently assessed by CS and MM using the Grading of Recommendations Assessment Development and Evaluation (GRADE) instrument (49). The overall strength of evidence was classified as either “high”, “moderate”, “low”, or “very low” (50).

### Data synthesis

#### Calculation of effect sizes

Given the different metrics used for the cognitive function outcomes, the standardized effect size (Hedge’s g) was calculated for each study by subtracting the change outcome differences in the exercise and control conditions, and then dividing by the pooled SD of the change outcome difference for the exercise and control conditions (51). For change score SDs that were not available, these were calculated from reported change outcome or treatment effect 95% confidence intervals (CI’s), pre- and post-SD values according to procedures developed by Follman et al. (51), or requested from study authors. If an outcome had at least five effect sizes, results were pooled using the inverse variance heterogeneity model (IVhet), per recent recommendations (53).

#### Pooled estimates for changes in outcomes

Where meta-analysis was not possible, results were narratively synthesized. Two-tailed z- alpha values less than or equal to 0.05 and non-overlapping 95%CIs were considered statistically significant. Heterogeneity and inconsistency were estimated using the Q (54) and *I²* statistics (55). An alpha level less than 0.10 for Q represented statistically significant heterogeneity while higher values for *I²* represent greater inconsistency (55). Absolute between-study heterogeneity was calculated using tau squared. Sensitivity analyses were conducted with each study deleted from the model once. Post hoc, a decision was made to also calculate 95% prediction intervals (PIs) to estimate what result one might expect if a new trial was conducted in similar populations. Small-study effects (publication bias, etc.) were assessed using the Doi plot and the Luis Furuya-Kanamori (LFK) index (56). LFK values within ±1, greater than ± 1 but within ± 2, and greater than ± 2 are considered to represent no, minor, and major asymmetry (57). Planned subgroup analyses were as follows: (1) domains of cognitive function (executive function, memory, motor memory, reaction time, information processing, attention, crystallised intelligence) (30), (2) type of CI (MCI versus participants with a condition known to affect cognition), (3) age (adults aged 18-64 versus those aged 65 years and above), (4) exercise session duration in minutes: <30 minutes versus ≥ 30 minutes), (5) risk of bias (some concerns versus high), (6) source of diagnosis (self-reported versus physician), and (7) RCT design (parallel groups versus crossover). Non-overlapping 95%CI’s between groups were considered statistically significant.

#### Protocol Amendments

Post hoc, data were meta-analysed for secondary outcomes only (i.e., indices of cognitive function) due to a lack of data for the primary outcome (i.e., global cognitive function). Furthermore, covariate analyses for group vs individual exercise, aerobic vs anaerobic exercise, high versus medium versus low intensity exercise and self-reported versus physician diagnosis could also not be conducted due to a lack of available data, neither could an outlier analysis. Finally, a cumulative meta-analysis was not conducted given evidence to suggest this requires a minimum of 15 studies to be accurate (58).

#### Software used for statistical analysis

All data were analysed using MetaXL (version 5.3) and Microsoft Excel (version 16.66.1, Microsoft Corporation; 2016).

## Results

### Study characteristics

Of the 9398 citations screened after removing duplicates both electronically and manually, 15 studies representing 18 exercise groups, 17 control groups and 500 participants (322 exercise, 313 control) met the criteria for inclusion. The precision of the search was 0.16% while the NNR was 626. A flow diagram that depicts the search process is shown in Figure 1 while a list of the 9383 excluded studies, including the reasons for exclusion can be found in Supplementary file 3.

**Figure 1.**
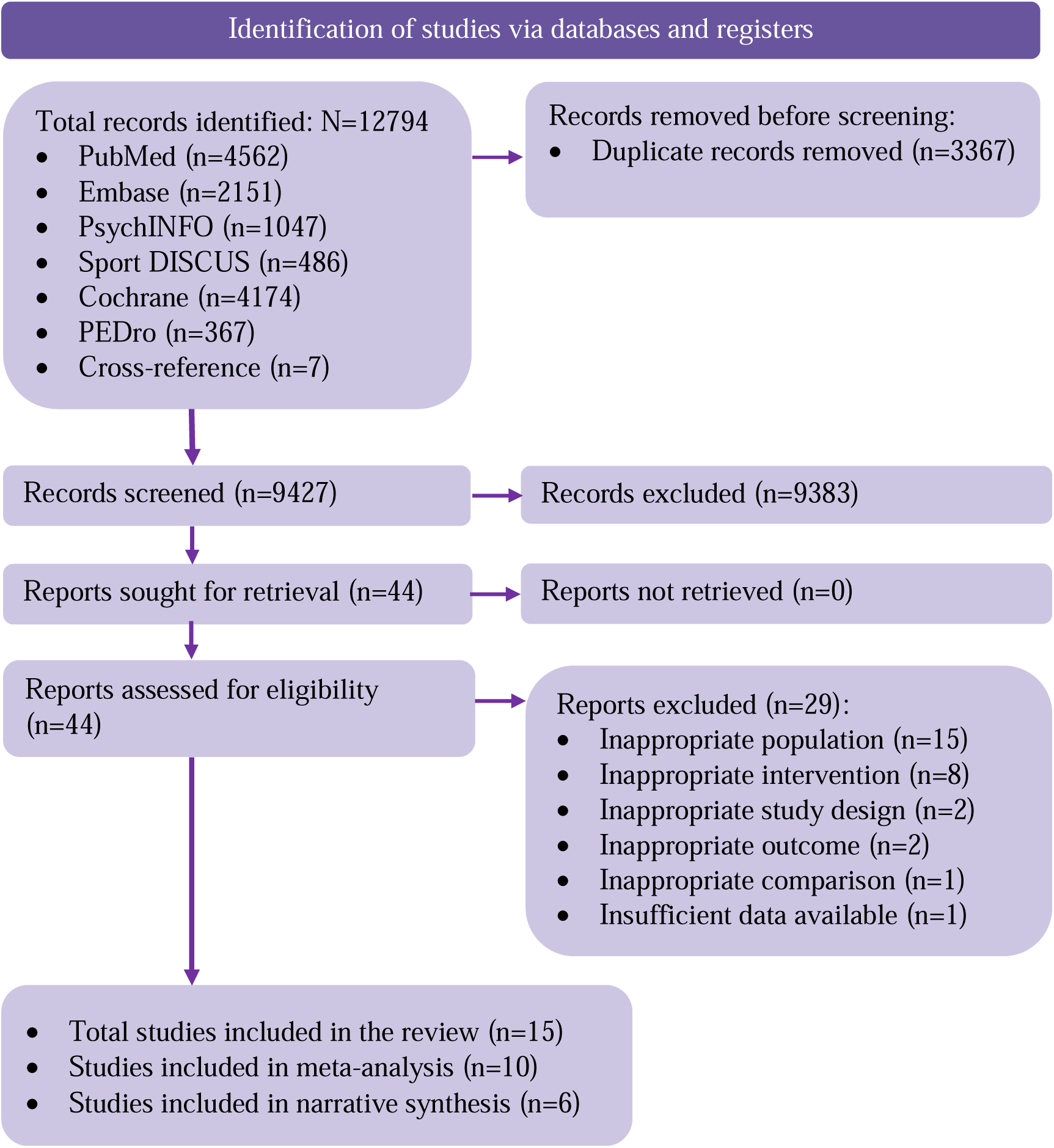
Flow diagram for selection of studies.

General study characteristics are shown in Supplementary file 4. The included studies were published in 15 different journals since 1988. Studies were conducted in seven different countries, five in the USA (33–36,59), three in Canada (37,60,61), two in Pakistan (62,63), one in Ireland (62), one in Taiwan (63), one in Italy (64), one in Germany (38) and one in Brazil (67). Of the 15 included studies, five were two-arm RCTs limited to one exercise and one control group that met all eligibility criteria (33,38,62–64). Eight trials reported a parallel arm RCT, and seven trials reported a cross-over RCT. Ten reported that all participants completed the study (34,37,38,59,61–65,67). Of the five studies with drop-outs, all used the per protocol approach in the analysis of their data.

### Participant characteristics

Baseline characteristics of the participants are shown in Supplementary file 4. Pooled mean age of participants was 61.97 (SD=16.44), and age range was 22.02 (SD=5.30) (33) to 83.00 (SD=4.00) (66). More than half the studies included both males and females (33,37,59–61,64–68), (33,37,38,59–61,64–66) while two were limited to females (34,35). Three did not describe the gender composition of their sample (34,62,63). For the five studies that reported race/ethnicity (35–37,62,63), participants were reported by authors as being multiple races, including Whites and Asians. Some studies included one or more participants with hypertension (61), cancer (35,36) and history of heart disease (66). For those studies in which data were available, one reported that some of the participants smoked cigarettes (37), none reported that participants consumed alcohol, and nine reported some participants to be overweight (33–38,61,65,66).

### Exercise intervention characteristics

Fourteen studies (93%) included solely aerobic exercise (33–38,59–64,66,67), while one included both aerobic and resistance exercise (65). The mean length of exercise sessions was 24 minutes (SD=7.85) (Median=20 minutes; Range=10-45 minutes). Methods for assessing the intensity of training for both aerobic and resistance exercise varied between the 13 studies that reported such information (33–38,60–65,67).The most commonly reported intensity was considered moderate (69). Specific activities performed included stationary cycling (37,38,59,62–65,67), treadmill walking (33–36), semi-recumbent stepping (61), a mixture of aerobic exercise and light stretching (60) and light ball games (tossing/catching ball) (66). For the one study that included a resistance training group (65), six different free weight exercises with two sets and 10 repetitions per exercise were used. All studies reported that exercise was supervised and facility based. Of the three (20%) studies to report the social setting of the exercise, two reported that exercise was conducted as part of a group (60,66) while another reported that exercise occurred individually (63).

### Risk-of-bias assessment

For the ten studies included in the meta-analysis, summary results using the Cochrane RoB2 Instrument are shown in Table 1, while study level results are shown in Supplementary file 4. Overall, six studies (33,35,60,64–66) were rated as being at a “high risk” of bias, while the remaining four studies (34,36,37,61) were considered to have “some concerns” for risk of bias. All studies were considered to have a low risk of bias for measurement of the outcome, and a majority (*n*=7) were considered to have low risk of bias for missing outcome data.

**Table 1.** Summary risk of bias assessment for crossover and parallel group studies included in the meta-analysis (N=10)

| Crossover studies (n=7) |  |  |  |  |  |  |  |
| --- | --- | --- | --- | --- | --- | --- | --- |
| Risk | Randomisation process | Bias arising from period and carryover effects | Deviations from intended interventions | Missing outcome data | Measurement of the outcome | Selection of the reported result | Overall bias |
| Low risk | 71.4% | 42.9 | 42.9% | 85.7% | 100% | 0% | 0% |
| Some concerns | 28.6% | 14.3% | 14.3% | 0% | 0% | 100% | 57.1% |
| High risk | 0% | 42.9% | 42.9% | 14.3% | 0% | 0% | 42.9% |
| Parallel group studies (n=3) |  |  |  |  |  |  |  |
| Risk | Randomisation process | Bias arising from period and carryover effects | Deviations from intended interventions | Missing outcome data | Measurement of the outcome | Selection of the reported result | Overall bias |
| Low risk | 66.6% | N/A | 0% | 33.3% | 66.6% | 0% | 0% |
| Some concerns | 33.3% | NA | 33.3% | 0% | 33.3% | 33.3% | 0% |
| High risk | 0% | NA | 66.6% | 66.6% | 0% | 66.6% | 100% |

### Data synthesis

Five trials were required to perform a meta-analysis for each cognitive function measure (52).

#### Primary outcomes

Only one study explored global changes in cognitive function (60). Total MMSE scores were compared before and after a control and acute exercise condition. An effect in the direction of benefit was identified (*g*=0.69).

#### Secondary outcomes

##### Executive function

Changes in executive function are shown in Figure 2 and associated plots in Supplementary file 7. Across all designs and categories, the overall pooled result for changes in executive function were statistically significant in the direction of benefit (*g*=0.33, 95%CI [0.07,0.59], *p*=0.01). No significant heterogeneity was observed (Q=10.92, *p*=0.21), with a low level of inconsistency but wide 95% confidence intervals (*I^2^*=26.77, 95%CI [00.00,65.82]). Tau-squared was 0.04. No asymmetry suggestive of small-study effects (publication bias, etc.) was observed (LFK=0.85). The 95%PI for what result one might expect if they conducted a new trial in populations similar to those included in the meta-analysis was -0.24-0.90. With each study deleted from the model once, changes remained statistically significant across all deletions, ranging from 0.23 (95%CI [0.002,0.46]) to 0.39 (95%CI [0.09,0.46]). One study (66) was close to identification as an outlier with a very large effect size in the direction of benefit (*g*=1.19). Upon inspection, it is plausible that this was due to a learning effect associated with repeated use of the test for severe impairment pre-post exercise.

**Figure 2.**
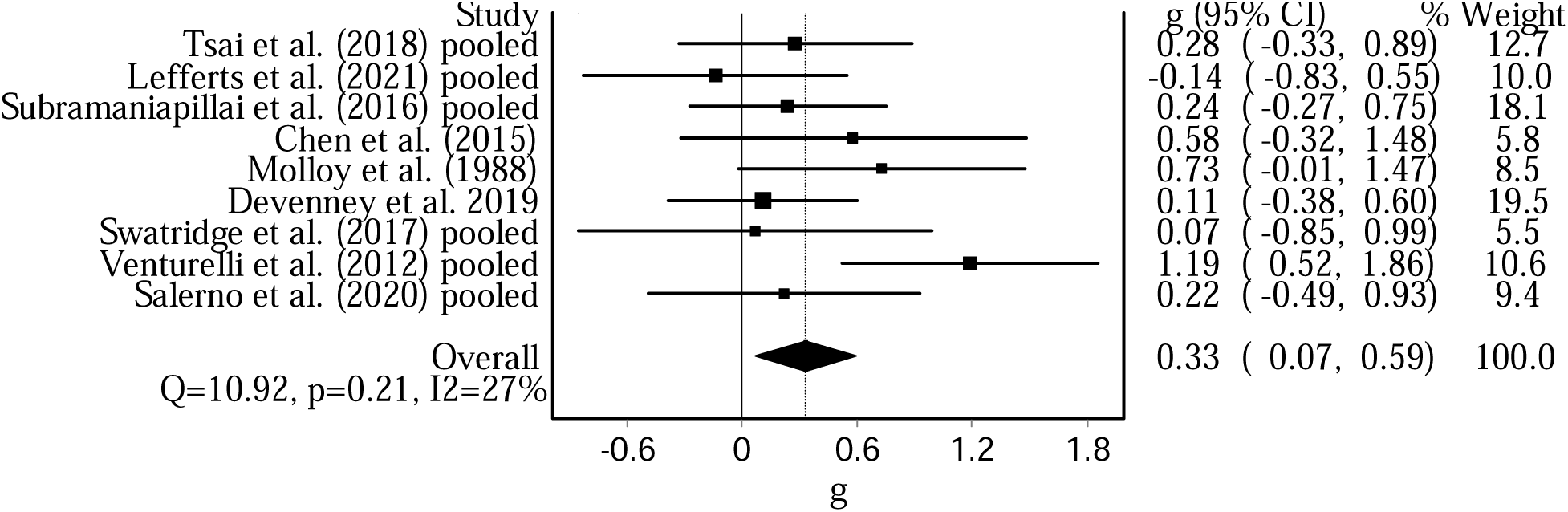
Forest plot for changes in executive function (N=9 studies).

##### Reaction time

Changes in reaction time are shown in Figure 3 and associated plots in Supplementary file 7. Across all designs and categories, the overall pooled result for changes in reaction time were in the direction of benefit but not statistically significant (*g*=0.17, 95%CI [-0.20, 0.54], *p*=0.36). Statistically significant heterogeneity was observed (Q=12.04, *p*=0.06) with moderate but wide 95% confidence intervals for inconsistency (*I^2^*=50.18, 95%CI {00.00,78.88]). Tau-squared was 0.12. No asymmetry suggestive of small-study effects (publication bias, etc.) was observed (LFK=-0.13). The 95%PI was -0.84 to 1.18. With each study deleted from the model once, changes ranged from 0.07 (95%CI, -0.30, 0.45) to 0.39 (95%CI, 0.10, 0.68).

**Figure 3.**
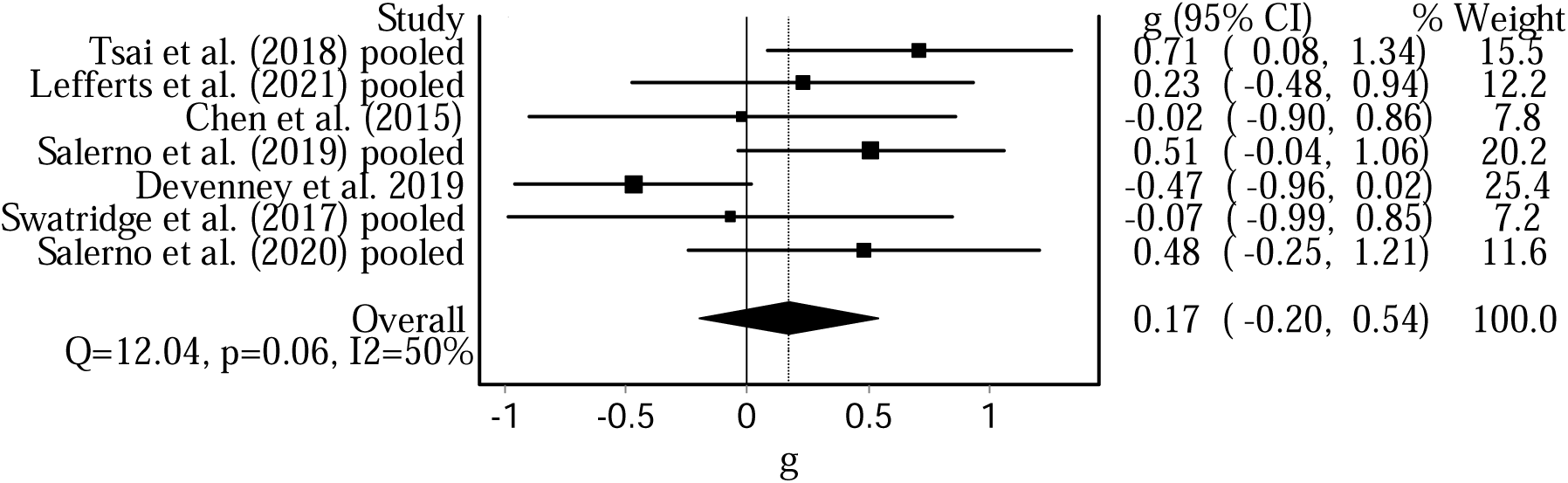
Forest plot for the reaction time outcome (N=7 studies)

##### Memory

Four studies explored the impact of acute exercise on memory. One found an effect in the direction away from benefit (*g*=-0.95; 64) while three found effects in the direction towards benefit *g*=0.41 (60), *g*=1.14 (35), *g*=5.05 (66).

##### Attention

Two studies explored the impact of acute exercise on attention. While one study (33) found an effect in the direction of benefit (*g*=0.59), another (64) found an effect in the direction away from benefit (*g*=-1.19).

##### Information Processing

Two studies explored the impact of acute exercise on information processing. One (60) found an effect in the direction away from benefit (*g*=-0.21) while another (35) found an effect in the direction towards benefit (*g*=0.43).

##### Motor memory

Two studies explored the impact of acute exercise on motor memory. One (66) found an effect in the direction of benefit (*g*=4.30) while another (38) found an effect in the direction away from benefit (*g*=-0.54).

##### Crystallised intelligence

One study explored the impact of acute exercise on crystallised intelligence and found an effect in the direction away from benefit (g=-1.47) (66).

#### Covariate and sensitivity analyses

Given that the impact of acute exercise was only found to be significant for executive function, covariate analyses were only conducted for this outcome. Age group (<65 vs ≥ 65) (*g*=-0.07, 95%CI [-0.81, 0.66], *p*=0.85), length of exercise session (<30 minutes vs ≥ 30 minutes), (g=0.77, 95%CI [-0.29, 0.67], *p*=0.44) and type of impairment (MCI vs condition known to affect cognition) (*g*=-0.27, 95%CI [-0.94, 0.41], *p*=0.44) were not found to be statistically significant covariates. However, the risk of bias (some concerns vs high risk) covariate trended towards statistical significance (*g*=-1.04, 95%CI [-2.07, 0.003], *p*=0.051) and study design (parallel vs crossover) was a statistically significant covariate (*g*=-0.84, 95%CI [-1.55, -0.13], *p*=0.021).

For studies with a high risk of bias (33,60,64–66), a statistically significant overall pooled result was found in the direction of benefit for improved executive function (*g*=0.49, 95%CI [0.08, 0.90], *p*=0.02, 95%PI= –0.72-1.70). However, for studies with some risk of bias (34,36,37,61), the overall pooled result was not significant but in the direction of benefit (*g*=0.13, 95%CI [0.20, 0.46], *p*=0.46). For studies with a crossover design (34,36,37,60,61,66), the overall pooled result was found to trend in direction of benefit for improved executive function (*g*=0.39, 95%CI, [-0.01, 0.79], *p*=0.051). However, for studies with a parallel groups design (33,64,65), the overall pooled result was not significant but in the direction of benefit (*g*=0.24, 95%CI [-0.11, 0.59], *p*=0.18]. See Supplementary file 7.

#### Narrative synthesis

Two studies were not deemed suitable for inclusion in the meta-analyses due to the use of varied EEG cognitive outcomes (62,63). Following acute aerobic exercise, a significant improvement was found in the slowness and complexity of EEG delta waves compared to the control group (62). A subsequent study explored the impact of acute exercise on Event Related Potentials. While a significant reduction in the N30 peak was identified, no significant effects were found for Somatosensory Evoked Potentials (63). An additional study used a non-standard control condition (59). Participants cycled (stationary), while either engaging on-screen with: (1) a bike tour (low cognitive demand) or (2) a video game (high cognitive demand task). Significant improvements in executive function were identified following exercise in the high cognitive demand task versus the low cognitive demand task. A final study lacked available pre-post effect sizes (difference scores computed instead; 67). No exercise-induced benefits were identified for implicit motor learning between skill acquisition and skill retention following a 20-minute bout of cycling.

### GRADE results

GRADE results for the strength/certainty of evidence for changes in global cognitive function and secondary cognitive function domains can be found in Supplementary file 6.

Strength/certainty of evidence was considered “very low” for global cognitive function and the sub-domains of motor memory, attention, information processing and reaction time.

Evidence was considered “low” for the memory and crystallised intelligence sub-domains, while evidence was “moderate” for executive function.

## Discussion

### Overall findings

The primary purpose of this systematic review and meta-analysis was to explore the impact of an acute bout of exercise on the global cognitive function of adults with cognitive impairment. Only one included study explored global changes in cognitive function as assessed via the MMSE (60). While an effect in the direction of benefit was identified, the mean change in score from 22 to 23 is unlikely to have resulted in a useful clinical effect. Further studies exploring global changes in cognitive function are essential, especially since these are more likely to be clinically relevant.

### Overall findings for secondary outcomes

All other included studies focused on specific domains of cognitive function, executive function, reaction time, memory, motor memory, attention, information processing and crystallised intelligence. Given the lack of available data, only two meta-analyses (i.e., gold-standard evidence) were conducted.

For executive function, a significant effect in the direction of benefit was identified (*p*=0.01) with a small effect size (*g*=0.33). Upon further exploration, age group, length of exercise session and degree of cognitive impairment were not found to be significant covariates. However, risk of bias and study design were identified as covariates whereby a significant effect of exercise was identified only in studies with a high risk of bias and cross-over design (plausibly due to having higher power). For reaction time, while an effect in the direction of benefit was identified, this was small and not statistically significant (*g*=0.17, *p*=0.36). While findings in the direction of benefit are encouraging, especially given the “moderate” strength of evidence for executive function and low risk of publication bias identified for both reaction time and executive function, the clinical importance may be questioned given the small effect sizes. Furthermore, the 95%PIs were overlapping, reducing confidence in the finding.

For the “low” / “very low” graded evidence narratively synthesised, a mixed picture was revealed. Evidence was limited (i.e., one or two studies) and conflicting for attention (33,64), motor memory (38,66), information processing (35,60) and crystallised intelligence (66). On the other hand, the impact of acute exercise was primarily (3 of 4 studies) found to be in the direction of benefit for memory (35,60,64,66).

### Implications for research

Included studies primarily evaluated the impact of acute exercise on cognitive function without exploring associated changes in brain activity. However, by adopting simultaneous measures of brain activity such as EEG and fMRI, the effects of acute exercise on cognitive function can be understood at the neuronal level (61,62). To definitively unpack the mechanisms underlying the acute exercise-cognition relationship, future research should adopt functional neuroimaging and explore biomarkers alongside standardised tests of cognitive function. In addition, it is important that researchers turn their attention to potential moderating/mediating factors that can affect the *strength* of this relationship. Studies included in the review primarily explored the impact of aerobic exercise conducted at medium intensity for 20 minutes. Therefore, a priori specified sub-group analyses split by mode of exercise, intensity, and duration were not possible. Existing research has also neglected to account for individual differences such as gender and baseline physical fitness. Clarification is now needed as to how exercise characteristics and individual differences may contribute to differential effects of acute exercise on cognitive function

### Implications for practice

Previous systematic reviews and meta-analyses have found good evidence in healthy populations for the beneficial impact of acute exercise on cognitive function (3–5). Our study has added to this evidence, tentatively suggesting that acute exercise may improve executive function, reaction time and memory sub-domains in adults with cognitive impairment.

Therefore, acute bouts of exercise may be warranted with respect to improving cognitive function among adults with cognitive impairment. As the population continues to age and the prevalence of mild cognitive impairment and conditions known to affect cognition are on the rise, early behavioural interventions are important for those seeking to remain independent and stave off cognitive decline (59,70). By strategically implementing exercise prior to daily activities that require a high degree of cognition (e.g., driving), this may increase the likelihood of success. Specifically, evidence suggest that acute exercise induced cognitive benefits may be particularly helpful for populations undergoing motor rehabilitation (e.g., in those with stroke). Post-exercise enhancements in cognitive control may enable patients to focus their attention for prolonged periods, optimise memory formation, improve sensori-motor integration, increase their processing speed and thus improve their overall performance in rehabilitation activities (38,61,63). Therefore, to optimise the benefits of therapy, physiotherapists may wish to encourage patients to engage in exercise directly before such rehabilitation activities.

### Implications for policy

Given the weak evidence base and high risk of bias of included studies, our results do not currently warrant any policy recommendations for the cognitive outcomes examined.

However, acute exercise has been shown to be safe and benefit other outcomes in a cognitive impaired population. For example, the improved performance in rehabilitation activities outlined above, which should be of interest to policy makers

### Strengths and potential limitations

A key strength of this study is that to the best of the authors’ knowledge, this is the first to comprehensively examine the impact of acute exercise on cognitive function in adults with cognitive impairment, using appropriate meta-analysis methodology. Findings add weight to the literature that acute exercise may improve cognitive function for this population, although further research is needed.

A number of potential study limitations need to be considered in the context of abovementioned findings. First, the GRADE strength/certainty of evidence was only “moderate” for executive function and “low”/“very low” for remaining outcomes. Second, risk of bias evaluated as “some concerns”/”high”, meaning that caution needs to be applied when interpreting overall treatment effects. Third, five studies with dropouts analysed their data per protocol as opposed to using intention to treat analyses, potentially reducing real world relevance of findings. Finally, a lack of statistically significant results for some of our findings may have been the result of the small sample sizes.

## Conclusions

In a healthy population, acute exercise is associated with improvements in multiple domains of cognitive function. In the present review exploring adults with cognitive impairment, we found acute exercise is associated with improvements in executive function, reaction time and memory. However, caution needs to be applied when interpreting these findings given the high risk of bias and overall weak strength of evidence.

## Data Availability

All data produced in the present study are available upon reasonable request to the authors

## Notes

### Competing Interest Statement

The authors have declared no competing interest.

### Funding Statement

This study did not receive any funding

