## Supplementary File 1 for "Effects of an acute bout of exercise on cognitive function in adults with cognitive impairment: A systematic review with meta-analysis of randomised controlled trials"

### PubMed

| Search | Actions | Details | Query | Results |
|---|---|---|---|---|
| #5 | ••• | > | Search: **((Cognition[Title/Abstract] OR Cognition Disorders[Title/Abstract] OR Parkinson Disease[Title/Abstract] OR Stroke[Title/Abstract] OR Dementia[Title/Abstract] OR MCI[Title/Abstract] OR Cognitive Decline[Title/Abstract] OR Stroke[Title/Abstract] OR alzheimer[Title/Abstract] OR Huntington[Title/Abstract]) AND (Exercise[Title/Abstract] OR Physical Activity[Title/Abstract] OR Fitness[Title/Abstract] OR Exergame[Title/Abstract])) AND (Acute[Title/Abstract] OR bout[Title/Abstract] OR single[Title/Abstract] OR chronic[Title/Abstract] OR aerobic[Title/Abstract] OR resistance[Title/Abstract] OR intensity[Title/Abstract] OR weight training[Title/Abstract])** Filters: **Journal Article, Humans, English, Adult: 19+ years** | 3,932 |

### Embase

| # ▲ | Searches | Results |
|---|---|---|
| 1 | Cognitive Dysfunction/ | 85968 |
| 2 | exp Cognition/ | 2353971 |
| 3 | exp Stroke/ | 219699 |
| 4 | Cognition Disorders.mp. | 2584 |
| 5 | exp Parkinson's Disease/ | 158842 |
| 6 | exp Dementia/ | 371676 |
| 7 | (mci or "cognitive impair*" or "cognitive decline" or cognition or "executive function" or dement* or stroke or alzheimer* or huntington* or parkinson*).mp. [mp=title, abstract, heading word, drug trade name, original title, device manufacturer, drug manufacturer, device trade name, keyword, floating subheading word, candidate term word] | 1312104 |
| 8 | 1 or 2 or 3 or 4 or 5 or 6 or 7 | 3345582 |
| 9 | exp Exercise/ | 352880 |
| 10 | (bout or acute or chronic or single or aerobic or resistance).mp. [mp=title, abstract, heading word, drug trade name, original title, device manufacturer, drug manufacturer, device trade name, keyword, floating subheading word, candidate term word] | 6628934 |
| 11 | 9 and 10 | 132004 |
| 12 | ((exercise or "physical exercise" or "physical activity" or fitness or exergam* or weight-training) adj3 (acute or bout or single or resistance or aerobic or chronic or intensity)).mp. [mp=title, abstract, heading word, drug trade name, original title, device manufacturer, drug manufacturer, device trade name, keyword, floating subheading word, candidate term word] | 71390 |
| 13 | 11 or 12 | 157468 |
| 14 | 13 and 8 | 26054 |
| 15 | limit 14 to (human and english language and exclude medline journals and english and article and journal and (adult <18 to 64 years> or aged <65+ years>)) | 993 |

### PsychInfo

| # ▲ | Searches | Results | Type |
|---|---|---|---|
| 1 | Cognitive Dysfunction/ | 37831 | Advanced |
| 2 | exp Cognition/ | 38650 | Advanced |
| 3 | exp Stroke/ | 21274 | Advanced |
| 4 | Cognition Disorders.mp. | 31765 | Advanced |
| 5 | exp Parkinson's Disease/ | 25275 | Advanced |
| 6 | exp Dementia/ | 78605 | Advanced |
| 7 | (mci or "cognitive impair*" or "cognitive decline" or cognition or "executive function" or dement* or stroke or alzheimer* or huntintington* or parkinson*).mp. [mp=title, abstract, heading word, table of contents, key concepts, original title, tests & measures, mesh] | 340272 | Advanced |
| 8 | 1 or 2 or 3 or 4 or 5 or 6 or 7 | 352136 | Advanced |
| 9 | exp Exercise/ | 26975 | Advanced |
| 10 | ((exercise or "physical exercise" or "physical activity*" or fitness or exergam*) adj3 (acute or bout or single or resistance or aerobic or chronic or intensity)).mp. [mp=title, abstract, heading word, table of contents, key concepts, original title, tests & measures, mesh] | 7478 | Advanced |
| 11 | (bout or acute or chronic or single).mp. [mp=title, abstract, heading word, table of contents, key concepts, original title, tests & measures, mesh] | 409173 | Advanced |
| 12 | 9 and 11 | 4550 | Advanced |
| 13 | 12 or 10 | 9970 | Advanced |
| 14 | 13 and 8 | 1543 | Advanced |
| 15 | limit 14 to (peer reviewed journal and human and english language and adulthood <18+ years> and "300 adulthood <age 18 yrs and older>" and "0110 peer-reviewed journal" and journal article and english and human) | 839 | Advanced |

### SPORT Discus

| Search ID# | Search Terms | Search Options | Actions |
|---|---|---|---|
| S1 | AB ( cognitive dysfunction or cognitive disord* or cognition or cognitive impair* or mild cognitive impair* or MCI or cognitive decline or dement or alzheimer* or stroke or parkinson* or Huntington or Amnes* ) AND AB ( exercis* or physical activity or fitness or exergam* ) AND AB ( acute or bout or chronic or aerobic or resistance or intensity or weight-training ) AND AB ( adults or adult or aged or elderly or middle aged or older person or geriatric or senior ) | **Limiters** - Peer Reviewed; Language: English; Publication Type: Academic Journal; Document Type: Article<br>**Expanders** - Apply related words; Apply equivalent subjects<br>**Search modes** - Boolean/Phrase | **View Results** (296) |

### Cochrane Central

#### Advanced Search

Search | Search manager | Medical terms (MeSH) | PICO search BETA

Save this search | View saved searches | Search help

View fewer lines | Print

| | Search | Results |
|---|---|---|
| #1 | Cognitive Dys* or Cognition or Stroke or Cognition Disord* or Parkin* or Cognitive Decline or MCI or Dement* or Cognitive Impair* or Impaired Cognition | 132211 |
| #2 | (Exercis* or physical exercise or physical act* or fitness or exergam* or weight training) adj3 (acute or bout or single or resistance or aerobic or chronic or intensity) | 1734 |
| #3 | Exercis* | 108859 |
| #4 | bout or acute or chronic or single or resistance or aerobic or intensity | 517490 |
| #5 | #3 AND #4 | 58347 |
| #6 | #1 AND #5 | 7763 |
| #7 | #1 AND #2 | 835 |
| #8 | #6 OR #7 | 8145 |
| #9 | Adult* | 669137 |
| #10 | #8 AND #9 | 4076 |

in Trials (Word variations have been searched)

### PEDro (239 hits)

| | |
|---|---|
| Abstract & Title: | Cognit*, impair*, exercis* |
| Therapy: | |
| Problem: | |
| Body Part: | |
| Subdiscipline: | |
| Topic: | |
| Method: | clinical trial |
| Author/Association: | |
| Title Only: | |
| Source: | |
| Published Since: | [YYYY] |
| New records added since: | [DD/MM/YYYY] |
| Score of at least: | [/10] |
| Return: | 50 records at a time |
| When Searching: | Match all search terms (AND) |
