## Supplementary File 3 for "Effects of an acute bout of exercise on cognitive function in adults with cognitive impairment: A systematic review with meta-analysis of randomised controlled trials"

| | Article Title | Exclusion reason |
|---|---|---|
| 1 | A Brief Review of the Effects of Physical Activity in Subjects with Cardiovascular Disease: An Interpretative Key | Inappropriate population |
| 2 | Acute Cardiometabolic Responses to Three Modes of Treadmill Exercise in Older Adults With Parkinson's Disease | Inappropriate population |
| 3 | Acute Effect of Exercise on Cognitive Performance in Middle-Aged Adults: Aerobic Versus Balance | Inappropriate study design |
| 4 | Acute Effects of Resistance Exercise on Cognitive Function in Healthy Adults: A Systematic Review with Multilevel Meta-Analysis | Inappropriate study design |
| 5 | An Analysis of Functional Outcome Measures After Treadmill Training in Older Adults With Parkinson's Disease | Inappropriate study design |
| 6 | Associations Between Physical Activity Intensities and Physical Function in Stroke Survivors | Inappropriate study design |
| 7 | Brain Health and Exercise in Older Adults | Inappropriate population |
| 8 | Combining perceptual regulation and exergaming for exercise prescription in low-active adults with and without cognitive impairment | Inappropriate study design |
| 9 | Dose-Response-Relationship between Physical Activity and Cognition in Elderly. / Dosis-Wirkungs-Beziehung zwischen körperlicher Aktivitätund Kognitionen bei Älteren | Inappropriate study design |
| 10 | Dulaglutide: A Review in Type 2 Diabetes | Inappropriate study design |
| 11 | Economic evaluations of physiotherapy interventions for neurological disorders: a systematic review | Inappropriate study design |
| 12 | Effect of 9 weeks continuous vs. interval aerobic training on plasma BDNF levels, aerobic fitness, cognitive capacity and quality of life among seniors with mild to moderate Alzheimer's disease: a randomized controlled trial | Inappropriate study design |
| 13 | Effect of acute aerobic exercise on cognitive performance: Role of cardiovascular fitness | Inappropriate study design |
| 14 | Effect of different exercise modes at high intensity on immediate learning and arousal | Inappropriate study design |
| 15 | Effects of 12-Week Resistance Exercise on Electroencephalogram Patterns and Cognitive Function in the Elderly With Mild Cognitive Impairment: A Randomized Controlled Trial | Inappropriate study design |
| 16 | Effects of a Single-Session Cognitive Enhancement Fitness Program on Serum Brain-Derived Neurotrophic Factor Levels and Cognitive Function in Middle-Aged Women | Inappropriate study design |
| 17 | Effects of an Exercise and Mental Activity Program for People With Dementia and Their Care Partners | Inappropriate study design |
| 18 | Effects of Dance/Movement Training vs. Aerobic Exercise Training on cognition, physical fitness and quality of life in older adults: A randomized controlled trial | Inappropriate population |
| 19 | Effects of Exercise Training Interventions on Executive Function in Older Adults: A Systematic Review and Meta-Analysis | Inappropriate population |
| 20 | Effects of Exergaming on Cognition and Gait in Older Adults at Risk for Falling | Inappropriate study design |
| 21 | Effects of high-velocity circuit resistance and treadmill training on cardiometabolic risk, blood markers, and quality of life in older adults | Inappropriate population |
| 22 | Effects of Tai Chi on Cerebral Hemodynamics and Health-Related Outcomes in Older Community Adults at Risk of Ischemic Stroke: A Randomized Controlled Trial | Inappropriate population |
| 23 | Exercise Programs Delivered According to Guidelines Improve Mobility in People With Stroke: A Systematic Review and Meta-analysis | Inappropriate study design |
| 24 | Expert's Choice: 2018's Most Exciting Research in the Field of Pediatric Exercise Science | Inappropriate study design |
| 25 | Health benefits of hard martial arts in adults: a systematic review | Inappropriate outcome |
| 26 | Healthy brain, healthy life: a review of diet and exercise interventions to promote brain health and reduce Alzheimer's disease risk | Inappropriate study design |
| 27 | Heart Rate and Lower Limb Muscle Activity on Cycle Ergometer. / Frecuencia CardÍaca Y Actividad Muscular De Los Miembros Inferiores En CicloergÓmetro | Inappropriate study design |
| 28 | International Congress - CIDESD 2019 | Inappropriate study design |
| 29 | Intrarater Reliability and Agreement of the Physioflow Bioimpedance Cardiography Device during Rest, Moderate and High-Intensity Exercise | Inappropriate population |
| 30 | Neuropsychological Benefits of Neuro-Exergaming for Older Adults: A Pilot Study of an Interactive Physical and Cognitive Exercise System (iPACES) | Inappropriate population |

| | | |
|---|---|---|
| 31 | Normative values of functional fitness in non-active postmenopausal women. / Valores normativos de aptidão funcional em mulheres não ativas na pós-menopausa | Inappropriate study design |
| 32 | One-year unsupervised individualized exercise training intervention enhances cardiorespiratory fitness but not muscle deoxygenation or glycemic control in adults with type 1 diabetes | Inappropriate population |
| 33 | Perceived dietary salt intake and the risk of primary liver cancer: a population-based prospective study | Inappropriate study design |
| 34 | Physical Activity: Cornucopia and Conundrums | Inappropriate population |
| 35 | Physical exercise in the prevention and treatment of Alzheimer's disease | Inappropriate study design |
| 36 | Poor physical fitness is independently associated with mild cognitive impairment in elderly Koreans | Inappropriate study design |
| 37 | Potential Indirect Mechanisms of Cognitive Enhancement After Long-Term Resistance Training in Older Adults | Inappropriate study design |
| 38 | Relationship Between Sedentary Behavior and Physical Activity at Work and Cognition and Mood | Inappropriate study design |
| 39 | Self-Efficacy and Affective Responses to Acute Exercise in Middle-Aged Adults | Inappropriate study design |
| 40 | Self-Paced Cycling Improves Cognition on Institutionalized Older Adults Without Known Cognitive Impairment: A 15-Month Randomized Controlled Trial | Inappropriate study design |
| 41 | The Effect of Combined Aerobic Exercise and Calorie Restriction on Mood, Cognition, and Motor Behavior in Overweight and Obese Women | Inappropriate study design |
| 42 | The Effect of Exercise on Cancer-Related Cognitive Impairment and Applications for Physical Therapy: Systematic Review of Randomized Controlled Trials | Inappropriate study design |
| 43 | The Immediate Effects of Acute Aerobic Exercise on Cognition in Healthy Older Adults: A Systematic Review | Inappropriate population |
| 44 | The influence of exercise and physical fitness status on attention: a systematic review | Inappropriate population |
| 45 | The physiological and perceptual responses of stand-up paddle board exercise in a laboratory- and field-setting | Inappropriate population |
| 46 | Therapeutic Effects of Exercise Training on Elderly Patients With Dementia: A Randomized Controlled Trial | Inappropriate study design |
| 47 | Twelve weeks of resistance training does not influence peripheral levels of neurotrophic growth factors or homocysteine in healthy adults: a randomized-controlled trial | Inappropriate study design |
| 48 | Walking activity in community-dwelling stroke survivors within 1 month after discharge from a rehabilitation setting | Inappropriate population |
| 49 | A Feasibility Study Examining Use of the FitBit Zip™ vs. the Accusplit Eagle AC 120 XL Pedometer to Increase Physical Activity for Persons with Mild Cognitive Disorder | Inappropriate population |
| 50 | Acute cardiovascular responses to multiple sets of high-velocity resistance exercise in healthy adults | Inappropriate study design |
| 51 | Acute exercise has a general facilitative effect on cognitive function: A combined ERP temporal dynamics and BDNF study | Inappropriate population |
| 52 | Adherence to Exercise Prescription and Improvements in the Clinical and Vascular Health of African Americans | Inappropriate study design |
| 53 | Aerobic exercise increases hippocampal volume in older women with probable mild cognitive impairment: a 6-month randomised controlled trial | Inappropriate population |
| 54 | Benefits of Motor And/or Cognitive Exercise: a Review with Recommendations for the Third Age. / Prednosti in Koristi Gibalne In/ali Kognitivne Vadbe: Pregled PodroČja S PriporoČili Za Tretje ŽIvljenjsko Obdobje | Inappropriate study design |
| 55 | Can Resistance Training Improve Upper Limb Postural Tremor, Force Steadiness and Dexterity in Older Adults? A Systematic Review | Inappropriate study design |
| 56 | Changes in body image during a 6-month lifestyle behaviour intervention in a sample of overweight and obese individuals | Inappropriate study design |
| 57 | Chronic participation in externally paced, but not self-paced sports is associated with the modulation of domain-general cognition | Inappropriate population |
| 58 | Does a cycling program combined with education and followed by coaching promote physical activity in subacute stroke patients? A randomized controlled trial | Inappropriate study design |
| 59 | Dose–response effects of years of self-reported physical activity on old females' motor and cognitive function | Inappropriate study design |
| 60 | Dual-Task Exercise as a Therapy for Executive Motor Function in Parkinson's Disease | Inappropriate study design |

| | | |
|---|---|---|
| 61 | Effect of acute exercise and cardiovascular fitness on cognitive function: An event-related cortical desynchronization study | Inappropriate population |
| 62 | Effect of Queen's College Step Test on Cognitive Performance among Young Adults | Inappropriate study design |
| 63 | Effects of passive Bi-axial ankle stretching while walking on uneven terrains in older adults with chronic stroke | Inappropriate study design |
| 64 | Effects of resistance training and chess playing on the quality of life and cognitive performance of elderly women: a randomized controlled trial | Inappropriate study design |
| 65 | Effects of Upright and Recumbent Cycling on Executive Function and Prefrontal Cortex Oxygenation in Young Healthy Men | Inappropriate study design |
| 66 | Engaging Cognitive Circuits to Promote Motor Recovery in Degenerative Disorders. Exercise as a Learning Modality | Inappropriate population |
| 67 | Exercise and Academic Achievement in Children: Effects of Acute Class-Based Circuit Training | Inappropriate study design |
| 68 | Exercise and lifestyle predictors of resting heart rate in healthy young adults | Inappropriate study design |
| 69 | Exogenous ketone salts do not improve cognitive responses after a high-intensity exercise protocol in healthy college-aged males | Inappropriate population |
| 70 | Fitness Level Moderates Executive Control Disruption During Exercise Regardless of Age | Inappropriate study design |
| 71 | Goal-oriented instructions increase the intensity of practice in stroke rehabilitation compared with non-specific instructions: a within-participant, repeated measures experimental study | Inappropriate study design |
| 72 | Hallmarks of Improved Immunological Responses in the Vaccination of More Physically Active Elderly Females | Inappropriate study design |
| 73 | HIIT Improves Left Ventricular Exercise Response in Adults with Type 2 Diabetes | Inappropriate population |
| 74 | Impact of a dual task intervention on physical performance of older adults who practice physical exercise. / Impacto de uma intervenção de dupla tarefa no desempenho físico de idosos praticantes de exercício físico | Inappropriate population |
| 75 | Influence of Acute High-Intensity Aerobic Interval Exercise Bout on Selective Attention and Short-Term Memory Tasks | Inappropriate outcome |
| 76 | Influence of water depth on energy expenditure during aquatic walking in people post stroke | Inappropriate population |
| 77 | International Society of Sports Nutrition position stand: safety and efficacy of creatine supplementation in exercise, sport, and medicine | Inappropriate study design |
| 78 | Low Back Pain--Related Disability in Parkinson Disease: Impact on Functional Mobility, Physical Activity, and Quality of Life | Inappropriate study design |
| 79 | Non-physical approaches to counteract age-related functional deterioration: Applications for rehabilitation and neural mechanisms | Inappropriate study design |
| 80 | Noteworthy | Inappropriate study design |
| 81 | Objective aerobic fitness level and neuropsychological functioning in healthy adolescents and emerging adults: Unique sex effects | Inappropriate study design |
| 82 | Physical activity and cognitive function in older persons | Inappropriate study design |
| 83 | Physical Activity in the Prevention of Alzheimer's Disease | Inappropriate study design |
| 84 | Physical activity is associated with better global cognition and frontal function in overweight/obese older adults with metabolic syndrome | Inappropriate population |
| 85 | Physical Activity Throughout the Adult Life Span and Domain-Specific Cognitive Function in Old Age: A Systematic Review of Cross-Sectional and Longitudinal Data | Inappropriate intervention |
| 86 | Physical Activity, Injurious Falls, and Physical Function in Aging: An Umbrella Review | Inappropriate population |
| 87 | Self-Efficacy, Self-Regulation, Social Support, and Outcomes Expectations for Daily Physical Activity in Adults with Chronic Stroke: A Descriptive, Exploratory Study | Inappropriate study design |
| 88 | Sitting and Activity Time in People With Stroke | Inappropriate population |
| 89 | Social Influence, Physical Activity, and Social Cognitions Among Adults With Physical Disability: A Meta-Analysis | Inappropriate study design |
| 90 | Systematic review of the relationships between objectively measured physical activity and health indicators in school-aged children and youth | Inappropriate study design |
| 91 | The effect of group exercises on balance, mobility, and depressive symptoms in older adults with mild cognitive impairment: a randomized controlled trial | Inappropriate study design |
| 92 | The effectiveness of group exercise for improving activity and participation in adult stroke survivors: a systematic review | Inappropriate study design |

| | | |
|---|---|---|
| 93 | The Effects of Blood Flow Restriction Training on Functional Improvements in an Active Single Subject with Parkinson Disease | Inappropriate study design |
| 94 | The Effects of Maximal Intensity Exercise on Cognitive Performance in Children | Inappropriate population |
| 95 | The Link Between Physical Activity and Cognitive Dysfunction in Alzheimer Disease | Inappropriate study design |
| 96 | Two Days of Measurement Provides Reliable Estimates of Physical Activity Poststroke: An Observational Study | Inappropriate study design |
| 97 | Vascular-ventricular coupling during exercise is not affected by exaggerated blood pressures in endurance-trained athletes | Inappropriate population |
| 98 | Water-Based Exercise and Resistance Training Improve Cognition in Older Adults. / Hidrogimnasia Y Entrenamiento Resistido Mejoran La CogniciÓn De Personas De La Tercera Edad | Inappropriate study design |
| 99 | A home program of strength training, movement strategy training and education did not prevent falls in people with Parkinson's disease: a randomised trial | Inappropriate intervention |
| 100 | A multifaceted investigation into the effects of acute exercise on indices of brain function | Inappropriate intervention |
| 101 | Accelerometry Shows Inpatients With Acute Medical or Surgical Conditions Spend Little Time Upright and Are Highly Sedentary: Systematic Review | Inappropriate study design |
| 102 | Acute Cardiovascular Response during Resistance Exercise with Whole-body Vibration in Sedentary Subjects: A Randomized Cross-over Trial | Inappropriate population |
| 103 | Acute Effect of Caffeine Intake on Hemodynamics after Resistance Exercise in Young Non-hypertensive Subjects | Inappropriate outcome |
| 104 | Acute Physical Exercise Affected Processing Efficiency in an Auditory Attention Task More Than Processing Effectiveness | Inappropriate population |
| 105 | Aerobic training as a means to enhance inhibition: what's yet to be studied? | Inappropriate population |
| 106 | Barriers to Exercise in People With Parkinson Disease | Inappropriate study design |
| 107 | Best Core Stabilization for Anticipatory Postural Adjustment and Falls in Hemiparetic Stroke | Inappropriate population |
| 108 | Body composition and physical function after progressive resistance and balance training among older adults after stroke: an exploratory randomized controlled trial | Inappropriate study design |
| 109 | Changes in Executive Function After Acute Bouts of Passive Cycling in Parkinson's Disease | Inappropriate population |
| 110 | Cognitive Effects and Sedation | Inappropriate population |
| 111 | Combinations of physical activity, sedentary behaviour and sleep: relationships with health indicators in school-aged children and youth | Inappropriate study design |
| 112 | Dietary fat and antioxidant vitamin intake in patients of neurodegenerative disease in a rural region of Jalisco, Mexico | Inappropriate study design |
| 113 | Early blood pressure response to isometric exercise is attenuated in obese individuals who have undergone bariatric surgery | Inappropriate study design |
| 114 | Effect of Neuromuscular Electrical Stimulation on Brain-derived Neurotrophic Factor | Inappropriate study design |
| 115 | Effects of acute resistance exercise on cognition in late middle-aged adults: General or specific cognitive improvement? | Inappropriate population |
| 116 | Effects of aging and Parkinson's disease on motor unit remodeling: influence of resistance exercise training | Inappropriate study design |
| 117 | Effects of Rehabilitation in the Form of Dance and Movement Therapy on Nitric Oxide Levels in Elderly Woman | Inappropriate study design |
| 118 | Examining Self-Training Procedures in Leisure Swimming | Inappropriate study design |
| 119 | Facilitating Community-Based Exercise for People With Stroke: Cross-Sectional e-Survey of Physical Therapist Practice and Perceived Needs | Inappropriate study design |
| 120 | Feasibility and Effectiveness of a Walking Program for Community-Dwelling Older Adults With Mild Cognitive Impairment | Inappropriate study design |
| 121 | Functional and/or structural brain changes in response to resistance exercises and resistance training lead to cognitive improvements – a systematic review | Inappropriate study design |
| 122 | High-Intensity Progressive Resistance Training Increases Strength With No Change in Cardiovascular Function and Autonomic Neural Regulation in Older Adults | Inappropriate study design |
| 123 | High-Speed Resistance Training Modifies Load-Velocity and Load-Power Relationships in Parkinson's Disease | Inappropriate population |
| 124 | How Physically Active Are People Following Stroke? Systematic Review and Quantitative Synthesis | Inappropriate study design |
| 125 | Impact of negative cognitions about body image on inflammatory status in relation to health | Inappropriate study design |

| | | |
|---|---|---|
| 126 | Interval and continuous exercise elicit equivalent postexercise hypotension in prehypertensive men, despite differences in regulation | Inappropriate study design |
| 127 | Near-infrared spectroscopy reveals link between chronic physical activity and anterior frontal oxygenated hemoglobin in healthy young women | Inappropriate study design |
| 128 | Objectively Measured Physical Activity and Cognitive Function in Older Adults | Inappropriate outcome |
| 129 | Occupational and leisure-time physical activity differentially predict 6-year incidence of stroke and transient ischemic attack in women | Inappropriate study design |
| 130 | Physical Activity and Fitness Effects on Cognition and Brain Health in Children and Older Adults | Inappropriate study design |
| 131 | Physical Activity and the Risk of Depression in Community-Dwelling Korean Adults With a History of Stroke | Inappropriate population |
| 132 | Physical Activity Levels and Their Associations With Postural Control in the First Year After Stroke | Inappropriate study design |
| 133 | Physiological Dynamic Apnea Responses in Relation to Apnea Capacity in Triathletes | Inappropriate study design |
| 134 | Reducing Sitting Time After Stroke: A Phase II Safety and Feasibility Randomized Controlled Trial | Inappropriate population |
| 135 | Research Report. Factors Contributing to 50-ft Walking Speed and Observed Ethnic Differences in Older Community-Dwelling Mexican Americans and European Americans | Inappropriate population |
| 136 | The Acute Effects of Yoga on Executive Function | Inappropriate population |
| 137 | The Association Between Decline in Physical Functioning and Atrophy of Medial Temporal Areas in Community-Dwelling Older Adults With Amnestic and Nonamnestic Mild Cognitive Impairment | Inappropriate study design |
| 138 | The discordance between subjectively and objectively measured physical function in women with fibromyalgia: association with catastrophizing and self-efficacy cognitions. The al-Ándalus project | Inappropriate population |
| 139 | The effect of 1 year of Alagebrium and moderate-intensity exercise training on left ventricular function during exercise in seniors: a randomized controlled trial | Inappropriate population |
| 140 | The effect of functional stretching exercises on functional outcomes in spastic stroke patients: A randomized controlled clinical trial | Inappropriate study design |
| 141 | The effectiveness of a video-supported group-based Otago exercise programme on physical performance in community-dwelling older adults: a preliminary study | Inappropriate study design |
| 142 | The Effects of Prolonged Physical Inactivity Induced by Bed Rest on Cognitive Functioning in Healthy Male Participants. / UČinki Dolgotrajne Gibalne Neaktivnosti V Pogojih Simulirane BrezteŽnosti Na Kognitivno Delovanje Pri Zdravih UdeleŽencih | Inappropriate population |
| 143 | The Healthy Mind, Healthy Mobility Trial: A Novel Exercise Program for Older Adults | Inappropriate population |
| 144 | The joint impact of habitual exercise and glycemic control on the incidence of chronic kidney disease (CKD) in middleaged and older males | Inappropriate study design |
| 145 | The Prevalence, Measurement, and Treatment of the Cognitive Dimension/Domain in Major Depressive Disorder | Inappropriate population |
| 146 | The relationship between cognitive and physical function among residents of a Czech senior home | Inappropriate population |
| 147 | Active Workstations to Fight Sedentary Behaviour | Inappropriate study design |
| 148 | Adaptive physical activity improves mobility function and quality of life in chronic hemiparesis | Inappropriate study design |
| 149 | Aerobic Exercise to Improve Cognitive Function in Adults With Neurological Disorders: A Systematic Review | Inappropriate study design |
| 150 | Bilateral upper-limb rehabilitation after stroke using a movement-based game controller | Inappropriate population |
| 151 | Cardiac Work Remains High after Strength Exercise in Elderly | Inappropriate study design |
| 152 | Cardio-pulmonary Responses to Increasing Workload Exercise on a Cycle Ergometer in Healthy Men | Inappropriate population |
| 153 | Cardiovascular drift in euhydrated prepubertal boys | Inappropriate outcome |
| 154 | Cardiovascular Responses During Downhill Treadmill Walking at Self-Selected Intensity in Older Adults | Inappropriate study design |
| 155 | Central and Peripheral Cardiovascular Impairments Limit V̇O2peak in Type 1 Diabetes | Inappropriate population |
| 156 | Cerebral/Peripheral Vascular Reactivity and Neurocognition in Middle-Age Athletes | Inappropriate population |
| 157 | Cerebrovascular reactivity is associated with maximal aerobic capacity in healthy older adults | Inappropriate population |

| | | |
|---|---|---|
| 158 | Determinants of Satisfaction With Community Reintegration in Older Adults With Chronic Stroke: Role of Balance Self-Efficacy | Inappropriate population |
| 159 | Effect of aging on hemodynamic response to metaboreflex activation | Inappropriate study design |
| 160 | Effect of Resistance-Exercise Training on Cognitive Function in Healthy Older Adults: A Review | Inappropriate study design |
| 161 | Effects of Intensive Whole-Body Vibration Training on Muscle Strength and Balance in Adults With Chronic Stroke: A Randomized Controlled Pilot Study | Inappropriate population |
| 162 | Effects of resistance training in gray matter density of elderly | Inappropriate study design |
| 163 | Effects of Task-Specific Locomotor and Strength Training in Adults Who Were Ambulatory After Stroke: Results of the STEPS Randomized Clinical Trial | Inappropriate study design |
| 164 | Effects of Twice-Weekly Intense Aerobic Exercise in Early Subacute Stroke: A Randomized Controlled Trial | Inappropriate outcome |
| 165 | Effects of two different intensities of aerobic exercise on elderly people with mild cognitive impairment: a randomized pilot study | Inappropriate study design |
| 166 | Endurance performance in masters athletes | Inappropriate study design |
| 167 | Exercise training at the maximal fat oxidation intensity improved health-related physical fitness in overweight middle-aged women | Inappropriate study design |
| 168 | Exercise, brain, and cognition across the life span | Inappropriate study design |
| 169 | Feasibility of Measuring Ventilatory Threshold in Adults With Stroke-Induced Hemiparesis: Implications for Exercise Prescription | Inappropriate study design |
| 170 | Fitness Effects on the Cognitive Function of Older Adults: A Meta–Analytic Study | Inappropriate study design |
| 171 | Improved Physical Fitness Correlates With Improved Cognition in Multiple Sclerosis | Inappropriate study design |
| 172 | Investigating the acute effect of an aerobic dance exercise program on neuro-cognitive function in the elderly | Inappropriate study design |
| 173 | Irregular 24-hour activity rhythms and the metabolic syndrome in older adults | Inappropriate outcome |
| 174 | Leg Muscle Activity during Whole-Body Vibration in Individuals with Chronic Stroke | Inappropriate study design |
| 175 | Long-term Mild Exercise Training Enhances Hippocampus-dependent Memory in Rats | Inappropriate population |
| 176 | Molecular mechanisms of life- and health-span extension: role of calorie restriction and exercise intervention | Inappropriate population |
| 177 | Patient Active Time During Therapy Sessions in Postacute Rehabilitation: Development and Validation of a New Measure | Inappropriate population |
| 178 | Patterns of Sedentary Behavior in US Middle-Age and Older Adults: The REGARDS Study | Inappropriate study design |
| 179 | Physical Activity and Cognition in Older Adults: The Potential of Tai Chi Chuan | Inappropriate study design |
| 180 | Physical activity and health: Metabolic and cardiovascular issues | Inappropriate population |
| 181 | Physical Inactivity During Leisure Time Among Older Adults-- Behavioral Risk Factor Surveillance System, 2005 | Inappropriate population |
| 182 | Resistance Exercise Enhances Cognitive Function in Mouse | Inappropriate study design |
| 183 | Resistance Training Does not have an Effect on Cognition or Related Serum Biomarkers in Nonagenarians: A Randomized Controlled Trial | Inappropriate population |
| 184 | Role of the Mirror-Neuron System in Cross-Education | Inappropriate study design |
| 185 | Sedentary versus active behavior in people after stroke | Inappropriate study design |
| 186 | Sex differences in ventricular-vascular coupling following endurance training | Inappropriate study design |
| 187 | Tai Ji Quan as an exercise modality to prevent and manage cardiovascular disease: A review | Inappropriate population |
| 188 | Tai Ji Quan, the brain, and cognition in older adults | Inappropriate study design |
| 189 | Taking up physical activity in later life and healthy ageing: the English longitudinal study of ageing | Inappropriate study design |
| 190 | The accumulation of healthy lifestyle behaviors prevents the incidence of chronic kidney disease (CKD) in middle-aged and older males | Inappropriate study design |
| 191 | The Role of Mobility as a Protective Factor of Cognitive Functioning in Aging Adults: A Review | Inappropriate population |
| 192 | The Structure of Walking Activity in People After Stroke Compared With Older Adults Without Disability: A Cross-Sectional Study | Inappropriate study design |
| 193 | Training in a shallow pool: Its effect on upper extremity strength and total body weight in postmenopausal women | Inappropriate population |
| 194 | Walking or vitamin B for cognition in older adults with mild cognitive impairment? A randomised controlled trial | Inappropriate study design |

| | | |
|---|---|---|
| 195 | A review of the acute cardiovascular responses to resistance exercise of healthy young and older adults | Inappropriate study design |
| 196 | Abstracts | Inappropriate population |
| 197 | Asthma and the Achievement Gap Among Urban Minority Youth | Inappropriate study design |
| 198 | Ballistic strength training compared with usual care for improving mobility following traumatic brain injury: protocol for a randomised, controlled trial | Inappropriate study design |
| 199 | Benefits of exercise for community-dwelling older adults | Inappropriate population |
| 200 | Blood pressure response to isometric exercise in patients with peripheral atherosclerotic disease | Inappropriate study design |
| 201 | Cardiac responses to progressive upright exercise in adult male cyclists | Inappropriate population |
| 202 | Cardiovascular fitness in premenarcheal girls and young women. / Etat de sante cardiovasculaire chez des filles prepuberes et des jeunes femmes | Inappropriate population |
| 203 | Cardiovascular Responses During Karate Exercise Regimen and Treadmill Exercise at Approximately 70% HR Intensity | Inappropriate study design |
| 204 | Cardiovascular responses to exercise in children | Inappropriate study design |
| 205 | Central and peripheral cardiovascular adaptations during a maximal cycle exercise in boys and men. / Adaptations cardiovasculaires centrales et peripheriques pendant un exercice maximal sur cycle chez les adolescents et les adultes de sexe masculin | Inappropriate study design |
| 206 | Changes in rowing technique over a routine one hour low intensity high volume training session | Inappropriate study design |
| 207 | Cognitive Performance in Older Women Relative to ApoE-[varepsilon]4 Genotype and Aerobic Fitness | Inappropriate population |
| 208 | Correlations Between Aerobic Capacity, Pulmonary and Cognitive Functioning in the Older Women | Inappropriate study design |
| 209 | Determinants of muscle metaboreflex and involvement of baroreflex in boys and young men | Inappropriate population |
| 210 | Dose response between total volume of physical activity and health and fitness. / La relation dose-effet entre le volume global de l ' activite physique, la sante et la condition physique | Inappropriate study design |
| 211 | Effect of Gravity on Robot-Assisted Motor Training After Chronic Stroke: A Randomized Trial | Inappropriate study design |
| 212 | Effect of Tai Chi on Physical Function, Fall Rates and Quality of Life Among Older Stroke Survivors | Inappropriate outcome |
| 213 | Effects of Acute Resistance Exercise on Late-Middle-Age Adults' Goal Planning | Inappropriate study design |
| 214 | Effects of home-based sensory and motor amplitude electrical stimulation on arm dysfunction in chronic stroke | Inappropriate population |
| 215 | Efficacy, Attributional, and Affective Responses to Exercise Participation | Inappropriate outcome |
| 216 | Energy Expenditure in Adults With Cerebral Palsy Playing Wii Sports | Inappropriate study design |
| 217 | Exercise and cognition in older adults: is there a role for resistance training programmes? | Inappropriate study design |
| 218 | Exercise Training is Beneficial for Alzheimer's Patients | Inappropriate study design |
| 219 | Food Intake Does Not Differ between Obese Women Who Are Metabolically Healthy or Abnormal | Inappropriate population |
| 220 | From Tibialis Anterior to Tai Chi: Biofeedback and Beyond | Inappropriate study design |
| 221 | Impact of resistance exercise program on functional capacity and muscular strength of knee extensor in pre-frail community-dwelling older women: a randomized crossover trial. / Efeito de um programa de resistência muscular na capacidade funcional e na força muscular dos extensores do joelho em idosas pré-frágeis da comunidade: ensaio clínico aleatorizado do tipo crossover | Inappropriate population |
| 222 | Individual Differences in Behaviorally Evoked Cardiovascular Response: Temporal Stability and Hemodynamic Patterning | Inappropriate study design |
| 223 | Nebivolol: Haemodynamic Effects and Clinical Significance of Combined β-Blockade and Nitric Oxide Release | Inappropriate study design |
| 224 | Pathogénie et épidémiologie de l’hypertension artérielle | Inappropriate study design |
| 225 | Physical activity and aging: retrospections and visions for the future | Inappropriate study design |
| 226 | Physical activity and mental well-being in older people participating in the Better Ageing Project | Inappropriate population |
| 227 | Physical Activity and Stroke | Inappropriate study design |
| 228 | Physical activity, body composition and healthy ageing. / Exercice, composition corporelle et vieillissement | Inappropriate population |

| | | |
|---|---|---|
| 229 | Promoting smoking cessation in the rehabilitation setting | Inappropriate population |
| 230 | Relationship of low plasma klotho with poor grip strength in older community-dwelling adults: the InCHIANTI study | Inappropriate population |
| 231 | Repeatability of Aerobic Capacity Measurements in Parkinson Disease | Inappropriate study design |
| 232 | Reproducibility of cardiac output measurements by Doppler echocardiography in prepubertal children and adults. / Reproductibilite des mesures du debit cardiaque sur une echocardiographie-doppler chez des pre-adolescents et des adultes | Inappropriate study design |
| 233 | Resistance training and gait function in patients with Parkinson's disease | Inappropriate population |
| 234 | Risk Due to Inactivity in Physically Capable Older Adults | Inappropriate study design |
| 235 | Routine physiotherapy does not induce a cardiorespiratory training effect post-stroke, regardless of walking ability | Inappropriate intervention |
| 236 | Strategies for Aging Well | Inappropriate population |
| 237 | Strength deficit of knee flexors is dependent on hip position in adults with chronic hemiparesis | Inappropriate study design |
| 238 | Stroke-related knowledge, lifestyle behaviours and health beliefs in Singaporean Chinese: Implications for health education | Inappropriate population |
| 239 | The Combination of Dietary Conjugated Linoleic Acid and Treadmill Exercise Lowers Gain in Body Fat Mass and Enhances Lean Body Mass in High Fat-Fed Male Balb/C Mice | Inappropriate study design |
| 240 | The effect of acute physical exercise on cognitive function during development | Inappropriate population |
| 241 | The Effects of Exercise on Cognition in Older Adults With and Without Cognitive Decline: A Systematic Review | Inappropriate outcome |
| 242 | The influence of diabetes mellitus on cardiovascular function in children and adolescents | Inappropriate population |
| 243 | A guideline for cardiopulmonary conditioning in the middle-aged recreational athlete | Inappropriate study design |
| 244 | Abstract 553 | Inappropriate population |
| 245 | Aerobic exercise enhances executive function and academic achievement in sedentary, overweight children aged 7–11 years | Inappropriate outcome |
| 246 | Aquatic exercise programs for patients with multiple sclerosis | Inappropriate population |
| 247 | Cardiac Function During Exercise in Obese Prepubertal Boys: Effect of Degree of Obesity | Inappropriate population |
| 248 | Cardiac responses to exercise in child distance runners | Inappropriate study design |
| 249 | Cardiac responses to progressive exercise in normal children: a synthesis. / Reponses cardiaques a l'effort progressif chez des enfants en bonne sante: synthese | Inappropriate study design |
| 250 | Cardiorespiratory fitness and walking ability in subacute stroke patients | Inappropriate population |
| 251 | Cardiovascular responses to static exercise in boys: insights from tissue Doppler imaging | Inappropriate population |
| 252 | Comparison of cardiorespiratory responses to open-water and simulated kayaking | Inappropriate population |
| 253 | Computerized Adaptive Testing for Follow-Up After Discharge From Inpatient Rehabilitation: I. Activity Outcomes | Inappropriate population |
| 254 | Dynamics of changes in the cardiovascular response to submaximal exercise during low-intensity endurance training with particular reference to the systolic time intervals | Inappropriate study design |
| 255 | Effect of exercise training on ambulatory blood pressure. / Effet de l ' entrainement physique sur la pression du sang ambulatoire | Inappropriate study design |
| 256 | Effect of head-out water immersion on response to exercise training | Inappropriate population |
| 257 | Effect of long-term intense swimming training on the upper body peak oxygen uptake of prepubertal girls. / Effet d ' un entrainement intense et de longue duree en natation sur le pic d ' oxygene du corps superieur de filles prepubertaires | Inappropriate study design |
| 258 | Effectiveness of training programmes for prepubescent children | Inappropriate study design |
| 259 | Effects of a short-term strength training on circulatory responses to standard static and dynamic exercises | Inappropriate study design |
| 260 | Evidence-informed physical activity guidelines for Canadian adults | Inappropriate outcome |
| 261 | From the Centers for Disease Control and Prevention (Cdc) | Inappropriate population |
| 262 | Hand dominance and side of stroke affect rehabilitation in chronic stroke | Inappropriate study design |

| | | |
|---|---|---|
| 263 | Health effects of recreational running in women. Some epidemiological and preventive aspects. / Effets de la course a pied de loisirs sur la sante des femmes - quelques aspects epidemiologiques et de la prevention | Inappropriate population |
| 264 | Influence of breathing technique on arterial blood pressure during heavy weight lifting | Inappropriate study design |
| 265 | Is It Possible to Individualize Intensity of Eccentric Cycling Exercise From Perceived Exertion on Concentric Test? | Inappropriate study design |
| 266 | Myocardial diastolic function and exercise. / Fonction diastolique du myocarde et exercice physique | Inappropriate study design |
| 267 | Neuroendocrine system and mental function in sedentary and endurance-trained elderly males. / Systeme neuroendocrinien et fonction mentale chez les hommes ages sedentaires et entraines | Inappropriate population |
| 268 | Other Headaches and Pain | Inappropriate population |
| 269 | Outcomes of progressive resistance strength training following stroke: a systematic review | Inappropriate study design |
| 270 | Physiological considerations in training young athletes | Inappropriate population |
| 271 | Poster Session 1-3: Diagnostic Tools and Risk Stratification Monday, April 3, 2006, 2:00–3:30pm | Inappropriate population |
| 272 | Rate and Amplitude of Adaptation to Two Intensities of Exercise in Men Aged 65-75 yr | Inappropriate population |
| 273 | The cardiovascular capacities of adults with Down syndrome: a comparative study. / Les capacites cardiovasculaires des adultes porteurs de la trisomie 21 : etude comparative | Inappropriate study design |
| 274 | The Effects of Exercise Programs on Cognition in Older Adults: A Review | Inappropriate study design |
| 275 | The Influence of Non-Paretic Leg Movement on Muscle Action in the Paretic Leg of Hemiplegic Patients | Inappropriate population |
| 276 | The role of the right ventricle during hypobaric hypoxic exercise: insights from patients after the Fontan operation | Inappropriate study design |
| 277 | Tracking and explanation of physical activity in young adults over a 7-year period. / Etude de l ' activite physique des jeunes adultes sur une periode de 7 ans | Inappropriate population |
| 278 | Treadmill training improves fitness reserve in chronic stroke patients | Inappropriate population |
| 279 | The effect of Hatha yoga intervention on students' creative ability | Inappropriate study design |
| 280 | Sensor-measured sedentariness and physical activity are differentially related to fluid and crystallized abilities in aging | Inappropriate population |
| 281 | Distinctive effects of aerobic and resistance exercise modes on neurocognitive and biochemical changes in individuals with mild cognitive impairment | Inappropriate study design |
| 282 | The effect of low-intensity aerobic training on cognitive functions of severely deconditioned subacute and chronic stroke patients: A randomized, controlled pilot study | Inappropriate study design |
| 283 | Does cardiorespiratory fitness influence the effect of acute aerobic exercise on executive function? [References]. | Inappropriate population |
| 284 | Depression in Parkinson's disease: Perspectives from an Australian cohort | Inappropriate population |
| 285 | Using intraindividual variability as an indicator of cognitive improvement in a physical exercise intervention of older women with mild cognitive impairment | Inappropriate population |
| 286 | Acute exercise increases circulating IGF-1 in Alzheimer's disease patients, but not in older adults without dementia | Inappropriate population |
| 287 | Comparison of the effects of running and badminton on executive function: A within-subjects design | Inappropriate study design |
| 288 | The effects of an aerobic training intervention on cognition, grey matter volumes and white matter microstructure | Inappropriate population |
| 289 | Acute aerobic exercise impairs aspects of cognitive function at high altitude | Inappropriate study design |
| 290 | Exercise intensity-dependent effects of arm and leg-cycling on cognitive performance | Inappropriate intervention |
| 291 | Moderators of cognitive outcomes from an exercise program in people with mild to moderate dementia | Inappropriate study design |
| 292 | Effects of laughter therapy on quality of life in patients with cancer: An open-label, randomized controlled trial | Inappropriate study design |
| 293 | Five-year cost-effectiveness analysis of the European Fans in Training (EuroFIT) physical activity intervention for men versus no intervention | Inappropriate study design |
| 294 | The effects of physical exercise on cognition: How heart rate variability can predict cognitive performances | Inappropriate population |
| 295 | Accelerometry-assessed physical activity and sedentary time and associations with chronic disease and hospital visits-A prospective cohort study with 15 years follow-up | Inappropriate study design |

| | | |
|---|---|---|
| 296 | Effect of acute moderate-intensity exercise on the mirror neuron system: Role of cardiovascular fitness level | Inappropriate study design |
| 297 | A single bout of moderate intensity exercise improves cognitive flexibility: Evidence from task-switching | Inappropriate study design |
| 298 | Effects of acute aerobic and resistance exercise on executive function: An ERP study | Inappropriate population |
| 299 | Aerobic exercise improves cognition and cerebrovascular regulation in older adults | Inappropriate study design |
| 300 | Walking time is associated with hippocampal volume in overweight and obese office workers | Inappropriate study design |
| 301 | Individuals with stroke improve anticipatory postural adjustments after a single session of targeted exercises | Inappropriate study design |
| 302 | Healthy lifestyle and the risk of Alzheimer dementia: Findings from 2 longitudinal studies | Inappropriate study design |
| 303 | Effects of exergaming on physical and cognitive functions in individuals with autism spectrum disorder: A systematic review | Inappropriate study design |
| 304 | Does aerobic fitness moderate age-related cognitive slowing? Evidence from the P3 and lateralized readiness potentials | Inappropriate study design |
| 305 | Better executive function is associated with faster on-transition aerobic metabolism among older adults | Inappropriate population |
| 306 | Brain perfusion change in patients with mild cognitive impairment after 12 months of aerobic exercise training | Inappropriate study design |
| 307 | Effects of aerobic exercise as add-on treatment for inpatients with moderate to severe depression on depression severity, sleep, cognition, psychological well-being, and biomarkers: Study protocol, description of study population, and manipulation check | Inappropriate study design |
| 308 | Physical activity and sedentary behaviour in a residential aged care facility | Inappropriate population |
| 309 | Tic frequency and behavioural measures of cognitive control are improved in individuals with Tourette syndrome by aerobic exercise training | Inappropriate population |
| 310 | Heart rate variability mainly relates to cognitive executive functions and improves through exergame training in older adults: A secondary analysis of a 6-month randomized controlled trial | Inappropriate study design |
| 311 | The positive effect of moderate-intensity exercise on the mirror neuron system: An fNIRS study | Inappropriate population |
| 312 | The immediate effect of stroboscopic visual training on information-processing time in people with multiple sclerosis: An exploratory study | Inappropriate population |
| 313 | Yoga for veterans with PTSD: Cognitive functioning, mental health, and salivary cortisol | Inappropriate population |
| 314 | ABCA7 genotype moderates the effect of aerobic exercise intervention on generalization of prior learning in healthy older African Americans | Inappropriate outcome |
| 315 | Empirical analysis on the positive correlation between physical exercise and the mental health of college students | Inappropriate population |
| 316 | Late-life depression and the risk of dementia in 14 countries: A 10-year follow-up study from the Survey of Health, Ageing and Retirement in Europe | Inappropriate study design |
| 317 | Effects of simultaneously performed dual-task training with aerobic exercise and working memory training on cognitive functions and neural systems in the elderly | Inappropriate population |
| 318 | A single 30 minutes bout of combination physical exercises improved inhibition and vigor-mood in middle-aged and older females: Evidence from a randomized controlled trial | Inappropriate study design |
| 319 | Ballroom dance for persons with multiple sclerosis: A pilot feasibility study | Inappropriate study design |
| 320 | Fitness level influences white matter microstructure in postmenopausal women | Inappropriate population |
| 321 | Longer term effects of diet and exercise on neurocognition: 1-year follow-up of the ENLIGHTEN trial | Inappropriate outcome |
| 322 | Conservative and disruptive modes of adolescent change in human brain functional connectivity | Inappropriate study design |
| 323 | Psychophysiological effects of dance movement therapy and physical exercise on older adults with mild dementia: A randomized controlled trial | Inappropriate study design |
| 324 | Resistance training in patients with schizophrenia: Concept and proof of principle trial | Inappropriate study design |
| 325 | The effects of acute exercise on retroactive memory interference | Inappropriate population |
| 326 | Post-exertional malaise in veterans with gulf war illness | Inappropriate study design |
| 327 | Acute effects of resistance and functional-task exercises on executive function of obese older adults: Two counterbalanced, crossover, randomized exploratory studies | Inappropriate population |

| | | |
|---|---|---|
| 328 | A multi-ingredient nutritional supplement in combination with resistance exercise and high-intensity interval training improves cognitive function and increases n-3 index in healthy older men: A randomized controlled trial | Inappropriate study design |
| 329 | The relationship between moderate to vigorous physical activity and cognitive rigidity in anorexia nervosa | Inappropriate study design |
| 330 | Aerobic exercise and cognitive functioning in schizophrenia: A pilot randomized controlled trial | Inappropriate study design |
| 331 | Effect of aerobic exercise on white matter microstructure in the aging brain | Inappropriate study design |
| 332 | Beneficial effects of acute high-intensity exercise on electrophysiological indices of attention processes in young adult men | Inappropriate study design |
| 333 | A single bout of hard RPE-based cycling exercise increases salivary alpha-amylase | Inappropriate population |
| 334 | Exercise, ageing and cognitive function-Effects of a personalized physical exercise program in the cognitive function of older adults | Inappropriate study design |
| 335 | Changes in moderate intensity physical activity are associated with better cognition in the Multilevel Intervention for Physical Activity in Retirement Communities (MIPARC) study | Inappropriate study design |
| 336 | Exercise and cognitive training as a strategy to improve neurocognitive outcomes in heart failure: A pilot study | Inappropriate study design |
| 337 | Effects of caffeine ingestion on the diurnal variation of cognitive and repeated high-intensity performances | Inappropriate population |
| 338 | Fractal properties and short-term correlations in motor control in cycling: Influence of a cognitive challenge | Inappropriate study design |
| 339 | Two days of calorie deprivation impairs high level cognitive processes, mood, and self-reported exertion during aerobic exercise: A randomized double-blind, placebo-controlled study | Inappropriate population |
| 340 | Reporting and methodological quality of clinical trials on exercise therapy for Parkinson's disease | Inappropriate study design |
| 341 | Intensity and workload related dose-response effects of acute resistance exercise on domain-specific cognitive function and affective response-A four-armed randomized controlled crossover trial | Inappropriate study design |
| 342 | Exercise training in amnestic mild cognitive impairment: A one-year randomized controlled trial | Inappropriate population |
| 343 | Brain Vitality Enhancement (BRAVE) program to promote brain health among persons with mild cognitive impairment: A study protocol | Inappropriate population |
| 344 | Technology, physical activity, loneliness, and cognitive functioning in old age | Inappropriate study design |
| 345 | Effects of a moderate-intensity aerobic exercise programme on the cognitive function and quality of life of community-dwelling elderly people with mild cognitive impairment: A randomised controlled trial | Inappropriate population |
| 346 | Association among 2-min step test, functional level and diagnosis of dementia | Inappropriate study design |
| 347 | Resting cerebral blood flow after exercise training in mild cognitive impairment | Inappropriate population |
| 348 | Yoga and meditation can help improve cognitive functioning in older adults with mild cognitive impairment and dementia | Inappropriate study design |
| 349 | Effects of exercise on symptoms of anxiety, cognitive ability and sick leave in patients with anxiety disorders in primary care: Study protocol for PHYSBI, a randomized controlled trial | Inappropriate study design |
| 350 | Intensity-dependent effects of acute exercise on executive function | Inappropriate study design |
| 351 | Light aerobic exercise modulates executive function and cortical excitability | Inappropriate study design |
| 352 | Improving fitness increases dentate gyrus/CA3 volume in the hippocampal head and enhances memory in young adults | Inappropriate study design |
| 353 | The Neurological Exercise Training (NExT) program: A pilot study of a community exercise program for survivors of stroke | Inappropriate population |
| 354 | Effects of an intensive exercise-based swallowing program for persons with Parkinson's disease and complex medical history: A single-case experiment | Inappropriate study design |
| 355 | Lifestyle and neurocognition in older adults with cognitive impairments: A randomized trial | Inappropriate study design |
| 356 | Survivor-perceived motivational facilitators and barriers to participation in cognitive exercise following chronic acquired brain injury | Inappropriate outcome |
| 357 | Acute effects of aerobic exercise on executive function and attention in adult patients with ADHD | Inappropriate study design |
| 358 | Exercise intervention in PTSD: A narrative review and rationale for implementation | Inappropriate study design |
| 359 | Evaluating the effects of exercise on cognitive function in hypertensive and diabetic patients using the mental test and training system | Inappropriate study design |

| | | |
|---|---|---|
| 360 | The effects of acute yoga versus aerobic exercise on executive function: A pilot study | Inappropriate population |
| 361 | Transferability and sustainability of motor-cognitive dual-task training in patients with dementia: A randomized controlled trial | Inappropriate study design |
| 362 | Influence of slow-paced breathing on inhibition after physical exertion | Inappropriate study design |
| 363 | Cognitive and physical factors affecting daily function in Alzheimer's disease: A cross-sectional analysis | Inappropriate intervention |
| 364 | Acute moderate-intensity exercise generally enhances attentional resources related to perceptual processing | Inappropriate study design |
| 365 | Aerobic exercise improves attention and quality of sleep among professional volleyball players | Inappropriate study design |
| 366 | Profiles in paint: Contrasting responses to a common artistic exercise by people with different dementias | Inappropriate study design |
| 367 | Ethnic differences in the cardiac responses to aerobic exercise | Inappropriate population |
| 368 | Exercise and quality of life in dementia: A systematic review and meta-analysis of randomized controlled trials | Inappropriate study design |
| 369 | Effects of mind-body exercises on cognitive function in older adults: A meta-analysis | Inappropriate study design |
| 370 | Effect of acute physical exercise on executive functions and emotional recognition: Analysis of moderate to high intensity in young adults | Inappropriate outcome |
| 371 | Effects of mindfulness yoga vs stretching and resistance training exercises on anxiety and depression for people with Parkinson disease a randomized clinical trial | Inappropriate study design |
| 372 | Changes in cognition over a 16.1 km cycling time trial using Think Aloud protocol: Preliminary evidence | Inappropriate study design |
| 373 | Effects of acute exercise on executive functioning: Testing the moderators | Inappropriate population |
| 374 | High-intensity interval training in people with Parkinson's disease: A randomized, controlled feasibility trial | Inappropriate population |
| 375 | Effects of combined intervention of physical exercise and cognitive training on cognitive function in stroke survivors with vascular cognitive impairment: A randomized controlled trial | Inappropriate study design |
| 376 | Progression of aphasia severity in the chronic stages of stroke | Inappropriate population |
| 377 | Aerobic exercise training improves cerebral blood flow and executive function: A randomized, controlled cross-over trial in sedentary older men | Inappropriate study design |
| 378 | Effects of a brief stair-climbing intervention on cognitive performance and mood states in healthy young adults | Inappropriate study design |
| 379 | Decrease in attentional performance after repeated bouts of high intensity exercise in association-football referees and assistant referees | Inappropriate study design |
| 380 | Differential treatment effects of an integrated motivational interviewing and exercise intervention on depressive symptom profiles and associated factors: A randomised controlled cross-over trial among youth with major depression | Inappropriate study design |
| 381 | Resistance training enhances delayed memory in healthy middle-aged and older adults: A randomised controlled trial | Inappropriate population |
| 382 | Study protocol: Does an acute intervention of high-intensity physical exercise followed by a brain training video game have immediate effects on brain activity of older people during Stroop task in fMRI?-A randomized controlled trial with crossover design | Inappropriate intervention |
| 383 | Effect of aerobic exercise on cognition in younger adults: A randomized clinical trial | Inappropriate study design |
| 384 | Effects of acute exercise duration on the inhibition aspect of executive function in late middle-aged adults | Inappropriate study design |
| 385 | Effects of exercise modes on neural processing of working memory in late middle-aged adults: An fMRI study | Inappropriate study design |
| 386 | A randomized controlled trial of a walking training with simultaneous cognitive demand (dual-task) in chronic stroke | Inappropriate study design |
| 387 | Differential acute effect of high-intensity interval or continuous moderate exercise on cognition in individuals with Parkinson's disease | Inappropriate study design |
| 388 | Social cognitive effects and mediators of a pilot telephone counseling intervention to increase aerobic exercise in hematologic cancer survivors | Inappropriate study design |
| 389 | Inpatient enhanced multidisciplinary care effects on the quality of life for Parkinson disease: A quasi-randomized controlled trial | Inappropriate population |
| 390 | Is improved fitness following a 12-week exercise program associated with decreased symptom severity, better wellbeing, and fewer sleep complaints in patients with major depressive disorders? A secondary analysis of a randomized controlled trial | Inappropriate population |

| | | |
|---|---|---|
| 391 | ABCA7 risk genotype diminishes the neuroprotective value of aerobic fitness in healthy older African Americans | Inappropriate outcome |
| 392 | Cognitive performance, aerobic fitness, motor proficiency, and brain function among children newly diagnosed with craniopharyngioma | Inappropriate study design |
| 393 | Feasibility of, adherence to, and satisfaction with video game versus traditional self-training of the upper extremity in people with chronic stroke: A pilot randomized controlled trial | Inappropriate study design |
| 394 | Pupillometric indices of locus-coeruleus activation are not modulated following single bouts of exercise | Inappropriate study design |
| 395 | Dynamic office environments improve brain activity and attentional performance mediated by increased motor activity | Inappropriate study design |
| 396 | Cognition in breast cancer survivors: A pilot study of interval and continuous exercise | Inappropriate study design |
| 397 | Older adults elicit a single-bout post-exercise executive benefit across a continuum of aerobically supported metabolic intensities | Inappropriate population |
| 398 | Physical activity pre- and post-dementia: English Longitudinal Study of Ageing | Inappropriate population |
| 399 | Effects of a multicomponent exercise program in institutionalized elders with Alzheimer's disease | Inappropriate study design |
| 400 | Aerobic exercise increases cortisol awakening response in older adults | Inappropriate population |
| 401 | Effects of acute aerobic exercise on cognition and constructs of decision-making in adults with and without hypertension | Inappropriate study design |
| 402 | Functional MRS with J-edited lactate in human motor cortex at 4 T | Inappropriate population |
| 403 | The influence of aerobic fitness on top-down and bottom-up mechanisms of interference control | Inappropriate study design |
| 404 | Effects of low and moderate acute resistance exercise on executive function in community-living older adults | Inappropriate study design |
| 405 | Endurance exercise enhances emotional valence and emotion regulation | Inappropriate study design |
| 406 | Two-day fasting evokes stress, but does not affect mood, brain activity, cognitive, psychomotor, and motor performance in overweight women | Inappropriate population |
| 407 | Supporting mindfulness practices with brain-sensing devices. Cognitive and electrophysiological evidences | Inappropriate study design |
| 408 | A home-based tele-rehabilitation protocol for patients with hip fracture called @ctivehip | Inappropriate population |
| 409 | Conscientiousness is associated with improvement in visuospatial working memory and mood following acute physical exercise: A randomized controlled trial | Inappropriate study design |
| 410 | Acute effect of three different exercise training modalities on executive function in overweight inactive men: A secondary analysis of the BrainFit study | Inappropriate population |
| 411 | Effects of aerobic training, resistance training, or both on brain-derived neurotrophic factor in adolescents with obesity: The hearty randomized controlled trial | Inappropriate study design |
| 412 | Impact of high intensity interval exercise on executive function and brain derived neurotrophic factor in healthy college aged males | Inappropriate population |
| 413 | Experimental effects of acute exercise and music listening on cognitive creativity | Inappropriate study design |
| 414 | Aerobic exercise is more effective than goal-based exercise for the treatment of cognition in Parkinson's disease | Inappropriate study design |
| 415 | The effect of acute cognitively engaging physical activity breaks on children's executive functions: Too much of a good thing? [References]. | Inappropriate study design |
| 416 | The acute effects of high-intensity interval training and moderate-intensity continuous exercise on declarative memory and inhibitory control | Inappropriate study design |
| 417 | Evaluating the effect of Brainfood groups for people with mild cognitive impairment and mild dementia: Preliminary mixed-methodology study | Inappropriate study design |
| 418 | Effects of simultaneous aerobic and cognitive training on executive functions, cardiovascular fitness and functional abilities in older adults with mild cognitive impairment | Inappropriate study design |
| 419 | Effects of physical activity on delayed memory measures in randomized controlled trials with nonclinical older, mild cognitive impairment, and dementia participants | Inappropriate study design |
| 420 | Can functional task exercise improve executive function and contribute to functional balance in older adults with mild cognitive impairment? A pilot study | Inappropriate study design |
| 421 | The Aerobic and Cognitive Exercise Study (ACES) for community-dwelling older adults with or at-risk for mild cognitive impairment (MCI): Neuropsychological, neurobiological and neuroimaging outcomes of a randomized clinical trial | Inappropriate population |

| | | |
|---|---|---|
| 422 | Dance training is superior to repetitive physical exercise in inducing brain plasticity in the elderly | Inappropriate population |
| 423 | Motivation to participate in high-intensity functional exercise compared with a social activity in older people with dementia in nursing homes | Inappropriate population |
| 424 | Exercise, the diurnal cycle of cortisol and cognitive impairment in older adults | Inappropriate study design |
| 425 | Frequency of resistance training does not affect inhibitory control or improve strength in well-trained young adults | Inappropriate study design |
| 426 | Acute effects of continuous and high-intensity interval exercise on executive function | Inappropriate outcome |
| 427 | Rehabilitation for improved cognition in patients with stress-related exhaustion disorder: RECO-A randomized clinical trial | Inappropriate study design |
| 428 | Characterizing beliefs about stroke and walking for exercise among seniors from four racial/ethnic minority communities | Inappropriate study design |
| 429 | Exercise effects on cognitive functioning in young adults with first-episode psychosis: FitForLife | Inappropriate population |
| 430 | A transferable high-intensity intermittent exercise improves executive performance in association with dorsolateral prefrontal activation in young adults | Inappropriate population |
| 431 | Simultaneous aerobic exercise and memory training program in older adults with subjective memory impairments | Inappropriate study design |
| 432 | Aerobic fitness level moderates the association between cannabis use and executive functioning and psychomotor speed following abstinence in adolescents and young adults | Inappropriate population |
| 433 | Leisure-time physical activity and the risk of incident dementia: The Mayo Clinic Study of Aging | Inappropriate intervention |
| 434 | Cortical neural arousal is differentially affected by type of physical exercise performed | Inappropriate study design |
| 435 | Association between aerobic fitness and cerebrovascular function with neurocognitive functions in healthy, young adults | Inappropriate population |
| 436 | Aerobic exercise lengthens telomeres and reduces stress in family caregivers: A randomized controlled trial-Curt Richter Award Paper 2018 | Inappropriate outcome |
| 437 | The effects of acute exercise on cognitive function: Solomon experimental design | Inappropriate study design |
| 438 | Implicit attitudes towards exercise and physical activity behaviour among in-patients with psychiatric disorders | Inappropriate study design |
| 439 | The association between depressive symptoms and cognitive functioning in older Hispanic/Latino adults enrolled in an exercise intervention: Results from the "Caminemos!" study | Inappropriate study design |
| 440 | Changes in cognitive function after a 12-week exercise intervention in adults with Down syndrome | Inappropriate population |
| 441 | Brief aerobic exercise immediately enhances visual attentional control and perceptual speed. Testing the mediating role of feelings of energy | Inappropriate study design |
| 442 | A simplified 10-step Tai-chi programme to enable people with dementia to improve their motor performance: A feasibility study | Inappropriate study design |
| 443 | The effect of acute exercise on encoding and consolidation of long-term memory | Inappropriate study design |
| 444 | "Shall we dance?" Older adults' perspectives on the feasibility of a dance intervention for cognitive function | Inappropriate study design |
| 445 | Balance and walking performance are improved after resistance and aerobic training in persons with chronic stroke | Inappropriate population |
| 446 | Aortic stiffness is associated with increased risk of incident dementia in older adults | Inappropriate study design |
| 447 | Effect of accelerometer-based feedback on physical activity in hospitalized patients with ischemic stroke: A randomized controlled trial | Inappropriate study design |
| 448 | Change in fitness and the relation to change in cognition and neuropsychiatric symptoms after aerobic exercise in patients with mild Alzheimer's disease | Inappropriate population |
| 449 | Compression garments and cerebral blood flow: Influence on cognitive and exercise performance | Inappropriate outcome |
| 450 | Objectively assessed physical activity and associated factors of sedentary behavior among survivors of stroke living in Cape Town, South Africa | Inappropriate population |
| 451 | Effects of a primary care-based multifactorial intervention on physical and cognitive function in frail, elderly individuals: A randomized controlled trial | Inappropriate study design |
| 452 | Experimental study on Taijiquan exercise improving university students' cognitive function | Inappropriate study design |
| 453 | Rating of perceived exertion with Borg scale in stroke over two common activities of the daily living | Inappropriate study design |
| 454 | The feasibility of an acute high-intensity exercise bout to promote locomotor learning after stroke | Inappropriate population |

| | | |
|---|---|---|
| 455 | Cognitive functioning in older people. Results of the first wave of Cognition of Older People, Education, Recreational Activities, Nutrition, Comorbidities, Functional Capacity Studies (COPERNICUS) | Inappropriate population |
| 456 | Effect of high-intensity treadmill exercise on motor symptoms inpatients with DeNovo Parkinson disease: A phase 2 randomized clinical trial | Inappropriate study design |
| 457 | Association of nonalcoholic fatty liver disease with lower brain volume in healthy middle-aged adults in the Framingham study | Inappropriate population |
| 458 | Relationships between aerobic fitness levels and cognitive performance in Swedish office workers | Inappropriate population |
| 459 | Efficacy of intensive multidisciplinary rehabilitation in Parkinson's disease: A randomised controlled study | Inappropriate study design |
| 460 | Improving memory inhibition: A study of retrieval induced forgetting, executive control, and chronic aerobic exercise | Inappropriate study design |
| 461 | Effect of chin tuck against resistance exercise on patients with dysphagia following stroke: A randomized pilot study | Inappropriate study design |
| 462 | A post-exercise facilitation of executive function is independent of aerobically supported metabolic costs | Inappropriate population |
| 463 | Hippocampal volume and vasculature before and after exercise in treatment-resistant schizophrenia | Inappropriate study design |
| 464 | Community-based activity and sedentary patterns are associated with cognitive performance in mobility-limited older adults | Inappropriate study design |
| 465 | Effect of volume in resistance training on inhibitory control in young adults: A randomized and crossover investigation | Inappropriate study design |
| 466 | A single bout of aerobic exercise improves motor skill consolidation in Parkinson's disease | Inappropriate population |
| 467 | Aerobic fitness explains individual differences in the functional brain connectome of healthy young adults | Inappropriate study design |
| 468 | Effects of combined physical and cognitive virtual reality-based training on cognitive impairment and oxidative stress in mci patients: A pilot study | Inappropriate study design |
| 469 | Dementia And Physical Activity (DAPA) trial of moderate to high intensity exercise training for people with dementia: Randomised controlled trial | Inappropriate study design |
| 470 | Association between physical exercise and mental health in 1.2 million individuals in the USA between 2011 and 2015: A cross-sectional study | Inappropriate population |
| 471 | A 16-week aerobic exercise intervention does not affect hippocampal volume and cortical thickness in mild to moderate Alzheimer's disease | Inappropriate study design |
| 472 | A 12-week cycling training regimen improves upper limb functions in people with Parkinson's disease | Inappropriate study design |
| 473 | Self-rated intensity of habitual physical activities is positively associated with dopamine D2/3 receptor availability and cognition | Inappropriate study design |
| 474 | Exercise training on locomotion in patients with Alzheimer's disease: A feasibility study | Inappropriate population |
| 475 | Effects of nurse-led home-based exercise & cognitive behavioral therapy on reducing cancer-related fatigue in patients with ovarian cancer during and after chemotherapy: A randomized controlled trial | Inappropriate study design |
| 476 | Accelerometer-determined physical activity and all-cause mortality in a national prospective cohort study of adults post-acute stroke | Inappropriate study design |
| 477 | Exercise-induced neuroplasticity: Balance training increases cortical thickness in visual and vestibular cortical regions | Inappropriate population |
| 478 | Moving forward in treatment of posttraumatic stress disorder: Innovations to exposure-based therapy | Inappropriate study design |
| 479 | Home-based step training using videogame technology in people with Parkinson's disease: A single-blinded randomised controlled trial | Inappropriate study design |
| 480 | Level of moderate-intensity leisure-time physical activity and reduced mortality in middle-aged and elderly Chinese | Inappropriate study design |
| 481 | Self-management and yoga for older adults with chronic stroke: A mixed-methods study of physical fitness and physical activity | Inappropriate population |
| 482 | Neural basis for reduced executive performance with hypoxic exercise | Inappropriate population |
| 483 | Fitness effects on the cognitive function of older adults: A meta-analytic study-revisited | Inappropriate study design |
| 484 | The association of aging and aerobic fitness with memory | Inappropriate population |
| 485 | The effects of exercise on memory function among young to middle-aged adults: Systematic review and recommendations for future research | Inappropriate study design |
| 486 | Effects of a sedentary intervention on cognitive function | Inappropriate population |
| 487 | Combining therapeutic approaches: rTMS and aerobic exercise in post-stroke depression: A case series | Inappropriate study design |

| | | |
|---|---|---|
| 488 | Brain network modularity predicts exercise-related executive function gains in older adults | Inappropriate study design |
| 489 | Exercise increases mTOR signaling in brain regions involved in cognition and emotional behavior | Inappropriate study design |
| 490 | Mild exercise in female subjects impairs complex learning independent of hydration status and emotion | Inappropriate population |
| 491 | The effects of low-intensity cycling on cognitive performance following sleep deprivation | Inappropriate study design |
| 492 | The positive cognitive impact of aerobic fitness is associated with peripheral inflammatory and brain-derived neurotrophic biomarkers in young adults | Inappropriate population |
| 493 | Using resting-state fMRI to assess the effect of aerobic exercise on functional connectivity of the DLPFC in older overweight adults | Inappropriate study design |
| 494 | The differential effects of prolonged exercise upon executive function and cerebral oxygenation | Inappropriate outcome |
| 495 | Aerobic exercise training ameliorates craving and inhibitory control in methamphetamine dependencies: A randomized controlled trial and event-related potential study | Inappropriate study design |
| 496 | Exercise training and functional connectivity changes in mild cognitive impairment and healthy elders | Inappropriate study design |
| 497 | Exercise to reduce work-related fatigue among employees: A randomized controlled trial | Inappropriate study design |
| 498 | No overt effects of a 6-week exergame training on sensorimotor and cognitive function in older adults. A preliminary investigation | Inappropriate outcome |
| 499 | Mediation of cognitive function improvements by strength gains after resistance training in older adults with mild cognitive impairment: Outcomes of the study of mental and resistance training | Inappropriate population |
| 500 | The moderating role of insulin-like growth factor 1 in the relationship between cognitive and aerobic endurance change: A preliminary study in elderly adults with mild cognitive impairment | Inappropriate population |
| 501 | Dancing or fitness sport? The effects of two training programs on hippocampal plasticity and balance abilities in healthy seniors | Inappropriate study design |
| 502 | Drug abusers have impaired cerebral oxygenation and cognition during exercise | Inappropriate study design |
| 503 | Affect during incremental exercise: The role of inhibitory cognition, autonomic cardiac function, and cerebral oxygenation | Inappropriate study design |
| 504 | The effects of a resistance vs. an aerobic single session on attention and executive functioning in adults | Inappropriate study design |
| 505 | Aerobic exercise for Alzheimer's disease: A randomized controlled pilot trial | Inappropriate intervention |
| 506 | Fitter women did not have attenuated hemodynamic responses to psychological stress compared with age-matched women with lower levels of fitness | Inappropriate study design |
| 507 | One bout of open skill exercise improves cross-modal perception and immediate memory in healthy older adults who habitually exercise | Inappropriate study design |
| 508 | Exercise modality is differentially associated with neurocognition in older adults | Inappropriate study design |
| 509 | Effects of moderate exercise on cortical resilience: A transcranial magnetic stimulation study targeting the dorsolateral prefrontal cortex | Inappropriate study design |
| 510 | Effects of physical activity on the P300 component in elderly people: A systematic review | Inappropriate study design |
| 511 | Resistance training does not alter same-day sleep architecture in institutionalized older adults | Inappropriate intervention |
| 512 | Feldenkrais method and functionality in Parkinson's disease: A randomized controlled clinical trial | Inappropriate study design |
| 513 | A randomised controlled trial of adjunctive yoga and adjunctive physical exercise training for cognitive dysfunction in schizophrenia | Inappropriate study design |
| 514 | Resist forgetting: Effects of aerobic and resistance exercise on prospective and retrospective memory | Inappropriate population |
| 515 | Increasing physical activity through principles of habit formation in new Gym members: A randomized controlled trial | Inappropriate study design |
| 516 | Aerobic exercise intervention, cognitive performance, and brain structure: Results from the physical influences on brain in aging (PHIBRA) study | Inappropriate study design |
| 517 | Factors of nocturnal sleep and daytime nap durations in community-dwelling elderly: A longitudinal population-based study | Inappropriate study design |
| 518 | Correlates of physical activity among depressed older people in six low-income and middle-income countries: A community-based cross-sectional study | Inappropriate population |
| 519 | Effect of aerobic exercise on cancer-associated cognitive impairment: A proof-of-concept RCT | Inappropriate population |

| | | |
|---|---|---|
| 520 | Changes in fMRI activation in anterior hippocampus and motor cortex during memory retrieval after an intense exercise intervention | Inappropriate study design |
| 521 | Acute effects of exercise posture on executive function in transient ischemic attack patients | Inappropriate study design |
| 522 | The effect of acute exercise on blood concentrations of brain-derived neurotrophic factor in healthy adults: A meta-analysis | Inappropriate study design |
| 523 | A group-mediated physical activity intervention in older knee osteoarthritis patients: Effects on social cognitive outcomes | Inappropriate study design |
| 524 | A systematic review of cognitive effects of exercise in depression | Inappropriate study design |
| 525 | Acute moderate exercise improves mnemonic discrimination in young adults | Inappropriate population |
| 526 | Physical exercise for late life depression: Effects on cognition and disability | Inappropriate study design |
| 527 | Effects of physical activity intervention on physical and cognitive function in sedentary adults with and without diabetes | Inappropriate population |
| 528 | Effects of sleep deprivation on cognitive and physical performance in university students | Inappropriate study design |
| 529 | Falls and physical activity in persons with mild to moderate dementia participating in an intensive motor training: Randomized controlled trial | Inappropriate study design |
| 530 | Determination of aerobic capacity via cycle ergometer exercise testing in Alzheimer's disease | Inappropriate study design |
| 531 | Dose-response effects of exercise duration and recovery on cognitive functioning | Inappropriate study design |
| 532 | Beliefs of rehabilitation professionals towards guided self-rehabilitation contracts for post stroke hemiparesis | Inappropriate study design |
| 533 | Control group paradigms in studies investigating acute effects of exercise on cognitive performance-An experiment on expectation-driven placebo effects | Inappropriate study design |
| 534 | Normobaric hypoxia and submaximal exercise effects on running memory and mood state in women | Inappropriate population |
| 535 | Effects of exercise on cognitive function in older people with dementia: A randomized controlled trial | Inappropriate study design |
| 536 | An acute bout of housework activities has beneficial effects on executive function | Inappropriate study design |
| 537 | Sex difference in aerobic exercise efficacy to improve cognition in older adults with vascular cognitive impairment: Secondary analysis of a randomized controlled trial | Inappropriate outcome |
| 538 | Constraining movement reveals motor capability in chronic stroke: An initial study | Inappropriate population |
| 539 | Effect of long-term body-mass-based resistance exercise on cognitive function in elderly people | Inappropriate population |
| 540 | Determining the potential benefits of yoga in chronic stroke care: A systematic review and meta-analysis | Inappropriate outcome |
| 541 | Improving cerebral oxygenation, cognition and autonomic nervous system control of a chronic alcohol abuser through a three-month running program | Inappropriate study design |
| 542 | Exercise training increases parietal lobe cerebral blood flow in chronic stroke: An observational study | Inappropriate outcome |
| 543 | Motor expertise modulates neural oscillations and temporal dynamics of cognitive control | Inappropriate study design |
| 544 | Comparative patient satisfaction and feasibility of a pilot parkinson's disease enrichment program | Inappropriate study design |
| 545 | Aerobic training for improved memory in patients with stress-related exhaustion: A randomized controlled trial | Inappropriate population |
| 546 | Combined dual-task gait training and aerobic exercise to improve cognition, mobility, and vascular health in community-dwelling older adults at risk for future cognitive decline | Inappropriate population |
| 547 | A systematic review of the effect of physical exercise on cognition in stroke and traumatic brain injury patients | Inappropriate study design |
| 548 | Upping the ante: Working harder to address physical inactivity in older adults | Inappropriate study design |
| 549 | Phase II randomised controlled trial of a 6-month self-managed community exercise programme for people with Parkinson's disease | Inappropriate population |
| 550 | Does aerobic exercise influence intrinsic brain activity? An aerobic exercise intervention among healthy old adults | Inappropriate study design |
| 551 | Daily physical activity patterns during the early stage of Alzheimer's disease | Inappropriate study design |
| 552 | A 3-month aerobic training program improves brain energy metabolism in mild Alzheimer's disease: Preliminary results from a neuroimaging study | Inappropriate study design |
| 553 | Pilot study of an exercise intervention for depressive symptoms and associated cognitive-behavioral factors in young adults with major depression | Inappropriate study design |

| | | |
|---|---|---|
| 554 | Physical exercise with music maintains activities of daily living in patients with dementia: Mihama-Kiho project part 2 | Inappropriate study design |
| 555 | Moderate physical activity is associated with cerebral glucose metabolism in adults at risk for Alzheimer's disease | Inappropriate study design |
| 556 | Long-term maintenance of executive- related oculomotor improvements in older adults with self-reported cognitive complaints following a 24-week multiple modality exercise program | Inappropriate population |
| 557 | A 24-week multi-modality exercise program improves executive control in older adults with a self-reported cognitive complaint: Evidence from the antisaccade task | Inappropriate study design |
| 558 | Lifestyle and neurocognition in older adults with cardiovascular risk factors and cognitive impairment | Inappropriate intervention |
| 559 | Resistance band exercises reduce depression and behavioral problems of wheelchair-bound older adults with dementia: A cluster-randomized controlled trial | Inappropriate population |
| 560 | Aerobic fitness, hippocampal viscoelasticity, and relational memory performance | Inappropriate population |
| 561 | Exercise-dependent BDNF as a modulatory factor for the executive processing of individuals in course of cognitive decline. A systematic review | Inappropriate study design |
| 562 | Exercise intensity-dependent effects on cognitive control function during and after acute treadmill running in young healthy adults | Inappropriate study design |
| 563 | Aerobic fitness is associated with cognitive control strategy in preadolescent children | Inappropriate study design |
| 564 | Factors related to meeting physical activity guidelines in active college students: A social cognitive perspective | Inappropriate study design |
| 565 | The effects of a 16-week aerobic exercise programme on cognitive function in people living with HIV | Inappropriate population |
| 566 | Replacing sedentary time with sleep, light, or moderate-to-vigorous physical activity: Effects on self-regulation and executive functioning | Inappropriate study design |
| 567 | A 12-week cycling training regimen improves gait and executive functions concomitantly in people with Parkinson's disease | Inappropriate study design |
| 568 | Left ventricular mechanics in untrained and trained males with tetraplegia | Inappropriate population |
| 569 | Estimated prestroke peak VO2 is related to circulating IGF-1 levels during acute stroke | Inappropriate population |
| 570 | After-effects of pedaling exercise on spinal excitability and spinal reciprocal inhibition in patients with chronic stroke | Inappropriate study design |
| 571 | The effects of aerobic exercise on cognition in schizophrenia: A 3-month follow-up study | Inappropriate study design |
| 572 | Effect of hypoxia on cerebrovascular and cognitive function during moderate intensity exercise | Inappropriate study design |
| 573 | Repeated high-intensity interval exercise shortens the positive effect on executive function during post-exercise recovery in healthy young males | Inappropriate population |
| 574 | Cognitive function at rest and during exercise following breakfast omission | Inappropriate study design |
| 575 | Greater impact of acute high-intensity interval exercise on post-exercise executive function compared to moderate-intensity continuous exercise | Inappropriate study design |
| 576 | Acute effects of varying intensities of treadmill walking exercise on inhibitory control in persons with multiple sclerosis: A pilot investigation | Inappropriate population |
| 577 | The effect of histamine on changes in mental energy and fatigue after a single bout of exercise | Inappropriate population |
| 578 | The relation between aerobic fitness and cognitive performance: Is it mediated by brain potentials? [References]. | Inappropriate study design |
| 579 | Acute aerobic activity enhances response inhibition for less than 30min | Inappropriate study design |
| 580 | An exploration of exercise-induced cognitive enhancement and transfer effects to dietary self-control | Inappropriate population |
| 581 | Premorbid exercise engagement and motor reserve in Parkinson's disease | Inappropriate study design |
| 582 | A randomized, controlled trial of a multi-modal exercise intervention in Huntington's disease | Inappropriate study design |
| 583 | Cortical and motor responses to acute forced exercise in Parkinson's disease | Inappropriate population |
| 584 | Physical and cognitive stimulation using an exergame in subjects with normal aging, mild and moderate cognitive impairment | Inappropriate study design |
| 585 | Aerobic fitness and intraindividual reaction time variability in middle and old age | Inappropriate study design |
| 586 | Acute resistance exercise facilitates attention control in adult males without an age-moderating effect | Inappropriate study design |
| 587 | Design and implementation of an intervention development study: Retaining Cognition while Avoiding Late-Life Depression (ReCALL) | Inappropriate study design |

| | | |
|---|---|---|
| 588 | Exercise plus cognitive performance over and above exercise alone in subjects with mild cognitive impairment | Inappropriate outcome |
| 589 | Combined omega-3 fatty acids, aerobic exercise and cognitive stimulation prevents decline in gray matter volume of the frontal, parietal and cingulate cortex in patients with mild cognitive impairment | Inappropriate study design |
| 590 | Aerobic fitness linked to cortical brain development in adolescent males: Preliminary findings suggest a possible role of BDNF genotype | Inappropriate population |
| 591 | Acute supramaximal exercise increases the brain oxygenation in relation to cognitive workload | Inappropriate study design |
| 592 | Do reported effects of acute aerobic exercise on subsequent higher cognitive performances remain if tested against an instructed self-myofascial release training control group? A randomized controlled trial | Inappropriate study design |
| 593 | Reach the person behind the dementia-Physical therapists' reflections and strategies when composing physical training | Inappropriate population |
| 594 | Effect of aerobic exercise on physical performance in patients with Alzheimer's disease | Inappropriate study design |
| 595 | The relationship between habitual physical activity status and executive function in individuals with Alzheimer's disease: A longitudinal, cross-lagged panel analysis | Inappropriate study design |
| 596 | Pilot study of intensive exercise on endurance, advanced mobility and gait speed in adults with chronic severe acquired brain injury | Inappropriate population |
| 597 | A RCT comparing daily mindfulness meditations, biofeedback exercises, and daily physical exercise on attention control, executive functioning, mindful awareness, self-compassion, and worrying in stressed young adults | Inappropriate population |
| 598 | Acute effects of 3G mobile phone radiations on frontal haemodynamics during a cognitive task in teenagers and possible protective value of Om chanting | Inappropriate study design |
| 599 | Prevalence and correlates of sleep problems among elderly Singaporeans | Inappropriate outcome |
| 600 | High-intensity resistance training in multiple sclerosis-An exploratory study of effects on immune markers in blood and cerebrospinal fluid, and on mood, fatigue, health-related quality of life, muscle strength, walking and cognition | Inappropriate study design |
| 601 | Executive functions improvement following a 5-month aquaerobics program in older adults: Role of cardiac vagal control in inhibition performance | Inappropriate population |
| 602 | Does physical exercise improve ADL capacities in people over 65 years with moderate or severe dementia hospitalized in an acute psychiatric setting? A multisite randomized clinical trial | Inappropriate population |
| 603 | Moderate-to-high intensity physical exercise in patients with Alzheimer's disease: A randomized controlled trial | Inappropriate intervention |
| 604 | Effects of a high-intensity functional exercise program on dependence in activities of daily living and balance in older adults with dementia | Inappropriate outcome |
| 605 | Relationships of peripheral IGF-1, VEGF and BDNF levels to exercise-related changes in memory, hippocampal perfusion and volumes in older adults | Inappropriate study design |
| 606 | Dose-response relationships between exercise intensity, cravings, and inhibitory control in methamphetamine dependence: An ERPs study | Inappropriate study design |
| 607 | A single bout of meditation biases cognitive control but not attentional focusing: Evidence from the global-local task | Inappropriate study design |
| 608 | Acute effects of moderate aerobic exercise on specific aspects of executive function in different age and fitness groups: A meta-analysis | Inappropriate study design |
| 609 | Influences of aerobic dance on cognitive performance in adults with schizophrenia | Inappropriate study design |
| 610 | Social cognitive theory correlates of moderate-intensity exercise among adults with type 2 diabetes | Inappropriate population |
| 611 | Caffeine does not affect improvements in cognition during prolonged high-intensity exercise in alert well-trained individuals | Inappropriate study design |
| 612 | Dose-response and time course effects of acute resistance exercise on executive function | Inappropriate study design |
| 613 | Examining the acute effects of hatha yoga and mindfulness meditation on executive function and mood | Inappropriate study design |
| 614 | A combined cognitive stimulation and physical exercise programme (MINDVital) in early dementia: Differential effects on single- and dual-task gait performance | Inappropriate population |
| 615 | Saving mental effort to maintain physical effort: A shift of activity within the prefrontal cortex in anticipation of prolonged exercise | Inappropriate study design |
| 616 | Feasibility and benefits of computerized cognitive exercise to adults with chronic moderate-to-severe cognitive impairments following an acquired brain injury: A pilot study | Inappropriate outcome |

| | | |
|---|---|---|
| 617 | Aerobic activity in the healthy elderly is associated with larger plasticity in memory related brain structures and lower systemic inflammation | Inappropriate outcome |
| 618 | Aerobic exercise and vascular cognitive impairment: A randomized controlled trial | Inappropriate population |
| 619 | A multimodal cognitive enhancement program for older adults: A case report of the implementation of Brain Training | Inappropriate study design |
| 620 | Sweat it out? The effects of physical exercise on cognition and behavior in children and adults with ADHD: A systematic literature review | Inappropriate population |
| 621 | Effect of a single session of a yogic meditation technique on cognitive performance in medical students: A randomized crossover trial | Inappropriate study design |
| 622 | Hippocampal sub-regional shape and physical activity in older adults | Inappropriate population |
| 623 | Effects of additional, dynamic supported standing practice on functional recovery in patients with sub-acute stroke: A randomized pilot and feasibility trial | Inappropriate population |
| 624 | Effects of a rapid-resisted elliptical training program on motor, cognitive and neurobehavioral functioning in adults with chronic traumatic brain injury | Inappropriate study design |
| 625 | Advances in the treatment of MELAS syndrome: Could cognitive rehabilitation have a role? [References]. | Inappropriate study design |
| 626 | Predictors of physical activity in persons with mental illness: Testing a social cognitive model | Inappropriate population |
| 627 | Aerobic exercise improves mood, cognition, and language function in Parkinson's disease: Results of a controlled study | Inappropriate population |
| 628 | The effects of assisted cycling therapy (ACT) and voluntary cycling on reaction time and measures of executive function in adolescents with Down syndrome | Inappropriate study design |
| 629 | Physical activity in chronic home-living and sub-acute hospitalized stroke patients using objective and self-reported measures | Inappropriate study design |
| 630 | Effect of intermittent normobaric hypoxia on aerobic capacity and cognitive function in older people | Inappropriate study design |
| 631 | Distinct brain and behavioral benefits from cognitive vs. physical training: A randomized trial in aging adults | Inappropriate population |
| 632 | An acute bout of exercise improves the cognitive performance of older adults | Inappropriate study design |
| 633 | Effectiveness of exercise- and cognitive-based treatments on salivary cortisol levels and sundowning syndrome symptoms in patients with Alzheimer's disease | Inappropriate study design |
| 634 | Cardiorespiratory fitness and cognitive functioning following short-term interventions in chronic stroke survivors with cognitive impairment: A pilot study | Inappropriate population |
| 635 | Intensive treadmill training in the acute phase after ischemic stroke | Inappropriate outcome |
| 636 | Affective evaluations of exercising: The role of automatic-reflective evaluation discrepancy | Inappropriate study design |
| 637 | Progressive resistance exercise restores some properties of the triphasic EMG pattern and improves bradykinesia: The PRET-PD randomized clinical trial | Inappropriate study design |
| 638 | A six-month cognitive-motor and aerobic exercise program improves executive function in persons with an objective cognitive impairment: A pilot investigation using the antisaccade task | Inappropriate study design |
| 639 | The effects of acute exercise on memory and brain-derived neurotrophic factor (BDNF) | Inappropriate study design |
| 640 | Effects of repetitive facilitative exercise with neuromuscular electrical stimulation, vibratory stimulation and repetitive transcranial magnetic stimulation of the hemiplegic hand in chronic stroke patients | Inappropriate population |
| 641 | Outcomes of repetitive facilitation exercises in convalescent patients after stroke with impaired health status | Inappropriate population |
| 642 | The effects of exercise frequency on executive function in individuals with Parkinson's disease | Inappropriate study design |
| 643 | Hatha yoga and executive function: A systematic review | Inappropriate population |
| 644 | Systematically developed pilot randomized controlled trial of exercise and cognition in persons with multiple sclerosis | Inappropriate study design |
| 645 | What influences acceptability and engagement with a high intensity exercise programme for people with stroke? A qualitative descriptive study | Inappropriate study design |
| 646 | Effects of exercise on Irisin, BDNF and IL-6 serum levels in patients with progressive multiple sclerosis | Inappropriate outcome |
| 647 | Evaluation of a physical activity intervention for adults with brain impairment: A controlled clinical trial | Inappropriate population |
| 648 | Perceptions of exercise for older people living with dementia in Bangkok, Thailand: An exploratory qualitative study | Inappropriate population |
| 649 | Yoga improves academic performance in urban high school students compared to physical education: A randomized controlled trial | Inappropriate study design |

| | | |
|---|---|---|
| 650 | Short-duration and intensive training improves long-term reaching performance in individuals with chronic stroke | Inappropriate study design |
| 651 | Neurophysiological and behavioral correlates of cognitive control during low and moderate intensity exercise | Inappropriate outcome |
| 652 | Hydrotherapy vs. Conventional land-based exercise for improving walking and balance after stroke: A randomized controlled trial | Inappropriate population |
| 653 | Variable intensive early walking poststroke (VIEWS): A randomized controlled trial | Inappropriate population |
| 654 | Accelerometer measured sedentary behavior and physical activity in White and Black adults: The REGARDS study | Inappropriate study design |
| 655 | Dose-response relationship between intensity of exercise and cognitive performance in individuals with Down syndrome: A preliminary study | Inappropriate study design |
| 656 | Entorhinal volume, aerobic fitness, and recognition memory in healthy young adults: A voxel-based morphometry study | Inappropriate population |
| 657 | Exercise mode moderates the relationship between mobility and basal ganglia volume in healthy older adults | Inappropriate study design |
| 658 | Group-based exercise combined with dual-task training improves gait but not vascular health in active older adults without dementia | Inappropriate population |
| 659 | Blood lactate levels as a biomarker of antipsychotic side effects in patients with schizophrenia | Inappropriate study design |
| 660 | Promoting policy and environmental change in faith-based organizations: Outcome evaluation of a mini-grants program | Inappropriate study design |
| 661 | Physical and psychomotor performance of Australian football and rugby league officials during a match simulation | Inappropriate population |
| 662 | Effects of a high-intensity functional exercise program on depressive symptoms among people with dementia in residential care: A randomized controlled trial | Inappropriate study design |
| 663 | Cost-effectiveness of exercise as a therapy for behavioural and psychological symptoms of dementia within the EVIDEM-E randomised controlled trial | Inappropriate population |
| 664 | Statistical learning is not affected by a prior bout of physical exercise | Inappropriate study design |
| 665 | Subjective memory impairment and well-being in community-dwelling older adults | Inappropriate population |
| 666 | High velocity circuit resistance training improves cognition, psychiatric symptoms and neuromuscular performance in overweight outpatients with severe mental illness | Inappropriate population |
| 667 | Objective measures of physical activity, white matter integrity and cognitive status in adults over age 80 | Inappropriate study design |
| 668 | The COMT val158met polymorphism in ultra-endurance athletes | Inappropriate population |
| 669 | Low intensity exercise does not impact cognitive function during exposure to normobaric hypoxia | Inappropriate study design |
| 670 | Does moderate hypoxia alter working memory and executive function during prolonged exercise? [References]. | Inappropriate study design |
| 671 | Exercise as potential countermeasure for the effects of 70 days of bed rest on cognitive and sensorimotor performance | Inappropriate population |
| 672 | Enhancing both motor and cognitive functioning in Parkinson's disease: Aerobic exercise as a rehabilitative intervention | Inappropriate study design |
| 673 | Capturing the post-exertional exacerbation of fatigue following physical and cognitive challenge in patients with chronic fatigue syndrome | Inappropriate population |
| 674 | Improved cardiorespiratory fitness is associated with increased cortical thickness in mild cognitive impairment | Inappropriate study design |
| 675 | Effects of endurance training combined with cognitive remediation on everyday functioning, symptoms, and cognition in multiepisode schizophrenia patients | Inappropriate population |
| 676 | Independent and combined effects of cognitive and physical activity on incident MCI | Inappropriate population |
| 677 | Moderate-intensity physical activity, Hippocampal volume, and memory in older adults with mild cognitive impairment | Inappropriate population |
| 678 | The effects of a brief, water-based exercise intervention on cognitive function in older adults | Inappropriate population |
| 679 | Hearts and minds: Linking vascular rigidity and aerobic fitness with cognitive aging | Inappropriate intervention |
| 680 | Higher cardiorespiratory fitness levels are associated with greater hippocampal volume in breast cancer survivors | Inappropriate study design |
| 681 | Antecedent acute cycling exercise affects attention control: An ERP study using attention network test | Inappropriate intervention |
| 682 | The Agewell trial: A pilot randomised controlled trial of a behaviour change intervention to promote healthy ageing and reduce risk of dementia in later life | Inappropriate population |

| | | |
|---|---|---|
| 683 | A single bout of moderate aerobic exercise improves motor skill acquisition | Inappropriate study design |
| 684 | Association between lifetime physical activity and cognitive functioning in middle-aged and older community dwelling adults: Results from the Brain in Motion Study | Inappropriate study design |
| 685 | Effect of structured physical activity on sleep-wake behaviors in sedentary elderly adults with mobility limitations | Inappropriate population |
| 686 | Effect of a 24-month physical activity intervention vs health education on cognitive outcomes in sedentary older adults: The LIFE randomized trial | Inappropriate population |
| 687 | Effects of a physical fitness program on memory and blood viscosity in sedentary elderly men | Inappropriate outcome |
| 688 | Impact of 6-month aerobic exercise on Alzheimer's symptoms | Inappropriate population |
| 689 | Pushing to the limits: The dynamics of cognitive control during exhausting exercise | Inappropriate population |
| 690 | Exercise practices in individuals at clinical high risk of developing psychosis | Inappropriate study design |
| 691 | A pilot test of the additive benefits of physical exercise to CBT for OCD | Inappropriate population |
| 692 | Long-term effects of resistance exercise training on cognition and brain volume in older women: Results from a randomized controlled trial | Inappropriate population |
| 693 | Acute exercise improves prefrontal cortex but not hippocampal function in healthy adults | Inappropriate study design |
| 694 | Age moderates the association of Aerobic exercise with initial learning of an online task requiring cognitive control | Inappropriate study design |
| 695 | Attitudes toward physical activity and exercise: Comparison of memory clinic patients and their caregivers and prediction of activity levels | Inappropriate population |
| 696 | Apolipoprotein E epsilon4 allele modulates the immediate impact of acute exercise on prefrontal function | Inappropriate population |
| 697 | An ecological approach to cognitive enhancement: Complex motor training | Inappropriate population |
| 698 | Intensive cycle ergometer training improves gait speed and endurance in patients with Parkinson's disease: A comparison with treadmill training | Inappropriate population |
| 699 | The effects of long-term resistance exercise on the relationship between neurocognitive performance and GH, IGF-1, and homocysteine levels in the elderly | Inappropriate study design |
| 700 | Physical exercise as a diagnostic, rehabilitation, and preventive tool: Influence on neuroplasticity and motor recovery after stroke | Inappropriate study design |
| 701 | Nervous facilitation in cardiodynamic response of exercising athletes to superimposed mental tasks: Implications in depressive disorder | Inappropriate study design |
| 702 | Impacts of dance on non-motor symptoms, participation, and quality of life in Parkinson disease and healthy older adults | Inappropriate population |
| 703 | Exercise in patients with subacute stroke: A randomized, controlled pilot study of home-based exercise in subacute stroke | Inappropriate study design |
| 704 | The effect of upper-extremity aerobic exercise on complex regional pain syndrome type I: A randomized controlled study on subacute stroke | Inappropriate study design |
| 705 | Post-exercise hypotensive responses following an acute bout of aquatic and overground treadmill walking in people post-stroke: A pilot study | Inappropriate study design |
| 706 | The effect of pelvic floor muscle training on sexual function in men with lower urinary tract symptoms after stroke | Inappropriate study design |
| 707 | Improving arm function in chronic stroke: a pilot study of sensory amplitude electrical stimulation via glove electrode during task-specific training | Inappropriate population |
| 708 | Exercise and fitness modulate cognitive function in older adults | Inappropriate study design |
| 709 | A 9-week aerobic and strength training program improves cognitive and motor function in patients with dementia: A randomized, controlled trial | Inappropriate population |
| 710 | Physical activity affects brain integrity in HIV+ individuals | Inappropriate population |
| 711 | Understanding how exercise promotes cognitive integrity in the aging brain | Inappropriate study design |
| 712 | Do cognitive measures and brain circuitry predict outcomes of exercise in Parkinson Disease: A randomized clinical trial | Inappropriate study design |
| 713 | Phase II pragmatic randomized controlled trial of patient-led therapies (mirror therapy and lower-limb exercises) during inpatient stroke rehabilitation | Inappropriate study design |
| 714 | Development and validation of the cognitive behavioral physical activity questionnaire | Inappropriate study design |
| 715 | Lag-lead based assessment and adaptation of exercise speed for stroke survivors | Inappropriate study design |
| 716 | Effect of two physical exercise protocols on cognition and depressive symptoms in oldest-old people: A randomized controlled trial | Inappropriate population |
| 717 | Forced aerobic exercise enhances motor recovery after stroke: A case report | Inappropriate population |

| | | |
|---|---|---|
| 718 | Strategy training shows promise for addressing disability in the first 6 months after stroke | Inappropriate study design |
| 719 | Effects of community exercise therapy on metabolic, brain, physical, and cognitive function following stroke: A randomized controlled pilot trial | Inappropriate study design |
| 720 | Ergometer cycling enhances executive control in task switching | Inappropriate population |
| 721 | The impact of aerobic exercise on brain-derived neurotrophic factor and neurocognition in individuals with schizophrenia: A single-blind, randomized clinical trial | Inappropriate population |
| 722 | Study protocol of "Worth the Walk": A randomized controlled trial of a stroke risk reduction walking intervention among racial/ethnic minority older adults with hypertension in community senior centers | Inappropriate population |
| 723 | Long-term mild-intensity exercise regimen preserves prefrontal cortical volume against aging | Inappropriate outcome |
| 724 | The acute effects of yoga on cognitive measures for women with premenstrual syndrome | Inappropriate population |
| 725 | Cardiovascular response to exercise training in the systemic right ventricle of adults with transposition of the great arteries | Inappropriate population |
| 726 | Aerobic fitness is associated with inhibitory control in persons with multiple sclerosis | Inappropriate study design |
| 727 | Exercise intensity modulates the change in cerebral blood flow following aerobic exercise in chronic stroke | Inappropriate population |
| 728 | Physical activity in adolescents with an orthopaedic limitation: A review of the literature | Inappropriate study design |
| 729 | Design of the Park-in-Shape study: A phase II double blind randomized controlled trial evaluating the effects of exercise on motor and non-motor symptoms in Parkinson's disease | Inappropriate study design |
| 730 | Low-intensity daily walking activity is associated with hippocampal volume in older adults | Inappropriate study design |
| 731 | Single bouts of exercise selectively sustain attentional processes | Inappropriate population |
| 732 | Motivators for uptake and maintenance of exercise: Perceptions of long-term stroke survivors and implications for design of exercise programmes | Inappropriate study design |
| 733 | Improvement in balance using a virtual reality-based stepping exercise: A randomized controlled trial involving individuals with chronic stroke | Inappropriate population |
| 734 | Acute effects of walking, cycling, and yoga exercise on cognition in persons with relapsing-remitting multiple sclerosis without impaired cognitive processing speed | Inappropriate population |
| 735 | A combination of supplements may reduce the risk of Alzheimer's disease in elderly Japanese with normal cognition | Inappropriate study design |
| 736 | Unilateral forced nostril breathing and aphasia-Exploring unilateral forced nostril breathing as an adjunct to aphasia treatment: A case series | Inappropriate intervention |
| 737 | Exercise testing and training in people with Huntington's disease | Inappropriate population |
| 738 | Combined effects of sleep deprivation and strenuous exercise on cognitive performances during the North Face Ultra Trail du Mont Blanc (UTMB) | Inappropriate population |
| 739 | Association between physical fitness and cognitive function in multiple sclerosis: Does disability status matter? [References]. | Inappropriate population |
| 740 | Testing a modified perceived exertion scale for Alzheimer's disease | Inappropriate study design |
| 741 | Neuroelectric adaptations to cognitive processing in virtual environments: An exercise-related approach | Inappropriate population |
| 742 | Isometric exercise and cognitive function: An investigation of acute dose-response effects during submaximal fatiguing contractions | Inappropriate study design |
| 743 | The effect of arm support combined with rehabilitation games on upper-extremity function in subacute stroke: A randomized controlled trial | Inappropriate study design |
| 744 | Two-year exercise program improves physical function in Parkinson's disease: The PRET-PD randomized clinical trial | Inappropriate study design |
| 745 | Improved gait adjustments after gait adaptability training are associated with reduced attentional demands in persons with stroke | Inappropriate population |
| 746 | Spin exercise improves semantic fluency in previously sedentary older adults | Inappropriate study design |
| 747 | A combination of physical and cognitive exercise improves reaction time in persons 61-84 years old | Inappropriate study design |
| 748 | A bilateral rehabilitation system for the lower limbs | Inappropriate study design |
| 749 | Components of late-life exercise and cognitive function: An 8-year longitudinal study | Inappropriate study design |
| 750 | Evidence cerebral blood-flow regulation mediates exercise-cognition links in healthy young adults | Inappropriate population |

| | | |
|---|---|---|
| 751 | Amplitude-oriented exercise in Parkinson's disease: A randomized study comparing LSVT-BIG and a short training protocol | Inappropriate study design |
| 752 | The impact of high intensity physical training on motor and non-motor symptoms in patients with Parkinson's disease (PIP): A preliminary study | Inappropriate study design |
| 753 | Interaction between serum BDNF and aerobic fitness predicts recognition memory in healthy young adults | Inappropriate study design |
| 754 | Does the inverted-u function disappear in expert athletes? An analysis of the attentional behavior under physical exercise of athletes and non-athletes | Inappropriate intervention |
| 755 | The Q223R polymorphism in the leptin receptor associates with objectively measured light physical activity in free-living Japanese | Inappropriate study design |
| 756 | Agentic extraversion moderates the effect of physical exercise on executive shifting performance | Inappropriate study design |
| 757 | Treadmill training as an augmentation treatment for Alzheimer's disease: A pilot randomized controlled study | Inappropriate population |
| 758 | Social cognitive correlates of physical activity among single mothers with young children | Inappropriate population |
| 759 | Acute exercise and aerobic fitness influence selective attention during visual search | Inappropriate study design |
| 760 | The diagnosis and management of mild cognitive impairment: A clinical review | Inappropriate study design |
| 761 | Optimizing function and physical activity among nursing home residents with dementia: Testing the impact of function-focused care | Inappropriate study design |
| 762 | Adapting mindfulness meditation for the older adult | Inappropriate study design |
| 763 | Older adults with amnestic mild cognitive impairment exhibit exacerbated gait slowing under dual-task challenges | Inappropriate outcome |
| 764 | Executive function and self-regulated exergaming adherence among older adults | Inappropriate intervention |
| 765 | BDNF mediates improvements in executive function following a 1-year exercise intervention | Inappropriate study design |
| 766 | Improvements to executive function during exercise training predict maintenance of physical activity over the following year | Inappropriate population |
| 767 | The effects of acute aerobic activity on cognition and cross-domain transfer to eating behavior | Inappropriate study design |
| 768 | Feasibility of a combined aerobic and strength training program and its effects on cognitive and physical function in institutionalized dementia patients. A pilot study | Inappropriate population |
| 769 | Positive effect of acute mild exercise on executive function via arousal-related prefrontal activations: An fNIRS study | Inappropriate study design |
| 770 | Effect of exercise and cognitive activity on self-reported sleep quality in community-dwelling older adults with cognitive complaints: A randomized controlled trial | Inappropriate study design |
| 771 | The relation of aerobic fitness to cognitive control and heart rate variability: A neurovisceral integration study | Inappropriate population |
| 772 | Energy expenditure and exercise intensity of interactive video gaming in individuals poststroke | Inappropriate population |
| 773 | Effects of caffeine and menthol on cognition and mood during simulated firefighting in the heat | Inappropriate population |
| 774 | The effects of an 8-week Hatha yoga intervention on executive function in older adults | Inappropriate study design |
| 775 | Effects of aerobic exercise on cognitive performance and individual psychopathology in depressive and schizophrenia patients | Inappropriate outcome |
| 776 | Aerobic fitness predicts relational memory but not item memory performance in healthy young adults | Inappropriate study design |
| 777 | Aerobic exercise influences quality of life of children and youngsters with congenital heart disease: A randomized controlled trial | Inappropriate study design |
| 778 | Rivastigmine transdermal patch and physical exercises for Alzheimer's disease: A randomized clinical trial | Inappropriate population |
| 779 | Impact of adapted physical activity and therapeutic patient education on functioning and quality of life in patients with postacute strokes | Inappropriate outcome |
| 780 | Moderate-to-high intensity aerobic exercise in patients with mild to moderate Alzheimer's disease: A pilot study | Inappropriate study design |
| 781 | An intensive exercise program improves motor performances in patients with dementia: Translational model of geriatric rehabilitation | Inappropriate study design |
| 782 | Improved cerebral oxygenation response and executive performance as a function of cardiorespiratory fitness in older women: A fNIRS study | Inappropriate outcome |
| 783 | Executive function and endocrinological responses to acute resistance exercise | Inappropriate study design |
| 784 | Tai Chi Chuan optimizes the functional organization of the intrinsic human brain architecture in older adults | Inappropriate study design |

| | | |
|---|---|---|
| 785 | Physical activity reduces hippocampal atrophy in elders at genetic risk for Alzheimer's disease | Inappropriate study design |
| 786 | Physical activity predicts microstructural integrity in memory-related networks in very old adults | Inappropriate study design |
| 787 | Cardiorespiratory fitness and accelerated cognitive decline with aging | Inappropriate population |
| 788 | Phase I/II randomized trial of aerobic exercise in Parkinson disease in a community setting | Inappropriate population |
| 789 | Physical activity and cognition in the Northern Manhattan Study | Inappropriate study design |
| 790 | A randomised controlled trial of an exercise plus behaviour change intervention in people with multiple sclerosis: The step it up study protocol | Inappropriate study design |
| 791 | The relation of childhood physical activity to brain health, cognition, and scholastic achievement: Abstract. | Inappropriate population |
| 792 | Physical activity and cognitive function in bariatric surgery candidates | Inappropriate study design |
| 793 | Protocol to evaluate the impact of yoga supplementation on cognitive function in schizophrenia: A randomised controlled trial | Inappropriate study design |
| 794 | Acute exercise increases oxygenated and deoxygenated hemoglobin in the prefrontal cortex | Inappropriate study design |
| 795 | Community-based exercise for chronic disease management: An Italian design for the United States? [References]. | Inappropriate population |
| 796 | Mean diffusivity as a potential diffusion tensor biomarker of motor rehabilitation after electrical stimulation incorporating task specific exercise in stroke: A pilot study | Inappropriate study design |
| 797 | Non-pharmacological strategies to delay cognitive decline | Inappropriate study design |
| 798 | Aerobic fitness and body mass index in individuals with schizophrenia: Implications for neurocognition and daily functioning | Inappropriate study design |
| 799 | Effects of cognitive training with and without aerobic exercise on cognitively demanding everyday activities | Inappropriate population |
| 800 | Physiotherapy and physical functioning post-stroke: Exercise habits and functioning 4 years later? Long-term follow-up after a 1-year long-term intervention period: A randomized controlled trial | Inappropriate population |
| 801 | A comprehensive exercise program for a young adult male with Down syndrome who experienced a stroke | Inappropriate study design |
| 802 | Clinical feasibility of the Nintendo WiiTM for balance training post-stroke: A phase II randomized controlled trial in an inpatient setting | Inappropriate study design |
| 803 | Body weight-supported treadmill training vs. overground walking training for persons with chronic stroke: A pilot randomized controlled trial | Inappropriate population |
| 804 | Partnering to increase access to community exercise programs for people with stroke, acquired brain injury, and multiple sclerosis | Inappropriate study design |
| 805 | Cognitive function during low-intensity walking: A test of the treadmill workstation | Inappropriate population |
| 806 | Effects of Tai Chi on balance and fall prevention in Parkinson's disease: A randomized controlled trial | Inappropriate study design |
| 807 | Exercise for depressive symptoms in stroke patients: A systematic review and meta-analysis | Inappropriate population |
| 808 | An enhanced protocol for constraint-induced aphasia therapy II: A case series | Inappropriate study design |
| 809 | Randomized controlled trial of yoga for chronic poststroke hemiparesis: Motor function, mental health, and quality of life outcomes | Inappropriate intervention |
| 810 | Cardiovascular resistance to orthostatic load in athletes after aerobic exercise | Inappropriate study design |
| 811 | Safety and feasibility of post-stroke care and exercise after minor ischemic stroke or transient ischemic attack: MotiveS & MoveIT | Inappropriate population |
| 812 | High-intensity physical activity, stable relationship, and high education level associate with decreasing risk of erectile dysfunction in 1,000 apparently healthy cardiovascular risk subjects | Inappropriate study design |
| 813 | Associations of daily walking and television viewing time with liver cancer mortality: Findings from the Japan Collaborative Cohort Study | Inappropriate population |
| 814 | The effect of exercise on behavioural and psychological symptoms of dementia: The EVIDEM-E randomised controlled clinical trial | Inappropriate study design |
| 815 | Interventions to reduce the number of falls among older adults with/without cognitive impairment: An exploratory meta-analysis | Inappropriate study design |
| 816 | Motor imagery group practice for gait rehabilitation in individuals with post-stroke hemiparesis: A pilot study | Inappropriate population |
| 817 | Acute bouts of assisted cycling improves cognitive and upper extremity movement functions in adolescents with Down syndrome | Inappropriate study design |

| | | |
|---|---|---|
| 818 | Feasibility and outcomes of a community-based, pedometer-monitored walking program in chronic stroke: A pilot study | Inappropriate study design |
| 819 | Does participation in standardized aerobic fitness training during inpatient stroke rehabilitation promote engagement in aerobic exercise after discharge? A cohort study | Inappropriate study design |
| 820 | Core stabilization exercise with real-time feedback for chronic hemiparetic stroke: A pilot randomized controlled trials | Inappropriate population |
| 821 | Community-based exercise programs as a strategy to optimize function in chronic disease: A systematic review | Inappropriate population |
| 822 | Cross-sectional comparison of executive attention function in normally aging long-term t'ai chi, meditation, and aerobic fitness practitioners versus sedentary adults | Inappropriate population |
| 823 | Family-mediated exercises (FAME): An exploration of participant's involvement in a novel form of exercise delivery after stroke | Inappropriate study design |
| 824 | Aerobic exercises enhance cognitive functions and brain derived neurotrophic factor in ischemic stroke patients | Inappropriate study design |
| 825 | Executive function and coping in stroke survivors | Inappropriate intervention |
| 826 | Leg muscle power is enhanced by training in people with Parkinson's disease: A randomized controlled trial | Inappropriate population |
| 827 | Executive function processes predict mobility outcomes in older adults | Inappropriate population |
| 828 | Impact of acute aerobic exercise and cardiorespiratory fitness on visuospatial attention performance and serum BDNF levels | Inappropriate study design |
| 829 | Acute aerobic exercise increases brain-derived neurotrophic factor levels in elderly with Alzheimer's disease | Inappropriate study design |
| 830 | Repetitive facilitative exercise under continuous electrical stimulation for severe arm impairment after sub-acute stroke: A randomized controlled pilot study | Inappropriate outcome |
| 831 | Maximal heart rate does not limit cardiovascular capacity in healthy humans: Insight from right atrial pacing during maximal exercise | Inappropriate study design |
| 832 | Intensive rehabilitation increases BDNF serum levels in Parkinsonian patients: A randomized study | Inappropriate study design |
| 833 | Cardiorespiratory responses during the 6-minute walk and ramp cycle ergometer tests and their relationship to physical activity in stroke | Inappropriate outcome |
| 834 | Randomized clinical trial of 3 types of physical exercise for patients with Parkinson disease | Inappropriate population |
| 835 | Reduced metaboreflex control of blood pressure during exercise in individuals with intellectual disability: A possible contributor to exercise intolerance | Inappropriate population |
| 836 | Decline in executive control during acute bouts of exercise as a function of exercise intensity and fitness level | Inappropriate population |
| 837 | Quantifying daily physical activity and determinants in sedentary patients with Parkinson's disease | Inappropriate study design |
| 838 | The immediate and sustained effects of acute exercise on planning aspect of executive function | Inappropriate population |
| 839 | Spiritual activity is associated with better cognitive function in old age | Inappropriate intervention |
| 840 | Aerobic fitness and cognitive function in midlife: An association mediated by plasma insulin | Inappropriate population |
| 841 | Semantic memory functional MRI and cognitive function after exercise intervention in mild cognitive impairment | Inappropriate outcome |
| 842 | An economic evaluation of resistance training and aerobic training versus balance and toning exercises in older adults with mild cognitive impairment | Inappropriate population |
| 843 | The effects of an aerobic and resistance exercise training program on cognition following stroke | Inappropriate study design |
| 844 | Exercise holds immediate benefits for affect and cognition in younger and older adults | Inappropriate study design |
| 845 | Preserving cognition, quality of life, physical health and functional ability in Alzheimer's disease: The effect of physical exercise (ADEX trial): Rationale and design | Inappropriate population |
| 846 | Developing physical activity interventions for adults with spinal cord injury. Part 2: Motivational counseling and peer-mediated interventions for people intending to be active | Inappropriate population |
| 847 | Development and initial testing of functional task exercise on older adults with cognitive impairment at risk of Alzheimer's disease-FcTSim programme-A feasibility study | Inappropriate study design |
| 848 | Benefits of regular aerobic exercise for executive functioning in healthy populations | Inappropriate population |

| | | |
|---|---|---|
| 849 | The effects of high-intensity intermittent exercise training on cardiovascular response to mental and physical challenge | Inappropriate population |
| 850 | Affecting cognition and quality of life via aerobic exercise in Alzheimer's disease | Inappropriate study design |
| 851 | The health benefits and challenges of exercise training in persons living with schizophrenia: A pilot study | Inappropriate study design |
| 852 | White matter hyperintensities, exercise, and improvement in gait speed: Does type of gait rehabilitation matter? [References]. | Inappropriate population |
| 853 | Shorter term aerobic exercise improves brain, cognition, and cardiovascular fitness in aging | Inappropriate study design |
| 854 | Effects of non-paretic arm exercises using a tubing band on abdominal muscle activity in stroke patients | Inappropriate study design |
| 855 | The role of Tai Chi, cultural dancing, playing a musical instrument and singing in the prevention of chronic disease in Chinese older adults: A mind-body meditative approach | Inappropriate study design |
| 856 | Perceptions of a videogame-based dance exercise program among individuals with Parkinson's disease | Inappropriate study design |
| 857 | How interventions might improve cognition in healthy older adults | Inappropriate study design |
| 858 | The dose-response relationship between resistance exercise intensity and cognitive performance: Does heart rate mediate this effect? [References]. | Inappropriate study design |
| 859 | Effects of simultaneously performed cognitive and physical training in older adults | Inappropriate outcome |
| 860 | Biomarkers and perfusion-training-induced changes after stroke (BAPTISe): Protocol of an observational study accompanying a randomized controlled trial | Inappropriate study design |
| 861 | Validity of rating of perceived exertion ranges in individuals in the subacute stage of stroke recovery | Inappropriate population |
| 862 | The influence of aerobic fitness on cerebral white matter integrity and cognitive function in older adults: Results of a one-year exercise intervention | Inappropriate population |
| 863 | Characteristics of exercise training interventions to improve cardiorespiratory fitness after stroke: A systematic review with meta-analysis | Inappropriate study design |
| 864 | Video game play (Dance Dance Revolution) as a potential exercise therapy in Huntington's disease: A controlled clinical trial | Inappropriate intervention |
| 865 | The cortical control of cycling exercise in stroke patients: An fNIRS study | Inappropriate study design |
| 866 | The effects of closed and open kinetic chain exercises on lower limb muscle activity and balance in stroke survivors | Inappropriate study design |
| 867 | Executive function performance is reduced during occupational burnout but can recover to the level of healthy controls | Inappropriate outcome |
| 868 | Executive function during acute exercise: The role of exercise intensity | Inappropriate study design |
| 869 | Perceptions of participating in high-intensity functional exercise among older people dependent in activities of daily living (ADL) | Inappropriate study design |
| 870 | Exercise treatment for postconcussion syndrome: A pilot study of changes in functional magnetic resonance imaging activation, physiology, and symptoms | Inappropriate study design |
| 871 | A randomized trial of two home-based exercise programmes to improve functional walking post-stroke | Inappropriate study design |
| 872 | Perceived recovery as a predictor of physical activity participation after mild stroke | Inappropriate study design |
| 873 | Low-dose propranolol and exercise capacity in postural tachycardia syndrome: A randomized study | Inappropriate population |
| 874 | Self-management intervention for chronic pain in older adults: A randomised controlled trial | Inappropriate study design |
| 875 | Effects of task-specific and impairment-based training compared with usual care on functional walking ability after inpatient stroke rehabilitation: LEAPS trial | Inappropriate study design |
| 876 | Eccentric versus concentric resistance training to enhance neuromuscular activation and walking speed following stroke | Inappropriate study design |
| 877 | Effects of aerobic cycling training on cardiovascular fitness and heart rate recovery in patients with chronic stroke | Inappropriate study design |
| 878 | Structural and functional brain changes related to different types of physical activity across the life span | Inappropriate study design |
| 879 | Effects of current physical activity on affective response to exercise: Physical and social-cognitive mechanisms | Inappropriate study design |
| 880 | Community-based group exercise for persons with Parkinson disease: A randomized controlled trial | Inappropriate study design |
| 881 | Aerobic fitness and the brain: Increased N-acetyl-aspartate and choline concentrations in endurance-trained middle-aged adults | Inappropriate study design |
| 882 | Multimodal exercise intervention improves frontal cognitive functions and gait in Alzheimer's disease: A controlled trial | Inappropriate population |

| | | |
|---|---|---|
| 883 | Correlation between moderate daily physical activity and neurocognitive variability in healthy elderly people | Inappropriate population |
| 884 | Exercise-induced noradrenergic activation enhances memory consolidation in both normal aging and patients with amnestic mild cognitive impairment | Inappropriate study design |
| 885 | Effects of forced use combined with scheduled home exercise program on upper extremity functioning in individuals with hemiparesis | Inappropriate population |
| 886 | High-intensity physical activity modulates diet effects on cerebrospinal amyloid-beta levels in normal aging and mild cognitive impairment | Inappropriate population |
| 887 | Study protocol: EXERcise and Cognition In Sedentary adults with Early-ONset dementia (EXERCISE-ON) | Inappropriate population |
| 888 | The association between aerobic fitness and executive function is mediated by prefrontal cortex volume | Inappropriate study design |
| 889 | Tai chi and postural stability in patients with Parkinson's disease | Inappropriate study design |
| 890 | Acute physical activity on cognitive function: A heart rate variability examination | Inappropriate population |
| 891 | The effects of yoga on physical functioning and health related quality of life in older adults: A systematic review and meta-analysis | Inappropriate study design |
| 892 | Feasibility, criterion validity and retest reliability of exercise testing using the Astrand-Rhyming test protocol with an adaptive ergometer in stroke patients | Inappropriate study design |
| 893 | Improvement in physiological and psychological parameters after 6 months of yoga practice | Inappropriate study design |
| 894 | Effects of weight training on cognitive functions in elderly with Alzheimer's disease | Inappropriate population |
| 895 | Neuroelectric indices of goal maintenance following a single bout of physical activity | Inappropriate population |
| 896 | Design and rationale for a randomized controlled trial testing the efficacy of aerobic exercise for patients with obsessive-compulsive disorder | Inappropriate study design |
| 897 | The relationship between physical activity and cognition in older Latinos | Inappropriate study design |
| 898 | Effects of acute physical exercise on executive functions: A comparison between aerobic and strength exercise | Inappropriate study design |
| 899 | Balance recovery through virtual stepping exercises using Kinect skeleton tracking: A follow-up study with chronic stroke patients | Inappropriate study design |
| 900 | Shifting the focus from quantitative to qualitative exercise characteristics in exercise and cognition research | Inappropriate population |
| 901 | Fitness and cognitive processing speed in persons with multiple sclerosis: A cross-sectional investigation | Inappropriate study design |
| 902 | Physical activity and neuropsychiatric symptoms of Parkinson disease | Inappropriate study design |
| 903 | Afferent stimulation provided by glove electrode during task-specific arm exercise following stroke | Inappropriate population |
| 904 | A single bout of exercise improves motor memory | Inappropriate study design |
| 905 | Treatment of hemispatial neglect in patients with posthemiparesis: A single-subject experimental design study using a whole-body tilt exercise plus mental practice | Inappropriate study design |
| 906 | Is there a bidirectional relationship between depressive symptoms and cognitive ability in older people? A prospective study using the English Longitudinal Study of Ageing | Inappropriate study design |
| 907 | Effects of fast functional exercise on muscle activity after stroke | Inappropriate population |
| 908 | Feasibility and perception of the impact from aerobic exercise in older adults with Alzheimer's disease | Inappropriate population |
| 909 | Home-based treadmill training for individuals with Parkinson's disease: A randomized controlled pilot trial | Inappropriate study design |
| 910 | Differential effects of power training versus functional task practice on compensation and restoration of arm function after stroke | Inappropriate study design |
| 911 | Correlations between walking exercise and Each of boned density, muscle volume, fluctuation of the center of gravity, and dementia in middle-aged and elderly women | Inappropriate population |
| 912 | Hemodynamic mechanisms of the attenuated blood pressure response to mental stress after a single bout of maximal dynamic exercise in healthy subjects | Inappropriate population |
| 913 | Changes in brain volume and cognition in a randomized trial of exercise and social interaction in a community-based sample of non-demented Chinese elders | Inappropriate study design |
| 914 | Resistance training and functional plasticity of the aging brain: A 12-month randomized controlled trial | Inappropriate study design |
| 915 | The influence of exercise-induced fatigue on cognitive function | Inappropriate study design |
| 916 | Declining physical capacity but maintained aerobic activity in early Alzheimer's disease | Inappropriate study design |

| | | |
|---|---|---|
| 917 | Neuropsychological and neurophysiological effects of strengthening exercise for early dementia: A pilot study | Inappropriate population |
| 918 | A four-week walking exercise programme in patients with knee osteoarthritis improves the ability of dual-task performance: A randomized controlled trial | Inappropriate population |
| 919 | Cognitive and neural correlates of aerobic fitness in obese older adults | Inappropriate outcome |
| 920 | The moderating role of exercise on stress-related effects on the hippocampus and memory in later adulthood | Inappropriate study design |
| 921 | Rehabilitation after stroke in older people | Inappropriate population |
| 922 | Chronic stroke survivors benefit from high-intensity aerobic treadmill exercise: A randomized control trial | Inappropriate population |
| 923 | The effect of energy drinks on cortisol levels, cognition and mood during a fire-fighting exercise | Inappropriate study design |
| 924 | Cardiovascular fitness modulates brain activation associated with spatial learning | Inappropriate population |
| 925 | Differential cognitive effects of cycling versus stretching/coordination training in middle-aged adults | Inappropriate population |
| 926 | Autism and exergaming: Effects on repetitive behaviors and cognition | Inappropriate population |
| 927 | Structured exercise does not stabilize cognitive function in individuals with mild cognitive impairment residing in a structured living facility | Inappropriate intervention |
| 928 | Aerobic fitness and multidomain cognitive function in advanced age | Inappropriate population |
| 929 | Comparison of physio ball and plinth trunk exercises regimens on trunk control and functional balance in patients with acute stroke: A pilot randomized controlled trial | Inappropriate study design |
| 930 | Effect of a yoga programme on an individual with Parkinson's disease: A single-subject design | Inappropriate study design |
| 931 | Maintaining physical fitness and function in Alzheimer's disease: A pilot study | Inappropriate study design |
| 932 | Classification of late-life leisure activities among elderly Chinese in Hong Kong | Inappropriate study design |
| 933 | The effects of single bouts of aerobic exercise, exergaming, and videogame play on cognitive control | Inappropriate study design |
| 934 | Exercise training increases size of hippocampus and improves memory | Inappropriate study design |
| 935 | Musculoskeletal pain is associated with incident mobility disability in community-dwelling elders | Inappropriate study design |
| 936 | Effects of acute exercise on executive function: A study with a Tower of London Task | Inappropriate study design |
| 937 | Assessing cognitive impairment following stroke | Inappropriate population |
| 938 | Predictors of physical activity in adults with type 2 diabetes | Inappropriate study design |
| 939 | Functioning of the attentional networks at rest vs. during acute bouts of aerobic exercise | Inappropriate study design |
| 940 | Does acute exercise switch off switch costs? A study with younger and older athletes | Inappropriate population |
| 941 | Thinking about maintaining exercise therapy: Does being positive or negative make a difference? [References]. | Inappropriate study design |
| 942 | A moderate intensity exercise program did not increase the oxidative stress in older adults | Inappropriate population |
| 943 | On the contribution of group III and IV muscle afferents to the circulatory response to rhythmic exercise in humans | Inappropriate outcome |
| 944 | Effect of a high-intensity functional exercise program on functional balance: Preplanned subgroup analyses of a randomized controlled trial in residential care facilities | Inappropriate study design |
| 945 | Influence of regular exercise on subjective sense of burden and physical symptoms in community-dwelling caregivers of dementia patients: A randomized controlled trial | Inappropriate population |
| 946 | Association between improvements in insulin resistance and changes in cognitive function in elderly diabetic patients with normal cognitive function | Inappropriate study design |
| 947 | Adherence to a home-based exercise program for individuals after stroke | Inappropriate study design |
| 948 | Current approaches to restoring walking in patients during the acute phase of cerebral stroke | Inappropriate study design |
| 949 | The effectiveness of an exercise programme for elders with dementia in a Taiwanese day-care centre | Inappropriate study design |
| 950 | Ankle training with a robotic device improves hemiparetic gait after a stroke | Inappropriate intervention |
| 951 | The interplay between cognitive and motor functioning in healthy older adults: Findings from dual-task studies and suggestions for intervention | Inappropriate outcome |

| | | |
|---|---|---|
| 952 | Bilateral and unilateral arm training improve motor function through differing neuroplastic mechanisms: A single-blinded randomized controlled trial | Inappropriate study design |
| 953 | Feasibility, safety and efficacy of an early aerobic rehabilitation program for patients after minor ischemic stroke: A pilot randomized controlled trial | Inappropriate study design |
| 954 | Two emerging concepts for elite athletes: The short-term effects of testosterone and cortisol on the neuromuscular system and the dose-response training role of these endogenous hormones | Inappropriate study design |
| 955 | The effects on cognitive functions of a movement-based intervention in patients with Alzheimer's type dementia: A pilot study | Inappropriate population |
| 956 | Acute and chronic exercise effects on attentional control in older road cyclists | Inappropriate outcome |
| 957 | Exercise and Parkinson's: Benefits for cognition and quality of life | Inappropriate study design |
| 958 | Exercise performance in adolescents with autonomic dysfunction | Inappropriate study design |
| 959 | Physical activity and memory functions: An interventional study | Inappropriate population |
| 960 | Effects of a controlled trial of aerobic exercise for mild cognitive impairment: A controlled trial | Inappropriate study design |
| 961 | Chronic endurance exercise training prevents aging-related cognitive decline in healthy older adults: A randomized controlled trial | Inappropriate intervention |
| 962 | The psychometric qualities of four observational pain tools (OPTs) for the assessment of pain in elderly people with osteoarthritic pain | Inappropriate study design |
| 963 | Dissociation between the facilitative effect of acute exercise on feeling states and on cognitive flexibility | Inappropriate population |
| 964 | Acute moderate exercise elicits increased dorsolateral prefrontal activation and improves cognitive performance with Stroop test | Inappropriate study design |
| 965 | Functional connectivity: A source of variance in the association between cardiorespiratory fitness and cognition? [References]. | Inappropriate population |
| 966 | Cognitive function during acute exercise: A test of the transient hypofrontality theory | Inappropriate study design |
| 967 | Plasticity of brain networks in a randomized intervention trial of exercise training in older adults | Inappropriate population |
| 968 | Neuronal and cognitive plasticity: A neurocognitive framework for ameliorating cognitive aging | Inappropriate study design |
| 969 | Intensity of recreational physical activity throughout life and later life cognitive | Inappropriate population |
| 970 | Feasibility of a 6-month exercise and recreation program to improve executive functioning and memory in individuals with chronic stroke | Inappropriate outcome |
| 971 | Hippocampal plasticity in response to exercise in schizophrenia | Inappropriate study design |
| 972 | Physical activity and functional limitations in older adults: A systematic review related to Canada's Physical Activity Guidelines | Inappropriate study design |
| 973 | Aerobic exercise and neurocognitive performance: A meta-analytic review of randomized controlled trials | Inappropriate study design |
| 974 | Program of rehabilitative exercise and education to avert vascular events after non-disabling stroke or transient ischemic attack (PREVENT Trial): A multi-centred, randomised controlled trial | Inappropriate study design |
| 975 | Aerobic exercise improves cognition for older adults with glucose intolerance, a risk factor for Alzheimer's disease | Inappropriate study design |
| 976 | Physical activity preferences and type 2 diabetes: Exploring demographic, cognitive, and behavioral differences | Inappropriate study design |
| 977 | Cognitive health benefits of strengthening exercise for community-dwelling older adults | Inappropriate population |
| 978 | Effects of cardiorespiratory fitness and cerebral blood flow on cognitive outcomes in older women | Inappropriate study design |
| 979 | Impact of aerobic exercise training on cognitive functions and affect associated to the COMT polymorphism in young adults | Inappropriate study design |
| 980 | The effect of a clinically practical exercise on levodopa bioavailability and motor response in patients with Parkinson disease | Inappropriate study design |
| 981 | Tai Chi exercise versus rehabilitation for the elderly with cerebral vascular disorder: A single-blinded randomized controlled trial | Inappropriate outcome |
| 982 | Feasibility and effects of adapted cardiac rehabilitation after stroke: A prospective trial | Inappropriate study design |
| 983 | Correlating heart rate and perceived exertion during aerobic exercise in Alzheimer's disease | Inappropriate intervention |
| 984 | Family participation can improve unilateral spatial neglect in patients with acute right hemispheric stroke | Inappropriate outcome |
| 985 | The relation of aerobic fitness to neuroelectric indices of cognitive and motor task preparation | Inappropriate study design |

| | | |
|---|---|---|
| 986 | Can exercise improve language and cognition in Parkinson's disease? A case report | Inappropriate population |
| 987 | Effects of a high-intensity functional exercise programme on depressive symptoms and psychological well-being among older people living in residential care facilities: A cluster-randomized controlled trial | Inappropriate study design |
| 988 | Predictors of response to treadmill exercise in stroke survivors | Inappropriate study design |
| 989 | Dual-task performances can be improved in patients with dementia: A randomized controlled trial | Inappropriate study design |
| 990 | The effects of treadmill exercise training on hip bone density and tibial bone geometry in stroke survivors: A pilot study | Inappropriate study design |
| 991 | Affect and self-efficacy responses during moderate-intensity exercise among low-active women: The effect of cognitive appraisal | Inappropriate population |
| 992 | Effects of exercise intensity and self-efficacy on state anxiety with breast cancer survivors | Inappropriate population |
| 993 | Seated bilateral leg exercise effects on hemiparetic lower extremity function in chronic stroke | Inappropriate study design |
| 994 | A community-based exercise and education scheme for stroke survivors: A randomized controlled trial and economic evaluation | Inappropriate population |
| 995 | The immediate and delayed effects of an acute bout of exercise on cognitive performance of healthy older adults | Inappropriate population |
| 996 | Exercise adoption among older, low-income women at risk for cardiovascular disease | Inappropriate study design |
| 997 | Aerobic fitness is associated with gray matter volume and white matter integrity in multiple sclerosis | Inappropriate population |
| 998 | Hydration effects on cognitive performance during military tasks in temperate and cold environments | Inappropriate outcome |
| 999 | A test of the catecholamines hypothesis for an acute exercise-cognition interaction | Inappropriate study design |
| 1000 | Progressive adaptive physical activity in stroke improves balance, gait, and fitness: Preliminary results | Inappropriate intervention |
| 1001 | The time course effect of moderate intensity exercise on response execution and response inhibition | Inappropriate study design |
| 1002 | Specific effects of acute moderate exercise on cognitive control | Inappropriate study design |
| 1003 | Benefits of physical exercise on executive functions in older people with Parkinson's Disease | Inappropriate study design |
| 1004 | Exercise programs improve mobility and balance in people with Parkinson's disease | Inappropriate study design |
| 1005 | High intensity eccentric resistance training decreases bradykinesia and improves quality of life in persons with Parkinson's disease: A preliminary study | Inappropriate population |
| 1006 | Factors associated with exercise counseling and program preferences among breast cancer survivors | Inappropriate population |
| 1007 | Effects of an acute bout of localized resistance exercise on cognitive performance in middle-aged adults: A randomized controlled trial study | Inappropriate study design |
| 1008 | Fitness's moderation of the facilitative effect of acute exercise on cognitive flexibility in older women | Inappropriate study design |
| 1009 | Prestroke physical activity and early functional status after stroke | Inappropriate study design |
| 1010 | Aerobic exercise enhances cognitive flexibility | Inappropriate population |
| 1011 | Exercise and health-related quality of life during the first year following acute stroke. A randomized controlled trial | Inappropriate study design |
| 1012 | Stable implicit motor processes despite aerobic locomotor fatigue | Inappropriate study design |
| 1013 | Exercise, fitness, and neurocognitive function in older adults: The "selective improvement" and "cardiovascular fitness" hypotheses | Inappropriate study design |
| 1014 | The effects of an exercise program on several abilities associated with driving performance in older adults | Inappropriate study design |
| 1015 | Acute aerobic exercise and information processing: Modulation of executive control in a random number generation task | Inappropriate study design |
| 1016 | Acute incremental exercise, performance of a central executive task, and sympathoadrenal system and hypothalamic-pituitary-adrenal axis activity | Inappropriate study design |
| 1017 | Effect of exercise on negative affect in residents in special care units with moderate to severe dementia | Inappropriate population |
| 1018 | The effects of a Simplified Tai-Chi Exercise Program (STEP) on the physical health of older adults living in long-term care facilities: A single group design with multiple time points | Inappropriate study design |

| | | |
|---|---|---|
| 1019 | An examination of potential mechanisms for exercise as a treatment for depression: A pilot study | Inappropriate population |
| 1020 | Community-based adaptive physical activity program for chronic stroke: Feasibility, safety, and efficacy of the Empoli model | Inappropriate study design |
| 1021 | Changes in cognitive function in a randomized trial of physical activity: Results of the lifestyle interventions and independence for elders pilot study | Inappropriate study design |
| 1022 | Regenerate: Assessing the feasibility of a strength-training program to enhance the physical and mental health of chronic post stroke patients with depression | Inappropriate outcome |
| 1023 | Influence of age on syncope following prolonged exercise: Differential responses but similar orthostatic intolerance | Inappropriate study design |
| 1024 | Protein and carbohydrate supplementation increases aerobic and thermoregulatory capacities | Inappropriate population |
| 1025 | Bed rest and cognition: Effects on executive functioning and reaction time | Inappropriate population |
| 1026 | Does physical activity intensity moderate social cognition and behavior relationships? [References]. | Inappropriate outcome |
| 1027 | Physical activity and risk of ischemic stroke in the Northern Manhattan Study | Inappropriate study design |
| 1028 | Delays in grip initiation and termination in persons with stroke: Effects of arm support and active muscle stretch exercise | Inappropriate study design |
| 1029 | Aerobic exercise improves cognition and motor function poststroke | Inappropriate study design |
| 1030 | Aerobic fitness is associated with hippocampal volume in elderly humans | Inappropriate study design |
| 1031 | The effect of a high-intensity functional exercise program on activities of daily living: A randomized controlled trial in residential care facilities | Inappropriate population |
| 1032 | Effect of acute exercise on cognitive control required during an Eriksen flanker task | Inappropriate study design |
| 1033 | Forced, not voluntary, exercise improves motor function in Parkinson's disease patients | Inappropriate study design |
| 1034 | A home programme for patients with cognitive dysfunction: A pilot study | Inappropriate population |
| 1035 | Short-form Tai Chi improves standing balance of people with chronic stroke | Inappropriate population |
| 1036 | A randomized controlled trial of gravity-supported, computer-enhanced arm exercise for individuals with severe hemiparesis | Inappropriate population |
| 1037 | Acute exercise performed close to the anaerobic threshold improves cognitive performance in elderly females | Inappropriate study design |
| 1038 | Effects of an aerobic exercise program on aerobic capacity, spatiotemporal gait parameters, and functional capacity in subacute stroke | Inappropriate outcome |
| 1039 | Aerobic endurance exercise benefits memory and affect in young adults | Inappropriate study design |
| 1040 | Social-cognitive theories for predicting physical activity behaviours of employed women with and without young children | Inappropriate study design |
| 1041 | Age, physical fitness, and attention: P3a and P3b | Inappropriate outcome |
| 1042 | Modality of physical exercise and cognitive function in Hong Kong older Chinese community | Inappropriate study design |
| 1043 | Haemodynamic responses to exercise, ATP infusion and thigh compression in humans: Insight into the role of muscle mechanisms on cardiovascular function | Inappropriate study design |
| 1044 | Impact of endurance exercise on levodopa-associated cortisol release and force increase in patients with Parkinson's disease | Inappropriate study design |
| 1045 | The exercise redox paradigm in the Down's syndrome: Improvements in motor function and increases in blood oxidative status in young adults | Inappropriate population |
| 1046 | Short-term exercise to exhaustion and its effects on cognitive function in young women | Inappropriate population |
| 1047 | A qualitative analysis of a progressive resistance exercise programme for people with Parkinson's disease | Inappropriate population |
| 1048 | Motivators for treadmill exercise after stroke | Inappropriate outcome |
| 1049 | Rehabilitation of balance after stroke with multisensorial training: A single-blind randomized controlled study | Inappropriate population |
| 1050 | Inflated perceptions of physical activity after stroke: Pairing self-report with physiologic measures | Inappropriate study design |
| 1051 | Physical exercise and psychological well-being among people with chronic illness and disability: A grounded approach | Inappropriate study design |
| 1052 | Movement in mind: The relationship of exercise with cognitive status for older adults in the Swedish National Study on Aging and Care (SNAC) | Inappropriate population |
| 1053 | Effects of acute exercise on executive processing, short-term and long-term memory | Inappropriate population |
| 1054 | Effects of nicotinamide adenine dinucleotide hydride on physical and mental performance | Inappropriate study design |

| | | |
|---|---|---|
| 1055 | A real-time assessment of the effect of exercise in chronic fatigue syndrome | Inappropriate intervention |
| 1056 | Creatine supplementation, sleep deprivation, cortisol, melatonin and behavior | Inappropriate study design |
| 1057 | The effectiveness of a leg-kicking training program on performance and physiological measures of competitive swimmers | Inappropriate study design |
| 1058 | Affective responses of inactive women to a maximal incremental exercise test: A test of the dual-mode model | Inappropriate population |
| 1059 | Neuromuscular function in survivors of the acute respiratory distress syndrome | Inappropriate population |
| 1060 | The interactive effect of exercise intensity and task difficulty on human cognitive processing | Inappropriate study design |
| 1061 | Arousal gradient hypothesis | Inappropriate study design |
| 1062 | Cardiac and vasomotor components of the carotid baroreflex control of arterial blood pressure during isometric exercise in humans | Inappropriate study design |
| 1063 | Physical Activity Participation by Presence and Type of Functional Deficits in Older Women: The Women's Health and Aging Studies | Inappropriate study design |
| 1064 | Aerobic Exercise Training Increases Brain Volume in Aging Humans | Inappropriate study design |
| 1065 | Exercise and working memory: An individual differences investigation | Inappropriate population |
| 1066 | Group physical activity for demented older adults--Feasibility and effectiveness | Inappropriate study design |
| 1067 | The influence of autonomic impairment on aerobic exercise outcome in stroke patients | Inappropriate study design |
| 1068 | Association of education and race/ethnicity with physical activity in insured urban women | Inappropriate study design |
| 1069 | Stroke: A randomized trial of exercise or relaxation | Inappropriate study design |
| 1070 | Effects of Stroke Severity and Training Duration on Locomotor Recovery After Stroke: A Pilot Study | Inappropriate study design |
| 1071 | Change in motor function and risk of mortality in older persons | Inappropriate population |
| 1072 | Interactive effects of fitness and hormone treatment on brain health in postmenopausal women | Inappropriate population |
| 1073 | Arg16Gly polymorphism of the beta2-adrenergic receptor is associated with differences in cardiovascular function at rest and during exercise in humans | Inappropriate population |
| 1074 | Body mass index in older persons is associated with Alzheimer disease pathology | Inappropriate study design |
| 1075 | State-specific prevalence of obesity among adults--United States, 2005. | Inappropriate population |
| 1076 | Perceived Exertion: Influence of Age and Cognitive Development | Inappropriate study design |
| 1077 | Psychological and Physical Activity Training for Older Persons: Who Does Not Attend? [References]. | Inappropriate outcome |
| 1078 | Prospective and Daily Measures of Acute Thoughts, Decisional Struggle, and Coping: Measurement Correspondence and the Prediction of Exercise in Young Adults | Inappropriate population |
| 1079 | Yearlong Physical Activity and Depressive Symptoms in Older Japanese Adults: Cross-Sectional Data from the Nakanojo Study | Inappropriate population |
| 1080 | The effect of visual feedback plus mental practice on symmetrical weight-bearing training in people with hemiparesis | Inappropriate population |
| 1081 | Age, aerobic fitness, executive function, and episodic memory | Inappropriate outcome |
| 1082 | Therapeutic Exercise and Depressive Symptoms After Stroke | Inappropriate population |
| 1083 | Student-led exercise sessions yield significant fitness gains for Alzheimer's patients | Inappropriate study design |
| 1084 | Effects of moderate-intensity exercise on physiological, behavioral, and emotional responses to family caregiving: A randomized controlled trial | Inappropriate study design |
| 1085 | Heart Rate Variability (HRV) parameters correlate with motor impairment and aerobic capacity in stroke patients | Inappropriate study design |
| 1086 | Individually Tailored Treatment Targeting Activity, Motor Behavior, and Cognition Reduces Pain-Related Disability: A Randomized Controlled Trial in Patients With Musculoskeletal Pain | Inappropriate population |
| 1087 | Effects of acute bouts of exercise on cognition | Inappropriate population |
| 1088 | Exercise level and cognitive decline: The MoVIES project | Inappropriate population |
| 1089 | Acute cardiovascular exercise and executive control function | Inappropriate study design |
| 1090 | Aerobic Endurance Exercise Improves Executive Functions in Depressed Patients | Inappropriate population |
| 1091 | The effect of weightlifting upon left ventricular function and markers of cardiomyocyte damage | Inappropriate study design |

| | | |
|---|---|---|
| 1092 | Effect of Intermittent Exercise on Multiple-choice Reaction Times of Soccer Players | Inappropriate population |
| 1093 | Cardiopulmonary and metabolic responses to maximum exercise and aerobic capacity in hemiplegic patients | Inappropriate study design |
| 1094 | Physical activity in relation to cognitive decline in elderly men: The FINE Study | Inappropriate study design |
| 1095 | Walking and Dementia in Physically Capable Elderly Men | Inappropriate population |
| 1096 | Physical Fitness Training: Outcomes for Adult Oncology Patients | Inappropriate study design |
| 1097 | Outcomes of an Exercise Program for Older Women Recruited Through Primary Care | Inappropriate population |
| 1098 | Mood alterations in mindful versus aerobic exercise modes | Inappropriate study design |
| 1099 | Cognitive and Psychological Outcomes of Exercise in a 1-Year Follow-Up Study of Patients With Chronic Obstructive Pulmonary Disease | Inappropriate population |
| 1100 | The effect of early aerobic training on independence six months post stroke | Inappropriate study design |
| 1101 | The effect of a perceptual cognitive task on exercise performance: The dual-task condition after brain injury | Inappropriate study design |
| 1102 | Does weight-shifting exercise improve postural symmetry in sitting in people with hemiplegia? [References]. | Inappropriate study design |
| 1103 | Cortical reorganization induced by task-oriented training in chronic hemiplegic stroke patients | Inappropriate population |
| 1104 | Cognitive behavioral therapy and aerobic exercise for Gulf war veterans' illnesses: A randomized controlled trial | Inappropriate population |
| 1105 | Acute thoughts, exercise consistency and coping self-efficacy | Inappropriate population |
| 1106 | Aerobic exercise intervention improves aerobic capacity and movement initiation in Parkinson's disease patients | Inappropriate population |
| 1107 | Systematic muscle building exercises in the rehabilitation of stroke patients | Inappropriate population |
| 1108 | A preliminary study on the reliability of physical performance measures in older day-care center clients with dementia | Inappropriate population |
| 1109 | Gait performance in dementia: The effects of a 6-week resistance training program in an adult day-care setting | Inappropriate study design |
| 1110 | Sympathetic nervous system and behavioral responses to stress following exercise training. | Inappropriate outcome |
| 1111 | An exploration of factors predicting the effects of aerobic conditioning on mood state. | Inappropriate study design |
| 1112 | Physical and mental training: Implications for cognitive functioning in old age | Inappropriate population |
| 1113 | Exercise cognition among elderly women. | Inappropriate population |
| 1114 | The feasibility of conducting strength and flexibility programs for elderly nursing home residents with dementia. | Inappropriate study design |
| 1115 | Effects of aerobic exercise on cognitive and psychosocial functioning in patients with mild hypertension | Inappropriate population |
| 1116 | Fluid intelligence in an older COPD sample after short- or long-term exercise | Inappropriate intervention |
| 1117 | Chronic fatigue in the elderly. | Inappropriate study design |
| 1118 | Chronic psychological effects of exercise and exercise plus cognitive strategies. | Inappropriate population |
| 1119 | Interactive effects of Type A personality and psychological and physical stressors on human cardiovascular functions. | Inappropriate population |
| 1120 | Aerobic capacity and cognitive performance in a cross-sectional aging study. | Inappropriate study design |
| 1121 | Is the glass half-full or half-empty? The relationship of big and little optimism with acute and longer term exercise-related social cognitions | Inappropriate intervention |
| 1122 | The mirror does not lie: Acute exercise and self-efficacy | Inappropriate study design |
| 1123 | Reliability and validity of questions about exercise in the Canadian Study of Health and Aging | Inappropriate study design |
| 1124 | Daughters and mothers exercising together (DAMET): A 12-week pilot project designed to improve physical self-perception and increase recreational physical activity | Inappropriate study design |
| 1125 | A randomized trial of exercise programs among older individuals living in two long-term care facilities: The FallsFREE Program | Inappropriate study design |
| 1126 | Cognitive compromise following exercise in monozygotic twins discordant for chronic fatigue syndrome: Fact or artifact? [References]. | Inappropriate study design |
| 1127 | Cardiovascular responses of women with chronic fatigue syndrome to stressful cognitive testing before and after strenuous exercise | Inappropriate population |
| 1128 | Disability in activities of daily living among the elderly. | Inappropriate outcome |

| | | |
|---|---|---|
| 1129 | Characteristics of exercise behavior among college students: Application of social cognitive theory to predicting stage of change. | Inappropriate population |
| 1130 | The benefits of exercise for the health of older people. | Inappropriate study design |
| 1131 | The development of health enhancing exercise groups adapted for hemiplegic patients: A pilot study | Inappropriate population |
| 1132 | Elder rehab: A student-supervised exercise program for Alzheimer's patients | Inappropriate population |
| 1133 | Ageing, fitness and neurocognitive function. | Inappropriate study design |
| 1134 | Increasing correct participation in an exercise class for adult day care clients | Inappropriate study design |
| 1135 | Decongestant effects on hemodynamics at rest, exercise, and recovery from exercise during -6" of head down tilt. | Inappropriate study design |
| 1136 | Effects of exercise on cognitive and motor function in chronic fatigue syndrome and depression. | Inappropriate population |
| 1137 | Psychological and cognitive outcomes of a randomized trial of exercise among patients with chronic obstructive pulmonary disease | Inappropriate study design |
| 1138 | Physical activity in chronic fatigue syndrome: Assessment and its role in fatigue. | Inappropriate population |
| 1139 | Aerobic exercise and early Parkinson's disease. | Inappropriate population |
| 1140 | Effect of aerobic training on the cognitive performance of elderly patients with senile dementia of Alzheimer type. | Inappropriate study design |
| 1141 | Hot temperatures, hostile affect, hostile cognition, and arousal: Tests of a general model of affective aggression. | Inappropriate population |
| 1142 | Exercise and the psychological state of institutionalized elderly: A review. | Inappropriate study design |
| 1143 | The effects of attendance and effort on outcomes among older adults in a long-term exercise program. | Inappropriate study design |
| 1144 | Effect of physical exercise on the performance of cognitive tasks. | Inappropriate study design |
| 1145 | The impact of long-term exercise training on psychological function in older adults. | Inappropriate study design |
| 1146 | Cardiovascular reactivity in Type A and B males to mental arithmetic and aerobic exercise at an equivalent oxygen uptake. | Inappropriate study design |
| 1147 | Long-term effects of exercise on psychological functioning in older men and women. | Inappropriate outcome |
| 1148 | Impact of exercise training on plasma adrenocorticotropin response to a well-learned vigilance task. | Inappropriate population |
| 1149 | Effects of moderate and low intensity long-term exercise by older adults. | Inappropriate study design |
| 1150 | Psychological and cognitive effects of an exercise program for community-residing older adults. | Inappropriate study design |
| 1151 | Age and fitness effects on EEG, ERPs, visual sensitivity, and cognition. | Inappropriate population |
| 1152 | Improving aerobic capacity in healthy older adults does not necessarily lead to improved cognitive performance | Inappropriate study design |
| 1153 | Effects of consistent aerobic exercise on the psychological functioning of women. | Inappropriate study design |
| 1154 | The relationship between aerobic activity and cognitive performance under stress. | Inappropriate study design |
| 1155 | Relationships between catecholamine or 3-methoxy 4-hydroxy phenylglycol changes and the mental performance under submaximal exercise in man. | Inappropriate study design |
| 1156 | Specific efficacy expectations mediate exercise compliance in patients with COPD | Inappropriate study design |
| 1157 | Psychophysiological response patterns to single, double, and triple stressors. | Inappropriate study design |
| 1158 | Effects of prolonged sleep deprivation, with and without chronic physical exercise, on mood and performance. | Inappropriate population |
| 1159 | Relieving the anxiety and fear in dementia. | Inappropriate population |
| 1160 | Effects of aerobic exercise upon affect and cognition. | Inappropriate study design |
| 1161 | Aerobic exercise training and improved neuropsychological function of older individuals. | Inappropriate population |
| 1162 | Effects of acute exercise and cigarette smoking on autonomic and neuromuscular responses to a cognitive stressor. | Inappropriate study design |
| 1163 | The WeReha Project for an Innovative Home-Based Exercise Training in Chronic Stroke Patients: A Clinical Study. | Inappropriate study design |
| 1164 | Feasibility of a 12 week physical intervention to prevent cognitive decline and disability in the at-risk elderly population in Korea. | Inappropriate study design |

| | | |
|---|---|---|
| 1165 | Therapist behaviours in a web-based mindfulness-based cognitive therapy (eMBCT) for chronic cancer-related fatigue - Analyses of e-mail correspondence. | Inappropriate study design |
| 1166 | Measurement site of inferior vena cava diameter affects the accuracy with which fluid responsiveness can be predicted in spontaneously breathing patients: a post hoc analysis of two prospective cohorts. | Inappropriate population |
| 1167 | Assessment of Fatigue Using Wearable Sensors: A Pilot Study. | Inappropriate study design |
| 1168 | Implementation and effects of information technology-based and print-based interventions to promote physical activity among community-dwelling older adults: Protocol for a randomized crossover trial. | Inappropriate population |
| 1169 | Development of cognitive and physical exercise systems, clinical recordings, large-scale data analytics, and virtual coaching for heart failure patients: Protocol for the biotechcoach-forall project. | Inappropriate study design |
| 1170 | A comparison of dry needling and kinesiotaping therapies in myofascial pain syndrome: A randomized clinical study. | Inappropriate population |
| 1171 | Importance of maximal strength and muscle-tendon mechanics for improving force steadiness in persons with parkinson's disease. | Inappropriate study design |
| 1172 | Patient and nurse perceptions of an innovative TV wellness channel in an acute medical ward: A feasibility study. | Inappropriate population |
| 1173 | Robotic Rehabilitation: An Opportunity to Improve Cognitive Functions in Subjects With Stroke. An Explorative Study. | Inappropriate population |
| 1174 | Cognitive and Psychosocial Outcomes of Self-Guided Executive Function Training and Low-Intensity Aerobic Exercise in Healthy Older Adults. | Inappropriate population |
| 1175 | Dose response of a novel exogenous ketone supplement on physiological, perceptual and performance parameters. | Inappropriate outcome |
| 1176 | Mild Physical Activity Does Not Improve Spatial Learning in a Virtual Environment. | Inappropriate study design |
| 1177 | Short- and long-term effects of a physical exercise intervention on work ability and work strain in symptomatic menopausal women. | Inappropriate population |
| 1178 | A Case report on impact of physiotherapy rehabilitation on partial claw hand secondary to borderline tuberculoid Hansen disease. | Inappropriate population |
| 1179 | Therapeutic effectiveness of a single exercise session combined with WalkAide functional electrical stimulation in post-stroke patients: A crossover design study. | Inappropriate population |
| 1180 | Development of a framework for pneumatic device selection for lymphedema treatment. | Inappropriate study design |
| 1181 | Comparative study of evolution and survival of patients with intermittent claudication, with or without limitation for exercises, followed in a specific outpatient setting. | Inappropriate study design |
| 1182 | Effectiveness of transcutaneous electrical stimulation of vagus nerve among post stroke urinary incontinence. | Inappropriate study design |
| 1183 | Advancing health-enhancing physical activity at workplace: Sport4Heath 2020 scientific forum. | Inappropriate study design |
| 1184 | The Effect of Virtual Reality Applications on Balance and Gait Speed in Individuals with Alzheimer Dementia: A Pilot Study. | Inappropriate outcome |
| 1185 | PasoDoble, a Proposed Dance/Music for People With Parkinson's Disease and Their Caregivers. | Inappropriate population |
| 1186 | Effect of aerobic training on selected cardiopulmonary parameters in myocardial infarction patients with stroke. | Inappropriate outcome |
| 1187 | Effects of individual progressive single- and dual-task training on gait and cognition among older healthy adults: a randomized-controlled comparison study. | Inappropriate study design |
| 1188 | Embodiment in virtual reality for the treatment of chronic low back pain: A case series. | Inappropriate outcome |
| 1189 | Determining the feasibility of a trial to evaluate the effectiveness of phototherapy versus placebo at reducing pain during physical activity for people with knee osteoarthritis: a pilot randomized controlled trial. | Inappropriate study design |
| 1190 | Aerobic and resistance exercise improves Reynolds risk score in overweight or obese breast cancer survivors. | Inappropriate study design |
| 1191 | Ecg dispersion mapping in preclinical diagnosis of cardiovascular diseases. | Inappropriate study design |
| 1192 | Effect of deep cervical flexor training on respiratory functions in chronic neck pain patients with forward head posture. | Inappropriate study design |
| 1193 | Superior effects of modified chen-style Tai Chi versus 24-style Tai Chi on cognitive function, fitness, and balance performance in adults over 55. | Inappropriate population |
| 1194 | Contextual interference in children with brain lesions: protocol of a pilot study investigating blocked vs. random practice order of an upper limb robotic exergame. | Inappropriate study design |

| | | |
|---|---|---|
| 1195 | Chronic asthma and the risk of cardiovascular disease. | Inappropriate population |
| 1196 | Study on the needs of health care for establishment of regional health care plan-focused on Cheonan city. | Inappropriate population |
| 1197 | Nutrition education intervention for the management of polycystic ovary syndrome (PCOS). | Inappropriate study design |
| 1198 | Cognitive Control Training as an Augmentation Strategy to CBT in the Treatment of Fear of Failure in Undergraduates. | Inappropriate population |
| 1199 | The IGNITE trial: Participant recruitment lessons prior to SARS-CoV-2. | Inappropriate population |
| 1200 | Adverse effects of COVID-19-related lockdown on pain, physical activity and psychological well-being in people with chronic pain. | Inappropriate study design |
| 1201 | Getting Physical: A Specific Boost for Cognition in Epilepsy?. | Inappropriate population |
| 1202 | The effects of exergaming on pain, postural control, technology acceptance and flow experience in older people with chronic musculoskeletal pain: A randomised controlled trial. | Inappropriate study design |
| 1203 | Aquatic exercise training for falls and potentially modifiable risk factors of falls in older people: A blinded randomized controlled trial protocol: Aquatic exercise protocol on falls. | Inappropriate study design |
| 1204 | The effect of aerobic exercises on cognition, physical fitness, and apolipoprotein E plasma among women with mild cognitive impairment. | Inappropriate study design |
| 1205 | Effects of Aerobic Exercise, Cognitive and Combined Training on Cognition in Physically Inactive Healthy Late-Middle-Aged Adults: The Projecte Moviment Randomized Controlled Trial. | Inappropriate outcome |
| 1206 | Wuqinxi exercise improves hand dexterity in patients with Parkinson's disease. | Inappropriate outcome |
| 1207 | Slow-Paced Breathing and Autonomic Function in People Post-stroke. | Inappropriate study design |
| 1208 | Impaired Firing Behavior of Individually Tracked Paretic Motor Units During Fatiguing Contractions of the Dorsiflexors and Functional Implications Post Stroke. | Inappropriate study design |
| 1209 | Development of a novel cognitive composite outcome to assess therapeutic effects of exercise in the EXERT trial for adults with MCI: The ADAS-Cog-Exec. | Inappropriate study design |
| 1210 | Effect of a multicomponent exercise intervention on brain metabolism: A randomized controlled trial on Alzheimer's pathology (Dementia-MOVE). | Inappropriate study design |
| 1211 | Feasibility of Imported Self-Management Program for Elderly People with Chronic Pain: A Single-Arm Confirmatory Trial. | Inappropriate study design |
| 1212 | Shedding light on the effects of moderate acute exercise on working memory performance in healthy older adults: An fnirs study. | Inappropriate population |
| 1213 | The effect of bio psychosocial model of rehabilitation on pain and quality of life after total knee replacement: A randomized controlled trial. | Inappropriate population |
| 1214 | Effectiveness of kinesio taping in bicipital tendinitis treatment: A randomized controlled trial kinesio taping in bicipital tendinitis. | Inappropriate study design |
| 1215 | Higher disease and pain severity and fatigue and lower balance skills are associated with higher prevalence of falling among individuals with the inflammatory disease of neuromyelitis optica spectrum disorder (NMOSD). | Inappropriate study design |
| 1216 | Rationale and Design of the PAIRED Trial: Partnered Dance Aerobic Exercise as a Neuroprotective, Motor, and Cognitive Intervention in Parkinson's Disease. | Inappropriate population |
| 1217 | Effects of the agility boot camp with cognitive challenge (ABC-C) exercise program for Parkinson's disease. | Inappropriate study design |
| 1218 | High-Intensity Interval Cycle Ergometer Training in Parkinson's Disease: Protocol for Identifying Individual Response Patterns Using a Single-Subject Research Design. | Inappropriate study design |
| 1219 | Physical Activity Pattern among Female College Students at Taibah University. | Inappropriate population |
| 1220 | Cardiovascular diseases risk factors among young adults: A stepwise-approach-based study in three university setups in morocco. | Inappropriate population |
| 1221 | Effects of Ramadan fasting on match-related changes in skill performance in elite Muslim badminton players. | Inappropriate study design |
| 1222 | The Effects of Nordic Walking With Poles With an Integrated Resistance Shock Absorber on Cognitive Abilities and Cardiopulmonary Efficiency in Postmenopausal Women. | Inappropriate study design |
| 1223 | Stretching intervention for pediatric caregivers in a university hospital. | Inappropriate population |
| 1224 | Pain threshold assessment in relation to neural mobilization therapy. | Inappropriate population |
| 1225 | High-intensity interval training improves cognitive flexibility in older adults. | Inappropriate study design |

| | | |
|---|---|---|
| 1226 | Closure Device-Related Thrombosis After Anticoagulation With Dabigatran in Patients Undergoing Percutaneous Left Atrial Appendage Closure: Case Reports and Literature Review. | Inappropriate study design |
| 1227 | Patterns of cardiopulmonary response to exercise in fibrotic ILD. | Inappropriate study design |
| 1228 | Effects of In-Bed Cycle Exercise in Patients With Acute Stroke: A Randomized Controlled Trial. | Inappropriate study design |
| 1229 | Greater discomfort with blood flow restriction training compared to standard low-load resistance exercise when both are performed to task failure. | Inappropriate population |
| 1230 | Do core stability exercises improve upper limb function in chronic stroke patients?. | Inappropriate study design |
| 1231 | Independent and Combined Effects of All-Out Sprint and Low-Intensity Continuous Exercise on Plasma Oxidative Stress Biomarkers in Trained Judokas. | Inappropriate study design |
| 1232 | Non-immersive virtual reality for post-stroke upper extremity rehabilitation: A small cohort randomized trial. | Inappropriate study design |
| 1233 | A cross-sectional study on practise of physical activity among medical students in Chennai. | Inappropriate population |
| 1234 | Acute moderate-intensity exercise generally enhances neural resources related to perceptual and cognitive processes: A randomized controlled ERP study. | Inappropriate outcome |
| 1235 | Walking for the assessment of patients with COPD. | Inappropriate outcome |
| 1236 | Physical Activity Patterns in People With Inflammatory Arthritis Indicate They Have not Received Recommendation-Based Guidance From Health Care Providers. | Inappropriate study design |
| 1237 | Effect of shooting experience on executive function: Differences between experts and novices. | Inappropriate study design |
| 1238 | Menstrual problems in undergraduate medical students: A cross-sectional study in a medical college of north india. | Inappropriate study design |
| 1239 | Validity of the Pregnancy Physical Activity Questionnaire for Maternal Recall. | Inappropriate study design |
| 1240 | Honest, Open, Proud (HOP)! Acceptability and feasibility of the French version of a programme to reduce self-stigma of mental illness. | Inappropriate study design |
| 1241 | Exercise intensity may not moderate the acute effects of functional circuit training on cognitive function: A randomized crossover trial. | Inappropriate population |
| 1242 | Effects of multicomponent exercise on cognitive function in elderly korean individuals. | Inappropriate study design |
| 1243 | Trajectories of body mass index and their associations with mortality among older adults in Korea: Analysis of the Korean longitudinal study of aging. | Inappropriate study design |
| 1244 | Effects of combining high-definition transcranial direct current stimulation with short-foot exercise on chronic ankle instability: A pilot randomized and double-blinded study. | Inappropriate study design |
| 1245 | The Effect of Aerobic Exercise on Physical and Cognitive Outcomes in a Small Cohort of Outpatients with Schizophrenia. | Inappropriate study design |
| 1246 | Cardiorespiratory Fitness Associates with Cerebral Vessel Pulsatility in a Cohort Enriched with Risk for Alzheimer's Disease. | Inappropriate study design |
| 1247 | Competition Enhances the Effectiveness and Motivation of Attention Rehabilitation After Stroke. A Randomized Controlled Trial. | Inappropriate intervention |
| 1248 | Classification of young healthy individuals with different exercise levels based on multiple musculoskeletal ultrasound images. | Inappropriate population |
| 1249 | Feasibility of Prehabilitation Prior to Breast Cancer Surgery: A Mixed-Methods Study. | Inappropriate study design |
| 1250 | Effect of comprehensive strength training intervention on middle-aged and elderly MCI patients of miao nationality and its mechanism. | Inappropriate study design |
| 1251 | Dynamic right ventricular function response to incremental exercise in pulmonary hypertension. | Inappropriate study design |
| 1252 | Variations and misconceptions of asthma terminology, triggers and therapy among the caregivers of asthmatic children in Makkah, Saudi Arabia. | Inappropriate population |
| 1253 | Influence of the nursing effect on the antiinflammatory effect of atorvastatin in acute cerebral infarction. | Inappropriate population |
| 1254 | Effect of adding motor imagery to task specific training on facilitation of sit to stand in hemiparetic patients. | Inappropriate study design |
| 1255 | The roles of some agility performance parameters on the linear, single sprint skills of young male basketball and handball players. | Inappropriate study design |
| 1256 | Free-weight resistance exercise is more effective in enhancing inhibitory control than machine-based training: A randomized, controlled trial. | Inappropriate study design |
| 1257 | Metastatic brain tumors: To treat or not to treat, and with what?. | Inappropriate study design |
| 1258 | Left Ventricular Structure and Function in Elite Swimmers and Runners. | Inappropriate population |

| | | |
|---|---|---|
| 1259 | Somatotypes trajectories during adulthood and their association with copd phenotypes. | Inappropriate population |
| 1260 | The Cone Flare Crush Modified-T (CFCT) stenting technique for coronary artery bifurcation lesions. | Inappropriate study design |
| 1261 | The predictive role of executive functions and psychological factors on chronic pain after orthopaedic surgery: A longitudinal cohort study. | Inappropriate population |
| 1262 | Clinical approach to diagnosis and assessment. | Inappropriate population |
| 1263 | The feasibility and acceptability of high-intensity interval training for adults with mental illness: A pilot study. | Inappropriate study design |
| 1264 | Regular aerobic exercise increases dispositional mindfulness in men: A randomized controlled trial. | Inappropriate study design |
| 1265 | Boxing to Improve Sleep Quality and Daytime Sleepiness in Individuals with Parkinson Disease: Pilot Study. | Inappropriate study design |
| 1266 | Validation of a method to estimate stroke volume from brachial-cuff derived pressure waveforms. | Inappropriate population |
| 1267 | To compare the effectiveness of laser, EMG biofeedback assisted core stability exercise versus laser and core stability exercise alone on pain and disability in patients with non-specific low back pain. | Inappropriate population |
| 1268 | The prediction of swim start performance based on squat jump force-time characteristics. | Inappropriate study design |
| 1269 | Awareness of physicians in yemen toward high blood pressure management according to the eighth joint national committee (Jnc 8) guideline. | Inappropriate population |
| 1270 | Association between accelerometer-measured physical activity and falls among community-dwelling older people living in cold, snowy areas. | Inappropriate study design |
| 1271 | Automated nociceptive withdrawal reflex measurements reveal normal reflex thresholds and augmented pain ratings in patients with fibromyalgia. | Inappropriate study design |
| 1272 | A questionnaire study on knowledge and awareness of metabolic syndrome and it's components in undergraduate medical students at entry level. | Inappropriate population |
| 1273 | Impact of Supervised Exercise on One-Year Medication Use in Older Veterans with Multiple Morbidities. | Inappropriate study design |
| 1274 | An analysis of the perceived causes leading to task-failure in resistance-exercises. | Inappropriate population |
| 1275 | A randomized controlled clinical trial of a core stability exercise program for the intrinsic motivation of Parkinson's patients. | Inappropriate study design |
| 1276 | Effects of air-perfused rucksack on physiological and perceptual strain during low-intensity exercise in a hot environment. | Inappropriate study design |
| 1277 | Walking speed is the sole determinant criterion of sarcopenia of mild cognitive impairment in japanese elderly patients with type 2 diabetes mellitus. | Inappropriate study design |
| 1278 | Research on the effect of Yoga on the recovery of motor and neurological functions of stroke patients. | Inappropriate study design |
| 1279 | Increased prefrontal activity and reduced motor cortex activity during imagined eccentric compared to concentric muscle actions. | Inappropriate population |
| 1280 | Fick principle and exercise pulmonary hemodynamic determinants of the six-minute walk distance in pulmonary hypertension. | Inappropriate population |
| 1281 | Research Topic: The realities of religious coping experiences of patients with diabetes mellitus: Implications for policy formulation in Ghana. | Inappropriate population |
| 1282 | Effect of a High-Intensity Tandem Bicycle Exercise Program on Clinical Severity, Functional Magnetic Resonance Imaging, and Plasma Biomarkers in Parkinson's Disease. | Inappropriate study design |
| 1283 | Peripheral BDNF Response to Physical and Cognitive Exercise and Its Association With Cardiorespiratory Fitness in Healthy Older Adults. | Inappropriate outcome |
| 1284 | Immersive virtual reality in stroke patients as a new approach for reducing postural disabilities and falls risk: A case series. | Inappropriate study design |
| 1285 | Effects of super-ultramarathon running on cardiac structure and function in middle-aged men. | Inappropriate population |
| 1286 | Are the antagonist muscles fatigued during a fatigue task of agonist muscles?. | Inappropriate population |
| 1287 | Effects of a single exercise workout on memory and learning functions in young adults-A systematic review. | Inappropriate study design |
| 1288 | Evaluating the prevalence and potential predictors for asthma among male university students: A randomized observational study. | Inappropriate study design |
| 1289 | Primary reinforcement with rectus abdominis muscle flap-a salvage technique for a tenuous post traumatic duodenal perforation- a case report. | Inappropriate outcome |
| 1290 | Short- and long-term changes in cognitive function after exercise-based rehabilitation in people with COPD: A pilot study. | Inappropriate study design |

| | | |
|---|---|---|
| 1291 | Quantitative ultrasound imaging differences in multifidus and thoracolumbar fasciae between athletes with and without chronic lumbopelvic pain: A case-control study. | Inappropriate population |
| 1292 | Effects of chronic exercise on temporal discounting among persons with methamphetamine use disorder. | Inappropriate study design |
| 1293 | A novel role of growth differentiation factor (GDF)-15 in overlap with sedentary lifestyle and cognitive risk in COPD. | Inappropriate study design |
| 1294 | Treadmill exercise buffers behavioral alterations related to ethanol binge-drinking in adolescent mice. | Inappropriate study design |
| 1295 | Effects of over-ground walking and cognitive rehabilitation on cognition, brain-derived neurotrophic factor, participation and quality of life among stroke survivors: a study protocol. | Inappropriate population |
| 1296 | The impact of a primary care, pharmacist-driven intervention in patients with chronic non-cancer pain-A pilot study. | Inappropriate population |
| 1297 | Machine learning detects pattern of differences in functional magnetic resonance imaging (FMRI) data between chronic fatigue syndrome (CFS) and gulf war illness (GWI). | Inappropriate study design |
| 1298 | Effects of rivastigmine hydrogen tartrate and donepezil hydrochloride on the cognitive function and mental behavior of patients with Alzheimer's disease. | Inappropriate study design |
| 1299 | Innovative long-dose neurorehabilitation for balance and mobility in chronic stroke: A preliminary case series. | Inappropriate study design |
| 1300 | Effects of mindfulness-based stress reduction on cervical cancer patients undergoing concurrent radiochemotherapy. | Inappropriate population |
| 1301 | Feasibility of a smartphone app to enhance physical activity in progressive MS: A pilot randomized controlled pilot trial over three months. | Inappropriate study design |
| 1302 | MNX (Medium Duration Nutrition and Resistance-Vibration Exercise) Bed-Rest: Effect of Resistance Vibration Exercise Alone or Combined With Whey Protein Supplementation on Cardiovascular System in 21-Day Head-Down Bed Rest. | Inappropriate study design |
| 1303 | Long-Term Spinal Cord Stimulation After Chronic Complete Spinal Cord Injury Enables Volitional Movement in the Absence of Stimulation. | Inappropriate population |
| 1304 | Effects of moderate intensity exercise on the cortical thickness and subcortical volumes of preclinical alzheimer's disease patients: A pilot study. | Inappropriate study design |
| 1305 | Tracheostomy ventilation outside the intensive care unit: Experience of a tertiary care hospital. | Inappropriate study design |
| 1306 | Findings of a Pilot Study Investigating the Effects of Mediterranean Diet and Aerobic Exercise on Cognition in Cognitively Healthy Older People Living Independently within Aged-Care Facilities: The Lifestyle Intervention in Independent Living Aged Care (LIILAC) Study. | Inappropriate study design |
| 1307 | Inclusion of People Poststroke in Cardiac Rehabilitation Programs in Canada: A Missed Opportunity for Referral. | Inappropriate study design |
| 1308 | Effect of tactile stimulation and joint integrity exercises on hemineglect in stroke survivors. | Inappropriate study design |
| 1309 | Psychometric properties of Hindi version of physical activity recall assessment for people with spinal cord injury. | Inappropriate study design |
| 1310 | The effect of participating in MedEx Wellness, a community-based chronic disease exercise rehabilitation programme, on physical, clinical and psychological health: A study protocol for a cohort trial. | Inappropriate study design |
| 1311 | Gross motor skills training leads to increased brain-derived neurotrophic factor levels in healthy older adults: A pilot study. | Inappropriate study design |
| 1312 | Prolonged sitting interrupted by 6-min of high-intensity exercise: Circulatory, metabolic, hormonal, thermal, cognitive, and perceptual responses. | Inappropriate study design |
| 1313 | Effect of Bosu Pilates on Primary Dysmenorrhea in Adolescent Girls. | Inappropriate population |
| 1314 | Sustained isometric wrist flexion and extension maximal voluntary contractions on corticospinal excitability to forearm muscles during low-intensity hand-gripping. | Inappropriate population |
| 1315 | Fitness-dependent effect of acute aerobic exercise on executive function. | Inappropriate intervention |
| 1316 | High-intensity training as a novel treatment for impaired awareness of hypoglycaemia in type 1 diabetes [HIT4HYPOS]: Protocol for a randomized parallel-group study. | Inappropriate population |
| 1317 | Exergaming With Integrated Head Turn Tasks Improves Compensatory Saccade Pattern in Some Patients With Chronic Peripheral Unilateral Vestibular Hypofunction. | Inappropriate population |
| 1318 | The effect of aerobic training and vitamin D supplements on the neurocognitive functions of elderly women with sleep disorders. | Inappropriate population |

| | | |
|---|---|---|
| 1319 | Cardiovascular Disease Risk Is Associated with Middle Cerebral Artery Blood Flow Velocity in Older Adults. | Inappropriate study design |
| 1320 | Spontaneous sternal fracture during labor in a healthy primigravida with female athlete triad: A case report. | Inappropriate outcome |
| 1321 | The Predicament of Caring: Work Interferences and Health of Family Caregivers of Persons With Multiple Chronic Conditions. | Inappropriate study design |
| 1322 | Does Cardiorespiratory Fitness Buffer Stress Reactivity and Stress Recovery in Police Officers? A Real-Life Study. | Inappropriate study design |
| 1323 | Successful rapid weight reduction and the use of liraglutide for morbid obesity in adolescent Prader-Willi syndrome. | Inappropriate study design |
| 1324 | Rhythmic Auditory Stimulation as an Adjuvant Therapy Improved Post-stroke Motor Functions of the Upper Extremity: A Randomized Controlled Pilot Study. | Inappropriate population |
| 1325 | Interindividual differences in the responses to aerobic exercise in Alzheimer's disease: Findings from the FIT-AD trial. | Inappropriate study design |
| 1326 | Disease self-management in patients with moderate COPD: a thematic analysis. | Inappropriate study design |
| 1327 | A Comprehensive Intervention for Promoting Successful Aging Amongst Older People With Diabetes With Below-Normal Cognitive Function-A Feasibility Study. | Inappropriate study design |
| 1328 | Case report: Liver Transplantation in a highly dependent Down Syndrome patient. | Inappropriate study design |
| 1329 | Oral Contraceptive Use Influences On-Kinetic Adaptations to Sprint Interval Training in Recreationally-Active Women. | Inappropriate study design |
| 1330 | Rehabilitative Impact of Exercise Training on Human Skeletal Muscle Transcriptional Programs in Parkinson's Disease. | Inappropriate study design |
| 1331 | Effects of a sit-stand-walk intervention on musculoskeletal discomfort, productivity, and perceived physical and mental fatigue, for computer-based work. | Inappropriate study design |
| 1332 | Soft Tissue: A Possible Source of Pain Pre and Post Minimally Invasive Spine Surgery. | Inappropriate population |
| 1333 | Acute remote ischemic preconditioning has no effect on quadriceps muscle endurance. | Inappropriate study design |
| 1334 | Walking Training Enhances Corticospinal Excitability in Progressive Multiple Sclerosis-A Pilot Study. | Inappropriate study design |
| 1335 | Stress and recovery perception, creatine kinase levels, and performance parameters of male volleyball athletes in a preseason for a championship. | Inappropriate study design |
| 1336 | Comparative study of the effectiveness of a low-pressure hyperbaric oxygen treatment and physical exercise in women with fibromyalgia: randomized clinical trial. | Inappropriate study design |
| 1337 | Physical Exercise May Increase Plasma Concentration of High-Density Lipoprotein-Cholesterol in Patients With Alzheimer's Disease. | Inappropriate population |
| 1338 | A low caffeine dose improves maximal strength, but not relative muscular endurance in either heavier-or lighter-loads, or perceptions of effort or discomfort at task failure in females. | Inappropriate population |
| 1339 | Exploring human trainability: Design and rationale of Studies of Twin Responses to Understand Exercise as a Therapy (STRUETH) study. | Inappropriate study design |
| 1340 | Effectiveness of relaxation training in addition to stabilization exercises in chronic neck pain: A randomized clinical trial. | Inappropriate study design |
| 1341 | Balloon pulmonary angioplasty with stent implantation as a treatment of proximal chronic thromboembolic pulmonary hypertension. | Inappropriate population |
| 1342 | The effect of a multicomponent dual-task exercise on cortical thickness in older adults with cognitive decline: A randomized controlled trial. | Inappropriate population |
| 1343 | Development of an integrative medicine program for breast cancer patients at the largest tertiary referral center in Thailand. | Inappropriate study design |
| 1344 | The role of the psychologist in the inpatient pain service: development and initial outcomes. | Inappropriate population |
| 1345 | A digital decision support system (selfBACK) for improved self-management of low back pain: A pilot study with 6-week follow-up. | Inappropriate population |
| 1346 | The frequency and influencing factors of common geriatric syndromes in hospitalized patients. | Inappropriate population |
| 1347 | Effect of high flow nasal cannula on peripheral muscle oxygenation and hemodynamic during paddling exercise in patients with chronic obstructive pulmonary disease: A randomized controlled trial. | Inappropriate population |
| 1348 | Influence of maternal aerobic exercise during pregnancy on fetal cardiac function and outflow. | Inappropriate study design |
| 1349 | Moderating effect of social interaction on enjoyment and perception of physical activity in young adults with autism spectrum disorders. | Inappropriate study design |

| | | |
|---|---|---|
| 1350 | Effect of Combined Mental Task and Metaboreflex Activation on Hemodynamics and Cerebral Oxygenation in Patients With Metabolic Syndrome. | Inappropriate study design |
| 1351 | Efficacy of a video-based physical therapy and rehabilitation system in patients with post-stroke hemiplegia: A randomized, controlled, pilot study. | Inappropriate population |
| 1352 | Protocol of Aerobic Exercise and Cognitive Health (REACH): A Pilot Study. | Inappropriate study design |
| 1353 | Transcranial direct current stimulation at 4 mA induces greater leg muscle fatigability in women compared to men. | Inappropriate study design |
| 1354 | Medication adherence in stroke patients in Brunei Darussalam Public Hospital: the cross-sectional study associated with chronic diseases, life style and potential barriers. | Inappropriate study design |
| 1355 | Cognitive inhibition correlates with exercise-induced hypoalgesia after aerobic bicycling in pain-free participants. | Inappropriate study design |
| 1356 | The influence of self-efficacy intervention on the mental resilience of patients with traumatic fractures. | Inappropriate study design |
| 1357 | Cognitive Load of Exercise Influences Cognition and Neuroplasticity of Healthy Elderly: An Exploratory Investigation. | Inappropriate study design |
| 1358 | Assessment of the Effects of Aerobic Fitness on Cerebrovascular Function in Young Adults Using Multiple Inversion Time Arterial Spin Labeling MRI. | Inappropriate study design |
| 1359 | The Impact of Rehabilitation-oriented Virtual Reality Device in Patients With Ischemic Stroke in the Early Subacute Recovery Phase: Study Protocol for a Phase III, Single-Blinded, Randomized, Controlled Clinical Trial. | Inappropriate population |
| 1360 | Acute effects of high-definition transcranial direct current stimulation on foot muscle strength, passive ankle kinesthesia, and static balance: A pilot study. | Inappropriate study design |
| 1361 | Ischemic Preconditioning Did Not Affect Central and Peripheral Factors of Performance Fatigability After Submaximal Isometric Exercise. | Inappropriate study design |
| 1362 | Asking an eyewitness to predict their later lineup performance could harm the confidence-accuracy relationship. | Inappropriate study design |
| 1363 | Characteristics of patients at initiation of renal replacement therapy - experience of a hemodialysis center. | Inappropriate study design |
| 1364 | Morning exercise improves cognitive performance decrements induced by partial sleep deprivation in elite athletes. | Inappropriate population |
| 1365 | Combined and isolated effects of acute exercise and brain stimulation on executive function in healthy young adults. | Inappropriate study design |
| 1366 | Acute effects of aerobic exercise on cognitive attention and memory performance: An investigation on duration-based dose-response relations and the impact of increased arousal levels. | Inappropriate study design |
| 1367 | Stability and predictors of poor 6-min walking test performance over 2 years in patients with COPD. | Inappropriate population |
| 1368 | Epidemiology of erectile dysfunction in men with diabetes mellitus: A study in a primary health care center in Indonesia. | Inappropriate study design |
| 1369 | The delayed fatigue effect in myalgic encephalomyelitis/chronic fatigue syndrome (ME/CFS). | Inappropriate population |
| 1370 | The effect of caffeine ingestion on coincidence anticipation timing, perceived exertion, and leg pain during submaximal cycling. | Inappropriate population |
| 1371 | Instrument-assisted soft tissue mobilisation versus stripping massage for upper trapezius myofascial trigger points. | Inappropriate population |
| 1372 | Effects of Physical Exercise on Working Memory and Attention-Related Neural Oscillations. | Inappropriate population |
| 1373 | Feasibility study of goal setting discussions between older adults and volunteers facilitated by an eHealth application: Development of the Health TAPESTRY approach. | Inappropriate study design |
| 1374 | Temporal associations between physical activity, mental activity and fatigue dimensions in knee osteoarthritis: an exploratory intensive longitudinal study. | Inappropriate study design |
| 1375 | Effectiveness of dysphagia exercises on swallowing ability among patients with cerebrovascular accidents. | Inappropriate study design |
| 1376 | South Korean study to prevent cognitive impairment and protect brain health through lifestyle intervention in at-risk elderly people: Protocol of a multicenter, randomized controlled feasibility trial. | Inappropriate study design |
| 1377 | Return to Everyday Activity in the Community and Home: A feasibility study for a lifestyle intervention to sit less, move more, and be strong. | Inappropriate study design |
| 1378 | Agility-based exercise training compared to traditional strength and balance training in older adults: A pilot randomized trial. | Inappropriate population |
| 1379 | How Do Patients With Axial Spondyloarthritis Experience High-Intensity Exercise?. | Inappropriate study design |
| 1380 | How to assess the benefits of pulmonary rehabilitation. | Inappropriate population |

| | | |
|---|---|---|
| 1381 | Effects of combined exercise on psychological and physiological variables in cancer patients: A pilot study. | Inappropriate study design |
| 1382 | The effect of rehabilitation nursing based on Roy's adaptation theory on the prognoses of acute cerebral infarct patients after intravascular intervention. | Inappropriate study design |
| 1383 | Functional outcome of joint mobilization added to task-oriented training on hand function in chronic stroke patients. | Inappropriate population |
| 1384 | Modelling Modifiable Predictors of Age-Related Cognitive Decline: Exercise, Aortic Stiffness, and the Importance of Physical Fitness. | Inappropriate intervention |
| 1385 | Study, Design, and Rationale of Noncommunicable Diseases in Nepal (NCD Nepal) Study: A Community-Based Prospective Epidemiological and Implementation Study in Rural Nepal. | Inappropriate population |
| 1386 | Measurement of pain, delayed onset of muscle soreness and muscle strengths after performing various eccentric exercise protocols with partial range of motion. | Inappropriate outcome |
| 1387 | Multi-sensorimotor training improves proprioception and balance in subacute stroke patients: A randomized controlled pilot trial. | Inappropriate study design |
| 1388 | The contributions of physical activity and fitness for the optimal health and wellness of the elderly people. | Inappropriate population |
| 1389 | Assessment of Exercise Stroke Volume and Its Prediction From Oxygen Pulse in Paralympic Athletes With Locomotor Impairments: Cardiac Long-Term Adaptations Are Possible. | Inappropriate outcome |
| 1390 | Acute and chronic effects of multivitamin/mineral supplementation on objective and subjective energy measures. | Inappropriate study design |
| 1391 | Pilot study on comparisons between the effectiveness of mobile video-guided and paper-based home exercise programs on improving exercise adherence, self-efficacy for exercise and functional outcomes of patients with stroke with 3-month follow-up: A single-blind randomized controlled trial. | Inappropriate study design |
| 1392 | Self-Efficacy to Engage in Physical Exercise and Walking Ability Best Predicted Exercise Adherence after Stroke. | Inappropriate study design |
| 1393 | Dynamic balance and gait speed improve in persons with Parkinson's disease after Lee Silverman Voice Treatment (LSVT)-BIG training: a single subject experimental design study. | Inappropriate study design |
| 1394 | Exercise and caffeine improve sustained attention following fatigue independent of fitness status. | Inappropriate study design |
| 1395 | Muscle Fatigue in the Three Heads of Triceps Brachii During Intensity and Speed Variations of Triceps Push-Down Exercise. | Inappropriate intervention |
| 1396 | Use of standardized patients in endocrinology fellowship programs to teach competent transgender care. | Inappropriate outcome |
| 1397 | Prison primary care and noncommunicable diseases: A data-linkage survey of prevalence and associated risk factors. | Inappropriate population |
| 1398 | Predicting Factors for Return to Prefracture Ambulatory Level in High Surgical Risk Elderly Patients Sustained Intertrochanteric Fracture and Treated With Proximal Femoral Nail Antirotation (PFNA) With and Without Cement Augmentation. | Inappropriate population |
| 1399 | Effects of individual task specific training verses group circuit training on balance and ambulation in sub-acute stroke. | Inappropriate population |
| 1400 | A Transfer Entropy Approach for the Assessment of the Impact of Inspiratory Muscle Training on the Cardiorespiratory Coupling of Amateur Cyclists. | Inappropriate study design |
| 1401 | Challenges associated with physical assessments for people living with dementia: Modifying standard assessment protocols. | Inappropriate population |
| 1402 | Pre-therapeutic device for post-stroke hemiplegic patients' wrist and finger rehabilitation. | Inappropriate population |
| 1403 | Effects of orofacial muscles exercise program on swallowing function and satisfaction in sub-acute stroke patients with dysphagia. | Inappropriate study design |
| 1404 | Effects of intensity and set method on iEMG of flexor carpi ulnaris and performance speed decrease point during barbell curl exercise. | Inappropriate study design |
| 1405 | Effect of immersive virtual reality on balance, mobility, and fatigue in patients with multiple sclerosis: A single-blinded randomized controlled trial. | Inappropriate study design |
| 1406 | Effect of Underwater Treadmill Gait Training With Water-Jet Resistance on Balance and Gait Ability in Patients With Chronic Stroke: A Randomized Controlled Pilot Trial. | Inappropriate study design |
| 1407 | Isolated Lateral Chronic Exertional Compartment Syndrome of the Leg: A New Entity?. | Inappropriate population |
| 1408 | Arthroscopic Treatment of First-Time Shoulder Dislocations in Younger Athletes. | Inappropriate study design |

| | | |
|---|---|---|
| 1409 | Using Pressure Massage for Achilles Tendinopathy: A Single-Blind, Randomized Controlled Trial Comparing a Novel Treatment Versus an Eccentric Exercise Protocol. | Inappropriate study design |
| 1410 | A Prospective Injury Surveillance Study on Ski Touring. | Inappropriate population |
| 1411 | Application of peer support models in respiratory rehabilitation of patients with chronic obstructive pulmonary disease. | Inappropriate population |
| 1412 | Can caffeine intake combined with aerobic exercise lead to improvement in attentional and psychomotor performance in trained individuals?. | Inappropriate population |
| 1413 | Using exercise to facilitate arousal reappraisal and reduce stress reactivity: A randomized controlled trial. | Inappropriate study design |
| 1414 | Effects of Combined Aerobic Exercise and Cognitive Training on Verbal Fluency in Older Adults. | Inappropriate study design |
| 1415 | Associations between Cardiorespiratory Fitness and Endocrine, Autonomous, and Psychological Stress Reactivity in Male Adolescents. | Inappropriate study design |
| 1416 | Effectiveness and cost-effectiveness of Pilates versus home-based exercises in individuals with chronic non-specific low back pain: randomised controlled trial protocol. | Inappropriate study design |
| 1417 | Barriers to physical activity in patients with pulmonary hypertension. | Inappropriate study design |
| 1418 | A comparison of the efficacy of nonweight-bearing and weight-bearing exercise programmes on function and pain pressure thresholds in knee osteoarthritis: a randomised study. | Inappropriate intervention |
| 1419 | The associations of self-reported physical fitness and physical activity with sleep quality in young adults: A population-based study. | Inappropriate study design |
| 1420 | Qualitative study of obesity risk perception, knowledge, and behavior among Hispanic taxi drivers in New York. | Inappropriate study design |
| 1421 | Automatic Detection of Compensation during Robotic Stroke Rehabilitation Therapy. | Inappropriate study design |
| 1422 | Adhering to a Tai Chi Chuan Exercise Program Improves Vascular Resistance and Cardiac Function. | Inappropriate population |
| 1423 | Cognitive effects of a 30-min aerobic exercise bout on adults with overweight/obesity and type 2 diabetes. | Inappropriate population |
| 1424 | Changes in metabolic syndrome status and risk of dementia. | Inappropriate population |
| 1425 | Bronchial asthma versus paroxysmal vocal-fold dysfunction in patients with exercise-induced respiratory symptoms. | Inappropriate study design |
| 1426 | Dual task effect on postural control in patients with degenerative cerebellar disorders. | Inappropriate study design |
| 1427 | Cognitive Remediation Therapy as a Feasible Treatment for a Young Person With Anorexia Nervosa and Autism Spectrum Disorder Comorbidity: A Case Study. | Inappropriate study design |
| 1428 | Acute and chronic effects of resistance exercise training among young adults with and without analogue Generalized Anxiety Disorder: A protocol for pilot randomized controlled trials. | Inappropriate intervention |
| 1429 | Acute Sprint Interval Exercise Increases Both Cognitive Functions and Peripheral Neurotrophic Factors in Humans: The Possible Involvement of Lactate. | Inappropriate population |
| 1430 | Original article comparison of cognitive auditory event related potentials and executive functions in adolescent athletes and non-athletes-a cross sectional study. | Inappropriate study design |
| 1431 | Effects of an In-home Multicomponent Exergame Training on Physical Functions, Cognition, and Brain Volume of Older Adults: A Randomized Controlled Trial. | Inappropriate outcome |
| 1432 | Effect of cardiovascular fitness on cognitive functions among male medical students of South Kerala. | Inappropriate study design |
| 1433 | An unusual cause of exertional dyspnea in a 55 years old man. | Inappropriate study design |
| 1434 | Effect of high intensity interval training compared to continuous training on cognitive performance in young healthy adults: A pilot study. | Inappropriate population |
| 1435 | Autonomic stress response and perceived effort jointly inform on dual tasking in aging. | Inappropriate intervention |
| 1436 | Intradialytic exercise increases cardiac power index. | Inappropriate study design |
| 1437 | The feasibility and effectiveness of a new practical multidisciplinary treatment for low-back pain: A randomized controlled trial. | Inappropriate study design |
| 1438 | Upper limb motor improvement in chronic stroke after combining botulinum toxin A injection and multi-joints robot-assisted therapy: A case report. | Inappropriate study design |

| | | |
|---|---|---|
| 1439 | Effects of acute aerobic exercise on motor response inhibition: An ERP study using the stop-signal task. | Inappropriate population |
| 1440 | Acute exercise is associated with specific executive functions in college students with ADHD: A preliminary study. | Inappropriate population |
| 1441 | Where are the limits of the effects of exercise intensity on cognitive control?. | Inappropriate study design |
| 1442 | The immunomodulary effects of systematic exercise in older adults and people with Parkinson's disease. | Inappropriate study design |
| 1443 | Does Structured Exercise Improve Cognitive Impairment in People with Mild to Moderate Dementia? A Cost-Effectiveness Analysis from a Confirmatory Randomised Controlled Trial: The Dementia and Physical Activity (DAPA) Trial. | Inappropriate study design |
| 1444 | The effect of the presence and severity of bronchiectasis on the respiratory functions, exercise capacity, dyspnea perception, and quality of life in patients with chronic obstructive pulmonary disease. | Inappropriate population |
| 1445 | The effect of a multicomponent exercise program on cognitive function and functional ability in community dwelling older adults. | Inappropriate study design |
| 1446 | Short Duration, Intensive Bed Mobility Training in Idiopathic Parkinson's Patients-One Group Pre-post Test Study Design. | Inappropriate study design |
| 1447 | Possibilities for Correcting Emotional and Behavioral Impairments in Stroke Patients during Rehabilitation Therapy. | Inappropriate study design |
| 1448 | Effectiveness of backward walking in the gait performance of stroke patients. | Inappropriate population |
| 1449 | Brain Glucose Metabolism, Cognition, and Cardiorespiratory Fitness following Exercise Training in Adults at Risk for Alzheimer's Disease. | Inappropriate population |
| 1450 | Caudate Volume Mediates the Interaction between Total Sleep Time and Executive Function after Acute Exercise in Healthy Older Adults. | Inappropriate population |
| 1451 | Modulation of Distinct Intrinsic Resting State Brain Networks by Acute Exercise Bouts of Differing Intensity. | Inappropriate population |
| 1452 | Cardiovascular diseases associated with pseudoexfoliation syndrome - A prospective cross-sectional study. | Inappropriate study design |
| 1453 | Improving memory and executive function of the elderly with mild congnitive impairment-a smart health care approach. | Inappropriate population |
| 1454 | Exercise and horticultural programs for older adults with depressive symptoms and memory problems: A randomized controlled trial. | Inappropriate study design |
| 1455 | Moderate exercise improves brain efficiency for executive functions in young adults. | Inappropriate study design |
| 1456 | Significant acute response of brain-derived neurotrophic factor following a session of extreme conditioning program is correlated with volume of specific exercise training in trained men. | Inappropriate study design |
| 1457 | The Impact of Exercise, Lifestyle, and Clinical Factors on Perceived Cognitive Function in Patients with Rheumatoid Arthritis: Results from a Prospective Cohort Study. | Inappropriate study design |
| 1458 | Psychosocial and physical factors associated with depression in older adults living in studio apartments: A cross-sectional study of a multi-ethnic Asian population. | Inappropriate study design |
| 1459 | How Does a Delay Between Temperate Running Exercise and Hot-Water Immersion Alter the Acute Thermoregulatory Response and Heat-Load?. | Inappropriate study design |
| 1460 | Exercise training enhances platelet mitochondrial bioenergetics in stroke patients: A randomized controlled trial. | Inappropriate population |
| 1461 | Physical Activity Reduces Epilepsy Incidence: a Retrospective Cohort Study in Swedish Cross-Country Skiers and an Experimental Study in Seizure-Prone Synapsin II Knockout Mice. | Inappropriate population |
| 1462 | The effects of different stretching and warm-up exercise protocols on 50-meter swimming performance in sub-elite women swimmers. | Inappropriate population |
| 1463 | Assessment of prescribing pattern of oral anti diabetic drugs in a tertiary care hospital. | Inappropriate study design |
| 1464 | Reactivate-A goal-orientated rehabilitation program for adolescent and young adult cancer survivors. | Inappropriate study design |
| 1465 | Sarc-F and muscle function in community dwelling adults with aged care service needs: Baseline and post-training relationship. | Inappropriate study design |
| 1466 | Factors associated with non-Attendance at exercise-based cardiac rehabilitation. | Inappropriate study design |
| 1467 | First Clinicogenetic Description of Parkinson's Disease Related to GBA Mutation S107L. | Inappropriate population |
| 1468 | Moderate-Intensity Aerobic Exercise Restores Appetite and Prefrontal Brain Activity to Images of Food Among Persons Dependent on Methamphetamine: A Functional Near-Infrared Spectroscopy Study. | Inappropriate study design |

| | | |
|---|---|---|
| 1469 | Acute Low-Intensity Aerobic Exercise Modulates Intracortical Inhibitory and Excitatory Circuits in an Exercised and a Non-exercised Muscle in the Primary Motor Cortex. | Inappropriate population |
| 1470 | Comparison of the effect of 8-week rebound therapy-based exercise program and weight-supported exercises on the range of motion, proprioception, and the quality of life in patients with Parkinson's disease. | Inappropriate study design |
| 1471 | Influence of acute combined physical and cognitive exercise on cognitive function: An NIRS study. | Inappropriate study design |
| 1472 | Effect of self-management program on health status of elderly patients with heart failure: A single-blind, randomized clinical trial. | Inappropriate population |
| 1473 | Muscular Tuberculosis: A New Case and a Review of the Literature. | Inappropriate population |
| 1474 | Prevalence of hypertension, diabetes, and dyslipidemia, and their additive effects on myocardial infarction and stroke: A cross-sectional study in Nanjing, China. | Inappropriate study design |
| 1475 | Feasibility of an at-home, web-based, interactive exercise program for older adults. | Inappropriate study design |
| 1476 | Acceptability, Feasibility and Preliminary Evaluation of a Novel, Personalised, Home-Based Physical Activity Intervention for Chronic Heart Failure (Active-at-Home-HF): a Pilot Study. | Inappropriate study design |
| 1477 | The Brain Health Champion study: Health coaching changes behaviors in patients with cognitive impairment. | Inappropriate outcome |
| 1478 | The responsiveness of the Manchester Chronic Obstructive Pulmonary Disease Fatigue Scale to pulmonary rehabilitation. | Inappropriate intervention |
| 1479 | Promoting independence in Lewy body dementia through exercise (PRIDE) study: Protocol for a pilot study. | Inappropriate study design |
| 1480 | Semi-Recumbent Vibration Exercise in Older Adults: A Pilot Study of Methodology, Feasibility, and Safety. | Inappropriate outcome |
| 1481 | Effective strategy of the multicomponent exercise program for older individuals in a depopulated rural region. | Inappropriate study design |
| 1482 | Effects of acute normobaric hypoxia on memory interference. | Inappropriate population |
| 1483 | Investigating performance in a strenuous physical task from the perspective of self-control. | Inappropriate population |
| 1484 | Behavioural interventions targeting physical activity improve psychocognitive outcomes in COPD. | Inappropriate study design |
| 1485 | Short-Term Music Therapy Attention and Arousal Regulation Treatment (SMAART) for Prisoners with Posttraumatic Stress Disorder: A Feasibility Study. | Inappropriate study design |
| 1486 | Relationship between oxygen pulse and arteriovenous oxygen difference in healthy subjects: Effect of exercise intensity. | Inappropriate study design |
| 1487 | Efficacy of dual-task training with two different priorities instructional sets on gait parameters in patients with chronic stroke. | Inappropriate study design |
| 1488 | Pro-health behaviors of women in given trimesters of pregnancy - A preliminary study. | Inappropriate population |
| 1489 | The negative effects of anterior cruciate ligament injury during stroke rehabilitation. | Inappropriate population |
| 1490 | Cognitive dysfunction among adults with type 2 diabetes mellitus in Karnataka, India. | Inappropriate population |
| 1491 | Effects of dual-task training with different intensity of aerobic exercise on cognitive function and neurotrophic factors in chronic stroke patients. | Inappropriate population |
| 1492 | Improvements in functional and cognitive status following short-term pulmonary rehabilitation in COPD lung transplant recipients: A pilot study. | Inappropriate population |
| 1493 | Restless leg syndrome in peripheral artery disease: Prevalence among patients with claudication and benefits from low-intensity exercise. | Inappropriate study design |
| 1494 | Pain symptomology, functional impact, and treatment of people with neurofibromatosis type 1. | Inappropriate study design |
| 1495 | Multidisciplinary pain management program for patients with chronic musculoskeletal pain in Japan: A cohort study. | Inappropriate population |
| 1496 | Development of an EMG-controlled knee exoskeleton to assist home rehabilitation in a game context. | Inappropriate study design |
| 1497 | Effects and Mechanisms of Cognitive, Aerobic Exercise, and Combined Training on Cognition, Health, and Brain Outcomes in Physically Inactive Older Adults: The Projecte Moviment Protocol. | Inappropriate study design |
| 1498 | Physical exercise: Bulking up neurogenesis in human adults. | Inappropriate population |
| 1499 | Long-Term Effects of Balance Training on Habitual Physical Activity in Older Adults with Parkinson's Disease. | Inappropriate outcome |
| 1500 | Brain-derived neurotrophic factor, epigenetics in stroke skeletal muscle, and exercise training. | Inappropriate study design |

| | | |
|---|---|---|
| 1501 | Use of Ayurveda in the Treatment of Type 2 Diabetes Mellitus. | Inappropriate population |
| 1502 | Development and Preliminary Validation of a Stroke Physical Activity Questionnaire. | Inappropriate study design |
| 1503 | Critical peripheral fatigue thresholds among different force-velocity conditions: An individual-based model approach. | Inappropriate study design |
| 1504 | Dynamic right ventricular-pulmonary arterial uncoupling during maximum incremental exercise in exercise pulmonary hypertension and pulmonary arterial hypertension. | Inappropriate study design |
| 1505 | Progression of blood flow restricted resistance training in older adults at risk of mobility limitations. | Inappropriate population |
| 1506 | The effects of superimposed whole-body electromyostimulation during short-term strength training on physical fitness in physically active females: A randomized controlled trial. | Inappropriate study design |
| 1507 | Tailored text message prompts to increase therapy homework adherence: A single-case randomised controlled study. | Inappropriate intervention |
| 1508 | Heat dissipating upper body compression garment: Thermoregulatory, cardiovascular, and perceptual responses. | Inappropriate study design |
| 1509 | Predictors of 30-day outcomes following mitral valve repair. | Inappropriate population |
| 1510 | Influence of a water physical rehabilitation program on the hemodynamic parameters in breast cancer survivors. | Inappropriate study design |
| 1511 | Assessment of metered-dose inhalers technique among patients with chronic respiratory disorders at al- hussein teaching hospital in al- nasiriyah city. | Inappropriate study design |
| 1512 | Cardiovascular response to exercise in hypertension - clinical characteristics of ASSECURE study participants. | Inappropriate study design |
| 1513 | Intermittent use of portable NIV increases exercise tolerance in COPD: A randomised, cross-over trial. | Inappropriate study design |
| 1514 | Comparative effect of driving side on low back pain due to Repetitive Ipsilateral Rotation. | Inappropriate population |
| 1515 | The effect of aerobic physical rehabilitation on the quality of life in patients with chronic atrial fibrillation; a randomized controlled clinical trial study. | Inappropriate population |
| 1516 | Inter-individual differences in cognitive response to a single bout of physical exercise-A randomized controlled cross-over study. | Inappropriate outcome |
| 1517 | The Interaction of Cognitive Interference, Standing Surface, and Fatigue on Lower Extremity Muscle Activity. | Inappropriate intervention |
| 1518 | The impact of physical exercise on brain-derived neurotrophic factor (BDNF) level in elderly population. | Inappropriate study design |
| 1519 | The effect of cycling through a projection-based virtual environment system on generalized anxiety disorder. | Inappropriate study design |
| 1520 | Combined intervention of physical activity, aerobic exercise, and cognitive exercise intervention to prevent cognitive decline for patients with mild cognitive impairment: A randomized controlled clinical study. | Inappropriate study design |
| 1521 | Impaired skeletal muscle kynurenine metabolism in patients with chronic obstructive pulmonary disease. | Inappropriate study design |
| 1522 | Ginsenoside Rg1 supplementation clears senescence-associated beta-galactosidase in exercising human skeletal muscle. | Inappropriate study design |
| 1523 | Nap opportunity during the daytime affects performance and perceived exertion in 5-m shuttle run test. | Inappropriate study design |
| 1524 | Foam rolling as a recovery tool following eccentric exercise: Potential mechanisms underpinning changes in jump performance. | Inappropriate population |
| 1525 | Effect of home-based high-intensity interval training in patients with lacunar stroke: A randomized controlled trial. | Inappropriate study design |
| 1526 | The effects of exercise and ambient temperature on dietary intake, appetite sensation, and appetite regulating hormone concentrations. | Inappropriate population |
| 1527 | Acute effects of the new method sarcoplasma stimulating training versus traditional resistance training on total training volume, lactate and muscle thickness. | Inappropriate study design |
| 1528 | The relationship between blood flow and motor unit firing rates in response to fatiguing exercise post-stroke. | Inappropriate study design |
| 1529 | Neuromuscular and muscle metabolic functions in melas before and after resistance training: A case study. | Inappropriate outcome |
| 1530 | In patients with multiple sclerosis, both objective and subjective sleep, depression, fatigue, and paresthesia improved after 3 weeks of regular exercise. | Inappropriate population |
| 1531 | Spasticity assessment based on the maximum isometrics voluntary contraction of upper limb muscles in post-stroke hemiplegia. | Inappropriate study design |

| | | |
|---|---|---|
| 1532 | Qualitative study on effective telehealtheducational method chronic obstructive pulmonary disease (COPD) in mojokerto indonesia. | Inappropriate study design |
| 1533 | Landing-takeoff asymmetries applied to running mechanics: A new perspective for performance. | Inappropriate study design |
| 1534 | Can a brief relaxation exercise modulate placebo or nocebo effects in a visceral pain model?. | Inappropriate population |
| 1535 | Fatiguing trunk flexor exercise decreases pain sensitivity in postpartum women. | Inappropriate population |
| 1536 | Improving self-management of people with type 2 diabetes in the first years after diagnosis: Development and pilot of a theory-based interactive group intervention. | Inappropriate study design |
| 1537 | Sprinting after having sprinted: Prior high-intensity stochastic cycling impairs the winning strike for gold. | Inappropriate study design |
| 1538 | High intensity physical rehabilitation later than 24 h post stroke is beneficial in patients: A pilot randomized controlled trial (RCT) study in mild to moderate ischemic stroke. | Inappropriate study design |
| 1539 | Psychosocial working conditions, asthma self-management at work and asthma morbidity: A cross-sectional study. | Inappropriate study design |
| 1540 | Exercises improves pelvic floor muscle function in healthy pregnant women - A randomized control trial. | Inappropriate study design |
| 1541 | Participants' perceptions of Kinesio tape for carpal tunnel syndrome: A qualitative study. | Inappropriate study design |
| 1542 | Implementing Trauma-Informed Care in Primary Medical Settings: Evidence-Based Rationale and Approaches. | Inappropriate study design |
| 1543 | T cell subsets' distribution in elite karate athletes as a response to physical effort. | Inappropriate study design |
| 1544 | The beneficial e_ect of acute exercise on motor memory consolidation is modulated by dopaminergic gene profile. | Inappropriate study design |
| 1545 | Effectiveness of dynamic balance training with and without visual feedback on balance in ambulatory stroke patients. | Inappropriate population |
| 1546 | Moderate- to high-intensity exercise does not modify cortical beta-amyloid in Alzheimer's disease. | Inappropriate population |
| 1547 | Digital thermal analysis as a performance-level indicator to evaluate two volleyball skills for female students during and after menstrual cycle. | Inappropriate study design |
| 1548 | A randomized controlled trial of exercise to prevent muscle mass and functional loss in elderly hemodialysis patients: Rationale, study design, and baseline sample. | Inappropriate study design |
| 1549 | Randomized Controlled Trial of Exercise in CKD-The RENEXC Study. | Inappropriate population |
| 1550 | Quality of life and social support in women with urinary incontinence. | Inappropriate study design |
| 1551 | Non-pharmacological methods and post-operative pain relief: An observational study. | Inappropriate study design |
| 1552 | Effect of exercises on quality of life in patients with postmenopausal osteoporosis - Randomized trial. | Inappropriate study design |
| 1553 | Examining a novel firefighter exercise training program on simulated fire ground test performance, cardiorespiratory endurance, and strength: A pilot investigation. | Inappropriate study design |
| 1554 | Predictors of hospital-based multidisciplinary rehabilitation effects in persons with multiple sclerosis: a large-scale, single-centre study. | Inappropriate study design |
| 1555 | Factors of academic success among undergraduate medical students in Taif University, Saudi Arabia: A cross-sectional study. | Inappropriate study design |
| 1556 | High-cadence cycling promotes sustained improvement in bradykinesia, rigidity, and mobility in individuals with mild-moderate Parkinson's disease. | Inappropriate study design |
| 1557 | Comparison of Indirect Calorimetry- and Accelerometry-Based Energy Expenditure during Object Project Skill Performance. | Inappropriate outcome |
| 1558 | Effect of physiohterapy on fatigue and psychological stress in cancer patients during the course of chemotherapy. | Inappropriate outcome |
| 1559 | Cognitive task during fatiguing exercise increases the sensory-motor performance in professional dancers. | Inappropriate population |
| 1560 | A conceptual model for quality of life among people with type 2 diabetes in the philippines. | Inappropriate population |
| 1561 | Patients with Alzheimer's disease who carry the APOE epsilon4 allele benefit more from physical exercise. | Inappropriate study design |
| 1562 | APOE epsilon4, the door to insulin-resistant dyslipidemia and brain fog? A case study. | Inappropriate outcome |
| 1563 | Use of dietary supplements among physicians at a hospital in Turkey. | Inappropriate population |
| 1564 | Prevalence and awareness of varicocele among athletes in Riyadh, Saudi Arabia. | Inappropriate study design |

| | | |
|---|---|---|
| 1565 | A longitudinal examination of the social-ecological correlates of exercise in men and women following cardiac rehabilitation. | Inappropriate study design |
| 1566 | How the results of a randomized trial of catheter-directed thrombolysis versus anticoagulation alone for submassive pulmonary embolism would affect patient and physician decision making: Report of an online survey. | Inappropriate study design |
| 1567 | Effects of Laser Acupuncture on Delayed Onset Muscle Soreness of the Biceps Brachii Muscle: A Randomized Controlled Trial. | Inappropriate study design |
| 1568 | Therapeutic management of closed central slip injuries: Outcome of a service evaluation. | Inappropriate population |
| 1569 | Effectiveness of web-based personalised e-Coaching lifestyle interventions. | Inappropriate population |
| 1570 | Using Prolonged Exposure With an Older Male U.S. Veteran With Childhood Sexual Abuse-Related PTSD. | Inappropriate study design |
| 1571 | Impact of chronic venous disease on quality of life: Results of vein alarm study. | Inappropriate study design |
| 1572 | Effects of aerobic exercise on sad emotion regulation in young women: an electroencephalograph study. | Inappropriate population |
| 1573 | Temple syndrome diagnosed in an adult patient with clinical autism spectrum disorder. | Inappropriate study design |
| 1574 | Measurement of transversus abdominis activation in chronic low back pain patients using a novel standardized real-time ultrasound imaging method. | Inappropriate study design |
| 1575 | Gender differences in self-care maintenance and its associations among patients with chronic heart failure. | Inappropriate study design |
| 1576 | Effects of antenatal exercise programme and education on health related quality of life: A randomised controlled trial. | Inappropriate study design |
| 1577 | Effect of spinal manipulation on pain sensitivity, postural sway, and health-related quality of life among patients with non-specific chronic low back pain: A randomised control trial. | Inappropriate study design |
| 1578 | Effect of HIIT on cognitive and physical performance. | Inappropriate study design |
| 1579 | Stress fractures of the lateral tibial plateau after open wedge high tibial osteotomy could be delayed type III lateral hinge fractures. | Inappropriate study design |
| 1580 | Inspiratory muscle training for advanced heart failure with lamin-related muscular dystrophy. | Inappropriate study design |
| 1581 | The effects of aerobic and resistance exercise on state anxiety and cognitive function. | Inappropriate population |
| 1582 | Associations between cardiorespiratory fitness, physical activity, intraindividual variability in behavior, and cingulate cortex in younger adults. | Inappropriate study design |
| 1583 | Effect of acute strength training on the posture control during dual tasking and executive function in older adults. A randomized controlled study. | Inappropriate population |
| 1584 | Mental imagery and acute exercise on episodic memory function. | Inappropriate population |
| 1585 | Acute exercise on memory reconsolidation. | Inappropriate study design |
| 1586 | Effects of Acute Aerobic Exercise on Cognitive Flexibility Required During Task-Switching Paradigm. | Inappropriate outcome |
| 1587 | How does a 4-week motor-cognitive training affect choice reaction, dynamic balance and cognitive performance ability? A randomized controlled trial in well-trained, young, healthy participants. | Inappropriate study design |
| 1588 | Vasectomy Simulation Curriculum and Trainer with Enhanced Face, Content and Construct Validity. | Inappropriate study design |
| 1589 | Exercise in Individuals With CKD: A Focus Group Study Exploring Patient Attitudes, Motivations, and Barriers to Exercise. | Inappropriate study design |
| 1590 | Adjustment effects of maximum intensity tolerance during whole-body electromyostimulation training. | Inappropriate study design |
| 1591 | Sex moderates the effect of aerobic exercise on some aspects of cognition in cognitively intact younger and middle-age adults. | Inappropriate population |
| 1592 | Effects of a tailored strength training program of the upper limb combined with transcranial direct current stimulation (tDCS) in chronic stroke patients: Study protocol for a randomised, double-blind, controlled trial. | Inappropriate study design |
| 1593 | Effect of multi-modal exercises after hemiarthroplasty of hip joint in a 72 year old male parkinson's disease participant. A case report. | Inappropriate population |
| 1594 | Comparative study on the effect of intense exercise and yoga on memory. | Inappropriate study design |
| 1595 | Recruitment of cognitive control regions during effortful self-control is associated with altered brain activity in control and reward systems in dieters during subsequent exposure to food commercials. | Inappropriate population |
| 1596 | Value of continuing nursing in chronic heart failure and its effect on patients' cardiac function improvement. | Inappropriate population |
| 1597 | The effect of passive infrared hemoencephalography on athlete performance. | Inappropriate study design |

| | | |
|---|---|---|
| 1598 | Effect of robotic erigo tilt table exercise on neurological parameters in hemiplegic patients: A pilot study. | Inappropriate intervention |
| 1599 | Impact of Exercise Counseling on Physical Function in Chronic Kidney Disease: An Observational Study. | Inappropriate population |
| 1600 | The effects of aerobic exercise on repetitive behaviors and task performance for adults with autism spectrum disorder and an intellectual disability. | Inappropriate population |
| 1601 | Factors associated with health-related quality of life of ambulatory stroke survivors in Jamaica. | Inappropriate population |
| 1602 | Effects of a combined aerobic and resistance exercise program on the quality of life and motor function of elderly men with Parkinson's disease. | Inappropriate population |
| 1603 | The naturopathic management of interstitial cystitis: A case study. | Inappropriate population |
| 1604 | Influence of isometric exercise on pressure pain sensitivity in knee osteoarthritis. | Inappropriate population |
| 1605 | Effect of neuromuscular electrical stimulation on labial and lingual weakness: A single-case preexperimental study. | Inappropriate outcome |
| 1606 | The effects of body weight-supported treadmill training on static and dynamic balance in stroke patients: A pilot, single-blind, randomized trial. | Inappropriate study design |
| 1607 | Sleep duration, lifestyles and chronic diseases: A cross-sectional population-based study. | Inappropriate population |
| 1608 | QT adaptation during exercise in cirrhosis. | Inappropriate study design |
| 1609 | A Comparative Study of Physical Health among Office and Technical Workers in Y Combined Cycle Power Plant in Korea. | Inappropriate population |
| 1610 | Effects of Six-Month Aerobic Exercise Intervention on Sleep in Healthy Older Adults in the Brain in Motion Study: A Pilot Study. | Inappropriate population |
| 1611 | An attributive component of the condition of people with the diagnosis of "ischemic heart disease. Effort angina". | Inappropriate study design |
| 1612 | The role of exercise-induced arousal and exposure to blue-enriched lighting on vigilance. | Inappropriate study design |
| 1613 | It's harder to push, when I have to push hard-physical exertion and fatigue changes reasoning and decision-making on hypothetical moral dilemmas in males. | Inappropriate population |
| 1614 | Exploring strategies to support medication adherence in patients with inflammatory arthritis: A patient-oriented qualitative study using an interactive focus group activity. | Inappropriate study design |
| 1615 | The effect of physiotherapy and acupuncture on psychocognitive, somatic, quality of life, and disability characteristics in TTH patients. | Inappropriate study design |
| 1616 | Knowledge, attitude, and practice on non-communicable diseases (NCDs) among the adult population in the urban area of Negeri Sembilan, Malaysia. | Inappropriate study design |
| 1617 | Yoga as a complementary therapy improves pulmonary functions in patients of bronchial asthma: A randomized controlled trial. | Inappropriate study design |
| 1618 | The experimental effects of acute exercise on long-term emotional memory. | Inappropriate study design |
| 1619 | The effects of yoga in patients suffering from subjective tinnitus. | Inappropriate study design |
| 1620 | Effectiveness of superficial radial nerve block on pain, function and quality of life in patients with hand osteoarthritis: A prospective, randomized and controlled single-blind study. | Inappropriate population |
| 1621 | Participants and developers experiences with a chronic pain self-management intervention under development: A qualitative study. | Inappropriate population |
| 1622 | Hyperbaric therapy influence on psychomotor abilities effectiveness development - Two cases study. | Inappropriate study design |
| 1623 | Short-term interval aerobic exercise training does not improve memory functioning in relapsing-remitting multiple sclerosis- A randomized controlled trial. | Inappropriate population |
| 1624 | Effects of isometric handgrip exercise training on systemic arterial stiffness, cardiovagal baroreflex sensitivity, and cognition in treated adults with hypertension: A pilot study. | Inappropriate study design |
| 1625 | Intensive multimodal training to improve gait resistance, mobility, balance and cognitive function in persons with multiple sclerosis: A pilot randomized controlled trial. | Inappropriate study design |
| 1626 | Exercise in african American and white colorectal cancer survivors: A mixed-methods approach. | Inappropriate population |
| 1627 | Awareness about antenatal and postnatal exercises amongst rural pregnant women. | Inappropriate study design |
| 1628 | Effect of proprioception training and backward walking training among diabetic neuropathy patients. | Inappropriate study design |
| 1629 | Normative data of the wingate anaerobic test in 1 year age groups of Male soccer players. | Inappropriate study design |

| | | |
|---|---|---|
| 1630 | Short-term cardiovascular response to short-radius centrifugation with and without ergometer exercise. | Inappropriate study design |
| 1631 | Changes of hemodynamic and cerebral oxygenation after exercise in normobaric and hypobaric hypoxia: Associations with acute mountain sickness. | Inappropriate population |
| 1632 | Immediate effect of slow right and left nostril deep breathing exercises on reaction time, pain sensitivity, and temperature of elderly population in Madurai. | Inappropriate study design |
| 1633 | Acute dehydration impairs endurance without modulating neuromuscular function. | Inappropriate study design |
| 1634 | Ratings of perceived exertion misclassify intensities for sedentary older adults during graded cycling test: Effect of supramaximal high-intensity interval training. | Inappropriate study design |
| 1635 | A case of coronary venous angioplasty to facilitate left ventricular lead placement. | Inappropriate outcome |
| 1636 | Different heart rate patterns during cardio-pulmonary exercise (CPX) testing in individuals with type 1 diabetes. | Inappropriate intervention |
| 1637 | Feasibility of a combined aerobic and cognitive training intervention on cognitive function in cancer survivors: A pilot investigation. | Inappropriate population |
| 1638 | The effects of oral smokeless tobacco administration on endurance performance. | Inappropriate population |
| 1639 | Effects of weight bearing exercises on bone mineral density of the first-year privates by quantitative ultrasound. | Inappropriate study design |
| 1640 | Exercise Stroke Volume in Adult Cystic Fibrosis: A Comparison of Acetylene Pulmonary Uptake and Oxygen Pulse. | Inappropriate population |
| 1641 | Oxygen consumptions of 30 task-oriented exercises for walking training in stroke. | Inappropriate study design |
| 1642 | Emerging themes in coping with lifetime stress and implication for stress management education. | Inappropriate intervention |
| 1643 | Early childhood trauma, alexithymia, dissociation and the power of the body self. | Inappropriate population |
| 1644 | Exercise for cognitive brain health in aging: A systematic review for an evaluation of dose. | Inappropriate study design |
| 1645 | The Impact of Whole Body Vibration Therapy on Spasticity and Disability of the Patients with Poststroke Hemiplegia. | Inappropriate population |
| 1646 | Thinking-While-Moving Exercises May Improve Cognition in Elderly with Mild Cognitive Deficits: A Proof-of-Principle Study. | Inappropriate study design |
| 1647 | Effects of Rhodiola rosea supplementation on mental performance, physical capacity, and oxidative stress biomarkers in healthy men. | Inappropriate study design |
| 1648 | Effectiveness of a multimodal low-moderate intensity exercise rehabilitation program for stroke survivors. | Inappropriate population |
| 1649 | The relationship among morningness-eveningness, sleep duration, social jetlag, and body mass index in Asian patients with prediabetes. | Inappropriate population |
| 1650 | Essential amino acids (EAA) mixture supplementation: Effects of an acute administration protocol on myoelectric manifestations of fatigue in the biceps brachii after resistance exercise. | Inappropriate study design |
| 1651 | Cerebrovascular reactivity and central arterial stiffness in habitually exercising healthy adults. | Inappropriate study design |
| 1652 | Beta-alanine supplementation improved 10-km running time trial in physically active adults. | Inappropriate outcome |
| 1653 | A comprehensive study through training to prevent dry eye syndrome among mobile phone addicts. | Inappropriate study design |
| 1654 | The influence of religious/spiritual exercises on well-being and quality of life in dermatological patients: A quasi-experimental study. | Inappropriate population |
| 1655 | Sleep duration and physical activity profiles associated with self-reported stroke in the United States: Application of Bayesian belief network modeling techniques. | Inappropriate study design |
| 1656 | Aerobic Exercise Recognition Through Sparse Representation Over Learned Dictionary by Using Wearable Inertial Sensors. | Inappropriate population |
| 1657 | Head-movement-emphasized rehabilitation in bilateral vestibulopathy. | Inappropriate study design |
| 1658 | A bout of high intensity interval training lengthened nerve conduction latency to the non-exercised affected limb in chronic stroke. | Inappropriate study design |
| 1659 | Nutritional aspects of a year-long wheelchair dance intervention in bedridden individuals with severe athetospastic cerebral palsy rated to GMFCS level v. | Inappropriate study design |
| 1660 | Experimental investigation of the time course effects of acute exercise on false episodic memory. | Inappropriate population |
| 1661 | To analyse the effectiveness of yoga, pilates and tai chi exercise for chronic mechanical neck pain -a randomized controlled trial. | Inappropriate study design |
| 1662 | Effect of training-induced changes in Achilles tendon stiffness on muscle-tendon behavior during landing. | Inappropriate population |

| | | |
|---|---|---|
| 1663 | Tailoring pain neuroscience education and exercise programme for people with fibromyalgia who also have cognitive deficits: a case series. | Inappropriate population |
| 1664 | Stress, adherence, and blood pressure control: A baseline examination of Black women with hypertension participating in the SisterTalk II intervention. | Inappropriate study design |
| 1665 | Perceived neighborhood disorder, racial-ethnic discrimination and leading risk factors for chronic disease among women: California Behavioral Risk Factor Surveillance System, 2013. | Inappropriate study design |
| 1666 | High intensity interval training negatively affects mood state in professional athletes. | Inappropriate study design |
| 1667 | Identifying the predisposing factors, signs and symptoms of overreaching and overtraining in physical education professionals. | Inappropriate population |
| 1668 | Comparison of the effectiveness of visual imagery technique and progressive relaxation technique on anxiety and depression in subjects with moderate chronic obstructive pulmonary disease. | Inappropriate study design |
| 1669 | The effects of aerobic exercise on cognitive performance and sleep quality haemodialysis patients. | Inappropriate study design |
| 1670 | Effect of acute augmented feedback on between limb asymmetries and eccentric knee flexor strength during the Nordic hamstring exercise. | Inappropriate population |
| 1671 | Effects of turning and cognitive training in fall prevention with dual task training in elderly with balance impairment. | Inappropriate study design |
| 1672 | Chagasic cardiomyopathy and pompe disease: Case report. | Inappropriate study design |
| 1673 | The prevalence of self-reported sleep problems and some factors affecting it among the elderly in Amirkola. | Inappropriate population |
| 1674 | Validity of multisensor array for measuring energy expenditure of an activity bout in early stroke survivors. | Inappropriate study design |
| 1675 | The effectiveness of transdiagnostic cognitive behavioral therapy for comorbid insomnia: A case report. | Inappropriate study design |
| 1676 | MAP Training My BrainTM: Meditation plus aerobic exercise lessens trauma of sexual violence more than either activity alone. | Inappropriate study design |
| 1677 | The relationship between physical fitness and simulated firefighting task performance. | Inappropriate study design |
| 1678 | Yoga for Persons with HIV-Related Distal Sensory Polyneuropathy: A Case Series. | Inappropriate study design |
| 1679 | Improvements in attention and cardiac autonomic modulation after a 2-weeks sprint interval training program: A fidelity approach. | Inappropriate study design |
| 1680 | Interdisciplinary assessment-oriented treatment of fibromyalgia: a case report. | Inappropriate outcome |
| 1681 | Dysphagia Screening Using High-resolution Impedance Manometry in Acute Stroke. | Inappropriate study design |
| 1682 | Protocol and pilot study of a short message service-guided training after acute stroke/transient ischemic attack to increase walking capacity and physical activity. | Inappropriate study design |
| 1683 | Interacting effects of exercise with breaks in sitting time on cognitive and metabolic function in older adults: Rationale and design of a randomised crossover trial. | Inappropriate study design |
| 1684 | Idiopathic pulmonary fibrosis patient supported with extracorporeal membrane oxygenation for 403 days while waiting for a lung transplant: A case report. | Inappropriate population |
| 1685 | Effects of a multicomponent exercise on dual-task performance and executive function among older adults. | Inappropriate outcome |
| 1686 | Effects of resisted sprint training and traditional power training on sprint, jump, and balance performance in healthy young adults: A randomized controlled trial. | Inappropriate study design |
| 1687 | Effects of a 12-Week Exercise Training Program on Physical Function in Institutionalized Frail Elderly. | Inappropriate study design |
| 1688 | Functional outcomes and quality of life after a 6-month early intervention program for oral cancer survivors: A single-arm clinical trial. | Inappropriate population |
| 1689 | Progressive abduction loading therapy with horizontal-plane viscous resistance targeting weakness and flexion synergy to treat upper limb function in chronic hemiparetic stroke: A randomized clinical trial. | Inappropriate study design |
| 1690 | Pharmacokinetics and scintigraphic imaging of the hypoxia-imaging agent [123I]IAZA in healthy adults following exercise-based cardiac stress. | Inappropriate population |
| 1691 | Moderate intensity physical activity associates with CSF biomarkers in a cohort at risk for Alzheimer's disease. | Inappropriate study design |
| 1692 | Kinect-based individualized upper extremity rehabilitation is effective and feasible for individuals with stroke using a transition from clinic to home protocol. | Inappropriate study design |

| | | |
|---|---|---|
| 1693 | Development and implementation of an inpatient multidisciplinary pain management program for patients with intractable chronic musculoskeletal pain in Japan: Preliminary report. | Inappropriate study design |
| 1694 | The effect of lower-body positive pressure on the cardiorespiratory response at rest and during submaximal running exercise. | Inappropriate study design |
| 1695 | The effects of posterior talar glide and dorsiflexion of the ankle plus mobilization with movement on balance and gait function in patient with chronic stroke: A randomized controlled trial. | Inappropriate study design |
| 1696 | Relationship between aortic compliance and impact of cerebral blood flow fluctuation to dynamic orthostatic challenge in endurance athletes. | Inappropriate population |
| 1697 | Functional strength training and movement performance therapy for upper limb recovery early poststroke-efficacy, Neural correlates, predictive markers, and cost-effectiveness: FAST-INdiCATE trial. | Inappropriate population |
| 1698 | Efficacy of internet-based guided treatment for genito-pelvic pain/penetration disorder: Rationale, treatment protocol, and design of a randomized controlled trial. | Inappropriate study design |
| 1699 | Aerobic and concentration training and allele 7 in the dopamine receptor D4 (D4DR) gene increase chances of smoking cessation in young Polish women. | Inappropriate study design |
| 1700 | Home exercise improves the quality of sleep and daytime sleepiness of elderlies: A randomized controlled trial. | Inappropriate study design |
| 1701 | An acute bout of housework activities has beneficial effects on executive function. | Inappropriate outcome |
| 1702 | An examination of the Counting Talk Test's ability to discern individual variability in exercise-induced metabolic stress. | Inappropriate population |
| 1703 | Combined endurance and resistance training during geriatric day care improve exercise capacity, balance and strength. | Inappropriate study design |
| 1704 | Perception of adults on electronic cigarettes (E-cigarette) in a malaysian tertiary care centre. | Inappropriate study design |
| 1705 | The Influence of Aerobic Exercise on Hippocampal Integrity and Function: Preliminary Findings of a Multi-Modal Imaging Analysis. | Inappropriate study design |
| 1706 | The impact of breaking up prolonged sitting on glucose metabolism and cognitive function when sleep is restricted. | Inappropriate population |
| 1707 | Yoga practitioners uniquely activate the superior parietal lobule and supramarginal gyrus during emotion regulation. | Inappropriate study design |
| 1708 | Acute effects of instructed and self-created variable rope skipping on EEG brain activity and heart rate variability. | Inappropriate study design |
| 1709 | High-intensity exercise mitigates cardiovascular deconditioning during long-duration bed rest. | Inappropriate study design |
| 1710 | Effect of task specific circuit training on gait parameters and mobility in stroke survivors. | Inappropriate study design |
| 1711 | The effects of task-oriented circuit training using rehabilitation tools on the upper-extremity functions and daily activities of patients with acute stroke: A randomized controlled pilot trial. | Inappropriate study design |
| 1712 | Acute exercise facilitates the N450 inhibition marker and P3 attention marker during stroop test in young and older adults. | Inappropriate study design |
| 1713 | Differences in Brain Structure and Function Among Yoga Practitioners and Controls. | Inappropriate study design |
| 1714 | The Impact of Blood Pressure Dipping Status on Cognition, Mobility, and Cardiovascular Health in Older Adults Following an Exercise Program. | Inappropriate study design |
| 1715 | Skipping and running improve short-term memory in young adults. | Inappropriate outcome |
| 1716 | Randomized controlled trial considering varied exercises for reducing proactive memory interference. | Inappropriate study design |
| 1717 | Experimental effects of acute exercise on iconic memory, short-term episodic, and long-term episodic memory. | Inappropriate study design |
| 1718 | Experimental effects of acute exercise and meditation on parameters of cognitive function. | Inappropriate intervention |
| 1719 | A 4-week intervention involving mobile-based daily 6-minute micro-sessions of functional high-intensity circuit training improves strength and quality of life, but not cardio-respiratory fitness of young untrained adults. | Inappropriate study design |
| 1720 | Measuring stroke patients' exercise preferences using a discrete choice experiment. | Inappropriate study design |
| 1721 | Comparative evaluation of mda levels during aerobic exercise in young trained and sedentary male subjects. | Inappropriate population |
| 1722 | Different intensities of basketball drills affect jump shot accuracy of expert and junior players. | Inappropriate study design |

| | | |
|---|---|---|
| 1723 | The effect of short-term outdoor taekwondo training on the concentration and mood of taekwondo players. | Inappropriate study design |
| 1724 | The reliability and minimal detectable change of the cardiovascular response and self-selected exercise intensity during forward and backward treadmill exercise in individuals with Parkinson disease. | Inappropriate study design |
| 1725 | Pilot of a Tailored Dance Intervention to Support Function in People With Cognitive Impairment Residing in Long-Term Care: A Brief Report. | Inappropriate study design |
| 1726 | Leap Motion-based virtual reality training for improving motor functional recovery of upper limbs and neural reorganization in subacute stroke patients. | Inappropriate study design |
| 1727 | Subjective assessment of outcomes of septoplasty. | Inappropriate study design |
| 1728 | Algorithm for Predicting Disease Likelihood From a Submaximal Exercise Test. | Inappropriate study design |
| 1729 | Health behavior and self-assessed health among some long-term unemployed living in Turku, Finland. | Inappropriate study design |
| 1730 | Impact of Mezieres Rehabilitative Method in Patients with Parkinson's Disease: A Randomized Controlled Trial. | Inappropriate population |
| 1731 | Explicating social values for resource allocation decisions on new cancer technologies: We, the jury, find. | Inappropriate study design |
| 1732 | The effects of resistance and endurance training on risk factors of vascular inflammation and atherogenesis in non-athlete men. | Inappropriate study design |
| 1733 | Aerobic-strength exercise improves metabolism and clinical state in Parkinson's disease patients. | Inappropriate study design |
| 1734 | Design and implementation of a combined observational and interventional study: Trends of prevalence, awareness, treatment and control hypertension and the effect of expanded chronic care model on control, treatment and self-care. | Inappropriate study design |
| 1735 | Physical capacity and energy expenditure of cavers. | Inappropriate study design |
| 1736 | Effects of dual-task management and resistance training on gait performance in older individuals: A randomized controlled trial. | Inappropriate study design |
| 1737 | Higher daily physical activity level is associated with lower RBC aggregation in carotid artery disease patients at high risk of stroke. | Inappropriate study design |
| 1738 | Age-related decline of wrist position sense and its relationship to specific physical training. | Inappropriate study design |
| 1739 | High repetitions of Salat (prayer) activity in stroke rehabilitation. | Inappropriate study design |
| 1740 | Treatment response in patients with idiopathic pulmonary arterial hypertension and a severely reduced diffusion capacity. | Inappropriate population |
| 1741 | Effects of weight-bearing exercise on a mini-trampoline on foot mobility, plantar pressure and sensation of diabetic neuropathic feet; a preliminary study. | Inappropriate study design |
| 1742 | The Acute Effects of Aerobic Exercise on the Functional Connectivity of Human Brain Networks. | Inappropriate study design |
| 1743 | Cardiac rehabilitation in patients who underwent primary percutaneous coronary intervention for acute myocardial infarction: Determinants of programme participation and completion. | Inappropriate population |
| 1744 | Intensifying functional task practice to meet aerobic training guidelines in stroke survivors. | Inappropriate study design |
| 1745 | Study protocol of the Intense Physical Activity and Cognition study: The effect of high-intensity exercise training on cognitive function in older adults. | Inappropriate study design |
| 1746 | Functional impact of exercise pulmonary hypertension in patients with borderline resting pulmonary arterial pressure. | Inappropriate study design |
| 1747 | A cross sectional study on complementary and alternative medicine use among a sample of Turkish hospital outpatients with chronic lower back pain. | Inappropriate population |
| 1748 | Effects of Exercise on Sport Concussion Assessment Tool-Third Edition Performance in Professional Athletes. | Inappropriate study design |
| 1749 | The Effects of Glycyrrhiza glabra L. extract use with aerobic training on inflammatory factors and cognitive state in elderly with mild cognitive impairment. | Inappropriate population |
| 1750 | Cardiac rehabilitation in patients with cardiovascular disease leads various hemodynamic parameters obtained using simple non-invasive tests to their appropriate levels. | Inappropriate study design |
| 1751 | Smart watch-based coaching with tiotropium and olodaterol ameliorates physical activity in patients with chronic obstructive pulmonary disease. | Inappropriate study design |
| 1752 | Effect of a short term pulmonary rehabilitation programme on exercise capacity, pulmonary function and health related quality of life in patients with COPD. | Inappropriate study design |
| 1753 | Branched-chain amino acids and arginine improve physical but not skill performance in two consecutive days of exercise. | Inappropriate study design |
| 1754 | Pre-discharge rehabilitation after hip surgery reduces 30-day readmissions in older adults: National Health Insurance Service-Senior Cohort (2007-2012). | Inappropriate population |

| | | |
|---|---|---|
| 1755 | Behavioral Activation for a Breast Cancer Patient with Major Depression and Coexistent Personality Disorder. | Inappropriate study design |
| 1756 | Muscle Ergoreflex Activity and Autonomic Balance Assessed in the Vertical and Horizontal Body Positions in Young Healthy Men. | Inappropriate study design |
| 1757 | Design of the ExCersion-VCI study: The effect of aerobic exercise on cerebral perfusion in patients with vascular cognitive impairment. | Inappropriate study design |
| 1758 | The effects of aerobic exercise with lavender essence use on cognitive state and serum brain-derived neurotrophic factor levels in elderly with mild cognitive impairment. | Inappropriate intervention |
| 1759 | The impact of aerobic exercise on fronto-parietal network connectivity and its relation to mobility: An exploratory analysis of a 6-month randomized controlled trial. | Inappropriate study design |
| 1760 | Outcomes of aerobic exercises on 6-minute walk test in post-stroke patients. | Inappropriate population |
| 1761 | The relationship between sustained attention and aerobic fitness in a group of young adults. | Inappropriate population |
| 1762 | Cortical substrate of supraspinal fatigue following exhaustive aerobic exercise localizes to a large cluster in the anterior premotor cortex. | Inappropriate study design |
| 1763 | Risk of shoulder injuries in sugarcane workers: Analysis based on the simulation of the movements. | Inappropriate study design |
| 1764 | Dose Response to One Week of Supplementation of a Multi-Ingredient Preworkout Supplement Containing Caffeine before Exercise. | Inappropriate population |
| 1765 | Gender, Success, and Drop-Out during a Resistance Exercise Program in Community Dwelling Old Adults. | Inappropriate population |
| 1766 | Physiological responses to firefighting in extreme temperatures do not compare to firefighting in temperate conditions. | Inappropriate study design |
| 1767 | Influence of waterpipe smoking on hematological parameters and cognitive function before and after supramaximal exercise. | Inappropriate study design |
| 1768 | Effect of cerebral vasomotion during physical exercise on associative memory, a near-infrared spectroscopy study. | Inappropriate population |
| 1769 | Pathophysiology of exercise intolerance in chronic diseases: The role of diminished cardiac performance in mitochondrial and heart failure patients. | Inappropriate study design |
| 1770 | The Reliability of a 5km Run Test on a Motorized Treadmill. | Inappropriate population |
| 1771 | Feasibility of a task-oriented training and muscle-strengthening programme to weight-bearing symmetry after stroke. | Inappropriate study design |
| 1772 | Effects of a lifestyle interventions program on quality of life in breast cancer survivors. | Inappropriate study design |
| 1773 | Muscle strength and cognition in ageing men and women: The DR's EXTRA study. | Inappropriate outcome |
| 1774 | Effects of dietary nitrate supplementation on physiological responses, cognitive function, and exercise performance at moderate and very-high simulated altitude. | Inappropriate population |
| 1775 | Strength training decreases inflammation and increases cognition and physical fitness in older women with cognitive impairment. | Inappropriate study design |
| 1776 | Exercise-induced muscle fatigue in the unaffected knee joint and its influence on postural control and lower limb kinematics in stroke patients. | Inappropriate study design |
| 1777 | Benefits of substituting sitting with standing and walking in free-living conditions for cardiometabolic risk markers, cognition and mood in overweight adults. | Inappropriate study design |
| 1778 | Distinctive steady-state heart rate and blood pressure responses to passive robotic leg exercise during head-up tilt: A pilot study in neurological patients. | Inappropriate study design |
| 1779 | A randomized trial of individual versus group-format exercise and self-management in individuals with Parkinson's disease and comorbid depression. | Inappropriate population |
| 1780 | Acute physiological and electrical accentuation of vagal tone has no effect on pain or gastrointestinal motility in chronic pancreatitis. | Inappropriate study design |
| 1781 | Phase-II clinical validation of a powered exoskeleton for the treatment of elbow spasticity. | Inappropriate outcome |
| 1782 | Effect of rehabilitation and prolotherapy on pain and functional performance in patients with chronic patellar tendinopathy. | Inappropriate study design |
| 1783 | Osteopathic manipulative treatment in soccer players with chronic groin pain: A pilot study. | Inappropriate study design |
| 1784 | Low frequency repetitive transcranial magnetic stimulation improves motor dysfunction after cerebral infarction. | Inappropriate study design |
| 1785 | Effect of physical exercise on markers of neuronal dysfunction in cerebrospinal fluid in patients with Alzheimer's disease. | Inappropriate population |
| 1786 | Influence of chair-based yoga on salivary anti-microbial proteins, functional fitness, perceived stress and well-being in older women: A randomized pilot controlled trial. | Inappropriate population |

| | | |
|---|---|---|
| 1787 | Effects of multicomponent exercise on functional and cognitive parameters of hypertensive patients: A quasi-experimental study. | Inappropriate study design |
| 1788 | The impact of indoor cycling training on hemodynamic parameters of the left ventricle of men with ischaemic heart disease or after myocardial infarction. | Inappropriate study design |
| 1789 | No influence of transcutaneous electrical nerve stimulation on exercise-induced pain and 5-Km cycling time-trial performance. | Inappropriate study design |
| 1790 | Fit to play: posture and seating position analysis with professional musicians - a study protocol. | Inappropriate study design |
| 1791 | Efficacy of Wii-Fit on Static and Dynamic Balance in Community Dwelling Older Veterans: A Randomized Controlled Pilot Trial. | Inappropriate study design |
| 1792 | Development and preliminary validation of the focused analgesia selection test to identify accurate pain reporters. | Inappropriate study design |
| 1793 | Acute effects of exercise on attentional bias in low and high anxious young adults. | Inappropriate study design |
| 1794 | The role of physical activity enjoyment on the acute mood experience of exercise among smokers with elevated depressive symptoms. | Inappropriate study design |
| 1795 | Exhaustive exercise alters thinking times in a tower of london task in a time-dependent manner. | Inappropriate population |
| 1796 | Relationship between body composition and physical fitness of rescue firefighter personnel in Selangor, Malaysia. | Inappropriate study design |
| 1797 | Effects of weight training time on bone mineral density of patients with secondary osteoporosis after hemiplegia. | Inappropriate study design |
| 1798 | Influence of exercise on visceral pain: An explorative study in healthy volunteers. | Inappropriate study design |
| 1799 | Associations of objectively measured moderate-to-vigorous-intensity physical activity and sedentary time with all-cause mortality in a population of adults at high risk of type 2 diabetes mellitus. | Inappropriate study design |
| 1800 | Quantifying mental arousal levels in daily living using additional heart rate. | Inappropriate population |
| 1801 | Cancer patients participating in a lifestyle intervention during chemotherapy greatly over-report their physical activity level: A validation study. | Inappropriate population |
| 1802 | A transient awakening of a patient with Alzheimer's disease that questions our practice. | Inappropriate population |
| 1803 | The Relationship Between Physical Activity and Depressive Symptoms in Healthy Older Women. | Inappropriate study design |
| 1804 | Effect of zumba dance on blood pressure. | Inappropriate study design |
| 1805 | Habitual and Low-Intensity Physical Activity in People with Multiple Sclerosis. | Inappropriate study design |
| 1806 | Ambulatory Activity in People with Early Parkinson's Disease. | Inappropriate study design |
| 1807 | Effect of different loading conditions on leg stiffness during hopping at different frequencies. | Inappropriate population |
| 1808 | Efficacy of external counterpulsation enhanced with outpatient cardiac rehabilitation. | Inappropriate study design |
| 1809 | Validity evidence for the state mindfulness scale for physical activity. | Inappropriate population |
| 1810 | Short-and long-term training effects on motor and functional performances of community-dwelling individuals with chronic stroke. | Inappropriate population |
| 1811 | Simulation of lifting motions using a novel multi-objective optimization approach. | Inappropriate study design |
| 1812 | Effect of pulmonary rehabilitation program with meditation on perceived self-efficacy, pulmonary rehabilitation behavior, exercise tolerance, and dyspnea in patients with chronic obstructive pulmonary disease. | Inappropriate population |
| 1813 | Chronic disease, medications and lifestyle: Perceptions from a regional Victorian Aboriginal community. | Inappropriate population |
| 1814 | Yoga-based pulmonary rehabilitation for the management of dyspnea in coal miners with chronic obstructive pulmonary disease: A randomized controlled trial. | Inappropriate population |
| 1815 | Exercise strengthens central nervous system modulation of pain in fibromyalgia. | Inappropriate intervention |
| 1816 | Suicidal ideation and suicide attempts among asthma. | Inappropriate study design |
| 1817 | Echocardiographic evaluation of the effects of high-intensity interval training on cardiac morphology and function. | Inappropriate study design |
| 1818 | Early-onset dementia in type 1 diabetes mellitus. | Inappropriate study design |
| 1819 | Effects of a dietary beetroot juice treatment on systemic and cerebral haemodynamics-A pilot study. | Inappropriate study design |
| 1820 | The effect of ginseng supplement on heart rate, systolic and diastolic blood pressure to resistance training in trained males. | Inappropriate population |

| | | |
|---|---|---|
| 1821 | Use of new guidance to profile 'equivalent minutes' of aerobic physical activity for adults in England reveals gender, geographical, and socio-economic inequalities in meeting public health guidance: A cross-sectional study. | Inappropriate study design |
| 1822 | Comparison of heart rate reserve-guided and ratings of perceived exertion-guided methods for high-intensity robot-assisted gait training in patients with chronic stroke focused on the motor function and gait ability. | Inappropriate study design |
| 1823 | Return to Elite Level of Play and Performance in Professional Golfers After Arthroscopic Hip Surgery. | Inappropriate study design |
| 1824 | Transcranial direct current stimulation combined with aerobic exercise to optimize analgesic responses in fibromyalgia: A randomized placebo-controlled clinical trial. | Inappropriate study design |
| 1825 | Dipyridamole in heart failure due to dilated cardiomyopathy: A pilot study. | Inappropriate study design |
| 1826 | Neuroscience-informed auditory training in schizophrenia: A final report of the effects on cognition and serum brain-derived neurotrophic factor. | Inappropriate study design |
| 1827 | Effect of PNF technique for knee muscles on lower limb performance in subacute stroke - An experimental study. | Inappropriate population |
| 1828 | Resistance Training, Lipid Profile, and Homocysteine in Patients with Alzheimer's Disease. | Inappropriate study design |
| 1829 | Case report on integrative cardiology and acupuncture treatment for hypertension and its cofactors such as daily, multiple, long lasting vertex and occipital migraines/headaches. | Inappropriate population |
| 1830 | Barriers, motivations, and preferences for physical activity among female African American older adults. | Inappropriate study design |
| 1831 | Short-term pulmonary rehabilitation for a female patient with chronic scleroderma under a single-case research design. | Inappropriate population |
| 1832 | Comparing the effectiveness of karate and fitness training on cognitive functioning in older adults-A randomized controlled trial. | Inappropriate population |
| 1833 | Examining the relationship between sport and health among USA women: An analysis of the Behavioral Risk Factor Surveillance System. | Inappropriate study design |
| 1834 | Closed-loop task difficulty adaptation during virtual reality reach-to-grasp training assisted with an exoskeleton for stroke rehabilitation. | Inappropriate study design |
| 1835 | Pacing profiles in competitive track races: Regulation of exercise intensity is related to cognitive ability. | Inappropriate outcome |
| 1836 | Endurance performance during severe-intensity intermittent cycling: Effect of exercise duration and recovery type. | Inappropriate study design |
| 1837 | Physical Fitness in Older People Recently Diagnosed with Cognitive Impairment Compared to Older People Recently Discharged from Hospital. | Inappropriate population |
| 1838 | Exploring the mechanisms of exercise-induced hypoalgesia using somatosensory and laser evoked potentials. | Inappropriate study design |
| 1839 | An individually tailored behavioral medicine treatment in physical therapy for tension-type headache - Two experimental case studies. | Inappropriate outcome |
| 1840 | Differences in resting state functional connectivity between young adult endurance athletes and healthy controls. | Inappropriate study design |
| 1841 | Aerobic Exercise Preserves Olfaction Function in Individuals with Parkinson's Disease. | Inappropriate study design |
| 1842 | Does Deep Cervical Flexor Muscle Training Affect Pain Pressure Thresholds of Myofascial Trigger Points in Patients with Chronic Neck Pain? A Prospective Randomized Controlled Trial. | Inappropriate study design |
| 1843 | Adaptations of prefrontal brain activity, executive functions, and gait in healthy elderly following exergame and balance training: A randomized-controlled study. | Inappropriate study design |
| 1844 | The effect of high and low exercise intensity periods on a simple memory recognition test. | Inappropriate study design |
| 1845 | Differential effects of assisted cycling therapy on short-term and working memory of adolescents with Down syndrome. | Inappropriate population |
| 1846 | Differential training facilitates early consolidation in motor learning. | Inappropriate population |
| 1847 | The effect of a horse riding simulator on energy expenditure, enjoyment, and task difficulty in the elderly. | Inappropriate study design |
| 1848 | Effectiveness of transcutaneous eletrical neuromuscular stimulation along with exercisces manoeuver in Dysphagia: Case series. | Inappropriate study design |
| 1849 | Cognitive fatigue influences time-on-task during bodyweight resistance training exercise. | Inappropriate study design |
| 1850 | Exercise portrayal in children's television programs: Analysis of the UK and Irish programming. | Inappropriate study design |
| 1851 | Awareness and prevalence of metabolic syndrome among high-risk individuals attending internal medicine clinics across Jordan. | Inappropriate population |

| | | |
|---|---|---|
| 1852 | Hybrid neuroprosthesis for the upper limb: Combining brain-controlled neuromuscular stimulation with a multi-joint arm exoskeleton. | Inappropriate study design |
| 1853 | Physical Exercise with Multicomponent Cognitive Intervention for Older Adults with Alzheimer's Disease: A 6-Month Randomized Controlled Trial. | Inappropriate study design |
| 1854 | Psychoeducation of adolescents with inflammatory bowel diseases and their families. | Inappropriate study design |
| 1855 | The psychometric properties of Chinese version of SCI Exercise Self-Efficacy Scale in patients with stroke. | Inappropriate study design |
| 1856 | Effect of aerobic exercises on stuttering. | Inappropriate study design |
| 1857 | Effect of aerobics exercise and dietary habits on blood pressure in hypertensives. | Inappropriate study design |
| 1858 | Lower cardiorespiratory fitness is associated with greater reduction in PTSD symptoms and anxiety sensitivity following aerobic exercise. | Inappropriate study design |
| 1859 | The Effects of POWER Training in Young and Older Adults after Stroke. | Inappropriate study design |
| 1860 | Focusing on Increasing Velocity during Heavy Resistance Knee Flexion Exercise Boosts Hamstring Muscle Activity in Chronic Stroke Patients. | Inappropriate study design |
| 1861 | The effect of brain-derived neurotrophic factor (BDNF) val66met polymorphism on the learning of complex motor skill. | Inappropriate population |
| 1862 | Achalasia: A case report on its effect during surgical decision making for laparoscopic sleeve gastrectomy in the young morbidly obese patient. | Inappropriate study design |
| 1863 | The upbeat association sandwiched between physical exercise and stress-free academic performance amid medical students of Universiti Sultan Zainal Abidin. | Inappropriate population |
| 1864 | Predictors of Adherence to a Falls Prevention Exercise Program for People with Parkinson's Disease. | Inappropriate outcome |
| 1865 | Re-training of type 2 diabetic patients for better adherence to diabetes care plan in oral anti-diabetics and plus insulin treatment groups. | Inappropriate study design |
| 1866 | Novel Motor-Assisted Elliptical Training Intervention Improves 6-Minute Walk Test and Oxygen Cost for an Individual with Progressive Supranuclear Palsy. | Inappropriate study design |
| 1867 | Reducing workplace burnout: The relative benefits of cardiovascular and resistance exercise. | Inappropriate study design |
| 1868 | Related factors and prevalence Of Parkinson's disease among Uygur residents in Hetian, Xinjiang Uygur autonomous region. | Inappropriate study design |
| 1869 | Failure to identify an acute exercise effect on executive function assessed by the wisconsin card sorting test. | Inappropriate outcome |
| 1870 | Satisfaction with chronic pain treatment. | Inappropriate population |
| 1871 | Study of knowledge, attitude, and practice of general population of guntur toward silent killer diseases: Hypertension and diabetes. | Inappropriate study design |
| 1872 | Effects of pioglitazone or exercise in older adults with mild cognitive impairment and insulin resistance: A pilot study. | Inappropriate population |
| 1873 | Aerobic physical exercise improved the cognitive function of elderly males but did not modify their blood homocysteine levels. | Inappropriate outcome |
| 1874 | Acute and Chronic Effect of Acoustic and Visual Cues on Gait Training in Parkinson's Disease: A Randomized, Controlled Study. | Inappropriate study design |
| 1875 | The rehabilitation of chronic neurological patients: An alternative answer to a growing demand. | Inappropriate study design |
| 1876 | Effects of aerobic exercise on physiological exercise intensity and perception of exercise intensity in young men. | Inappropriate study design |
| 1877 | Measuring physical activity in older adults: Calibrating cut-points for the MotionWatch 8. ©. | Inappropriate study design |
| 1878 | Effects of the bilateral isokinetic strengthening training on functional parameters, gait, and the quality of life in patients with stroke. | Inappropriate population |
| 1879 | Therapeutic efficacy of intensified walk training under the electrocardiogram telemetry in stroke induced lower limb dysfunction patients with heart failure. | Inappropriate population |
| 1880 | Weight and blood pressure reduction among participants engaged in a cancer awareness and prevention program. | Inappropriate study design |
| 1881 | Dyspnea: When the preliminary imaging is unconvincing. | Inappropriate study design |
| 1882 | Effects of moderate exercise on relieving mental load of elementary school teachers. | Inappropriate study design |
| 1883 | Mental fatigue induced by prolonged self-regulation does not exacerbate central fatigue during subsequent whole-body endurance exercise. | Inappropriate population |
| 1884 | Acute short-term dim light exposure can lower muscle strength endurance. | Inappropriate study design |
| 1885 | Sex differences in neuroanatomy of the human mirror neuron system: Impact on functional recovery of ischemic hemiparetic patients. | Inappropriate study design |
| 1886 | Exercise-mode-related changes in task-switching performance in the elderly. | Inappropriate population |

| | | |
|---|---|---|
| 1887 | High-intensity, unilateral resistance training of a non-paretic muscle group increases active range of motion in a severely paretic upper extremity muscle group after stroke. | Inappropriate population |
| 1888 | Self-reported physical activity and objective aerobic fitness: Differential associations with gray matter density in healthy aging. | Inappropriate study design |
| 1889 | Simulated firefighting task performance and physiology under very hot conditions. | Inappropriate population |
| 1890 | Task-oriented training and lower limb strengthening to improve balance and function after stroke: A pilot study. | Inappropriate population |
| 1891 | Effect of Acute Caffeine Supplementation on Anaerobic Power and Blood Lactate Levels in Female Athletes. | Inappropriate study design |
| 1892 | Evaluation of short-term use of nocturnal nasal continuous positive airway pressure for a clinical profile and exercise capacity in adult patients with obstructive sleep apnea-hypopnea syndrome. | Inappropriate study design |
| 1893 | "Like walking into an empty room": Effects of eurythmy therapy on stress perception in comparison with a sports intervention from the subjects' perspective - A qualitative study. | Inappropriate study design |
| 1894 | The effect of physical activity on adiponectin and osteocalcin in overweight young females. | Inappropriate study design |
| 1895 | Perceived personal importance of exercise and fears of re-injury: A longitudinal study of psychological factors related to activity after anterior cruciate ligament reconstruction. | Inappropriate study design |
| 1896 | Promoting an obesity education program among minority patients in a single urban pediatric Emergency Department (ED). | Inappropriate study design |
| 1897 | Qualitative evaluation of baduanjin (Traditional Chinese Qigong) on health promotion among an elderly community population at risk for ischemic stroke. | Inappropriate population |
| 1898 | Validation of perceptual strain index to evaluate the thermal strain in experimental hot conditions. | Inappropriate population |
| 1899 | Multidimensional unconscious structural changes in Davanloo's intensive short-term dynamic psychotherapy. Part I. | Inappropriate population |
| 1900 | Fruit and vegetable consumption and its determinants among Saudi university students. | Inappropriate outcome |
| 1901 | Effects of Functional Limb Overloading on Symmetrical Weight Bearing, Walking Speed, Perceived Mobility, and Community Participation among Patients with Chronic Stroke. | Inappropriate population |
| 1902 | Are the Measurements of Attention Allocation and Perceived Exertion Trustworthy?. | Inappropriate population |
| 1903 | Eagle's syndrome - A non-perceived differential diagnosis of temporomandibular disorder. | Inappropriate study design |
| 1904 | Associations between lower extremity muscle mass and metabolic parameters related to obesity in Japanese obese patients with type 2 diabetes. | Inappropriate study design |
| 1905 | Cross-Country differences in the additive effects of socioeconomics, health behaviors and medical comorbidities on disability among older adults with heart disease. | Inappropriate population |
| 1906 | Effect of total sleep deprivation on egocentric distance estimation following a fatiguing task. | Inappropriate outcome |
| 1907 | The determinants of choosing traditional Korean medicine or conventional medicine: Findings from the Korea health panel. | Inappropriate study design |
| 1908 | A rare case of anomalous venous drainage of the right upper lobe. | Inappropriate intervention |
| 1909 | Exercise training before surgical revascularisation-influence on short term outcomes. | Inappropriate population |
| 1910 | Bimanual elbow robotic orthoses: Preliminary investigations on an impairment force-feedback rehabilitation method. | Inappropriate study design |
| 1911 | A single-session preliminary evaluation of an affordable BCI-controlled arm exoskeleton and motor-proprioception platform. | Inappropriate study design |
| 1912 | An investigation into the effects of different types of exercise on the maintenance of approach motivation levels. | Inappropriate study design |
| 1913 | Associations of EDNRA and EDN1 polymorphisms with carotid intima media thickness through interactions with gender, regular exercise, and obesity in subjects in Taiwan: Taichung Community Health Study (TCHS). | Inappropriate population |
| 1914 | Speaking on behalf of the body and activity: Investigating the activity experiences of Canadian women living with anorexia nervosa. | Inappropriate population |
| 1915 | The importance of community consultation and social support in adhering to an obesity reduction program: Results from the Healthy Weights Initiative. | Inappropriate intervention |
| 1916 | A study of cardiovascular autonomic response among young adults before and after isometric exercise. | Inappropriate study design |

| | | |
|---|---|---|
| 1917 | Glucose but not insulin or insulin resistance is associated with memory performance in middle-aged non-diabetic women: A cross sectional study. | Inappropriate population |
| 1918 | TNF-alpha and TNFR1 responses to recovery therapies following acute resistance exercise. | Inappropriate outcome |
| 1919 | Identifying chronic conditions and other selected factors that motivate physical activity in world senior games participants and the general population. | Inappropriate study design |
| 1920 | Evaluation of blood prothrombotic parameters under the influence of physical activity. | Inappropriate population |
| 1921 | Efficacy and safety of single Botulinum toxin Type A (Botox) injection for relief of upper trapezius myofascial trigger point: A randomized, double-blind, placebo-controlled study. | Inappropriate study design |
| 1922 | The influence of physical therapy on oropharyngeal dysphagia in acute stroke patients. | Inappropriate study design |
| 1923 | Feasibility of an exercise program for older depressive inpatients: A pilot study. | Inappropriate study design |
| 1924 | The impact of a videogame-based pilot physical activity program in older adults with schizophrenia on subjectively and objectively measured physical activity. | Inappropriate study design |
| 1925 | Acupuncture for the treatment of hyperhidrosis: A case report. | Inappropriate study design |
| 1926 | Influence of kinesitherapy on gait in patients with ischemic stroke in the chronic period. | Inappropriate population |
| 1927 | Continuous cardiopulmonary resuscitation training compared to single training by laypersons. | Inappropriate study design |
| 1928 | Role of ultrasound in determining the severity of hallux valgus: Ultrasonic characteristics of abductor hallucis muscle. | Inappropriate population |
| 1929 | Social-ecological, motivational and volitional factors for initiating and maintaining physical activity in the context of HIV. | Inappropriate study design |
| 1930 | Effects of Tyrosine on Parkinson's Disease: A Randomized, Double-Blind, Placebo-Controlled Trial. | Inappropriate study design |
| 1931 | Parameters associated with elevated systolic blood pressure levels in greek military pilots. | Inappropriate study design |
| 1932 | Strength training with stretch-shortening cycle exercises optimizes neuromuscular economy during functional tasks in elderly women. | Inappropriate population |
| 1933 | 5 Hz repetitive transcranial magnetic stimulation over the ipsilesional sensory cortex enhances motor learning after stroke. | Inappropriate study design |
| 1934 | The role of sense of effort on self-selected cycling power output. | Inappropriate study design |
| 1935 | A rehabilitation program for lung cancer patients during postthoracotomy chemotherapy. | Inappropriate population |
| 1936 | Physical functioning after occupational rehabilitation and returning to work among employees with chronic musculoskeletal pain and comorbid depressive symptoms. | Inappropriate population |
| 1937 | A comparison between a Tai Chi program and a usual medical care program in chronic cardiovascular disease participants in quality of life, psychological health, resilience, blood pressure and body mass index. | Inappropriate study design |
| 1938 | Electroacupuncture and transcranial direct current stimulation: Simultaneous treatment for patients with chronic poststroke symptoms. | Inappropriate study design |
| 1939 | The acute effects of mental arithmetic, cold pressor and maximal voluntary contraction on arterial stiffness in young healthy subjects. | Inappropriate population |
| 1940 | PainVision apparatus is effective for assessing low back pain after fusion surgery. | Inappropriate population |
| 1941 | Changes in brain activation in stroke patients after mental practice and physical exercise: A functional MRI study. | Inappropriate study design |
| 1942 | Warm-up durations and time-of-day impacts on rate of perceived exertion after short-term maximal performance. | Inappropriate study design |
| 1943 | Effects of periodic task-specific test feedback on physical performance in older adults undertaking band-based resistance exercise. | Inappropriate population |
| 1944 | Bacterial prints in human infectious diseases. | Inappropriate study design |
| 1945 | Self management and factors associated with the impact of insomnia among older adults with chronic medical illnesses at outpatient clinic. | Inappropriate study design |
| 1946 | Is platelet-rich plasma superior to whole blood in the management of chronic tennis elbow: One year randomized clinical trial. | Inappropriate outcome |
| 1947 | "Es como uno bomba de tiempo [It's like a time bomb]": A qualitative analysis of perceptions of diabetes among first-degree relatives of latino patients with diabetes. | Inappropriate population |
| 1948 | Effects of chronic exercise on severity, quality of life and functionality in an elderly Parkinson's disease patient: Case report. | Inappropriate study design |

| | | |
|---|---|---|
| 1949 | Awareness in patients with COPD about the disease and pulmonary rehabilitation: A survey. | Inappropriate study design |
| 1950 | Effect of high temperature yoga exercise on improving physical and mental well-being of overweight middle-aged and young women. | Inappropriate study design |
| 1951 | Bilateral repetitive transcranial magnetic stimulation combined with intensive swallowing rehabilitation for chronic stroke dysphagia: A case series study. | Inappropriate study design |
| 1952 | Inhaled iloprost for the treatment of patient with Fontan circulation. | Inappropriate study design |
| 1953 | Cardiovascular effects of hemoglobin response in patients receiving epoetin alfa and oral iron in heart failure with a preserved ejection fraction. | Inappropriate population |
| 1954 | Effect of a health coaching self-management program for older adults with multimorbidity in nursing homes. | Inappropriate study design |
| 1955 | Chronic exercise keeps working memory and inhibitory capacities fit. | Inappropriate study design |
| 1956 | Physical activity of patients with chronic pain receiving acceptance and commitment therapy or cognitive behavioural therapy. | Inappropriate study design |
| 1957 | Study on therapeutic outcome, attitude and awareness about type-2 diabetes among type-2 diabetic female patients in aseer diabetic center. | Inappropriate study design |
| 1958 | Sport specificity background affects the principal component structure ofvertical squat jump performance of young adult female athletes. | Inappropriate population |
| 1959 | Should art be integrated into cognitive behavioral therapy for anxiety disorders?. | Inappropriate study design |
| 1960 | P-wave dispersion and its relationship to aortic stiffness in patients with acute myocardial infarction after cardiac rehabilitation. | Inappropriate study design |
| 1961 | Treatment of Concussion in High School Athletes: A Proposed Protocol for Athletic and Academic Return to Activity. | Inappropriate study design |
| 1962 | Non-conscious visual cues related to affect and action alter perception of effort and endurance performance. | Inappropriate population |
| 1963 | PARIS: A good start for exercise in HFPEF. | Inappropriate population |
| 1964 | Treadmill training improves overground walking economy in Parkinson's disease: A randomized, controlled pilot study. | Inappropriate study design |
| 1965 | Long-term glutamine supplementation in elite gymnasts. | Inappropriate study design |
| 1966 | Reliability of 1RM test in detrained men with previous resistance training experience. | Inappropriate study design |
| 1967 | Effects of aquatic physiotherapy on the improvement of balance and corporal symmetry in stroke. | Inappropriate study design |
| 1968 | Effects of using the Nintendo Wii Fit Plus Platform in the sensorimotor training of gait disorders in Parkinson's disease. | Inappropriate population |
| 1969 | Influence of affective changes on behavioral and cognitive performances after acute bout of exhaustive exercise. | Inappropriate population |
| 1970 | Enhancing muscular performance in women: Compound versus complex, traditional resistance and plyometric training alone. | Inappropriate population |
| 1971 | The effects of caffeine on rugby passing accuracy while performing the Reactive Agility Test. | Inappropriate study design |
| 1972 | Experience with exercise right heart catheterization in the diagnosis of pulmonary hypertension: A retrospective study. | Inappropriate study design |
| 1973 | Effect of short-term hyperoxia on model exercise in basketball. | Inappropriate population |
| 1974 | Yoga effects on physical activity and sexual satisfaction among the Iranian women with multiple sclerosis: A randomized controlled trial. | Inappropriate study design |
| 1975 | Regular exercise is associated with emotional resilience to acute stress in healthy adults. | Inappropriate population |
| 1976 | Relationship between exercise-induced heart rate increase and the formation of microbubbles and high-intensity transient signals in mechanical heart valve implanted patients. | Inappropriate outcome |
| 1977 | VR mobile solutions for chronic stress reduction in young adults. | Inappropriate study design |
| 1978 | Low frequency repetitive transcranial magnetic stimulation to improve motor function and grip force of upper limbs of patients with hemiplegia. | Inappropriate population |
| 1979 | Cerebral responses to innocuous somatic pressure stimulation following aerobic exercise rehabilitation in chronic pain patients: A functional magnetic resonance imaging study. | Inappropriate study design |
| 1980 | Physical activity habits and preferences in the month prior to a first-ever stroke. | Inappropriate population |
| 1981 | Making use of equity sensitive QALYs: A case study on identifying the worse off across diseases. | Inappropriate population |
| 1982 | Effects of an exercise program on physiological functions in postmenopausal women with metabolic syndrome. | Inappropriate outcome |

| | | |
|---|---|---|
| 1983 | Time to exhaustion at and above critical power in trained cyclists: The relationship between heavy and severe intensity domains. | Inappropriate intervention |
| 1984 | Stress-induced immune-related diseases and health outcomes of pharmacy students: A pilot study. | Inappropriate population |
| 1985 | Use of self-care and practitioner-based forms of complementary and alternative medicine before and after a diagnosis of breast cancer. | Inappropriate study design |
| 1986 | Effects of qigong exercise on fatigue, anxiety, and depressive symptoms of patients with chronic fatigue syndrome-like illness: A randomized controlled trial. | Inappropriate study design |
| 1987 | Effects of resistance training on cognitive functions in the elderly. | Inappropriate study design |
| 1988 | Altered muscle coordination when pedaling with independent cranks. | Inappropriate population |
| 1989 | Effects of aerobic fitness on aging-related changes of interhemispheric inhibition and motor performance. | Inappropriate population |
| 1990 | Overall reductions in functional brain activation are associated with falls in older adults: An fMRI study. | Inappropriate study design |
| 1991 | Effect of fasting ramadan in diabetes control status-application of extensive diabetes education, serum creatinine with hba1c statistical anova and regression models to prevent hypoglycemia. | Inappropriate population |
| 1992 | Mediators of yoga and stretching for chronic low back pain. | Inappropriate study design |
| 1993 | Effect of diet and exercise on blood glucose level in non insulin dependent diabetics. | Inappropriate study design |
| 1994 | Randomized controlled trial of mindfulness meditation and exercise for the prevention of acute respiratory infection: Possible mechanisms of action. | Inappropriate population |
| 1995 | Exploring outcome measures for exercise intervention in people with Parkinson's disease. | Inappropriate study design |
| 1996 | Effects of time of day on resistance exercise-induced anabolic signaling in skeletal muscle. | Inappropriate study design |
| 1997 | Case of acquired idiopathic cold-induced hyperhidrosis. | Inappropriate study design |
| 1998 | Educating patients about hypoglycemia prevention and self-management. | Inappropriate study design |
| 1999 | Self-directed mindfulness training and improvement in blood pressure, migraine frequency, and quality of life. | Inappropriate population |
| 2000 | Syncope during resistance exercise in an individual with type 1 diabetes. | Inappropriate study design |
| 2001 | Effects of ovarian cycle on hemodynamic responses during dynamic exercise in sedentary women. | Inappropriate population |
| 2002 | The acute effects of a single bout of moderate-intensity aerobic exercise on cognitive functions in healthy adult males. | Inappropriate study design |
| 2003 | Physical activity improves verbal and spatial memory in older adults with probable mild cognitive impairment: A 6-month randomized controlled trial. | Inappropriate study design |
| 2004 | Association between smoking, exercise habits and dental checkups among community residents in Japan. | Inappropriate study design |
| 2005 | Physical function and self-efficacy-Important aspects of health-related quality of life in individuals with hip osteoarthritis. | Inappropriate population |
| 2006 | Clarifying the link between distress intolerance and exercise: Elevated anxiety sensitivity predicts less vigorous exercise. | Inappropriate study design |
| 2007 | Insight for learning and stability of one repetition maximum test in subjects with or without experience on resistance training. | Inappropriate study design |
| 2008 | The role of 6-week hydrotherapy and land-based therapy plus ankle taping in a preseason rehabilitation program for athletes with chronic ankle instability. | Inappropriate study design |
| 2009 | Improvement of processing speed in executive function immediately following an increase in cardiovascular activity. | Inappropriate outcome |
| 2010 | A pulmonary rehabilitation program reduces levels of anxiety and depression in COPD patients. | Inappropriate population |
| 2011 | Multidisciplinary stroke rehabilitation delivered by a humanoid robot: Interaction between speech and physical therapies. | Inappropriate population |
| 2012 | Acid-base balance at lactate threshold in resistance exercise. | Inappropriate intervention |
| 2013 | Task-oriented physical exercise using postural re-alignment with body weight support in chronic stroke. | Inappropriate study design |
| 2014 | The impact of physical exercise on convergent and divergent thinking. | Inappropriate study design |
| 2015 | Does combined cognitive training and physical activity training enhance cognitive abilities more than either alone? A four-condition randomized controlled trial among healthy older adults. | Inappropriate study design |
| 2016 | Taekwondo training improves balance in volunteers over 40. | Inappropriate population |
| 2017 | The influence of systematic pulse-limited physical exercise on the parameters of the cardiovascular system in patients over 65 years of age. | Inappropriate study design |

| | | |
|---|---|---|
| 2018 | Exercise improves cognitive control: Evidence from the stop signal task. | Inappropriate study design |
| 2019 | Use of noninvasive gas exchange to track pulmonary vascular responses to exercise in heart failure. | Inappropriate population |
| 2020 | The effects of acclimatization on blood clotting parameters in exertional heat stress. | Inappropriate study design |
| 2021 | Adult hippocampal neurogenesis reduces memory interference in humans: Opposing effects of aerobic exercise and depression. | Inappropriate study design |
| 2022 | Responsiveness of measurements of lower-limb muscle strength obtained with a hand-held dynamometer from patients with stroke. | Inappropriate study design |
| 2023 | Cardiovascular responses associated with daily walking in subacute stroke. | Inappropriate intervention |
| 2024 | Assessment of cardiorespiratory fitness using submaximal protocol in older adults with mood disorder and Parkinson's disease. | Inappropriate study design |
| 2025 | Virtual Reality Versus Conventional Treatment of Reaching Ability in Chronic Stroke: Clinical Feasibility Study. | Inappropriate study design |
| 2026 | Do cognitive reappraisal and diaphragmatic breathing augment interoceptive exposure for anxiety sensitivity?. | Inappropriate study design |
| 2027 | Intensity of recreational physical activity in different life periods in relation to breast cancer among women in the region of Western Pomerania. | Inappropriate population |
| 2028 | Cognitive impairment and the associated risk factors among the elderly in the Shanghai urban area: A pilot study from China. | Inappropriate study design |
| 2029 | Reliability in one-repetition maximum performance in people with Parkinson's disease. | Inappropriate study design |
| 2030 | Randomized cont rolled trial of mindfulness-based stress reduction versus aerobic exercise: Effects on the self-referential brain network in social anxiety disorder. | Inappropriate study design |
| 2031 | Effects of a cardiac rehabilitation program on systolic function and left ventricular mass in patients after myocardial infarction and revascularization. | Inappropriate population |
| 2032 | Recreational activities and body exercise among secondary school students in Kwara state, Nigeria. | Inappropriate study design |
| 2033 | Barriers are not the limiting factor to participation in physical activity in canadian seniors. | Inappropriate population |
| 2034 | Physiological and mechanical exercise intensity determination based on perceived exertion. | Inappropriate population |
| 2035 | Effects of stretching and strength exercises on speed and power abilities in male basketball players. | Inappropriate study design |
| 2036 | Hand robotics rehabilitation: Feasibility and preliminary results of a robotic treatment in patients with hemiparesis. | Inappropriate study design |
| 2037 | The effect of three months of aerobic training on stroop performance in older adults. | Inappropriate study design |
| 2038 | Feasibility, safety, and compliance in a randomized controlled trial of physical therapy for parkinson's disease. | Inappropriate study design |
| 2039 | The effect of caffeine on performance in collegiate tennis players. | Inappropriate study design |
| 2040 | Aerobic exercises improve blood flow and cognitive functions in anterior circulation ischemic strokes. | Inappropriate study design |
| 2041 | The association between vascular factors and subjective memory impairment in older people: The HUNT study, Norway. | Inappropriate study design |
| 2042 | Training-induced improvement of response selection and error detection in aging assessed by task switching: Effects of cognitive, physical, and relaxation training. | Inappropriate population |
| 2043 | The influence of resistance exercise training on the levels of anxiety in ischemic stroke. | Inappropriate population |
| 2044 | Functional exercise and physical fitness post stroke: The importance of exercise maintenance for motor control and physical fitness after stroke. | Inappropriate study design |
| 2045 | The effects of chronic aerobic and anaerobic exercise on blood nitric oxide levels. | Inappropriate study design |
| 2046 | Rheumatoid arthritis: An integrated acupuncture and energetic approach. | Inappropriate study design |
| 2047 | Effect of time-of-day of aerobic maximal exercise on the sleep quality of trained subjects. | Inappropriate population |
| 2048 | Weighing the pros and cons in family-based pediatric obesity intervention: Parent and child decisional balance as a predictor of child outcomes. | Inappropriate study design |
| 2049 | Torque-angle relationship are better preserved during eccentric compared to concentric contractions in patients with stroke. | Inappropriate study design |
| 2050 | Oxygen consumption and peak heart rate in stroke patients during the completion of the Modified Rivermead Mobility Index (MRMI). | Inappropriate study design |
| 2051 | Exercise preferences are different after stroke. | Inappropriate population |

| | | |
|---|---|---|
| 2052 | Aerobic exercise training increases cerebral blood flow in postmenopausal women. | Inappropriate population |
| 2053 | People getting a grip on arthritis: A knowledge transfer strategy to empower patients with rheumatoid arthritis and osteoarthritis. | Inappropriate study design |
| 2054 | Genetic influences on physiological and subjective responses to an aerobic exercise session among sedentary adults. | Inappropriate population |
| 2055 | ENGAGE: Guided activity-based gaming in neurorehabilitation after stroke: A pilot study. | Inappropriate population |
| 2056 | Life events, perceived stress and depressive symptoms in a physical activity intervention with young adult women. | Inappropriate outcome |
| 2057 | Physical activity across frailty phenotypes in females with Parkinson's disease. | Inappropriate population |
| 2058 | Cardiorespiratory response to aerobic exercise programs with different intensity: 20 weeks longitudinal study. | Inappropriate population |
| 2059 | The effect of Tai Chi intervention on balance in older males. | Inappropriate population |
| 2060 | Cycling and walking for transport: Estimating net health effects from comparison of different transport mode users' self-reported physical activity. | Inappropriate outcome |
| 2061 | The effect of lifestyle practices on blood glucose levels and the development of diabetic complications in women diagnosed with type II diabetes mellitus in Guyana. | Inappropriate study design |
| 2062 | An epidemiological survey of arthritis in the population of north Gujarat, India. | Inappropriate study design |
| 2063 | TECNOB study: Ad interim results of a randomized controlled trial of a multidisciplinary telecare intervention for obese patients with type-2 diabetes. | Inappropriate study design |
| 2064 | Relationship among different tests of evaluating low back core stability. | Inappropriate outcome |
| 2065 | Age-related ventricular remodeling is an independent risk for heart failure symptoms in subjects with preserved systolic function. | Inappropriate population |
| 2066 | Sensory electrical nerve stimulation for training dynamic balance responses in a chronic stroke patient. | Inappropriate population |
| 2067 | Hatha yoga practices: Energy expenditure, respiratory changes and intensity of exercise. | Inappropriate study design |
| 2068 | Comparison of electrical stimulation and isometric training on isokinetic strength of knee extensors: A randomized clinical trial. | Inappropriate study design |
| 2069 | Walking before study enhances free recall but not judgement-of-learning magnitude. | Inappropriate population |
| 2070 | Effects of a flexibility and relaxation programme, walking, and nordic walking on parkinson's disease. | Inappropriate study design |
| 2071 | Geriatric orthopaedics:Acetabular fractures in the elderly. | Inappropriate population |
| 2072 | Perspectives on the Expert Patient Programme delivered by bilingual tutors. | Inappropriate study design |
| 2073 | Three cases of exercise-induced rhabdomyolysis: A potential hazard in university students participating in orientation programs. | Inappropriate population |
| 2074 | Therapeutic hypertension system based on a microbreathing pressure sensor system. | Inappropriate population |
| 2075 | The effect of associative and dissociative attentional focus strategies on muscle activity and heart rate during a weight training exercise. | Inappropriate population |
| 2076 | Effects of two modes of exercise on physical fitness and endothelial function in the elderly: Exercise with a flexible stick versus Tai Chi. | Inappropriate outcome |
| 2077 | Occupational therapy utilization for individuals with brain cancer following a craniotomy: A descriptive study. | Inappropriate population |
| 2078 | Fatigue and multidimensional disease severity in chronic obstructive pulmonary disease. | Inappropriate study design |
| 2079 | Factors impacting on the outcomes of Greek intervention programmes for children with autistic spectrum disorders. | Inappropriate study design |
| 2080 | The effectiveness of two energy drinks on selected indices of maximal cardiorespiratory fitness and blood lactate levels in male athletes. | Inappropriate population |
| 2081 | Osteoporosis in frail older people. | Inappropriate population |
| 2082 | Stressful life events and habitual physical activity in older adults: 1-Year accelerometer data from the Nakanojo study. | Inappropriate intervention |
| 2083 | Maximal power output during incremental exercise by resistance and endurance trained athletes. | Inappropriate study design |
| 2084 | Effect of different musical tempo on post-exercise recovery in young adults. | Inappropriate population |
| 2085 | A pilot study of aerobic exercise as an adjunctive treatment for drug dependence. | Inappropriate study design |
| 2086 | Determining emotion management skills of adults taking physical exercise and individuals leading a sedentary life. | Inappropriate study design |

| | | |
|---|---|---|
| 2087 | Effects of moderate-volume, high-load lower-body resistance training on strength and function in persons with parkinson's disease: A pilot study. | Inappropriate outcome |
| 2088 | Exercise, fitness and cognition - A randomised controlled trial in older individuals: The DR's EXTRA study. | Inappropriate population |
| 2089 | Electrophysiological correlates of listening effort: Neurodynamical modeling and measurement. | Inappropriate study design |
| 2090 | Modalities of empowerment and strengthening of ex-servicemen. | Inappropriate study design |
| 2091 | Ancient wisdom meets modern ailment - Traditional Asian medicine improves psychological vigor in stressed subjects. | Inappropriate study design |
| 2092 | The association of exercise with sleep, sex steroids, sexual activities and aging in Asian men. | Inappropriate study design |
| 2093 | Accelerometry to assess preschooler's free-play: Issues with count thresholds and epoch durations. | Inappropriate study design |
| 2094 | The potential effects of a biofeedback writing exercise on radial artery blood flow and neck mobility. | Inappropriate population |
| 2095 | The multistage shuttle run: The influence of verbal cues. | Inappropriate study design |
| 2096 | Functional fitness and blood pressure in low-income Brazilian elderly women. | Inappropriate study design |
| 2097 | Older women's assessment and management of chronic pain in their daily lives. | Inappropriate study design |
| 2098 | Assessing cardiovascular risk in the 10-minute consultation. | Inappropriate study design |
| 2099 | Sustainable participation in regular exercise amongst older people: Developing an action research approach. | Inappropriate population |
| 2100 | Effect of carbohydrate drink intake patterns on exercise in heat. | Inappropriate study design |
| 2101 | Comparison of prevalence of premenstrual syndrome in swimmer and non-swimmer students: A historical cohort study. | Inappropriate study design |
| 2102 | Effect of an exercise programme on stroke volume in patients with cystic fibrosis. | Inappropriate study design |
| 2103 | Ginseng supplementation does not change lactate threshold and physical performances in physically active Thai men. | Inappropriate intervention |
| 2104 | Cognitive impairment in elderly women: The relative importance of selected genes, lifestyle factors, and comorbidities. | Inappropriate population |
| 2105 | Effect of different multipliers and their interactions during manual lifting operations. | Inappropriate intervention |
| 2106 | Illness beliefs in chronic fatigue syndrome: A study involving affected adolescents and their parents. | Inappropriate study design |
| 2107 | Type 2 diabetics without insulin therapy in Germany: Epidemiological characteristics. | Inappropriate study design |
| 2108 | Effects of lesion laterality on cardiovascular responses of post stroke hemiparetic patients to a single bout of exercise. | Inappropriate study design |
| 2109 | A pilot falls-prevention programme for older people with dementia from a predominantly Italian background. | Inappropriate population |
| 2110 | Diabetes in older people. | Inappropriate study design |
| 2111 | Effect of a comprehensive yoga-based lifestyle modification program on lipid peroxidation. | Inappropriate population |
| 2112 | Grip strength and endurance: Influences of anthropometric variation, hand dominance, and gender. | Inappropriate study design |
| 2113 | Central hypoventilation: A complex presentation in an 83 year old. | Inappropriate population |
| 2114 | Effectiveness of constraint-induced movement therapy in chronic stroke patients. | Inappropriate study design |
| 2115 | Smoking attenuates regular aerobic exercise benefits to episodic free recall immediately following strenuous physical activity. | Inappropriate study design |
| 2116 | Effect of branched-chain amino acid supplementation on human growth hormone and cortisol secretion after exercise. | Inappropriate outcome |
| 2117 | Energy cost and cardiorespiratory changes during the practice of Surya Namaskar. | Inappropriate study design |
| 2118 | A qualitative study into the informational needs of coronary heart disease patients. | Inappropriate population |
| 2119 | Oxygen saturation response to exercise VO2 at 2100 m and 4350 m in women mountaineering trainees. | Inappropriate study design |
| 2120 | Effect of branched-chain amino acid supplementation on body composition measured with SBIA: An advanced BIA. | Inappropriate population |
| 2121 | Different training modes for the rotator cuff muscle group. A comparative study. | Inappropriate study design |
| 2122 | The effects of four weeks of ribose supplementation on body composition and exercise performance in healthy, young, male recreational bodybuilders: A double-blind, placebo-controlled trial. | Inappropriate study design |

| | | |
|---|---|---|
| 2123 | Knowledge of asthma and its management: A study in primary schoolteachers in Hong Kong. | Inappropriate outcome |
| 2124 | Management of stroke in the new millennium. | Inappropriate study design |
| 2125 | Resisted exercise training of institutionalized older adults for improved strength and functional mobility: A review. | Inappropriate population |
| 2126 | Cardiac rehabilitation (CR): Use of manual materials handling. | Inappropriate study design |
| 2127 | Effect of Sahaja yoga practice on stress management in patients of epilepsy. | Inappropriate study design |
| 2128 | Cardiovascular response patterning and type A behavior in middle aged men. | Inappropriate outcome |
| 2129 | Physiological activity during a prolonged mental stress task: Evidence for a shift in the control of pressor reactions. | Inappropriate study design |
| 2130 | An exploratory randomized sub-study of light-to-moderate intensity exercise on cognitive function, depression symptoms and inflammation in older adults with heart failure [with consumer summary] | Inappropriate population |
| 2131 | Is a stepped-care intervention effective in overweight and obese people with medial tibiofemoral osteoarthritis? The STrEAMline study: a randomised controlled trial [with consumer summary] | Inappropriate study design |
| 2132 | Scapular exercise combined with cognitive functional therapy is more effective at reducing chronic neck pain and kinesiophobia than scapular exercise alone: a randomized controlled trial [with consumer summary] | Inappropriate study design |
| 2133 | Combined exercise training improves cognitive functions in multiple sclerosis patients with cognitive impairment: a single-blinded randomized controlled trial [with consumer summary] | Inappropriate outcome |
| 2134 | Synergistic benefits of combined aerobic and cognitive training on fluid intelligence and the role of IGF-1 in chronic stroke | Inappropriate population |
| 2135 | Aerobic and strength training exercise programme for cognitive impairment in people with mild to moderate dementia: the DAPA RCT [with consumer summary] | Inappropriate study design |
| 2136 | Reducing the burden of dizziness in middle-aged and older people: a multifactorial, tailored, single-blind randomized controlled trial [with consumer summary] | Inappropriate study design |
| 2137 | Dual-task exercise reduces cognitive-motor interference in walking and falls after stroke | Inappropriate outcome |
| 2138 | Social dancing and incidence of falls in older adults: a cluster randomised controlled trial [with consumer summary] | Inappropriate population |
| 2139 | Exercise training and recreational activities to promote executive functions in chronic stroke: a proof-of-concept study | Inappropriate study design |
| 2140 | Effect of a multifactorial, interdisciplinary intervention on risk factors for falls and fall rate in frail older people: a randomised controlled trial [with consumer summary] | Inappropriate study design |
| 2141 | The effects of multimodal exercise on cognitive and physical functioning and brain-derived neurotrophic factor in older women: a randomised controlled trial [with consumer summary] | Inappropriate study design |
| 2142 | Effects of functional tasks exercise on older adults with cognitive impairment at risk of Alzheimer's disease: a randomised controlled trial [with consumer summary] | Inappropriate study design |
| 2143 | Effects of a multimodal exercise program on balance, functional mobility and fall risk in older adults with cognitive impairment: a randomized controlled single-blind study | Inappropriate outcome |
| 2144 | Effect of a multifactorial interdisciplinary intervention on mobility-related disability in frail older people: randomised controlled trial | Inappropriate study design |
| 2145 | Feasibility and effects of a physical exercise programme in adults with myotonic dystrophy type 1: a randomized controlled pilot study | Inappropriate study design |
| 2146 | Mobility training after hip fracture: a randomised controlled trial [with consumer summary] | Inappropriate study design |
| 2147 | Cognitive behavioral therapy or graded exercise therapy compared with usual care for severe fatigue in patients with advanced cancer during treatment: a randomized controlled trial [with consumer summary] | Inappropriate study design |
| 2148 | Combining cognitive stimulation therapy and fall prevention exercise (CogEx) in older adults with mild to moderate dementia: a feasibility randomised controlled trial | Inappropriate population |
| 2149 | Effects and mediating mechanisms of a structured limbs-exercise program on general cognitive function in older adults with mild cognitive impairment: a randomized controlled trial | Inappropriate outcome |

| | | |
|---|---|---|
| 2150 | The effects of virtual kayak paddling exercise on postural balance, muscle performance, and cognitive function in older adults with mild cognitive impairment: a randomized controlled trial | Inappropriate population |
| 2151 | An augmented prescribed exercise program (APEP) to improve mobility of older acute medical patients -- a randomized, controlled pilot and feasibility trial | Inappropriate study design |
| 2152 | Multidomain lifestyle intervention benefits a large elderly population at risk for cognitive decline and dementia regardless of baseline characteristics: the FINGER trial [with consumer summary] | Inappropriate study design |
| 2153 | Effects of a specially designed aerobic dance routine on mild cognitive impairment | Inappropriate population |
| 2154 | The effects of group-based versus individual-based Tai Chi training on nonmotor symptoms in patients with mild to moderate Parkinson's disease: a randomized controlled pilot trial | Inappropriate population |
| 2155 | Effects of momentum-based dumbbell training on cognitive function in older adults with mild cognitive impairment: a pilot randomized controlled trial | Inappropriate study design |
| 2156 | Would older adults with mild cognitive impairment adhere to and benefit from a structured lifestyle activity intervention to enhance cognition? A cluster randomized controlled trial | Inappropriate study design |
| 2157 | Effects of resistance exercise on fatigue and quality of life in breast cancer patients undergoing adjuvant chemotherapy: a randomized controlled trial [with consumer summary] | Inappropriate population |
| 2158 | A 2 year multidomain intervention of diet, exercise, cognitive training, and vascular risk monitoring versus control to prevent cognitive decline in at-risk elderly people (FINGER): a randomised controlled trial [with consumer summary] | Inappropriate study design |
| 2159 | Tai Chi training may reduce dual task gait variability, a potential mediator of fall risk, in healthy older adults: cross-sectional and randomized trial studies | Inappropriate population |
| 2160 | Dose-response of aerobic exercise on cognition: a community-based, pilot randomized controlled trial | Inappropriate population |
| 2161 | Feasibility and safety of early combined cognitive and physical therapy for critically ill medical and surgical patients: the Activity and Cognitive Therapy in ICU (ACT-ICU) trial | Inappropriate study design |
| 2162 | Home-based walking exercise in peripheral artery disease: 12-month follow-up of the goals randomized trial | Inappropriate intervention |
| 2163 | A randomized controlled pilot study of home-based step training in older people using videogame technology | Inappropriate study design |
| 2164 | The mental activity and exercise (MAX) trial: a randomized controlled trial to enhance cognitive function in older adults | Inappropriate study design |
| 2165 | Home-based walking exercise intervention in peripheral artery disease: a randomized clinical trial | Inappropriate population |
| 2166 | Effectiveness of an upper extremity exercise device and text message reminders to exercise in adults with spina bifida: a pilot study | Inappropriate population |
| 2167 | Effects of exercise intervention on vascular risk factors in older adults with mild cognitive impairment: a randomized controlled trial | Inappropriate study design |
| 2168 | Effects of high-intensity exercise and protein supplement on muscle mass in ADL dependent older people with and without malnutrition -- a randomized controlled trial | Inappropriate study design |
| 2169 | Patient education integrated with acupuncture for relief of cancer-related fatigue randomized controlled feasibility study | Inappropriate study design |
| 2170 | An exploratory cluster randomized controlled trial of group exercise on mobility and depression in care home residents [with consumer summary] | Inappropriate study design |
| 2171 | A randomized trial of a physical conditioning program to enhance the driving performance of older persons | Inappropriate study design |
| 2172 | Effects of a fall prevention program including exercise on mobility and falls in frail older people living in residential care facilities | Inappropriate study design |
| 2173 | Zumba dancing and aerobic exercise can improve working memory, motor function, and depressive symptoms in female patients with fibromyalgia [with consumer summary] | Inappropriate study design |
| 2174 | Effects of Tai Chi Chuan on inhibitory control in elderly women: an fNIRS study | Inappropriate study design |
| 2175 | Frequent karaoke training improves frontal executive cognitive skills, tongue pressure, and respiratory function in elderly people: pilot study from a randomized controlled trial | Inappropriate study design |
| 2176 | Effects of a 16-week multimodal exercise program on gait performance in individuals with dementia: a multicenter randomized controlled trial | Inappropriate study design |

| | | |
|---|---|---|
| 2177 | Lifestyle risk factors and cognitive outcomes from the multidomain dementia risk reduction randomized controlled trial, body brain life for cognitive decline (BBL-CD) | Inappropriate study design |
| 2178 | Effectiveness of a multicomponent treatment for fibromyalgia based on pain neuroscience education, exercise therapy, psychological support, and nature exposure (NAT-FM): a pragmatic randomized controlled trial | Inappropriate study design |
| 2179 | Sardinian folk dance for individuals with Parkinson's disease: a randomized controlled pilot trial | Inappropriate study design |
| 2180 | The effects of different types of physical exercise on physical and cognitive function in frail institutionalized older adults with mild to moderate cognitive impairment. A randomized controlled trial [with consumer summary] | Inappropriate study design |
| 2181 | Effect of exercise intervention on functional decline in very elderly patients during acute hospitalization: a randomized clinical trial [with consumer summary] | Inappropriate study design |
| 2182 | Assessing the impact of physical exercise on cognitive function in older medical patients during acute hospitalization: secondary analysis of a randomized trial | Inappropriate outcome |
| 2183 | Effects of traditional versus iPad-enhanced aerobic exercise on wayfinding efficacy and cognition: a pilot randomized controlled trial | Inappropriate study design |
| 2184 | Dual-task walking capacity mediates Tai Ji Quan impact on physical and cognitive function | Inappropriate population |
| 2185 | Effects of a modified Tai Chi program on older people with mild dementia: a randomized controlled trial | Inappropriate population |
| 2186 | Randomized controlled trial of increasing physical activity on objectively measured and self-reported cognitive functioning among breast cancer survivors: the memory and motion study | Inappropriate study design |
| 2187 | The Mental Activity and eXercise (MAX) trial: effects on physical function and quality of life among older adults with cognitive complaints | Inappropriate intervention |
| 2188 | Ground kayak paddling exercise improves postural balance, muscle performance, and cognitive function in older adults with mild cognitive impairment: a randomized controlled trial | Inappropriate outcome |
| 2189 | Effects of golf training on cognition in older adults: a randomised controlled trial [with consumer summary] | Inappropriate study design |
| 2190 | Exercise training effects on memory and hippocampal viscoelasticity in multiple sclerosis: a novel application of magnetic resonance elastography | Inappropriate study design |
| 2191 | Economic evaluation of aerobic exercise training in older adults with vascular cognitive impairment: PROMoTE trial [with consumer summary] | Inappropriate study design |
| 2192 | Multiple-modality exercise and mind-motor training to improve cardiovascular health and fitness in older adults at risk for cognitive impairment: a randomized controlled trial | Inappropriate study design |
| 2193 | Baseline telomere length and effects of a multidomain lifestyle intervention on cognition: the FINGER randomized controlled trial | Inappropriate study design |
| 2194 | Open- and closed-skill exercise interventions produce different neurocognitive effects on executive functions in the elderly: a 6-month randomized, controlled trial | Inappropriate population |
| 2195 | Effects of a long-term aerobic exercise intervention on institutionalized patients with dementia [with consumer summary] | Inappropriate study design |
| 2196 | Fascial manipulation associated with standard care compared to only standard postsurgical care for total hip arthroplasty: a randomized controlled trial | Inappropriate outcome |
| 2197 | Effect of a combining cognitive and balanced training on the cognitive, postural and functional status of seniors with a mild cognitive deficit in a randomized, controlled trial | Inappropriate study design |
| 2198 | Impact of Tai Chi on falls among preclinically disabled older people. A randomized controlled trial | Inappropriate study design |
| 2199 | In-patient multidisciplinary rehabilitation for Parkinson's disease: a randomized controlled trial | Inappropriate population |
| 2200 | Physical fitness exercise versus cognitive behavior therapy on reducing the depressive symptoms among community-dwelling elderly adults: a randomized controlled trial [with consumer summary] | Inappropriate population |
| 2201 | Aerobic exercise and yoga improve neurocognitive function in women with early psychosis | Inappropriate study design |
| 2202 | Mental and physical activities delay cognitive decline in older persons with dementia | Inappropriate intervention |
| 2203 | Effect of music-based multitask training on cognition and mood in older adults [with consumer summary] | Inappropriate outcome |
| 2204 | Effect of weight loss, exercise, or both on cognition and quality of life in obese older adults | Inappropriate population |

| | | |
|---|---|---|
| 2205 | A randomized controlled trial of multicomponent exercise in older adults with mild cognitive impairment | Inappropriate population |
| 2206 | A behavioral weight-loss intervention in persons with serious mental illness | Inappropriate study design |
| 2207 | Multicenter, randomized controlled trial of yoga for sleep quality among cancer survivors | Inappropriate study design |
| 2208 | Exergaming and older adult cognition: a cluster randomized clinical trial | Inappropriate population |
| 2209 | Impact of Tai Chi on impairment, functional limitation, and disability among preclinically disabled older people: a randomized controlled trial | Inappropriate intervention |
| 2210 | Efficacy of a multimodal cognitive rehabilitation including psychomotor and endurance training in Parkinson's disease | Inappropriate population |
| 2211 | Exercise training improves selected aspects of daytime functioning in adults with obstructive sleep apnea [with consumer summary] | Inappropriate study design |
| 2212 | Effects of multicomponent exercise on cognitive function in older adults with amnestic mild cognitive impairment: a randomized controlled trial | Inappropriate study design |
| 2213 | Does a multicomponent exercise program improve dual-task performance in amnestic mild cognitive impairment? A randomized controlled trial | Inappropriate study design |
| 2214 | Effects of resistance training exercise on cognitive performance in elderly individuals with memory impairment: results of a controlled trial | Inappropriate study design |
| 2215 | Integration of motor imagery and physical practice in group treatment applied to subjects with Parkinson's disease | Inappropriate population |
| 2216 | Evaluation of an individualised asthma programme directed at behavioural change | Inappropriate population |
| 2217 | Effectiveness of behavioral therapy to treat incontinence in homebound older adults | Inappropriate study design |
| 2218 | Comparison of cognitive-behavioral group treatment and an alternative non-psychological treatment for chronic low back pain | Inappropriate population |
| 2219 | Distinct effects of acute exercise and breaks in sitting on working memory and executive function in older adults: a three-arm, randomised cross-over trial to evaluate the effects of exercise with and without breaks in sitting on cognition [with consumer summary] | Inappropriate outcome |
| 2220 | A randomised controlled study of high intensity exercise as a dishabituating stimulus to improve hypoglycaemia awareness in people with type 1 diabetes: a proof-of-concept study [with consumer summary] | Inappropriate population |
| 2221 | Falls risk in relation to activity exposure in high risk older adults | Inappropriate study design |
| 2222 | Is physical exercise and dietary therapy a feasible alternative to cognitive behavior therapy in treatment of eating disorders? A randomized controlled trial of two group therapies | Inappropriate population |
| 2223 | Safety and feasibility of a Dalcroze eurhythmics and a simple home exercise program among older adults with mild cognitive impairment (MCI) or mild dementia: the MOVE for your MIND pilot trial | Inappropriate population |
| 2224 | Metabolic and neurocognitive changes following lifestyle modification: examination of biomarkers from the ENLIGHTEN randomized clinical trial | Inappropriate population |
| 2225 | Improved functional performance in individuals with dementia after a moderate-intensity home-based exercise program: a randomized controlled trial | Inappropriate study design |
| 2226 | Breath Qigong improves recognition in seniors with vascular cognitive impairment | Inappropriate study design |
| 2227 | The effects of high-intensity aerobic exercise on cognitive performance after stroke: a pilot randomised controlled trial | Inappropriate study design |
| 2228 | Aerobic exercise sustains performance of instrumental activities of daily living in early-stage Alzheimer disease | Inappropriate population |
| 2229 | The effect of multidomain lifestyle intervention on daily functioning in older people | Inappropriate population |
| 2230 | Xbox 360 Kinect cognitive games improve slowness, complexity of EEG, and cognitive functions in subjects with mild cognitive impairment: a randomized control trial | Inappropriate comparison |
| 2231 | Contemporary dance practice improves motor function and body representation in Huntington's disease: a pilot study | Inappropriate study design |
| 2232 | Brain volumes and cortical thickness on MRI in the Finnish Geriatric Intervention Study to Prevent Cognitive Impairment and Disability (FINGER) | Inappropriate study design |
| 2233 | Exercise training induced changes in nuclear magnetic resonance-measured lipid particles in mild cognitively impaired elderly African American volunteers: a pilot study | Inappropriate study design |
| 2234 | Promoting Activity, Independence and Stability in Early Dementia (PrAISED): a, multisite, randomised controlled, feasibility trial | Inappropriate study design |

| | | |
|---|---|---|
| 2235 | The effect of aerobic exercise on white matter hyperintensity progression may vary by sex | Inappropriate study design |
| 2236 | Effects of physical activity in nursing home residents with dementia: a randomized controlled trial | Inappropriate population |
| 2237 | Effects of BEAT Cancer randomized physical activity trial on subjective memory impairments in breast cancer survivors [with consumer summary] | Inappropriate study design |
| 2238 | Weight-loss outcomes from a pilot study of African dance in older African Americans | Inappropriate population |
| 2239 | Effects of exercise on the cognition of older women treated with lovastatin | Inappropriate population |
| 2240 | Effects of feedback-based balance and core resistance training versus Pilates training on cognitive functions in older women with mild cognitive impairment: a pilot randomized controlled trial | Inappropriate study design |
| 2241 | Happy despite pain: a randomized controlled trial of an 8-week internet-delivered positive psychology intervention for enhancing well-being in patients with chronic pain | Inappropriate study design |
| 2242 | A paleolithic diet with and without combined aerobic and resistance exercise increases functional brain responses and hippocampal volume in subjects with type 2 diabetes | Inappropriate population |
| 2243 | Resistance training with instability for patients with Parkinson's disease | Inappropriate study design |
| 2244 | Influence of yoga and aerobics exercise on fatigue, pain and psychosocial status in patients with multiple sclerosis: a randomized trial | Inappropriate population |
| 2245 | Effect of chair stand exercise on activity of daily living: a randomized controlled trial in hemodialysis patients | Inappropriate study design |
| 2246 | Exercise augmentation of exposure therapy for PTSD: rationale and pilot efficacy data | Inappropriate population |
| 2247 | The effects of aerobic exercise on cognitive function of Alzheimer's disease patients | Inappropriate population |
| 2248 | Novel occupational therapy interventions may improve quality of life in older adults with dementia | Inappropriate outcome |
| 2249 | Does treadmill training improve lower-extremity tasks in Parkinson disease? A randomized controlled trial | Inappropriate population |
| 2250 | Physical activity and executive functions in the elderly with mild cognitive impairment | Inappropriate study design |
| 2251 | Fall and injury prevention in residential care -- effects in residents with higher and lower levels of cognition | Inappropriate population |
| 2252 | The mediating effect of different exercise programs on the immune profile of frail older women with cognitive impairment | Inappropriate population |
| 2253 | Effects of single-task versus dual-task training on balance performance in elderly patients with knee osteoarthritis | Inappropriate outcome |
| 2254 | Three months of multimodal training contributes to mobility and executive function in elderly individuals with mild cognitive impairment, but not in those with Alzheimer's disease: a randomized controlled trial [with consumer summary] | Inappropriate study design |
| 2255 | Aerobic exercise promotes executive functions and impacts functional neural activity among older adults with vascular cognitive impairment [with consumer summary] | Inappropriate study design |
| 2256 | Effects of resistance exercise training on cognitive function and physical performance in cognitive frailty: a randomized controlled trial | Inappropriate study design |
| 2257 | Feasibility and efficacy of a multi-factorial intervention to prevent falls in older adults with cognitive impairment living in residential care (ProF-Cog). A feasibility and pilot cluster randomised controlled trial | Inappropriate study design |
| 2258 | Effects of pelvic floor muscle exercise on urinary incontinence in elderly women with cognitive impairment | Inappropriate study design |
| 2259 | Feasible modalities and long-term effects of elastic band exercises in nursing home older adults in wheelchairs: a cluster randomized controlled trial [with consumer summary] | Inappropriate study design |
| 2260 | Taoist Tai Chi and memory intervention for individuals with mild cognitive impairment | Inappropriate study design |
| 2261 | Elastic band exercises improved activities of daily living and functional fitness of wheelchair-bound older adults with cognitive impairment: a cluster randomized controlled trial | Inappropriate population |
| 2262 | Does multicomponent physical exercise with simultaneous cognitive training boost cognitive performance in older adults? A 6-month randomized controlled trial with a 1-year follow-up | Inappropriate study design |

| | | |
|---|---|---|
| 2263 | A standardized randomized 6-month aerobic exercise-training down-regulated pro-inflammatory genes, but up-regulated anti-inflammatory, neuron survival and axon growth-related genes | Inappropriate outcome |
| 2264 | Resistance exercise may improve spatial awareness and visual reaction in older adults [with consumer summary] | Inappropriate population |
| 2265 | Virtual games and quality of life in Parkinson's disease: a randomised controlled trial | Inappropriate study design |
| 2266 | Adherence to exercise programs and determinants of maintenance in older adults with mild cognitive impairment | Inappropriate population |
| 2267 | Effect of medical Qigong on cognitive function, quality of life, and a biomarker of inflammation in cancer patients: a randomized controlled trial | Inappropriate study design |
| 2268 | Sensory-specific balance training in older adults: effect on proprioceptive reintegration and cognitive demands | Inappropriate study design |
| 2269 | Operant-behavioural and cognitive-behavioural treatment for chronic low back pain | Inappropriate study design |
| 2270 | Action observation improves sit-to-walk in patients with Parkinson's disease and freezing of gait. Biomechanical analysis of performance [with consumer summary] | Inappropriate study design |
| 2271 | Mind-body exercise improves cognitive function and modulates the function and structure of the hippocampus and anterior cingulate cortex in patients with mild cognitive impairment [with consumer summary] | Inappropriate study design |
| 2272 | Comparative evaluation of weight bearing exercise and vestibular stimulation on balance in children with Down syndrome | Inappropriate study design |
| 2273 | The role of long-term physical exercise on performance and brain activation during the Stroop colour word task in fibromyalgia patients | Inappropriate study design |
| 2274 | Preliminary evidence of improved cognitive performance following vestibular rehabilitation in children with combined ADHD (cADHD) and concurrent vestibular impairment | Inappropriate study design |
| 2275 | Multimodal exercise training during myeloablative chemotherapy: a prospective randomized pilot trial | Inappropriate population |
| 2276 | An exercise program to prevent falls in institutionalized elderly with cognitive deficits: a crossover pilot study | Inappropriate outcome |
| 2277 | A phase II exploratory cluster randomized controlled trial of a group mobility training and staff education intervention to promote urinary continence in UK care homes [with consumer summary] | Inappropriate study design |
| 2278 | Comparison of integrated group therapy and group relaxation training for fibromyalgia | Inappropriate population |
| 2279 | Functional incidental training, mobility performance, and incontinence care with nursing home residents | Inappropriate population |
| 2280 | Regional brain volumes moderate, but do not mediate, the effects of group-based exercise training on reductions in loneliness in older adults | Inappropriate population |
| 2281 | (Effect of regular physical exercise on elderly with mild cognitive impairment) [Chinese - simplified characters] | Inappropriate population |
| 2282 | The Group Oriented Arterial Leg Study (GOALS) to improve walking performance in patients with peripheral arterial disease | Inappropriate intervention |
| 2283 | Tailored exercise and home hazard reduction for fall prevention in older people with cognitive impairment: the i-FOCIS randomized controlled trial | Inappropriate study design |
| 2284 | Rehabilitation of ADHD children by sport intervention: a Tunisian experience | Inappropriate study design |
| 2285 | (Effect of action observation and imitation on improving the functional activities indices in hemiplegic patients based on mirror neurons theory) [Persian] | Inappropriate population |
| 2286 | Effectiveness of stepped-care intervention in overweight and obese patients with medial tibiofemoral osteoarthritis: a randomized controlled trial [with consumer summary] | Inappropriate population |
| 2287 | Feasibility and acceptability of a preoperative exercise program for patients undergoing major cancer surgery: results from a pilot randomized controlled trial | Inappropriate population |
| 2288 | Cost-effectiveness and cost-utility of a home-based exercise program in geriatric patients with cognitive impairment | Inappropriate intervention |
| 2289 | E-health StandingTall balance exercise for fall prevention in older people: results of a two year randomised controlled trial [with consumer summary] | Inappropriate intervention |
| 2290 | Implementing an online virtual falls prevention intervention during a public health pandemic for older adults with mild cognitive impairment: a feasibility trial | Inappropriate outcome |
| 2291 | Effect of aerobic exercise on amyloid accumulation in preclinical Alzheimer's: a 1-year randomized controlled trial | Inappropriate intervention |
| 2292 | Effects of physical-cognitive training on physical and psychological functions among older adults with type 2 diabetes and balance impairment: a randomized controlled trial | Inappropriate population |

| | | |
|---|---|---|
| 2293 | A novel cognitive-motor exercise program delivered via a tablet to improve mobility in older people with cognitive impairment -- StandingTall cognition and mobility [with consumer summary] | Inappropriate population |
| 2294 | Effects of aerobic exercise on cognitive function in women with methamphetamine dependence in a detoxification program in Tianjin, China: a randomized controlled trial | Inappropriate intervention |
| 2295 | Short- and long-term changes in cognitive function after exercise-based rehabilitation in people with COPD: a pilot study | Inappropriate population |
| 2296 | The effectiveness of Tai Chi Chuan on fear of movement, prevention of falls, physical activity, and cognitive status in older adults with mild cognitive impairment: a randomized controlled trial | Inappropriate population |
| 2297 | Efficacy of high-intensity aerobic exercise on cognitive performance in people with multiple sclerosis: a randomized controlled trial | Inappropriate intervention |
| 2298 | Effect of a home-based exercise program on indices of physical function and quality of life in elderly maintenance hemodialysis patients | Inappropriate population |
| 2299 | Neurocognitive plasticity is associated with cardiorespiratory fitness following physical exercise in older adults with amnestic mild cognitive impairment | Inappropriate intervention |
| 2300 | The effect of physical activity with and without cognitive demand on the improvement of executive functions and behavioral symptoms in children with ADHD | Inappropriate study design |
| 2301 | Clinical efficacy of aerobic exercise combined with computer-based cognitive training in stroke: a multicenter randomized controlled trial | Inappropriate intervention |
| 2302 | Is Tai Chi an effective intervention for enhancing health-related quality of life in older people with mild cognitive impairment? An interventional study [with consumer summary] | Inappropriate outcome |
| 2303 | Effect of exergaming versus combined exercise on cognitive function and brain activation in frail older adults: a randomized controlled trial [with consumer summary] | Inappropriate intervention |
| 2304 | A pilot randomized controlled trial of robotic exoskeleton-assisted exercise rehabilitation in multiple sclerosis [with consumer summary] | Inappropriate study design |
| 2305 | One-year aerobic exercise altered cerebral vasomotor reactivity in mild cognitive impairment [with consumer summary] | Inappropriate intervention |
| 2306 | Multi-component intervention program on habitual physical activity parameters and cognitive function in patients with mild cognitive impairment: a randomized controlled trial | Inappropriate intervention |
| 2307 | Change in CAIDE dementia risk score and neuroimaging biomarkers during a 2-year multidomain lifestyle randomized controlled trial: results of a post-hoc subgroup analysis | Inappropriate intervention |
| 2308 | Effect of different types of exercises on psychological and cognitive features in people with Parkinson's disease: a randomized controlled trial | Inappropriate intervention |
| 2309 | The influence of a 16-week exercise program, APOE status, and age on executive function task performance: a randomized trial [with consumer summary] | Inappropriate intervention |
| 2310 | Effect of exoskeleton-assisted rehabilitation over prefrontal cortex in multiple sclerosis patients: a neuroimaging pilot study | Inappropriate intervention |
| 2311 | Mind-body exercise modulates locus coeruleus and ventral tegmental area functional connectivity in individuals with mild cognitive impairment | Inappropriate intervention |
| 2312 | Effect of cognitive behavior therapy in patients with chronic nonspecific low back pain | Inappropriate population |
| 2313 | Nursing effects of finger exercise on cognitive function and others for cerebral ischemic stroke patients | Inappropriate study design |
| 2314 | One-year aerobic exercise reduced carotid arterial stiffness and increased cerebral blood flow in amnestic mild cognitive impairment | Inappropriate intervention |
| 2315 | Multi-component exercise training improves the physical and cognitive function of the elderly with mild cognitive impairment: a six-month randomized controlled trial | Inappropriate intervention |
| 2316 | The effect of Chinese square dance exercise on cognitive function in older women with mild cognitive impairment: the mediating effect of mood status and quality of life | Inappropriate intervention |
| 2317 | White matter plasticity in healthy older adults: the effects of aerobic exercise | Inappropriate population |
| 2318 | Comparative effects of physical exercise and other behavioral interventions on functional status outcomes in mild cognitive impairment | Inappropriate study design |
| 2319 | Cognitive impairment and stroke in elderly patients | Inappropriate study design |
| 2320 | Menopause and exercise | Inappropriate study design |

| | | |
|---|---|---|
| 2321 | Exercise interventions for cognitive function in adults older than 50: a systematic review with meta-analysis | Inappropriate study design |
| 2322 | Health benefits of tai chi: What is the evidence? | Inappropriate study design |
| 2323 | Aerobic exercise to improve cognitive function in older people without known cognitive impairment | Inappropriate study design |
| 2324 | Physical Activity Interventions in Preventing Cognitive Decline and Alzheimer-Type Dementia: A Systematic Review | Inappropriate population |
| 2325 | Physical exercise and cognitive performance in the elderly: current perspectives | Inappropriate study design |
| 2326 | Effects of Acute and Chronic Exercise on Immunological Parameters in the Elderly Aged: Can Physical Activity Counteract the Effects of Aging? | Inappropriate study design |
| 2327 | High- and low-intensity exercise do not improve cognitive function after stroke: A randomized controlled trial | Inappropriate study design |
| 2328 | Role of physical exercise on cognitive function in healthy older adults: A systematic review of randomized clinical trials | Inappropriate study design |
| 2329 | Effects of low-intensity endurance and resistance training on mobility in chronic stroke survivors: a pilot randomized controlled study | Inappropriate population |
| 2330 | Physical activity improves cognition: possible explanations | Inappropriate study design |
| 2331 | Cognitive plasticity in older adults: effects of cognitive training and physical exercise | Inappropriate study design |
| 2332 | The effect of physical exercise on cerebral blood flow in Alzheimer's disease | Inappropriate study design |
| 2333 | Aerobic exercise effects upon cognition in Alzheimer's Disease: A systematic review of randomized controlled trials | Inappropriate study design |
| 2334 | The effects of an extensive exercise programme on the progression of Mild Cognitive Impairment (MCI): study protocol for a randomised controlled trial | Inappropriate study design |
| 2335 | Copenhagen Consensus statement 2019: physical activity and ageing | Inappropriate study design |
| 2336 | Physical exercise for prevention of dementia (EPD) study: background, design and methods | Inappropriate study design |
| 2337 | Can Exercise Improve Cognitive Symptoms of Alzheimer's Disease? | Inappropriate population |
| 2338 | The benefits of physical activities on cognitive and mental health in healthy and pathological aging | Inappropriate study design |
| 2339 | Exercise intensity affects acute neurotrophic and neurophysiological responses poststroke | Inappropriate study design |
| 2340 | Combined Aerobic and Resistance Training for Cardiorespiratory Fitness, Muscle Strength, and Walking Capacity after Stroke: A Systematic Review and Meta-Analysis | Inappropriate study design |
| 2341 | The effect of aerobic exercise on stroke rehabilitation | Inappropriate study design |
| 2342 | The Relationship Between Physical Activity and Dementia: A Systematic Review and Meta-Analysis of Prospective Cohort Studies | Inappropriate population |
| 2343 | Physical exercise as a preventive or disease-modifying treatment of dementia and brain aging | Inappropriate population |
| 2344 | The effect of an interactive cycling training on cognitive functioning in older adults with mild dementia: study protocol for a randomized controlled trial | Inappropriate population |
| 2345 | A Single Bout of High-Intensity Interval Training Improves Motor Skill Retention in Individuals With Stroke | Inappropriate study design |
| 2346 | Potentially modifiable lifestyle factors, cognitive reserve, and cognitive function in later life: A cross-sectional study | Inappropriate population |
| 2347 | Physical fitness training for stroke patients | Inappropriate study design |
| 2348 | Different types of exercise in Multiple Sclerosis: Aerobic exercise or Pilates, a single-blind clinical study | Inappropriate population |
| 2349 | Physical activity and enhanced fitness to improve cognitive function in older people without known cognitive impairment | Inappropriate study design |
| 2350 | Effects of Caffeine and Acute Aerobic Exercise on Working Memory and Caffeine Withdrawal | Inappropriate population |
| 2351 | Cognitive-motor exergaming for reducing fall risk in people with chronic stroke: A randomized controlled trial | Inappropriate study design |
| 2352 | Efficacy and mechanisms of combined aerobic exercise and cognitive training in mild cognitive impairment: study protocol of the ACT trial | Inappropriate population |
| 2353 | Potential Benefits of Physical Activity in MCI and Dementia | Inappropriate study design |
| 2354 | Effects of Physical Exercise on Working Memory and Prefrontal Cortex Function in Post-Stroke Patients | Inappropriate population |
| 2355 | Effects of Acute Aerobic and Resistance Exercise on Cognitive Function and Salivary Cortisol Responses | Inappropriate population |
| 2356 | Lifestyle Markers Predict Cognitive Function | Inappropriate study design |

| | | |
|---|---|---|
| 2357 | Acute Impact of Different Exercise Modalities on Arterial and Platelet Function | Inappropriate study design |
| 2358 | Physical activity as treatment for alcohol use disorders (FitForChange): study protocol for a randomized controlled trial | Inappropriate population |
| 2359 | Acute resistance exercise with blood flow restriction in elderly hypertensive women: haemodynamic, rating of perceived exertion and blood lactate | Inappropriate study design |
| 2360 | SYNERGIC TRIAL (SYNchronizing Exercises, Remedies in Gait and Cognition) a multi-Centre randomized controlled double blind trial to improve gait and cognition in mild cognitive impairment | Inappropriate study design |
| 2361 | Metabolomics of Aerobic Exercise in Chronic Stroke Survivors: A Pilot Study | Inappropriate study design |
| 2362 | Response heterogeneity in fitness, mobility and cognition with exercise-training in MS | Inappropriate outcome |
| 2363 | Vascular biomarkers to predict response to exercise in Alzheimer's disease: the study protocol | Inappropriate study design |
| 2364 | Cerebral changes improved by physical activity during cognitive decline: A systematic review on MRI studies | Inappropriate study design |
| 2365 | Acute Exercise Intensity and Memory Function: Evaluation of the Transient Hypofrontality Hypothesis | Inappropriate study design |
| 2366 | A single exercise bout and locomotor learning after stroke: physiological, behavioural, and computational outcomes | Inappropriate study design |
| 2367 | Body, Brain, Life for Cognitive Decline (BBL-CD): protocol for a multidomain dementia risk reduction randomized controlled trial for subjective cognitive decline and mild cognitive impairment | Inappropriate population |
| 2368 | Acute effects of aerobic exercise on cognitive function in individuals with Parkinson's disease | Inappropriate population |
| 2369 | Higher Cardiorespiratory Fitness Is Associated With Better Verbal Generativity in Community-Dwelling Older Adults | Inappropriate study design |
| 2370 | Study protocol for Vitality: a proof-of-concept randomised controlled trial of exercise training or complex mental and social activities to promote cognition in adults with chronic stroke | Inappropriate study design |
| 2371 | Self-Reported Physical Activity and Cardiovascular Disease Risk Factors in Patients with Lacunar Stroke | Inappropriate study design |
| 2372 | Cardiorespiratory fitness and cognitive processing speed in multiple sclerosis: The possible roles of psychological symptoms | Inappropriate study design |
| 2373 | Aerobic training session length affects arterial elasticity | Inappropriate population |
| 2374 | Effect of aerobic exercise prior to modified constraint-induced movement therapy outcomes in individuals with chronic hemiparesis: a study protocol for a randomized clinical trial | Inappropriate population |
| 2375 | The Influence of Cognitive Impairment on the Fitness-Cognition Relationship in Multiple Sclerosis | Inappropriate outcome |
| 2376 | The quest for synergy between physical exercise and cognitive stimulation via exergaming in people with dementia: a randomized controlled trial | Inappropriate study design |
| 2377 | Driving Fitness in Different Forms of Dementia: An Update | Inappropriate population |
| 2378 | Foot Inertial Sensing for Combined Cognitive-Motor Exercise of the Sustained Attention Domain | Inappropriate study design |
| 2379 | Effect of a multicomponent exercise programme (VIVIFRAIL) on functional capacity in frail community elders with cognitive decline: study protocol for a randomized multicentre control trial | Inappropriate study design |
| 2380 | Aerobic With Resistance Training or Aerobic Training Alone Poststroke: A Secondary Analysis From a Randomized Clinical Trial | Inappropriate study design |
| 2381 | The Acute Effect of High-Intensity Exercise on Executive Function: A Meta-Analysis | Inappropriate study design |
| 2382 | "Muscle-Gut-Brain Axis": Can Physical Activity Help Patients with Alzheimer's Disease Due to Microbiome Modulation? | Inappropriate population |
| 2383 | An integrative review of exercise interventions among community-dwelling adults with Alzheimer's disease | Inappropriate population |
| 2384 | Comparing self-reported and objective monitoring of physical activity in Parkinson disease | Inappropriate study design |
| 2385 | The Relation Between Physical Activity and Cognitive Change in Older Latinos | Inappropriate study design |
| 2386 | Intense resistance exercise increases peripheral brain-derived neurotrophic factor | Inappropriate intervention |
| 2387 | Practical applications of physical activity for successful cognitive aging | Inappropriate study design |
| 2388 | High levels of incidental physical activity are positively associated with cognition and EEG activity in aging | Inappropriate population |

| | | |
|---|---|---|
| 2389 | Exergaming as a Physical Exercise Strategy Reduces Frailty in People With Dementia: A Randomized Controlled Trial | Inappropriate study design |
| 2390 | Effects of aerobic training on physical activity in people with stroke: A randomized controlled trial | Inappropriate population |
| 2391 | Aftereffects of Cognitively Demanding Acute Aerobic Exercise on Working Memory | Inappropriate study design |
| 2392 | High-Intensity Intermittent Exercise: Effect on Young People's Cardiometabolic Health and Cognition | Inappropriate population |
| 2393 | Physical activity for cognitive health: what advice can we give to older adults with subjective cognitive decline and mild cognitive impairment? | Inappropriate study design |
| 2394 | Exercise Test Performance Reveals Evidence of the Cardiorespiratory Fitness Hypothesis | Inappropriate study design |
| 2395 | Aerobic Stimulus Induced by Virtual Reality Games in Stroke Survivors | Inappropriate outcome |
| 2396 | Acute high-intensity exercise-induced cognitive enhancement and brain-derived neurotrophic factor in young, healthy adults | Inappropriate population |
| 2397 | Cardiovascular and hemodynamic responses to adapted physical exercises in very old adults | Inappropriate study design |
| 2398 | Executive function: Responses to aerobic exercise in Alzheimer's disease | Inappropriate population |
| 2399 | A short bout of high-intensity exercise alters ipsilesional motor cortical excitability post-stroke | Inappropriate study design |
| 2400 | Physical frailty and health outcomes of fitness, hormones, psychological and disability in institutionalized older women: an exploratory association study | Inappropriate study design |
| 2401 | Flavanol-rich cocoa consumption enhances exercise-induced executive function improvements in humans | Inappropriate study design |
| 2402 | Physiotherapists' perspectives on aerobic exercise early after stroke: A preliminary study | Inappropriate population |
| 2403 | Exercise and Episodic Specificity Induction on Episodic Memory Function | Inappropriate study design |
| 2404 | The Energy Cost of Steady State Physical Activity in Acute Stroke | Inappropriate study design |
| 2405 | Assessing kinematics and kinetics of functional electrical stimulation rowing | Inappropriate study design |
| 2406 | Experimental effects of exercise on memory function among mild cognitive impairment: systematic review and meta-analysis | Inappropriate study design |
| 2407 | Investigating Gains in Neurocognition in an Intervention Trial of Exercise (IGNITE): Protocol | Inappropriate intervention |
| 2408 | How Exercise Influences Cognitive Performance When Mild Cognitive Impairment Exists: A Literature Review | Inappropriate population |
| 2409 | Effects of Aerobic Exercise on Cognitive Performance Among Young Adults in a Higher Education Setting | Inappropriate intervention |
| 2410 | Ketogenic diet, high intensity interval training (HIIT) and memory training in the treatment of mild cognitive impairment: A case study | Inappropriate study design |
| 2411 | Neurocognition in treatment-resistant hypertension: profile and associations with cardiovascular biomarkers | Inappropriate study design |
| 2412 | Effects of combined physical and cognitive training on fitness and neuropsychological outcomes in healthy older adults | Inappropriate study design |
| 2413 | Cerebrovascular Pulsatility During Rest and Exercise Reflects Hemodynamic Impairment in Stroke and Cerebral Small Vessel Disease | Inappropriate study design |
| 2414 | The MMSE should not be the sole indicator of fitness to drive in mild Alzheimer's dementia | Inappropriate study design |
| 2415 | What is the Relationship between Health, Mood, and Mild Cognitive Impairment? | Inappropriate study design |
| 2416 | Towards ageing well: Use it or lose it: Exercise, epigenetics and cognition | Inappropriate study design |
| 2417 | Systematic review of cardiopulmonary exercise testing post stroke: Are we adhering to practice recommendations? | Inappropriate study design |
| 2418 | Exercise, Dopamine, and Cognition in Older Age | Inappropriate study design |
| 2419 | The Effect of Stroke on Middle Cerebral Artery Blood Flow Velocity Dynamics During Exercise | Inappropriate study design |
| 2420 | Efficacy of different types of exercises on global cognition in adults with mild cognitive impairment: a network meta-analysis | Inappropriate population |
| 2421 | Patients' perspectives on aerobic exercise early after stroke | Inappropriate population |
| 2422 | Dose-dependent association of accelerometer-measured physical activity and sedentary time with brain perfusion in aging | Inappropriate population |
| 2423 | Early functional improvement after stroke correlates with cardiovascular fitness | Inappropriate population |
| 2424 | Effects of Cardiorespiratory Exercise on Cognition in Older Women Exposed to Air Pollution | Inappropriate population |

| | | |
|---|---|---|
| 2425 | Physical fitness in institutionalized older adults with dementia: association with cognition, functional capacity and quality of life | Inappropriate population |
| 2426 | Effects of combined training vs aerobic training on cognitive functions in COPD: a randomized controlled trial | Inappropriate outcome |
| 2427 | Predicting cognitive performance from physical activity and fitness in adolescents and young adults in Botswana relative to HIV status | Inappropriate intervention |
| 2428 | Relation between aerobic fitness and brain structures in amnestic mild cognitive impairment elderly | Inappropriate study design |
| 2429 | Home-Based Physical Behavior in Late Stage Parkinson Disease Dementia: Differences between Cognitive Subtypes | Inappropriate study design |
| 2430 | Experimental Effects of Acute Exercise in Attenuating Memory Interference: Considerations by Biological Sex | Inappropriate study design |
| 2431 | The Impact of Different Types of Exercise Training on Peripheral Blood Brain-Derived Neurotrophic Factor Concentrations in Older Adults: A Meta-Analysis | Inappropriate population |
| 2432 | The effect of acute exercise on the performance of verbal fluency in adolescents and young adults with Down syndrome: a pilot study | Inappropriate study design |
| 2433 | Interactive effects of physical fitness and body mass index on risk of stroke: A national cohort study | Inappropriate population |
| 2434 | The Role of S100B in Aerobic Training Efficacy in Older Adults with Mild Vascular Cognitive Impairment: Secondary Analysis of a Randomized Controlled Trial | Inappropriate population |
| 2435 | Commentary on: "Effects of Regular Physical Activity on the Cognitive Performance of Type 2 Diabetic Patients: A Systematic Review" by Podolski et al. (Metab Syndr Relat Disord 2017;15:481-493) | Inappropriate study design |
| 2436 | Ten years younger: Practice of chronic aerobic exercise improves attention and spatial memory functions in ageing | Inappropriate study design |
| 2437 | Intensity and Types of Physical Exercise in Relation to Dementia Risk Reduction in Community-Living Older Adults | Inappropriate study design |
| 2438 | Interventional programmes to improve cognition during healthy and pathological ageing: Cortical modulations and evidence for brain plasticity | Inappropriate population |
| 2439 | Dose-response relation between exercise duration and cognition | Inappropriate population |
| 2440 | Prescribing Aerobic Exercise Intensity without a Cardiopulmonary Exercise Test Post Stroke: Utility of the Six-Minute Walk Test | Inappropriate study design |
| 2441 | Effects of a 6-Week Aquatic Treadmill Exercise Program on Cardiorespiratory Fitness and Walking Endurance in Subacute Stroke Patients: A PILOT TRIAL | Inappropriate study design |
| 2442 | The Applicability of a High-Intensity Functional Exercise Program Among Older People With Dementia Living in Nursing Homes | Inappropriate population |
| 2443 | Within-session responses to high-intensity interval training in chronic stroke | Inappropriate study design |
| 2444 | A single session of moderate intensity walking increases brain-derived neurotrophic factor (BDNF) in the chronic post-stroke patients | Inappropriate study design |
| 2445 | Hemodynamic, Autonomic, Ventilatory, and Metabolic Alterations After Resistance Training in Patients With Coronary Artery Disease: A Randomized Controlled Trial | Inappropriate population |
| 2446 | Cognitive Effects of Intentional Weight Loss in Elderly Obese Individuals With Mild Cognitive Impairment | Inappropriate population |
| 2447 | The effect of Active Plus, a computer-tailored physical activity intervention, on cognitive functioning of elderly people with chronic illness(es) - study protocol for a randomized controlled trial | Inappropriate study design |
| 2448 | Decreases in daily physical activity predict acute decline in attention and executive function in heart failure | Inappropriate population |
| 2449 | SMART: physical activity and cerebral metabolism in older people: study protocol for a randomised controlled trial | Inappropriate study design |
| 2450 | Physical Fitness Assessment Across the Disability Spectrum in Persons With Multiple Sclerosis: A Comparison of Testing Modalities | Inappropriate population |
| 2451 | Multiple stages of information processing are modulated during acute bouts of exercise | Inappropriate study design |
| 2452 | Acute Exercise Effects Predict Training Change in Cognition and Connectivity | Inappropriate population |
| 2453 | Physical activity, fitness, and gray matter volume | Inappropriate population |
| 2454 | Severe Hypoxia Does Not Offset the Benefits of Exercise on Cognitive Function in Sedentary Young Women | Inappropriate population |
| 2455 | Feasibility of a multi-modal exercise program on cognition in older adults with Type 2 diabetes - a pilot randomised controlled trial | Inappropriate study design |
| 2456 | Prescribing physical activity as a preventive measure for middle-aged Australians with dementia risk factors | Inappropriate population |

| | | |
|---|---|---|
| 2457 | Rating of perceived exertion - a valid method for monitoring light to vigorous exercise intensity in individuals with subjective and mild cognitive impairment? | Inappropriate study design |
| 2458 | Physical exercise improves peripheral BDNF levels and cognitive functions in mild cognitive impairment elderly with different bdnf Val66Met genotypes | Inappropriate study design |
| 2459 | Obtaining Reliable Estimates of Ambulatory Physical Activity in People with Parkinson's Disease | Inappropriate study design |
| 2460 | Effects of resistance training and aerobic training on ambulation in chronic stroke | Inappropriate study design |
| 2461 | A review of chronic and acute physical activity participation on neuroelectric measures of brain health and cognition during childhood | Inappropriate population |
| 2462 | Ketogenic diet rescues cognition in ApoE4+ patient with mild Alzheimer's disease: A case study | Inappropriate study design |
| 2463 | Relations between Recent Past Leisure Activities with Risks of Dementia and Cognitive Functions after Stroke | Inappropriate outcome |
| 2464 | Acute Effects of a Short Bout of Physical Activity on Cognitive Function in Sport Students | Inappropriate study design |
| 2465 | Gait in ageing and associated dementias; its relationship with cognition | Inappropriate study design |
| 2466 | Carotid artery wall mechanics in young males with high cardiorespiratory fitness | Inappropriate outcome |
| 2467 | Effects of Aerobic Training on Cognition and Brain Glucose Metabolism in Subjects with Mild Cognitive Impairment | Inappropriate study design |
| 2468 | Diagnosing and managing vascular dementia | Inappropriate study design |
| 2469 | Guiding research and practice: a conceptual model for aerobic exercise training in Alzheimer's disease | Inappropriate study design |
| 2470 | Effect of a 24-month physical activity program on brain changes in older adults at risk of Alzheimer's disease: the AIBL active trial | Inappropriate population |
| 2471 | The use of aerobic exercise training in improving aerobic capacity in individuals with stroke: a meta-analysis | Inappropriate population |
| 2472 | Alzheimer disease alters the relationship of cardiorespiratory fitness with brain activity during the stroop task | Inappropriate study design |
| 2473 | The impact of exercise on the cognitive functioning of healthy older adults: a systematic review and meta-analysis | Inappropriate study design |
| 2474 | Cerebrospinal Fluid Amyloid Beta and Tau Concentrations Are Not Modulated by 16 Weeks of Moderate- to High-Intensity Physical Exercise in Patients with Alzheimer Disease | Inappropriate study design |
| 2475 | Aerobic exercise prior to task-specific training to improve poststroke motor function: A case series | Inappropriate study design |
| 2476 | Efficacy of exercise intervention programs on cognition in people suffering from multiple sclerosis, stroke and Parkinson's disease: A systematic review and meta-analysis of current evidence | Inappropriate study design |
| 2477 | Exercise at 65 and beyond | Inappropriate study design |
| 2478 | Effects of different types of physical activity on the cognitive functions and attention in older people: A randomized controlled study | Inappropriate population |
| 2479 | Differential effects of acute and regular physical exercise on cognition and affect | Inappropriate study design |
| 2480 | Determining Maximal Tolerable Aerobic Training Intensity in the Acute Phase after Stroke: a Novel Dose Ranging Trial Protocol | Inappropriate study design |
| 2481 | Total daily physical activity and the risk of AD and cognitive decline in older adults | Inappropriate study design |
| 2482 | A New Adaptive Home-based Exercise Technology among Older Adults Living in Nursing Home: A Pilot Study on Feasibility, Acceptability and Physical Performance | Inappropriate study design |
| 2483 | A randomized controlled trial of physical activity with individual goal-setting and volunteer mentors to overcome sedentary lifestyle in older adults at risk of cognitive decline: the INDIGO trial protocol | Inappropriate study design |
| 2484 | Aerobic training for older men with Alzheimer's disease: individual examples of progression | Inappropriate intervention |
| 2485 | Physical activity, inflammation, and volume of the aging brain | Inappropriate population |
| 2486 | Predicting Heart Rate at the Ventilatory Threshold for Aerobic Exercise Prescription in Persons With Chronic Stroke | Inappropriate population |
| 2487 | Comparison of Effect of Two Exercise Programs on Activities of Daily Living in Individuals with Dementia: A 9-Week Randomized, Controlled Trial | Inappropriate study design |
| 2488 | Exercise intensity of robot-assisted walking versus overground walking in nonambulatory stroke patients | Inappropriate population |
| 2489 | German recommendations for physical activity and physical activity promotion in adults with noncommunicable diseases | Inappropriate study design |

| | | |
|---|---|---|
| 2490 | The effects of different aerobic exercise intensities on serum serotonin concentrations and their association with Stroop task performance: a randomized controlled trial | Inappropriate study design |
| 2491 | The LIFE Cognition Study: design and baseline characteristics | Inappropriate population |
| 2492 | Cognitive dysfunction and physical activity after stroke: the Gothenburg cognitive stroke study in the elderly | Inappropriate study design |
| 2493 | Exercise and executive function in individuals with chronic stroke: a pilot study | Inappropriate population |
| 2494 | The effect of acute aerobic and resistance exercise on working memory | Inappropriate population |
| 2495 | Exercise in the prevention of stroke | Inappropriate study design |
| 2496 | Exercise capacity and cardiovascular adaptations to aerobic training early after stroke | Inappropriate population |
| 2497 | Aerobic fitness and skewness of frequency distribution of continuously measured heart rate in adults with brain injury | Inappropriate population |
| 2498 | Relationships among diet, physical activity and other lifestyle factors and debilitating diseases in the elderly | Inappropriate outcome |
| 2499 | Assessing for unique immunomodulatory and neuroplastic profiles of physical activity subtypes: a focus on psychiatric disorders | Inappropriate study design |
| 2500 | What is the role played by physical activity and exercise in the frailty syndrome? Perspectives for future research | Inappropriate study design |
| 2501 | APOEε4 impacts up-regulation of brain-derived neurotrophic factor after a six-month stretch and aerobic exercise intervention in mild cognitively impaired elderly African Americans: A pilot study | Inappropriate population |
| 2502 | A community-based approach to trials of aerobic exercise in aging and Alzheimer's disease | Inappropriate population |
| 2503 | Exercise-induced changes in cardiovascular function after stroke: a randomized controlled trial | Inappropriate study design |
| 2504 | Patient characteristics that influence enrollment and attendance in aerobic exercise early after stroke | Inappropriate study design |
| 2505 | A single bout of walking exercise enhances endogenous fibrinolysis in stroke patients | Inappropriate population |
| 2506 | Is the effect of aerobic exercise on cognition a placebo effect? | Inappropriate population |
| 2507 | Efficacy of lifestyle and psychosocial interventions in reducing cognitive decline in older people: Systematic review | Inappropriate population |
| 2508 | Blood pressure changes following aerobic exercise in Caucasian and Chinese descendants | Inappropriate study design |
| 2509 | Facilitating aerobic exercise training in older adults with Alzheimer's disease | Inappropriate intervention |
| 2510 | Effects of physical exercise on cognitive function of older adults with mild cognitive impairment: A systematic review and meta-analysis | Inappropriate study design |
| 2511 | Efficacy of Feedback-Controlled Robotics-Assisted Treadmill Exercise to Improve Cardiovascular Fitness Early After Stroke: A Randomized Controlled Pilot Trial | Inappropriate study design |
| 2512 | Effects of physical activity on health status in older adults. II. Intervention studies | Inappropriate study design |
| 2513 | Physical activity and dementia risk in the elderly: findings from a prospective Italian study | Inappropriate outcome |
| 2514 | Evaluation of exercise on individuals with dementia and their carers: a randomised controlled trial | Inappropriate study design |
| 2515 | Effects of aerobic fitness on cognitive motor interference during self-paced treadmill walking in older adults | Inappropriate population |
| 2516 | Resting and post exercise arterial-ventricular coupling in endurance-trained men and women | Inappropriate study design |
| 2517 | The role of aerobic fitness and exercise intensity on endurance performance in uncompensable heat stress conditions | Inappropriate population |
| 2518 | Increased self-efficacy: the experience of high-intensity exercise of nursing home residents with dementia - a qualitative study | Inappropriate study design |
| 2519 | Aerobic exercise training in stroke survivors | Inappropriate outcome |
| 2520 | Acute effect of a single bout of aerobic exercise on vascular and baroreflex function of young males with a family history of hypertension | Inappropriate population |
| 2521 | Heart rate variability in stroke patients submitted to an acute bout of aerobic exercise | Inappropriate outcome |
| 2522 | The impact of exercise on patients with dementia: A 2-year follow-up | Inappropriate study design |
| 2523 | Ambulatory activity intensity profiles, fitness, and fatigue in chronic stroke | Inappropriate intervention |
| 2524 | Effect of aerobic and resistance exercise on central hemodynamic responses in severe chronic heart failure | Inappropriate study design |

| | | |
|---|---|---|
| 2525 | Sensor-derived physical activity parameters can predict future falls in people with dementia | Inappropriate study design |
| 2526 | Integrating aerobic training within subacute stroke rehabilitation: a feasibility study | Inappropriate population |
| 2527 | Treadmill aerobic training improves glucose tolerance and indices of insulin sensitivity in disabled stroke survivors: a preliminary report | Inappropriate population |
| 2528 | Multiple roads lead to Rome: combined high-intensity aerobic and strength training vs. gross motor activities leads to equivalent improvement in executive functions in a cohort of healthy older adults | Inappropriate population |
| 2529 | Elevated aerobic fitness sustained throughout the adult lifespan is associated with improved cerebral hemodynamics | Inappropriate study design |
| 2530 | Treadmill aerobic exercise training reduces the energy expenditure and cardiovascular demands of hemiparetic gait in chronic stroke patients. A preliminary report | Inappropriate population |
| 2531 | A meta-regression to examine the relationship between aerobic fitness and cognitive performance | Inappropriate study design |
| 2532 | Absolute versus relative intensity of physical activity in a dose-response context | Inappropriate population |
| 2533 | Oxygen uptake kinetics: associations with ambulatory activity and physical functional performance in stroke survivors | Inappropriate study design |
| 2534 | Modified high-intensity interval training increases peak cardiac power output in patients with heart failure | Inappropriate population |
| 2535 | Findings from a strength-based moderate-intensity exercise interventions for individuals with dementia (innovative practice) | Inappropriate study design |
| 2536 | Sex differences in the effects of exercise on cognition post-stroke: Secondary analysis of a randomized controlled trial | Inappropriate study design |
| 2537 | Right ventricular myocardial responses to progressive exercise in young adult males | Inappropriate outcome |
| 2538 | A single weekly bout of exercise may reduce cardiovascular mortality: how little pain for cardiac gain? 'The HUNT study, Norway' | Inappropriate population |
| 2539 | Challenges with cost-utility analyses of behavioural interventions among older adults at risk for dementia | Inappropriate study design |
| 2540 | Are patients 1 year post-stroke active enough to improve their physical health? | Inappropriate population |
| 2541 | Treadmill exercise activates subcortical neural networks and improves walking after stroke: a randomized controlled trial | Inappropriate intervention |
| 2542 | Post-exercise hypotension and cardiovascular responses to moderate orthostatic stress in endurance-trained males | Inappropriate study design |
| 2543 | Effect of progressive resistance training on muscle performance after chronic stroke | Inappropriate study design |
| 2544 | Physical activity guidelines for older adults | Inappropriate study design |
| 2545 | Pilot safety and feasibility study of treadmill aerobic exercise in Parkinson disease with gait impairment | Inappropriate population |
| 2546 | Is the magnitude of acute post-exercise hypotension mediated by exercise intensity or total work done? | Inappropriate study design |
| 2547 | Factors associated with change in aerobic capacity following an exercise program for individuals with stroke | Inappropriate population |
| 2548 | The brain-in-motion study: effect of a 6-month aerobic exercise intervention on cerebrovascular regulation and cognitive function in older adults | Inappropriate population |
| 2549 | The benefits of staying active in old age: physical activity counteracts the negative influence of PICALM, BIN1, and CLU risk alleles on episodic memory functioning | Inappropriate study design |
| 2550 | Aerobic exercise attenuates an exaggerated exercise blood pressure response in normotensive young adult African-American men | Inappropriate study design |
| 2551 | Late-life social activity and cognitive decline in old age | Inappropriate study design |
| 2552 | Survey of fitness facilities for individuals post-stroke in the Greater Toronto Area | Inappropriate study design |
| 2553 | Effect of intensive outpatient physical training on gait performance and cardiovascular health in people with hemiparesis after stroke | Inappropriate intervention |
| 2554 | Protocol for a randomized controlled trial evaluating the effect of physical activity on delaying the progression of white matter changes on MRI in older adults with memory complaints and mild cognitive impairment: the AIBL Active trial | Inappropriate study design |
| 2555 | Exercise-induced changes of the upper extremity in chronic stroke survivors | Inappropriate population |
| 2556 | Comparison of effect of aerobic cycle training and progressive resistance training on walking ability after stroke: a randomized sham exercise-controlled study | Inappropriate study design |

| | | |
|---|---|---|
| 2557 | Responses to exercise differ for chronic fatigue syndrome patients with fibromyalgia | Inappropriate population |
| 2558 | Effects of moderate-intensity aerobic cycling and swim exercise on post-exertional blood pressure in healthy young untrained and triathlon-trained men and women | Inappropriate study design |
| 2559 | Protocol for Project FACT: a randomised controlled trial on the effect of a walking program and vitamin B supplementation on the rate of cognitive decline and psychosocial wellbeing in older adults with mild cognitive impairment [ISRCTN19227688 | Inappropriate population |
| 2560 | Can individuals participating in cardiac rehabilitation achieve recommended exercise training levels following stroke? | Inappropriate population |
| 2561 | Evolution of cardiorespiratory fitness after stroke: a 1-year follow-up study. Influence of prestroke patients' characteristics and stroke-related factors | Inappropriate intervention |
| 2562 | Chronic psychological distress and risk of Alzheimer's disease in old age | Inappropriate study design |
| 2563 | A remotely supervised home-based aerobic exercise programme is feasible for patients with Parkinson's disease: results of a small randomised feasibility trial | Inappropriate study design |
| 2564 | Maximal cardiorespiratory fitness testing in individuals with chronic stroke with cognitive impairment: practice test effects and test-retest reliability | Inappropriate study design |
| 2565 | Effects of early exercise engagement on vascular risk in patients with transient ischemic attack and nondisabling stroke | Inappropriate study design |
| 2566 | The relationship between fatigue, aerobic fitness, and motor control in people with chronic stroke: a pilot study | Inappropriate study design |
| 2567 | Aerobic exercise to improve executive function in Parkinson disease: a case series | Inappropriate outcome |
| 2568 | A preliminary study to examine the effects of aerobic and therapeutic (nonaerobic) exercise on cardiorespiratory fitness and coronary risk reduction in stroke survivors | Inappropriate study design |
| 2569 | Intensive aerobic cycling training with lower limb weights in Chinese patients with chronic stroke: discordance between improved cardiovascular fitness and walking ability | Inappropriate study design |
| 2570 | Translating an Evidence-based Multicomponent Intervention for Older Adults With Dementia and Caregivers | Inappropriate population |
| 2571 | Postexercise hypotension. Key features, mechanisms, and clinical significance | Inappropriate study design |
| 2572 | Postexercise hypotension in an endurance-trained population of men and women following high-intensity interval and steady-state cycling | Inappropriate population |
| 2573 | Augmented Performance Feedback during Robotic Gait Therapy Results in Moderate Intensity Cardiovascular Exercise in Subacute Stroke | Inappropriate population |
| 2574 | Conference on "Multidisciplinary approaches to nutritional problems". Symposium on "Performance, exercise and health". Exercise in improving health v. performance | Inappropriate outcome |
| 2575 | Haemodynamic changes induced by submaximal exercise before a dive and its consequences on bubble formation | Inappropriate outcome |
| 2576 | Acute and chronic hemodynamic effects of nicardipine hydrochloride in patients with heart failure | Inappropriate population |
| 2577 | Water-based exercise for cardiovascular fitness in people with chronic stroke: a randomized controlled trial | Inappropriate study design |
| 2578 | No exercise-induced increase in serum BDNF after cycling near a major traffic road | Inappropriate population |
| 2579 | Silent cardiac dysfunction and exercise intolerance in HIV+ men receiving combined antiretroviral therapies | Inappropriate study design |
| 2580 | Impaired leg vasodilatory function after stroke: adaptations with treadmill exercise training | Inappropriate population |
| 2581 | Acute exercise reduces vascular reactivity to mental challenge in offspring of hypertensive families | Inappropriate study design |
| 2582 | Determinants of improvement in walking capacity among individuals with chronic stroke following a multi-dimensional exercise program | Inappropriate study design |
| 2583 | Does fitness level modulate the cardiovascular hemodynamic response to exercise? | Inappropriate study design |
| 2584 | Attenuation of exaggerated exercise blood pressure response in African-American women by regular aerobic physical activity | Inappropriate population |
| 2585 | Cardiovascular hemodynamics with increasing exercise intensities in postmenopausal women | Inappropriate study design |
| 2586 | Effect of exercise training on VO2peak and left ventricular systolic function in recent cardiac transplant recipients | Inappropriate population |

| | | |
|---|---|---|
| 2587 | Hemodynamic and hormonal responses to lower body negative pressure in men with varying profiles of strength and aerobic power | Inappropriate study design |
| 2588 | Cardiorespiratory responses to low-intensity heart rate-controlled exercise in female subjects | Inappropriate study design |
| 2589 | Arterial compliance increases after moderate-intensity cycling | Inappropriate study design |
| 2590 | Effects of aerobic fitness training on human cortical function: a proposal | Inappropriate study design |
| 2591 | High-intensity cycling exercise after a stroke: a single case study | Inappropriate study design |
| 2592 | Aerobic interval training reduces vascular resistances during submaximal exercise in obese metabolic syndrome individuals | Inappropriate population |
| 2593 | Association of Chronic Periodontitis on Alzheimer's Disease or Vascular Dementia | Inappropriate study design |
| 2594 | Physical activity correlates in people with mild cognitive impairment: findings from six low- and middle-income countries | Inappropriate study design |
| 2595 | Physical fitness interventions for nonambulatory stroke survivors: A mixed-methods systematic review and meta-analysis | Inappropriate study design |
| 2596 | Adherence to a Long-Term Physical Activity and Exercise Program After Stroke Applied in a Randomized Controlled Trial | Inappropriate population |
| 2597 | The effects of poststroke aerobic exercise on neuroplasticity: a systematic review of animal and clinical studies | Inappropriate study design |
| 2598 | Evidence-based risk assessment and recommendations for physical activity clearance: stroke and spinal cord injury | Inappropriate study design |
| 2599 | Applicability and effects of physical exercise on physical and cognitive functions and activities of daily living among people with dementia: a systematic review | Inappropriate population |
| 2600 | Role of aerobic fitness and aging on cerebral white matter integrity | Inappropriate study design |
| 2601 | Conducting Maximal and Submaximal Endurance Exercise Testing to Measure Physiological and Biological Responses to Acute Exercise in Humans | Inappropriate population |
| 2602 | Determining Safe Participation in Aerobic Exercise Early After Stroke Through a Graded Submaximal Exercise Test | Inappropriate study design |
| 2603 | Subclinical effects of aerobic training in urban environment | Inappropriate study design |
| 2604 | Effect of a bout of leg cycling with electrical stimulation on reduction of hypertonia in patients with stroke | Inappropriate population |
| 2605 | Association of physical fitness and fatness with cognitive function in women with fibromyalgia | Inappropriate outcome |
| 2606 | Physical Fitness Training in Patients with Subacute Stroke (PHYS-STROKE): multicentre, randomised controlled, endpoint blinded trial | Inappropriate study design |
| 2607 | A High-Intensity Exercise Boot Camp for Persons With Parkinson Disease: A Phase II, Pragmatic, Randomized Clinical Trial of Feasibility, Safety, Signal of Efficacy, and Disease Mechanisms | Inappropriate study design |
| 2608 | Dementia Patients Are More Sedentary and Less Physically Active than Age- and Sex-Matched Cognitively Healthy Older Adults | Inappropriate study design |
| 2609 | Overview of resistance training, diet, hormone replacement and nutritional supplements on age-related sarcopenia--a minireview | Inappropriate study design |
| 2610 | The Impact of Dietary Factors on Indices of Chronic Disease in Older People: A Systematic Review | Inappropriate population |
| 2611 | Acute increases in brain-derived neurotrophic factor following high or moderate-intensity exercise is accompanied with better cognition performance in obese adults | Inappropriate population |
| 2612 | Higher Treadmill Training Intensity to Address Functional Aerobic Impairment after Stroke | Inappropriate population |
| 2613 | Shiatsu as an adjuvant therapy for depression in patients with Alzheimer's disease: A pilot study | Inappropriate population |
| 2614 | Differential effects of differing intensities of acute exercise on speed and accuracy of cognition: a meta-analytical investigation | Inappropriate population |
| 2615 | Electromyographic Comparison of Elastic Resistance and Machine Exercises for High-Intensity Strength Training in Patients With Chronic Stroke | Inappropriate study design |
| 2616 | Step-rate threshold for physical activity intensity in Parkinson's disease | Inappropriate study design |
| 2617 | Combined Aerobic Exercise and Task Practice Improve Health-Related Quality of Life Poststroke: A Preliminary Analysis | Inappropriate study design |
| 2618 | Blood pressure and hemodynamic responses after exercise in older hypertensives | Inappropriate study design |
| 2619 | Aerobic fitness and the cardiovascular response to stress | Inappropriate population |
| 2620 | The association between mastication and mild cognitive impairment in Korean adults | Inappropriate population |

| | | |
|---|---|---|
| 2621 | Effects of a primary rehabilitation programme on arterial vascular adaptations in an individual with paraplegia | Inappropriate study design |
| 2622 | Exertion fatigue and chronic fatigue are two distinct constructs in people post-stroke | Inappropriate study design |
| 2623 | Postexercise hypotension and hemodynamics: the role of exercise intensity | Inappropriate population |
| 2624 | Short-term and long-term effects of a progressive resistance and balance exercise program in individuals with chronic stroke: a randomized controlled trial | Inappropriate outcome |
| 2625 | Circulatory response evoked by a 3 s bout of dynamic leg exercise in humans | Inappropriate study design |
| 2626 | Obesity Modifies the Effect of Fitness on Heart Rate Indices during Exercise Stress Testing in Asymptomatic Individuals | Inappropriate population |
| 2627 | Preventive effects of multiple domain interventions on lifestyle and risk factor changes in stroke survivors: Evidence from a two-year randomized trial | Inappropriate study design |
| 2628 | Risk factors for dementia in highly educated elderly people in Tianjin, China | Inappropriate population |
| 2629 | Group-based exercise and cognitive-physical training in older adults with self-reported cognitive complaints: The Multiple-Modality, Mind-Motor (M4) study protocol | Inappropriate population |
| 2630 | Structured physical exercise improves neuropsychiatric symptoms in acute dementia care: a hospital-based RCT | Inappropriate study design |
| 2631 | Characterizing differences in mortality at the low end of the fitness spectrum in individuals with cardiovascular disease | Inappropriate study design |
| 2632 | Experimental Effects of Acute Exercise on Episodic Memory Function: Considerations for the Timing of Exercise | Inappropriate study design |
| 2633 | Measuring the impact of exercise on cognitive aging: methodological issues | Inappropriate study design |
| 2634 | Exercise and medication effects on persons with Parkinson disease across the domains of disability: a randomized clinical trial | Inappropriate study design |
| 2635 | Two-week virtual reality training for dementia: Single case feasibility study | Inappropriate study design |
| 2636 | Cardiovascular Stress Induced by Whole-Body Vibration Exercise in Individuals With Chronic Stroke | Inappropriate population |
| 2637 | A 1-year randomized controlled trial comparing mind body exercise (Tai Chi) with stretching and toning exercise on cognitive function in older Chinese adults at risk of cognitive decline | Inappropriate population |
| 2638 | A Pilot Study: The Beneficial Effects of Combined Statin-exercise Therapy on Cognitive Function in Patients with Coronary Artery Disease and Mild Cognitive Decline | Inappropriate population |
| 2639 | Physical activity and onset of acute ischemic stroke: the stroke onset study | Inappropriate study design |
| 2640 | Assessment of compliance with prescribed activity by hemiplegic stroke patients after an exercise programme and physical activity education | Inappropriate study design |
| 2641 | Impaired Hyperemic Response to Exercise Post Stroke | Inappropriate study design |
| 2642 | Monitoring exercise delivery to increase participation adherence in older adults with Alzheimer's disease | Inappropriate outcome |
| 2643 | Acute exhaustive rowing exercise reduces skin microvascular dilator function in young adult rowing athletes | Inappropriate study design |
| 2644 | Poststroke Physical Activity Levels No Higher in Rehabilitation than in the Acute Hospital | Inappropriate study design |
| 2645 | Physical activity and exercise as countermeasures to physical frailty and sarcopenia | Inappropriate study design |
| 2646 | Family reports of medically impaired drivers in Missouri: cognitive concerns and licensing outcomes | Inappropriate population |
| 2647 | Relationship between maximal exercise capacity and walking capacity in adult hemiplegic stroke patients | Inappropriate study design |
| 2648 | Aerobic exercise in subacute stroke improves cardiovascular health and physical performance | Inappropriate population |
| 2649 | Interventions to Improve Gait in Older Adults with Cognitive Impairment: A Systematic Review | Inappropriate intervention |
| 2650 | Effects of Combined Physical and Cognitive Exercises on Cognition and Mobility in Patients With Mild Cognitive Impairment: A Randomized Clinical Trial | Inappropriate outcome |
| 2651 | Effect of long-term high intensity aerobic training on left ventricular volume during maximal upright exercise | Inappropriate study design |
| 2652 | The Effects of Functional Training, Bicycle Exercise, and Exergaming on Walking Capacity of Elderly Patients With Parkinson Disease: A Pilot Randomized Controlled Single-blinded Trial | Inappropriate study design |
| 2653 | Ventriculoarterial coupling during exercise in normal human subjects | Inappropriate outcome |

| | | |
|---|---|---|
| 2654 | Does clinical rehabilitation impose sufficient cardiorespiratory strain to improve aerobic fitness? | Inappropriate population |
| 2655 | Effects of exercise intervention on patients with stroke with prior coronary artery disease: aerobic capacity, functional ability, and lipid profile: a pilot study | Inappropriate intervention |
| 2656 | Aerobic exercise moderates the effect of heavy alcohol consumption on white matter damage | Inappropriate population |
| 2657 | Promoting physical activity after stroke via self-management: a feasibility study | Inappropriate study design |
| 2658 | Review of effects of physical activity on strength, balance, mobility and ADL performance in elderly subjects with dementia | Inappropriate study design |
| 2659 | Multicomponent exercise and the hallmarks of frailty: Considerations on cognitive impairment and acute hospitalization | Inappropriate study design |
| 2660 | Evaluation and facilitation of intervention fidelity in community exercise programs through an adaptation of the TIDier framework | Inappropriate study design |
| 2661 | Predictors of gait velocity among community-dwelling stroke survivors | Inappropriate population |
| 2662 | Matched dose interval and continuous exercise training induce similar cardiorespiratory and metabolic adaptations in patients with heart failure | Inappropriate study design |
| 2663 | Hippocampal Response to a 24-Month Physical Activity Intervention in Sedentary Older Adults | Inappropriate study design |
| 2664 | The Feasibility of Remotely Delivered Exercise Session in Adults With Alzheimer's Disease and Their Caregivers | Inappropriate study design |
| 2665 | Feasibility of Three Novel Forms of Passive Exercise in a Multisensory Environment in Vulnerable Institutionalized Older Adults with Dementia | Inappropriate population |
| 2666 | Energy expenditure in chronic stroke patients playing Wii Sports: a pilot study | Inappropriate intervention |
| 2667 | Cognitive decline and the default American lifestyle | Inappropriate study design |
| 2668 | Shall We Dance? Dancing Modulates Executive Functions and Spatial Memory | Inappropriate study design |
| 2669 | Understanding physical activity in Veterans with Parkinson disease: A mixed-methods approach | Inappropriate population |
| 2670 | Exercise responses during functional electrical stimulation cycling in individuals with spinal cord injury | Inappropriate study design |
| 2671 | Cardiac MRI: a new gold standard for ventricular volume quantification during high-intensity exercise | Inappropriate study design |
| 2672 | Building a Bridge to the Community: An Integrated Knowledge Translation Approach to Improving Participation in Community-Based Exercise for People After Stroke | Inappropriate study design |
| 2673 | Cardiovascular Drift during Training for Fitness in Patients with Metabolic Syndrome | Inappropriate population |
| 2674 | Exercise as a potential modulator of inflammation in patients with Alzheimer's disease measured in cerebrospinal fluid and plasma | Inappropriate study design |
| 2675 | Near-maximal exercise responses in health and disease in middle-aged Chinese men | Inappropriate study design |
| 2676 | Effects of acute and chronic ibopamine administration on resting and exercise hemodynamics, plasma catecholamines and functional capacity of patients with chronic congestive heart failure | Inappropriate population |
| 2677 | Blood volume and its relation to peak O(2) consumption and physical activity in patients with chronic fatigue | Inappropriate population |
| 2678 | The physical performance test predicts aerobic capacity sufficient for independence in early-stage Alzheimer disease | Inappropriate population |
| 2679 | Frequent, short bouts of light-intensity exercises while standing decreases systolic blood pressure: Breaking Up Sitting Time after Stroke (BUST-Stroke) trial | Inappropriate population |
| 2680 | The effects of aerobic fitness and beta1-adrenergic receptor blockade on cardiac work during dynamic exercise | Inappropriate study design |
| 2681 | The effects of exercise under hypoxia on cognitive function | Inappropriate study design |
| 2682 | Factors Influencing the Delivery of Intensive Rehabilitation in Stroke: Patient Perceptions Versus Rehabilitation Therapist Perceptions | Inappropriate population |
| 2683 | Stroke volume during recovery from upright bicycle exercise | Inappropriate population |
| 2684 | Improving Recovery and Outcomes Every Day after the ICU (IMPROVE): study protocol for a randomized controlled trial | Inappropriate study design |
| 2685 | Improving recruitment, retention, and adherence to 6-month cycling in Alzheimer's disease | Inappropriate population |
| 2686 | Aerobic exercise improves quality of life, psychological well-being and systemic inflammation in subjects with Alzheimer's disease | Inappropriate intervention |
| 2687 | Effect of acute and long-term aerobic exercise on arterial stiffness in the elderly | Inappropriate outcome |

| | | |
|---|---|---|
| 2688 | Accelerometer-assessed light physical activity is protective of future cognitive ability: A longitudinal study among community dwelling older adults | Inappropriate study design |
| 2689 | The effect of resistance training on cognitive function in the older adults: a systematic review of randomized clinical trials | Inappropriate population |
| 2690 | Aerobic physical exercise as a possible treatment for neurocognitive dysfunction in bipolar disorder | Inappropriate study design |
| 2691 | Social resources and cognitive decline in a population of older African Americans and whites | Inappropriate population |
| 2692 | Acute physiological responses in healthy men during whole-body vibration | Inappropriate study design |
| 2693 | Associations of Chronic Kidney Disease Markers with Cognitive Function: A 12-Year Follow-Up Study | Inappropriate study design |
| 2694 | Factors Affecting Attendance at an Adapted Cardiac Rehabilitation Exercise Program for Individuals with Mobility Deficits Poststroke | Inappropriate study design |
| 2695 | The Norwegian Cognitive impairment after stroke study (Nor-COAST): study protocol of a multicentre, prospective cohort study | Inappropriate study design |
| 2696 | A review of multidomain interventions to support healthy cognitive ageing | Inappropriate study design |
| 2697 | Effects of the dietary approaches to stop hypertension diet, exercise, and caloric restriction on neurocognition in overweight adults with high blood pressure | Inappropriate study design |
| 2698 | "Task-oriented" exercise improves hamstring strength and spastic reflexes in chronic stroke patients | Inappropriate study design |
| 2699 | Haemodynamic effects of hydralazine at rest and during exercise in patients with chronic heart failure | Inappropriate study design |
| 2700 | Associations of arterial stiffness and cognitive function with physical fitness in patients with chronic stroke | Inappropriate study design |
| 2701 | Cost-effectiveness of a randomised trial of physical activity in Alzheimer's disease: a secondary analysis exploring patient and proxy-reported health-related quality of life measures in Denmark | Inappropriate study design |
| 2702 | Study of Mental Activity and Regular Training (SMART) in at risk individuals: a randomised double blind, sham controlled, longitudinal trial | Inappropriate study design |
| 2703 | Quantification of exercise capability and evaluation of physical capacity in man | Inappropriate study design |
| 2704 | Hand strengthening exercises in chronic stroke patients: Dose-response evaluation using electromyography | Inappropriate study design |
| 2705 | A randomised controlled trial investigating the effects of Mediterranean diet and aerobic exercise on cognition in cognitively healthy older people living independently within aged care facilities: the Lifestyle Intervention in Independent Living Aged Care (LIILAC) study protocol [ACTRN12614001133628 | Inappropriate study design |
| 2706 | Effects of a Single Session of High Intensity Interval Treadmill Training on Corticomotor Excitability following Stroke: Implications for Therapy | Inappropriate population |
| 2707 | Respiratory training as strategy to prevent cognitive decline in aging: a randomized controlled trial | Inappropriate study design |
| 2708 | Anaerobic threshold measurements of elite oarsmen | Inappropriate study design |
| 2709 | Recommended physical activity and all cause and cause specific mortality in US adults: prospective cohort study | Inappropriate population |
| 2710 | Technology-enhanced multi-domain at home continuum of care program with respect to usual care for people with cognitive impairment: the Ability-TelerehABILITation study protocol for a randomized controlled trial | Inappropriate population |
| 2711 | High-intensity aerobic interval training for patients 3-9 months after stroke: a feasibility study | Inappropriate population |
| 2712 | Training effects of wheelchair dance on aerobic fitness in bedridden individuals with severe athetospastic cerebral palsy rated to GMFCS level V | Inappropriate study design |
| 2713 | Dose-response relationship between exercise and cognitive function in older adults with and without cognitive impairment: A systematic review and meta-analysis | Inappropriate study design |
| 2714 | Daily Total Physical Activity and Incident Stroke: The Japan Public Health Center-Based Prospective Study | Inappropriate study design |
| 2715 | Relationship Between Dual-Task Gait Speed and Walking Activity Poststroke | Inappropriate study design |
| 2716 | Association of physical activity on the functional connectivity of the hippocampal-orbitofrontal pathway | Inappropriate study design |
| 2717 | Effects of combining exercise with long-chain polyunsaturated fatty acid supplementation on cognitive function in the elderly: a randomised controlled trial | Inappropriate study design |
| 2718 | The feasibility and the effects of cycloergometer interval-training on aerobic capacity and walking performance after stroke. Preliminary study | Inappropriate study design |

| | | |
|---|---|---|
| 2719 | Effect of physical exercise on cognitive function and brain measures after chemotherapy in patients with breast cancer (PAM study): protocol of a randomised controlled trial | Inappropriate study design |
| 2720 | Approach to the problems of the aged | Inappropriate outcome |
| 2721 | Associations between physical function, dual-task performance and cognition in patients with mild Alzheimer's disease | Inappropriate outcome |
| 2722 | Intermittent maximal exercise improves attentional performance only in physically active students | Inappropriate study design |
| 2723 | Effect of Whole-Body Vibration on Neuromuscular Activation of Leg Muscles During Dynamic Exercises in Individuals With Stroke | Inappropriate study design |
| 2724 | Evidence of a secondary hypervolemia in trained man following acute high intensity exercise | Inappropriate population |
| 2725 | Accelerometry: A feasible method to monitor physical activity during sub-acute rehabilitation of persons with stroke | Inappropriate study design |
| 2726 | Physical performance is associated with working memory in older people with mild to severe cognitive impairment | Inappropriate population |
| 2727 | Frequent and intensive physical activity reduces risk of cardiovascular events in type 1 diabetes | Inappropriate study design |
| 2728 | The association between bouted and non-bouted physical activity on retinopathy prevalence | Inappropriate study design |
| 2729 | Brain-derived neurotrophic factor mediates cognitive improvements following acute exercise | Inappropriate study design |
| 2730 | Effect of Physical Activity on the Progression of Alzheimer's Disease: The Clinical Research Center for Dementia of South Korea Study | Inappropriate study design |
| 2731 | Using Implementation Frameworks to Provide Proactive Physical Therapy for People With Parkinson Disease: Case Report | Inappropriate outcome |
| 2732 | Effects of Vibration Intensity, Exercise, and Motor Impairment on Leg Muscle Activity Induced by Whole-Body Vibration in People With Stroke | Inappropriate study design |
| 2733 | Does Replacing Sodium Excreted in Sweat Attenuate the Health Benefits of Physical Activity? | Inappropriate study design |
| 2734 | Community exercise: a vital component to healthy aging | Inappropriate outcome |
| 2735 | Effects of exercise training on cognition in chronic obstructive pulmonary disease: A systematic review | Inappropriate population |
| 2736 | Level of physical activity in men and women with chronic stroke | Inappropriate population |
| 2737 | Rhythm experience and Africana culture trial (REACT!): A culturally salient intervention to promote neurocognitive health, mood, and well-being in older African Americans | Inappropriate population |
| 2738 | Muscle Oxygenation Responses to Low-intensity Steady Rate Concentric and Eccentric Cycling | Inappropriate outcome |
| 2739 | Combined short-arm centrifuge and aerobic exercise training improves cardiovascular function and physical working capacity in humans | Inappropriate population |
| 2740 | Acute and chronic hemodynamic effects of drugs with different actions on adrenergic receptors: a comparison between alpha blockers and different types of beta blockers with and without vasodilating effect | Inappropriate study design |
| 2741 | Approach to assessing fitness to drive in patients with cardiac and cognitive conditions | Inappropriate population |
| 2742 | Cardiovascular dynamics after acute and long-term alpha- and beta-adrenoceptor blockade at rest, supine and standing, and during exercise | Inappropriate population |
| 2743 | Physical activity mediates the association between striatal dopamine transporter availability and cognition in Parkinson's disease | Inappropriate population |
| 2744 | Genotype, resilience and function and physical activity post hip fracture | Inappropriate study design |
| 2745 | Plantar flexion training primes peripheral arterial disease patients for improvements in cardiac function | Inappropriate study design |
| 2746 | Acute Modulation of Cortical Glutamate and GABA Content by Physical Activity | Inappropriate study design |
| 2747 | Effectiveness of High-Intensity Interval Training for Fitness and Mobility Post Stroke: A Systematic Review | Inappropriate study design |
| 2748 | Post-exercise cognitive testing to assess persisting alterations in athletes with a history of concussion | Inappropriate study design |
| 2749 | The intestinal microbiome and its relevance for functionality in older persons | Inappropriate outcome |
| 2750 | Acute effects of exercise on cerebrovascular response and cognitive performance in individuals with stable coronary heart disease | Inappropriate study design |
| 2751 | Personalized motor-cognitive exergame training in chronic stroke patients—A feasibility study | Inappropriate intervention |

| | | |
|---|---|---|
| 2752 | Feasibility of virtual reality augmented cycling for health promotion of people poststroke | Inappropriate population |
| 2753 | Physical and cognitive profiles in motoric cognitive risk syndrome in an older population from Southern Italy | Inappropriate population |
| 2754 | Comparison between the effects of continuous and intermittent light-intensity aerobic dance exercise on mood and executive functions in older adults | Inappropriate population |
| 2755 | Does treadmill exercise improve performance of cognitive or upper-extremity tasks in people with chronic stroke? A randomized cross-over trial | Inappropriate outcome |
| 2756 | Acute exercise increases circulating IGF-1 in Alzheimer’s disease patients, but not in older adults without dementia | Inappropriate study design |
| 2757 | Acute exercise improves cognition in the depressed elderly: the effect of dual-tasks | Inappropriate population |
| 2758 | Health-related quality of life after first-ever acute ischemic stroke: Associations with cardiovascular health metrics | Inappropriate study design |
| 2759 | Acute effects of different exercise intensities on executive function and oculomotor performance in middle-aged and older adults: Moderate-intensity continuous exercise vs High-intensity interval exercise | Inappropriate population |
| 2760 | Effects of Progressive Resistance Training on Cognition and IGF-1 Levels in Elder Women Who Live in Areas with High Air Pollution | Inappropriate study design |
| 2761 | Exercise intensity-specific changes to cerebral blood velocity do not modulate a postexercise executive function benefit | Inappropriate study design |
| 2762 | Effects of a single physical or mindfulness intervention on mood, attention, and executive functions: Results from two randomized controlled studies in university classes | Inappropriate population |
| 2763 | A multisession evaluation of an adaptive competitive arm rehabilitation game | Inappropriate study design |
| 2764 | A longitudinal randomized controlled trial protocol to evaluate the effects of Wuqinxi on dynamic functional connectivity in Parkinson’s disease patients | Inappropriate study design |
| 2765 | A multimodal risk network predicts executive function trajectories in non-demented aging | Inappropriate study design |
| 2766 | Physical Activity and Risk of Coronary Heart Disease and Stroke in Older Adults: The Cardiovascular Health Study | Inappropriate population |
| 2767 | Test-retest reliability and four-week changes in cardiopulmonary fitness in stroke patients: evaluation using a robotics-assisted tilt table | Inappropriate population |
| 2768 | Should strengthening bonds be a public health priority? A population-based analysis | Inappropriate study design |
| 2769 | Psychological interventions for reducing fear avoidance beliefs among people with chronic back pain | Inappropriate study design |
| 2770 | Physical activity effects on the individual alpha peak frequency of older adults with and without genetic risk factors for Alzheimer's Disease: A MEG study | Inappropriate study design |
| 2771 | The positive influence of sense of control on physical, behavioral, and psychosocial health in older adults: An outcome-wide approach | Inappropriate study design |
| 2772 | Effects of short-term exercise and exercise training on cognitive function among patients with cardiac disease | Inappropriate population |
| 2773 | Validating accelerometry as a measure of physical activity and energy expenditure in chronic stroke | Inappropriate study design |
| 2774 | Accuracy and precision of wrist-worn actigraphy for measuring steps taken during over-ground and treadmill walking in adults with Parkinson's disease | Inappropriate population |
| 2775 | The effect of acute exercise for reducing cognitive alterations associated with individuals high in anxiety | Inappropriate population |
| 2776 | Effects of 2 Years of Exercise on Gait Impairment in People With Parkinson Disease: The PRET-PD Randomized Trial | Inappropriate population |
| 2777 | Golf as a physical activity to improve walking speed and cognition in older adults: A non-randomized, pre-post, pilot study | Inappropriate population |
| 2778 | Physical activity on prescription at the time of stroke or transient ischemic attack diagnosis – From a patient perspective | Inappropriate study design |
| 2779 | Effects of treadmill exercise on transcranial magnetic stimulation-induced excitability to quadriceps after stroke | Inappropriate study design |
| 2780 | Effects of physical and cognitive training on gait speed and cognition in older adults: A randomized controlled trial | Inappropriate intervention |
| 2781 | Factors related to post-stroke depression during the first 2 weeks after stroke | Inappropriate study design |
| 2782 | Psychophysiological responses to robot training in different recovery phases after stroke | Inappropriate study design |
| 2783 | The acute effect of moderate-intensity exercise on inhibitory control and activation of prefrontal cortex in younger and older adults | Inappropriate study design |

| | | |
|---|---|---|
| 2784 | The effects of an activity-based lifestyle intervention on moderate sleep complaints among older adults: study protocol for a randomized controlled trial | Inappropriate study design |
| 2785 | Relationship between acute physical fatigue and cognitive function during orthostatic challenge in men and women: A neuroergonomics investigation | Inappropriate population |
| 2786 | Physical exercise intensity during submersion selectively affects executive functions | Inappropriate population |
| 2787 | Modifiable risk factors for poor cognitive function in former American-style football players: Findings from the Harvard football players health study | Inappropriate study design |
| 2788 | Influence of Intradialytic Aerobic Training in Cerebral Blood Flow and Cognitive Function in Patients with Chronic Kidney Disease: A Pilot Randomized Controlled Trial | Inappropriate study design |
| 2789 | Repeated exposure to illusory sense of body ownership and agency over a moving virtual body improves executive functioning and increases prefrontal cortex activity in the elderly | Inappropriate population |
| 2790 | Reduced ambulatory activity after stroke: the role of balance, gait, and cardiovascular fitness | Inappropriate intervention |
| 2791 | Perceived barriers to exercise reported by individuals with stroke, who are able to walk in the community | Inappropriate study design |
| 2792 | Physical and cognitive activity and exercise for older adults: a review | Inappropriate study design |
| 2793 | Influence of visual stimulation-induced passive reproduction of motor images in the brain on motor paralysis after stroke | Inappropriate study design |
| 2794 | Robotic restoration of gait function in patients in the early recovery period of stroke | Inappropriate population |
| 2795 | Predictors of outcomes in exercisers with Parkinson disease: A two-year longitudinal cohort study | Inappropriate study design |
| 2796 | The effect of aerobic exercise on concussion recovery: A pilot clinical trial | Inappropriate intervention |
| 2797 | Effects of aerobic fitness on cognitive performance as a function of dual-task demands in older adults | Inappropriate population |
| 2798 | Sustained effects of acute resistance exercise on executive function in healthy middle-aged adults | Inappropriate population |
| 2799 | 'Masterstroke: a pilot group stroke prevention program for community dwelling stroke survivors' | Inappropriate outcome |
| 2800 | Virtual reality cognitive training among individuals with alcohol use disorder undergoing residential treatment: Pilot randomized controlled trial | Inappropriate intervention |
| 2801 | Association between the caregiver's burden and physical activity in community-dwelling caregivers of dementia patients | Inappropriate study design |
| 2802 | Daily activities: The impact of COPD and cognitive dysfunction | Inappropriate study design |
| 2803 | Leisure-time physical activity associates with cognitive decline: The Northern Manhattan Study | Inappropriate study design |
| 2804 | The feasibility, usability and acceptability of a tablet-based aphasia therapy in the acute phase following stroke | Inappropriate study design |
| 2805 | Behavioural activation therapy to improve participation in adults with depression following brain injury: A single-case experimental design study | Inappropriate study design |
| 2806 | Cognitive functioning enhancement in older adults: is there an advantage of multicomponent training over Nordic walking? | Inappropriate intervention |
| 2807 | Prominent health problems, socioeconomic deprivation, and higher brain age in lonely and isolated individuals: A population-based study | Inappropriate study design |
| 2808 | Levels of homocysteine are inversely associated with cardiovascular fitness in women, but not in men: data from the National Health and Nutrition Examination Survey 1999-2002 | Inappropriate study design |
| 2809 | Investigation of the relationship between physical activity, kinesiophobia and fear of falling in older adults with chronic pain | Inappropriate study design |
| 2810 | Primary prevention for risk factors of ischemic stroke with Baduanjin exercise intervention in the community elder population: study protocol for a randomized controlled trial | Inappropriate population |
| 2811 | Comparison of three instruments for activity disability in acute ischemic stroke survivors | Inappropriate study design |
| 2812 | Effect of static handgrip on plasma adrenomedullin concentration in patients with heart failure and in healthy subjects | Inappropriate study design |
| 2813 | E-health StandingTall balance exercise for fall prevention in older people: Results of a two year randomised controlled trial | Inappropriate population |
| 2814 | Motor-cognitive effects of a computerized game-based training method in people with dementia: a randomized controlled trial | Inappropriate study design |

| | | |
|---|---|---|
| 2815 | Is dynapenia associated with the onset and persistence of depressive and anxiety symptoms among older adults? Findings from the Irish longitudinal study on ageing | Inappropriate intervention |
| 2816 | Dynamic content reactivation supports naturalistic autobiographical recall in humans | Inappropriate study design |
| 2817 | Association of Exercise and Metabolic Equivalent of Task (MET) Score with Survival Outcomes after Out-of-Hospital Cardiac Arrest of Young and Middle Age | Inappropriate population |
| 2818 | Physical Activity for the Treatment and Prevention of Depression: A Rapid Review of Meta-Analyses. / Körperliche Aktivität für die Behandlung und Prävention der Depression: Übersichtsarbeit von Meta-Analysen | Inappropriate study design |
| 2819 | A Critical Systematic Review of Current Evidence on the Effects of Physical Exercise on Whole/Regional Grey Matter Brain Volume in Populations at Risk of Neurodegeneration | Inappropriate study design |
| 2820 | Effects of exercise training on left ventricular function and peripheral resistance in patients with chronic heart failure: A randomized trial | Inappropriate population |
| 2821 | Long-term beneficial effects of endralazine, a new arteriolar vasodilator at rest and during exercise capacity in chronic congestive heart failure | Inappropriate study design |
| 2822 | Acute Exercise Ameliorates Inhibition And Error Monitoring Among Drug Abusers: An ERPs Study | Inappropriate population |
| 2823 | Physical Activity, Aging and Brain Health. / Körperliche Aktivität, Altern und kognitive Fitness | Inappropriate study design |
| 2824 | High impact running improves learning | Inappropriate study design |
| 2825 | The Relations Between Physical Activity Level, Executive Function, and White Matter Microstructure in Older Adults | Inappropriate study design |
| 2826 | Physical Exercise and Cognitive Function in Post-stroke Patients: A Systematic Review with Meta-Analysis | Inappropriate study design |
| 2827 | Prevention of diseases after menopause | Inappropriate study design |
| 2828 | Effects of different exercise interventions on heart rate variability and cardiovascular health factors in older adults: a systematic review | Inappropriate study design |
| 2829 | A pilot randomized controlled trial to evaluate the benefit of the cardiac rehabilitation paradigm for the non-acute ischaemic stroke population | Inappropriate population |
| 2830 | Acute effects of Square Stepping Exercise on cognitive and social functions in sedentary young adults: a home-based online trial | Inappropriate population |
| 2831 | BAILAMOS: Development, Pilot Testing, and Future Directions of a Latin Dance Program for Older Latinos | Inappropriate population |
| 2832 | Trends in Meeting the Aerobic Physical Activity Guideline Among Adults With and Without Select Chronic Health Conditions, United States, 1998–2018 | Inappropriate study design |
| 2833 | Efficacy and Safety of Individualized Coaching After Stroke: the LAST Study (Life After Stroke): A Pragmatic Randomized Controlled Trial | Inappropriate population |
| 2834 | The Potential of Split-Belt Treadmill Walking as a Therapeutic Tool in Parkinson's Disease: a Narrative Review. / Das Potenzial des Split-Belt Treadmill Walking als therapeutisches Mittel bei Morbus Parkinson: eine Übersichtsarbeit | Inappropriate study design |
| 2835 | The Effect of Light-Intensity Cycling on Mood and Working Memory in Response to a Randomized, Placebo-Controlled Design | Inappropriate population |
| 2836 | Acute Effects of Frequent Light-Intensity Standing-Based Exercises That Interrupt 8 Hours of Prolonged Sitting on Postprandial Glucose in Stroke Survivors: A Dose-Escalation Trial | Inappropriate study design |
| 2837 | The Effects of Exercise on Patient-Reported Outcomes and Performance-Based Physical Function in Adults With Acute Leukemia Undergoing Induction Therapy: Exercise and Quality of Life in Acute Leukemia (EQUAL) | Inappropriate study design |
| 2838 | REducing SEDENTary Behavior Among Mild to Moderate Cognitively Impaired Assisted Living Residents: A Pilot Randomized Controlled Trial (RESEDENT Study) | Inappropriate intervention |
| 2839 | Influence of antenatal physical exercise on haemodynamics in pregnant women: a flexible randomisation approach | Inappropriate population |
| 2840 | Kinematic Analysis of Dance-Based Exergaming: A Cross-Sectional Study | Inappropriate study design |
| 2841 | Long-term hemodynamic effects of antihypertensive treatment | Inappropriate population |
| 2842 | The Effects of Interrupting Prolonged Sitting With Frequent Bouts of Light-Intensity Standing Exercises on Blood Pressure in Stroke Survivors: A Dose Escalation Trial | Inappropriate study design |
| 2843 | Effectiveness of Tai Chi for health promotion for adults with health conditions: a scoping review of Meta-analyses | Inappropriate study design |
| 2844 | Development of a Self-Efficacy Questionnaire for Walking in Patients with Mild Ischemic Stroke | Inappropriate population |

| | | |
|---|---|---|
| 2845 | Effects of Aerobic Exercise on Serum Biomarkers of Neuroplasticity and Brain Repair in Stroke: A Systematic Review | Inappropriate study design |
| 2846 | Effects of premorbid physical activity on stroke severity and post-stroke functioning | Inappropriate study design |
| 2847 | Elevated cerebral perfusion and preserved cognition in elite Brazilian Jiu-Jitsu athletes: Evidence for neuroprotection | Inappropriate study design |
| 2848 | Two Months of Using Global Recommendations for Physical Activity Had No Impact on Cognitive or Motor Functions in Overweight and Obese Middle-Aged Women | Inappropriate population |
| 2849 | Response of brain-derived neurotrophic factor to combining cognitive and physical exercise | Inappropriate population |
| 2850 | Exercise intensity when adjusted for an individual's maximal aerobic power positively affects executive functions in young adults | Inappropriate intervention |
| 2851 | Feedback-controlled robotics-assisted treadmill exercise to assess and influence aerobic capacity early after stroke: a proof-of-concept study | Inappropriate study design |
| 2852 | Few Structural Brain Changes Associated With Moderate-Intensity Interval Training and Low-Intensity Continuous Training in a Randomized Trial of Fitness and Older Adults | Inappropriate population |
| 2853 | Comparison of Activity and Participation, and Mental and Physical Functions of Immediate Post-Acute and Distant Post-Acute Adults with Chronic Cardiac Conditions | Inappropriate study design |
| 2854 | The early prevention of metabolic syndrome by physical exercise | Inappropriate study design |
| 2855 | The Effects of Low-Intensity Multimodal Proprioceptive Exercise on Cognitive Function in Older Adults | Inappropriate population |
| 2856 | Influence of previous physical activity on the outcome of patients treated by thrombolytic therapy for stroke | Inappropriate study design |
| 2857 | A randomized controlled trial into the cognitive effects of a computer-tailored physical activity intervention in older adults with chronic disease(s) | Inappropriate intervention |
| 2858 | Management of Parkinson's Disease during the COVID-19 Pandemic: Challenges and Opportunities | Inappropriate study design |
| 2859 | Key role of left ventricular untwisting in endurance cyclists at onset of exercise | Inappropriate study design |
| 2860 | Home-based multimodal exercise program in older people with Alzheimer disease: Randomized controlled trial protocol | Inappropriate study design |
| 2861 | Physical activity patterns of acute stroke patients managed in a rehabilitation focused stroke unit | Inappropriate outcome |
| 2862 | The effect of the Take Charge intervention on mood, motivation, activation and risk factor management: Analysis of secondary data from the Taking Charge after Stroke (TaCAS) trial | Inappropriate intervention |
| 2863 | Use of Accelerometer-Based Feedback of Walking Activity for Appraising Progress With Walking-Related Goals in Inpatient Stroke Rehabilitation: A Randomized Controlled Trial | Inappropriate population |
| 2864 | High-intensity functional exercise in older adults with dementia: A systematic review and meta-analysis | Inappropriate study design |
| 2865 | Levels of physical activity and quality of life among community-dwelling adults with stroke in a developing country | Inappropriate study design |
| 2866 | Post-concurrent exercise hemodynamics and cardiac autonomic modulation | Inappropriate study design |
| 2867 | L-lactate induces neurogenesis in the mouse ventricular-subventricular zone via the lactate receptor HCA1 | Inappropriate study design |
| 2868 | Psychological responses to HIIT and MICT over a 2-week progressive randomized trial among individuals at risk of type 2 diabetes | Inappropriate population |
| 2869 | Comparison of 2 types of therapeutic exercise: jaw opening exercise and head lift exercise for dysphagic stroke: A pilot study | Inappropriate study design |
| 2870 | Associations of Objectively Measured Physical Activity and Sedentary Time with the Risk of Stroke, Myocardial Infarction or All-Cause Mortality in 70-Year-Old Men and Women: A Prospective Cohort Study | Inappropriate population |
| 2871 | The association between obesity and fluid intelligence impairment is mediated by chronic low-grade inflammation | Inappropriate outcome |
| 2872 | Virtual Reality Exercises for Alzheimer's or Dementia | Inappropriate study design |
| 2873 | 'PDSAFE' - a multi-dimensional model of falls-rehabilitation for people with Parkinson's. A mixed methods analysis of therapists' delivery and experience | Inappropriate study design |
| 2874 | Vital signs: disability and physical activity--United States, 2009-2012 | Inappropriate population |
| 2875 | Predictive Relationships Between Parkinson's Disease Outcome Measures and Participation in Long-Term HIIT Exercise | Inappropriate intervention |

| | | |
|---|---|---|
| 2876 | Can lifestyle modification improve neurocognition? Rationale and design of the ENLIGHTEN clinical trial | Inappropriate study design |
| 2877 | How Adults with Stroke Conceptualize Physical Activity: An Exploratory Qualitative Study | Inappropriate population |
| 2878 | Effect of physical activity on cognitive flexibility, depression and RBD in healthy elderly | Inappropriate study design |
| 2879 | Home-based Walking And Global Cognitive Function In Patients With Renal Disease And Subjective Cognitive Complaints | Inappropriate population |
| 2880 | Physical activity is associated with higher cognitive function among adults at risk for Alzheimer's disease | Inappropriate study design |
| 2881 | Fall Risk Analysis: Implications For Exercise Prescription Among Older Adults | Inappropriate population |
| 2882 | Reliability Of Cycle-ergometer Based Cardiopulmonary Exercise Testing In Older Adults With Mild Alzheimer's Dementia | Inappropriate study design |
| 2883 | Leisure activities and the risk of dementia in the elderly | Inappropriate study design |
| 2884 | The New Age of Cardiac Rehab: Do Adult Protocols Meet the Needs of Children? | Inappropriate study design |
| 2885 | Characterization of an Algorithm for Autonomous, Closed-Loop Neuromodulation During Stroke Rehabilitation | Inappropriate study design |
| 2886 | Physical therapists' guideline adherence on early mobilization and intensity of practice at dutch acute stroke units: a country-wide survey | Inappropriate study design |
| 2887 | Fractionized Exercise Improves Cognitive Performance Under Conditions Of Normal And Long Duration Sleep | Inappropriate population |
| 2888 | Physical fitness: benefits of exercise for the older patient. 2 | Inappropriate study design |
| 2889 | Noninvasive detection of anaerobic threshold by digital plethysmography: theoretical background | Inappropriate study design |
| 2890 | A case report of idiosyncratic hyperthermia and review of U.S. Army heat stroke hospitalizations | Inappropriate study design |
| 2891 | Immediate Effects of Passive Ankle Joint Movement with Electrical Muscle Stimulation on Walking Speed in Older Adults | Inappropriate study design |
| 2892 | Can neuromuscular strength and function in people with dementia be rehabilitated using resistance-exercise training? Results from a preliminary intervention study | Inappropriate outcome |
| 2893 | Depression, Dementia And Psychiatric Disorders: The Heart Of The Problem | Inappropriate study design |
| 2894 | Effects of Tai Chi Practice on Inflammatory Biomarkers – A Systematic Review | Inappropriate study design |
| 2895 | Combination Effects Of Electrical Muscle Stimulation And Passive Cycle-ergometer On Metabolic Response In Healthy Adults | Inappropriate population |
| 2896 | Comparing Accuracy of Step Count Detection Algorithm at Different Activity Monitor Wear Locations | Inappropriate study design |
| 2897 | Clinical Predictors Of Vo2Max Response To Endurance Training: Heritage Family Study | Inappropriate study design |
| 2898 | Changes in the physical activity of acute stroke survivors between inpatient and community living with early supported discharge: an observational cohort study | Inappropriate population |
| 2899 | Using A Mixed Methods Design To Better Understand Older Adults' Responses To Resistance Training | Inappropriate intervention |
| 2900 | Effects Of Quenching Thirst Versus Full-Fluid Replenishment On Mood And Cognitive Changes After Mild-Hypohydration | Inappropriate study design |
| 2901 | Difference Between Fit Vs. Unfit In Heart Rate And Peripheral Vascular Resistance Incrementation During Orthostasis | Inappropriate study design |
| 2902 | Randomized clinical trial of therapeutic exercise in subacute stroke | Inappropriate population |
| 2903 | Association Of Vigorous-intensity Physical Activity With The Risk Of Dementia | Inappropriate study design |
| 2904 | The effects of lower extremity cross-training on gait and balance in stroke patients: a double-blinded randomized controlled trial | Inappropriate intervention |
| 2905 | Adherence to a Mediterranean diet, cognitive decline, and risk of dementia | Inappropriate study design |
| 2906 | Cognitive Effects of Aerobic Exercise in Alzheimer's Disease: A Pilot Randomized Controlled Trial | Inappropriate intervention |
| 2907 | Levels of physical activity in acute stroke patients treated at a stroke unit: A prospective, observational study | Inappropriate population |
| 2908 | Physical exercise for individuals with dementia: potential benefits perceived by formal caregivers | Inappropriate study design |
| 2909 | Increased serum brain-derived neurotrophic factor with high-intensity interval training in stroke patients: A randomized controlled trial | Inappropriate intervention |

| | | |
|---|---|---|
| 2910 | Aerobic endurance training to improve cognition and enhance recovery in schizophrenia: design and methodology of a multicenter randomized controlled trial | Inappropriate study design |
| 2911 | High-intensity exercise and cognitive function in cognitively normal older adults: a pilot randomised clinical trial | Inappropriate population |
| 2912 | Non-Exercise Based Estimation of Cardiorespiratory Fitness Mediates Associations between Comorbidities and Health-Related Quality of Life in Older Korean Adults with Diabetes | Inappropriate study design |
| 2913 | Inter-individual differences in the responses to aerobic exercise in Alzheimer's disease: Findings from the FIT-AD trial | Inappropriate study design |
| 2914 | Cytokine modulation in response to acute and chronic aquatic therapy intervention in Parkinson disease individuals: A pilot study | Inappropriate study design |
| 2915 | Effects of resistance training using elastic bands on muscle strength with or without a leucine supplement for 48 weeks in elderly patients with type 2 diabetes | Inappropriate intervention |
| 2916 | Effect of aerobic exercise (walking) training on functional status and health-related quality of life in chronic stroke survivors: a randomized controlled trial | Inappropriate study design |
| 2917 | Feasibility and adherence to moderate intensity cardiovascular fitness training following stroke: a pilot randomized controlled trial | Inappropriate intervention |
| 2918 | Exercise and cancer recovery | Inappropriate study design |
| 2919 | Immediate effects of a single session of physical exercise on cognition and cerebral blood flow: A randomized controlled study of older adults | Inappropriate study design |
| 2920 | Impact of Cardiorespiratory Fitness on Frequency of Atrial Fibrillation, Stroke, and All-Cause Mortality | Inappropriate study design |
| 2921 | Cross training effects of non-paralytic dorsiflexion muscle strengthening exercise on paralytic dorsiflexor muscle activity, gait ability, and balancing ability in patients with chronic stroke: A randomized, controlled, pilot trial | Inappropriate intervention |
| 2922 | Effects of aerobic treadmill training on gait velocity, cadence, and gait symmetry in chronic hemiparetic stroke: a preliminary report | Inappropriate population |
| 2923 | Graded motor imagery training as a home exercise program for upper limb motor function in patients with chronic stroke: A randomized controlled trial | Inappropriate intervention |
| 2924 | Improved cerebral vasomotor reactivity after exercise training in hemiparetic stroke survivors | Inappropriate population |
| 2925 | Exercise, diet, and cognition in a 4-year randomized controlled trial: Dose-Responses to Exercise Training (DR's EXTRA) | Inappropriate population |
| 2926 | What's keeping people after stroke from walking outdoors to become physically active? A qualitative study, using an integrated biomedical and behavioral theory of functioning and disability | Inappropriate study design |
| 2927 | Task-specific training for improving propulsion symmetry and gait speed in people in the chronic phase after stroke: a proof-of-concept study | Inappropriate intervention |
| 2928 | A methodology for an acute exercise clinical trial called dementia risk and dynamic response to exercise | Inappropriate study design |
| 2929 | Examining the Inter-relations of Depression, Physical Function, and Cognition with Subjective Sleep Parameters among Stroke Survivors: A Cross-sectional Analysis | Inappropriate study design |
| 2930 | Effect of combined home-based, overground robotic-assisted gait training and usual physiotherapy on clinical functional outcomes in people with chronic stroke: A randomized controlled trial | Inappropriate intervention |
| 2931 | Effects of electric stimulation-assisted cycling training in people with chronic stroke | Inappropriate population |
| 2932 | Comparison of neuromuscular and cardiovascular exercise intensity and enjoyment between standard of care, off-the-shelf and custom active video games for promotion of physical activity of persons post-stroke | Inappropriate population |
| 2933 | Swimming-related effects on cerebrovascular and cognitive function | Inappropriate population |
| 2934 | The Effects of Aerobic Exercise on Psychological Functioning in Family Caregivers: Secondary Analyses of a Randomized Controlled Trial | Inappropriate intervention |
| 2935 | Association of leisure-time physical activity with cognition by apolipoprotein-E genotype in persons aged 60 years and over: the National Health and Nutrition Examination Survey (NHANES-III) | Inappropriate outcome |
| 2936 | Statistical analysis plan for management of hypertension and multiple risk factors to enhance cardiovascular health in Singapore: the SingHypertension pragmatic cluster randomized controlled trial | Inappropriate population |
| 2937 | Association of Midlife Cardiorespiratory Fitness With Incident Depression and Cardiovascular Death After Depression in Later Life | Inappropriate study design |
| 2938 | Exercise effects on backward walking speed in people with dementia: A randomized controlled trial | Inappropriate intervention |

| | | |
|---|---|---|
| 2939 | Use of the Frank-Starling mechanism during exercise is linked to exercise-induced changes in arterial load | Inappropriate study design |
| 2940 | Effects of Online Bodyweight High-Intensity Interval Training Intervention and Health Education on the Mental Health and Cognition of Sedentary Young Females | Inappropriate population |
| 2941 | Conditioning leisure time physical activity and cardiorespiratory fitness in sociodemographic groups of middle-ages men in eastern Finland | Inappropriate study design |
| 2942 | Home-based rehabilitation programs on postural balance, walking, and quality of life in patients with stroke: A single-blind, randomized controlled trial | Inappropriate population |
| 2943 | Robot-assisted gait training improves brachial-ankle pulse wave velocity and peak aerobic capacity in subacute stroke patients with totally dependent ambulation: Randomized controlled trial | Inappropriate population |
| 2944 | Impact of aerobic exercise on clinical and magnetic resonance imaging biomarkers in persons with multiple sclerosis: An exploratory randomized controlled trial | Inappropriate intervention |
| 2945 | Metabolic syndrome marks early risk for cognitive decline with APOE4 gene variation: A case study | Inappropriate population |
| 2946 | Physiological Responses and Perceived Exertion During Robot-Assisted and Body Weight-Supported Gait After Stroke | Inappropriate study design |
| 2947 | Effects of intensive multiplanar trunk training coupled with dual-task exercises on balance, mobility, and fall risk in patients with stroke: a randomized controlled trial | Inappropriate population |
| 2948 | The Effect of Aerobic Exercise on Concussion Recovery: A Pilot Clinical Trial | Inappropriate intervention |
| 2949 | Absolute and relative reliability of acute effects of aerobic exercise on executive function in seniors | Inappropriate population |
| 2950 | Randomized Controlled Trial on the Effects of a Combined Intervention of Computerized Cognitive Training Preceded by Physical Exercise for Improving Frailty Status and Cognitive Function in Older Adults | Inappropriate intervention |
| 2951 | Reduced leg blood flow during submaximal exercise in type 2 diabetes | Inappropriate study design |
| 2952 | Feasibility and effect of interactive telerehabilitation on balance in individuals with chronic stroke: a pilot study | Inappropriate study design |
| 2953 | Ventilatory threshold may be a more specific measure of aerobic capacity than peak oxygen consumption rate in persons with stroke | Inappropriate population |
| 2954 | Effect of a Coordinated Community and Chronic Care Model Team Intervention vs Usual Care on Systolic Blood Pressure in Patients With Stroke or Transient Ischemic Attack: The SUCCEED Randomized Clinical Trial | Inappropriate intervention |
| 2955 | Protocol for the WARM Hearts study: examining cardiovascular disease risk in middle-aged and older women - a prospective, observational cohort study | Inappropriate study design |
| 2956 | Post-resistance exercise hypotension, hemodynamics, and heart rate variability: influence of exercise intensity | Inappropriate population |
| 2957 | Prediction of fear of falling at 6 months after stroke based on 279 individuals from the Fall Study of Gothenburg | Inappropriate study design |
| 2958 | For non-exercising people, the number of steps walked is more strongly associated with health than time spent walking | Inappropriate population |
| 2959 | 12 weeks of strength training improves fluid cognition in older adults: A nonrandomized pilot trial | Inappropriate study design |
| 2960 | Breaking up sitting time after stroke (BUST-stroke) | Inappropriate study design |
| 2961 | Decreased exercise capacity in young athletes using self-adapted mouthguards | Inappropriate population |
| 2962 | Sildenafil improves exercise hemodynamics in Fontan patients | Inappropriate study design |
| 2963 | Cognitive Performance Does not Limit Physical Activity Participation in the Lifestyle Interventions and Independence for Elders Pilot Study (LIFE-P) | Inappropriate population |
| 2964 | The Effects of Sodium Phosphate Supplementation on the Cardiorespiratory System and Gross Efficiency during Exercise under Hypoxia in Male Cyclists: A Randomized, Placebo-Controlled, Cross-Over Study | Inappropriate population |
| 2965 | Building your best day for healthy brain aging-The neuroprotective effects of optimal time use | Inappropriate population |
| 2966 | Traditional Chinese medicine training for cardiac rehabilitation: a randomized comparison with aerobic and resistance training | Inappropriate population |
| 2967 | A family care model for older persons with hip-fracture and cognitive impairment: a randomized controlled trial | Inappropriate intervention |
| 2968 | Determining optimal poststroke exercise: Study protocol for a randomized controlled trial investigating therapeutic intensity and dose on functional recovery during stroke inpatient rehabilitation | Inappropriate study design |

| | | |
|---|---|---|
| 2969 | Memory enhancement by multidomain group cognitive training in patients with Parkinson's disease and mild cognitive impairment: long-term effects of a multicenter randomized controlled trial | Inappropriate intervention |
| 2970 | Cognitive effects of acute aerobic exercise: exploring the influence of exercise duration, exhaustion, task complexity and expectancies in endurance-trained individuals | Inappropriate study design |
| 2971 | The interaction of acute physical fatigue with three traditional functional performance tests and the reactive balance test | Inappropriate population |
| 2972 | The Effects of the Combination of High-Intensity Interval Training with 3D-Multiple Object Tracking Task on Perceptual-Cognitive Performance: a Randomized Controlled Intervention Trial | Inappropriate population |
| 2973 | Verapamil acutely reduces ventricular-vascular stiffening and improves aerobic exercise performance in elderly individuals | Inappropriate study design |
| 2974 | Effect of Acute Exercise and Cycling Desk on Energy Intake and Appetite Response to Mental Work: the CORTEX Study | Inappropriate population |
| 2975 | Frailty, dependency and mortality predictors in a cohort of Cuban older adults, 2003-2011 | Inappropriate study design |
| 2976 | Effects of a Strength-Dominated Exercise Program on Physical Fitness and Cognitive Performance in Preschool Children | Inappropriate intervention |
| 2977 | Is the Effect of a High-Intensity Functional Exercise Program on Functional Balance Influenced by Applicability and Motivation among Older People with Dementia in Nursing Homes? | Inappropriate study design |
| 2978 | Effects of very low-carbohydrate vs. high-carbohydrate weight loss diets on psychological health in adults with obesity and type 2 diabetes: a 2-year randomized controlled trial | Inappropriate population |
| 2979 | Domain dependent associations between cognitive functioning and regular voluntary exercise behavior | Inappropriate outcome |
| 2980 | A pilot randomized controlled trial of robotic exoskeleton-assisted exercise rehabilitation in multiple sclerosis | Inappropriate population |
| 2981 | An analysis of the determinants of exercise performance in congestive heart failure | Inappropriate study design |
| 2982 | Effect of Differing Durations of High-Intensity Intermittent Activity on Cognitive Function in Adolescents | Inappropriate population |
| 2983 | Effects of leg blood flow restriction during walking on cardiovascular function | Inappropriate population |
| 2984 | Effects of a 10-week active recess program in school setting on physical fitness, school aptitudes, creativity and cognitive flexibility in elementary school children. A randomised-controlled trial | Inappropriate population |
| 2985 | Lifting cognition: a meta-analysis of effects of resistance exercise on cognition | Inappropriate study design |
| 2986 | Effects of Short-Interval and Long-Interval Swimming Protocols on Performance, Aerobic Adaptations, and Technical Parameters: A Training Study | Inappropriate population |
| 2987 | Prestroke physical activity is associated with good functional outcome and arterial recanalization after stroke due to a large vessel occlusion | Inappropriate population |
| 2988 | Norwegian Psychomotor Physiotherapy versus Cognitive Patient Education and active physiotherapy-A randomized controlled trial | Inappropriate population |
| 2989 | Exercise intolerance in comorbid COPD and heart failure: the role of impaired aerobic function | Inappropriate study design |
| 2990 | Acute aerobic exercise to recover from mental exhaustion - a randomized controlled trial | Inappropriate population |
| 2991 | Cardiorespiratory fitness attenuates age-associated aggregation of white matter hyperintensities in an at-risk cohort | Inappropriate population |
| 2992 | Dual-task walking and automaticity after Stroke: insights from a secondary analysis and imaging sub-study of a randomised controlled trial | Inappropriate intervention |
| 2993 | Can supine recovery mitigate the exercise intensity dependent attenuation of post-exercise heat loss responses? | Inappropriate population |
| 2994 | Novel evidence on the effect of tramadol on self-paced high-intensity cycling | Inappropriate study design |
| 2995 | Cyclical blood flow restriction resistance exercise: a potential parallel to remote ischemic preconditioning? | Inappropriate study design |
| 2996 | Effects of Two Physical Training Programs on the Cognitive Status of a Group of Older Adults in Chile | Inappropriate intervention |
| 2997 | Leisure activities and the risk of amnestic mild cognitive impairment in the elderly | Inappropriate study design |
| 2998 | The Effects of 12-Week Beta-Hydroxy-Beta-Methylbutyrate Supplementation in Patients with Liver Cirrhosis: results from a Randomized Controlled Single-Blind Pilot Study | Inappropriate population |

| | | |
|---|---|---|
| 2999 | A randomized controlled trial comparing community lifestyle interventions to improve adherence to diet and physical activity recommendations: the VitalUM study | Inappropriate intervention |
| 3000 | Blunted Maximal and Submaximal Responses to Cardiopulmonary Exercise Tests in Patients With Parkinson Disease | Inappropriate study design |
| 3001 | Effects of Exergame-Based Dual-Task Training on Executive Function and Dual-Task Performance in Community-Dwelling Older People: a Randomized-Controlled Trial | Inappropriate intervention |
| 3002 | Serum levels of insulin-like growth factor-1 and brain-derived neurotrophic factor as potential recovery biomarkers in stroke | Inappropriate study design |
| 3003 | Multivariate radiomics models based on 18F-FDG hybrid PET/MRI for distinguishing between Parkinson's disease and multiple system atrophy | Inappropriate study design |
| 3004 | Walk, Talk and Listen: a pilot randomised controlled trial targeting functional fitness and loneliness in older adults with hearing loss | Inappropriate study design |
| 3005 | Computer Game Assisted Task Specific Exercises in the Treatment of Motor and Cognitive Function and Quality of Life in Stroke: a Randomized Control Study | Inappropriate intervention |
| 3006 | Therapeutically relevant structural and functional mechanisms triggered by physical and cognitive exercise | Inappropriate study design |
| 3007 | Mental Fatigue From Smartphone Use Reduces Volume-Load in Resistance Training: a Randomized, Single-Blinded Cross-Over Study | Inappropriate population |
| 3008 | Central hemodynamic effects at rest and during exercise of acute and chronic treatment with nisoldipine in essential hypertension | Inappropriate study design |
| 3009 | Mindful Self-Compassion program for chronic pain patients: a randomized controlled trial | Inappropriate population |
| 3010 | The Long-Term Dietitian and Psychological Support of Obese Patients Who Have Reduced Their Weight Allows Them to Maintain the Effects | Inappropriate population |
| 3011 | Improved exercise capacity in patients after minor ischemic stroke undergoing a supervised exercise training program | Inappropriate outcome |
| 3012 | Effects of Dual-Task Gait Treadmill Training on Gait Ability, Dual-Task Interference, and Fall Efficacy in People With Stroke: a Randomized Controlled Trial | Inappropriate population |
| 3013 | Haemodynamics at rest and during isometric exercise under long-term treatment with captopril in dilated and ischaemic cardiomyopathy | Inappropriate study design |
| 3014 | Postexercise Hemodynamic Responses in Lean and Obese Men | Inappropriate study design |
| 3015 | Effects of resistance training on metabolic and cardiovascular responses to a maximal cardiopulmonary exercise test in Parkinson`s disease | Inappropriate intervention |
| 3016 | Perturbation training to promote safe independent mobility post-stroke: study protocol for a randomized controlled trial | Inappropriate study design |
| 3017 | Effects of Beetroot Juice Supplementation on Cognitive Function, Aerobic and Anaerobic Performances of Trained Male Taekwondo Athletes: a Pilot Study | Inappropriate population |
| 3018 | Hemodynamic effects of unloading the old heart | Inappropriate study design |
| 3019 | Ain't Just Imagination! Effects of Motor Imagery Training on Strength and Power Performance of Athletes during Detraining | Inappropriate population |
| 3020 | Effects of stroke resistance on rowing economy in club rowers post-season | Inappropriate study design |
| 3021 | Effectiveness of robotic balance training on postural instability in patients with mild Parkinson's disease: a pilot, single blind, randomized controlled trial | Inappropriate intervention |
| 3022 | Associations between Physical Activity and Cognitive Functioning among Middle-Aged and Older Adults | Inappropriate study design |
| 3023 | Predicting Working Memory Training Responsiveness in Parkinson's Disease: both "System Hardware" and Room for Improvement Are Needed | Inappropriate intervention |
| 3024 | Effect analysis of kinetic energy progressive exercise in patients with acute myocardial infarction after percutaneous coronary intervention: a randomized trial | Inappropriate intervention |
| 3025 | Hemodynamic evaluation during exercise test after acute and chronic ibopamine treatment in patients with congestive heart failure | Inappropriate study design |
| 3026 | Trait Energy and Fatigue Modify the Effects of Caffeine on Mood, Cognitive and Fine-Motor Task Performance: a Post-Hoc Study | Inappropriate population |
| 3027 | Energy cost and psychological impact of robotic-assisted gait training in people with spinal cord injury: effect of two different types of devices | Inappropriate study design |
| 3028 | Evaluating and Treating Frailty in Cardiac Rehabilitation | Inappropriate study design |
| 3029 | The effects of neuroplasticity-based auditory information processing remediation in adults with chronic traumatic brain injury | Inappropriate intervention |
| 3030 | Higher vitamin D dietary intake is associated with lower risk of alzheimer's disease: a 7-year follow-up | Inappropriate population |

| | | |
|---|---|---|
| 3031 | Predictors of adherence to an internet-based cognitive behavioral therapy program for individuals with chronic pain and comorbid psychological distress | Inappropriate population |
| 3032 | Predicting physical activity change in men and women in two New England communities | Inappropriate study design |
| 3033 | Balance rehabilitation with a virtual reality protocol for patients with hereditary spastic paraplegia: protocol for a clinical trial | Inappropriate study design |
| 3034 | The effects of hydration status and ice-water dousing on physiological and performance indices during a simulated soccer match in the heat | Inappropriate population |
| 3035 | The effects of acute dopamine reuptake inhibition on cognitive function during passive hyperthermia | Inappropriate population |
| 3036 | Neuromuscular Electric Stimulation in Addition to Exercise Therapy in Patients with Lower Extremity Paresis Due to Acute Ischemic Stroke. A proof-of-concept randomised controlled trial | Inappropriate intervention |
| 3037 | Effects of task-oriented training combined with aerobic training on serum BDNF, GDNF, IGF-1, VEGF, TNF-α, and IL-1β levels in people with Parkinson's disease: a randomized controlled study | Inappropriate intervention |
| 3038 | Cognition and motor impairment correlates with exercise test performance after stroke | Inappropriate study design |
| 3039 | Conventional chemotherapy for acute myeloid leukemia in older adults: impact on nutritional, cognitive, and functional status | Inappropriate study design |
| 3040 | A novel cognitive-motor exercise program delivered via a tablet to improve mobility in older people with cognitive impairment - StandingTall Cognition and Mobility | Inappropriate intervention |
| 3041 | Physiologic basis for the age-related decline in aerobic work capacity | Inappropriate study design |
| 3042 | One-year aerobic exercise altered cerebral vasomotor reactivity in mild cognitive impairment | Inappropriate intervention |
| 3043 | Efficacy of high-intensity aerobic exercise on cognitive performance in people with multiple sclerosis: a randomized controlled trial | Inappropriate intervention |
| 3044 | Efficacy and feasibility of HIIT training for university students: The Uni-HIIT RCT | Inappropriate study design |
| 3045 | Neck-related function and its connection with disability in chronic whiplash-associated disorders: secondary analysis of a randomized controlled study | Inappropriate population |
| 3046 | Executive functions, physical fitness and mobility in well-functioning older adults | Inappropriate outcome |
| 3047 | The effects of lingual exercise in stroke patients with dysphagia | Inappropriate outcome |
| 3048 | Aerobic fitness in women and responses to lower body negative pressure | Inappropriate population |
| 3049 | Improvement in cognitive impairment following a 12-week aerobic exercise intervention in individuals with non-cirrhotic chronic hepatitis C | Inappropriate intervention |
| 3050 | Reduced hip bone mineral density is related to physical fitness and leg lean mass in ambulatory individuals with chronic stroke | Inappropriate study design |
| 3051 | Functional electrical stimulation in-bed cycle ergometry in mechanically ventilated patients: a multicentre randomised controlled trial | Inappropriate population |
| 3052 | Associations between handgrip strength and mild cognitive impairment in middle-aged and older adults in six low- and middle-income countries | Inappropriate population |
| 3053 | High-intensity training as a novel treatment for impaired awareness of hypoglycaemia in type 1 diabetes [HIT4HYPOS]: protocol for a randomized parallel-group study | Inappropriate study design |
| 3054 | The effect of a one-year vigorous physical activity intervention on fitness, cognitive performance and mental health in young adolescents: the Fit to Study cluster randomised controlled trial | Inappropriate population |
| 3055 | Reliability of intensity-based physical activity measurement using an activity monitor in people with subacute stroke in the hospital setting: a cross-sectional study | Inappropriate population |
| 3056 | Effects of Two Randomized and Controlled Multi-Component Interventions Focusing On 24-Hour Movement Behavior among Office Workers: a Compositional Data Analysis | Inappropriate population |
| 3057 | Results of the Optimune trial: a randomized controlled trial evaluating a novel Internet intervention for breast cancer survivors | Inappropriate population |
| 3058 | Gains of cardiorespiratory fitness with arm-crank training in spinally disabled men | Inappropriate population |
| 3059 | Effects of Aerobic, Resistance, or Combined Exercise Training Among Older Adults with Subjective Memory Complaints: a Randomized Controlled Trial | Inappropriate intervention |
| 3060 | Quantifying physical activity in early Parkinson disease using a commercial activity monitor | Inappropriate study design |
| 3061 | High-intensity exercise and cognitive function in cognitively normal older adults: a pilot randomised clinical trial | Inappropriate study design |

| | | |
|---|---|---|
| 3062 | Long-term effects of a 12 weeks high-intensity functional exercise program on physical function and mental health in nursing home residents with dementia: a single blinded randomized controlled trial | Inappropriate outcome |
| 3063 | Twelve-month findings of the MOVE Frankston randomised controlled trial of interventions to increase recreation facility usage and physical activity among adults | Inappropriate population |
| 3064 | Exercise Intervention for Academic Achievement Among Children: a Randomized Controlled Trial | Inappropriate population |
| 3065 | Obesity as a risk factor for various diseases: necessity of lifestyle changes for healthy aging | Inappropriate study design |
| 3066 | Effects of Exercise Dose and Detraining Duration on Mobility at Late Midlife: a Randomized Clinical Trial | Inappropriate study design |
| 3067 | Central circulatory and peripheral O2 extraction changes as interactive facilitators of pulmonary O2 uptake during a repeated high-intensity exercise protocol in humans | Inappropriate study design |
| 3068 | Feasibility of reducing frailty components in older adults with Alzheimer's dementia: a randomized controlled home-based exercise trial (AD-HOMEX) | Inappropriate study design |
| 3069 | Implementing physical exercise and music interventions for patients suffering from dementia on an acute psychogeriatric inpatient ward | Inappropriate outcome |
| 3070 | The impact of exercise intensity on neurophysiological indices of food-related inhibitory control and cognitive control: a randomized crossover event-related potential (ERP) study | Inappropriate population |
| 3071 | Exercise, diet, and cognition in a 4-year randomized controlled trial: dose-Responses to Exercise Training (DR's EXTRA) | Inappropriate population |
| 3072 | Prominent physical inactivity in acute dementia care: Psychopathology seems to be more important than the dose of sedative medication | Inappropriate study design |
| 3073 | The performance of balance exercises during daily tooth brushing is not sufficient to improve balance and muscle strength in healthy older adults | Inappropriate intervention |
| 3074 | The effect of regular walks on various health aspects in older people with dementia: protocol of a randomized-controlled trial | Inappropriate population |
| 3075 | The influence of a 16-week exercise program, APOE status, and age on executive function task performance: a randomized trial | Inappropriate intervention |
| 3076 | Multicomponent exercise to improve motor functions, cognition and well-being for nursing home residents who are unable to walk - A randomized controlled trial | Inappropriate intervention |
| 3077 | Effects of a "test in-train out" walking program versus supervised standard rehabilitation in chronic stroke patients: a feasibility and pilot randomized study | Inappropriate population |
| 3078 | Effects of a Health Promotion Intervention on Physical Activity in African American Men Living with HIV: randomized Controlled Trial | Inappropriate population |
| 3079 | The differential relationship of an afterschool physical activity intervention on brain function and cognition in children with obesity and their normal weight peers | Inappropriate study design |
| 3080 | E-health StandingTall balance exercise for fall prevention in older people: results of a two year randomised controlled trial | Inappropriate population |
| 3081 | The Effectiveness of an Active Learning Program in Promoting a Healthy Lifestyle among Older Adults with Low Health Literacy: a Randomized Controlled Trial | Inappropriate intervention |
| 3082 | Objectively measured physical activity, brain atrophy, and white matter lesions in older adults with mild cognitive impairment | Inappropriate population |
| 3083 | Impact of prolonged sitting and physical activity breaks on cognitive performance, perceivable benefits, and cardiometabolic health in overweight/obese adults: the role of meal composition | Inappropriate population |
| 3084 | The impact of Tai Chi and mind-body breathing in COPD: insights from a qualitative sub-study of a randomized controlled trial | Inappropriate study design |
| 3085 | Effects of yoga on well-being and healthy ageing: study protocol for a randomised controlled trial (FitForAge) | Inappropriate study design |
| 3086 | Effect of Aerobic Exercise on Cardiopulmonary Responses and Predictors of Change in Individuals With Parkinson's Disease | Inappropriate intervention |
| 3087 | A Controlled Clinical Trial on the Effects of Exercise on Cognition and Mobility in Adults With Multiple Sclerosis | Inappropriate population |
| 3088 | Effects of aerobic walking on cognitive function in patients with schizophrenia: a randomized controlled trial | Inappropriate intervention |
| 3089 | Cardiovascular responses to exercise. Effects of aging and exercise training in healthy men | Inappropriate study design |
| 3090 | A dismantling study on imaginal retraining in smokers | Inappropriate population |

| | | |
|---|---|---|
| 3091 | Effects of a Tailored Exercise Intervention in Acutely Hospitalized Oldest Old Diabetic Adults: an Ancillary Analysis | Inappropriate outcome |
| 3092 | Current and past leisure time physical activity in relation to risk of Alzheimer's disease in older adults | Inappropriate study design |
| 3093 | Time Course of Relapse Following Acute Treatment for Anorexia Nervosa | Inappropriate population |
| 3094 | Does raising heart rate prior to a behavioural test enhance learning in cognitive therapy for anxiety? An experimental test for the treatment of fear of heights using virtual reality | Inappropriate population |
| 3095 | Walking Aids Moderate Exercise Effects on Gait Speed in People With Dementia: A Randomized Controlled Trial | Inappropriate outcome |
| 3096 | A randomized controlled trial of exercise on augmenting the effects of cognitive remediation in persons with severe mental illness | Inappropriate intervention |
| 3097 | Work capacity, exercise responses and body composition of professional pilots in relation to age | Inappropriate population |
| 3098 | Beyond Physical Capacity: factors Associated With Real-world Walking Activity After Stroke | Inappropriate study design |
| 3099 | Psychological training to improve psychosocial function in patients with major depressive disorder: a randomised clinical trial | Inappropriate population |
| 3100 | Influence of Baseline Kinesiophobia Levels on Treatment Outcome in People With Chronic Spinal Pain | Inappropriate study design |
| 3101 | Effect of age on exercise-induced alterations in cognitive executive function: relationship to cerebral perfusion | Inappropriate study design |
| 3102 | FEASibility testing a randomized controlled trial of an exercise program to improve cognition for T2DM patients (the FEAST trial): a study protocol | Inappropriate study design |
| 3103 | Cardiac function assessed by exercise echocardiography on the first morning after coronary artery bypass grafting | Inappropriate population |
| 3104 | Exercise, Processing Speed, and Subsequent Falls: a Secondary Analysis of a 12-Month Randomized Controlled Trial | Inappropriate population |
| 3105 | The effects of barbell resistance exercise on information processing speed and conflict-related ERP in older adults: a crossover randomized controlled trial | Inappropriate population |
| 3106 | Effectiveness of a community-based low intensity exercise programme for ambulatory stroke survivors | Inappropriate population |
| 3107 | Urinary sodium excretion and dietary sources of sodium intake in community-dwelling Chinese adults aged 50 years and over | Inappropriate intervention |
| 3108 | Postexercise hypotension in moderately trained athletes after maximal exercise | Inappropriate intervention |
| 3109 | Does muscular or mental fatigue have an influence on the nociceptive flexion reflex? A randomized cross-over study in healthy people | Inappropriate population |
| 3110 | Effects of aerobic exercise and gender on visual and auditory P300, reaction time, and accuracy | Inappropriate study design |
| 3111 | Feasibility and preliminary effect of anabolic steroids in addition to strength training and nutritional supplement in rehabilitation of patients with hip fracture: a randomized controlled pilot trial (HIP-SAP1 trial) | Inappropriate population |
| 3112 | Exercise cardiac function in young through elderly endurance trained women | Inappropriate outcome |
| 3113 | Contemplative Mental Training Reduces Hair Glucocorticoid Levels in a Randomized Clinical Trial | Inappropriate population |
| 3114 | Physical activity and risk of stroke in women | Inappropriate study design |
| 3115 | Recumbent cycling to improve outcomes in people with hip fracture: a feasibility randomized trial | Inappropriate population |
| 3116 | Pulmonary Vascular Function and Aerobic Exercise Capacity at Moderate Altitude | Inappropriate study design |
| 3117 | Contribution of systemic arterial compliance and systemic vascular resistance to effective arterial elastance changes during exercise in humans | Inappropriate study design |
| 3118 | The Daily Activity Study of Health (DASH): a pilot randomized controlled trial to enhance physical activity in sedentary older adults | Inappropriate population |
| 3119 | Acute hemodynamic responses of spinal cord injured individuals to functional neuromuscular stimulation-induced knee extension exercise | Inappropriate study design |
| 3120 | Tai Chi training's effect on lower extremity muscle co-contraction during single- and dual-task gait: cross-sectional and randomized trial studies | Inappropriate study design |
| 3121 | Cognitive response to testosterone replacement added to intensive lifestyle intervention in older men with obesity and hypogonadism: prespecified secondary analyses of a randomized clinical trial | Inappropriate population |
| 3122 | Cognitive Impairment Along the Course of Depression: Non-Pharmacological Treatment Options | Inappropriate study design |

| | | |
|---|---|---|
| 3123 | The effects of a physical and cognitive training intervention vs. physical training alone on older adults' physical activity: a randomized controlled trial with extended follow-up during COVID-19 | Inappropriate intervention |
| 3124 | Dual-task exercise improves walking ability in chronic stroke: a randomized controlled trial | Inappropriate intervention |
| 3125 | Association of Urine Biomarkers of Kidney Tubule Injury and Dysfunction With Frailty Index and Cognitive Function in Persons With CKD in SPRINT | Inappropriate study design |
| 3126 | Physiological responses and enjoyment of Kinect-based exergames in older adults at risk for falls: A feasibility study | Inappropriate population |
| 3127 | Cardiovascular responses during prolonged exercise at ventilatory threshold in boys and men | Inappropriate study design |
| 3128 | Physical activity modifies the association between depression and cognitive function in older adults | Inappropriate study design |
| 3129 | Noninvasive assessment of acute effects of nifedipine on rest and exercise hemodynamics and cardiac function in patients with aortic regurgitation | Inappropriate study design |
| 3130 | Protective effects of acute exercise prior to doxorubicin on cardiac function of breast cancer patients: A proof-of-concept RCT | Inappropriate study design |
| 3131 | Effects of acute and chronic minoxidil administration on rest and exercise hemodynamics and clinical status in patients with severe, chronic heart failure | Inappropriate study design |
| 3132 | Influence of light physical activity on cardiac responses during recovery from exercise in humans | Inappropriate population |
| 3133 | Low intensity exercise training in patients with chronic heart failure | Inappropriate study design |
| 3134 | The influence of physical exercise and leisure activity on neuropsychological functioning in older adults | Inappropriate outcome |
| 3135 | The Effects of Exercise on Falls in Older People With Dementia Living in Nursing Homes: A Randomized Controlled Trial | Inappropriate study design |
| 3136 | Recovery of ambulation activity across the first six months post-stroke | Inappropriate study design |
| 3137 | Investigating measures of intensity during a structured upper limb exercise program in stroke rehabilitation: an exploratory study | Inappropriate study design |
| 3138 | Relationship between cardiopulmonary response to exercise and adiposity in survivors of childhood malignancy | Inappropriate intervention |
| 3139 | WISE-2005: adrenergic responses of women following 56-days, 6 degrees head-down bed rest with or without exercise countermeasures | Inappropriate population |
| 3140 | Central hemodynamic responses during acute high-intensity interval exercise and moderate continuous exercise in patients with heart failure | Inappropriate population |
| 3141 | The association of single and combined factors of sedentary behavior and physical activity with subjective cognitive complaints among community-dwelling older adults: Cross-sectional study | Inappropriate study design |
| 3142 | Higher Doses Improve Walking Recovery During Stroke Inpatient Rehabilitation | Inappropriate population |
| 3143 | Acute hemodynamic effects of an alpha- and beta-receptor blocking agent (AH 5158) on the systemic and pulmonary circulation at rest and during exercise in hypertensive patients | Inappropriate study design |
| 3144 | Energetic Profile in Forehand Loop Drive Practice with Well-Trained, Young Table Tennis Players | Inappropriate population |
| 3145 | Effect of resistance-type exercise training with or without protein supplementation on cognitive functioning in frail and pre-frail elderly: secondary analysis of a randomized, double-blind, placebo-controlled trial | Inappropriate study design |
| 3146 | Central hemodynamics during progressive upper- and lower-body exercise and recovery | Inappropriate study design |
| 3147 | Running effects on cognition and plasticity (ReCaP): study protocol of a longitudinal examination of multimodal adaptations of marathon running | Inappropriate population |
| 3148 | From bedside to bench: does mental and physical activity promote cognitive vitality in late life? | Inappropriate population |
| 3149 | An education programme influencing health professionals to recommend exercise to their type 2 diabetes patients - understanding the processes: a case study from Oxfordshire, UK | Inappropriate population |
| 3150 | Resistance training improves cardiac output, exercise capacity and tolerance to positive airway pressure in Fontan physiology | Inappropriate study design |
| 3151 | Is there influence of the load addition during treadmill training on cardiovascular parameters and gait performance in patients with stroke? A randomized clinical trial | Inappropriate study design |
| 3152 | The effects of multidisciplinary rehabilitation in patients with early-to-middle-stage Huntington's disease: a pilot study | Inappropriate population |
| 3153 | Resistance exercise training improves heart function and physical fitness in stable patients with heart failure | Inappropriate outcome |

| | | |
|---|---|---|
| 3154 | Haemodynamic parameters in postmenopausal women - beneficial effect of moderate continuous exercise training | Inappropriate population |
| 3155 | Perimenopausal risk factors and future health | Inappropriate study design |
| 3156 | Sony PlayStation EyeToy elicits higher levels of movement than the Nintendo Wii: implications for stroke rehabilitation | Inappropriate study design |
| 3157 | Impaired ventricular filling limits cardiac reserve during submaximal exercise in people with type 2 diabetes | Inappropriate outcome |
| 3158 | Gender differences in the longitudinal associations of depressive symptoms and leisure-time physical activity with cognitive decline in ≥57year-old Taiwanese | Inappropriate study design |
| 3159 | Hemodynamics during active and passive recovery from a single bout of supramaximal exercise | Inappropriate study design |
| 3160 | Effects of acute and chronic alacepril treatment on exercise capacity and hemodynamics in patients with heart failure: a preliminary study | Inappropriate study design |
| 3161 | Lifestyle modifications to prevent and control hypertension. 4. Recommendations on physical exercise training. Canadian Hypertension Society, Canadian Coalition for High Blood Pressure Prevention and Control, Laboratory Centre for Disease Control at Health Canada, Heart and Stroke Foundation of Canada | Inappropriate study design |
| 3162 | Relationship between ambulatory capacity and cardiorespiratory fitness in chronic stroke: influence of stroke-specific impairments | Inappropriate population |
| 3163 | Inactive lifestyles and sedentary behavior in persons with chronic aneurysmal subarachnoid hemorrhage: evidence from accelerometer-based activity monitoring | Inappropriate study design |
| 3164 | Changes in bone density and geometry of the radius in chronic stroke and related factors: a one-year prospective study | Inappropriate study design |
| 3165 | Prior moderate and heavy exercise accelerate oxygen uptake and cardiac output kinetics in endurance athletes | Inappropriate study design |
| 3166 | Cardiovascular biomarkers and carotid IMT scores as predictors of cognitive function | Inappropriate study design |
| 3167 | Low-level dynamic exercises for earlier cardiac rehabilitation: aerobic and hemodynamic responses | Inappropriate study design |
| 3168 | The effect of acute hypoxia on left ventricular function during exercise | Inappropriate population |
| 3169 | Effect of treadmill exercise training on spatial and temporal gait parameters in subjects with chronic stroke: a preliminary report | Inappropriate outcome |
| 3170 | Potential Effects on Mortality of Replacing Sedentary Time With Short Sedentary Bouts or Physical Activity: A National Cohort Study | Inappropriate intervention |
| 3171 | Impact of physical activity on cardiovascular events in patients with chronic heart failure. A multicenter prospective cohort study | Inappropriate study design |
| 3172 | Relationship between maximal oxygen uptake and left ventricular function in exercise | Inappropriate population |
| 3173 | Cardiovascular responses to exhaustive upright cycle exercise in highly trained older men | Inappropriate population |
| 3174 | Similar barriers and facilitators to physical activity across different clinical groups experiencing lower limb spasticity | Inappropriate study design |
| 3175 | The effects of exercise on cognitive function and brain plasticity - a feasibility trial | Inappropriate study design |
| 3176 | Taekwondo Enhances Cognitive Function as a Result of Increased Neurotrophic Growth Factors in Elderly Women | Inappropriate study design |
| 3177 | Differential Post-Exercise Blood Pressure Responses between Blacks and Caucasians | Inappropriate intervention |
| 3178 | Risk factors assessed in adolescence and the later risk of stroke in men: a 33-year follow-up study | Inappropriate study design |
| 3179 | Nisoldipine. Central haemodynamics at rest and during exercise in essential hypertension: acute and chronic studies | Inappropriate population |
| 3180 | Aerobic exercise training in healthy postmenopausal women: effects of hormone therapy | Inappropriate study design |
| 3181 | Cardiovascular fitness is improved post-stroke with upper-limb Wii-based Movement Therapy but not dose-matched constraint therapy | Inappropriate study design |
| 3182 | Arterial blood pressure response to rowing | Inappropriate study design |
| 3183 | Resistance training reduces the blood pressure response of older men during submaximum aerobic exercise | Inappropriate population |
| 3184 | Attitudes toward exercise following participation in an exercise intervention study | Inappropriate study design |
| 3185 | Evaluation of a Physical Activity Behavior Change Program for Individuals With a Brain Injury | Inappropriate study design |
| 3186 | Quantitative measurement of physical activity in acute ischemic stroke and transient ischemic attack | Inappropriate study design |

| | | |
|---|---|---|
| 3187 | Sleep Duration, Sedentary Behavior, Physical Activity, and Quality of Life after Inpatient Stroke Rehabilitation | Inappropriate study design |
| 3188 | Short-term recovery of limb muscle strength after acute stroke | Inappropriate population |
| 3189 | What maintains energy supply at peak aerobic exercise in trained and untrained older men? | Inappropriate study design |
| 3190 | Addressing the nonexercise part of the activity continuum: a more realistic and achievable approach to activity programming for adults with mobility disability? | Inappropriate population |
| 3191 | Effect of resistance training on blood oxidative stress in Parkinson disease | Inappropriate study design |
| 3192 | The effect of critical speed and exercise intensity on stroke phase duration and bilateral asymmetry in 200-m front crawl swimming | Inappropriate study design |
| 3193 | Fatigue, as measured using the Modified Fatigue Impact Scale, is a predictor of processing speed improvement induced by exercise in patients with multiple sclerosis: data from a randomized controlled trial | Inappropriate study design |
| 3194 | COPD is associated with cognitive dysfunction and poor physical fitness in heart failure | Inappropriate population |
| 3195 | Self-Reported Omega-3 Supplement Use Moderates the Association between Age and Exercising Cerebral Blood Flow Velocity in Older Adults | Inappropriate study design |
| 3196 | Feasibility of a virtual exercise coach to promote walking in community-dwelling persons with Parkinson disease | Inappropriate population |
| 3197 | The role of postural control in the association between aerobic capacity and walking capacity in chronic stroke: a cross-sectional analysis | Inappropriate study design |
| 3198 | Lifelong Physical Activity Regardless of Dose Is Not Associated With Myocardial Fibrosis | Inappropriate population |
| 3199 | Cardiac power during exercise and the risk of stroke in men | Inappropriate study design |
| 3200 | Feasibility of adapted aerobic cycle ergometry tasks to encourage paretic limb use after stroke: a case series | Inappropriate study design |
| 3201 | Relationship between arterial baroreflex sensitivity and exercise capacity in patients with acute myocardial infarction | Inappropriate population |
| 3202 | Strength Training for Skeletal Muscle Endurance after Stroke | Inappropriate study design |
| 3203 | Intermittent exercise test in chronic obstructive pulmonary disease patients: how do the pulmonary hemodynamics adapt? | Inappropriate population |
| 3204 | Modifiable Factors Associated With Poststroke Physical Activity at Discharge From Rehabilitation: Prospective Cohort Study | Inappropriate outcome |
| 3205 | Right ventricular dysfunction and the exercise limitation of chronic obstructive pulmonary disease | Inappropriate study design |
| 3206 | Anaemia and reduced exercise capacity in patients on chronic haemodialysis | Inappropriate study design |
| 3207 | Acute and long-term effects of vasodilator therapy on resting and exercise hemodynamics and exercise tolerance | Inappropriate study design |
| 3208 | Randomized controlled trial of home-based exercise training to evaluate cardiac functional gains | Inappropriate population |
| 3209 | Attenuation of neuropsychiatric symptoms and caregiver burden in Alzheimer's disease by motor intervention: a controlled trial | Inappropriate population |
| 3210 | Increased Brain Glucose Uptake After 12 Weeks of Aerobic High-Intensity Interval Training in Young and Older Adults | Inappropriate population |
| 3211 | Hemodynamic responses during leg press exercise in patients with chronic congestive heart failure | Inappropriate population |
| 3212 | Are personal characteristics associated with exercise participation in patients with rheumatoid arthritis? A cross-sectional explorative survey | Inappropriate study design |
| 3213 | Cardiovascular Responses During Resistance Exercise in Patients With Parkinson Disease | Inappropriate population |
| 3214 | Experiences of older people with dementia participating in a high-intensity functional exercise program in nursing homes: "While it's tough, it's useful" | Inappropriate study design |
| 3215 | Consumer-Based Physical Activity Monitor as a Practical Way to Measure Walking Intensity During Inpatient Stroke Rehabilitation | Inappropriate study design |
| 3216 | Efficacy of felodipine in chronic congestive heart failure: a placebo controlled haemodynamic study at rest and during exercise and orthostatic stress | Inappropriate study design |
| 3217 | Acute haemodynamic effects of metoprolol in hypertensive patients | Inappropriate study design |
| 3218 | Central artery stiffness, neuropsychological function, and cerebral perfusion in sedentary and endurance-trained middle-aged adults | Inappropriate population |
| 3219 | The Association of Sport and Exercise Activities With Cardiovascular Disease Risk: The Atherosclerosis Risk in Communities (ARIC) Study | Inappropriate population |
| 3220 | Risk stratification for large artery or chronic coronary artery disease-related ischemic stroke in octogenarians undergoing exercise stress myocardial perfusion imaging: A cohort study | Inappropriate population |

| | | |
|---|---|---|
| 3221 | Aerobic exercise training improves right- and left ventricular systolic function in patients with COPD | Inappropriate study design |
| 3222 | Increased heart rate variability and executive performance after aerobic training in the elderly | Inappropriate population |
| 3223 | Combined effects of lactotripeptide and aerobic exercise on cognitive function and cerebral oxygenation in middle-aged and older adults | Inappropriate study design |
| 3224 | Effects of running on adiponectin, insulin and cytokines in cerebrospinal fluid in healthy young individuals | Inappropriate study design |
| 3225 | Evaluation of exercise capacity after severe stroke using robotics-assisted treadmill exercise: a proof-of-concept study | Inappropriate study design |
| 3226 | Physical activity participation may offset some of the negative impact of diabetes on cognitive function | Inappropriate study design |
| 3227 | Comparative effects of acute volume expansion and short-term training on thermal and cardiovascular responses to prolonged exercise | Inappropriate population |
| 3228 | A questionnaire survey to assess lower urinary tract symptoms in patients with chronic stroke | Inappropriate population |
| 3229 | Oxygen uptake and cardiac output during submaximal and maximal exercise in adult subjects with totally corrected tetralogy of fallot | Inappropriate population |
| 3230 | Strength Training and the Risk of Type 2 Diabetes and Cardiovascular Disease | Inappropriate population |
| 3231 | Handgrip exercise by the non-affected hand increases venous return in the contralateral axillary vein in patients with stroke: a pilot study | Inappropriate study design |
| 3232 | Effects of an exercise program to increase hip abductor muscle strength and improve lateral stability following stroke: a single subject design | Inappropriate population |
| 3233 | Gradual reduction in exercise capacity in chronic kidney disease is associated with systemic oxygen delivery factors | Inappropriate population |
| 3234 | Relation of cognitive activity to risk of developing Alzheimer disease | Inappropriate study design |
| 3235 | "Graded Cycling Test with Talk Test" Is a Reliable Test to Monitor Cardiovascular Fitness in Patients with Minor Stroke | Inappropriate study design |
| 3236 | Initial blood pressure fall on stand up and exercise explained by changes in total peripheral resistance | Inappropriate population |
| 3237 | Stair climbing test post-stroke: feasibility, convergent validity and metabolic, cardiac, and respiratory responses | Inappropriate study design |
| 3238 | An exercise intervention during acute hospitalization improved function and cognition at discharge in older adults | Inappropriate study design |
| 3239 | Cardiorespiratory response during exercise in patients with cyanotic congenital heart disease with and without a Fontan operation and in patients with congestive heart failure | Inappropriate study design |
| 3240 | Is There Evidence That Active Videogames Increase Energy Expenditure and Exercise Intensity for People Poststroke and with Cerebral Palsy? | Inappropriate population |
| 3241 | Cardiorespiratory and cerebrovascular responses to head-up tilt II: influence of age, training status and acute exercise | Inappropriate study design |
| 3242 | Trajectories in Leisure-Time Physical Activity and Risk of Stroke in Women in the California Teachers Study | Inappropriate study design |
| 3243 | Cardiovascular responses to progressive cycle exercise in healthy children and adults | Inappropriate population |
| 3244 | The association between objectively measured physical activity, depression, cognition, and health-related quality of life in Parkinson's disease | Inappropriate study design |
| 3245 | Continuum of cardiovascular performance across a broad range of fitness levels in healthy older men | Inappropriate study design |
| 3246 | Cardiorespiratory responses to arm cranking and electrical stimulation leg cycling in people with paraplegia | Inappropriate study design |
| 3247 | Levels and Patterns of Physical Activity and Sedentary Behavior in Elderly People With Mild to Moderate Parkinson Disease | Inappropriate study design |
| 3248 | Haemodynamic long-term effects of beta-receptor-blocking agents in hypertension: a comparison between alprenolol, atenolol, metoprolol and timolol | Inappropriate population |
| 3249 | The basis for disease-modifying treatments for Alzheimer's disease: the Sixth Annual Mild Cognitive Impairment Symposium | Inappropriate population |
| 3250 | Association of Accelerometer-Measured Light-Intensity Physical Activity With Brain Volume: The Framingham Heart Study | Inappropriate population |
| 3251 | Hemodynamic Responses to Blood Flow Restriction and Resistance Exercise to Muscular Failure | Inappropriate study design |
| 3252 | Functional electrical stimulation leg cycle ergometer exercise: training effects on cardiorespiratory responses of spinal cord injured subjects at rest and during submaximal exercise | Inappropriate study design |

| | | |
|---|---|---|
| 3253 | Effect of high intensity exercise training on central hemodynamic responses to exercise in men with reduced left ventricular function | Inappropriate study design |
| 3254 | Effects of Combined Resistance and Power Training on Cognitive Function in Older Women: A Randomized Controlled Trial | Inappropriate population |
| 3255 | Circadian profile of systemic hemodynamics | Inappropriate population |
| 3256 | Effects of a Single Hand-Exercise Session on Manual Dexterity and Strength in Persons with Parkinson Disease: A Randomized Controlled Trial | Inappropriate study design |
| 3257 | Polygenic burden associated to oligodendrocyte precursor cells and radial glia influences the hippocampal volume changes induced by aerobic exercise in schizophrenia patients | Inappropriate study design |
| 3258 | Changes in the haemodynamic responses to exercise, posture and nitrates after treatment of congestive heart failure with an arteriolar vasodilator | Inappropriate outcome |
| 3259 | Respiratory muscle strength predicts decline in mobility in older persons | Inappropriate study design |
| 3260 | Impact of chronic exercise training on the blood pressure response to orthostatic stimulation | Inappropriate outcome |
| 3261 | Hemodynamic and antiischemic effects of intravenous elgodipine, a new dihydropyridine calcium channel blocker, in patients with chronic stable angina | Inappropriate intervention |
| 3262 | Individualized exercise program for the treatment of severe fatigue in patients after allogeneic hematopoietic stem-cell transplant: a pilot study | Inappropriate population |
| 3263 | Working mechanisms of a behavioural intervention promoting physical activity in persons with subacute spinal cord injury | Inappropriate study design |
| 3264 | Exertional heat stroke and acute renal failure in a young woman | Inappropriate study design |
| 3265 | A Multidomain Intervention for Modifying Lifestyle Habits Reduces the Dementia Risk in Community-Dwelling Older Adults: A Single-Blinded Randomized Controlled Pilot Study | Inappropriate population |
| 3266 | Cardiovascular, metabolic, and ventilatory responses of women to equivalent cycle ergometer and treadmill exercise | Inappropriate population |
| 3267 | The effects of exergaming and treadmill training on gait, balance, and cognition in a person with Parkinson's disease: A case study | Inappropriate study design |
| 3268 | Sildenafil treatment in COPD does not affect stroke volume or exercise capacity | Inappropriate population |
| 3269 | A meta-analytic review of the effects of exercise on brain-derived neurotrophic factor | Inappropriate study design |
| 3270 | Arterial vasodilator, systemic and coronary hemodynamic effects of nisoldipine in congestive heart failure secondary to ischemic or dilated cardiomyopathy | Inappropriate study design |
| 3271 | Cardiopulmonary responses to exercise in swimmer using a swim bench and a leg-kicking ergometer | Inappropriate study design |
| 3272 | Rest and exercise cardiovascular effects of terazosin in congestive heart failure | Inappropriate population |
| 3273 | Out-of-hospital cardiac arrest survivors with cognitive impairments have lower exercise capacity | Inappropriate study design |
| 3274 | Cardiac rehabilitation is associated with lasting improvements in cognitive function in older adults with heart failure | Inappropriate population |
| 3275 | Cognitive effects of weight-shifting controlled exergames in patients with chronic stroke: a pilot randomized comparison trial | Inappropriate study design |
| 3276 | Physical activity and 5-year cognitive decline in the Doetinchem cohort study | Inappropriate study design |
| 3277 | Effect of Physical Fatigue Elicited by Continuous and Intermittent Exercise on Motor Imagery Ability | Inappropriate study design |
| 3278 | Intensity, but not duration, of physical activities is related to cognitive function | Inappropriate study design |
| 3279 | Cardiovascular adaptations to exercise training after uncomplicated acute myocardial infarction | Inappropriate study design |
| 3280 | Acute intravenous and sustained oral treatment with the beta1 agonist prenalterol in patients with chronic severe cardiac failure | Inappropriate study design |
| 3281 | Control of skin blood flow during exercise by thermal reflexes and baroreflexes | Inappropriate outcome |
| 3282 | Maintaining health and wellness in the face of dementia: an exploratory analysis of individuals attending a rural and remote memory clinic | Inappropriate study design |
| 3283 | Experiences of an exercise referral scheme from the perspective of people with chronic stroke: a qualitative study | Inappropriate population |
| 3284 | Exercise performance and VO2 kinetics during upright and recumbent high-intensity cycling exercise | Inappropriate study design |
| 3285 | Randomized Controlled Trial Evaluating Aerobic Training and Common Sport-Related Concussion Outcomes in Healthy Participants | Inappropriate population |
| 3286 | Increased masticatory activity and quality of life in elderly persons with dementia--a longitudinal matched cluster randomized single-blind multicenter intervention study | Inappropriate study design |

| | | |
|---|---|---|
| 3287 | Preliminary study of exercise capacity in post-acute stroke survivors | Inappropriate population |
| 3288 | Exercise training to improve motor performance in chronic stroke: effects of a community-based exercise program | Inappropriate intervention |
| 3289 | Contemporary thinking in stroke prevention and management | Inappropriate study design |
| 3290 | Improvements in gait characteristics after intensive resistance and functional training in people with dementia: a randomised controlled trial | Inappropriate population |
| 3291 | The effects of exercise with TENS on spasticity, balance, and gait in patients with chronic stroke: a randomized controlled trial | Inappropriate population |
| 3292 | Longitudinal changes in exercise capacity after stroke | Inappropriate study design |
| 3293 | Transient autonomic responses during sustained attention in high and low fit young adults | Inappropriate study design |
| 3294 | Physical Activity and Cognitive Function among Older Adults with an Elevated Gamma Gap | Inappropriate study design |
| 3295 | Mild cognitive impairment and depressive symptoms in elderly patients with diabetes: prevalence, risk factors, and comorbidity | Inappropriate population |
| 3296 | Cardiovascular reserve capacity in healthy older humans | Inappropriate population |
| 3297 | Effects of chronic exercise on cardiac output and its determinants | Inappropriate study design |
| 3298 | Effects of chronic atenolol therapy on cardiovascular response to isometric exercise in essential hypertension | Inappropriate study design |
| 3299 | Virtual reality exergaming as adjunctive therapy in a sub-acute stroke rehabilitation setting: facilitators and barriers | Inappropriate population |
| 3300 | A Twelve-Week Moderate Exercise Programme Improved Symptoms of Depression, Insomnia, and Verbal Learning in Post-Aneurysmal Subarachnoid Haemorrhage Patients: A Comparison with Meningioma Patients and Healthy Controls | Inappropriate study design |
| 3301 | Pathophysiology of Noncardiac Syncope in Athletes | Inappropriate population |
| 3302 | Inactive and alone: physical activity within the first 14 days of acute stroke unit care | Inappropriate study design |
| 3303 | Acute increases in brain-derived neurotrophic factor in plasma following physical exercise relates to subsequent learning in older adults | Inappropriate study design |
| 3304 | Chronic stroke patients show early and robust improvements in muscle and functional performance in response to eccentric-overload flywheel resistance training: a pilot study | Inappropriate population |
| 3305 | The stroke volume in juvenile borderline hypertension | Inappropriate study design |
| 3306 | Detection of left ventricular functional reserve by supine exercise hemodynamics in patients with severe, chronic heart failure | Inappropriate population |
| 3307 | Change of Objectively-Measured Physical Activity during Geriatric Rehabilitation | Inappropriate study design |
| 3308 | Cardiorespiratory fitness: A predictor of cortical plasticity in multiple sclerosis | Inappropriate study design |
| 3309 | Repeated Valsalva maneuvers promote symptomatic manifestations of cerebral microhemorrhages: implications for the pathogenesis of vascular cognitive impairment in older adults | Inappropriate study design |
| 3310 | Physical activity in incident patients with pulmonary arterial and chronic thromboembolic hypertension | Inappropriate study design |
| 3311 | Association between physical activity and stroke in a middle-aged and elderly Chinese population | Inappropriate intervention |
| 3312 | Chronic exercise preserves brain function in masters athletes when compared to sedentary counterparts | Inappropriate population |
| 3313 | Glycemic control and cardiopulmonary function in patients with insulin-dependent diabetes mellitus | Inappropriate study design |
| 3314 | Patterns of non-response to a mail survey | Inappropriate study design |
| 3315 | High Intensity Exercise for Walking Competency in Individuals with Stroke: A Systematic Review and Meta-Analysis | Inappropriate study design |
| 3316 | Effortful swallow with resistive electrical stimulation training improves pharyngeal constriction in patients post-stroke with dysphagia | Inappropriate population |
| 3317 | Study protocol: a randomised controlled trial of the effects of a multi-modal exercise program on cognition and physical functioning in older women | Inappropriate outcome |
| 3318 | Pulmonary vascular responsiveness at rest and during exercise in idiopathic pulmonary fibrosis: effects of oxygen and nifedipine | Inappropriate study design |
| 3319 | Association between mid- to late life physical fitness and dementia: evidence from the CAIDE study | Inappropriate population |
| 3320 | Enhancing memory in late-life depression: the effects of a combined psychoeducation and cognitive training program | Inappropriate study design |

| | | |
|---|---|---|
| 3321 | Single and combined therapy for systemic hypertension with propranolol, hydralazine and hydrochlorothiazide: hemodynamic and neuroendocrine mechanisms of action | Inappropriate study design |
| 3322 | Sedentary behavior in the first year after stroke: a longitudinal cohort study with objective measures | Inappropriate population |
| 3323 | Central cardiac limit to aerobic capacity in patients with exertional pulmonary venous hypertension: implications for heart failure with preserved ejection fraction | Inappropriate outcome |
| 3324 | Outcomes in people after stroke attending an adapted cardiac rehabilitation exercise program: does time from stroke make a difference? | Inappropriate study design |
| 3325 | Auditory and visual cueing modulate cycling speed of older adults and persons with Parkinson's disease in a Virtual Cycling (V-Cycle) system | Inappropriate study design |
| 3326 | Stepping towards prevention of bone loss after stroke: a systematic review of the skeletal effects of physical activity after stroke | Inappropriate study design |
| 3327 | Aerobic exercise and consecutive task-specific training (AExaCTT) for upper limb recovery after stroke: A randomized controlled pilot study | Inappropriate study design |
| 3328 | The effect of long-term aerobic exercise on maximal oxygen consumption, left ventricular function and serum lipids in elderly women | Inappropriate population |
| 3329 | Improved cardiac performance by salbutamol, a selective beta 2-agonist, in chronic cor pulmonale | Inappropriate study design |
| 3330 | The obesity epidemic: incidence and prevalence | Inappropriate population |
| 3331 | The right ventricle: physiologic and pathophysiologic considerations | Inappropriate outcome |
| 3332 | Effects of Moderate-To-Vigorous Physical Activity on Cancer-Related Fatigue in Patients with Colorectal Cancer: A Systematic Review and Meta-Analysis | Inappropriate study design |
| 3333 | Little therapy, little physical activity: rehabilitation within the first 14 days of organized stroke unit care | Inappropriate study design |
| 3334 | Physical activity frequency and risk of incident stroke in a national US study of blacks and whites | Inappropriate study design |
| 3335 | Methods to improve constraint-induced movement therapy | Inappropriate study design |
| 3336 | Managing atrial fibrillation to prevent its major complication: ischemic stroke | Inappropriate population |
| 3337 | How much exercise does the enhanced gait-oriented physiotherapy provide for chronic stroke patients? | Inappropriate population |
| 3338 | Short- and long-term (six-year) hemodynamic effects of labetalol in essential hypertension | Inappropriate study design |
| 3339 | Differences in echocardiography, blood pressure, stroke volume, maximal power and profile of genes related to cardiac hypertrophy in elite road cyclists | Inappropriate intervention |
| 3340 | Training dual-task walking in community-dwelling adults within 1 year of stroke: a protocol for a single-blind randomized controlled trial | Inappropriate study design |
| 3341 | Blood pressure and heart rate adjustment following acute Frenkel's ambulatory exercise in chronic hemiparetics stroke survivors: a comparative study | Inappropriate study design |
| 3342 | Effects of a high-intensity task-oriented training on gait performance early after stroke: a pilot study | Inappropriate study design |
| 3343 | Evaluation of dementia-prevention classes for community-dwelling older adults with mild cognitive impairment | Inappropriate population |
| 3344 | An assessment of the relationship of physical activity, obesity, and chronic diseases/conditions between active/obese and sedentary/ normal weight American women in a national sample | Inappropriate population |
| 3345 | The effect of water-based exercises on balance in persons post-stroke: a randomized controlled trial | Inappropriate study design |
| 3346 | Physical activity and stroke incidence: the Harvard Alumni Health Study | Inappropriate study design |
| 3347 | Peak exercise stroke volume: associations with cardiac structure and diastolic function | Inappropriate population |
| 3348 | Cardiorespiratory fitness and nonfatalcardiovascular events: A population-based follow-up study | Inappropriate study design |
| 3349 | Attenuation of cardiovascular adaptations to exercise in frail octogenarians | Inappropriate study design |
| 3350 | Single limb exercise: pilot study of physiological and functional responses to forced use of the hemiparetic lower extremity | Inappropriate study design |
| 3351 | Oral N-acetylcysteine and exercise tolerance in mild chronic obstructive pulmonary disease | Inappropriate study design |
| 3352 | Strengthening interventions increase strength and improve activity after stroke: a systematic review | Inappropriate study design |
| 3353 | Physical activity correlates with arterial stiffness in community-dwelling individuals with stroke | Inappropriate study design |

| | | |
|---|---|---|
| 3354 | A 19-week exercise program for people with chronic stroke enhances bone geometry at the tibia: a peripheral quantitative computed tomography study | Inappropriate study design |
| 3355 | The home stroke rehabilitation and monitoring system trial: a randomized controlled trial | Inappropriate study design |
| 3356 | Usability evaluation of a kinematics focused Kinect therapy program for individuals with stroke | Inappropriate study design |
| 3357 | Is there a preference for PET or SPECT brain imaging in diagnosing dementia? The views of people with dementia, carers, and healthy controls | Inappropriate study design |
| 3358 | Effect of Baduanjin exercise on cognitive function in patients with post-stroke cognitive impairment: study protocol for a randomised controlled trial | Inappropriate intervention |
| 3359 | Similarities among factors associated with components of frailty in elderly: SABE Study | Inappropriate population |
| 3360 | Cardiovascular response to treadmill testing in Parkinson disease | Inappropriate outcome |
| 3361 | Association between physical activity and sedentary behaviour on carotid atherosclerotic plaques: an epidemiological and histological study in 90 asymptomatic patients | Inappropriate study design |
| 3362 | Effect of 24-month physical activity on cognitive frailty and the role of inflammation: the LIFE randomized clinical trial | Inappropriate study design |
| 3363 | The short physical performance battery as a predictor of functional capacity after stroke | Inappropriate study design |
| 3364 | Progressive left ventricular dysfunction and remodeling after myocardial infarction. Potential mechanisms and early predictors | Inappropriate study design |
| 3365 | ACE insertion/deletion polymorphism and submaximal exercise hemodynamics in postmenopausal women | Inappropriate study design |
| 3366 | Ambulatory activity of stroke survivors: measurement options for dose, intensity, and variability of activity | Inappropriate study design |
| 3367 | A yoga-based exercise program for people with chronic poststroke hemiparesis | Inappropriate study design |
| 3368 | A nurse manager's guide to support patients with dementia | Inappropriate population |
| 3369 | Leisure-Time Physical Activity and Cardiovascular Mortality in an Elderly Population in Northern Manhattan: A Prospective Cohort Study | Inappropriate study design |
| 3370 | High-volume plasma exchange in a patient with acute liver failure due to non-exertional heat stroke in a sauna | Inappropriate study design |
| 3371 | Preliminary safety analysis of high-intensity interval training (HIIT) in persons with chronic stroke | Inappropriate study design |
| 3372 | Cardiac resynchronization therapy modulation of exercise left ventricular function and pulmonary $O_2$ uptake in heart failure | Inappropriate population |
| 3373 | Regular aerobic exercise and the age-related increase in carotid artery intima-media thickness in healthy men | Inappropriate study design |
| 3374 | After-effects of exercise on haemodynamics and muscle sympathetic nerve activity in young patients with dilated cardiomyopathy | Inappropriate population |
| 3375 | Diabetes in old age: an emerging epidemic | Inappropriate study design |
| 3376 | Health-enhancing physical activity programme (HEPAP) for transient ischaemic attack and non-disabling stroke: recruitment and compliance | Inappropriate population |
| 3377 | Sensory-motor and cardiorespiratory sensory rehabilitation associated with transcranial photobiomodulation in patients with central nervous system injury: Trial protocol for a single-center, randomized, double-blind, and controlled clinical trial | Inappropriate outcome |
| 3378 | Cardiac paraganglioma presenting with acute myocardial infarction and stroke | Inappropriate study design |
| 3379 | Older People with Dementia Have Reduced Daily-Life Activity and Impaired Daily-Life Gait When Compared to Age-Sex Matched Controls | Inappropriate population |
| 3380 | Captopril in primary pulmonary hypertension | Inappropriate population |
| 3381 | The Burden of Stroke Scale (BOSS) provides valid and reliable score estimates of functioning and well-being in stroke survivors with and without communication disorders | Inappropriate study design |
| 3382 | Rationale and design of a single-blind, randomised controlled trial of exercise training for managing learning and memory impairment in persons with multiple sclerosis | Inappropriate population |
| 3383 | Physical fitness and cardiovascular regulation: mechanisms of orthostatic intolerance | Inappropriate study design |
| 3384 | Effect of total, domain-specific, and intensity-specific physical activity on all-cause and cardiovascular mortality among hypertensive adults in China | Inappropriate study design |
| 3385 | Altered hemodynamics during exercise in older essential hypertensive subjects | Inappropriate study design |
| 3386 | Short and Long-Term Trainability in Older Adults: Training and Detraining Following Two Years of Multicomponent Cognitive-Physical Exercise Training | Inappropriate outcome |

| | | |
|---|---|---|
| 3387 | Enhanced left ventricular performance in endurance trained older men | Inappropriate outcome |
| 3388 | Hemodynamic Responses and Perceived Exertion During Continuous and Discontinuous Resistance Exercise | Inappropriate study design |
| 3389 | Stroke volume increase to exercise in chronic obstructive pulmonary disease is limited by increased pulmonary artery pressure | Inappropriate study design |
| 3390 | The Feasibility and Longitudinal Effects of a Home-Based Sedentary Behavior Change Intervention After Stroke | Inappropriate intervention |
| 3391 | Blunted cerebrovascular response is associated with elevated beta-amyloid | Inappropriate study design |
| 3392 | Mediterranean Lifestyle in Relation to Cognitive Health: Results from the HELIAD Study | Inappropriate population |
| 3393 | Cognitive Tests and Determining Fitness to Drive in Dementia: A Systematic Review | Inappropriate study design |
| 3394 | Ascending aortic blood flow dynamics following intense exercise | Inappropriate study design |
| 3395 | Dystypia in acute stroke not attributable to aphasia or neglect | Inappropriate study design |
| 3396 | Frequency and duration of interval training programs and changes in aerobic power | Inappropriate population |
| 3397 | Role of calcium antagonists in the treatment of essential hypertension | Inappropriate study design |
| 3398 | Treadmill training to improve mobility for people with sub-acute stroke: a phase II feasibility randomized controlled trial | Inappropriate intervention |
| 3399 | Reported Prestroke Physical Activity Is Associated with Vascular Endothelial Growth Factor Expression and Good Outcomes after Stroke | Inappropriate population |
| 3400 | Vasodilators as first-line therapy for congestive heart failure: a comparative hemodynamic study of hydralazine, digoxin, and their combination | Inappropriate study design |
| 3401 | The relevance of nutrition for the concept of cognitive frailty | Inappropriate study design |
| 3402 | The role of caregiver involvement in upper-limb treatment in individuals with subacute stroke | Inappropriate study design |
| 3403 | Vestibular rehabilitation in acute central vestibulopathy: a randomized controlled trial | Inappropriate population |
| 3404 | The feasibility of cardiopulmonary exercise testing for prescribing exercise to people after stroke | Inappropriate population |
| 3405 | Cardiorespiratory fitness of post-stroke patients: as inpatients and as outpatients | Inappropriate study design |
| 3406 | Oxygen uptake and cardiac output at rest and during exercise after surgery for coarctation of the aorta | Inappropriate study design |
| 3407 | Self-reported free-living physical activity and executive control in young adults | Inappropriate population |
| 3408 | Robotic upper-limb neurorehabilitation in chronic stroke patients | Inappropriate population |
| 3409 | Age and cardiac output during cycle exercise in thermoneutral and warm environments | Inappropriate study design |
| 3410 | Response to upright exercise after myocardial infarction | Inappropriate population |
| 3411 | Dynamic Resistance Training Improves Cardiac Autonomic Modulation and Oxidative Stress Parameters in Chronic Stroke Survivors: A Randomized Controlled Trial | Inappropriate study design |
| 3412 | Attenuated relationship between cardiac output and oxygen uptake during high-intensity exercise | Inappropriate outcome |
| 3413 | Increasing motivation in robot-aided arm rehabilitation with competitive and cooperative gameplay | Inappropriate study design |
| 3414 | Physical Activity Frequency and the Risk of Stroke: A Nationwide Cohort Study in Korea | Inappropriate population |
| 3415 | Physical activity and risk of cognitive impairment among older persons living in the community | Inappropriate study design |
| 3416 | Whole-Body Vibration Intensities in Chronic Stroke: A Randomized Controlled Trial | Inappropriate study design |
| 3417 | Arthritis and cognitive impairment in older adults | Inappropriate population |
| 3418 | Use of an ankle-foot orthosis improves aerobic capacity in subacute hemiparetic stroke patients | Inappropriate study design |
| 3419 | Acute hypervolemia, cardiac performance, and aerobic power during exercise | Inappropriate study design |
| 3420 | Physical activity and executive function in aging: the MOBILIZE Boston Study | Inappropriate study design |
| 3421 | Hemodynamic response to oxygen therapy in chronic obstructive pulmonary disease | Inappropriate study design |
| 3422 | Measurement properties of maximal cardiopulmonary exercise tests protocols in persons after stroke: A systematic review | Inappropriate population |
| 3423 | Exercise testing and training of the elderly patient | Inappropriate intervention |

| | | |
|---|---|---|
| 3424 | Early prediction of physical inactivity in 215 individuals 6 months after a mild stroke: The fall study of Gothenburg | Inappropriate outcome |
| 3425 | Alteration of circadian periodicity in core body temperatures of patients with acute stroke | Inappropriate study design |
| 3426 | Leisure-time physical activity and ischemic stroke risk: the Northern Manhattan Stroke Study | Inappropriate study design |
| 3427 | Lifestyle and vascular risk effects on MRI-based biomarkers of Alzheimer's disease: a cross-sectional study of middle-aged adults from the broader New York City area | Inappropriate study design |
| 3428 | A randomized controlled trial of an activity specific exercise program for individuals with Alzheimer disease in long-term care settings | Inappropriate study design |
| 3429 | A step activity monitoring program improves real world walking activity post stroke | Inappropriate population |
| 3430 | Assessing Chinese coach drivers' fitness to drive: The development of a toolkit based on cognition measurements | Inappropriate study design |
| 3431 | Impact of aerobic interval training and continuous training on left ventricular geometry and function: a SAINTEX-CAD substudy | Inappropriate study design |
| 3432 | How should long-term free-living physical activity be targeted after stroke? A systematic review and narrative synthesis | Inappropriate intervention |
| 3433 | Left ventricular function at rest and during exercise in acute hypothyroidism | Inappropriate study design |
| 3434 | Effects of dynamic exercise on cardiovascular regulation during lower body negative pressure | Inappropriate population |
| 3435 | Impact of Function Focused Care and Physical Activity on Falls in Assisted Living Residents | Inappropriate population |
| 3436 | Preliminary trial of symmetry-based resistance in individuals with post-stroke hemiparesis | Inappropriate study design |
| 3437 | Haemodynamic responses to exercise and acute beta-receptor blockade in renin sub-types of essential hypertension | Inappropriate study design |
| 3438 | Association of diet, exercise, and smoking modification with risk of early cardiovascular events after acute coronary syndromes | Inappropriate population |
| 3439 | Is a coded physical activity diary valid for assessing physical activity level and energy expenditure in stroke patients? | Inappropriate study design |
| 3440 | Sedentary behavior and cardiovascular disease in older women: The Objective Physical Activity and Cardiovascular Health (OPACH) Study | Inappropriate population |
| 3441 | Acute haemodynamic effects of carvedilol in essential hypertension at rest and during exercise | Inappropriate study design |
| 3442 | Socio-demographic and health-related factors associated with cognitive impairment in the elderly in Taiwan | Inappropriate study design |
| 3443 | Physical Activity, Physical Function, and Quadriceps Muscle Thickness in Male Patients with Sub-Acute Stroke during Hospitalization: A Pilot Study | Inappropriate population |
| 3444 | WISE 2005: stroke volume changes contribute to the pressor response during ischemic handgrip exercise in women | Inappropriate population |
| 3445 | Motivation as a mechanism underpinning exercise-based falls prevention programmes for older adults with cognitive impairment: a realist review | Inappropriate population |
| 3446 | Crossfit training changes brain-derived neurotrophic factor and irisin levels at rest, after wingate and progressive tests, and improves aerobic capacity and body composition of young physically active men and women | Inappropriate study design |
| 3447 | Spatial Navigation in the Elderly with Alzheimer's Disease: A Cross-Sectional Study | Inappropriate population |
| 3448 | Cardiovascular response to submaximal stationary cycling during hemodialysis | Inappropriate study design |
| 3449 | Correlation between regional cerebral blood flow and oxidative metabolism. In vivo studies in man | Inappropriate outcome |
| 3450 | Metabolic and hemodynamic responses to concurrent voluntary arm crank and electrical stimulation leg cycle exercise in quadriplegics | Inappropriate study design |
| 3451 | The effects of movement stimulation on activities of daily living performance and quality of life in nursing home residents with dementia: a randomized controlled trial | Inappropriate study design |
| 3452 | Predictive Power of Head Impact Intensity Measures for Recognition Memory Performance | Inappropriate population |
| 3453 | Active-Arm Passive-Leg Exercise Improves Cardiovascular Function in Spinal Cord Injury | Inappropriate outcome |
| 3454 | Cardiovascular response to static handgrip in trained and untrained men | Inappropriate study design |
| 3455 | A high-intensity functional weight-bearing exercise program for older people dependent in activities of daily living and living in residential care facilities: evaluation of the applicability with focus on cognitive function | Inappropriate population |

| | | |
|---|---|---|
| 3456 | Ergometer rowing with and without slides | Inappropriate study design |
| 3457 | Cardiac effects of ACE inhibition | Inappropriate population |
| 3458 | Device-assessed physical activity and sedentary behavior in a community-based cohort of older adults | Inappropriate outcome |
| 3459 | Acute volume loading and exercise capacity in postural tachycardia syndrome | Inappropriate intervention |
| 3460 | The First 10 Years of Aerobic Exercise Responses to Long-Duration ISS Flights | Inappropriate intervention |
| 3461 | Resistance training and executive functions: a 12-month randomized controlled trial | Inappropriate study design |
| 3462 | Activity monitors for increasing physical activity in adult stroke survivors | Inappropriate study design |
| 3463 | Effects of angiotensin antagonism with tasosartan on regional and systemic haemodynamics in hypertensive patients | Inappropriate study design |
| 3464 | Cardiovascular responses during static exercise. Studies in patients with complete heart block and dual chamber pacemakers | Inappropriate study design |
| 3465 | Acute recovery profile of lung volumes and function after running 5 miles | Inappropriate study design |
| 3466 | Acute effects of sildenafil on exercise pulmonary hemodynamics and capacity in patients with COPD | Inappropriate study design |
| 3467 | Endurance training in older men and women. I. Cardiovascular responses to exercise | Inappropriate study design |
| 3468 | Upper-extremity functional electric stimulation-assisted exercises on a workstation in the subacute phase of stroke recovery | Inappropriate population |
| 3469 | The development and feasibility of treadmill-induced fall recovery training applied to individuals with chronic stroke | Inappropriate population |
| 3470 | Cardiac perfusion and function after high-intensity exercise training in late premenopausal and recent postmenopausal women: an MRI study | Inappropriate study design |
| 3471 | Electromyography-controlled exoskeletal upper-limb-powered orthosis for exercise training after stroke | Inappropriate population |
| 3472 | Role of a single session of ball throwing exercise on postural control in older adults with mild cognitive impairment | Inappropriate study design |
| 3473 | Mechanism of overshoot in cardiac function during recovery from submaximal exercise in man | Inappropriate population |
| 3474 | Task-oriented intervention in chronic stroke: changes in clinical and laboratory measures of balance and mobility | Inappropriate study design |
| 3475 | Stroke while jogging | Inappropriate study design |
| 3476 | Aquatic Exercise Therapy for People With Parkinson Disease: A Randomized Controlled Trial | Inappropriate study design |
| 3477 | Haemodynamic and radionuclide effects of amlodipine in coronary artery disease | Inappropriate study design |
| 3478 | Aging well: Processing speed inhibition and working memory related to balance and aerobic endurance | Inappropriate study design |
| 3479 | Predictors of care dependency in nursing home residents with moderate to severe dementia: A cross-sectional study | Inappropriate study design |
| 3480 | Repeated Treadmill Sprints Impair Cognitive Performance in Amateur Team-Sport Athletes When Performed in Normobaric Hypoxia | Inappropriate study design |
| 3481 | Combined health risks of cigarette smoking and low levels of physical activity: a prospective cohort study in England with 12-year follow-up | Inappropriate study design |
| 3482 | Association of Financial Literacy With Hospitalization in Community-dwelling Older Adults | Inappropriate study design |
| 3483 | Comparative Effect of Power Training and High-Speed Yoga on Motor Function in Older Patients With Parkinson Disease | Inappropriate study design |
| 3484 | Lokomat training in vascular dementia: motor improvement and beyond! | Inappropriate study design |
| 3485 | A 30-year follow-up of the Dallas Bedrest and Training Study: II. Effect of age on cardiovascular adaptation to exercise training | Inappropriate study design |
| 3486 | Cardiovascular responses to exercise when increasing skin temperature with narrowing of the core-to-skin temperature gradient | Inappropriate outcome |
| 3487 | Hemodynamic and thermal responses to a 30-minute constant-workload aerobic exercise in middle- or old-aged patients with cardiovascular diseases | Inappropriate population |
| 3488 | Muscle, functional and cognitive adaptations after flywheel resistance training in stroke patients: a pilot randomized controlled trial | Inappropriate population |
| 3489 | Cardiovascular response to exercise after the Mustard operation for simple and complex transposition of the great vessels | Inappropriate study design |
| 3490 | Acute increase in exercise capacity with milrinone: lack of correlation with resting hemodynamic responses | Inappropriate outcome |

| | | |
|---|---|---|
| 3491 | Effects of xamoterol on resting and exercise haemodynamics in patients with chronic heart failure | Inappropriate population |
| 3492 | Leisure activities and cognitive function in elderly community-dwelling individuals in Japan: a 5-year prospective cohort study | Inappropriate population |
| 3493 | Associations between cerebral amyloid and changes in cognitive function and falls risk in subcortical ischemic vascular cognitive impairment | Inappropriate population |
| 3494 | Hemodynamic adaptations to stress with advancing age | Inappropriate study design |
| 3495 | The effect of acute caffeine ingestion on upper body anaerobic exercise and cognitive performance | Inappropriate population |
| 3496 | Physical training improves motor performance in people with dementia: a randomized controlled trial | Inappropriate study design |
| 3497 | Gender differences in cardiovascular regulation during recovery from exercise | Inappropriate study design |
| 3498 | Haemodynamic effects of combined alpha- and beta-adrenoreceptor blockade after intravenous labetalol in hypertensive patients at rest and during exercise | Inappropriate population |
| 3499 | Cardiac output responses of primigravid women during exercise determined by the direct Fick technique | Inappropriate study design |
| 3500 | Severe decrements in cognition function and mood induced by sleep loss, heat, dehydration, and undernutrition during simulated combat | Inappropriate population |
| 3501 | Cardiovascular response to dynamic exercise in patients with chronic symptomatic mild-to-moderate and severe aortic regurgitation | Inappropriate study design |
| 3502 | Hemodynamic and hormonal responses to a short-term low-intensity resistance exercise with the reduction of muscle blood flow | Inappropriate study design |
| 3503 | Addressing Stroke Literacy in Nigeria Through Music: A Qualitative Study of Community Perspectives | Inappropriate study design |
| 3504 | No association between ACE I/D polymorphism and cardiovascular hemodynamics during exercise in young women | Inappropriate study design |
| 3505 | Body surface potential mapping in anterior myocardial infarction--a longitudinal study in acute, convalescent and chronic phases | Inappropriate population |
| 3506 | Isometric exercise in the denervated heart: a Doppler echocardiographic study | Inappropriate study design |
| 3507 | Physical frailty in older persons is associated with Alzheimer disease pathology | Inappropriate study design |
| 3508 | Relationship between electrocardiographic RR and QT interval variabilities and indices of ventricular function in healthy subjects | Inappropriate study design |
| 3509 | Bilateral upper limb trainer with virtual reality for post-stroke rehabilitation: case series report | Inappropriate population |
| 3510 | Dose dependence of tolerance during treatment with mononitrates | Inappropriate study design |
| 3511 | Reduction of the plasma concentration of C-reactive protein following nine months of endurance training | Inappropriate study design |
| 3512 | Passive Leg Raising Correlates with Future Exercise Capacity after Coronary Revascularization | Inappropriate population |
| 3513 | Left ventricular energetics: new insight into the plasticity of regional contributions at rest and during exercise | Inappropriate population |
| 3514 | Acute ingestion of neuromuscular enhancement supplements do not improve power output, work capacity, and cognition | Inappropriate study design |
| 3515 | Is transcutaneous peroneal stimulation beneficial to patients with chronic stroke using an ankle-foot orthosis? A within-subjects study of patients' satisfaction, walking speed and physical activity level | Inappropriate population |
| 3516 | Comparative hemodynamic and neurohormonal effects of intravenous captopril and digoxin and their combinations in patients with severe heart failure | Inappropriate population |
| 3517 | Effectiveness of Active Cycling in Subacute Stroke Rehabilitation: A Randomized Controlled Trial | Inappropriate study design |
| 3518 | Effect of nicardipine on rest and exercise hemodynamics in chronic congestive heart failure | Inappropriate population |
| 3519 | Resistance-based, reciprocal upper and lower limb locomotor training in chronic stroke: a randomized, controlled crossover study | Inappropriate study design |
| 3520 | Cardiovascular reactivity in Black South-African males of different age groups: the influence of urbanization | Inappropriate study design |
| 3521 | Exercise cardiac MRI unmasks right ventricular dysfunction in acute hypoxia and chronic pulmonary arterial hypertension | Inappropriate population |
| 3522 | Paradox of improved exercise but not resting hemodynamics with short-term prazosin in chronic heart failure | Inappropriate population |
| 3523 | Doppler echocardiography and ultrafast cine computed tomography during dynamic exercise in chronic parenchymal pulmonary disease | Inappropriate study design |

| | | |
|---|---|---|
| 3524 | Non-invasive assessment of cardiac output during exercise in chronic obstructive pulmonary disease: comparison of the CO2-rebreathing method and electrical impedance cardiography | Inappropriate study design |
| 3525 | High Prestroke Physical Activity Is Associated with Reduced Infarct Growth in Acute Ischemic Stroke Patients Treated with Intravenous tPA and Randomized to Remote Ischemic Perconditioning | Inappropriate population |
| 3526 | Sex-specific impact of aging on the blood pressure response to exercise | Inappropriate study design |
| 3527 | Cardiac autonomic dysfunction in chronic stroke women is attenuated after submaximal exercise test, as evaluated by linear and nonlinear analysis | Inappropriate study design |
| 3528 | Acute vascular and cardiovascular responses to blood flow-restricted exercise | Inappropriate study design |
| 3529 | Physical inactivity is correlated with levels of quantitative C-reactive protein in serum, independent of obesity: results of the national surveillance of risk factors of non-communicable diseases in Iran | Inappropriate population |
| 3530 | Assessment tools predicting fitness to drive in older adults: a systematic review | Inappropriate study design |
| 3531 | Difference between the response of the pulmonary and systemic circulation during exercise in congestive heart failure | Inappropriate population |
| 3532 | Haemodynamic effects of felodipine in congestive heart failure | Inappropriate population |
| 3533 | Creatine supplementation in the aging population: effects on skeletal muscle, bone and brain | Inappropriate study design |
| 3534 | A multidisciplinary group programme in rural settings for community-dwelling chronic stroke survivors and their carers: a pilot randomized controlled trial | Inappropriate study design |
| 3535 | Assessment of peak oxygen consumption, lactate and ventilatory thresholds and correlation with resting and exercise hemodynamic data in chronic congestive heart failure | Inappropriate population |
| 3536 | Prestroke physical activity could influence acute stroke severity (part of PAPSIGOT) | Inappropriate study design |
| 3537 | Peak Cardiorespiratory Responses of Patients with Subacute Stroke During Land and Aquatic Treadmill Exercise | Inappropriate study design |
| 3538 | Multidimensional evaluation of changes in limb function following botulinum toxin injection in persons with stroke | Inappropriate population |
| 3539 | Self-reported sleep disturbance and incidence of dementia in ageing men | Inappropriate study design |
| 3540 | Reductions in resting blood pressure after 4 weeks of isometric exercise training | Inappropriate population |
| 3541 | The effect of tyramine infusion and exercise on blood flow, coagulation and clot microstructure in healthy individuals | Inappropriate population |
| 3542 | Hemodynamic and metabolic basis of impaired exercise tolerance in patients with severe left ventricular dysfunction | Inappropriate population |
| 3543 | Method for Muscle Tone Monitoring During Robot-Assisted Therapy of Hand Function: A Proof of Concept | Inappropriate study design |
| 3544 | Hydralazine therapy in severe chronic heart failure: inability of radionuclide left ventricular ejection fraction measurement to predict the hemodynamic response | Inappropriate study design |
| 3545 | Exercise-related cognitive effects on sensory-motor control in athletes and drummers compared to non-athletes and other musicians | Inappropriate population |
| 3546 | Multi-organ damage in exertional heat stroke | Inappropriate study design |
| 3547 | Beta blockade in congestive heart failure: persistent adverse haemodynamic effects during chronic treatment with subsequent doses | Inappropriate study design |
| 3548 | Human cardiorespiratory responses to acute cold exposure | Inappropriate study design |
| 3549 | Haemodynamic dose-response effects of intravenous nisoldipine in coronary artery disease | Inappropriate population |
| 3550 | Dynamics of middle cerebral artery blood flow velocity during moderate-intensity exercise | Inappropriate study design |
| 3551 | Protocol for the "Chemobrain in Motion - study" (CIM - study): a randomized placebo-controlled trial of the impact of a high-intensity interval endurance training on cancer related cognitive impairments in women with breast cancer receiving first-line chemotherapy | Inappropriate outcome |
| 3552 | Association between recent sports activity, sports activity in young adulthood, and stroke | Inappropriate study design |
| 3553 | A model to estimate the potential for a physical activity-induced reduction in healthcare costs for the elderly, based on pedometer/accelerometer data from the Nakanojo Study | Inappropriate population |
| 3554 | Short bouts of mild-intensity physical exercise improve spatial learning and memory in aging rats: involvement of hippocampal plasticity via AKT, CREB and BDNF signaling | Inappropriate population |

| | | |
|---|---|---|
| 3555 | Improved exercise tolerance of the polycythemic lung patient following phlebotomy | Inappropriate study design |
| 3556 | The effects of exercise during pregnancy on the newborn's brain: study protocol for a randomized controlled trial | Inappropriate population |
| 3557 | Isometric exercise for the evaluation of vasodilatory therapy in severe congestive heart failure | Inappropriate population |
| 3558 | Confirmatory factor analysis of the depressive cognition scale | Inappropriate population |
| 3559 | Physical activity, sleep, and fatigue in community dwelling Stroke Survivors | Inappropriate study design |
| 3560 | Changes in physical activity and related functional and disability levels in the first six months after stroke: a longitudinal follow-up study | Inappropriate population |
| 3561 | Mechanisms responsible for enhanced stroke volume after exercise training in coronary heart disease | Inappropriate population |
| 3562 | Effects of acute exercise on hemorheological, endothelial, and platelet markers in patients with chronic heart failure in sinus rhythm | Inappropriate population |
| 3563 | The effect of an exercise-based balance intervention on physical and cognitive performance for older adults: a pilot study | Inappropriate study design |
| 3564 | Importance of considering ventricular function when prescribing exercise after acute myocardial infarction | Inappropriate study design |
| 3565 | Pre-Frail Multicomponent Training Intervention project for complexity of biological signals, functional capacity and cognition improvement in pre-frail older adults: A blinded randomized controlled study protocol | Inappropriate population |
| 3566 | Forearm vasoconstriction during dynamic leg exercise in patients with chronic heart failure | Inappropriate population |
| 3567 | Implementing frailty into clinical practice: a cautionary tale | Inappropriate population |
| 3568 | Role of walnuts in maintaining brain health with age | Inappropriate population |
| 3569 | Improvement of Upper Limb Motor Control and Function After Competitive and Noncompetitive Volleyball Exercises in Chronic Stroke Survivors: A Randomized Clinical Trial | Inappropriate study design |
| 3570 | Stroke volume decreases during mild dynamic and static exercise in supine humans | Inappropriate study design |
| 3571 | Beneficial effects of hydralazine on rest and exercise hemodynamics in patients with chronic severe aortic insufficiency | Inappropriate study design |
| 3572 | Association of Self-Leadership and Planning With Performing an Exercise in Patients With Colorectal Cancer: Cross-Sectional Study | Inappropriate study design |
| 3573 | Interactive effects of physical activity and APOE-ε4 on white matter tract diffusivity in healthy elders | Inappropriate study design |
| 3574 | Family-mediated exercise intervention (FAME): evaluation of a novel form of exercise delivery after stroke | Inappropriate population |
| 3575 | Scoping review report: obesity in older adults | Inappropriate study design |
| 3576 | Multicomponent Exercise Improves Physical Functioning but Not Cognition and Hemodynamic Parameters in Elderly Osteoarthritis Patients Regardless of Hypertension | Inappropriate population |
| 3577 | Measurement of stroke volume with impedance cardiography | Inappropriate study design |
| 3578 | A cholinergic contribution to the circulatory responses evoked at the onset of handgrip exercise in humans | Inappropriate study design |
| 3579 | Relationships between heart rate, exercise tolerance and cardiac output in atrial fibrillation: the effects of treatment with digoxin, verapamil and diltiazem | Inappropriate intervention |
| 3580 | Difficulty with learning of exercise instructions associated with 'working memory' dysfunction and frontal glucose hypometabolism in a patient with very mild subcortical vascular dementia with knee osteoarthritis | Inappropriate study design |
| 3581 | Multidisciplinary treatment of chronic pulmonary insufficiency. 3. The effect of physical training on cardiopulmonary performance in patients with chronic obstructive pulmonary disease | Inappropriate population |
| 3582 | Aerobic fitness does not contribute to prediction of orthostatic intolerance | Inappropriate population |
| 3583 | Exercise-training-induced changes in metabolic capacity with age: the role of central cardiovascular plasticity | Inappropriate population |
| 3584 | Designing clinical trials for assessing the effects of cognitive training and physical activity interventions on cognitive outcomes: the Seniors Health and Activity Research Program Pilot (SHARP-P) study, a randomized controlled trial | Inappropriate population |
| 3585 | Chronic exercise and left ventricular structure and function in healthy human subjects | Inappropriate study design |
| 3586 | Left ventricular function in endurance runners during exercise | Inappropriate study design |

| | | |
|---|---|---|
| 3587 | Yoga and exercise for symptoms of depression and anxiety in people with poststroke disability: a randomized, controlled pilot trial | Inappropriate population |
| 3588 | Resistance training with creatine monohydrate improves upper-body strength in patients with Parkinson disease: a randomized trial | Inappropriate study design |
| 3589 | Trimazosin in chronic congestive heart failure: improved left ventricular function at rest and during exercise | Inappropriate study design |
| 3590 | Exercise leads to faster postural reflexes, improved balance and mobility, and fewer falls in older persons with chronic stroke | Inappropriate study design |
| 3591 | Exercise frequency is related to psychopathology but not neurocognitive function | Inappropriate study design |
| 3592 | Illness cognition as a predictor of exercise habits and participation in cardiac prevention and rehabilitation programs after acute coronary syndrome | Inappropriate study design |
| 3593 | Frailty in childhood cancer survivors | Inappropriate study design |
| 3594 | Determinants of walking function after stroke: differences by deficit severity | Inappropriate study design |
| 3595 | Objectively measured physical activity is related to cognitive function in older adults | Inappropriate population |
| 3596 | Cardiac output but not stroke volume is similar in a Wingate and VO2peak test in young men | Inappropriate study design |
| 3597 | Effects of a new second generation calcium channel blocker, nilvadipine (FR34235), on exercise-induced hemodynamic changes in stable angina pectoris | Inappropriate population |
| 3598 | Chronic training with static and dynamic exercise: cardiovascular adaptation, and response to exercise | Inappropriate population |
| 3599 | Muscle torque preservation and physical activity in individuals with stroke | Inappropriate study design |
| 3600 | Impaired central hemodynamics in chronic obstructive pulmonary disease during submaximal exercise | Inappropriate study design |
| 3601 | A non-invasively determined surrogate of cardiac power ('circulatory power') at peak exercise is a powerful prognostic factor in chronic heart failure | Inappropriate study design |
| 3602 | Use of Accelerometers to Examine Sedentary Time on an Acute Stroke Unit | Inappropriate population |
| 3603 | Persistent Impairment in Cardiopulmonary Fitness after Breast Cancer Chemotherapy | Inappropriate population |
| 3604 | Facilitation of corticospinal excitability by virtual reality exercise following anodal transcranial direct current stimulation in healthy volunteers and subacute stroke subjects | Inappropriate study design |
| 3605 | NOS3 gene polymorphisms and exercise hemodynamics in postmenopausal women | Inappropriate study design |
| 3606 | Patterns of exercise response in patients with severe left ventricular dysfunction: radionuclide ejection fraction and hemodynamic cardiac performance evaluations | Inappropriate study design |
| 3607 | Fatigue after stroke: relationship to mobility, fitness, ambulatory activity, social support, and falls efficacy | Inappropriate study design |
| 3608 | Feasibility, reproducibility and validity of the 10 meter Shuttle Test in mild to moderately impaired people with stroke | Inappropriate study design |
| 3609 | Nifedipine in congestive heart failure: effects on resting and exercise hemodynamics and regional blood flow | Inappropriate population |
| 3610 | Effectiveness of coordination exercise in improving cognitive function in older adults: a prospective study | Inappropriate study design |
| 3611 | What do acute stroke physiotherapists do to treat postural control and mobility? An exploration of the content of therapy in the UK | Inappropriate study design |
| 3612 | Hemodynamic effects of alcohol at rest and during upright exercise in coronary artery disease | Inappropriate study design |
| 3613 | Effect of pulmonary endarterectomy for chronic thromboembolic pulmonary hypertension on stroke volume response to exercise | Inappropriate study design |
| 3614 | Syncope during exercise, documented with continuous blood pressure monitoring during ergometer testing | Inappropriate study design |
| 3615 | Endurance and neuromuscular changes in world-class level kayakers during a periodized training cycle | Inappropriate population |
| 3616 | Mechanical efficiency and propulsion technique after 7 weeks of low-intensity wheelchair training | Inappropriate population |
| 3617 | Abnormal systolic blood pressure response during exercise recovery in patients with angina pectoris | Inappropriate study design |
| 3618 | Exercise training in recently hospitalized heart failure patients enrolled in a disease management programme: design of the EJECTION-HF randomized controlled trial | Inappropriate study design |
| 3619 | Phase I dynamics of cardiac output, systemic O2 delivery, and lung O2 uptake at exercise onset in men in acute normobaric hypoxia | Inappropriate study design |

| | | |
|---|---|---|
| 3620 | Swim specialty affects energy cost and motor organization | Inappropriate study design |
| 3621 | Acute haemodynamic effects of perindoprilat in essential hypertension at rest and during exercise | Inappropriate population |
| 3622 | Variation in renal responses to exercise in the heat with progressive acclimatisation | Inappropriate study design |
| 3623 | Chronic use of benzodiazepines and latent cognitive decline in the elderly: results from the Three-city study | Inappropriate study design |
| 3624 | The effect of gender on aerobic power and exercise hemodynamics in hypertensive adults | Inappropriate study design |
| 3625 | Effects of long-term therapy with oral piroximone on resting hemodynamics, peak aerobic capacity, and the anaerobic threshold in patients with heart failure | Inappropriate study design |
| 3626 | Symptoms, haemodynamics, and exercise capacity during long term treatment of chronic heart failure. Experience with pirbuterol | Inappropriate study design |
| 3627 | Quantitative radionuclide angiography in assessment of hemodynamic changes during upright exercise: observations in normal subjects, patient with coronary artery disease and patients with aortic regurgitation | Inappropriate study design |
| 3628 | Anti-ischaemic and haemodynamic effects of elgodipine, a potent new generation calcium antagonist, in chronic stable angina | Inappropriate study design |
| 3629 | Dynamics of cardiac, respiratory, and metabolic function in men in response to step work load | Inappropriate study design |
| 3630 | Influence of fitness and gender on blood pressure responses during active or passive stress | Inappropriate study design |
| 3631 | Dysfunctions associated with dementia and their treatment | Inappropriate study design |
| 3632 | Stroke prevention and management in older adults | Inappropriate intervention |
| 3633 | A comparison of functional and impairment-based robotic training in severe to moderate chronic stroke: a pilot study | Inappropriate study design |
| 3634 | Incremental prognostic value of exercise hemodynamic variables in chronic congestive heart failure secondary to coronary artery disease or to dilated cardiomyopathy | Inappropriate study design |
| 3635 | High-intensity endurance training in 20- to 30- and 60- to 70-yr-old healthy men | Inappropriate study design |
| 3636 | Differential contribution of central command to the cardiovascular responses during static exercise of ankle dorsal and plantar flexion in humans | Inappropriate study design |
| 3637 | Patient Engagement Is Related to Impairment Reduction During Digital Game-Based Therapy in Stroke | Inappropriate study design |
| 3638 | Blood pressure and muscle sympathetic nerve activity during cold pressor stress: fitness and gender | Inappropriate population |
| 3639 | Hemodynamic effects of nifedipine in primary pulmonary hypertension | Inappropriate study design |
| 3640 | Exercise training effects on elderly and middle-age patients with chronic heart failure after acute decompensation: A randomized, controlled trial | Inappropriate study design |
| 3641 | Fatal rhabdomyolysis during mountaineering | Inappropriate population |
| 3642 | The Frequency and Health Benefits of Physical Activity for Older Adults | Inappropriate study design |
| 3643 | Diastolic properties of the normal left ventricle during supine exercise | Inappropriate population |
| 3644 | A High-Intensity Multicomponent Agility Intervention Improves Parkinson Patients' Clinical and Motor Symptoms | Inappropriate study design |
| 3645 | Hemodynamic changes and prognosis in patients with hypertrophic cardiomyopathy and abnormal blood pressure responses during exercise | Inappropriate study design |
| 3646 | Acute effects of exergames on cognitive function of institutionalized older persons: a single-blinded, randomized and controlled pilot study | Inappropriate study design |
| 3647 | Cardiac response to exercise in young, normal weight and overweight men and women | Inappropriate outcome |
| 3648 | Influence of dynamic training on hemodynamic, neurohormonal responses to static exercise and on inflammatory markers in patients after coronary artery bypass grafting | Inappropriate study design |
| 3649 | Effect of physical exhaustion on cognitive functioning | Inappropriate study design |
| 3650 | Metaboreflex activity in multiple sclerosis patients | Inappropriate study design |
| 3651 | Effects of an Exercise Protocol for Improving Handgrip Strength and Walking Speed on Cognitive Function in Patients with Chronic Stroke | Inappropriate population |
| 3652 | Strategies to improve recruitment and retention of older stroke survivors to a randomized clinical exercise trial | Inappropriate study design |
| 3653 | Diagnostic Performance of Pulmonary Capacitance at Rest and During Exercise in Idiopathic Pulmonary Arterial Hypertension | Inappropriate population |

| | | |
|---|---|---|
| 3654 | Potential mediators of ethnic differences in physical activity in older Mexican Americans and European Americans: results from the San Antonio Longitudinal Study of Aging | Inappropriate population |
| 3655 | Insulin resistance syndrome predicts the risk of coronary heart disease and stroke in healthy middle-aged men: the 22-year follow-up results of the Helsinki Policemen Study | Inappropriate population |
| 3656 | Ask the doctor. I am a 70-year-old man with high blood pressure that I control with medication, diet, and exercise. My mother, aunt, and maternal grandfather all died in their 50s after a single stroke. Is there an inherited tendency to hemorrhagic stroke? Are there any precautions I can take? | Inappropriate study design |
| 3657 | Parastomal hernia and physical activity. Are patients getting the right advice? | Inappropriate outcome |
| 3658 | Reducing the impact of physical inactivity: evidence to support the case for targeting people with chronic mental and physical conditions | Inappropriate study design |
| 3659 | Late cardiovascular drift observable during ultraendurance exercise | Inappropriate population |
| 3660 | Life-long aerobic exercise preserved baseline cerebral blood flow but reduced vascular reactivity to CO2 | Inappropriate outcome |
| 3661 | Additive effects of non-invasive ventilation to hyperoxia on cerebral oxygenation in COPD patients with exercise-related O2 desaturation | Inappropriate study design |
| 3662 | Hemodynamic response to graded exercise after chronic beta-adrenergic blockade | Inappropriate outcome |
| 3663 | Functional mitral regurgitation at rest determines the acute hemodynamic response to cardiac resynchronization therapy during exercise: an acute exercise echocardiographic study | Inappropriate population |
| 3664 | Endurance exercise training augments diastolic filling at rest and during exercise in healthy young and older men | Inappropriate study design |
| 3665 | Effectiveness of exercise programs to reduce falls in older people with dementia living in the community: a systematic review and meta-analysis | Inappropriate population |
| 3666 | Behavioural Risk Factors in Mid-Life Associated with Successful Ageing, Disability, Dementia and Frailty in Later Life: A Rapid Systematic Review | Inappropriate study design |
| 3667 | Left ventricular function during exercise in athletes and in sedentary men | Inappropriate population |
| 3668 | Influence of endurance and resistance exercise order on the postexercise hemodynamic responses in hypertensive women | Inappropriate population |
| 3669 | Influence of short-term oral hydralazine therapy on exercise hemodynamics in patients with severe chronic heart failure | Inappropriate population |
| 3670 | Effect of reduced training volume on cardiac function, VO2 max, and running performance | Inappropriate study design |
| 3671 | Feasibility of gait training for acute stroke patients using FNS with implanted electrodes | Inappropriate study design |
| 3672 | Chronic neurological disorders and related comorbidities: Role of age-associated physiological changes | Inappropriate population |
| 3673 | Respiratory muscle training increases cycling endurance without affecting cardiovascular responses to exercise | Inappropriate population |
| 3674 | Pharmacotherapy guidelines for the aged by family doctors for the use of family doctors: Part D Basic conditions supporting drug treatment | Inappropriate population |
| 3675 | Non-linear relationships between central cardiovascular variables and VO2 during incremental cycling exercise in endurance-trained individuals | Inappropriate population |
| 3676 | A multicomponent exercise intervention to improve physical functioning, cognition and psychosocial well-being in elderly nursing home residents: a study protocol of a randomized controlled trial in the PROCARE (prevention and occupational health in long-term care) project | Inappropriate population |
| 3677 | Effects of high-intensity progressive resistance training and targeted multidisciplinary treatment of frailty on mortality and nursing home admissions after hip fracture: a randomized controlled trial | Inappropriate outcome |
| 3678 | Acute Mild Hypoxic Hypoxia Effects on Cognitive and Simulated Aircraft Pilot Performance | Inappropriate population |
| 3679 | Rowing increases stroke volume and cardiac output to a greater extent than cycling | Inappropriate population |
| 3680 | Prevention and treatment of the metabolic syndrome in the elderly | Inappropriate study design |
| 3681 | People with Dementia Can Learn Compensatory Movement Maneuvers for the Sit-to-Stand Task: A Randomized Controlled Trial | Inappropriate study design |
| 3682 | Hypotensive and regional haemodynamic effects of exercise, fasted and after food, in human sympathetic denervation | Inappropriate population |
| 3683 | Effect of digoxin and vasodilators on left ventricular function in aortic regurgitation | Inappropriate study design |

| | | |
|---|---|---|
| 3684 | Chronic haemodynamic effects of carvedilol in essential hypertension at rest and during exercise | Inappropriate study design |
| 3685 | Movement behavior remains stable in stroke survivors within the first two months after returning home | Inappropriate study design |
| 3686 | Cognitive decline in patients with long-term domiciliary oxygen therapy | Inappropriate study design |
| 3687 | Self-rated physical fitness and estimated maximal oxygen uptake in relation to all-cause and cause-specific mortality | Inappropriate population |
| 3688 | Physical activity and risk of Alzheimer disease: A 2-sample mendelian randomization study | Inappropriate population |
| 3689 | Energy Expenditure and Cost During Walking After Stroke: A Systematic Review | Inappropriate study design |
| 3690 | Histamine H2 receptor blockade augments blood pressure responses to acute submaximal exercise in males | Inappropriate population |
| 3691 | Improved exercise tolerance and cardiac function in severe chronic heart failure patients undergoing a supervised exercise program | Inappropriate study design |
| 3692 | Afterload reduction with vasodilators and diuretics decreases mitral regurgitation during upright exercise in advanced heart failure | Inappropriate study design |
| 3693 | Effects of a multidisciplinary supervised exercise program on motor performance and quality of life in community-dwelling chronic stroke survivors in Korean | Inappropriate study design |
| 3694 | Lokomat: a therapeutic chance for patients with chronic hemiplegia | Inappropriate study design |
| 3695 | Physical activity and risk of myocardial infarction, cerebral stroke and death: a longitudinal study in Eastern Finland | Inappropriate population |
| 3696 | Effect of exercise on indices of valvular aortic stenosis | Inappropriate study design |
| 3697 | Heart Rate Reserve in Fontan Patients: Chronotropic Incompetence or Hemodynamic Limitation? | Inappropriate study design |
| 3698 | Randomized controlled trial of a coordinated care intervention to improve risk factor control after stroke or transient ischemic attack in the safety net: Secondary stroke prevention by Uniting Community and Chronic care model teams Early to End Disparities (SUCCEED) | Inappropriate study design |
| 3699 | A comparison of dance interventions in people with Parkinson disease and older adults | Inappropriate study design |
| 3700 | Efficacy of Trunk Regimes on Balance, Mobility, Physical Function, and Community Reintegration in Chronic Stroke: A Parallel-Group Randomized Trial | Inappropriate population |
| 3701 | Favorable effects of hydralazine on the hemodynamic response to isometric exercise in chronic severe aortic regurgitation | Inappropriate study design |
| 3702 | Haemodynamics in hypertensive patients before and during guanfacine treatment | Inappropriate outcome |
| 3703 | Positive effects of resistance training in frail elderly patients with dementia after long-term physical restraint | Inappropriate study design |
| 3704 | Compromised bone strength index in the hemiparetic distal tibia epiphysis among chronic stroke patients: the association with cardiovascular function, muscle atrophy, mobility, and spasticity | Inappropriate population |
| 3705 | Community-based exercise for upper limb paresis: a controlled trial with telerehabilitation | Inappropriate study design |
| 3706 | Perspectives on the prospective development of stroke-specific lower extremity wearable monitoring technology: a qualitative focus group study with physical therapists and individuals with stroke | Inappropriate study design |
| 3707 | Single limb exercise induces femoral artery remodeling and improves blood flow in the hemiparetic leg poststroke | Inappropriate study design |
| 3708 | Supervised exercise to reduce agitation in severely cognitively impaired persons | Inappropriate population |
| 3709 | The importance of the muscle and ventilatory blood pumps during exercise in patients without a subpulmonary ventricle (Fontan operation) | Inappropriate population |
| 3710 | Relationship between cognitive function and 6-minute walking test in older outpatients with chronic heart failure | Inappropriate study design |
| 3711 | Illuminating the black box: investigating prefrontal cortical hemodynamics during exercise with near-infrared spectroscopy | Inappropriate study design |
| 3712 | Functional impairment associated with acute poststroke depression: the Stroke Data Bank Study | Inappropriate population |
| 3713 | Cardiorespiratory responses and myocardial function within incremental exercise in healthy unmedicated older vs. young men and women | Inappropriate study design |
| 3714 | Physical activity and 10-year mortality from cardiovascular diseases and all causes: The Zutphen Elderly Study | Inappropriate study design |
| 3715 | Haemodynamic effects of proportional assist ventilation during high-intensity exercise in patients with chronic obstructive pulmonary disease | Inappropriate population |

| | | |
|---|---|---|
| 3716 | Myocardial infarct size and cardiac performance at exercise soon after myocardial infarction | Inappropriate study design |
| 3717 | Prior TIA, lipid-lowering drug use, and physical activity decrease ischemic stroke severity | Inappropriate population |
| 3718 | Trial Implementation of a Telerehabilitation Exercise System in Residential Aged Care | Inappropriate population |
| 3719 | Hemodynamic responses to indoramin at rest and during exercise in congestive heart failure | Inappropriate population |
| 3720 | Progressive Resistance and Balance Training for Falls Prevention in Long-Term Residential Aged Care: A Cluster Randomized Trial of the Sunbeam Program | Inappropriate study design |
| 3721 | Pilot randomized controlled trial to assess a physical therapy program on upper extremity function to counteract inactivity in chronic stroke | Inappropriate study design |
| 3722 | Modified constraint-induced therapy in chronic stroke: results of a single-blinded randomized controlled trial | Inappropriate study design |
| 3723 | Physical activity-related beliefs and discrepancies between beliefs and physical activity behavior for various chronic diseases | Inappropriate study design |
| 3724 | Effect of long-term diltiazem treatment on central haemodynamics and exercise endurance in essential hypertension | Inappropriate population |
| 3725 | Postural specificity of cardiovascular adaptations to exercise training | Inappropriate population |
| 3726 | Assessment of Motivational Cognitions in Diabetes Self-Care: the Motivation Thought Frequency Scales for Glucose Testing, Physical Activity and Healthy Eating | Inappropriate population |
| 3727 | Measuring activity levels at an acute stroke ward: comparing observations to a device | Inappropriate study design |
| 3728 | Exercise haemodynamics and maximal exercise capacity during beta-adrenoceptor blockade in normotensive and hypertensive subjects | Inappropriate study design |
| 3729 | Influence of verapamil therapy on left ventricular performance at rest and during exercise in hypertrophic cardiomyopathy | Inappropriate population |
| 3730 | Cardio-protection afforded by β-blockade is maintained during resistance exercise | Inappropriate population |
| 3731 | Risk factors for probable REM sleep behavior disorder: A community-based study | Inappropriate study design |
| 3732 | Noninvasive estimation of pulmonary vascular resistance by stroke index measurement with an inert gas rebreathing technique | Inappropriate study design |
| 3733 | Noninvasive assessment of cardiac output from arterial pressure profiles during exercise | Inappropriate outcome |
| 3734 | Cardiopulmonary function after pulmonary lobectomy in patients with lung cancer | Inappropriate study design |
| 3735 | Effect of aquatic versus land motor dual task training on balance and gait of patients with chronic stroke: A randomized controlled trial | Inappropriate outcome |
| 3736 | Mapping Movement: Applying Motion Measurement Technologies to the Psychiatric Care of Older Adults | Inappropriate study design |
| 3737 | Determinants of fatigue after first-ever ischemic stroke during acute phase | Inappropriate study design |
| 3738 | Immediate effects of unaffected arm exercise in poststroke patients with spastic upper limb hemiparesis | Inappropriate study design |
| 3739 | Frailty Among Older Decompensated Heart Failure Patients: Prevalence, Association With Patient-Centered Outcomes, and Efficient Detection Methods | Inappropriate study design |
| 3740 | Multicomponent Exercise on Physical Function, Cognition and Hemodynamic Parameters of Community-Dwelling Older Adults: A Quasi-Experimental Study | Inappropriate study design |
| 3741 | Mitral regurgitation determined by radionuclide cardiography: dependence on posture and exercise | Inappropriate study design |
| 3742 | The haemodynamic importance of atrioventricular synchrony and rate increase at rest and during exercise | Inappropriate population |
| 3743 | Cardiorespiratory response to walking in trained and sedentary pregnant women | Inappropriate outcome |
| 3744 | Lung volumes in swimmers performing different styles of swimming | Inappropriate population |
| 3745 | Hemodynamic and cardiorespiratory responses to various arm cycling regimens in men with spinal cord injury | Inappropriate population |
| 3746 | High intensity interval training protects the heart during increased metabolic demand in patients with type 2 diabetes: a randomised controlled trial | Inappropriate study design |
| 3747 | Prior transient ischemic attack is independently associated with lesser in-hospital case fatality in acute stroke | Inappropriate study design |
| 3748 | Limitations to systemic and locomotor limb muscle oxygen delivery and uptake during maximal exercise in humans | Inappropriate population |

| | | |
|---|---|---|
| 3749 | Effect of a multimodal exercise program on sleep disturbances and instrumental activities of daily living performance on Parkinson's and Alzheimer's disease patients | Inappropriate study design |
| 3750 | Promoting physical activity in hospitalized patients with mild ischemic stroke: a pilot study | Inappropriate population |
| 3751 | Evaluation of spastic muscle in stroke survivors using magnetic resonance imaging and resistance to passive motion | Inappropriate population |
| 3752 | Effects of a meal on hemodynamic function at rest and during exercise in patients with hypertrophic cardiomyopathy | Inappropriate study design |
| 3753 | Comparison of epidemiology, emergency care, and outcomes of acute ischemic stroke between young adults and elderly in Korean population: a multicenter observational study | Inappropriate study design |
| 3754 | Recurrence risk after noncardioembolic mild ischemic stroke in a Japanese population | Inappropriate study design |
| 3755 | Influence of combined physical and cognitive training on cognition: a systematic review | Inappropriate study design |
| 3756 | Effects of acute cardioselective and non-selective beta-adrenergic blockade on left-ventricular volumes and vascular resistance at rest and during exercise | Inappropriate intervention |
| 3757 | Circulatory regulation during exercise in different ambient temperatures | Inappropriate study design |
| 3758 | Development of a patient reported outcome measures for measuring the impact of visual impairment following stroke | Inappropriate study design |
| 3759 | Not all stroke units are the same: a comparison of physical activity patterns in Melbourne, Australia, and Trondheim, Norway | Inappropriate population |
| 3760 | Mechanisms for an abnormal radionuclide left ventricular ejection fraction response to exercise in patients with chronic, severe aortic regurgitation | Inappropriate study design |
| 3761 | Physical Activity Intensity and Cardiovascular Disease Prevention-From the Seguimiento Universidad de Navarra Study | Inappropriate population |
| 3762 | Interventions incorporating physical and cognitive elements to reduce falls risk in cognitively impaired older adults: a systematic review | Inappropriate population |
| 3763 | Effect of pedaling exercise on the hemiplegic lower limb | Inappropriate population |
| 3764 | Community-based Yang-style Tai Chi is safe and feasible in chronic stroke: a pilot study | Inappropriate study design |
| 3765 | Flow dependence of measures of aortic stenosis severity during exercise | Inappropriate study design |
| 3766 | Exercise tolerance and cardiorespiratory response to exercise after the Fontan operation for tricuspid atresia or functional single ventricle | Inappropriate study design |
| 3767 | Proficiency of balance in children and youth who have had acute lymphoblastic leukemia | Inappropriate population |
| 3768 | Individualized treadmill and strength training for chronic stroke rehabilitation: effects of imbalance | Inappropriate outcome |
| 3769 | Hemodynamic adjustments in hypertensive patients developing circulatory congestion during beta-adrenergic blockade | Inappropriate population |
| 3770 | Moderate-to-vigorous physical activity and the risk of stroke recurrence in patients with a history of minor ischemic stroke in Japan: a retrospective analysis | Inappropriate outcome |
| 3771 | Secondary prevention of new vascular events with lifestyle intervention in patients with noncardioembolic mild ischemic stroke: a single-center randomized controlled trial | Inappropriate intervention |
| 3772 | Accelerometer monitoring of home- and community-based ambulatory activity after stroke | Inappropriate study design |
| 3773 | Associations between vigorous physical activity and chronic diseases in older adults: a study in 13 European countries | Inappropriate population |
| 3774 | Improving executive function deficits by playing interactive video-games: secondary analysis of a randomized controlled trial for individuals with chronic stroke | Inappropriate study design |
| 3775 | Haemodynamic effects of short-term treatment with bopindolol in essential hypertension | Inappropriate study design |
| 3776 | Inter-individual variability in response to exercise intervention or usual care in hospitalized older adults | Inappropriate population |
| 3777 | Effect of isometric exercise on cardiac performance and mitral regurgitation in patients with severe congestive heart failure | Inappropriate study design |
| 3778 | Early hemodynamic adaptations to physical training in patients with healed myocardial infarction | Inappropriate population |
| 3779 | Geography, Race/Ethnicity, and Physical Activity Among Men in the United States | Inappropriate study design |
| 3780 | Physical Activity and Incident Cardiovascular Disease in Women: Is the Relation Modified by Level of Global Cardiovascular Risk? | Inappropriate outcome |

| | | |
|---|---|---|
| 3781 | Autonomic cardiovascular control changes in recent heart transplant recipients lead to physiological limitations in response to orthostatic challenge and isometric exercise | Inappropriate study design |
| 3782 | Beneficial Physiological Effects With Blackcurrant Intake in Endurance Athletes | Inappropriate study design |
| 3783 | Physical activity and cardiovascular disease: evidence for a dose response | Inappropriate population |
| 3784 | Keep moving without hurting: The interaction between physical activity and pain in determining cognitive function at the population level | Inappropriate study design |
| 3785 | Central and peripheral hemodynamics in exercising humans: leg vs arm exercise | Inappropriate population |
| 3786 | Response to changing plasma concentrations of isosorbide-bound NO2 during acute and sustained treatment with isosorbide dinitrate in patients with coronary artery disease | Inappropriate population |
| 3787 | Haemodynamics in juvenile hypertension at rest and during exercise | Inappropriate intervention |
| 3788 | Hemodynamic effects of felodipine in congestive heart failure | Inappropriate study design |
| 3789 | Daily physical activity and its contribution to the health-related quality of life of ambulatory individuals with chronic stroke | Inappropriate study design |
| 3790 | Hemodynamic determinants of exercise-induced abnormal blood pressure response in hypertrophic cardiomyopathy | Inappropriate population |
| 3791 | Low Stroke Volume during Exercise with Hot Skin Is Due to Elevated Heart Rate | Inappropriate study design |
| 3792 | Influence of training on biomechanics of wheelchair propulsion | Inappropriate study design |
| 3793 | Practical tips for prescribing exercise for fall prevention | Inappropriate study design |
| 3794 | Interval and strength training in CAD patients | Inappropriate study design |
| 3795 | Water-Based Exercise on Functioning and Quality of Life in Poststroke Persons: A Systematic Review and Meta-Analysis | Inappropriate population |
| 3796 | Systolic function during exercise in patients with coronary artery disease | Inappropriate population |
| 3797 | Are heat stroke and physical exhaustion underestimated causes of acute hepatic failure? | Inappropriate outcome |
| 3798 | Sex-related differences in the normal cardiac response to upright exercise | Inappropriate study design |
| 3799 | Dietary and Serum Omega-6/Omega-3 Fatty Acids Are Associated with Physical and Metabolic Function in Stroke Survivors | Inappropriate population |
| 3800 | Intrapulmonary arteriovenous anastomoses in humans with chronic obstructive pulmonary disease: implications for cryptogenic stroke? | Inappropriate study design |
| 3801 | Static exercise with congestive heart failure and the response to vasodilating drugs | Inappropriate study design |
| 3802 | Haemodynamic effects of isosorbide dinitrate in patients with congestive cardiac failure at rest and during submaximal supine exercise | Inappropriate population |
| 3803 | The Manitoba Personalized Lifestyle Research (TMPLR) study protocol: a multicentre bidirectional observational cohort study with administrative health record linkage investigating the interactions between lifestyle and health in Manitoba, Canada | Inappropriate population |
| 3804 | The effects of multi-stage exercise with and without concurrent cognitive performance on cardiorespiratory and cerebral haemodynamic responses | Inappropriate study design |
| 3805 | Physical activity, adiponectin, and cardiovascular structure and function | Inappropriate study design |
| 3806 | Hemodynamic and radionuclide effects of acute captopril therapy for heart failure: changes in left and right ventricular volumes and function at rest and during exercise | Inappropriate study design |
| 3807 | Health promotion in older adults: the role of lifestyle in the metabolic syndrome | Inappropriate study design |
| 3808 | Evidence for improved cardiac performance after beta-blockade in patients with coronary artery disease | Inappropriate population |
| 3809 | Efficacy of a Chronic Care-Based Intervention on Secondary Stroke Prevention Among Vulnerable Stroke Survivors: A Randomized Controlled Trial | Inappropriate study design |
| 3810 | A comparison of electromyography and stroke kinematics during ergometer and on-water rowing | Inappropriate study design |
| 3811 | Effect of weightlifting upon left ventricular function and markers of cardiomyocyte damage | Inappropriate outcome |
| 3812 | Augmentation index immediately after maximal exercise in patients with type 2 diabetes mellitus | Inappropriate population |
| 3813 | The Physical Activity and Disability Survey -- Revised (PADS-R): an evaluation of a measure of physical activity in people with chronic neurological conditions | Inappropriate population |
| 3814 | Detection and Quantitation of Circulating Human Irisin by Tandem Mass Spectrometry | Inappropriate study design |

| | | |
|---|---|---|
| 3815 | Cardiorespiratory response to absolute and relative work intensity in untrained men | Inappropriate study design |
| 3816 | Effects of different heel-raise-lower exercise interventions on the strength of plantarflexion, balance, and gait parameters in stroke survivors | Inappropriate study design |
| 3817 | Exercise pathophysiology and sildenafil effects in chronic thromboembolic pulmonary hypertension | Inappropriate study design |
| 3818 | Left ventricular performance in alcoholic patients without chronic liver disease | Inappropriate study design |
| 3819 | Contributions of Stepping Intensity and Variability to Mobility in Individuals Poststroke | Inappropriate study design |
| 3820 | Effects of nitroglycerin on isometric exercise | Inappropriate population |
| 3821 | Twenty-year follow-up of hemodynamics in essential hypertension during rest and exercise | Inappropriate study design |
| 3822 | Effect of a Combined Tai Chi, Resistance Training and Dietary Intervention on Cognitive Function in Obese Older Women | Inappropriate population |
| 3823 | A cross-sectional examination of age and physical activity on performance and event-related brain potentials in a task switching paradigm | Inappropriate intervention |
| 3824 | Reversing the Cardiac Effects of Sedentary Aging in Middle Age-A Randomized Controlled Trial: Implications For Heart Failure Prevention | Inappropriate population |
| 3825 | Haemodynamic and metabolic responses to prolonged exercise after chronic beta 1-adrenoceptor blockade in hypertensive man | Inappropriate population |
| 3826 | Hypoxia attenuates cardiopulmonary reflex control of sympathetic nerve activity during mild dynamic leg exercise | Inappropriate study design |
| 3827 | Haemodynamic effects of diltiazem at rest and during exercise in patients with previous myocardial infarction | Inappropriate study design |
| 3828 | The correlation of cardiac power output to exercise capacity in chronic heart failure | Inappropriate study design |
| 3829 | Improved lower extremity pedaling mechanics in individuals with stroke under maximal workloads | Inappropriate study design |
| 3830 | Stress resilience in male adolescents and subsequent stroke risk: cohort study | Inappropriate population |
| 3831 | Symptom-limited exercise testing, ST depressions and long-term coronary heart disease mortality in apparently healthy middle-aged men | Inappropriate population |
| 3832 | Continuous Cardiac Autonomic and Hemodynamic Responses to Isometric Exercise | Inappropriate population |
| 3833 | Exergames-based intervention for assisted living residents: a pilot study | Inappropriate population |
| 3834 | Where did the clot go? An unusual complication of mechanical thrombectomy caused by malignancy related subclavian steal phenomenon in a patient with acute basilar artery occlusion | Inappropriate population |
| 3835 | A hemodynamic comparison of young and older endurance athletes during exercise | Inappropriate study design |
| 3836 | Time spent lying, sitting, and upright during hospitalization after stroke: a prospective observation study | Inappropriate study design |
| 3837 | Pilot testing of the restorative care intervention for the cognitively impaired | Inappropriate study design |
| 3838 | Ascending aortic Doppler velocity and the prediction of exercise capacity in post-infarction left ventricular dysfunction | Inappropriate outcome |
| 3839 | Circulatory changes in acute glomerulonephritis at rest and during exercise | Inappropriate study design |
| 3840 | A comparison of cardiopulmonary adaptations to exercise in pregnancy at sea level and altitude | Inappropriate outcome |
| 3841 | Acute cocoa flavanol improves cerebral oxygenation without enhancing executive function at rest or after exercise | Inappropriate population |
| 3842 | Effect of exercise and of prolonged oral administration of propranolol on haemodynamic variables, plasma renin concentration, plasma aldosterone and c-AMP | Inappropriate population |
| 3843 | Right ventricular pressure-volume relations in chronic lung disease | Inappropriate study design |
| 3844 | Cardiovascular drift and Vo2max during cycling and walking in a temperate environment | Inappropriate study design |
| 3845 | Designing personal exercise monitoring employing multiple modes of delivery: implications from a qualitative study on heart rate monitoring | Inappropriate study design |
| 3846 | Sodium Nitrite Improves Exercise Hemodynamics and Ventricular Performance in Heart Failure With Preserved Ejection Fraction | Inappropriate population |
| 3847 | Exercise for reducing fear of falling in older people living in the community | Inappropriate population |
| 3848 | Development and preliminary evaluation of a novel low cost VR-based upper limb stroke rehabilitation platform using Wii technology | Inappropriate outcome |

| | | |
|---|---|---|
| 3849 | Focal Muscle Vibration and Progressive Modular Rebalancing with neurokinetic facilitations in post- stroke recovery of upper limb | Inappropriate population |
| 3850 | Impact of left ventricular diastolic function on exercise capacity in patients with chronic mitral regurgitation: an exercise echocardiography study | Inappropriate population |
| 3851 | Radionuclide absolute left ventricular volumes during upright exercise: validation in normal subjects by simultaneous hemodynamic measurements | Inappropriate study design |
| 3852 | Effects of low-intensity exercise conditioning on blood pressure, heart rate, and autonomic modulation of heart rate in men and women with hypertension | Inappropriate study design |
| 3853 | MELAS without ragged red fibers or lactic acidosis diagnosed by mitochondrial DNA testing | Inappropriate population |
| 3854 | Effects of passive leg movement on the oxygenation level of lower limb muscle in chronic stroke patients | Inappropriate study design |
| 3855 | Early Mobilization After Stroke Is Not Associated With Cognitive Outcome | Inappropriate study design |
| 3856 | The prevalence of co-morbidities and their impact on physical activity in people with inflammatory rheumatic diseases compared with the general population: results from the UK Biobank | Inappropriate population |
| 3857 | Haemodynamic contributions to post-exercise hypotension in young adults with hypertension and rapid resting heart rates | Inappropriate study design |
| 3858 | Physical activity, metabolic factors, and the incidence of coronary heart disease and type 2 diabetes | Inappropriate outcome |
| 3859 | Coarctation of the aorta. A long-term follow-up study after surgery | Inappropriate study design |
| 3860 | Inhaled Sodium Nitrite Improves Rest and Exercise Hemodynamics in Heart Failure With Preserved Ejection Fraction | Inappropriate population |
| 3861 | Hemodynamic effects of lidoflazine during graded levels of bicycle exercise in normal subjects | Inappropriate study design |
| 3862 | Effects of endoscopic transthoracic sympathicotomy on hemodynamic and neurohumoral responses to exercise in humans | Inappropriate study design |
| 3863 | Cardiovascular response to physical stress in offspring of hypertensive parents: Dutch Hypertension and Offspring Study | Inappropriate population |
| 3864 | Vascular elasticity and grip strength are associated with bone health of the hemiparetic radius in people with chronic stroke: implications for rehabilitation | Inappropriate study design |
| 3865 | Cardiovagal baroreflex and aortic hemodynamic responses to isometric exercise and post-exercise muscle ischemia in resistance trained men | Inappropriate study design |
| 3866 | Game analysis and clinical use of the Xbox-Kinect for stroke rehabilitation | Inappropriate study design |
| 3867 | Self-regulatory biofeedback training: an intervention to reduce school burnout and improve cardiac functioning in college students | Inappropriate study design |
| 3868 | Chronic Disease Diagnosis as a Teachable Moment for Health Behavior Changes Among Middle-Aged and Older Adults | Inappropriate population |
| 3869 | Central haemodynamics in essential hypertension at rest and during exercise: a 20-year follow-up study | Inappropriate population |
| 3870 | A pilot study of robotic-assisted exercise for hand weakness after stroke | Inappropriate study design |
| 3871 | Adjustments of cardiac function during prolonged exercise relative to lactate threshold | Inappropriate study design |
| 3872 | Exertional Heat Stroke and Susceptibility to Malignant Hyperthermia in an Athlete: Evidence for a Link? | Inappropriate outcome |
| 3873 | Comparative physiological responses of normotensive and essentially hypertensive men to exercise in the heat | Inappropriate intervention |
| 3874 | Ventriculo-arterial coupling detects occult RV dysfunction in chronic thromboembolic pulmonary vascular disease | Inappropriate study design |
| 3875 | Maximal aerobic power in essential hypertension | Inappropriate intervention |
| 3876 | Cardiovascular responses to head-out water immersion in Korean women breath-hold divers | Inappropriate intervention |
| 3877 | Secondary stroke prevention: challenges and solutions | Inappropriate intervention |
| 3878 | Submaximal exercise in persons with stroke: test-retest reliability and concurrent validity with maximal oxygen consumption | Inappropriate study design |
| 3879 | Acute heat stroke deaths | Inappropriate population |
| 3880 | Design and implementation of a training strategy in chronic stroke with an arm robotic exoskeleton | Inappropriate study design |
| 3881 | Dynamic and static exercise haemodynamics after acute myocardial infarction | Inappropriate population |
| 3882 | Carotid endarterectomy: improved clinical outcome and reduced length of hospitalization | Inappropriate population |
| 3883 | Exercise modality modulates body temperature regulation during exercise in uncompensable heat stress | Inappropriate outcome |

| | | |
|---|---|---|
| 3884 | A Tandem Cycling Program: Feasibility and Physical Performance Outcomes in People With Parkinson Disease | Inappropriate population |
| 3885 | Relations of pulmonary diffusing capacity to ventilation and haemodynamics in healthy subjects | Inappropriate population |
| 3886 | Maximum Walking Speed at Discharge Could Be a Prognostic Factor for Vascular Events in Patients With Mild Stroke: A Cohort Study | Inappropriate study design |
| 3887 | Analysis of the association between selected factors and outcomes of treadmill gait training with biofeedback in patients with chronic stroke | Inappropriate study design |
| 3888 | Glycemic status affects cardiopulmonary exercise response in athletes with type I diabetes | Inappropriate population |
| 3889 | The Volunteering-in-Place (VIP) Program: Providing meaningful volunteer activity to residents in assisted living with mild cognitive impairment | Inappropriate study design |
| 3890 | Physical activity as a mediator of the impact of chronic conditions on quality of life in older adults | Inappropriate population |
| 3891 | Formerly preeclamptic women with a subnormal plasma volume are unable to maintain a rise in stroke volume during moderate exercise | Inappropriate outcome |
| 3892 | Exercising in a hot environment with muscle damage: effects on acute kidney injury biomarkers and kidney function | Inappropriate study design |
| 3893 | Chronotropic Incompetence in Chronic Heart Failure | Inappropriate intervention |
| 3894 | Exercise capacity and stroke volume are preserved late after tetralogy repair, despite severe right ventricular dilatation | Inappropriate study design |
| 3895 | Physical activity is associated with incident disability in community-based older persons | Inappropriate study design |
| 3896 | Pulmonary vascular reserve and exercise capacity at sea level and at high altitude | Inappropriate population |
| 3897 | Determinants of functional capacity in chronic mitral regurgitation unassociated with coronary artery disease or left ventricular dysfunction | Inappropriate study design |
| 3898 | Intracyclic velocity variations and arm coordination during exhaustive exercise in front crawl stroke | Inappropriate outcome |
| 3899 | Expanded blood volumes contribute to the increased cardiovascular performance of endurance-trained older men | Inappropriate intervention |
| 3900 | Comparison of cognitive function between patients on chronic hemodialysis who carry out assisted physical activity and inactive ones | Inappropriate study design |
| 3901 | Effects of hydraulic circuit training following coronary artery bypass surgery | Inappropriate intervention |
| 3902 | Nitric oxide does not significantly contribute to changes in pulse pressure amplification during light aerobic exercise | Inappropriate population |
| 3903 | Task-related circuit training improves performance of locomotor tasks in chronic stroke: a randomized, controlled pilot trial | Inappropriate study design |
| 3904 | Robot-assisted exercise for hand weakness after stroke: a pilot study | Inappropriate outcome |
| 3905 | Association between skeletal muscle mass and cardiorespiratory fitness in community-dwelling elderly men | Inappropriate study design |
| 3906 | Perceptions and beliefs about the role of physical activity and nutrition on brain health in older adults | Inappropriate population |
| 3907 | Exercise capacity and systolic and diastolic ventricular function after recovery from acute dilated cardiomyopathy | Inappropriate population |
| 3908 | Cardiovascular response to exercise in pacemaker implanted patients with fixed heart rate | Inappropriate population |
| 3909 | Endurance exercise performance in Masters athletes: age-associated changes and underlying physiological mechanisms | Inappropriate study design |
| 3910 | Response of left ventricular diastolic filling to graded exercise relative to the lactate threshold | Inappropriate population |
| 3911 | Actigraphic analysis of the sleep-wake cycle and physical activity level in patients with stroke: implications for clinical practice | Inappropriate population |
| 3912 | Use of an elliptical machine for improving functional walking capacity in individuals with chronic stroke: a case series | Inappropriate study design |
| 3913 | Comparison of Training Monitoring and Prescription Methods in Sprint Kayaking | Inappropriate study design |
| 3914 | Cardiovascular reserve and risk profile of postmenopausal women after chemoendocrine therapy for hormone receptor--positive operable breast cancer | Inappropriate study design |
| 3915 | Long-term haemodynamic effects of amlodipine at rest and during exercise in essential hypertension | Inappropriate study design |
| 3916 | Effect of prostacyclin and prazosin in the treatment of congestive heart failure; with special reference to the sympathetic nervous system | Inappropriate population |
| 3917 | Left ventricular function after exercise training in young men | Inappropriate population |
| 3918 | Stress echocardiography in hyperthyroidism | Inappropriate population |

| | | |
|---|---|---|
| 3919 | Effects of autonomic blockade on cardiac function at rest and during upright exercise in humans | Inappropriate intervention |
| 3920 | Gait training with a robotic leg brace after stroke: a randomized controlled pilot study | Inappropriate study design |
| 3921 | Functional electrical stimulation assisted cycling of patients with subacute stroke: kinetic and kinematic analysis | Inappropriate study design |
| 3922 | Health behaviour changes after diagnosis of chronic illness among Canadians aged 50 or older | Inappropriate study design |
| 3923 | Multivariate logistic analysis of risk factors for stroke in Tilburg, The Netherlands | Inappropriate study design |
| 3924 | Epidemiological aspects of ageing | Inappropriate population |
| 3925 | Right ventricular performance and central circulatory hemodynamics during upright exercise in patients with chronic obstructive pulmonary disease | Inappropriate study design |
| 3926 | Effects of the calcium antagonist, isradipine, and nifedipine on resting and exercise haemodynamics and the neurohumoral system in patients with stable chronic angina | Inappropriate study design |
| 3927 | Effectiveness of psychological interventions in chronic stage of stroke: a systematic review | Inappropriate study design |
| 3928 | Cardiac output is a sensitive indicator of difference in exercise performance between single and dual sensor pacemakers | Inappropriate study design |
| 3929 | Acute care for exercise-induced hyperthermia to avoid adverse outcome from exertional heat stroke | Inappropriate study design |
| 3930 | Segmentation of Exercise Repetitions Enabling Real-Time Patient Analysis and Feedback Using a Single Exemplar | Inappropriate population |
| 3931 | Effects of acute intradialytic exercise on cardiovascular responses in hemodialysis patients | Inappropriate study design |
| 3932 | Cardiovascular responses to rowing | Inappropriate study design |
| 3933 | Effects of the self-contained breathing apparatus on left-ventricular function at rest and during graded exercise | Inappropriate study design |
| 3934 | Postexercise blood pressure reduction in elderly hypertensive patients | Inappropriate study design |
| 3935 | The cell cycle hypothesis of Alzheimer's disease: suggestions for drug development | Inappropriate outcome |
| 3936 | Mitral valve replacement for mitral regurgitation with and without preservation of chordae tendineae | Inappropriate population |
| 3937 | A video-game group intervention: Experiences and perceptions of adults with chronic stroke and their therapists: Intervention de groupe à l'aide de jeux vidéo : Expériences et perceptions d'adultes en phase chronique d'un accident vasculaire cérébral et de leurs ergothérapeutes | Inappropriate outcome |
| 3938 | Haemodynamic effect of metaboreflex activation in men after running above and below the velocity of the anaerobic threshold | Inappropriate population |
| 3939 | Influence of Previous-Year Physical Activity on the Cognition of COPD Older Patients During Exacerbation | Inappropriate population |
| 3940 | Cardiovascular responses to handgrip isometric exercise in patients following cardiac transplantation | Inappropriate population |
| 3941 | Predictive impact of daily physical activity on new vascular events in patients with mild ischemic stroke | Inappropriate study design |
| 3942 | Individualized home-based exercise programs for older people to reduce falls and improve physical performance: A systematic review and meta-analysis | Inappropriate population |
| 3943 | Integrative rehabilitation of residents chronic post-stroke in skilled nursing facilities: the design and evaluation of the BrightArm Duo | Inappropriate population |
| 3944 | Awareness of chronic disease related health benefits of physical activity among residents of a rural South Indian region: a cross-sectional study | Inappropriate study design |
| 3945 | Differential cardiovascular effects of propranolol, atenolol, and pindolol measured by impedance cardiography | Inappropriate study design |
| 3946 | Effects of daily mild supine exercise on physical performance after 20 days bed rest in young persons | Inappropriate study design |
| 3947 | Interactive effects of physical activity and APOE-ε4 on BOLD semantic memory activation in healthy elders | Inappropriate population |
| 3948 | Increased proximal arterial stiffness and cardiac response with moderate exercise in patients with heart failure and preserved ejection fraction | Inappropriate study design |
| 3949 | Are changes in upper extremity use during sub-acute rehabilitation after stroke associated with physical, cognitive, and social activities? An observational cohort pilot study | Inappropriate study design |
| 3950 | Effects of a Self-Exercise Program on Activities of Daily Living in Patients After Acute Stroke: A Propensity Score Analysis Based on the Japan Association of Rehabilitation Database | Inappropriate study design |

| | | |
|---|---|---|
| 3951 | Updating the Evidence for Physical Activity: Summative Reviews of the Epidemiological Evidence, Prevalence, and Interventions to Promote "Active Aging" | Inappropriate population |
| 3952 | Predictors of physical activity in older adults early in an emergency hospital admission: a prospective cohort study | Inappropriate population |
| 3953 | Attenuated responses of Doppler-derived hemodynamic parameters during supine bicycle exercise in heart transplant recipients | Inappropriate outcome |
| 3954 | Which dietary and lifestyle behaviours may be important in the aetiology (and prevention) of stroke? | Inappropriate intervention |
| 3955 | Ventilatory and circulatory responses at the onset of exercise in man following heart or heart-lung transplantation | Inappropriate study design |
| 3956 | Pulmonary hemodynamics during a strenuous intermittent exercise in healthy subjects | Inappropriate study design |
| 3957 | The cardiac autonomic nervous system response to different daily demands among patients at the sub-acute phase post ischemic stroke and healthy controls | Inappropriate outcome |
| 3958 | Left ventricular response to dynamic and static exercise: evaluation by radiocardiography in healthy men | Inappropriate study design |
| 3959 | Protocol for the Locomotor Experience Applied Post-stroke (LEAPS) trial: a randomized controlled trial | Inappropriate population |
| 3960 | Relationship Between Nutritional Factors and Hip Bone Density in Individuals with Chronic Stroke | Inappropriate population |
| 3961 | Physical activity participation and the risk of chronic diseases among South Asian adults: protocol for a systematic review and meta-analysis | Inappropriate study design |
| 3962 | Lifestyles of older adults: can we influence cardiovascular risk in older adults? | Inappropriate study design |
| 3963 | Exercise-induced rhabdomyolysis | Inappropriate study design |
| 3964 | Weight-shift training improves trunk control, proprioception, and balance in patients with chronic hemiparetic stroke | Inappropriate study design |
| 3965 | Effect of exercise training on biologic vascular age in healthy seniors | Inappropriate study design |
| 3966 | The association between changes in lifestyle behaviors and the incidence of chronic kidney disease (CKD) in middle-aged and older men | Inappropriate study design |
| 3967 | A systematic review of the efficacy of self-management programs for increasing physical activity in community-dwelling adults with acquired brain injury (ABI) | Inappropriate population |
| 3968 | Hemodynamic effects of intravenous elgodipine in coronary artery disease during rest and exercise, and basic pharmacokinetic parameters | Inappropriate study design |
| 3969 | Quantitative analysis of skeletal muscle bioenergetics and proton efflux in migraine and cluster headache | Inappropriate population |
| 3970 | Multisystem Balance Training Reduces Injurious Fall Risk in Parkinson Disease: A Randomized Trial | Inappropriate study design |
| 3971 | Brain-derived neurotrophic factor, insulin like growth factor-1 and inflammatory cytokine responses to continuous and intermittent exercise in patients with type 1 diabetes | Inappropriate study design |
| 3972 | Effectiveness of a Stroke Risk Self-Management Intervention for Adults with Prehypertension | Inappropriate outcome |
| 3973 | Muscle metaboreflex-induced increases in stroke volume | Inappropriate population |
| 3974 | The effects of a long-term care walking program on balance, falls and well-being | Inappropriate population |
| 3975 | Limitations to VO2max in humans after blood retransfusion | Inappropriate population |
| 3976 | Relationship between objectively assessed physical activity and fatigue in patients with rheumatoid arthritis: inverse correlation of activity and fatigue | Inappropriate study design |
| 3977 | Hemodynamic and ventilatory responses during exercise in chronic thromboembolic disease | Inappropriate population |
| 3978 | Sildenafil and diastolic dysfunction after acute myocardial infarction in patients with preserved ejection fraction: the Sildenafil and Diastolic Dysfunction After Acute Myocardial Infarction (SIDAMI) trial | Inappropriate population |
| 3979 | Importance of heart rate modulation on the cardiac hemodynamics during postexercise recovery | Inappropriate population |
| 3980 | Falls prevention through physical and cognitive training (falls PACT) in older adults with mild cognitive impairment: a randomized controlled trial protocol | Inappropriate study design |
| 3981 | Acute plasma expansion: left ventricular hemodynamics and endocrine function during exercise | Inappropriate study design |
| 3982 | Tele-Supervised FES-Assisted Exercise for Hemiplegic Upper Limb | Inappropriate study design |
| 3983 | RV Myocardial Strain During Pre-Load Augmentation Is Associated With Exercise Capacity in Patients With Chronic HF | Inappropriate study design |

| | | |
|---|---|---|
| 3984 | Cardiovascular responses to isometric handgrip exercise: an invasive study in pregnancy-induced hypertension | Inappropriate outcome |
| 3985 | Intensity of leg and arm training after primary middle-cerebral-artery stroke: a randomised trial | Inappropriate study design |
| 3986 | Operation Everest II: preservation of cardiac function at extreme altitude | Inappropriate population |
| 3987 | Strength, power, and muscular endurance exercise and elite rowing ergometer performance | Inappropriate study design |
| 3988 | Paretic hand rehabilitation with constraint-induced movement therapy after stroke | Inappropriate study design |
| 3989 | Cardiac function after orthotopic heart transplantation: response to postural changes, exercise, and beta-adrenergic blockade | Inappropriate study design |
| 3990 | Two-Year Agility Maintenance Training Slows the Progression of Parkinsonian Symptoms | Inappropriate population |
| 3991 | Treatment of chronic lymphocytic leukemia in older adults | Inappropriate study design |
| 3992 | Effects of Gait Training With Body Weight Support on a Treadmill Versus Overground in Individuals With Stroke | Inappropriate study design |
| 3993 | Frailty Screening in the Community Using the FRAIL Scale | Inappropriate study design |
| 3994 | Haemodynamic and humoral responses to chronic ketanserin treatment in essential hypertension | Inappropriate population |
| 3995 | Stroke rehabilitation at home using virtual reality, haptics and telemedicine | Inappropriate population |
| 3996 | Acute haemodynamic effects of felodipine in congestive heart failure | Inappropriate study design |
| 3997 | Association Between Active Commuting and Incident Cardiovascular Diseases in Chinese: A Prospective Cohort Study | Inappropriate population |
| 3998 | The effect of an intensive exercise programme on leg function in chronic stroke patients: a pilot study with one-year follow-up | Inappropriate outcome |
| 3999 | Pulmonary vascular response to exercise in symptomatic heart failure with reduced ejection fraction and pulmonary hypertension | Inappropriate study design |
| 4000 | High Level Physical Activity and Prevalence of Cardiovascular Disease Using the Korea National Health and Nutrition Examination Survey Data, 2007-2013 | Inappropriate population |
| 4001 | Bone mineral density in acute stroke patients: low bone mineral density may predict first stroke in women | Inappropriate intervention |
| 4002 | The haemodynamic response to exercise in chronic obstructive pulmonary disease: assessment by impedance cardiography | Inappropriate population |
| 4003 | Clinical experience of rehabilitation therapists with chronic diseases: a quantitative approach | Inappropriate population |
| 4004 | Hemodynamic effects of atrial synchronization in acute and long-term ventricular pacing | Inappropriate study design |
| 4005 | Haemodynamic effects of intravenous diltiazem at rest and exercise in patients with coronary artery disease | Inappropriate population |
| 4006 | Abnormal blood-pressure response to exercise and oxygen consumption in patients with hypertrophic cardiomyopathy | Inappropriate study design |
| 4007 | Comparison of left ventricular function using isometric exercise Doppler echocardiography in competitive runners and weightlifters versus sedentary individuals | Inappropriate population |
| 4008 | Cardiovascular responses to acute exercise in patients with cerebrovascular accidents | Inappropriate population |
| 4009 | Integrated cardiovascular rehabilitation: how do noncardiac and polyvascular disease patients fare? | Inappropriate population |
| 4010 | Heart rate is lower during ergometer rowing than during treadmill running | Inappropriate study design |
| 4011 | The six-minute walk test as a fall risk screening tool in community programs for persons with stroke: a cross-sectional analysis | Inappropriate study design |
| 4012 | Ottawa panel evidence-based clinical practice guidelines for post-stroke rehabilitation | Inappropriate study design |
| 4013 | A randomized feasibility study of a 12-week community-based exercise program for people with Huntington's disease | Inappropriate study design |
| 4014 | Assessment of patients' medical fitness to drive by primary care physicians: A cross-sectional study | Inappropriate outcome |
| 4015 | Exercise performance during captopril and atenolol treatment in hypertensive patients | Inappropriate population |
| 4016 | Effect of changes in arterial oxygen content on circulation and physical performance | Inappropriate study design |
| 4017 | Kinetics of cardiorespiratory response to dynamic and rhythmic-static exercise in men | Inappropriate population |
| 4018 | Echocardiographic evaluation of stress test for determining safety of participation in strength training | Inappropriate study design |

| | | |
|---|---|---|
| 4019 | Tai chi for upper limb rehabilitation in stroke patients: the patient's perspective | Inappropriate population |
| 4020 | Exercise unmasks distinct pathophysiologic features in heart failure with preserved ejection fraction and pulmonary vascular disease | Inappropriate intervention |
| 4021 | Successful treatment of post-exertion acute myocardial infarction by primary angioplasty and stenting in a patient with antiphospholipid antibody syndrome | Inappropriate study design |
| 4022 | Sustained haemodynamic effects of felodipine in patients with chronic cardiac failure | Inappropriate population |
| 4023 | Impact of contractile reserve on acute response to cardiac resynchronization therapy | Inappropriate study design |
| 4024 | Characterization of exercise limitations by evaluating individual cardiac output patterns: a prospective cohort study in patients with chronic heart failure | Inappropriate outcome |
| 4025 | Postprandial hyperglycemia is a possible contributor to paroxysmal atrial fibrillation: a case report | Inappropriate study design |
| 4026 | Impaired pulmonary oxygen uptake kinetics and reduced peak aerobic power during small muscle mass exercise in heart transplant recipients | Inappropriate population |
| 4027 | Biopsychosocial Predictors of Fall Events Among Older African Americans | Inappropriate study design |
| 4028 | The efficacy of ergometry determined heart rates for flatwater kayak training | Inappropriate study design |
| 4029 | Age-associated changes in cardiovascular structure and function: a fertile milieu for future disease | Inappropriate study design |
| 4030 | Exploring the relationship between exercise-induced arousal and cognition using fractionated response time | Inappropriate population |
| 4031 | Strength of the respiratory and lower limb muscles and functional capacity in chronic stroke survivors with different physical activity levels | Inappropriate population |
| 4032 | Ventilatory capacity and risk for dementia | Inappropriate population |
| 4033 | Sociodemographic Patterns of Chronic Disease: How the Mid-South Region Compares to the Rest of the Country | Inappropriate study design |
| 4034 | Oxygen uptake response to cycle ergometry in post-acute stroke patients with different severity of hemiparesis | Inappropriate study design |
| 4035 | Evaluation of a home-based exercise and training programme to improve sit-to-stand in patients with chronic stroke | Inappropriate population |
| 4036 | DREAMS-START (Dementia RElAted Manual for Sleep; STrAtegies for RelaTives) for people with dementia and sleep disturbances: a single-blind feasibility and acceptability randomized controlled trial | Inappropriate study design |
| 4037 | Quadriceps muscle strength is a discriminant predictor of dependence in daily activities in nursing home residents | Inappropriate population |
| 4038 | Left ventricular performance of the athletic heart during upright exercise: a heart rate-controlled study | Inappropriate population |
| 4039 | Isometric exercise in patients with chronic advanced heart failure: hemodynamic and neurohumoral evaluation | Inappropriate intervention |
| 4040 | Evaluation of myocardial performance during wheelchair ergometer exercise | Inappropriate population |
| 4041 | Polymorphisms of the DNA repair gene EXO1 modulate cognitive aging in old adults in a Taiwanese population | Inappropriate study design |
| 4042 | Captopril or nifedipine? Comparison of rest and exercise acute effects and long-term therapy in chronic isolated asymptomatic moderate to severe aortic regurgitation | Inappropriate study design |
| 4043 | POWER training in chronic stroke individuals: differences between responders and nonresponders | Inappropriate population |
| 4044 | IAP National Task Force for Childhood Prevention of Adult Diseases: insulin resistance and Type 2 diabetes mellitus in childhood | Inappropriate study design |
| 4045 | Oxygen utilization and ventilation during exercise in patients with chronic cardiac failure | Inappropriate study design |
| 4046 | Factors affecting the exercise capacity of pediatric patients with aortic regurgitation | Inappropriate population |
| 4047 | Variation and achievement of ambulatory activity among patients with chronic stroke | Inappropriate study design |
| 4048 | Exercise training mode affects the hemodynamic responses to lower body negative pressure in women | Inappropriate study design |
| 4049 | Effects of left ventricular diastolic dysfunction on exercise capacity three to six weeks after acute myocardial infarction in men | Inappropriate study design |
| 4050 | Left atrial function: bridge to central and hormonal determinants of exercise capacity in patients with chronic heart failure | Inappropriate study design |
| 4051 | Left ventricular volumes and hemodynamic responses at onset of dynamic exercise with reduced venous return | Inappropriate study design |
| 4052 | Maximal exercise test results in subacute stroke | Inappropriate study design |

| | | |
|---|---|---|
| 4053 | Development of a Sensor-Based Behavioral Monitoring Solution to Support Dementia Care | Inappropriate population |
| 4054 | Step by step: a proof of concept study of C-Mill gait adaptability training in the chronic phase after stroke | Inappropriate study design |
| 4055 | Long-term effects of physical training on coronary patients with impaired ventricular function | Inappropriate study design |
| 4056 | Sympatho-vagal changes induced by physical training in cardiac patients | Inappropriate study design |
| 4057 | Arterial vasodilatory and ventricular diastolic reserves determine the stroke volume response to exercise in elderly female hypertensive patients | Inappropriate population |
| 4058 | Does the use of TENS increase the effectiveness of exercise for improving walking after stroke? A randomized controlled clinical trial | Inappropriate study design |
| 4059 | Single-breath DLCO maneuver causes cardiac output to fall during and after cycling | Inappropriate study design |
| 4060 | Association between actigraphy-derived physical activity and cognitive performance in patients with schizophrenia | Inappropriate study design |
| 4061 | Low baseline urine creatinine excretion rate predicts poor outcomes among critically ill acute stroke patients | Inappropriate study design |
| 4062 | The effect of repeated supine exercise on exercise tolerance and left ventricular pump function in patients with angina pectoris | Inappropriate study design |
| 4063 | Effects of robotic-aided rehabilitation on recovery of upper extremity function in chronic stroke: a single case study | Inappropriate study design |
| 4064 | Prevalence and effect factors of dementia among the community elderly in Chongqing, China | Inappropriate study design |
| 4065 | Carotid chemoreceptors have a limited role in mediating the hyperthermia-induced hyperventilation in exercising humans | Inappropriate population |
| 4066 | Hemodynamic effects of calcium channel blockers at rest and during exercise in essential hypertension | Inappropriate study design |
| 4067 | Resistive training improves insulin sensitivity after stroke | Inappropriate population |
| 4068 | The acute effects of intravenous xamoterol ('Corwin', I.C.I. 118, 587) on resting and exercise haemodynamics in patients with mild to moderate heart failure | Inappropriate population |
| 4069 | Blood pressure responses during exercise testing-is up best for prognosis? | Inappropriate study design |
| 4070 | Combined prevalence of frailty and mild cognitive impairment in a population of elderly Japanese people | Inappropriate outcome |
| 4071 | Dietary nitrate supplementation and exercise tolerance in patients with heart failure with reduced ejection fraction | Inappropriate population |
| 4072 | Physiologic responses to prolonged electrically stimulated leg-cycle exercise in the spinal cord injured | Inappropriate study design |
| 4073 | Effects of electroacupuncture combined with computer-based cognitive rehabilitation on mild cognitive impairment: study protocol for a pilot randomized controlled trial | Inappropriate study design |
| 4074 | Motor Performance and Physical Activity as Predictors of Prospective Falls in Community-Dwelling Older Adults by Frailty Level: Application of Wearable Technology | Inappropriate intervention |
| 4075 | Clinical factors related to post-stroke fatigue within the first 3 month after acute cerebrovascular events | Inappropriate study design |
| 4076 | Social cognitive role of schizophrenia candidate gene GABRB2 | Inappropriate population |
| 4077 | Virtual reality-enhanced stroke rehabilitation | Inappropriate study design |
| 4078 | Stress-induced cardiomyopathy caused by heat stroke | Inappropriate population |
| 4079 | Is Physical Activity Causally Associated With Symptoms of Attention-Deficit/Hyperactivity Disorder? | Inappropriate population |
| 4080 | Clinical utility of the AlphaFIM® instrument in stroke rehabilitation | Inappropriate study design |
| 4081 | Effect of intensive rehabilitation on physical function and arterial function in community-dwelling chronic stroke survivors | Inappropriate outcome |
| 4082 | Noninvasive assessment of the pulmonary artery pressure response to exercise after uncomplicated heart transplantation | Inappropriate outcome |
| 4083 | Effects of Physical Exercise Interventions on Dual-Task Gait Speed Following Stroke: A Systematic Review and Meta-Analysis | Inappropriate population |
| 4084 | Protein and amino acid supplementation in older humans | Inappropriate population |
| 4085 | Acute and chronic effects of nitrendipine on hemodynamics and myocardial ischemia in patients with combined angina pectoris and hypertension | Inappropriate population |
| 4086 | Determinants of fast marathon performance: low basal sympathetic drive, enhanced postcompetition vasodilatation and preserved cardiac performance after competition | Inappropriate study design |

| | | |
|---|---|---|
| 4087 | Relationship of cardiac allograft size and pulmonary vascular resistance to long-term cardiopulmonary function | Inappropriate population |
| 4088 | Functional Reach, Depression Scores, and Number of Medications Are Associated With Number of Falls in People With Chronic Stroke | Inappropriate study design |
| 4089 | Health-related quality of life in long-term survivors of acute lymphoblastic leukemia in childhood and adolescence | Inappropriate study design |
| 4090 | A haemodynamic and radionuclide assessment of diltiazem in coronary heart disease | Inappropriate study design |
| 4091 | An evaluation of two different exercise regimes during the first year following stroke: a randomised controlled trial | Inappropriate study design |
| 4092 | Impact of protein and carbohydrate supplementation on plasma volume expansion and thermoregulatory adaptation by aerobic training in older men | Inappropriate population |
| 4093 | Risk factors for stroke in Thai patients | Inappropriate outcome |
| 4094 | Neuro-mechanics of muscle coordination during recumbent pedaling in post-acute stroke patients | Inappropriate population |
| 4095 | Association of physical activity and appetite with visual function related to driving competence in older adults | Inappropriate study design |
| 4096 | Hemodynamic effects of beta-blocking compounds possessing vasodilating activity: a review of labetalol, prizidilol, and dilevalol | Inappropriate study design |
| 4097 | Stressing the critically ill patient: the cardiopulmonary and metabolic responses to an acute increase in oxygen consumption | Inappropriate outcome |
| 4098 | A longitudinal analysis on the validity and reliability of ratings of perceived exertion for elite swimmers | Inappropriate population |
| 4099 | Remote Management of Poststroke Patients With a Smartphone-Based Management System Integrated in Clinical Care: Prospective, Nonrandomized, Interventional Study | Inappropriate study design |
| 4100 | Risk of Stroke-Associated Pneumonia With Acid-Suppressive Drugs: A Population-Based Cohort Study | Inappropriate population |
| 4101 | Prevalence of chronic disease and health practices in a retirement community | Inappropriate outcome |
| 4102 | Prolonged recovery of cardiac output after maximal exercise in patients with chronic heart failure | Inappropriate study design |
| 4103 | Effects of nicardipine on cardiac volume at rest and during exercise-induced angina | Inappropriate study design |
| 4104 | Combined captopril and hydrochlorothiazide therapy in severe hypertension: long-term haemodynamic changes at rest and during exercise | Inappropriate intervention |
| 4105 | Electrically induced recovery of gait components for older patients with chronic stroke | Inappropriate population |
| 4106 | Effect of swimming intensity on subsequent cycling and overall triathlon performance | Inappropriate population |
| 4107 | Central and peripheral hemodynamics during maximal leg extension exercise | Inappropriate outcome |
| 4108 | Physical activity mediates the relationship between fruit and vegetable consumption and cognitive functioning: a cross-sectional analysis | Inappropriate study design |
| 4109 | Use of Doppler ultrasound to assess femoral artery adaptations in the hemiparetic limb in people with stroke | Inappropriate population |
| 4110 | Acute effects of combined alpha/beta-adrenoceptor blockade v combined beta-receptor and slow channel calcium blockade in ischemic heart disease complicated by hypertension. Hemodynamic and adrenergic responses | Inappropriate study design |
| 4111 | Effects of rehabilitation on quality of life in patients with chronic stroke | Inappropriate population |
| 4112 | Long-term effects of 6-week whole-body vibration on balance recovery and activities of daily living in the postacute phase of stroke: a randomized, controlled trial | Inappropriate population |
| 4113 | Acute effect of a high nitrate diet on brain perfusion in older adults | Inappropriate study design |
| 4114 | Employment status among the Singapore elderly and its correlates | Inappropriate outcome |
| 4115 | Effects of Progressive Resistance Training on Cardiovascular Autonomic Regulation in Patients With Parkinson Disease: A Randomized Controlled Trial | Inappropriate study design |
| 4116 | Reduced skeletal muscle capillarization and glucose intolerance | Inappropriate study design |
| 4117 | Prevalence of 3 Healthy Lifestyle Behaviors Among US Adults With and Without History of Stroke | Inappropriate study design |
| 4118 | The effects of arm crank training on the physiological responses to submaximal wheelchair ergometry | Inappropriate population |
| 4119 | The haemodynamic response to submaximal exercise during isovolaemic haemodialysis | Inappropriate population |
| 4120 | Orthostatic hypotension after 10 days of exercise-heat acclimation and 28 hours of sleep loss | Inappropriate population |

| | | |
|---|---|---|
| 4121 | Influence of ergometer exercise on aortic pressure-diameter relationship in healthy men | Inappropriate outcome |
| 4122 | Improving Physical Activity Ensures the Long-Term Survival of Pneumonia Patients in a Super-Aged Society: A Retrospective Study in an Acute-Care Hospital in Japan | Inappropriate population |
| 4123 | Effects of High-Intensity Interval Training After Stroke (the HIIT-Stroke Study): A Multicenter Randomized Controlled Trial | Inappropriate study design |
| 4124 | Physical fitness and cardiopulmonary baroreflex to passive head-up tilt | Inappropriate study design |
| 4125 | Exercise hemodynamics and oxygen delivery in human hypertension. Response to verapamil | Inappropriate study design |
| 4126 | Cardiopulmonary function after lobectomy or pneumonectomy for pulmonary neoplasm | Inappropriate study design |
| 4127 | Specific exercise heat stress protocol for a triathlete's return from exertional heat stroke | Inappropriate population |
| 4128 | Arterial vasodilator therapy in heart failure: haemodynamic response patterns predictive of changes in exercise capacity | Inappropriate outcome |
| 4129 | Concomitant application of sprint and high-intensity interval training on maximal oxygen uptake and work output in well-trained cyclists | Inappropriate study design |
| 4130 | An exercise hemodynamic comparison of verapamil, diltiazem, and amlodipine in coronary artery disease | Inappropriate outcome |
| 4131 | What maintains the metabolic cost at maximal exercise in heart transplant recipients and coronary artery disease patients? | Inappropriate population |
| 4132 | Electromyogram-controlled assistive exercise for the motor recovery of shoulder in chronic hemiplegia: A pilot study | Inappropriate study design |
| 4133 | Changes in Cognitive Performance With Age in Adults With Multiple Sclerosis | Inappropriate study design |
| 4134 | Hemodynamic and neurohumoral responses to exercise in patients with congestive heart failure | Inappropriate study design |
| 4135 | Decreased cutaneous vasodilatation to isometric handgrip exercise in Alzheimer's disease | Inappropriate study design |
| 4136 | Effect of exercise training on left ventricular performance in older women free of cardiopulmonary disease | Inappropriate study design |
| 4137 | Does perturbation-based balance training prevent falls among individuals with chronic stroke? A randomised controlled trial | Inappropriate population |
| 4138 | Prediction of peak oxygen uptake in men using pulmonary and hemodynamic variables during exercise | Inappropriate study design |
| 4139 | Physiological comparison between non-athletes, endurance, power and team athletes | Inappropriate population |
| 4140 | Arterial oxygenation and arterial oxygen transport in chronic myocardial failure at rest, during exercise and after hydralazine treatment | Inappropriate study design |
| 4141 | Improvement in exercise haemodynamics by isosorbide dinitrate in patients with severe congestive cardiac failure secondary to ischaemic heart disease | Inappropriate intervention |
| 4142 | Left ventricular adaptations following short-term endurance training | Inappropriate population |
| 4143 | Caregiver burden and fatigue in caregivers of people with dementia: Measuring human herpesvirus (HHV)-6 and -7 DNA levels in saliva | Inappropriate study design |
| 4144 | The Effects of Supplementation with a Vitamin and Mineral Complex with Guaraná Prior to Fasted Exercise on Affect, Exertion, Cognitive Performance, and Substrate Metabolism: A Randomized Controlled Trial | Inappropriate study design |
| 4145 | Noninvasive positive pressure ventilation enhances the effects of aerobic training on cardiopulmonary function | Inappropriate study design |
| 4146 | Stroke volume response to cycle ergometry in trained and untrained older men | Inappropriate population |
| 4147 | Hemodynamic effects of moricizine at rest and during supine bicycle exercise: results in patients with ventricular tachycardia and left ventricular dysfunction | Inappropriate population |
| 4148 | A community-based group exercise program for persons with chronic stroke | Inappropriate population |
| 4149 | Felodipine improves pulmonary hemodynamics in chronic obstructive pulmonary disease | Inappropriate study design |
| 4150 | Immersion in warm water induces improvement in cardiac function in patients with chronic heart failure | Inappropriate study design |
| 4151 | Effect of changes in intrathoracic pressure on cardiac function at rest and during moderate exercise in health and heart failure | Inappropriate study design |
| 4152 | Low-dose captopril in chronic heart failure: acute haemodynamic effects and long-term treatment | Inappropriate study design |
| 4153 | Nutrition in the elderly: nutritional aspects of chronic diseases | Inappropriate study design |
| 4154 | Physiological responses to prolonged exercise in ultramarathon athletes | Inappropriate study design |

| | | |
|---|---|---|
| 4155 | The Effect of Habitual Physical Training on Left Ventricular Function During Exercise Assessed by Three-Dimensional Echocardiography | Inappropriate outcome |
| 4156 | Effect on spasticity after performance of dynamic-repeated-passive ankle joint motion exercise in chronic stroke patients | Inappropriate study design |
| 4157 | Does Pain Neuroscience Education and Cognition-Targeted Motor Control Training Improve Cervical Motor Output? Secondary Analysis of a Randomized Clinical Trial | Inappropriate study design |
| 4158 | EX-MET study: exercise in prevention on of metabolic syndrome - a randomized multicenter trial: rational and design | Inappropriate outcome |
| 4159 | Self-Care Self-Efficacy Correlates with Independence in Basic Activities of Daily Living in Individuals with Chronic Stroke | Inappropriate population |
| 4160 | Effectiveness of creatine monohydrate in mitochondrial encephalomyopathies | Inappropriate study design |
| 4161 | The electromyographic activity of the multifidus muscles during the execution of two pilates exercises--swan dive and breast stroke--for healthy people | Inappropriate study design |
| 4162 | Physical activity and risk of cerebrovascular disease in the European Prospective Investigation into Cancer and Nutrition-Spain study | Inappropriate study design |
| 4163 | Gender differences in cardiovascular and metabolic responses to cold and exercise | Inappropriate study design |
| 4164 | A hypothalamic stroke producing recurrent hemihyperhidrosis | Inappropriate study design |
| 4165 | Technology-Assisted Ankle Rehabilitation Improves Balance and Gait Performance in Stroke Survivors: A Randomized Controlled Study With 1-Month Follow-Up | Inappropriate study design |
| 4166 | Cardiac output in single-lead VDD pacing versus rate-matched VVIR pacing | Inappropriate study design |
| 4167 | Long-term central hemodynamic effects at rest and during exercise of losartan in essential hypertension | Inappropriate study design |
| 4168 | Post-resistance exercise hemodynamic and autonomic responses: Comparison between normotensive and hypertensive men | Inappropriate study design |
| 4169 | Reversal of cognitive impairment in a hypotensive elderly population using a passive exercise intervention | Inappropriate population |
| 4170 | Clinical management provided by board-certificated physiatrists in early rehabilitation is a significant determinant of functional improvement in acute stroke patients: a retrospective analysis of Japan rehabilitation database | Inappropriate study design |
| 4171 | Diabetes mellitus and cerebrovascular disease | Inappropriate population |
| 4172 | Biological markers of cardiac damage are not related to measures of cardiac systolic and diastolic function using cardiovascular magnetic resonance and echocardiography after an acute bout of prolonged endurance exercise | Inappropriate study design |
| 4173 | Systemic arterial response to exercise in patients with aortic valve stenosis | Inappropriate population |
| 4174 | Acute effect of static exercise in patients with aortic regurgitation assessed by cardiovascular magnetic resonance: role of left ventricular remodelling | Inappropriate population |
| 4175 | Contribution of Established Stroke Risk Factors to the Burden of Stroke in Young Adults | Inappropriate population |
| 4176 | Cardiovascular regulation during water immersion | Inappropriate study design |
| 4177 | Effect of graded erythrocythemia on cardiovascular and metabolic responses to exercise | Inappropriate outcome |
| 4178 | Exercise stroke volume relative to plasma-volume expansion | Inappropriate study design |
| 4179 | Effect of betaxolol on the hemodynamic, gas exchange, and cardiac output response to exercise in chronic atrial fibrillation | Inappropriate population |
| 4180 | Clinical and haemodynamic results following mitral valve replacement with the new Lillehei-Kaster pivoting disc valve prosthesis | Inappropriate study design |
| 4181 | Effects of posture on upper and lower limb peripheral resistance following submaximal cycling | Inappropriate population |
| 4182 | FES-assisted Cycling Improves Aerobic Capacity and Locomotor Function Postcerebrovascular Accident | Inappropriate study design |
| 4183 | COPD patients' self-reported adherence, psychosocial factors and mild cognitive impairment in pulmonary rehabilitation | Inappropriate study design |
| 4184 | Retraining postural responses with exercises emphasizing speed poststroke | Inappropriate population |
| 4185 | Prevention of heart failure in older adults may require higher levels of physical activity than needed for other cardiovascular events | Inappropriate study design |
| 4186 | Modest antihypertensive effect of epanolol, a beta 1-selective receptor blocker with beta 1 agonist activity: an acute and long-term hemodynamic study at rest and during exercise and double crossover comparison with atenolol on ambulatory blood pressure | Inappropriate outcome |

| | | |
|---|---|---|
| 4187 | Sensor-based arm skill training in chronic stroke patients: results on treatment outcome, patient motivation, and system usability | Inappropriate study design |
| 4188 | Usefulness of thoracic electrical bioimpedance in detection of ejection fraction changes | Inappropriate study design |
| 4189 | Limited Ventricular Preload is the Main Reason for Reduced Stress Reserve After Atrial Baffle Repair | Inappropriate study design |
| 4190 | Multiple factors limit exercise capacity in cystic fibrosis | Inappropriate study design |
| 4191 | Physical activity monitoring in stroke: SenseWear Pro2 activity accelerometer versus Yamax Digi-Walker SW-200 pedometer | Inappropriate study design |
| 4192 | The normal right ventricular response to supine exercise | Inappropriate study design |
| 4193 | Low-frequency electrical stimulation combined with a cooling vest improves recovery of elite kayakers following a simulated 1000-m race in a hot environment | Inappropriate study design |
| 4194 | Correlates of physical activity among community-dwelling adults aged 50 or over in six low- and middle-income countries | Inappropriate study design |
| 4195 | Effect of an anabolic steroid (metandienone) on central and peripheral blood flow in well-trained male athletes | Inappropriate study design |
| 4196 | Association of total daily physical activity with disability in community-dwelling older persons: a prospective cohort study | Inappropriate study design |
| 4197 | Improvement of left ventricular contractility and relaxation with the beta 1-adrenergic receptor partial agonist xamoterol at rest and during exercise in patients with postinfarction left ventricular dysfunction. A placebo-controlled randomized trial | Inappropriate population |
| 4198 | Lacunar stroke in a hypertensive football player: implications in managing hypertension in athletes | Inappropriate population |
| 4199 | Differing effects of airway obstruction on physical work capacity and ventilation in men and women with COPD | Inappropriate study design |
| 4200 | Impact of Tai Chi Yuttari-exercise on arteriosclerosis and physical function in older people | Inappropriate study design |
| 4201 | Postexercise decrease in arterial blood pressure, total peripheral resistance and in circulatory responses to brief hyperoxia in subjects with mild essential hypertension | Inappropriate population |
| 4202 | Pulmonary issues in the older adult | Inappropriate study design |
| 4203 | Evaluation of a long-acting converting enzyme inhibitor (enalapril) for the treatment of chronic congestive heart failure | Inappropriate study design |
| 4204 | Improved ventilatory response to exercise after cardioversion of chronic atrial fibrillation to sinus rhythm | Inappropriate study design |
| 4205 | Stroking parameters in front crawl swimming and maximal lactate steady state speed | Inappropriate population |
| 4206 | Application of combined botulinum toxin type A and modified constraint-induced movement therapy for an individual with chronic upper-extremity spasticity after stroke | Inappropriate study design |
| 4207 | Group III/IV muscle afferents impair limb blood in patients with chronic heart failure | Inappropriate study design |
| 4208 | An exploration of factors influencing physical activity levels amongst a cohort of people living in the community after stroke in the south of England | Inappropriate study design |
| 4209 | Older Adults with Dementia Are Sedentary for Most of the Day | Inappropriate study design |
| 4210 | The influence of endurance training on the transient haemodynamic response to orthostatic manoeuvre | Inappropriate study design |
| 4211 | Impairment-oriented training or Bobath therapy for severe arm paresis after stroke: a single-blind, multicentre randomized controlled trial | Inappropriate population |
| 4212 | Physical chronic conditions, multimorbidity and sedentary behavior amongst middle-aged and older adults in six low- and middle-income countries | Inappropriate study design |
| 4213 | Transient ischemic attacks in the elderly: diagnosis and treatment | Inappropriate study design |
| 4214 | Effects of intensive therapy using gait trainer or floor walking exercises early after stroke | Inappropriate outcome |
| 4215 | Acute heat stroke. Epidemiologic, biochemical, renal, and coagulation studies | Inappropriate population |
| 4216 | Improved cardiac performance during exercise following hydralazine treatment in chronic heart failure | Inappropriate study design |
| 4217 | Hypertension in the elderly patient | Inappropriate study design |
| 4218 | Effects of combined robotic therapy and repetitive-task practice on upper-extremity function in a patient with chronic stroke | Inappropriate study design |
| 4219 | Haemodynamic dose-response effects of i.v. nicardipine in coronary artery disease | Inappropriate population |

| | | |
|---|---|---|
| 4220 | Energy expenditure during front crawl swimming: predicting success in middle-distance events | Inappropriate population |
| 4221 | Impact of age on the cardiovascular response to dynamic upright exercise in healthy men and women | Inappropriate population |
| 4222 | Bone-density changes after stroke | Inappropriate study design |
| 4223 | Identifying factors associated with changes in physical functioning in an older population | Inappropriate population |
| 4224 | Independent associations of fasting insulin, glucose, and glycated haemoglobin with stroke and coronary heart disease in older women | Inappropriate population |
| 4225 | Whole-body intensive rehabilitation is feasible and effective in chronic stroke survivors: a retrospective data analysis | Inappropriate outcome |
| 4226 | Aerobic fitness does not influence the biventricular response to whole body passive heat stress | Inappropriate intervention |
| 4227 | Expiratory loading improves cardiac output during exercise in heart failure | Inappropriate study design |
| 4228 | Association between Carotid Artery Stenosis and Cognitive Impairment in Stroke Patients: A Cross-Sectional Study | Inappropriate population |
| 4229 | The time use and activity levels of inpatients in a co-located acute and rehabilitation stroke unit: an observational study | Inappropriate population |
| 4230 | Effect of exercise on placental blood flow in pregnancies complicated by hypertension, diabetes or intrahepatic cholestasis | Inappropriate outcome |
| 4231 | Captopril does not Potentiate Post-Exercise Hypotension: A Randomized Crossover Study | Inappropriate study design |
| 4232 | Environment, epigenetics and neurodegeneration: Focus on nutrition in Alzheimer's disease | Inappropriate study design |
| 4233 | Hemodynamic effects of nifedipine during upright exercise in stable angina pectoris and either normal or severely impaired left ventricular function | Inappropriate study design |
| 4234 | Effects of long use of a beta-blocker, carteolol, on exercise hemodynamics in hypertensives | Inappropriate study design |
| 4235 | Hypertension in the elderly | Inappropriate population |
| 4236 | Feasibility of cardiopulmonary exercise testing and training using a robotics-assisted tilt table in dependent-ambulatory stroke patients | Inappropriate population |
| 4237 | Post-Exercise Hypotension and Its Mechanisms Differ after Morning and Evening Exercise: A Randomized Crossover Study | Inappropriate study design |
| 4238 | Effects of propranolol and metoprolol on haemodynamic and respiratory indices and on perceived exertion during exercise in hypertensive patients | Inappropriate outcome |
| 4239 | Residual risk for acute stroke in patients with type 2 diabetes and hypertension in primary care: Skaraborg Hypertension and Diabetes Project | Inappropriate study design |
| 4240 | Changes in disability in older adults with generalized radiographic osteoarthritis: A complex relationship with physical activity | Inappropriate study design |
| 4241 | Benefits of training at moderate altitude versus sea level training in amateur runners | Inappropriate population |
| 4242 | Carvedilol in hypertension: effects on hemodynamics and 24-hour blood pressure | Inappropriate study design |
| 4243 | Hemodynamic effects of alinidine (ST 567) at rest and during exercise in patients with chronic congestive heart failure | Inappropriate study design |
| 4244 | Metabolomic correlates of aerobic capacity among elderly adults | Inappropriate study design |
| 4245 | Cardiac performance assessment during cardiopulmonary exercise test can improve the management of children with repaired congenital heart disease | Inappropriate population |
| 4246 | Peripheral chemoreceptor control of cardiovascular function at rest and during exercise in heart failure patients | Inappropriate intervention |
| 4247 | The effect of physical activity on mortality and cardiovascular disease in 130 000 people from 17 high-income, middle-income, and low-income countries: the PURE study | Inappropriate study design |
| 4248 | Driving in stroke survivors aged 18-65 years: The Psychosocial Outcomes In StrokE (POISE) Cohort Study | Inappropriate study design |
| 4249 | Mediating effects of body mass index, physical activity, and emotional distress on the relationship between short sleep and cardiovascular disease | Inappropriate study design |
| 4250 | Physical activity early after stroke and its association to functional outcome 3 months later | Inappropriate population |
| 4251 | Speed and duration of walking and other leisure time physical activity and the risk of heart failure: a prospective cohort study from the Copenhagen City Heart Study | Inappropriate population |
| 4252 | MDL 17,043: short- and long-term cardiopulmonary and clinical effects in patients with heart failure | Inappropriate study design |

| | | |
|---|---|---|
| 4253 | Effect of kinesthetic illusion induced by visual stimulation on ankle dorsiflexion dysfunction in a stroke patient: ABAB single-case design | Inappropriate study design |
| 4254 | Post-exercise Doppler-echocardiographic haemodynamics in idiopathic dilated cardiomyopathy after metoprolol infusion | Inappropriate outcome |
| 4255 | Predictors of thrombolysis in the telestroke and non telestroke settings for hypertensive acute ischemic stroke patients | Inappropriate study design |
| 4256 | Toward Understanding Ambulatory Activity Decline in Parkinson Disease | Inappropriate study design |
| 4257 | Lymphocyte beta adrenoceptor upregulation and improved cardiac response to adrenergic stimulation following converting enzyme inhibition in congestive heart failure | Inappropriate study design |
| 4258 | Associations between active travel and physical multi-morbidity in six low- and middle-income countries among community-dwelling older adults: A cross-sectional study | Inappropriate study design |
| 4259 | Resistive Training and Molecular Regulators of Vascular-Metabolic Risk in Chronic Stroke | Inappropriate population |
| 4260 | Efficacy and safety of ibopamine in congestive heart failure | Inappropriate study design |
| 4261 | Effect of inhaled iloprost on the exercise function of Fontan patients: a demonstration of concept | Inappropriate study design |
| 4262 | The Association Between Unhealthy Lifestyle Behaviors and the Prevalence of Chronic Kidney Disease (CKD) in Middle-Aged and Older Men | Inappropriate intervention |
| 4263 | Effect of work rate on cardiorespiratory response to rhythmic-static exercise | Inappropriate study design |
| 4264 | Temporal stability of ambulatory stroke volume and cardiac output measured by impedance cardiography | Inappropriate study design |
| 4265 | Improving memory in Parkinson's disease: a healthy brain ageing cognitive training program | Inappropriate study design |
| 4266 | Effect of infarct site on diastolic time during exercise | Inappropriate population |
| 4267 | Comparative haemodynamic effects of labetalol, timolol, prazosin and the combination of tolamolol and prazosin | Inappropriate study design |
| 4268 | Effects of exercise on left ventricular performance determined by echocardiography in chronic, severe mitral regurgitation secondary to mitral valve prolapse | Inappropriate population |
| 4269 | Impaired dynamic cerebral autoregulation at rest and during isometric exercise in type 2 diabetes patients | Inappropriate study design |
| 4270 | Hemodynamic response to exercise after beta-adrenergic blockade with propranolol in patients with mitral valve obstruction | Inappropriate study design |
| 4271 | Physiological responses to 1000-m ergometer time-trial performance in outrigger canoeing | Inappropriate study design |
| 4272 | Cardiovascular sensitivity to epinephrine in the trained and untrained states | Inappropriate population |
| 4273 | Exercise challenge in Gulf War Illness reveals two subgroups with altered brain structure and function | Inappropriate population |
| 4274 | Shorter intervals at peak SV vs.V̇O2max may yield high SV with less physiological stress | Inappropriate population |
| 4275 | Reduced stroke volume during exercise in postural tachycardia syndrome | Inappropriate study design |
| 4276 | Impaired chronotropic and vasodilator reserves limit exercise capacity in patients with heart failure and a preserved ejection fraction | Inappropriate study design |
| 4277 | Immediate and sustained hemodynamic and clinical improvement in chronic heart failure by an oral angiotensin-converting enzyme inhibitor | Inappropriate population |
| 4278 | Minimally assistive robot training for proprioception enhancement | Inappropriate population |
| 4279 | Cardiopulmonary Responses During Clinical and Laboratory Gait Assessments in People With Chronic Stroke | Inappropriate study design |
| 4280 | A pet therapy intervention with geriatric psychiatry inpatients | Inappropriate population |
| 4281 | Effects of intensive gait-oriented physiotherapy during early acute phase of stroke | Inappropriate population |
| 4282 | Impact of cardiac rehabilitation on peripheral circulation as assessed by impedance plethysmography: a randomized clinical trial | Inappropriate study design |
| 4283 | Hemodynamic-inotropic response to beta-blocker with intrinsic sympathomimetic activity in patients with congestive cardiomyopathy | Inappropriate study design |
| 4284 | A bicentric controlled study on the effects of aquatic Ai Chi in Parkinson disease | Inappropriate study design |
| 4285 | The hemodynamics and hemodynamic effects of indenolol in mild hypertension | Inappropriate population |
| 4286 | Longitudinal association between lifetime workforce participation and risk of self-reported cognitive decline in community-dwelling older adults | Inappropriate outcome |
| 4287 | Long-term indoramin therapy in congestive heart failure: a double-blind, randomized, parallel placebo-controlled trial | Inappropriate population |

| | | |
|---|---|---|
| 4288 | Effects of progressive backward body weight suppoted treadmill training on gait ability in chronic stroke patients: A randomized controlled trial | Inappropriate study design |
| 4289 | Gravity-supported exercise with computer gaming improves arm function in chronic stroke | Inappropriate study design |
| 4290 | Cardiac output response to exercise in chronic cardiac failure patients | Inappropriate population |
| 4291 | Promoting cognitive health and vitality: a review of clinical implications | Inappropriate study design |
| 4292 | Increases in intramuscular pressure raise arterial blood pressure during dynamic exercise | Inappropriate population |
| 4293 | Influence of heat stress and exercise intensity on vastus lateralis muscle and prefrontal cortex oxygenation | Inappropriate population |
| 4294 | Effect of sex on wasted left ventricular effort following maximal exercise | Inappropriate population |
| 4295 | Pet Ownership and the Risk of Dying from Cardiovascular Disease Among Adults Without Major Chronic Medical Conditions | Inappropriate population |
| 4296 | Balance ability, not muscle strength and exercise endurance, determines the performance of hemiparetic subjects on the timed-sit-to-stand test | Inappropriate study design |
| 4297 | Exercise rehabilitation restores physiological cardiovascular responses to short-term head-out water immersion in patients with chronic heart failure | Inappropriate study design |
| 4298 | Factors contributing to upper limb recovery after stroke: a survey of stroke survivors in Queensland Australia | Inappropriate population |
| 4299 | Interaction between nifedipine and atenolol: pharmacokinetics and pharmacodynamics in normotensive volunteers | Inappropriate study design |
| 4300 | Central hemodynamic changes of calcium antagonists at rest and during exercise in essential hypertension | Inappropriate study design |
| 4301 | A dose-response relationship between amount of weight-bearing exercise and walking outcome following cerebrovascular accident | Inappropriate population |
| 4302 | Acute inotropic stimulation with dopamine in severe congestive heart failure: beneficial hemodynamic effect at rest but not during maximal exercise | Inappropriate population |
| 4303 | Tibial bone geometry in chronic stroke patients: influence of sex, cardiovascular health, and muscle mass | Inappropriate study design |
| 4304 | Aerobic interval training compensates age related decline in cardiac function | Inappropriate population |
| 4305 | The role of insulin and neurotrophic factor signaling in brain aging and Alzheimer's Disease | Inappropriate population |
| 4306 | Genome-wide autozygosity is associated with lower general cognitive ability | Inappropriate population |
| 4307 | Most functional outcomes are similar for men and women after hip fracture: a secondary analysis of the enhancing mobility after hip fracture trial | Inappropriate population |
| 4308 | Greater stroke severity predominates over all other factors for the worse outcome of cardioembolic stroke | Inappropriate study design |
| 4309 | Mechanisms for increasing stroke volume during static exercise with fixed heart rate in humans | Inappropriate population |
| 4310 | Effects of exercise and horticultural intervention on the brain and mental health in older adults with depressive symptoms and memory problems: study protocol for a randomized controlled trial [UMIN000018547 | Inappropriate study design |
| 4311 | Simultaneous determination of the kinetics of cardiac output, systemic O(2) delivery, and lung O(2) uptake at exercise onset in men | Inappropriate study design |
| 4312 | Stroke and dyslipidemia: clinical risk factors in the telestroke versus non-telestroke | Inappropriate study design |
| 4313 | Haemodynamic effects of saluretic treatment and beta-receptor blockade in patients with essential hypertension | Inappropriate outcome |
| 4314 | A Multicomponent Behavioral Intervention to Reduce Stroke Risk Factor Behaviors: The Stroke Health and Risk Education Cluster-Randomized Controlled Trial | Inappropriate population |
| 4315 | Hemodynamic response to postural shift in women with multiple chemical sensitivities | Inappropriate study design |
| 4316 | Will a healthy lifestyle help prevent Alzheimer's disease? | Inappropriate outcome |
| 4317 | Association between heart rate variability and haemodynamic response to exercise in chronic heart failure | Inappropriate study design |
| 4318 | Cardiac output in paraplegic subjects at high exercise intensities | Inappropriate study design |
| 4319 | Balancing the playing field: collaborative gaming for physical training | Inappropriate population |
| 4320 | Hemodynamic alterations in alcohol-related transitory hypertension | Inappropriate study design |
| 4321 | Evaluation of inert gas rebreathing for determination of cardiac output: influence of age, gender and body size | Inappropriate intervention |
| 4322 | Postural effect on cardiac output, oxygen uptake and lactate during cycle exercise of varying intensity | Inappropriate intervention |

| | | |
|---|---|---|
| 4323 | Cerebral Blood Flow Responses to Aquatic Treadmill Exercise | Inappropriate study design |
| 4324 | An EEG Tool for Monitoring Patient Engagement during Stroke Rehabilitation: A Feasibility Study | Inappropriate study design |
| 4325 | Impaired central hemodynamic response and exaggerated vasoconstriction during muscle metaboreflex activation in heart failure patients | Inappropriate study design |
| 4326 | Decreased peak arteriovenous oxygen difference during treadmill exercise testing in individuals infected with the human immunodeficiency virus | Inappropriate study design |
| 4327 | Exercise training enhances endogenous fibrinolysis in peripheral arterial disease | Inappropriate study design |
| 4328 | Primary prevention of dementia: from modifiable risk factors to a public brain health agenda? | Inappropriate study design |
| 4329 | Cardiovascular and hormonal responses to bicycle exercise during lower body negative pressure | Inappropriate study design |
| 4330 | Cardiovascular response to passive cycle exercise | Inappropriate study design |
| 4331 | Lower risk of incident dementia among Chinese older adults having three servings of vegetables and two servings of fruits a day | Inappropriate population |
| 4332 | Influence of combined exercise and gravity transients and apnea on hemodynamics | Inappropriate population |
| 4333 | Validity of the adaptive filter for accurate measurement of cardiac output in impedance cardiography | Inappropriate outcome |
| 4334 | Public Awareness of Stroke and the Appropriate Responses in China: A Cross-Sectional Community-Based Study (FAST-RIGHT) | Inappropriate study design |
| 4335 | Cardiovascular effects of prenalterol on rest and exercise haemodynamics in patients with chronic congestive cardiac failure | Inappropriate population |
| 4336 | Radiofrequency maze ablation for atrial fibrillation in a patient undergoing atrial septal defect repair | Inappropriate population |
| 4337 | Differential MicroRNA expression following head-down tilt bed rest: implications for cardiovascular responses to microgravity | Inappropriate population |
| 4338 | First clinical trial with etomoxir in patients with chronic congestive heart failure | Inappropriate study design |
| 4339 | Effects of inclined treadmill training on functional and cardiovascular parameters of stroke patients: study protocol for a randomized controlled trial | Inappropriate study design |
| 4340 | Detecting abnormalities in left ventricular function during exercise by respiratory measurement | Inappropriate study design |
| 4341 | Exercise testing in pulmonary arterial hypertension and in chronic heart failure | Inappropriate intervention |
| 4342 | Time Course of Flow-Mediated Dilation and Vascular Endothelial Growth Factor following Acute Stroke | Inappropriate study design |
| 4343 | Cardiac output and associated left ventricular hypertrophy in pediatric chronic kidney disease | Inappropriate outcome |
| 4344 | Exercise tolerance in a hot and humid climate in heat-acclimatized girls and women | Inappropriate study design |
| 4345 | Effect of biventricular pacing on the exercise pathophysiology of heart failure | Inappropriate study design |
| 4346 | The use of a treadmill with biofeedback function in assessment of relearning walking skills in post-stroke hemiplegic patients--a preliminary report | Inappropriate study design |
| 4347 | Effects of therapeutic Tai Chi on balance, gait, and quality of life in chronic stroke patients | Inappropriate study design |
| 4348 | Exercise-induced and nitroglycerin-induced myocardial preconditioning improves hemodynamics in patients with angina | Inappropriate study design |
| 4349 | Depression, fatigue and disability are independently associated with quality of life in patients with multiple Sclerosis: Results of a cross-sectional study | Inappropriate population |
| 4350 | The effectiveness of body weight-supported gait training and floor walking in patients with chronic stroke | Inappropriate population |
| 4351 | Impaired left ventricular function during exercise in coronary artery disease and exertional hypotension | Inappropriate population |
| 4352 | Effects of movement music therapy with a percussion instrument on physical and frontal lobe function in older adults with mild cognitive impairment: a randomized controlled trial | Inappropriate study design |
| 4353 | Preservation of directly stimulated muscle strength in hemiplegia due to stroke | Inappropriate study design |
| 4354 | Effect of exercise on maternal hemodynamics and placental blood flow in healthy women | Inappropriate study design |
| 4355 | The effects of a protein enriched diet with lean red meat combined with a multi-modal exercise program on muscle and cognitive health and function in older adults: study protocol for a randomised controlled trial | Inappropriate study design |
| 4356 | Cardiovascular responses during recovery from exercise and thermal stress | Inappropriate study design |

| | | |
|---|---|---|
| 4357 | The effects of a multicomponent dyadic intervention on the mood, behavior, and physical health of people with dementia: a randomized controlled trial | Inappropriate study design |
| 4358 | Knee-high compression socks minimize head-up tilt-induced cerebral and cardiovascular responses following dynamic exercise | Inappropriate study design |
| 4359 | Assessment of Physical Activity of Hospitalised Older Adults: A Systematic Review | Inappropriate population |
| 4360 | Physical multimorbidity and sedentary behavior in older adults: Findings from the Irish longitudinal study on ageing (TILDA) | Inappropriate population |
| 4361 | Exercise training prevents decline in stroke volume during exercise in young healthy subjects | Inappropriate study design |
| 4362 | Cardiovascular function during early recovery from acute myocardial infarction | Inappropriate study design |
| 4363 | Cost effectiveness of community-based physical activity interventions | Inappropriate outcome |
| 4364 | The influence of thoracic epidural analgesia on the circulation at rest and during physical exercise in man | Inappropriate study design |
| 4365 | The QT interval and cycle length: the influence of atropine, hyoscine and exercise | Inappropriate study design |
| 4366 | Leisure time physical activity and change in body mass index: an 11-year follow-up study of 9357 normal weight health women 20-49 years old | Inappropriate study design |
| 4367 | The Rehabilitation of Patients With Advanced Heart Failure After Left Ventricular Assist Device Placement: A Narrative Review | Inappropriate study design |
| 4368 | Cardiovascular adjustments in breath-hold diving: comparison between divers and non-divers in simulated dynamic apnoea | Inappropriate population |
| 4369 | Cardiac responses to exercise distinguish postural orthostatic tachycardia syndrome variants | Inappropriate study design |
| 4370 | Exercise tolerance and quality of life in elderly patients with chronic atrial fibrillation | Inappropriate population |
| 4371 | Cardiovascular response to orthostatic stress: effects of exercise training modality | Inappropriate population |
| 4372 | Prenalterol, an oral beta-1 adrenoceptor agonist, in the treatment of chronic heart failure | Inappropriate population |
| 4373 | Turn the Beat Around: A Stroke Prevention Program for African-American Churches | Inappropriate study design |
| 4374 | Effects of physiological salt water injection after 20 days of head down tilt bed rest on maximal aerobic power in young males | Inappropriate study design |
| 4375 | Type 2 diabetes: causes, complications, and new screening recommendations. I | Inappropriate population |
| 4376 | Sympathoadrenal-circulatory regulation during sustained isometric exercise in young and older men | Inappropriate population |
| 4377 | optimizing self-exercise scheduling in motor stroke using Challenge Point Framework theory | Inappropriate study design |
| 4378 | Muscle activity and vibration transmissibility during whole-body vibration in chronic stroke | Inappropriate population |
| 4379 | Splenic constriction during isometric handgrip exercise in humans | Inappropriate population |
| 4380 | Associations between cardiorespiratory responses to exercise and the C34T AMPD1 gene polymorphism in the HERITAGE Family Study | Inappropriate population |
| 4381 | Systemic, pulmonary, and coronary hemodynamic effects of labetalol in hypertensive subjects | Inappropriate study design |
| 4382 | Hemodynamic performance during exercise in patients with severe chronic congestive heart failure before and after a single dose of pimobendan | Inappropriate study design |
| 4383 | Augmented mean systolic ejection rate in coronary patients treated with diltiazem | Inappropriate study design |
| 4384 | Intracerebral haemorrhage and Qigong | Inappropriate intervention |
| 4385 | Haemodynamic response to exercise in healthy young and elderly subjects | Inappropriate study design |
| 4386 | Mind over matter--what do we know about neuroplasticity in adults? | Inappropriate study design |
| 4387 | Circulatory responses at the onset of handgrip exercise in patients with Parkinson's disease | Inappropriate study design |
| 4388 | Challenging rehabilitation environment for older patients | Inappropriate study design |
| 4389 | Interpretation of high wedge pressure on exercise in patients with chronic obstructive pulmonary disease | Inappropriate population |
| 4390 | Physical activity participation and the risk of chronic diseases among South Asian adults: a systematic review and meta-analysis | Inappropriate study design |
| 4391 | Robot-assisted Guitar Hero for finger rehabilitation after stroke | Inappropriate population |
| 4392 | Forearm vascular responses to baroreceptor unloading at the onset of dynamic exercise | Inappropriate study design |
| 4393 | Health behavior change following chronic illness in middle and later life | Inappropriate study design |

| | | |
|---|---|---|
| 4394 | Treatment patterns, risk factor control and functional capacity in patients with cardiovascular and chronic kidney disease in the cardiac rehabilitation setting | Inappropriate population |
| 4395 | Potential benefits of creatine monohydrate supplementation in the elderly | Inappropriate outcome |
| 4396 | Effects of weight-shift training on walking ability, ambulation, and weight distribution in individuals with chronic stroke: a pilot study | Inappropriate population |
| 4397 | Subarachnoid Haemorrhage and Sports | Inappropriate population |
| 4398 | Effects of dual-task aquatic exercises on functional mobility, balance and gait of individuals with Parkinson's disease: A randomized clinical trial with a 3-month follow-up | Inappropriate study design |
| 4399 | The effect of exercise of the affected foot in stroke patients--a randomized controlled pilot trial | Inappropriate study design |
| 4400 | Nutrient intake and dietary changes during a 2-year multi-domain lifestyle intervention among older adults: secondary analysis of the Finnish Geriatric Intervention Study to Prevent Cognitive Impairment and Disability (FINGER) randomised controlled trial | Inappropriate population |
| 4401 | Lifestyle behaviors, chronic diseases, and ratings of health between black and white adults with pre-diabetes | Inappropriate study design |
| 4402 | Blunted responses of doppler-derived aortic flow parameters during whole-body heavy isometric exercise in heart transplant recipients | Inappropriate population |
| 4403 | Effectiveness of the physical activity intervention program in the PREDIMED-Plus study: a randomized controlled trial | Inappropriate population |
| 4404 | Cardiovascular control during voluntary static exercise in humans with tetraplegia | Inappropriate study design |
| 4405 | The development and implementation of stroke risk prediction model in National Health Insurance Service's personal health record | Inappropriate population |
| 4406 | Effect of acute alterations of blood volume on circulatory performance in humans | Inappropriate population |
| 4407 | Is there a relationship between insulin resistance and frailty syndrome? | Inappropriate population |
| 4408 | The role of reduced fitness to drive due to medical impairments in explaining crashes involving older drivers | Inappropriate study design |
| 4409 | Enhanced life-role participation in response to comprehensive gait training in chronic-stroke survivors | Inappropriate outcome |
| 4410 | The Link between Fetal Programming, Inflammation, Muscular Strength, and Blood Pressure | Inappropriate population |
| 4411 | Singapore Tele-technology Aided Rehabilitation in Stroke (STARS) trial: protocol of a randomized clinical trial on tele-rehabilitation for stroke patients | Inappropriate study design |
| 4412 | Is impact of fatigue an independent factor associated with physical activity in patients with idiopathic Parkinson's disease? | Inappropriate study design |
| 4413 | Exploratory longitudinal cohort study of associations of fatigue after stroke | Inappropriate population |
| 4414 | Implementation of High-Intensity Stepping Training During Inpatient Stroke Rehabilitation Improves Functional Outcomes | Inappropriate intervention |
| 4415 | Disability and its correlates with chronic morbidities among U.S. adults aged 50-<65 years | Inappropriate population |
| 4416 | The effect of resistance training on left ventricular function and structure of patients with chronic heart failure | Inappropriate study design |
| 4417 | Continuous haemodynamic ambulatory monitoring in essential hypertension | Inappropriate study design |
| 4418 | Feasibility and effectiveness of circuit training in acute stroke rehabilitation | Inappropriate population |
| 4419 | β-Adrenergic Responsiveness in the Type 2 Diabetic Heart: Effects on Cardiac Reserve | Inappropriate study design |
| 4420 | Improving stroke prevention in the French West Indies: limits to lay knowledge of risk factors | Inappropriate study design |
| 4421 | Balance and Gait After First Minor Ischemic Stroke in People 70 Years of Age or Younger: A Prospective Observational Cohort Study | Inappropriate intervention |
| 4422 | Comparison between group and personal rehabilitation for dementia in a geriatric health service facility: single-blinded randomized controlled study | Inappropriate study design |
| 4423 | Long-term outcomes of lercanidipine versus other calcium channel blockers in newly diagnosed hypertension: a nationwide cohort study | Inappropriate intervention |
| 4424 | Early markers of kidney dysfunction and cognitive impairment among older adults | Inappropriate population |
| 4425 | Increased risk of stress fractures of the ribs in elite rowers | Inappropriate study design |
| 4426 | Warm-up intensity and duration's effect on traditional rowing time-trial performance | Inappropriate study design |

| | | |
|---|---|---|
| 4427 | Dose-response relationship between physical activity and mortality in people with non-communicable diseases: a study protocol for the systematic review and meta-analysis of cohort studies | Inappropriate study design |
| 4428 | Age and gender affect ventricular-vascular coupling during aerobic exercise | Inappropriate study design |
| 4429 | Cardiovascular deconditioning occurs during a 7-day saturation dive at 31 ATA | Inappropriate study design |
| 4430 | Autonomic function is impaired in elderly stroke survivors | Inappropriate population |
| 4431 | Genome-wide linkage scan for exercise stroke volume and cardiac output in the HERITAGE Family Study | Inappropriate population |
| 4432 | Moderate intensity supine exercise causes decreased cardiac volumes and increased outer volume variations: a cardiovascular magnetic resonance study | Inappropriate study design |
| 4433 | Evaluation of upper extremity robot-assistances in subacute and chronic stroke subjects | Inappropriate study design |
| 4434 | Comprehensive measurement of stroke gait characteristics with a single accelerometer in the laboratory and community: a feasibility, validity and reliability study | Inappropriate study design |
| 4435 | The use of commercially available games for a combined physical and cognitive challenge during exercise for individuals with Parkinson's disease - a case series report | Inappropriate study design |
| 4436 | Effectiveness of force application in manual wheelchair propulsion in persons with spinal cord injuries | Inappropriate population |
| 4437 | The effects of exercise at high altitude on high-sensitivity cardiac troponin release and associated biventricular cardiac function | Inappropriate study design |
| 4438 | Muscle sympathetic nerve responses to passive and active one-legged cycling: insights into the contributions of central command | Inappropriate study design |
| 4439 | Functional mitral regurgitation during exercise in patients with heart failure | Inappropriate outcome |
| 4440 | Robot-assisted arm trainer for the passive and active practice of bilateral forearm and wrist movements in hemiparetic subjects | Inappropriate study design |
| 4441 | Effects of chin tuck against resistance exercise versus Shaker exercise on dysphagia and psychological state after cerebral infarction | Inappropriate study design |
| 4442 | Blunted hemodynamic response and reduced oxygen delivery with exercise in anemic heart failure patients with systolic dysfunction | Inappropriate study design |
| 4443 | Quality of life in post-stroke patients: self-evaluation of physical and mental health during six months | Inappropriate study design |
| 4444 | Effect of stroke rate reduction on swimming technique during paced exercise | Inappropriate population |
| 4445 | Eating behaviors and their relationship with cardiovascular disease. A case/case-control study | Inappropriate study design |
| 4446 | Sex differences in blood pressure regulation during ischemic isometric exercise: the role of the β-adrenergic receptors | Inappropriate study design |
| 4447 | Right ventricular dysfunction after major pulmonary resection | Inappropriate study design |
| 4448 | Small unrepaired atrial septal defects display impaired exercise capacity compared with healthy peers | Inappropriate study design |
| 4449 | Capturing ambulatory activity decline in Parkinson's disease | Inappropriate study design |
| 4450 | Robotic Exoskeleton for Wrist and Fingers Joint in Post-Stroke Neuro-Rehabilitation for Low-Resource Settings | Inappropriate population |
| 4451 | High-Intensity Respiratory Muscle Training Improves Strength and Dyspnea Poststroke: A Double-Blind Randomized Trial | Inappropriate population |
| 4452 | Does post-exercise ST depression reflect local ischemia or some global effect in the left ventricle? | Inappropriate population |
| 4453 | Endurance training in the elderly nursing home patient | Inappropriate study design |
| 4454 | Symmetric cardiac enlargement in highly trained endurance athletes: a two-dimensional echocardiographic study | Inappropriate study design |
| 4455 | Physical activity, smoking, and alcohol consumption in association with incidence of type 2 diabetes among middle-aged and elderly Chinese men | Inappropriate study design |
| 4456 | Pharmacokinetic, haemodynamic and radionuclide studies with nicardipine in coronary artery disease | Inappropriate study design |
| 4457 | A cooperative multicenter study of captopril in congestive heart failure: hemodynamic effects and long-term response | Inappropriate study design |
| 4458 | Insulin resistance in non-diabetic subjects is associated with increased incidence of myocardial infarction and death | Inappropriate study design |
| 4459 | What stroke patients want to know and what medical professionals think they should know about stroke: Korean perspectives | Inappropriate study design |
| 4460 | Effects of exercise in normobaric hypoxia on hemodynamics during muscle metaboreflex activation in normoxia | Inappropriate study design |

| | | |
|---|---|---|
| 4461 | The effects of four months' treatment with spironolactone on systemic blood pressure, cardiac output and plasma renin activity in hypertensive patients | Inappropriate population |
| 4462 | Blood flow in the brachial artery increases after intense cycling exercise | Inappropriate study design |
| 4463 | Captopril versus placebo in congestive heart failure: effects on oxygen delivery to exercising skeletal muscle | Inappropriate outcome |
| 4464 | The hemodynamic responses to upright exercise after orthotopic cardiac transplant | Inappropriate study design |
| 4465 | Isometric muscle function of knee extensors and the relation with functional performance in patients with stroke | Inappropriate study design |
| 4466 | Alterations in ventricular mass and performance induced by exercise training in man evaluated by echocardiography | Inappropriate study design |
| 4467 | A Novel Framework for Estimating Time-Varying Multivariate Autoregressive Models and Application to Cardiovascular Responses to Acute Exercise | Inappropriate study design |
| 4468 | Cardiorespiratory reactions to static, isometric exercise in man | Inappropriate study design |
| 4469 | Relationship between functional vision and balance and mobility performance in community-dwelling older adults | Inappropriate study design |
| 4470 | Relationships between activity and well-being in people with parkinson's disease | Inappropriate study design |
| 4471 | Increasing access to chronic disease self-management programs in rural and remote communities using telehealth | Inappropriate intervention |
| 4472 | The relationship of walking intensity to total and cause-specific mortality. Results from the National Walkers' Health Study | Inappropriate study design |
| 4473 | Exercise cardiac output following Skylab missions: the second manned Skylab mission | Inappropriate study design |
| 4474 | Left ventricular peak power during exercise: a noninvasive approach for assessment of contractile reserve | Inappropriate study design |
| 4475 | Divergent pathways to influence: Cognition and behavior differentially mediate the effects of optimism on physical and mental quality of life in Chinese university students | Inappropriate study design |
| 4476 | Factors Associated With Responsiveness to Gait and Balance Training in People With Parkinson Disease | Inappropriate population |
| 4477 | The stroke-related effects of hip flexion fatigue on over ground walking | Inappropriate study design |
| 4478 | Relationships between daily acute glucose fluctuations and cognitive performance among aged type 2 diabetic patients | Inappropriate population |
| 4479 | Associations of cognition with physical functioning and health-related quality of life among COPD patients | Inappropriate study design |
| 4480 | Acute and long-term effects of captopril on exercise cardiac performance and exercise capacity in congestive heart failure | Inappropriate study design |
| 4481 | Cerebral palsy improvement achieved by coordination dynamics therapy | Inappropriate study design |
| 4482 | Impedance cardiography for cardiac output measurement: an evaluation of accuracy and limitations | Inappropriate study design |
| 4483 | Effect of endurance training on the determinants of peak exercise oxygen consumption in elderly patients with stable compensated heart failure and preserved ejection fraction | Inappropriate study design |
| 4484 | Exercise with a twist: left ventricular twist and recoil in healthy young and middle-aged men, and middle-aged endurance-trained men | Inappropriate study design |
| 4485 | Cardiovascular circulatory adjustments and renal function in acute heart failure | Inappropriate population |
| 4486 | Relationship of exercise capacity to resting left ventricular performance and basal plasma norepinephrine levels in patients with congestive heart failure | Inappropriate study design |
| 4487 | Cardiac function at rest and during exercise in normals and in patients with coronary heart disease: evaluation by radionuclide angiocardiography | Inappropriate outcome |
| 4488 | Insulin resistance is associated with the pathology of Alzheimer disease: the Hisayama study | Inappropriate study design |
| 4489 | The Rutgers Arm, a rehabilitation system in virtual reality: a pilot study | Inappropriate study design |
| 4490 | Cognition and Indicators of Dietary Habits in Older Adults from Southern Brazil | Inappropriate population |
| 4491 | Participants' experiences of an Education, self-management and upper extremity eXercise Training for people with Rheumatoid Arthritis programme (EXTRA) | Inappropriate population |
| 4492 | Efficacy of Computer-Based Telephone Counseling on Long-Term Adherence to Strength Training in Elderly Patients With Knee Osteoarthritis: A Randomized Trial | Inappropriate outcome |
| 4493 | Above-moderate physical activity reduces both incident and persistent late-life depression in rural Koreans | Inappropriate population |
| 4494 | The costs of caring for stroke patients in a GP-led community hospital: an application of programme budgeting and marginal analysis | Inappropriate population |

| | | |
|---|---|---|
| 4495 | Combined oral contraceptives do not influence post-exercise hypotension in women | Inappropriate intervention |
| 4496 | Effects of training with a passive hand orthosis and games at home in chronic stroke: a pilot randomised controlled trial | Inappropriate study design |
| 4497 | Effect of wheelchair stroke pattern on mechanical efficiency | Inappropriate study design |
| 4498 | Exercise training in patients with severe chronic heart failure: impact on left ventricular performance and cardiac size. A retrospective analysis of the Leipzig Heart Failure Training Trial | Inappropriate intervention |
| 4499 | Insulin resistance and cognitive test performance in elderly adults: National health and nutrition examination survey (NHANES) | Inappropriate population |
| 4500 | Effect of New Zealand Blackcurrant Extract on Physiological Responses at Rest and during Brisk Walking in Southeast Asian Men: A Randomized, Double-Blind, Placebo-Controlled, Crossover Study | Inappropriate study design |
| 4501 | Lifelong Exercise Patterns and Cardiovascular Health | Inappropriate study design |
| 4502 | Chronic hemodynamic effects of tiapamil and felodipine in essential hypertension at rest and during exercise | Inappropriate population |
| 4503 | Risk factors for stroke in different levels of cerebral arterial disease | Inappropriate study design |
| 4504 | Response of blood pressure, cardiac output, peripheral resistance, and exercise performance to substitution of calcium blocker for beta-blocker plus thiazide diuretic therapy in patients with both systemic hypertension and mild stable angina | Inappropriate study design |
| 4505 | Exercise haemodynamics in patients over 65 years of age with recent myocardial infarction | Inappropriate outcome |
| 4506 | Comparison of exercise training effect with different robotic devices for upper limb rehabilitation: a retrospective study | Inappropriate study design |
| 4507 | Pulmonary artery hemodynamics in primary pulmonary hypertension | Inappropriate study design |
| 4508 | Skeletal muscle changes following stroke: a systematic review and comparison to healthy individuals | Inappropriate study design |
| 4509 | Late hemodynamic effects of the preserved papillary muscles during mitral valve replacement | Inappropriate study design |
| 4510 | Use of an abbreviated neuroscience education approach in the treatment of chronic low back pain: a case report | Inappropriate study design |
| 4511 | Right ventricular metabolic adaptations to high-intensity interval and moderate-intensity continuous training in healthy middle-aged men | Inappropriate study design |
| 4512 | Evaluation of the impact of exercise of gait on a treadmill on balance of people who suffered from cerebral stroke | Inappropriate study design |
| 4513 | Vasoactive mediators and renal haemodynamics in exertional heat stroke complicated by acute renal failure | Inappropriate study design |
| 4514 | A randomized controlled trial of functional neuromuscular stimulation in chronic stroke subjects | Inappropriate study design |
| 4515 | Long-term digitalis therapy improves left ventricular function in heart failure | Inappropriate population |
| 4516 | Factors influencing Doppler indexes of left ventricular filling in healthy persons | Inappropriate study design |
| 4517 | Short-term and long-term hemodynamic and clinical effects of metoprolol alone and combined with amlodipine in patients with chronic heart failure | Inappropriate intervention |
| 4518 | Benzodiazepines: tolerability in elderly patients | Inappropriate study design |
| 4519 | Does central fatigue explain reduced cycling after complete sleep deprivation? | Inappropriate study design |
| 4520 | Hemodynamic responses to isolated increments in heart rate by atrial pacing after a Fontan procedure | Inappropriate study design |
| 4521 | Thermal and circulatory responses to repeated bouts of prolonged running | Inappropriate study design |
| 4522 | Comparison between multicomponent and simultaneous dual-task exercise interventions in long-term nursing home residents: the Ageing-ONDUAL-TASK randomized controlled study | Inappropriate population |
| 4523 | Hemodynamic responses to exercise after lung transplantation | Inappropriate study design |
| 4524 | Mitochondrial myopathy with anemia, cardiomyopathy, and lactic acidosis: a distinct late onset mitochondrial disorder | Inappropriate outcome |
| 4525 | Effectiveness of preload reserve as a determinant of clinical status in patients with left ventricular systolic dysfunction. The SOLVD Investigators | Inappropriate population |
| 4526 | Temporal distribution of death among oncology patients: environmental links | Inappropriate outcome |
| 4527 | Psychiatric morbidity and acute hospitalization in elderly people | Inappropriate outcome |
| 4528 | Vasodilator treatment for mild to moderate heart failure. Effect of Ro 12-4713, a minoxidil analogue | Inappropriate study design |
| 4529 | Modifiable lifestyle factors associated with metabolic syndrome in patients with psoriasis | Inappropriate outcome |

| | | |
|---|---|---|
| 4530 | The effect of dietary flaxseed on improving symptoms of cardiovascular disease in patients with peripheral artery disease: rationale and design of the FLAX-PAD randomized controlled trial | Inappropriate population |
| 4531 | Cardiovascular adjustments to exercise distributed between the upper and lower body | Inappropriate outcome |
| 4532 | Isometric stress in cardiovascular magnetic resonance-a simple and easily replicable method of assessing cardiovascular differences not apparent at rest | Inappropriate intervention |
| 4533 | Heart failure and movement-induced hemodynamics: partitioning the impact of central and peripheral dysfunction | Inappropriate intervention |
| 4534 | Smoking, drinking, diet and physical activity-modifiable lifestyle risk factors and their associations with age to first chronic disease | Inappropriate study design |
| 4535 | Increased oxygen pulse after lung volume reduction surgery is associated with reduced dynamic hyperinflation | Inappropriate population |
| 4536 | Is improvement in depression in patients attending cardiac rehabilitation with new-onset depressive symptoms determined by patient characteristics? | Inappropriate outcome |
| 4537 | PUFA supplements and type 2 diabetes in the elderly | Inappropriate population |
| 4538 | Blood pressure regulation during cardiac autonomic blockade: effect of fitness | Inappropriate population |
| 4539 | Differences within Elite Female Tennis Players during an Incremental Field Test | Inappropriate study design |
| 4540 | Multimorbidity in Middle-Aged Adults with Cerebral Palsy | Inappropriate study design |
| 4541 | Effect of lean red meat combined with a multicomponent exercise program on muscle and cognitive function in older adults: a 6-month randomized controlled trial | Inappropriate study design |
| 4542 | Energetic adaptation to chronic disease in the elderly | Inappropriate outcome |
| 4543 | Low resonance frequency vibration affects strength of paretic and non-paretic leg differently in patients with stroke | Inappropriate study design |
| 4544 | Participation in activity and risk for incident delirium | Inappropriate study design |
| 4545 | Baroreflex function in endurance- and static exercise-trained men | Inappropriate study design |
| 4546 | Migraine in women | Inappropriate population |
| 4547 | Systemic hemodynamic determinants of blood pressure in women: age, physical activity, and hormone replacement | Inappropriate study design |
| 4548 | Clinical implications of pathophysiologic changes in the midlife hypertensive patient | Inappropriate study design |
| 4549 | Forced single-nostril breathing and cognition | Inappropriate study design |
| 4550 | Afterload reduction therapy in patients following intraatrial baffle operation for transposition of the great arteries | Inappropriate study design |
| 4551 | Neuroimaging Characteristics and Post-Stroke Fatigue within the First 6 Months After Ischemic Strokes | Inappropriate study design |
| 4552 | Cortical excitability changes following grasping exercise augmented with electrical stimulation | Inappropriate study design |
| 4553 | Influence of physical activity during leisure time in patients in the follow-up two years after CABG | Inappropriate study design |
| 4554 | Energy Balance in Huntington's Disease | Inappropriate study design |
| 4555 | Daily kiwifruit consumption did not improve blood pressure and markers of cardiovascular function in men with hypercholesterolemia | Inappropriate study design |
| 4556 | Hemodynamic effects of incremental lung hyperinflation | Inappropriate study design |
| 4557 | Self-body split: issues of identity in physical recovery following a stroke | Inappropriate study design |
| 4558 | Post-stroke bilateral heterotopic ossification: An acute problem with long-lasting consequences | Inappropriate study design |
| 4559 | Warm water immersion in patients with chronic heart failure: a pilot study : Shah immerse: HF | Inappropriate study design |
| 4560 | A treadmill and overground walking program improves walking in persons residing in the community after stroke: a placebo-controlled, randomized trial | Inappropriate population |
| 4561 | Stroking parameters during continuous and intermittent exercise in regional-level competitive swimmers | Inappropriate study design |
| 4562 | Relationship of Aortic Stiffness to Exercise and Ventricular Volumes in Single Ventricles | Inappropriate study design |
| 4563 | Design and test of a Microsoft Kinect-based system for delivering adaptive visual feedback to stroke patients during training of upper limb movement | Inappropriate study design |
| 4564 | Association of physical activity on body composition, cardiometabolic risk factors, and prevalence of cardiovascular disease in the Korean population (from the fifth Korea national health and nutrition examination survey, 2008-2011) | Inappropriate study design |

| | | |
|---|---|---|
| 4565 | Prevalence of smoking in a psychiatric hospital and its relationship with respiratory symptoms and the prevalence of COPD | Inappropriate study design |
| 4566 | Assessment of energy requirements in elderly populations | Inappropriate population |
| 4567 | Clinical feasibility of exercise-based A-V interval optimization for cardiac resynchronization: a pilot study | Inappropriate study design |
| 4568 | Upper motor neuron lesions: their effect on muscle performance and appearance in stroke patients with minor motor impairment | Inappropriate study design |
| 4569 | Important differences between short- and long-term hemodynamic effects of amiodarone in patients with chronic ischemic heart disease at rest and during ischemia-induced left ventricular dysfunction | Inappropriate study design |
| 4570 | Cholinergic stimulation improves autonomic and hemodynamic profile during dynamic exercise in patients with heart failure | Inappropriate study design |
| 4571 | Acute postoperative digitalization of patients with arteriosclerotic heart disease after major surgery. A randomized haemodynamic study and proposal for therapy | Inappropriate study design |
| 4572 | Lung diffusing capacity and exercise in subjects with previous high altitude pulmonary oedema | Inappropriate population |
| 4573 | Quantifying the effects of four weeks of low-volume high-intensity sprint interval training on V̇O2max through assessment of hemodynamics | Inappropriate study design |
| 4574 | The effects of strength training on cognitive performance in elderly women | Inappropriate outcome |
| 4575 | Prospects and possible pitfalls of a preventive Polypill: confessions of a health promotion convert | Inappropriate population |
| 4576 | The intra- and interobserver variability of impedance cardiography in patients at rest and during exercise | Inappropriate outcome |
| 4577 | Comorbidities in the Elderly and Their Possible Influence on Vaccine Response | Inappropriate intervention |
| 4578 | Prevalence and Risk Factors of Atrial Fibrillation in Chinese Elderly: Results from the Chinese Longitudinal Healthy Longevity Survey | Inappropriate study design |
| 4579 | Implementation of preventive strength training in residential geriatric care: a multi-centre study protocol with one year of interventions on multiple levels | Inappropriate study design |
| 4580 | The effects of physical exercise with music on cognitive function of elderly people: Mihama-Kiho project | Inappropriate population |
| 4581 | Exercise tolerance and thermoregulatory responses during cycling in boys and men | Inappropriate population |
| 4582 | Long-term haemodynamic effects of sustained-release trimazosin at rest and during exercise in essential hypertension | Inappropriate study design |
| 4583 | Cardiovascular, metabolic and plasma catecholamine responses to passive and active exercises | Inappropriate study design |
| 4584 | Haemodynamic adaptation during exercise in fontan patients at a long-term follow-up | Inappropriate study design |
| 4585 | A kinematic comparison of alterations to knee and ankle angles from resting measures to active pedaling during a graded exercise protocol | Inappropriate outcome |
| 4586 | The Prevalence of Chronic Conditions and Poor Health Among People With and Without Vision Impairment, Aged ≥65 Years, 2010-2014 | Inappropriate study design |
| 4587 | Oxygen Delivery and Muscle Deoxygenation during Continuous, Long- and Short-Interval Exercise | Inappropriate study design |
| 4588 | Prizidilol in essential hypertension: long-term effects on plasma volume, extracellular fluid volume, and central hemodynamics at rest and during exercise | Inappropriate study design |
| 4589 | Thermoregulatory responses in wheelchair tennis players: a pilot study | Inappropriate study design |
| 4590 | Kinetics of exercise-induced neural activation; interpretive dilemma of altered cerebral perfusion | Inappropriate study design |
| 4591 | Muscular strength and physical function | Inappropriate outcome |
| 4592 | "Reps" Aren't Enough: Augmenting Functional Electrical Stimulation With Behavioral Supports Significantly Reduces Impairment in Moderately Impaired Stroke | Inappropriate outcome |
| 4593 | Disease severity and health-related quality of life across different chronic conditions | Inappropriate population |
| 4594 | Relationship of physical activity and body mass index to the risk of hypertension: a prospective study in Finland | Inappropriate population |
| 4595 | Whole body vibration improves cognition in healthy young adults | Inappropriate population |
| 4596 | Growth hormone (GH)-releasing hormone and GH secretagogues in normal aging: Fountain of Youth or Pool of Tantalus? | Inappropriate population |
| 4597 | The role of rehabilitation psychology in stroke care described through case examples | Inappropriate study design |

| | | |
|---|---|---|
| 4598 | Effectiveness of a combination of cognitive behavioral therapy and task-oriented balance training in reducing the fear of falling in patients with chronic stroke: study protocol for a randomized controlled trial | Inappropriate study design |
| 4599 | Haemodynamic changes in man during immersion in water at different temperatures | Inappropriate outcome |
| 4600 | The cardiac model of rehabilitation for reducing cardiovascular risk factors post transient ischaemic attack and stroke: a randomized controlled trial | Inappropriate population |
| 4601 | The different radionuclide angiography (RNA) responses to upright bicycle ergometer exercise in a population with low probability coronary disease | Inappropriate population |
| 4602 | Predicting the transition from acute to persistent low back pain | Inappropriate population |
| 4603 | A beat-by-beat analysis of cardiovascular responses to dry resting and exercise apnoeas in elite divers | Inappropriate study design |
| 4604 | Leisure-time physical activity over the life course and cognitive functioning in late mid-adult years: a cohort-based investigation | Inappropriate population |
| 4605 | Use of cardiopulmonary exercise testing with hemodynamic monitoring in the prognostic assessment of ambulatory patients with chronic heart failure | Inappropriate intervention |
| 4606 | Modulation of cardiac contractility by muscle metaboreflex following efforts of different intensities in humans | Inappropriate study design |
| 4607 | Haemodynamic responses to physical activity and body posture during everyday life | Inappropriate study design |
| 4608 | Right atrial pressure and ANP release during prolonged exercise in a hot environment | Inappropriate study design |
| 4609 | The effect of group-based exercise on cognitive performance and mood in seniors residing in intermediate care and self-care retirement facilities: a randomised controlled trial | Inappropriate population |
| 4610 | Cardiac responses of young women to conditioning for a 10 kilometer race | Inappropriate study design |
| 4611 | Importance of wash riding in kayaking training and competition | Inappropriate outcome |
| 4612 | Vagus Nerve Stimulation Paired With Upper Limb Rehabilitation After Chronic Stroke | Inappropriate study design |
| 4613 | Eligibility, Enrollment, and Completion of Exercise-Based Cardiac Rehabilitation Following Stroke Rehabilitation: What Are the Barriers? | Inappropriate population |
| 4614 | Assessing the human cardiovascular response to moderate exercise: feature extraction by support vector regression | Inappropriate outcome |
| 4615 | Haemodynamic effects of nifedipine in essential hypertension at rest and during exercise | Inappropriate population |
| 4616 | Learning, retention, and slacking: a model of the dynamics of recovery in robot therapy | Inappropriate population |
| 4617 | Exercise to prevent falls in older adults: an updated systematic review and meta-analysis | Inappropriate population |
| 4618 | Sustained effectiveness of chronic prazosin therapy in severe chronic congestive heart failure | Inappropriate population |
| 4619 | Adjustment of cardiac output to step exercise in heart transplant recipients | Inappropriate population |
| 4620 | Cardiovascular effects of 1 year of progressive and vigorous exercise training in previously sedentary individuals older than 65 years of age | Inappropriate population |
| 4621 | Pharmacokinetics and hemodynamic effects of tiapamil: exercise performance, thallium stress scintigraphy, and radionuclide ventriculography | Inappropriate study design |
| 4622 | Is adiposity associated with objectively measured physical activity and sedentary behaviors in older adults? | Inappropriate population |
| 4623 | Role of hypoxemia for the cardiovascular responses to apnea during exercise | Inappropriate study design |
| 4624 | Long-term haemodynamic effects of enalapril at rest and during exercise in essential hypertension | Inappropriate study design |
| 4625 | Effect of strengthening exercise in addition to task-specific gait training after stroke: a randomised trial | Inappropriate intervention |
| 4626 | Loss of strength contributes more to physical disability after stroke than loss of dexterity | Inappropriate study design |
| 4627 | Energy expenditure, cigarette smoking, and blood pressure level as related to death from specific diseases | Inappropriate study design |
| 4628 | Effect of a robotic seal on the motor activity and sleep patterns of older people with dementia, as measured by wearable technology: A cluster-randomised controlled trial | Inappropriate study design |
| 4629 | Preventing recurrence of thromboembolic events through coordinated treatment in the District of Columbia | Inappropriate population |
| 4630 | Effect of increasing heart rate on finger photoplethysmography fitness index (PPGF) in subjects with implanted cardiac pacemakers | Inappropriate population |

| | | |
|---|---|---|
| 4631 | Muscle fatigability and depressive symptoms in later life | Inappropriate study design |
| 4632 | Continuous wave Doppler measurements of aortic bloodflow during exercise in patients with chronic heart failure | Inappropriate population |
| 4633 | A comparison of regular rehabilitation and regular rehabilitation with supported treadmill ambulation training for acute stroke patients | Inappropriate population |
| 4634 | Skill transfer from symmetric and asymmetric bimanual training using a robotic system to single limb performance | Inappropriate study design |
| 4635 | Comparison of ventricular volumes in normal and post-myocardial infarction subjects | Inappropriate outcome |
| 4636 | Feasibility of Ballistic Strength Training in Subacute Stroke: A Randomized, Controlled, Assessor-Blinded Pilot Study | Inappropriate intervention |
| 4637 | Pulmonary arterial compliance at rest and exercise in normal humans | Inappropriate study design |
| 4638 | Value of carvedilol in congestive heart failure secondary to coronary artery disease | Inappropriate study design |
| 4639 | Frailty and its potential relevance to cardiovascular care | Inappropriate study design |
| 4640 | Cardiac remodeling in response to 1 year of intensive endurance training | Inappropriate population |
| 4641 | Pressure-flow relations in the pulmonary artery during myocardial ischaemia: implications for right ventricular function in coronary disease | Inappropriate population |
| 4642 | Cardiovascular drift is related to reduced maximal oxygen uptake during heat stress | Inappropriate outcome |
| 4643 | Operation Everest II | Inappropriate study design |
| 4644 | The effect of electric stimulation treatment on the functional rehabilitation of acute geriatric patients with stroke--a preliminary study | Inappropriate outcome |
| 4645 | Early versus delayed rehabilitation treatment in hemiplegic patients with ischemic stroke: proprioceptive or cognitive approach? | Inappropriate study design |
| 4646 | Hemodynamic effects of epinephrine: concentration-effect study in humans | Inappropriate study design |
| 4647 | How Older Adults Cope with Cognitive Complexity and Environmental Constraints during Dual-Task Walking: The Role of Executive Function Involvement | Inappropriate study design |
| 4648 | Impact Of Peak Oxygen Pulse On Patients With Chronic Obstructive Pulmonary Disease | Inappropriate study design |
| 4649 | Circadian blood pressure and systemic haemodynamics during 42 days of 6 degrees head-down tilt | Inappropriate study design |
| 4650 | Effects of oral lacidipine on cardiopulmonary function at rest and during exercise in normal subjects | Inappropriate population |
| 4651 | Stroke volume response to progressive exercise in athletes engaged in different types of training | Inappropriate population |
| 4652 | Extremely low frequency electromagnetic field reduces oxidative stress during the rehabilitation of post-acute stroke patients | Inappropriate population |
| 4653 | Hemodynamic differences between supine and upright exercise in patients with congestive heart failure | Inappropriate study design |
| 4654 | Defeating Urinary Incontinence with Exercise Training: Results of a Pilot Study in Frail Older Women | Inappropriate study design |
| 4655 | Effects of different sprint cycling bouts on left ventricular function in top cyclists | Inappropriate population |
| 4656 | Correlation between self-perceived burden and self-management behavior in elderly stroke survivors: A longitudinal observational study | Inappropriate study design |
| 4657 | Physiologic effects of electrical stimulation leg cycle exercise training in spinal cord injured persons | Inappropriate study design |
| 4658 | Insulin resistance and executive dysfunction in older persons | Inappropriate population |
| 4659 | Hemodynamic consequences of long-term beta-blocker therapy: a 5-year follow-up study of atenolol | Inappropriate population |
| 4660 | Impact of enhanced secondary prevention on health behaviour in patients following minor stroke and transient ischaemic attack: a randomized controlled trial | Inappropriate study design |
| 4661 | High cytochrome c oxidase expression links to severe skeletal energy failure by (31)P-MRS spectroscopy in mitochondrial encephalomyopathy, lactic acidosis, and stroke-like episodes | Inappropriate study design |
| 4662 | Functional walk tests in individuals with stroke: relation to perceived exertion and myocardial exertion | Inappropriate study design |
| 4663 | Valve replacement for chronic mitral insufficiency; long-term follow up using technetium pertechnetate scintigraphy | Inappropriate population |
| 4664 | The hemodynamics of calcium-channel antagonists in hypertension: vascular and myocardial responses | Inappropriate population |

| | | |
|---|---|---|
| 4665 | Respiratory muscle training in athletes with cervical spinal cord injury: effects on cardiopulmonary function and exercise capacity | Inappropriate intervention |
| 4666 | Exercise and extracorporeal blood cooling during hemodialysis | Inappropriate study design |
| 4667 | Greater fluid loss does not fully explain the divergent hemodynamic balance mediating postexercise hypotension in endurance-trained men | Inappropriate study design |
| 4668 | Acute effects of firefighting on cardiac performance | Inappropriate study design |
| 4669 | Influence of individual and combined healthy behaviours on successful aging | Inappropriate outcome |
| 4670 | Cognitive decline and cardiometabolic risk among Hispanic and non-Hispanic white adults in the San Luis Valley Health and Aging Study | Inappropriate population |
| 4671 | Yoga leads to multiple physical improvements after stroke, a pilot study | Inappropriate study design |
| 4672 | Vastus lateralis oxygenation during prolonged cycling in healthy males | Inappropriate study design |
| 4673 | Effects of multi-domain interventions in (pre)frail elderly on frailty, functional, and cognitive status: a systematic review | Inappropriate study design |
| 4674 | Perceptions and attitudes toward risk and personal responsibility in the context of medical fitness to drive | Inappropriate study design |
| 4675 | Elastic, viscous, and mass load effects on poststroke muscle recruitment and co-contraction during reaching: a pilot study | Inappropriate outcome |
| 4676 | Comparison of functional autonomy with associated sociodemographic factors, lifestyle, chronic diseases (CD) and neuropsychiatric factors in elderly patients with or without the metabolic syndrome (MS) | Inappropriate population |
| 4677 | Post-stroke fatigue: a deficit in corticomotor excitability? | Inappropriate population |
| 4678 | Ventricular-Arterial Coupling and Exercise-Induced Pulmonary Hypertension During Low-Level Exercise in Heart Failure With Preserved or Reduced Ejection Fraction | Inappropriate study design |
| 4679 | Efficacy and tolerance of a neurological restoration program in stroke patients | Inappropriate outcome |
| 4680 | Calcium channel blocker and isosorbide 5-mononitrate in the management of chronic cardiac failure | Inappropriate intervention |
| 4681 | Hemodynamic observations on a yogic breathing technique claimed to help eliminate and prevent heart attacks: a pilot study | Inappropriate study design |
| 4682 | Sleep loss affects vigilance: effects of chronic insomnia and sleep therapy | Inappropriate population |
| 4683 | Whole-body vibration has no effect on neuromotor function and falls in chronic stroke | Inappropriate study design |
| 4684 | Evaluation of left ventricular function in chronic pulmonary disease by exercise gated equilibrium radionuclide angiography | Inappropriate population |
| 4685 | Haemodynamic effects of atenolol, labetalol, pindolol and captopril: a comparison in hypertensive patients with special reference to changes in limb blood flow, heart rate and left ventricular function | Inappropriate study design |
| 4686 | Obesity and the metabolic syndrome | Inappropriate population |
| 4687 | Influence of cardiac failure on atrial natriuretic peptide responses in patients undergoing vascular surgery | Inappropriate population |
| 4688 | Physical activity and vascular disease in a prospective cohort study of older men: The Health In Men Study (HIMS) | Inappropriate population |
| 4689 | Home-based Computer Assisted Arm Rehabilitation (hCAAR) robotic device for upper limb exercise after stroke: results of a feasibility study in home setting | Inappropriate population |
| 4690 | Reliability and Validity of the Resistiveness to Care Scale Among Cognitively Impaired Older Adults | Inappropriate study design |
| 4691 | Hemodynamic changes during dipyridamole stress in patients with aortic stenosis | Inappropriate study design |
| 4692 | How valid is the determination of hematocrit values to detect blood manipulations? | Inappropriate population |
| 4693 | Lifestyle in progression from hypertensive disorders of pregnancy to chronic hypertension in Nurses' Health Study II: observational cohort study | Inappropriate study design |
| 4694 | Left ventricular contractile performance in the early stage of hypertension in humans | Inappropriate study design |
| 4695 | Cardiovascular response to trigeminal nerve stimulation at rest and during exercise in humans: does sex matter? | Inappropriate study design |
| 4696 | Lower body positive and negative pressure alter thermal and hemodynamic responses after exercise | Inappropriate study design |
| 4697 | Is sodium restriction effective treatment of borderline and mild essential hypertension? A long-term haemodynamic study at rest and during exercise | Inappropriate study design |
| 4698 | Hemodynamic and clinical significance of the pulmonary vascular response to long-term captopril therapy in patients with severe chronic heart failure | Inappropriate population |
| 4699 | Neurobehavioral improvement after brief rehabilitation in patients with chronic obstructive pulmonary disease | Inappropriate study design |

| | | |
|---|---|---|
| 4700 | Monitoring of cardiac output during exercise in coronary patients: a Doppler study | Inappropriate intervention |
| 4701 | Long-term effects on central haemodynamics and body fluid volumes of ketanserin in essential hypertension studies at rest and during dynamic exercise | Inappropriate study design |
| 4702 | Risk Factors for Bone Loss in Patients Having Clinical Conditions That Restrict Physical Activity | Inappropriate population |
| 4703 | Hemodynamic changes following correction of severe aortic stenosis using the Cutter-Smeloff prosthesis | Inappropriate study design |
| 4704 | Outcome quality of in-patient cardiac rehabilitation in elderly patients--identification of relevant parameters | Inappropriate population |
| 4705 | Effects of diltiazem and long-term beta 1-adrenergic blockade on hemodynamics and blood flow during exercise in patients with stable angina pectoris | Inappropriate study design |
| 4706 | Echocardiographic evaluation of space shuttle crewmembers | Inappropriate outcome |
| 4707 | In-hospital initiation of secondary stroke prevention therapies yields high rates of adherence at follow-up | Inappropriate population |
| 4708 | Cardiovascular responses to military antishock trouser inflation during standing arm exercise | Inappropriate study design |
| 4709 | Comparative effects of metoprolol and celiprolol on cardiac hemodynamics and left ventricular volume at rest and during exercise-induced angina | Inappropriate outcome |
| 4710 | Cognitive Dysfunction and Malnutrition Are Independent Predictor of Dysphagia in Patients with Acute Exacerbation of Congestive Heart Failure | Inappropriate study design |
| 4711 | Visual impairment, uncorrected refractive error, and accelerometer-defined physical activity in the United States | Inappropriate outcome |
| 4712 | Healthy living is the best revenge: findings from the European Prospective Investigation Into Cancer and Nutrition-Potsdam study | Inappropriate study design |
| 4713 | Haemodynamic long-term effects of metoprolol at rest and during exercise in essential hypertension | Inappropriate study design |
| 4714 | Randomized trial of a robotic assistive device for the upper extremity during early inpatient stroke rehabilitation | Inappropriate population |
| 4715 | Neurophysiological signatures of hand motor response to dual-transcranial direct current stimulation in subacute stroke: a TMS and MEG study | Inappropriate study design |
| 4716 | Cardiovascular strain impairs prolonged self-paced exercise in the heat | Inappropriate population |
| 4717 | Effects of acceleration in the Gz axis on human cardiopulmonary responses to exercise | Inappropriate population |
| 4718 | The development of an adaptive upper-limb stroke rehabilitation robotic system | Inappropriate study design |
| 4719 | Impaired skeletal muscle vasodilation during exercise in heart failure with preserved ejection fraction | Inappropriate study design |
| 4720 | Risk factors for ischaemic and intracerebral haemorrhagic stroke in 22 countries (the INTERSTROKE study): a case-control study | Inappropriate outcome |
| 4721 | Acute physiological effects of whole body vibration (WBV) on central hemodynamics, muscle oxygenation and oxygen consumption in individuals with chronic spinal cord injury | Inappropriate study design |
| 4722 | Diastolic heart failure in the elderly | Inappropriate study design |
| 4723 | Perceptions of Heart-Healthy Behaviors among African American Adults: A Mixed Methods Study | Inappropriate population |
| 4724 | Dog Ownership and Survival After a Major Cardiovascular Event: A Register-Based Prospective Study | Inappropriate population |
| 4725 | An overview of diabetes mellitus in older persons | Inappropriate study design |
| 4726 | Examining the Disability Model From the International Classification of Functioning, Disability, and Health Using a Large Data Set of Community-Dwelling Malaysian Older Adults | Inappropriate study design |
| 4727 | Left ventricular diastolic function in trained male weight lifters at rest and during isometric exercise | Inappropriate study design |
| 4728 | Diabetes mellitus in older persons | Inappropriate population |
| 4729 | Acceleration with exercise during head-down bed rest preserves upright exercise responses | Inappropriate intervention |
| 4730 | Influence of nebivolol on the cardiovascular hemodynamics during postural changes and isometric exercise | Inappropriate population |
| 4731 | Habitual alcohol consumption is associated with lower cardiovascular stress responses--a novel explanation for the known cardiovascular benefits of alcohol? | Inappropriate study design |
| 4732 | Right heart size and function significantly correlate in patients with pulmonary arterial hypertension - a cross-sectional study | Inappropriate study design |

| | | |
|---|---|---|
| 4733 | Lower body negative pressure exercise plus brief postexercise lower body negative pressure improve post-bed rest orthostatic tolerance | Inappropriate study design |
| 4734 | Physiological and cardiovascular changes associated with deep water running in the young. Possible implications for the elderly | Inappropriate study design |
| 4735 | Reliability of treadmill exercise testing in older patients with chronic hemiparetic stroke | Inappropriate population |
| 4736 | Modifiable disease risk, readiness to change, and psychosocial functioning improve with integrative medicine immersion model | Inappropriate study design |
| 4737 | Modelflow estimates of cardiac output compared with Doppler ultrasound during acute changes in vascular resistance in women | Inappropriate study design |
| 4738 | Effects of implantable peroneal nerve stimulation on gait quality, energy expenditure, participation and user satisfaction in patients with post-stroke drop foot using an ankle-foot orthosis | Inappropriate study design |
| 4739 | Rate response and cardiac resynchronisation therapy in chronic heart failure: higher cardiac output does not acutely improve exercise performance: a pilot trial | Inappropriate outcome |
| 4740 | Cardiovascular and diabetes burden attributable to physical inactivity in Mexico | Inappropriate study design |
| 4741 | Walking speed as a predictor of community mobility and quality of life after stroke | Inappropriate population |
| 4742 | Cardiac outputs of post-myocardial infarction patients during swimming and cyclic | Inappropriate study design |
| 4743 | Responses to dynamic leg exercise in man as influenced by changes in muscle perfusion pressure | Inappropriate population |
| 4744 | L-arginine supplementation improves exercise capacity after a heart transplant | Inappropriate intervention |
| 4745 | Development of robots for rehabilitation therapy: the Palo Alto VA/Stanford experience | Inappropriate intervention |
| 4746 | Forebrain regions associated with postexercise differences in autonomic and cardiovascular function during baroreceptor unloading | Inappropriate population |
| 4747 | Progressive shoulder abduction loading is a crucial element of arm rehabilitation in chronic stroke | Inappropriate study design |
| 4748 | Validation of a novel activity monitor in impaired, slow-walking, crutch-supported patients | Inappropriate outcome |
| 4749 | Effect of exercise intensity on relationship between VO2max and cardiac output | Inappropriate population |
| 4750 | Association and interaction effects of Alzheimer's disease-associated genes and lifestyle on cognitive aging in older adults in a Taiwanese population | Inappropriate population |
| 4751 | Differential hemodynamic effects of exercise and volume expansion in people with and without heart failure | Inappropriate population |
| 4752 | Recovery in the Severely Impaired Arm Post-Stroke After Mirror Therapy: A Randomized Controlled Study | Inappropriate study design |
| 4753 | Prediction of long-term survival by preoperative exercise testing in patients with depressed ejection fraction undergoing myocardial revascularization | Inappropriate study design |
| 4754 | Telomere length is associated with decline in grip strength in older persons aged 65 years and over | Inappropriate population |
| 4755 | Respiration-dependent ventricular pacing compared with fixed ventricular and atrial-ventricular synchronous pacing: aerobic and hemodynamic variables | Inappropriate study design |
| 4756 | Left ventricular response in healthy young men during heavy-intensity weight-lifting exercise | Inappropriate study design |
| 4757 | The validity and reliability of the Incremental Shuttle Walk Test and Six-minute Walk Test compared to an Incremental Cycle Test for people who have had a mild-to-moderate stroke | Inappropriate study design |
| 4758 | Effects of head-down bed rest on the executive functions and emotional response | Inappropriate study design |
| 4759 | Erectile dysfunction is not independently associated with cardiovascular death: data from the vitamins and lifestyle (VITAL) study | Inappropriate population |
| 4760 | PROFILE PD: profile of function and impairment level experience with Parkinson disease--clinimetric properties of a rating scale for physical therapist practice | Inappropriate study design |
| 4761 | Dynamics of cardiac output and systolic time intervals in supine and upright exercise | Inappropriate study design |
| 4762 | Long-term disability after first-ever stroke and related prognostic factors in the Perth Community Stroke Study, 1989-1990 | Inappropriate population |
| 4763 | Assessment of circulatory adjustments during underwater apnoea in elite divers by means of a portable device | Inappropriate population |
| 4764 | Effect of Lower Body Negative Pressure on Phase I Cardiovascular Responses at Exercise Onset | Inappropriate study design |
| 4765 | Hemodynamic responses to brief hyperoxia in healthy and in mild hypertensive human subjects in rest and during dynamic exercise | Inappropriate population |

| | | |
|---|---|---|
| 4766 | Cardiovascular and Lifestyle Risk Factors and Cognitive Function in Patients With Stable Coronary Heart Disease | Inappropriate study design |
| 4767 | Left ventricular function in response to the transition from aerobic to anaerobic metabolism | Inappropriate intervention |
| 4768 | Feasibility of robot-based perturbed-balance training during treadmill walking in a high-functioning chronic stroke subject: a case-control study | Inappropriate population |
| 4769 | Oxygen pulse is not predictive of stroke volume in heart failure | Inappropriate study design |
| 4770 | Determination of stroke volume and cardiac output during exercise: comparison of two-dimensional and Doppler echocardiography, Fick oximetry, and thermodilution | Inappropriate population |
| 4771 | AMOBES (Active Mobility Very Early After Stroke): A Randomized Controlled Trial | Inappropriate population |
| 4772 | Effect of beta-adrenergic blockade on thermoregulation during exercise | Inappropriate study design |
| 4773 | Chronic medical conditions and metabolic syndrome as risk factors for incidence of major depressive disorder: A longitudinal study based on 4.7 million adults in South Korea | Inappropriate study design |
| 4774 | Physiologic responses of paraplegics and quadriplegics to passive and active leg cycle ergometry | Inappropriate intervention |
| 4775 | Functional electrical therapy for hemiparesis alleviates disability and enhances neuroplasticity | Inappropriate study design |
| 4776 | Mesencephalic corticospinal atrophy predicts baseline deficit but not response to unilateral or bilateral arm training in chronic stroke | Inappropriate study design |
| 4777 | Does total volume of physical activity matter more than pattern for onset of CVD? A prospective cohort study of older British men | Inappropriate study design |
| 4778 | Atrial fibrillation is associated with a lower exercise capacity in male chronic heart failure patients | Inappropriate study design |
| 4779 | Cardiopulmonary exercise response in patients with left ventricular dysfunction or heart failure: a noninvasive study by gas exchange and impedance cardiography monitoring | Inappropriate outcome |
| 4780 | Translation and Validation of a Chinese Version of the Stroke Self-Efficacy Questionnaire in Community-Dwelling Stroke Survivors | Inappropriate study design |
| 4781 | Utilization of phase II cardiac rehabilitation in Veterans Administration patients | Inappropriate population |
| 4782 | An analysis of the physiological strain of submaximal exercise in patients with chronic obstructive bronchitis | Inappropriate outcome |
| 4783 | Difference in human cardiovascular response between upright and supine recovery from upright cycle exercise | Inappropriate intervention |
| 4784 | Cereal, fruit, and vegetable fiber intake and the risk of cardiovascular disease in elderly individuals | Inappropriate population |
| 4785 | Systemic and regional hemodynamic response to activation of the exercise pressor reflex in patients with peripheral artery disease | Inappropriate study design |
| 4786 | Prevalence of frailty and its association with sociodemographic and clinical characteristics, and resource utilization in a population of Singaporean older adults | Inappropriate study design |
| 4787 | Haemodynamic adjustments to mental stress in normotensives and subjects with mildly elevated blood pressure | Inappropriate population |
| 4788 | Deep flexion activity training in a patient with stroke using task-oriented exercise: a case report | Inappropriate study design |
| 4789 | A rule-based, dose-finding design for use in stroke rehabilitation research: methodological development | Inappropriate study design |
| 4790 | Comparison of Three Popular Exercise Modalities on V˙O2max in Overweight and Obese | Inappropriate study design |
| 4791 | The Effects of a Cycling Warm-up Including High-Intensity Heavy-Resistance Conditioning Contractions on Subsequent 4-km Time Trial Performance | Inappropriate study design |
| 4792 | Predictors of chewing ability among community-residing older adults in Korea | Inappropriate study design |
| 4793 | Gamma-glutamyltransferase and cardiovascular mortality in Korean adults: A cohort study | Inappropriate population |
| 4794 | Effect of levodopa in combination with physiotherapy on functional motor recovery after stroke: a prospective, randomised, double-blind study | Inappropriate study design |
| 4795 | Pulmonary thallium uptake: correlation with systolic and diastolic left ventricular function at rest and during exercise | Inappropriate study design |
| 4796 | Dynamic cerebral autoregulation is impaired during submaximal isometric handgrip in patients with heart failure | Inappropriate population |
| 4797 | Exercise training improves baroreflex sensitivity in type 2 diabetes | Inappropriate population |

| | | |
|---|---|---|
| 4798 | Day healthcare services for family caregivers of older people with stroke: needs and satisfaction | Inappropriate study design |
| 4799 | Chronic medical conditions and risk of fall injury events at home in older adults | Inappropriate population |
| 4800 | Effects of different training amplitudes on heart rate and heart rate variability in young rowers | Inappropriate study design |
| 4801 | The association between neighborhood greenness and cardiovascular disease: an observational study | Inappropriate population |
| 4802 | Psychomotor speed and functional brain MRI 2 years after completing a physical activity treatment | Inappropriate population |
| 4803 | Acute antianginal hemodynamic effects of nicardipine in coronary artery disease | Inappropriate study design |
| 4804 | Cardiovascular responses at the onset of static exercise in patients with dual-chamber pacemakers | Inappropriate study design |
| 4805 | Doppler echocardiographic assessment of the pressure gradient during bicycle ergometry in hypertrophic cardiomyopathy | Inappropriate population |
| 4806 | Nonthermoregulatory control of cutaneous vascular conductance and sweating during recovery from dynamic exercise in women | Inappropriate population |
| 4807 | Inter-individual variability in right ventricle adaptation after an endurance race | Inappropriate outcome |
| 4808 | Afterload reduction by nifedipine--the acute haemodynamic response to exercise in hypertensive subjects | Inappropriate population |
| 4809 | The influence of recovery posture on post-exercise hypotension in normotensive men | Inappropriate population |
| 4810 | Effects of detraining on cardiovascular responses to exercise: role of blood volume | Inappropriate study design |
| 4811 | Causes and hemodynamic findings in chronic severe pulmonary regurgitation | Inappropriate outcome |
| 4812 | Hypohydration effects on thermoregulation during moderate exercise in the cold | Inappropriate study design |
| 4813 | Stochastic resonance stimulation for upper limb rehabilitation poststroke | Inappropriate population |
| 4814 | Maximal cardiac output determines 6 minutes walking distance in pulmonary hypertension | Inappropriate population |
| 4815 | Nifedipine pharmacodynamics and pharmacokinetics in treatment of congestive heart failure | Inappropriate study design |
| 4816 | Automated impedance cardiography for detecting ischemic left ventricular dysfunction during exercise testing | Inappropriate study design |
| 4817 | Myocardial adaption to HI(R)T in previously untrained men with a randomized, longitudinal cardiac MR imaging study (Physical adaptions in Untrained on Strength and Heart trial, PUSH-trial) | Inappropriate study design |
| 4818 | Progressive Resistance Training Improves Bradykinesia, Motor Symptoms and Functional Performance in Patients with Parkinson's Disease | Inappropriate study design |
| 4819 | Management of the older person with atrial fibrillation | Inappropriate population |
| 4820 | Effects of a new vasodilator, nicorandil, on exercise-induced impairment of left ventricular function in patients with old myocardial infarction | Inappropriate study design |
| 4821 | Acute effects of oral propranolol on hemodynamic responses to upright exercise | Inappropriate study design |
| 4822 | Trunk muscle exercises as a means of improving postural stability in people with Parkinson's disease: a protocol for a randomised controlled trial | Inappropriate population |
| 4823 | Stroke increases ischemia-related decreases in motor unit discharge rates | Inappropriate study design |
| 4824 | myMoves Program: Feasibility and Acceptability Study of a Remotely Delivered Self-Management Program for Increasing Physical Activity Among Adults With Acquired Brain Injury Living in the Community | Inappropriate study design |
| 4825 | Cost-effectiveness analysis of offering free leisure centre memberships to physically inactive members of the public receiving state benefits: a case study | Inappropriate study design |
| 4826 | Lifestyle factors and risk of cerebral infarction | Inappropriate study design |
| 4827 | Cogniphobia in Mild Traumatic Brain Injury | Inappropriate study design |
| 4828 | Livelihood vs. life: the occupational well-being of women musician survivors of breast cancer | Inappropriate study design |
| 4829 | Self-management support interventions for persons with chronic disease: an evidence-based analysis | Inappropriate study design |
| 4830 | High frequency of migraine-only patients negative for the 3243 A>G tRNALeu mtDNA mutation in two MELAS families | Inappropriate population |
| 4831 | Vitamin D and cognitive function: The Tromsø Study | Inappropriate intervention |
| 4832 | Lay beliefs about risk: relation to risk behaviors and to probabilistic risk perceptions | Inappropriate study design |
| 4833 | The hemodynamics of the aging cardiovascular system | Inappropriate study design |

| | | |
|---|---|---|
| 4834 | Beneficial effects of the calcium antagonist PN 200-110 in patients with congestive heart failure | Inappropriate study design |
| 4835 | Management of newly detected atrial fibrillation: a clinical practice guideline from the American Academy of Family Physicians and the American College of Physicians | Inappropriate population |
| 4836 | Pain management in the elderly | Inappropriate study design |
| 4837 | Post-walking exercise hypotension in patients with intermittent claudication | Inappropriate study design |
| 4838 | Fit elderly men can also stand: orthostatic tolerance and autonomic cardiovascular control in elderly endurance athletes | Inappropriate study design |
| 4839 | Evaluating the effects of an exercise program (Staying UpRight) for older adults in long-term care on rates of falls: study protocol for a randomised controlled trial | Inappropriate outcome |
| 4840 | The role of the community nurse in psoriatic comorbidities interventions | Inappropriate study design |
| 4841 | Global and regional effects of potentially modifiable risk factors associated with acute stroke in 32 countries (INTERSTROKE): a case-control study | Inappropriate study design |
| 4842 | Caregiver-mediated exercises with e-health support for early supported discharge after stroke (CARE4STROKE): A randomized controlled trial | Inappropriate population |
| 4843 | Self-Reported Fatigue Predicts Incident Stroke in a General Population: EPIC-Norfolk Prospective Population-Based Study | Inappropriate population |
| 4844 | The impact of non-compliance with the therapeutic regimen on the development of stroke among hypertensive men and women in Gaza, Palestine | Inappropriate study design |
| 4845 | Effects of oxyfedrine on left ventricular function in patients several months after myocardial infarction | Inappropriate population |
| 4846 | Protective effect of fruits and vegetables on development of stroke in men | Inappropriate population |
| 4847 | Rest and exercise cardiac output and diffusing capacity assessed by a single slow exhalation of methane, acetylene, and carbon monoxide | Inappropriate outcome |
| 4848 | An open-circuit method for determining lung diffusing capacity during exercise: comparison to rebreathe | Inappropriate study design |
| 4849 | The dimensionality of fatigue in Parkinson's disease | Inappropriate study design |
| 4850 | Plasma klotho and mortality risk in older community-dwelling adults | Inappropriate intervention |
| 4851 | The metabolic syndrome and insulin resistance: relationship to haemostatic and inflammatory markers in older non-diabetic men | Inappropriate study design |
| 4852 | Low body weight and weight loss in the aged | Inappropriate population |
| 4853 | Tracking motor improvement at the subtask level during robot-aided neurorehabilitation of stroke patients | Inappropriate study design |
| 4854 | Relation of consumption of vitamin E, vitamin C, and carotenoids to risk for stroke among men in the United States | Inappropriate outcome |
| 4855 | Common geriatric conditions and disabilities in older persons with chronic obstructive pulmonary disease | Inappropriate study design |
| 4856 | Uremic myopathy limits aerobic capacity in hemodialysis patients | Inappropriate population |
| 4857 | Cognitive impairment, chronic disease burden, and functional disability: a population study of older Italians | Inappropriate study design |
| 4858 | A randomized controlled trial on Stroke telerehabilitation: The effects on falls self-efficacy and satisfaction with care | Inappropriate study design |
| 4859 | Insulin resistance and the development of cardiovascular disease in a Japanese community: the Hisayama study | Inappropriate study design |
| 4860 | Differential effects of beta-blockers in patients with heart failure: A prospective, randomized, double-blind comparison of the long-term effects of metoprolol versus carvedilol | Inappropriate intervention |
| 4861 | Disproportionate exercise load and remodeling of the athlete's right ventricle | Inappropriate population |
| 4862 | Exercise in heart failure: should aqua therapy and swimming be allowed? | Inappropriate population |
| 4863 | Supervised exercise training improves exercise cardiovascular function in idiopathic pulmonary fibrosis | Inappropriate population |
| 4864 | Morning or evening activity improves neuropsychological performance and subjective sleep quality in older adults | Inappropriate population |
| 4865 | Effects of brain natriuretic peptide on exercise hemodynamics and neurohormones in isolated diastolic heart failure | Inappropriate study design |
| 4866 | Muscle mass gain is positively associated with functional recovery in patients with sarcopenia after stroke | Inappropriate study design |
| 4867 | Effect of wearing clothes on oxygen uptake and ratings of perceived exertion while swimming | Inappropriate study design |
| 4868 | Associations of adiponectin with metabolic and vascular risk parameters in the British Regional Heart Study reveal stronger links to insulin resistance-related than to coronory heart disease risk-related parameters | Inappropriate outcome |

| | | |
|---|---|---|
| 4869 | Should obesity be blamed for the high prevalence rates of hypertension in black South African women? | Inappropriate population |
| 4870 | A comparison of the acute and long-term hemodynamic effects of ventricular inhibited and atrial synchronous ventricular inhibited pacing | Inappropriate population |
| 4871 | Effectiveness of Respiratory Muscle Training for Pulmonary Function and Walking Ability in Patients with Stroke: A Systematic Review with Meta-Analysis | Inappropriate study design |
| 4872 | Haemodynamic responses following intermittent supramaximal exercise in athletes | Inappropriate outcome |
| 4873 | Reduction in LBNP tolerance following prolonged endurance exercise training | Inappropriate population |
| 4874 | Effects of atenolol on left ventricular hypertrophy and early left ventricular function in essential hypertension | Inappropriate population |
| 4875 | A path model of chronic stress, the metabolic syndrome, and coronary heart disease | Inappropriate study design |
| 4876 | O2 Pulse Patterns in Male Master Athletes with Normal and Abnormal Exercise Tests | Inappropriate population |
| 4877 | H(2)FPEF Score for the Prediction of Exercise Intolerance and Abnormal Hemodynamics in Japanese - Evaluation by Exercise Stress Echocardiography Combined With Cardiopulmonary Exercise Testing | Inappropriate outcome |
| 4878 | Objective Assessment of Upper-Limb Mobility for Poststroke Rehabilitation | Inappropriate population |
| 4879 | Cardiovascular responses to isometric exercise in patients with mitral stenosis. Comparison with normal subjects and patients with depressed ejection fraction | Inappropriate study design |
| 4880 | Cardiovascular and ventilatory responses during formalized T'ai Chi Chuan exercise | Inappropriate outcome |
| 4881 | Impaired Pulmonary Diffusion in Heart Failure With Preserved Ejection Fraction | Inappropriate study design |
| 4882 | Cardiovascular response of trained and untrained old men to mental challenge | Inappropriate population |
| 4883 | Self-rated health over the first five years after stroke | Inappropriate population |
| 4884 | Cardiovascular responses to arm cranking and FNS-induced leg exercise in paraplegics | Inappropriate population |
| 4885 | The healthy diver: A cross-sectional survey to evaluate the health status of recreational scuba diver members of Divers Alert Network (DAN) | Inappropriate study design |
| 4886 | Plasma catecholamines and hyperglycaemia influence thermoregulation in man during prolonged exercise in the heat | Inappropriate population |
| 4887 | Mumbai stroke registry (2005-2006)--surveillance using WHO steps stroke instrument--challenges and opportunities | Inappropriate population |
| 4888 | Facial cooling does not benefit cardiac dynamics during recovery from exercise hyperthermia | Inappropriate study design |
| 4889 | Post-operative myocardial dysfunction does not affect the physiological response to early mobilization after coronary artery bypass grafting | Inappropriate population |
| 4890 | Postmenopausal hormone therapy and risk of stroke: A pooled analysis of data from population-based cohort studies | Inappropriate study design |
| 4891 | Exercise training-induced alterations of cardiac morphology | Inappropriate study design |
| 4892 | Hemodynamic abnormalities during muscle metaboreflex activation in patients with type 2 diabetes mellitus | Inappropriate outcome |
| 4893 | Right Ventricular-Arterial Uncoupling During Exercise in Heart Failure With Preserved Ejection Fraction: Role of Pulmonary Vascular Dysfunction | Inappropriate outcome |
| 4894 | Inverse association of dietary fat with development of ischemic stroke in men | Inappropriate study design |
| 4895 | Development of a 3D, networked multi-user virtual reality environment for home therapy after stroke | Inappropriate study design |
| 4896 | Genetic risk, incident stroke, and the benefits of adhering to a healthy lifestyle: cohort study of 306 473 UK Biobank participants | Inappropriate outcome |
| 4897 | Relevance of water gymnastics in rehabilitation programs in patients with chronic heart failure or coronary artery disease with normal left ventricular function | Inappropriate population |
| 4898 | Response to upright exercise after cardiac transplantation | Inappropriate population |
| 4899 | Effects of self-contained breathing apparatus on ventricular function during strenuous exercise | Inappropriate intervention |
| 4900 | Bone marrow cell transplantation improves cardiac, autonomic, and functional indexes in acute anterior myocardial infarction patients (Cardiac Study) | Inappropriate study design |
| 4901 | Granulocyte Colony Stimulating Factor and Physiotherapy after Stroke: Results of a Feasibility Randomised Controlled Trial: Stem Cell Trial of Recovery EnhanceMent after Stroke-3 (STEMS-3 ISRCTN16714730) | Inappropriate study design |
| 4902 | The effect of an anti-G suit on cardiovascular responses to exercise in persons with paraplegia | Inappropriate study design |

| | | |
|---|---|---|
| 4903 | Strong linear relationship between heart rate and mean pulmonary artery pressure in exercising patients with severe precapillary pulmonary hypertension | Inappropriate study design |
| 4904 | Relationships between functional fitness and cognitive impairment in Chinese community-dwelling older adults: a cross-sectional study | Inappropriate population |
| 4905 | Physical Activity and Lifetime Risk of Cardiovascular Disease and Cancer | Inappropriate study design |
| 4906 | The influence of depression on risk development of acute cardiovascular diseases in the female population aged 25-64 in Russia | Inappropriate study design |
| 4907 | Stroke follow-up in primary care: a prospective cohort study on guideline adherence | Inappropriate study design |
| 4908 | Whole body bioimpedance monitoring for outpatient chronic heart failure follow up | Inappropriate study design |
| 4909 | Rehabilitation of reaching after stroke: task-related training versus progressive resistive exercise | Inappropriate study design |
| 4910 | Working capacity and cardiopulmonary function after extensive lung resections | Inappropriate study design |
| 4911 | Brain Mechanisms of Anticipated Painful Movements and Their Modulation by Manual Therapy in Chronic Low Back Pain | Inappropriate study design |
| 4912 | Cardiovascular reactivity to physical and psychologic stress during long-term treatment with nitrendipine in essential hypertension | Inappropriate population |
| 4913 | Older Aboriginal Australians' Health Concerns and Preferences for Healthy Ageing Programs | Inappropriate study design |
| 4914 | Gender differences in autonomic functions associated with blood pressure regulation | Inappropriate outcome |
| 4915 | Case Study: Physical Capacity and Nutritional Status Before and After a Single-Handed Yacht Race | Inappropriate study design |
| 4916 | Cardiopulmonary blood volumes at rest and during muscular exercise measured by 113mIn radiocardiography | Inappropriate study design |
| 4917 | A randomized pilot study of aortic waveform guided therapy in chronic heart failure | Inappropriate study design |
| 4918 | Performance and blood lactate on Gjessing and Concept II rowing ergometers | Inappropriate study design |
| 4919 | Cardiovascular reflexes during isometric exercise--role of muscle mass and gravitational stress | Inappropriate population |
| 4920 | Ventilatory response at the onset of voluntary exercise and passive movement in endurance runners | Inappropriate study design |
| 4921 | A Hybrid Robotic System for Arm Training of Stroke Survivors: Concept and First Evaluation | Inappropriate population |
| 4922 | A haemodynamic study of the effects of combined slow-calcium channel blockade (nisoldipine) and beta-blockade (metoprolol) in coronary heart disease | Inappropriate outcome |
| 4923 | Impact of atrial fibrillation on exercise capacity in heart failure with preserved ejection fraction: a RELAX trial ancillary study | Inappropriate study design |
| 4924 | Physical activity and cause-specific mortality: the Rotterdam Study | Inappropriate population |
| 4925 | Beneficial effects of intravenous and oral carvedilol treatment in acute myocardial infarction. A placebo-controlled, randomized trial | Inappropriate study design |
| 4926 | Role of skeletal muscles impairment and brain oxygenation in limiting oxidative metabolism during exercise after bed rest | Inappropriate population |
| 4927 | Prognostic value of an increase in post-exercise ankle-brachial index | Inappropriate study design |
| 4928 | Usefulness of noninvasive Doppler measurement of ascending aortic blood velocity and acceleration in detecting impairment of the left ventricular functional response to exercise three weeks after acute myocardial infarction | Inappropriate study design |
| 4929 | Percutaneous coronary angioplasty compared with exercise training in patients with stable coronary artery disease: a randomized trial | Inappropriate study design |
| 4930 | Patterns and predictors of sleep quality before, during, and after hospitalization in older adults | Inappropriate population |
| 4931 | Hemodynamic basis of exercise limitation in patients with heart failure and normal ejection fraction | Inappropriate study design |
| 4932 | Prevalence and risk factors of arthritis in a middle-aged and older Chinese population: the China health and retirement longitudinal study | Inappropriate population |
| 4933 | Hemodynamic Responses to Resistance Exercise in Patients with Coronary Artery Disease | Inappropriate study design |
| 4934 | Management of atrial fibrillation in the elderly | Inappropriate population |
| 4935 | Prospective evaluation of haemoglobin oxygen saturation at rest and after exercise in paediatric sickle cell disease patients | Inappropriate population |
| 4936 | The impact of prolonged exercise in a cold environment upon cardiac function | Inappropriate study design |
| 4937 | Troponin release following endurance exercise: is inflammation the cause? a cardiovascular magnetic resonance study | Inappropriate study design |

| | | |
|---|---|---|
| 4938 | Methodology and baseline characteristics for the Sarcopenia and Hip Fracture study: a 5-year prospective study | Inappropriate study design |
| 4939 | Pilot study: Computer-based virtual anatomical interactivity for rehabilitation of individuals with chronic acquired brain injury | Inappropriate study design |
| 4940 | Low-magnitude whole body vibration with resistive exercise as a countermeasure against cardiovascular deconditioning after 60 days of head-down bed rest | Inappropriate study design |
| 4941 | Tai Chi Chuan exercise related change in brain function as assessed by functional near-infrared spectroscopy | Inappropriate intervention |
| 4942 | Evaluation of cardiovascular risks of spaceflight does not support the NASA bioastronautics critical path roadmap | Inappropriate population |
| 4943 | "You've got to keep moving, keep going": Understanding older patients' experiences and perceptions of delirium and nonpharmacological delirium prevention strategies in the acute hospital setting | Inappropriate population |
| 4944 | Hana kai ii: a 17-day dry saturation dive at 18.6 ATA. V. Maximal oxygen uptake | Inappropriate population |
| 4945 | Cognitive function modifies the effect of physiological function on the risk of multiple falls--a population-based study | Inappropriate population |
| 4946 | Early physiologic patterns in acute illness and accidents: toward a concept of circulatory dysfunction and shock based on invasive and noninvasive hemodynamic monitoring | Inappropriate population |
| 4947 | Bilateral spontaneous dissection of the internal carotid arteries--a case report | Inappropriate study design |
| 4948 | Effect of mental practice on the improvement of function and daily activity performance of the upper extremity in patients with subacute stroke: a randomized clinical trial | Inappropriate study design |
| 4949 | Impaired atrioventricular transport in patients with transposition of the great arteries palliated by atrial switch and preserved systolic right ventricular function: A magnetic resonance imaging study | Inappropriate population |
| 4950 | Spousal correlations for lifestyle factors and selected diseases in Chinese couples | Inappropriate study design |
| 4951 | A stair-climbing test for measuring mechanical efficiency of ambulation in adults with chronic stroke | Inappropriate study design |
| 4952 | Arterial blood pressure and cardiovascular responses to yoga practice | Inappropriate intervention |
| 4953 | Effect of Water Immersion on Dual-task Performance: Implications for Aquatic Therapy | Inappropriate population |
| 4954 | Plasma leptin: associations with metabolic, inflammatory and haemostatic risk factors for cardiovascular disease | Inappropriate outcome |
| 4955 | The modified rivermead mobility index: validity and reliability | Inappropriate study design |
| 4956 | Diabetes among Alaska Natives: a review | Inappropriate study design |
| 4957 | Association of behavioral risk factors with self-reported and symptom or measured chronic diseases among adult population (18-69 years) in India: evidence from SAGE study | Inappropriate study design |
| 4958 | Evidence for restricted muscle blood flow during speed skating | Inappropriate study design |
| 4959 | Healthy lifestyles reduce the incidence of chronic diseases and dementia: evidence from the Caerphilly cohort study | Inappropriate study design |
| 4960 | Associations of resting heart rate with insulin resistance, cardiovascular events and mortality in chronic kidney disease | Inappropriate study design |
| 4961 | Strength training improves submaximum cardiovascular performance in older men | Inappropriate study design |
| 4962 | Post-Exercise Hypotension and Reduced Cardiac Baroreflex after Half-Marathon Run: In Men, but Not in Women | Inappropriate study design |
| 4963 | Effects of treadmill incline and speed on peroneus longus muscle activity in persons with chronic stroke and healthy subjects | Inappropriate study design |
| 4964 | Task- and Context-Specific Balance Training Program Enhances Dynamic Balance and Functional Performance in Parkinsonian Nonfallers: A Randomized Controlled Trial With Six-Month Follow-Up | Inappropriate outcome |
| 4965 | Effects of a computer-based cognitive exercise program on age-related cognitive decline | Inappropriate study design |
| 4966 | Economic evaluation of pioglitazone hydrochloride in the management of type 2 diabetes mellitus in Canada | Inappropriate study design |
| 4967 | Pilot trial of a tele-rehab intervention to improve outcomes after stroke in Ghana: A feasibility and user satisfaction study | Inappropriate outcome |
| 4968 | A multi-centre, parallel-group, randomised controlled trial to assess the efficacy and safety of eurythmy therapy and tai chi in comparison with standard care in chronically ill elderly patients with increased risk of falling (ENTAiER): a trial protocol | Inappropriate study design |

| | | |
|---|---|---|
| 4969 | Usefulness of paradoxical systolic blood pressure increase after exercise as a predictor of cardiovascular mortality | Inappropriate population |
| 4970 | Health-related factors correlate with behavior trends in physical activity level in old age: longitudinal results from a population in São Paulo, Brazil | Inappropriate population |
| 4971 | Long-term secondary prevention of acute myocardial infarction (SEPAT) - guidelines adherence and outcome | Inappropriate study design |
| 4972 | The upper limit of alveolar capillary recruitment in a young man with lung growth impairment | Inappropriate study design |
| 4973 | Clinimetric properties of the motor activity log for the assessment of arm use in hemiparetic patients | Inappropriate study design |
| 4974 | Left ventricular systolic function and diastolic filling after intermittent high intensity team sports | Inappropriate study design |
| 4975 | Determinants of maximal oxygen uptake in severe acute hypoxia | Inappropriate study design |
| 4976 | A Systematic Review With Meta-Analysis of Mindful Exercises on Rehabilitative Outcomes Among Poststroke Patients | Inappropriate study design |
| 4977 | Enhanced ventilatory and exercise performance in athletes with slight expiratory resistive loading | Inappropriate study design |
| 4978 | Relationship of low-circulating "anti-aging" klotho hormone with disability in activities of daily living among older community-dwelling adults | Inappropriate population |
| 4979 | Determinants of mobility in populations of older adults: Results from a cross-sectional study in Finland, Poland and Spain | Inappropriate population |
| 4980 | Influence of respiratory pressure support on hemodynamics and exercise tolerance in patients with COPD | Inappropriate intervention |
| 4981 | Triad of impairment in older people with diabetes-reciprocal relations and clinical implications | Inappropriate study design |
| 4982 | Risk prediction models for mortality in patients with cardiovascular disease: The BioBank Japan project | Inappropriate intervention |
| 4983 | Incidence, reversibility, risk factors and the protective effect of high body mass index against sarcopenia in community-dwelling older Chinese adults | Inappropriate population |
| 4984 | The effect of temperature on hand function in patients with tremor | Inappropriate population |
| 4985 | Women and exertional heat illness: identification of gender specific risk factors | Inappropriate study design |
| 4986 | Combining risk communication strategies to simultaneously convey the risks of four diseases associated with physical inactivity to socio-demographically diverse populations | Inappropriate study design |
| 4987 | EMAS position statement: Diet and health in midlife and beyond | Inappropriate study design |
| 4988 | Restoration of the work capacity of the skeletal muscle with electrical myostimulation | Inappropriate study design |
| 4989 | Metaboreceptor activation in heart failure with reduced ejection fraction: Linking cardiac and peripheral vascular haemodynamics | Inappropriate population |
| 4990 | Tongue-based biofeedback for balance in stroke: results of an 8-week pilot study | Inappropriate population |
| 4991 | Prevalence of and factors related to mild and substantial dizziness in community-dwelling older adults: a cross-sectional study | Inappropriate population |
| 4992 | Effects of Wearable Sensor-Based Balance and Gait Training on Balance, Gait, and Functional Performance in Healthy and Patient Populations: A Systematic Review and Meta-Analysis of Randomized Controlled Trials | Inappropriate study design |
| 4993 | Elevated serum total bilirubin levels are negatively associated with major diabetic complications among Chinese senile diabetic patients | Inappropriate study design |
| 4994 | Effects of a 12-week intervention period with football and running for habitually active men with mild hypertension | Inappropriate population |
| 4995 | Exercise for preventing falls in older people living in the community | Inappropriate study design |
| 4996 | Impaired mitochondrial function and insulin resistance of skeletal muscle in mitochondrial diabetes | Inappropriate outcome |
| 4997 | The effects of urapidil therapy on hemodynamics and gas exchange in exercising patients with chronic obstructive pulmonary disease and pulmonary hypertension | Inappropriate population |
| 4998 | 'Real-world' management and outcomes of patients with paroxysmal vs. non-paroxysmal atrial fibrillation in Europe: the EURObservational Research Programme-Atrial Fibrillation (EORP-AF) General Pilot Registry | Inappropriate population |
| 4999 | The influence of menstrual cycle phase upon postexercise hypotension | Inappropriate population |
| 5000 | The impact of physical exercise on cognitive and affective functions and serum levels of brain-derived neurotrophic factor in nursing home residents: A randomized controlled trial | Inappropriate population |
| 5001 | Concomitant tuberculous meningitis and Lutembacher syndrome with multiple atrial septal defects and infective endocarditis | Inappropriate population |

| | | |
|---|---|---|
| 5002 | Cardiovascular and hormonal responses to static handgrip in young and older healthy men | Inappropriate study design |
| 5003 | Deleterious effects of beta-blockers on exercise capacity and hemodynamics in patients with portopulmonary hypertension | Inappropriate population |
| 5004 | Longterm effects of pulmonary resection on cardiopulmonary function | Inappropriate study design |
| 5005 | Vasodepressor effects of prazosin during insulin-induced hypoglycemia in hypertensive patients | Inappropriate study design |
| 5006 | Non-pharmacological interventions for spasticity in adults: An overview of systematic reviews | Inappropriate study design |
| 5007 | High altitude pulmonary edema and exercise at 4,400 meters on Mount McKinley. Effect of expiratory positive airway pressure | Inappropriate study design |
| 5008 | Effect of Pain Neuroscience Education Combined With Cognition-Targeted Motor Control Training on Chronic Spinal Pain: A Randomized Clinical Trial | Inappropriate study design |
| 5009 | The DEFIANT study of left ventricular function and exercise performance after acute myocardial infarction. Doppler Flow and Echocardiology in Functional Cardiac Insufficiency: Assessment of Nisoldipine Therapy Study Group | Inappropriate intervention |
| 5010 | Optimal management of older patients with atrial fibrillation | Inappropriate population |
| 5011 | Women show a closer association between educational level and hypertension or diabetes mellitus than males: a secondary analysis from the Austrian HIS | Inappropriate study design |
| 5012 | Feasibility of physical training in post-infarct patients with left ventricular aneurysm: a haemodynamic study | Inappropriate study design |
| 5013 | Gender difference in the proportionality factor between the mass specific VO2max and the ratio between HR(max) and HR(rest) | Inappropriate study design |
| 5014 | Left ventricular volume response to exercise in normal and coronary artery disease patients | Inappropriate study design |
| 5015 | Cardiorespiratory capacity after weight-supported treadmill training in patients with traumatic brain injury | Inappropriate study design |
| 5016 | Prediction of heat tolerance from heart rate and rectal temperature in a temperate environment | Inappropriate study design |
| 5017 | Exercise-Induced Cardiac Troponin I Increase and Incident Mortality and Cardiovascular Events | Inappropriate population |
| 5018 | The effects of cigarette smoking on C-reactive protein concentrations in men and women and its modification by exogenous oral hormones in women | Inappropriate study design |
| 5019 | Disproportionate shortening of diastolic time and cardiac performance in asymptomatic patients with aortic regurgitation | Inappropriate study design |
| 5020 | A survey of the prevalence of modifiable health risk behaviours among carers of people with a mental illness | Inappropriate study design |
| 5021 | Effects of acute beta-adrenergic receptor blockade on age-associated changes in cardiovascular performance during dynamic exercise | Inappropriate study design |
| 5022 | Effect of exposure to evening light on sleep initiation in the elderly: a longitudinal analysis for repeated measurements in home settings | Inappropriate study design |
| 5023 | Pulmonary function, muscle strength and mortality in old age | Inappropriate study design |
| 5024 | Mechanisms and mitigating factors for venous thromboembolism in chronic kidney disease: the REGARDS study | Inappropriate study design |
| 5025 | Hemodynamic comparison of twice daily metoprolol tartrate with once daily metoprolol succinate in congestive heart failure | Inappropriate population |
| 5026 | Cohort Profile: The Saguenay Youth Study (SYS) | Inappropriate study design |
| 5027 | Clinical relevance of apolipoprotein E genotyping based on a family history of Alzheimer's disease | Inappropriate study design |
| 5028 | Physical activity interventions are delivered consistently across hospitalized older adults but multimorbidity is associated with poorer rehabilitation outcomes: A population-based cohort study | Inappropriate study design |
| 5029 | Circulating neuropeptide Y in humans: relation to changes in catecholamine levels and changes in hemodynamics | Inappropriate study design |
| 5030 | Reduced Right Ventricular Output Reserve in Patients With Systemic Sclerosis and Mildly Elevated Pulmonary Artery Pressure | Inappropriate study design |
| 5031 | Postexercise heat loss and hemodynamic responses during head-down tilt are similar between genders | Inappropriate population |
| 5032 | Development and Clinical Evaluation of a Web-Based Upper Limb Home Rehabilitation System Using a Smartwatch and Machine Learning Model for Chronic Stroke Survivors: Prospective Comparative Study | Inappropriate study design |
| 5033 | Moderate exercise training reduces arterial chemoreceptor reflex drive in mild hypertension | Inappropriate population |

| | | |
|---|---|---|
| 5034 | Assessment of left ventricular preload by cardiac magnetic resonance imaging predicts exercise capacity in adult operated tetralogy of Fallot: a retrospective study | Inappropriate population |
| 5035 | Chronic organ failure in adult sickle cell disease | Inappropriate study design |
| 5036 | Reproducibility of the exponential rise technique of CO(2) rebreathing for measuring P(v)CO(2) and C(v)CO(2 )to non-invasively estimate cardiac output during incremental, maximal treadmill exercise | Inappropriate study design |
| 5037 | Efficiency of work production by spastic muscles | Inappropriate outcome |
| 5038 | Anaerobic capacity and maximal oxygen uptake during arm stroke, leg kicking and whole body swimming | Inappropriate population |
| 5039 | Hemorheological disturbances in cerebrovascular diseases | Inappropriate study design |
| 5040 | Loss and recovery of independence among seniors | Inappropriate study design |
| 5041 | Usefulness of maximal oxygen pulse in timing of pulmonary valve replacement in patients with isolated pulmonary regurgitation | Inappropriate study design |
| 5042 | Physical Activity, Vitamin D, and Incident Atherosclerotic Cardiovascular Disease in Whites and Blacks: The ARIC Study | Inappropriate study design |
| 5043 | Incidence of postoperative atrial fibrillation recurrence in patients discharged in sinus rhythm after cardiac surgery: a systematic review and meta-analysis | Inappropriate study design |
| 5044 | Comparison of different interdialytic intervals among hemodialysis patients on their echocardiogram-based cardiovascular parameters | Inappropriate outcome |
| 5045 | Effect of short-term endurance training on exercise capacity, haemodynamics and atrial natriuretic peptide secretion in heart transplant recipients | Inappropriate study design |
| 5046 | Concomitant carotid and coronary artery reconstruction | Inappropriate study design |
| 5047 | Reducing Potentially Excess Deaths from the Five Leading Causes of Death in the Rural United States | Inappropriate study design |
| 5048 | Cinnamon users with prediabetes have a better fasting working memory: a cross-sectional function study | Inappropriate study design |
| 5049 | Development of a new method for assessing global risk of Alzheimer's disease for use in population health approaches to prevention | Inappropriate population |
| 5050 | Factors of non-compliance with the therapeutic regimen among hypertensive men and women: a case-control study to investigate risk factors of stroke | Inappropriate study design |
| 5051 | Effect of Combined Physical and Cognitive Interventions on Executive Functions in OLDER Adults: A Meta-Analysis of Outcomes | Inappropriate study design |
| 5052 | Blackcurrant Alters Physiological Responses and Femoral Artery Diameter during Sustained Isometric Contraction | Inappropriate study design |
| 5053 | Cardiovascular and pulmonary responses to increased acceleration forces during rest and exercise | Inappropriate study design |
| 5054 | Effect of adrenergic blockade on plasma adrenomedullin concentration during static handgrip in patients with heart failure | Inappropriate study design |
| 5055 | Non-pharmacological interventions for preventing secondary vascular events after stroke or transient ischemic attack | Inappropriate population |
| 5056 | Silent myocardial ischemia during rehabilitation for cerebrovascular disease | Inappropriate intervention |
| 5057 | The association of income with health behavior change and disease monitoring among patients with chronic disease | Inappropriate study design |
| 5058 | A self-administered Graded Repetitive Arm Supplementary Program (GRASP) improves arm function during inpatient stroke rehabilitation: a multi-site randomized controlled trial | Inappropriate study design |
| 5059 | Atrial fibrillation | Inappropriate population |
| 5060 | Cardiovascular Events in a Physical Activity Intervention Compared With a Successful Aging Intervention: The LIFE Study Randomized Trial | Inappropriate outcome |
| 5061 | Safety and efficacy of beta blockade in patients with chronic congestive heart failure awaiting transplantation | Inappropriate study design |
| 5062 | Self-care agency, lifestyle, and physical condition predict future frailty in community-dwelling older people | Inappropriate intervention |
| 5063 | Adaptive rate pacing controlled by the right ventricular preejection interval: clinical experience with a physiological pacing system | Inappropriate study design |
| 5064 | Mirror therapy for involuntary movement due to chronic thalamic hemorrhage: a case report | Inappropriate study design |
| 5065 | Cardiovascular function during sleep of active older adults and the effects of exercise | Inappropriate population |
| 5066 | Early Pulmonary Vascular Disease in Young Adults Born Preterm | Inappropriate population |
| 5067 | Risk factors associated with multiple correlated health outcomes in the 500 Cities Project | Inappropriate study design |

| | | |
|---|---|---|
| 5068 | Dehydration markedly impairs cardiovascular function in hyperthermic endurance athletes during exercise | Inappropriate study design |
| 5069 | Polymorphisms of the beta(2)-adrenergic receptor determine exercise capacity in patients with heart failure | Inappropriate population |
| 5070 | Echocardiographic dimensions and function in adults with primary growth hormone resistance (Laron syndrome) | Inappropriate population |
| 5071 | Cardiac effects of short term arm crank training in paraplegics: echocardiographic evidence | Inappropriate study design |
| 5072 | Left ventricular strain modifications after maximal exercise in athletes: a speckle tracking study | Inappropriate study design |
| 5073 | Anistreplase versus alteplase in acute myocardial infarction: comparative effects on left ventricular function, morbidity and 1-day coronary artery patency. The TEAM-3 Investigators | Inappropriate population |
| 5074 | Effect of walking distance on 8-year incident depressive symptoms in elderly men with and without chronic disease: the Honolulu-Asia Aging Study | Inappropriate study design |
| 5075 | Hemodynamic effects of nebivolol in men: comparison of radionuclide angiocardiography with systolic time intervals | Inappropriate study design |
| 5076 | An exaggerated systolic blood pressure response to exercise is associated with cardiovascular remodeling in subjects with prehypertension | Inappropriate study design |
| 5077 | Serum uric acid is inversely proportional to estimated stroke volume and cardiac output in a large sample of pharmacologically untreated subjects: data from the Brisighella Heart Study | Inappropriate population |
| 5078 | Mortality rates in major and subthreshold depression: 10-year follow-up of a Singaporean population cohort of older adults | Inappropriate study design |
| 5079 | Physiological and clinical consequences of relief of right ventricular outflow tract obstruction late after repair of congenital heart defects | Inappropriate study design |
| 5080 | Childhood adversity heightens the impact of later-life caregiving stress on telomere length and inflammation | Inappropriate study design |
| 5081 | Oral contraceptive cycle phase does not affect 200-m swim time trial performance | Inappropriate study design |
| 5082 | Concurrent validity of the GAITRite electronic walkway and the 10-m walk test for measurement of walking speed after stroke | Inappropriate study design |
| 5083 | Smoking modifies the associated increased risk of future cardiovascular disease by genetic variation on chromosome 9p21 | Inappropriate population |
| 5084 | Changes in right ventricular structure and function assessed using cardiac magnetic resonance imaging in bosentan-treated patients with pulmonary arterial hypertension | Inappropriate study design |
| 5085 | Design and preliminary evaluation of the FINGER rehabilitation robot: controlling challenge and quantifying finger individuation during musical computer game play | Inappropriate study design |
| 5086 | Capability of 2 gait measures for detecting response to gait training in stroke survivors: Gait Assessment and Intervention Tool and the Tinetti Gait Scale | Inappropriate study design |
| 5087 | Heavy snoring is a risk factor for case fatality and poor short-term prognosis after a first acute myocardial infarction | Inappropriate study design |
| 5088 | Clinical and haemodynamic effects of low dose captopril in severe chronic heart failure | Inappropriate study design |
| 5089 | Accelerometer-triggered electrical stimulation for reach and grasp in chronic stroke patients: a pilot study | Inappropriate study design |
| 5090 | Effects of modality change and transplant on peak oxygen uptake in patients with kidney failure | Inappropriate population |
| 5091 | Predictors of 1000-m outrigger canoeing performance | Inappropriate population |
| 5092 | Prediction of maximal cardiac output in preoperative patients with coronary artery disease | Inappropriate study design |
| 5093 | Relationship between Physical Inactivity and Long-term Outcome in Patients Aged≥80 Years with Acute Coronary Syndrome | Inappropriate study design |
| 5094 | Differential Clinical Profiles, Exercise Responses, and Outcomes Associated With Existing HFpEF Definitions | Inappropriate population |
| 5095 | Determinants of clinical response and survival in patients with congestive heart failure treated with captopril | Inappropriate study design |
| 5096 | Chinese Version of the Assessment of Survivor Concerns Scale for Gynecological Cancer Survivors: A Psychometric Study in Taiwan | Inappropriate study design |
| 5097 | The prognostic value of pre-discharge exercise testing after myocardial infarction treated with either primary PCI or fibrinolysis: a DANAMI-2 sub-study | Inappropriate population |
| 5098 | Thermoregulatory responses to low-intensity prolonged swimming in water at various temperatures and treadmill walking on land | Inappropriate study design |

| | | |
|---|---|---|
| 5099 | Tennis for physical health: acute age- and gender-based physiological responses to cardio tennis | Inappropriate study design |
| 5100 | Evaluation of a body-worn sensor system to measure physical activity in older people with impaired function | Inappropriate outcome |
| 5101 | Radionuclide ventriculography and two dimensional echocardiography as predictors of left ventricular performance during aortic vascular surgery | Inappropriate study design |
| 5102 | Ramipril and captopril in patients with heart failure: effects on hemodynamics and vasoconstrictor systems | Inappropriate population |
| 5103 | Energy metabolism in exertional heat stroke with acute renal failure | Inappropriate population |
| 5104 | Coronary artery disease is highly prevalent among patients with premature peripheral vascular disease | Inappropriate study design |
| 5105 | Participation in leisure activities in adolescents with congenital heart defects | Inappropriate study design |
| 5106 | Left atrial myocardial dysfunction after chronic abuse of anabolic androgenic steroids: a speckle tracking echocardiography analysis | Inappropriate study design |
| 5107 | Echocardiographic predictors of exercise capacity and mortality in chronic obstructive pulmonary disease | Inappropriate study design |
| 5108 | Strategies to prevent falls and fractures in hospitals and care homes and effect of cognitive impairment: systematic review and meta-analyses | Inappropriate study design |
| 5109 | Association between obesity and selected morbidities: a study of BRICS countries | Inappropriate outcome |
| 5110 | Tissue plasminogen activator, fibrin D-dimer, and insulin resistance in the relatives of patients with premature coronary artery disease | Inappropriate study design |
| 5111 | Short- and intermediate-term clinical outcomes from direct myocardial laser revascularization guided by biosense left ventricular electromechanical mapping | Inappropriate population |
| 5112 | Phase I and Phase II oxygen uptake kinetics during atrioventricular dyssynchrony in chronotropically competent pacemaker patients | Inappropriate population |
| 5113 | Alcohol consumption, Lewis phenotypes, and risk of ischaemic heart disease | Inappropriate study design |
| 5114 | Living With Chronic Obstructive Pulmonary Disease: The Process of Self-Managing Chronic Obstructive Pulmonary Disease | Inappropriate study design |
| 5115 | Interdisciplinary cardiovascular and neurologic outpatient rehabilitation in patients surviving transient ischemic attack or stroke with minor or no residual deficits | Inappropriate study design |
| 5116 | Effects of a community-based intensive motor training program combined with early supported discharge after treatment in a comprehensive stroke unit: a randomized, controlled trial | Inappropriate study design |
| 5117 | Pelvic floor muscle training for urinary incontinence in female stroke patients: a randomized, controlled and blinded trial | Inappropriate study design |
| 5118 | Evidence suggesting that a chronic disease self-management program can improve health status while reducing hospitalization: a randomized trial | Inappropriate study design |
| 5119 | A Case of Adult-onset Pompe Disease with Cerebral Stroke and Left Ventricular Hypertrophy | Inappropriate study design |
| 5120 | Stability of relative oxygen pulse curve during repeated maximal cardiopulmonary testing in professional soccer players | Inappropriate study design |
| 5121 | The role of physical activity in the development of first cardiovascular disease event: a tree-structured survival analysis of the Danish ADDITION-PRO cohort | Inappropriate population |
| 5122 | Eigen Posture Based Fall Risk Assessment System Using Kinect | Inappropriate outcome |
| 5123 | Patterns of Oxygen Pulse Curve in Response to Incremental Exercise in Patients with Chronic Obstructive Pulmonary Disease - An Observational Study | Inappropriate population |
| 5124 | Macroangiopathy in adults and children with diabetes: risk factors (part 2) | Inappropriate population |
| 5125 | Coronary heart disease from a life-course approach: findings from the health and retirement study, 1998-2004 | Inappropriate population |
| 5126 | Natural history and evolution of peripheral obstructive arterial disease | Inappropriate study design |
| 5127 | Health-related behaviours of people with diabetes and those with cardiometabolic risk factors: results from SHIELD | Inappropriate study design |
| 5128 | Age hemodynamics and exercise in essential hypertension: difference between beta-blockers and dihydropyridine calcium antagonists | Inappropriate study design |
| 5129 | Prognostic Role of Hypertensive Response to Exercise in Patients With Repaired Coarctation of Aorta | Inappropriate study design |
| 5130 | Glucosuria and all-cause mortality among general screening participants | Inappropriate study design |
| 5131 | A support programme for secondary prevention in patients with transient ischaemic attack and minor stroke (INSPiRE-TMS): an open-label, randomised controlled trial | Inappropriate outcome |
| 5132 | Determinants of quality of life of patients with heart failure and iron deficiency treated with ferric carboxymaltose: FAIR-HF sub-analysis | Inappropriate population |

| | | |
|---|---|---|
| 5133 | Healthy lifestyle and risk of kidney disease progression, atherosclerotic events, and death in CKD: findings from the Chronic Renal Insufficiency Cohort (CRIC) Study | Inappropriate population |
| 5134 | Unexpected dual task benefits on cycling in Parkinson disease and healthy adults: a neuro-behavioral model | Inappropriate population |
| 5135 | Surgical revascularization after acute myocardial infarction. Does timing make a difference? | Inappropriate population |
| 5136 | Death from dialysis termination | Inappropriate study design |
| 5137 | Fluid retention index predicts the 30-day mortality in geriatric care | Inappropriate study design |
| 5138 | Echocardiography during preload stress for evaluation of right ventricular contractile reserve and exercise capacity in pulmonary hypertension | Inappropriate population |
| 5139 | Is autonomic support of arterial blood pressure related to habitual exercise status in healthy men? | Inappropriate study design |
| 5140 | Cardiac event rate in a lifestyle modification program for patients with chronic coronary artery disease | Inappropriate population |
| 5141 | Effects of cholinergic and beta-adrenergic blockade on orthostatic tolerance in healthy subjects | Inappropriate study design |
| 5142 | Cardiovascular and renal effects of growth hormone | Inappropriate study design |
| 5143 | Factors Contributing to 50-ft Walking Speed and Observed Ethnic Differences in Older Community-Dwelling Mexican Americans and European Americans | Inappropriate study design |
| 5144 | Low Incidence of Death and Renal Failure in United States Military Service Members Hospitalized with Exertional Heat Stroke: A Retrospective Cohort Study | Inappropriate study design |
| 5145 | The impact of a healthy lifestyle on Disability-Adjusted Life Years: a prospective cohort study | Inappropriate study design |
| 5146 | The contribution of osteoarthritis to disability: preliminary data from the Women's Health and Aging Study | Inappropriate study design |
| 5147 | Missed opportunities in preventive counseling for cardiovascular disease--United States, 1995 | Inappropriate population |
| 5148 | Central circulation during exercise after venesection and reinfusion of red blood cells | Inappropriate study design |
| 5149 | Is it worth offering cardiovascular disease prevention to the elderly? | Inappropriate study design |
| 5150 | Association of anemia correction with health related quality of life in patients not on dialysis | Inappropriate population |
| 5151 | Development of co-selected single nucleotide polymorphisms in the viral promoter precedes the onset of human immunodeficiency virus type 1-associated neurocognitive impairment | Inappropriate study design |
| 5152 | Comparison of the cardiac and hemodynamic effects of lisinopril and atenolol in patients with hypertension: therapeutic implications | Inappropriate intervention |
| 5153 | Risk of mortality following hip fracture in Japan | Inappropriate study design |
| 5154 | Evaluation of left ventricular performance in acutely ill patients with chronic obstructive lung disease | Inappropriate study design |
| 5155 | Non-supportive interactions in the experience of women family caregivers | Inappropriate study design |
| 5156 | Improving cognitive and physical function through 12-weeks of resistance training in older adults: Randomized controlled trial | Inappropriate population |
| 5157 | Hemodynamic and neuroendocrine responses to changes in sodium intake in compensated heart failure | Inappropriate outcome |
| 5158 | The impact of chronic diseases on the health-related quality of life (HRQOL) of Chinese patients in primary care | Inappropriate population |
| 5159 | WITHDRAWN: Interventions for fatigue and weight loss in adults with advanced progressive illness | Inappropriate intervention |
| 5160 | Therapy for triggered acute risk prevention: a study of feasibility | Inappropriate outcome |
| 5161 | Interventions for fatigue and weight loss in adults with advanced progressive illness | Inappropriate outcome |
| 5162 | Prevalence of selected risk behaviors and chronic diseases--Behavioral Risk Factor Surveillance System (BRFSS), 39 steps communities, United States, 2005 | Inappropriate study design |
| 5163 | Using an upper extremity exoskeleton for semi-autonomous exercise during inpatient neurological rehabilitation- a pilot study | Inappropriate population |
| 5164 | Physiologic improvement following coronary artery bypass surgery | Inappropriate population |
| 5165 | The influence of a mild thermal challenge and severe hypoxia on exercise performance and serum BDNF | Inappropriate study design |
| 5166 | Long-term follow-up after surgical repair of ostium primum atrial septal defects in adults | Inappropriate population |

| | | |
|---|---|---|
| 5167 | An exploratory study of parents' approaches to health promotion in families of adolescents with physical disabilities | Inappropriate population |
| 5168 | Haemodynamic benefit of a rate-adapted A-V delay in dual chamber pacing | Inappropriate population |
| 5169 | Evaluation of the Effectiveness of Compression Garments on Autonomic Nervous System Recovery After Exercise | Inappropriate outcome |
| 5170 | Association Between Life's Simple 7 and Noncardiovascular Disease: The Multi-Ethnic Study of Atherosclerosis | Inappropriate study design |
| 5171 | Cause-specific mortality in osteoarthritis of peripheral joints | Inappropriate study design |
| 5172 | Association of the magnitude of weight loss and changes in physical fitness with long-term cardiovascular disease outcomes in overweight or obese people with type 2 diabetes: a post-hoc analysis of the Look AHEAD randomised clinical trial | Inappropriate population |
| 5173 | Association of a diabetes risk score with risk of myocardial infarction, stroke, specific types of cancer, and mortality: a prospective study in the European Prospective Investigation into Cancer and Nutrition (EPIC)-Potsdam cohort | Inappropriate population |
| 5174 | The influence of chronic disease, physical function, and lifestyle on health transition among the middle-aged and older persons in Taiwan | Inappropriate study design |
| 5175 | Early Supported Discharge by Caregiver-Mediated Exercises and e-Health Support After Stroke: A Proof-of-Concept Trial | Inappropriate study design |
| 5176 | Impact of sex on cardiovascular outcome in patients at high cardiovascular risk: analysis of the Telmisartan Randomized Assessment Study in ACE-Intolerant Subjects With Cardiovascular Disease (TRANSCEND) and the Ongoing Telmisartan Alone and in Combination With Ramipril Global End Point Trial (ONTARGET) | Inappropriate population |
| 5177 | Assessment of muscle function using hybrid PET/MRI: comparison of (18)F-FDG PET and T2-weighted MRI for quantifying muscle activation in human subjects | Inappropriate study design |
| 5178 | Effect of simulated altitude erythrocythemia in women on hemoglobin flow rate during exercise | Inappropriate population |
| 5179 | Cardiovascular function in patients with progressive systemic sclerosis (scleroderma) | Inappropriate study design |
| 5180 | Cardiovascular adaptations to intense swim training in sedentary middle-aged men and women | Inappropriate population |
| 5181 | β-Blockers and ivabradine differentially affect cardiopulmonary function and left ventricular filling index | Inappropriate population |
| 5182 | Association between renal insufficiency and inducible ischemia in patients with coronary artery disease: the heart and soul study | Inappropriate study design |
| 5183 | Two dimensional and real-time three dimensional ultrasound measurements of left ventricular diastolic function after marathon running: results from a substudy of the BeMaGIC trial | Inappropriate study design |
| 5184 | Study of the acute effect of AR-L 115 BS, a new positive-inotropic agent in patients with exercise-induced heart failure | Inappropriate population |
| 5185 | Impaired cardiac and peripheral hemodynamic responses to inhaled $\beta_2$-agonist in cystic fibrosis | Inappropriate study design |
| 5186 | Left ventricular diastolic filling performance in older male athletes | Inappropriate study design |
| 5187 | Factors related to gait and balance deficits in older adults | Inappropriate population |
| 5188 | Intra-ocular pressure changes during maximal isometric contraction: does this reflect intra-cranial pressure or retinal venous pressure? | Inappropriate population |
| 5189 | Cardiodynamic responses during seated and supine recovery from supramaximal exercise | Inappropriate study design |
| 5190 | Nifedipine improves left ventricular function in patients with hypertension | Inappropriate intervention |
| 5191 | Long term haemodynamic effects of pinacidil and hydralazine in arterial hypertension | Inappropriate study design |
| 5192 | Effects of lacidipine on peak oxygen consumption, neurohormones and invasive haemodynamics in patients with mild to moderate chronic heart failure | Inappropriate study design |
| 5193 | Haemodynamic effects of physical training in essential hypertension | Inappropriate study design |
| 5194 | Altered hemodynamics during muscle metaboreflex in young type 1 diabetes patients | Inappropriate study design |
| 5195 | Relationship between sleep duration and self-reported health-related quality of life among US adults with or without major chronic diseases, 2014 | Inappropriate study design |
| 5196 | Falls in elderly | Inappropriate population |
| 5197 | Peak expiratory flow rate in an elderly population | Inappropriate study design |
| 5198 | Effect of Transcutaneous Electrical Nerve Stimulation on Plantar Flexor Muscle Spasticity and Walking Speed in Stroke Patients | Inappropriate study design |
| 5199 | Observing the onset of disability in older adults | Inappropriate study design |

| | | |
|---|---|---|
| 5200 | Association of CKD with disability in the United States | Inappropriate population |
| 5201 | Tissue kallikrein preventing the restenosis after stenting of symptomatic MCA atherosclerotic stenosis (KPRASS) | Inappropriate study design |
| 5202 | Impact of cognitive performance on disease-related knowledge six months after multi-component rehabilitation in patients after an acute cardiac event | Inappropriate study design |
| 5203 | Favourable interaction of calcium antagonist plus ACE inhibitor on cardiac haemodynamics in treating hypertension: rest and effort evaluation | Inappropriate study design |
| 5204 | Antibodies to Human Herpesviruses in Myalgic Encephalomyelitis/Chronic Fatigue Syndrome Patients | Inappropriate population |
| 5205 | An Exertional Heat Stroke Survivor's Return to Running: An Integrated Approach on the Treatment, Recovery, and Return to Activity | Inappropriate population |
| 5206 | Determinants of pulmonary perfusion measured by electrical impedance tomography | Inappropriate study design |
| 5207 | Discriminating Circulatory Problems From Deconditioning: Echocardiographic and Cardiopulmonary Exercise Test Analysis | Inappropriate study design |
| 5208 | Left ventricular volumes and hemodynamic responses to postexercise ischemia in healthy humans | Inappropriate population |
| 5209 | Physiological consequences of percutaneous pulmonary valve implantation: the different behaviour of volume- and pressure-overloaded ventricles | Inappropriate study design |
| 5210 | Cardiac adaptation to increased systemic blood pressure in borderline hypertensive men | Inappropriate outcome |
| 5211 | Effects of a home-based step training programme on balance, stepping, cognition and functional performance in people with multiple sclerosis--a randomized controlled trial | Inappropriate study design |
| 5212 | Transposed firing activation of motor units | Inappropriate study design |
| 5213 | Effects of prenalterol administered orally in patients with congestive heart failure | Inappropriate study design |
| 5214 | Effects of angiotensin converting enzyme inhibitors in hypertensive patients with aortic valve stenosis: a drug withdrawal study | Inappropriate study design |
| 5215 | Normalization of right ventricular performance and remodeling evaluated by magnetic resonance imaging at late follow-up of heart transplantation: relationship between function, exercise capacity and pulmonary vascular resistance | Inappropriate population |
| 5216 | Estimated Glomerular Filtration Rate Decline and Incident Frailty in Older Adults | Inappropriate study design |
| 5217 | Ventilatory responses and entrainment of breathing during rowing | Inappropriate study design |
| 5218 | Long-term health outcomes and cost-effectiveness of a computer-tailored physical activity intervention among people aged over fifty: modelling the results of a randomized controlled trial | Inappropriate outcome |
| 5219 | Cardiac origins of the postural orthostatic tachycardia syndrome | Inappropriate study design |
| 5220 | Impact of blood pressure on the Doppler echocardiographic assessment of severity of aortic stenosis | Inappropriate study design |
| 5221 | Middle cerebral artery blood flow velocity in elite power athletes during maximal weight-lifting | Inappropriate population |
| 5222 | Insulin resistance as estimated by homeostasis model assessment predicts incident symptomatic cardiovascular disease in caucasian subjects from the general population: the Bruneck study | Inappropriate population |
| 5223 | Improving activity recognition using a wearable barometric pressure sensor in mobility-impaired stroke patients | Inappropriate study design |
| 5224 | Perceptions of body size, obesity threat and the willingness to lose weight among black South African adults: a qualitative study | Inappropriate study design |
| 5225 | Metaboreflex-mediated hemodynamic abnormalities in individuals with coronary artery disease without overt signs or symptoms of heart failure | Inappropriate study design |
| 5226 | Metabolic syndrome in coronary artery and occlusive vascular diseases: a systematic review | Inappropriate population |
| 5227 | Therapy for triggered acute risk prevention in subjects at increased cardiovascular risk | Inappropriate study design |
| 5228 | QRS duration in Fontan circulation in adults: a predictor of aerobic capacity | Inappropriate study design |
| 5229 | Left ventricular end-diastolic dimension and septal e' are predictors of cardiac index at rest, while tricuspid annular plane systolic excursion is a predictor of peak oxygen uptake in patients with pulmonary hypertension | Inappropriate study design |
| 5230 | Cardiac responses to maximal anisotonic isometric contractions during handgrip and leg extension | Inappropriate study design |
| 5231 | Improved work capacity but unchanged peak oxygen uptake during primary rehabilitation in tetraplegic patients | Inappropriate study design |

| | | |
|---|---|---|
| 5232 | Cardiovascular function following reduced aerobic activity | Inappropriate population |
| 5233 | Effects of a short-term training programme in post-infarct patients with residual myocardial ischaemia | Inappropriate population |
| 5234 | Successful aging: development and testing of a multidimensional model using data from a large sample of older australians | Inappropriate study design |
| 5235 | A haemodynamic dose finding study with a new slow-calcium channel blocker (amlodipine) in coronary artery disease | Inappropriate study design |
| 5236 | Epidemiology of coronary artery disease and stroke and associated risk factors in Gaza community -Palestine | Inappropriate study design |
| 5237 | Prevalence of multimorbidity among Bangladeshi adult population: a nationwide cross-sectional study | Inappropriate population |
| 5238 | Handgrip strength, chronic physical conditions and physical multimorbidity in middle-aged and older adults in six low- and middle income countries | Inappropriate study design |
| 5239 | The effect of a telephone follow-up intervention on illness perception and lifestyle after myocardial infarction in China: a randomized controlled trial | Inappropriate population |
| 5240 | The Association of Coronary Artery Calcium With Noncardiovascular Disease: The Multi-Ethnic Study of Atherosclerosis | Inappropriate study design |
| 5241 | Randomized, double-blind, placebo-controlled, multicenter study to evaluate the safety and tolerability of metrifonate in patients with probable Alzheimer disease. The Metrifonate Study Group | Inappropriate study design |
| 5242 | Dietary Patterns and Blood Pressure in Adults: A Systematic Review and Meta-Analysis of Randomized Controlled Trials | Inappropriate population |
| 5243 | Comorbidities of diabetes and hypertension: mechanisms and approach to target organ protection | Inappropriate study design |
| 5244 | Correlates of sedentary behavior in the general population: A cross-sectional study using nationally representative data from six low- and middle-income countries | Inappropriate study design |
| 5245 | Measures of adiposity in the identification of metabolic abnormalities in elderly men | Inappropriate study design |
| 5246 | Early famine exposure and adult disease risk based on a 10-year prospective study of Chinese adults | Inappropriate population |
| 5247 | Predictors of impaired heart rate recovery: a myocardial perfusion SPECT study | Inappropriate study design |
| 5248 | Modulation of nutritional state in Parkinsonian patients with bilateral subthalamic nucleus stimulation | Inappropriate population |
| 5249 | Improving the health of Oklahomans through clinical prevention. Part 1: Counseling to decrease major risk factors | Inappropriate population |
| 5250 | Estimating right ventricular stroke work and the pulsatile work fraction in pulmonary hypertension | Inappropriate study design |
| 5251 | What Do We Know about Diet and Markers of Cardiovascular Health in Children: A Review | Inappropriate study design |
| 5252 | Incidence of cardiovascular events in patients with stabilized coronary heart disease: the EUROASPIRE IV follow-up study | Inappropriate population |
| 5253 | Acute Cardiovascular Adaptation to Strenuous Exercise: An Integrative Ultrasound Study | Inappropriate population |
| 5254 | Major adverse cardiac events during endurance sports | Inappropriate study design |
| 5255 | Protocol for the HAPPY Hearts study: cardiovascular screening for the early detection of future adverse cardiovascular outcomes in middle-aged and older women: a prospective, observational cohort study | Inappropriate study design |
| 5256 | A Multicomponent Exercise Intervention that Reverses Frailty and Improves Cognition, Emotion, and Social Networking in the Community-Dwelling Frail Elderly: A Randomized Clinical Trial | Inappropriate population |
| 5257 | Modified Fontan operation for complex cardiac anomalies--postoperative hemodynamics, cardiac function and clinical status | Inappropriate population |
| 5258 | Prevalence of memory disorders in ambulatory patients aged ≥70 years with chronic heart failure (from the EFICARE study) | Inappropriate population |
| 5259 | Multifactorial intervention with nurse practitioners does not change cardiovascular outcomes in patients with chronic kidney disease | Inappropriate study design |
| 5260 | Double-blind, placebo-controlled study of the long-term efficacy of carvedilol in patients with severe chronic heart failure | Inappropriate study design |
| 5261 | Influence of mental stress on ventricular pump function in postinfarction patients. An invasive hemodynamic investigation | Inappropriate study design |
| 5262 | Quality of life (QOL) of the community-dwelling elderly and associated factors: a population-based study in urban areas of China | Inappropriate study design |
| 5263 | Measuring quality outcomes across hospital systems: Using a claims data model for risk adjustment of mortality rates | Inappropriate study design |

| | | |
|---|---|---|
| 5264 | Contralaterally controlled functional electrical stimulation for recovery of elbow extension and hand opening after stroke: a pilot case series study | Inappropriate study design |
| 5265 | Plasma fibrinogen and its association with cardiovascular risk factors in apparently healthy Japanese subjects | Inappropriate population |
| 5266 | Walking and leisure-time activity and risk of hip fracture in postmenopausal women | Inappropriate study design |
| 5267 | Ambient fine particulate matter pollution and years of life lost from cardiovascular diseases in 48 large Chinese cities: Association, effect modification, and additional life gain | Inappropriate study design |
| 5268 | Feasibility of lower-limb muscle power training to enhance locomotor function poststroke | Inappropriate study design |
| 5269 | Gender differences in tolerance to lower body negative pressure | Inappropriate population |
| 5270 | Absence of training-specific cardiac adaptation in paraplegic athletes | Inappropriate study design |
| 5271 | Large therapeutic studies in elderly patients with hypertension | Inappropriate study design |
| 5272 | Predictive factors of self-care capacity in veterans care institution residents | Inappropriate population |
| 5273 | Determinants of right ventricular ejection fraction in pulmonary arterial hypertension | Inappropriate study design |
| 5274 | Cardiovascular responses to active and passive cycling movements | Inappropriate population |
| 5275 | Association Between Sickle Cell Trait With Selected Chronic Medical Conditions in U.S. Service Members | Inappropriate study design |
| 5276 | Comparison of the effectiveness of active and passive neuromuscular electrical stimulation of hemiplegic upper extremities: a randomized, controlled trial | Inappropriate study design |
| 5277 | Right ventricular function before and after percutaneous balloon mitral valvuloplasty | Inappropriate study design |
| 5278 | Effect of muscle metaboreflex activation on central hemodynamics and cardiac function in humans | Inappropriate population |
| 5279 | Clinical impact of a structured secondary cardiovascular prevention program following acute coronary syndromes: A prospective multicenter healthcare intervention | Inappropriate study design |
| 5280 | Factors related to falls, weight-loss and pressure ulcers--more insight in risk assessment among nursing home residents | Inappropriate study design |
| 5281 | Haemodynamic responses of wearing low-pressure sports compression tights during an orthostatic challenge in healthy individuals | Inappropriate study design |
| 5282 | Peak exercise oxygen pulse and prognosis in chronic heart failure | Inappropriate study design |
| 5283 | Acute cardiac functional and mechanical responses to isometric exercise in prehypertensive males | Inappropriate outcome |
| 5284 | Effect of parasympathetic impairment on the haemodynamic response to handgrip in Chagas's heart disease | Inappropriate study design |
| 5285 | Non-invasive assessment of myocardial recovery on chronic left ventricular assist device: results associated with successful device removal | Inappropriate study design |
| 5286 | Validity of ratings of perceived exertion as an index of exercise intensity in swimming training | Inappropriate study design |
| 5287 | Stimulus-specific functional remodeling of the left ventricle in endurance and resistance-trained men | Inappropriate study design |
| 5288 | Clinical, socioeconomic, and behavioural factors at age 50 years and risk of cardiometabolic multimorbidity and mortality: A cohort study | Inappropriate outcome |
| 5289 | Leisure-time cross-country skiing is associated with lower incidence of hypertension: a prospective cohort study | Inappropriate population |
| 5290 | Temporal trends and associated factors of inpatient cardiac rehabilitation in patients with acute myocardial infarction: a community-wide perspective | Inappropriate intervention |
| 5291 | Myocardial infarction in the Wisconsin Longitudinal Study: the interaction among environmental, health, social, behavioural and genetic factors | Inappropriate population |
| 5292 | Association between left ventricular structure and cardiac performance during effort in two morphological forms of athlete's heart | Inappropriate population |
| 5293 | Effect of metabolic syndrome or type II diabetes mellitus on the occurrence of recurrent vascular events in hypertensive patients | Inappropriate study design |
| 5294 | Prevalence and determinants of metabolic syndrome among women in Chinese rural areas | Inappropriate study design |
| 5295 | Echocardiographic evidence of right ventricular functional improvement after balloon pulmonary angioplasty in chronic thromboembolic pulmonary hypertension | Inappropriate population |
| 5296 | Inter-regional comparisons of the prevalence of cardiometabolic risk factors in patients with hypertension in Europe: the GOOD survey | Inappropriate study design |

| | | |
|---|---|---|
| 5297 | A critical comparison of echocardiographic measurements used for optimizing cardiac resynchronization therapy: stroke distance is best | Inappropriate study design |
| 5298 | Epidemiologic study of type 2 diabetes in Taiwan | Inappropriate population |
| 5299 | A global clinical measure of fitness and frailty in elderly people | Inappropriate population |
| 5300 | Excitability changes in intracortical neural circuits induced by differentially controlled walking patterns | Inappropriate study design |
| 5301 | Diet quality as a predictor of cardiometabolic disease-free life expectancy: the Whitehall II cohort study | Inappropriate study design |
| 5302 | Chronic hepatitis B virus infection and risk of chronic kidney disease: a population-based prospective cohort study of 0.5 million Chinese adults | Inappropriate outcome |
| 5303 | Characterization of the work-time relationship during cross-country ski ergometry | Inappropriate study design |
| 5304 | Interrelations between power, force production and energy metabolism in maximal leg work using a modified rowing ergometer | Inappropriate population |
| 5305 | Bioenergetic characteristics of swimmers determined during an arm-ergometer test and during swimming | Inappropriate population |
| 5306 | Rest and exercise performance of the Medtronic Advantage bileaflet valve in the aortic position | Inappropriate study design |
| 5307 | The influence of dual-task conditions on movement in young adults with and without Down syndrome | Inappropriate population |
| 5308 | Implicit associations and compensatory health beliefs in smokers: exploring their role for behaviour and their change through warning labels | Inappropriate study design |
| 5309 | Characteristics of physical measurement consent in a population-based survey of older adults | Inappropriate study design |
| 5310 | Serum Angiopoietin-Like Protein 2 Is a Novel Risk Factor for Cardiovascular Disease in the Community: The Hisayama Study | Inappropriate study design |
| 5311 | Chronic fatigue syndrome: illness severity, sedentary lifestyle, blood volume and evidence of diminished cardiac function | Inappropriate population |
| 5312 | Adverse systemic arterial function in patients with selenium deficiency | Inappropriate study design |
| 5313 | Decreased muscle mass in Korean subjects with intracranial arterial stenosis: The Kangbuk Samsung Health Study | Inappropriate population |
| 5314 | Fear of falling and its predictors among community-living older adults in Korea | Inappropriate population |
| 5315 | Left ventricular dynamics and plasma catecholamines during isometric exercise in patients following cardiac transplantation | Inappropriate study design |
| 5316 | Mitochondrial dysfunction in Gulf War illness revealed by 31Phosphorus Magnetic Resonance Spectroscopy: a case-control study | Inappropriate study design |
| 5317 | Comparison of energy supplements during prolonged exercise for maintenance of cardiac function: carbohydrate only versus carbohydrate plus whey or casein hydrolysate | Inappropriate outcome |
| 5318 | Self-perception of health status, mental health and quality of life among adults with diabetes residing in a metropolitan area | Inappropriate population |
| 5319 | Pilot Randomised Controlled Trial of a Web-Based Intervention to Promote Healthy Eating, Physical Activity and Meaningful Social Connections Compared with Usual Care Control in People of Retirement Age Recruited from Workplaces | Inappropriate study design |
| 5320 | Vitamin D and intact PTH status in patients with hip fracture | Inappropriate study design |
| 5321 | Therapeutic Yoga: Symptom Management for Multiple Sclerosis | Inappropriate study design |
| 5322 | Centrifuge training increases presyncopal orthostatic tolerance in ambulatory men | Inappropriate study design |
| 5323 | Association between low plasma levels of ophthalmic timolol and haemodynamics in glaucoma patients | Inappropriate outcome |
| 5324 | Effects of oral nisoldipine in ischemic heart disease. A radioisotopic study | Inappropriate population |
| 5325 | Which arterial and cardiac parameters best predict left ventricular mass? | Inappropriate population |
| 5326 | Performance changes in world-class kayakers following two different training periodization models | Inappropriate population |
| 5327 | Diastolic aortic pressure rise during percutaneous transluminal coronary angioplasty: an index of left ventricular systolic dysfunction | Inappropriate study design |
| 5328 | Psychological distress, caregiving, and metabolic variables | Inappropriate population |
| 5329 | Oral modified release morphine for breathlessness in chronic heart failure: a randomized placebo-controlled trial | Inappropriate study design |
| 5330 | Association Between Protein Intake and Mortality in Hypertensive Patients Without Chronic Kidney Disease in the OLD-HTA Cohort | Inappropriate study design |
| 5331 | Improved neurobehavioral functioning in emphysema patients following medical therapy | Inappropriate study design |

| | | |
|---|---|---|
| 5332 | Impact of Diabetes Mellitus on Human Erythrocytes: Atomic Force Microscopy and Spectral Investigations | Inappropriate population |
| 5333 | Impact of tachycardia and sympathetic stimulation by cold pressor test on cardiac diastology and arterial function in elderly females | Inappropriate study design |
| 5334 | Supplement use and mortality: the SENECA study | Inappropriate study design |
| 5335 | Underappreciated statin-induced myopathic weakness causes disability | Inappropriate study design |
| 5336 | Factors associated with grip strength decline in older adults | Inappropriate study design |
| 5337 | Brain-derived neurotrophic factor and its receptor tropomyosin-related kinase B in the mechanism of action of antidepressant therapies | Inappropriate study design |
| 5338 | Relationship between higher estradiol levels and 9-year mortality in older women: the Invecchiare in Chianti study | Inappropriate study design |
| 5339 | A combination of healthy lifestyle factors is associated with a decreased incidence of chronic kidney disease: a population-based cohort study | Inappropriate population |
| 5340 | Prevalence of mild cognitive impairment in employable patients after acute coronary event in cardiac rehabilitation | Inappropriate study design |
| 5341 | Balloon pulmonary angioplasty improves interventricular dyssynchrony in patients with inoperable chronic thromboembolic pulmonary hypertension: a cardiac MR imaging study | Inappropriate study design |
| 5342 | C-reactive protein, insulin resistance and risk of cardiovascular disease: a population-based study | Inappropriate study design |
| 5343 | The BALANCE Study: clinical benefit and long-term outcome after intracoronary autologous bone marrow cell transplantation in patients with acute myocardial infarction | Inappropriate intervention |
| 5344 | Rigidity and bradykinesia reduce interlimb coordination in Parkinsonian gait | Inappropriate study design |
| 5345 | Association between sarcopenia and depressive mood in urban-dwelling older adults: A cross-sectional study | Inappropriate study design |
| 5346 | Determinants and prognostic value of cardiovascular autonomic function in coronary artery disease patients with and without type 2 diabetes | Inappropriate study design |
| 5347 | Dietary Patterns and Incident Heart Failure in U.S. Adults Without Known Coronary Disease | Inappropriate population |
| 5348 | Relationship between periodontal disease, tooth loss, and carotid artery plaque: the Oral Infections and Vascular Disease Epidemiology Study (INVEST) | Inappropriate population |
| 5349 | Prevalence of overweight and obesity in an Italian Prison and relation with average term of detention: a pilot study | Inappropriate study design |
| 5350 | Nutrition and cognitive deficit in the elderly: a population study | Inappropriate study design |
| 5351 | Risk of Incident Coronary Heart Disease Events in Men Compared to Women by Menopause Type and Race | Inappropriate study design |
| 5352 | Associations between healthy lifestyles and health outcomes among older Koreans | Inappropriate population |
| 5353 | Serum 25-hydroxyvitamin D and pulmonary function in older disabled community-dwelling women | Inappropriate study design |
| 5354 | The cardiac evaluation of liver transplant recipients: a single center's experience | Inappropriate population |
| 5355 | A randomized controlled trial of community health workers using patient stories to support hypertension management: Study protocol | Inappropriate study design |
| 5356 | Ambulation and functional outcome after major lower extremity amputation | Inappropriate study design |
| 5357 | Hemodynamic response to muscle reflex is abnormal in patients with heart failure with preserved ejection fraction | Inappropriate outcome |
| 5358 | Safety assessment of an anti-obesity drug (sibutramine): a retrospective cohort study | Inappropriate population |
| 5359 | Effects of moderate physical training on blood pressure variability and hemodynamic pattern in mildly hypertensive subjects | Inappropriate study design |
| 5360 | Hemodynamic and autonomic changes induced by Ironman: prediction of competition time by blood pressure variability | Inappropriate population |
| 5361 | Rehabilitation Interventions for Older Individuals With Cognitive Impairment Post-Hip Fracture: A Systematic Review | Inappropriate population |
| 5362 | Stress response pattern in obesity and systemic hypertension | Inappropriate study design |
| 5363 | Cross-sectional study examining the association between metabolic syndrome and cognitive function among the oldest old | Inappropriate study design |
| 5364 | The effect of cognitive-motor dual-task training on cognitive function and plasma amyloid β peptide 42/40 ratio in healthy elderly persons: a randomized controlled trial | Inappropriate study design |

| | | |
|---|---|---|
| 5365 | Functional Changes after Recombinant Human Growth Hormone Replacement in Patients with Chronic Traumatic Brain Injury and Abnormal Growth Hormone Secretion | Inappropriate study design |
| 5366 | Unbalanced risk factors, could compromise the effectiveness of physical training in patients with intermittent claudication? | Inappropriate study design |
| 5367 | Long-term hemodynamic benefit of atrial synchronization with A2A2D or A2A2T pacing in sinus node syndrome after orthotopic heart transplantation | Inappropriate study design |
| 5368 | Crouch gait can be an effective form of forced-use/no constraint exercise for the paretic lower limb in stroke | Inappropriate study design |
| 5369 | Caregiver-mediated exercises for improving outcomes after stroke | Inappropriate population |
| 5370 | Cardiovascular safety of exenatide BID: an integrated analysis from controlled clinical trials in participants with type 2 diabetes | Inappropriate study design |
| 5371 | Risk factors and clinical outcomes in chronic coronary and peripheral artery disease: An analysis of the randomized, double-blind COMPASS trial | Inappropriate outcome |
| 5372 | The prevalence and impact of self-reported hip fracture in elderly community-dwelling women: the Women's Health and Aging Study | Inappropriate study design |
| 5373 | Phenotypic and genetic relationships between kidney function and carotid intima-media thickness in Koreans: the Healthy Twin Study | Inappropriate population |
| 5374 | Muscle strength in the Mataró aging study participants and its relationship to successful aging | Inappropriate study design |
| 5375 | Feasibility of combining multi-channel functional neuromuscular stimulation with weight-supported treadmill training | Inappropriate population |
| 5376 | Frailty and safety: the example of diabetes | Inappropriate outcome |
| 5377 | Weight gain after 20 years of age is associated with prevalence of chronic kidney disease | Inappropriate population |
| 5378 | Multicenter trial of dynamic cardiomyoplasty for chronic heart failure. The American Cardiomyoplasty Group | Inappropriate study design |
| 5379 | Psychosocial stress moderates the relationship of cancer history with natural killer cell activity | Inappropriate study design |
| 5380 | Association between life space and risk of mortality in advanced age | Inappropriate population |
| 5381 | Determinants of maximal oxygen uptake in moderate acute hypoxia in endurance athletes | Inappropriate population |
| 5382 | Hemodynamic effects of calcium antagonists in cardiac patients | Inappropriate study design |
| 5383 | Isolated systolic hypertension in older patients | Inappropriate study design |
| 5384 | Relationship between neuropsychological tests and driver's license renewal tests in Parkinson's disease | Inappropriate population |
| 5385 | Impaired oral health status on admission is associated with poor clinical outcomes in post-acute inpatients: A prospective cohort study | Inappropriate study design |
| 5386 | Myocardial and Cardiocirculatory Reserve in Asymptomatic Aortic Stenosis and Preserved Ejection Fraction | Inappropriate population |
| 5387 | Behavioral risk factors associated with overweight and obesity among older adults: the 2005 National Health Interview Survey | Inappropriate study design |
| 5388 | Pulsed Doppler tissue imaging in endurance athletes: relation between left ventricular preload and myocardial regional diastolic function | Inappropriate study design |
| 5389 | All-cause mortality and risk factors in a cohort of retired military male veterans, Xi'an, China: an 18-year follow up study | Inappropriate study design |
| 5390 | Asymptomatic atrial fibrillation: clinical correlates, management, and outcomes in the EORP-AF Pilot General Registry | Inappropriate study design |
| 5391 | Healthy behaviors, risk factor control and awareness of chronic kidney disease | Inappropriate outcome |
| 5392 | Association of serum uric acid and cardiovascular disease in healthy adults | Inappropriate population |
| 5393 | Temporary loss of cardiac autonomic innervation after the maze procedure | Inappropriate outcome |
| 5394 | An assessment of non-communicable diseases, diabetes, and related risk factors in the Republic of the Marshall Islands, Kwajelein Atoll, Ebeye Island: a systems perspective | Inappropriate study design |
| 5395 | Why does lung function predict mortality? Results from the Whitehall II Cohort Study | Inappropriate population |
| 5396 | Late-life depressive symptoms: prediction models of change | Inappropriate population |
| 5397 | Factors associated with impairment in gait speed in older people with clinically normal gait. A cross-sectional study | Inappropriate study design |
| 5398 | Cortical reorganization induced by body weight-supported treadmill training in patients with hemiparesis of different stroke durations | Inappropriate study design |
| 5399 | Enhanced rehabilitation of gait after stroke: a case report of a therapeutic approach using multichannel functional electrical stimulation | Inappropriate study design |

| | | |
|---|---|---|
| 5400 | An improved method of right ventricular gated equilibrium blood pool radionuclide ventriculography | Inappropriate population |
| 5401 | Investigating the interference pattern of dual tasks using serial decomposition | Inappropriate study design |
| 5402 | Dietary nitrate modulates cerebral blood flow parameters and cognitive performance in humans: A double-blind, placebo-controlled, crossover investigation | Inappropriate population |
| 5403 | Callosal hyperintensities and gait speed gain from two types of mobility interventions in older adults | Inappropriate study design |
| 5404 | WHO guidelines for a healthy diet and mortality from cardiovascular disease in European and American elderly: the CHANCES project | Inappropriate study design |
| 5405 | Pharmacological cardioversion for atrial fibrillation and flutter | Inappropriate population |
| 5406 | Rhythm control versus rate control in patients with persistent atrial fibrillation. Results of the HOT CAFE Polish Study | Inappropriate population |
| 5407 | Serum levels of irisin predict short-term outcomes in ischemic stroke | Inappropriate intervention |
| 5408 | WITHDRAWN: Pharmacological cardioversion for atrial fibrillation and flutter | Inappropriate study design |
| 5409 | A low plasma volume in formerly preeclamptic women predisposes to the recurrence of hypertensive complications in the next pregnancy | Inappropriate study design |
| 5410 | The American Heart Association Ideal Cardiovascular Health and Incident Type 2 Diabetes Mellitus Among Blacks: The Jackson Heart Study | Inappropriate population |
| 5411 | The global impact of noncommunicable diseases: estimates and projections | Inappropriate population |
| 5412 | Gated single photon emission tomography MIBI stress perfusion imaging for assessing cardiac output and index in obese and non-obese patients | Inappropriate study design |
| 5413 | A blinded, randomized, placebo-controlled trial of percutaneous laser myocardial revascularization to improve angina symptoms in patients with severe coronary disease | Inappropriate population |
| 5414 | Practice-based skill acquisition of pushrim-activated power-assisted wheelchair propulsion versus regular handrim propulsion in novices | Inappropriate study design |
| 5415 | Health related quality of life in coronary patients and its association with their cardiovascular risk profile: results from the EUROASPIRE III survey | Inappropriate population |
| 5416 | Reference values for handgrip strength and their association with intrinsic capacity domains among older adults | Inappropriate outcome |
| 5417 | Onset of electrical stimulation leg cycling in individuals with paraplegia | Inappropriate study design |
| 5418 | Heart Failure with Preserved Ejection Fraction: Diagnosis and Management | Inappropriate population |
| 5419 | Erythropoietin in heart failure | Inappropriate population |
| 5420 | Multicentre, randomised controlled trial of PDSAFE, a physiotherapist-delivered fall prevention programme for people with Parkinson's | Inappropriate study design |
| 5421 | Surveillance of certain health behaviors and conditions among states and selected local areas --- Behavioral Risk Factor Surveillance System, United States, 2009 | Inappropriate population |
| 5422 | Calcium antagonists in the elderly. A risk-benefit analysis | Inappropriate population |
| 5423 | Inflammation biomarkers and risk of all-cause mortality in the Reasons for Geographic And Racial Differences in Stroke cohort | Inappropriate study design |
| 5424 | First-in-man study of Heartech® percutaneous left ventricular partitioning device for treatment of heart failure postmyocardial infarction | Inappropriate study design |
| 5425 | The marked decline in coronary heart disease mortality rates in the United States, 1968-1981; summary of findings and possible explanations | Inappropriate study design |
| 5426 | Randomised controlled trial to evaluate the efficacy and usability of a computerised phone-based lifestyle coaching system for primary and secondary prevention of stroke | Inappropriate population |
| 5427 | Differences in hemodynamic response to metaboreflex activation between obese patients with metabolic syndrome and healthy subjects with obese phenotype | Inappropriate population |
| 5428 | The relationships between casual and ambulatory blood pressure measurements and central hemodynamics in essential human hypertension | Inappropriate study design |
| 5429 | Association of vitamin D deficiency with cognitive impairment in older women: cross-sectional study | Inappropriate outcome |
| 5430 | Social inequalities in multimorbidity, frailty, disability, and transitions to mortality: a 24-year follow-up of the Whitehall II cohort study | Inappropriate intervention |
| 5431 | Immediate haemodynamic effects of a novel partial agonist, beta 1-adrenoceptor blocking drug ICI 141,292 after intravenous administration to healthy young volunteers and patients with ischaemic heart disease | Inappropriate study design |
| 5432 | Studies with felodipine in congestive heart failure | Inappropriate study design |
| 5433 | Regression of left ventricular hypertrophy in hypertensive heart transplant recipients treated with enalapril, furosemide, and verapamil | Inappropriate study design |

| | | |
|---|---|---|
| 5434 | The impact of the Polish national Programme of Cardiovascular Disease Prevention on the quality of primary cardiovascular disease prevention in clinical practice | Inappropriate population |
| 5435 | Heart disease and stroke statistics--2011 update: a report from the American Heart Association | Inappropriate study design |
| 5436 | Waist circumference and neutrophil gelatinase-associated lipocalin in late-life depression | Inappropriate study design |
| 5437 | Gender differences in vascular compliance in young, healthy subjects assessed by pulse contour analysis | Inappropriate population |
| 5438 | Addressing rural health disparities through policy change in the stroke belt | Inappropriate study design |
| 5439 | Physical and psychological burden of chronic kidney disease among older adults | Inappropriate study design |
| 5440 | Chronic disease and lifestyle factors associated with change in sleep duration among older adults in the Singapore Chinese Health Study | Inappropriate study design |
| 5441 | EPIC-Heart: the cardiovascular component of a prospective study of nutritional, lifestyle and biological factors in 520,000 middle-aged participants from 10 European countries | Inappropriate study design |
| 5442 | Patients with knee osteoarthritis undergoing total knee arthroplasty have a lower risk of subsequent severe cardiovascular events: propensity score and instrumental variable analysis | Inappropriate study design |
| 5443 | Interaction of BDNF rs6265 variants and energy and protein intake in the risk for glucose intolerance and type 2 diabetes in middle-aged adults | Inappropriate study design |
| 5444 | Chronic obstructive pulmonary disease as an independent risk factor for cardiovascular morbidity | Inappropriate study design |
| 5445 | Musculoskeletal manifestations of Fabry disease: A retrospective study | Inappropriate population |
| 5446 | Beta-adrenergic blockade and training in healthy men--effects on central circulation | Inappropriate study design |
| 5447 | Regurgitant volume informs rate of progressive cardiac dysfunction in asymptomatic patients with chronic aortic or mitral regurgitation | Inappropriate population |
| 5448 | Severity Weighting of Postoperative Adverse Events in Orthopedic Surgery | Inappropriate study design |
| 5449 | Prevalence and natural history of heart disease in adults with primary mitochondrial respiratory chain disease | Inappropriate population |
| 5450 | Extent of myocardial viability predicts response to biventricular pacing in ischemic cardiomyopathy | Inappropriate study design |
| 5451 | Staphylococcus aureus CC30 Lineage and Absence of sed,j,r-Harboring Plasmid Predict Embolism in Infective Endocarditis | Inappropriate population |
| 5452 | Response of patients with coronary artery disease stratified by ejection fraction following short-term training | Inappropriate outcome |
| 5453 | The relationship between body mass index and number of days spent in hospital in Scotland | Inappropriate study design |
| 5454 | RV adaptation to increased afterload in congenital heart disease and pulmonary hypertension | Inappropriate study design |
| 5455 | Health-related quality of life of type 2 diabetes patients hospitalized for a diabetes-related complication | Inappropriate study design |
| 5456 | A practical approach to assess depression risk and to guide risk reduction strategies in later life | Inappropriate population |
| 5457 | Transmyocardial holmium laser revascularization: feasibility of a thoracoscopic approach | Inappropriate study design |
| 5458 | Sleep-time blood pressure: prognostic value and relevance as a therapeutic target for cardiovascular risk reduction | Inappropriate study design |
| 5459 | Upper limb muscle activation during sports video gaming of persons with spinal cord injury | Inappropriate population |
| 5460 | Feeling Better at This Age? Investigating Three Explanations for Self-Rated Health Improvements Among the Oldest-Old | Inappropriate study design |
| 5461 | Circulating mononuclear superoxide production and inflammatory markers for long-term prognosis in patients with cardiac syndrome X | Inappropriate study design |
| 5462 | Prehospital and Intrahospital Temporal Intervals in Patients Requiring Emergent Trauma Craniotomy. A 6-Year Observational Study in a Level 1 Trauma Center | Inappropriate study design |
| 5463 | Homeostasis model assessment of insulin resistance in relation to the incidence of cardiovascular disease: the San Antonio Heart Study | Inappropriate study design |
| 5464 | Ambulatory blood pressure thresholds for diagnosis of hypertension in patients with and without type 2 diabetes based on cardiovascular outcomes | Inappropriate population |
| 5465 | Contracts, covenants and advance care planning: an empirical study of the moral obligations of patient and proxy | Inappropriate population |
| 5466 | Cardiovascular and autonomic responses to physiological stressors before and after six hours of water immersion | Inappropriate intervention |

| | | |
|---|---|---|
| 5467 | Impact of Low Cardiovascular Risk Profiles on Geriatric Outcomes: Evidence From 421,000 Participants in Two Cohorts | Inappropriate population |
| 5468 | Chronic physical activity mitigates cerebral hypoperfusion during central hypovolemia in elderly humans | Inappropriate study design |
| 5469 | Coronary Artery Calcification and Risk of Cardiovascular Disease and Death Among Patients With Chronic Kidney Disease | Inappropriate study design |
| 5470 | Depression as a Clinical Determinant of Dependence and Low Quality of Life in Elderly Patients with Cardiovascular Disease | Inappropriate study design |
| 5471 | Predictors of discharge destination in a geriatric population after undergoing rehabilitation | Inappropriate population |
| 5472 | Probability of Heat Intolerance: Standardized Interpretation of Heat-Tolerance Testing Results Versus Specialist Judgment | Inappropriate study design |
| 5473 | Are all people with diabetes and cardiovascular risk factors or microvascular complications at very high risk? Findings from the Risk and Prevention Study | Inappropriate study design |
| 5474 | The role of conventional and novel mechanisms in explaining increased risk of cardiovascular events in offspring with positive parental history | Inappropriate study design |
| 5475 | Relation of leptin to left ventricular hypertrophy (from the Multi-Ethnic Study of Atherosclerosis) | Inappropriate population |
| 5476 | Inflammatory potential of diet is associated with cognitive function in an older adult Korean population | Inappropriate study design |
| 5477 | Ethnic differences in the prognostic value of stress technetium-99m tetrofosmin gated single-photon emission computed tomography myocardial perfusion imaging | Inappropriate population |
| 5478 | Calcified amorphous tumor of the heart with mitral annular calcification: a case report | Inappropriate study design |
| 5479 | Age-associated cardiovascular changes in health: impact on cardiovascular disease in older persons | Inappropriate study design |
| 5480 | Effects of oxygen fraction in inspired air on force production and electromyogram activity during ergometer rowing | Inappropriate study design |
| 5481 | Unexpected sudden death due to recreational swimming and diving in men in Croatia in a 14-year period | Inappropriate study design |
| 5482 | Endothelial Function Assessed by Digital Volume Plethysmography Predicts the Development and Progression of Type 2 Diabetes Mellitus | Inappropriate study design |
| 5483 | Effect of eight weeks of endurance exercise training on right and left ventricular volume and mass in untrained obese subjects: a longitudinal MRI study | Inappropriate study design |
| 5484 | Physical disability in older adults: a physiological approach. Cardiovascular Health Study Research Group | Inappropriate study design |
| 5485 | Hemodynamic effects of long-term cardiac resynchronization therapy: analysis by pressure-volume loops | Inappropriate outcome |
| 5486 | Necessity of Removing American Football Uniforms From Humans With Hyperthermia Before Cold-Water Immersion | Inappropriate intervention |
| 5487 | The relationships between body composition characteristics and cognitive functioning in a population-based sample of older British men | Inappropriate outcome |
| 5488 | Using impairment and cognitions to predict walking in osteoarthritis: A series of n-of-1 studies with an individually tailored, data-driven intervention | Inappropriate population |
| 5489 | Intima-media thickness and age of first depressive episode | Inappropriate study design |
| 5490 | Development and Validation of Novel Dietary and Lifestyle Inflammation Scores | Inappropriate study design |
| 5491 | Health-Related Quality of Life of Hypertension Patients: A Population-Based Cross-Sectional Study in Chongqing, China | Inappropriate study design |
| 5492 | Changes to oral anticoagulant therapy and risk of death over a 3-year follow-up of a contemporary cohort of European patients with atrial fibrillation final report of the EURObservational Research Programme on Atrial Fibrillation (EORP-AF) pilot general registry | Inappropriate study design |
| 5493 | 25-Hydroxyvitamin D deficiency is independently associated with cardiovascular disease in the Third National Health and Nutrition Examination Survey | Inappropriate population |
| 5494 | Health literacy is associated with health behaviour and self-reported health: A large population-based study in individuals with cardiovascular disease | Inappropriate intervention |
| 5495 | Gender-based differences in the management and prognosis of acute coronary syndrome in Korea | Inappropriate study design |
| 5496 | Long-term follow-up after percutaneous septal ablation in hypertrophic obstructive cardiomyopathy | Inappropriate population |
| 5497 | Anticytomegalovirus antibody titres are not associated with caregiving burden in younger caregivers | Inappropriate study design |

| | | |
|---|---|---|
| 5498 | Identification of Potential Type II Diabetes in a Chinese Population with a Sensitive Decision Tree Approach | Inappropriate study design |
| 5499 | State-level income inequality and the odds for meeting fruit and vegetable recommendations among US adults | Inappropriate population |
| 5500 | Depression and health service utilization from age 70 to 85: the Jerusalem Longitudinal Study | Inappropriate study design |
| 5501 | All cause and disease specific mortality in patients with knee or hip osteoarthritis: population based cohort study | Inappropriate population |
| 5502 | Intentional coverage of the left subclavian artery during endovascular repair of traumatic descending thoracic aortic transection | Inappropriate outcome |
| 5503 | Risk factors for injurious falls leading to hospitalization or death in a cohort of 19,500 adults | Inappropriate study design |
| 5504 | Manipal lifestyle modification score to predict major adverse cardiac events in postcoronary angioplasty patients | Inappropriate study design |
| 5505 | Frailty Status and Outcomes After Transcatheter Aortic Valve Implantation | Inappropriate study design |
| 5506 | Prospective association of a genetic risk score and lifestyle intervention with cardiovascular morbidity and mortality among individuals with type 2 diabetes: the Look AHEAD randomised controlled trial | Inappropriate population |
| 5507 | Genetic variation and decreased risk for obesity in the Atherosclerosis Risk in Communities Study | Inappropriate study design |
| 5508 | Treatment of functional mitral valve regurgitation with the permanent percutaneous transvenous mitral annuloplasty system: results of the multicenter international Percutaneous Transvenous Mitral Annuloplasty System to Reduce Mitral Valve Regurgitation in Patients with Heart Failure trial | Inappropriate population |
| 5509 | Association between body mass index and mortality in the Korean elderly: A nationwide cohort study | Inappropriate study design |
| 5510 | Design strategies to improve patient motivation during robot-aided rehabilitation | Inappropriate study design |
| 5511 | Obesity, diabetes, and risk of cardiovascular disease in the elderly | Inappropriate study design |
| 5512 | Relationship of serum C3 to fasting insulin, risk factors and previous ischaemic events in middle-aged men | Inappropriate study design |
| 5513 | Precise estimation of human corticospinal excitability associated with the levels of motor imagery-related EEG desynchronization extracted by a locked-in amplifier algorithm | Inappropriate intervention |
| 5514 | The influence of adrenergic stimulation on sex differences in left ventricular twist mechanics | Inappropriate study design |
| 5515 | Endoluminal repair of abdominal aortic aneurysm: the Middlemore Hospital experience | Inappropriate study design |
| 5516 | Poor self-rated health is significantly associated with elevated C-reactive protein levels in women, but not in men, in the Japanese general population | Inappropriate outcome |
| 5517 | Statins improve myocardial perfusion in metabolic syndrome patients who have perfusion defects on myocardial perfusion imaging and angiographically normal coronary arteries | Inappropriate population |
| 5518 | Volumetric and functional assessment of the left atrium in young competitive athletes without left ventricular hypertrophy: the MAGYAR-Sport Study | Inappropriate study design |
| 5519 | Influence of water immersion, water gymnastics and swimming on cardiac output in patients with heart failure | Inappropriate study design |
| 5520 | Associations of metabolic syndrome with inflammation in CKD: results From the Third National Health and Nutrition Examination Survey (NHANES III) | Inappropriate study design |
| 5521 | Baseline health-related quality of life and 10-year all-cause mortality among 1739 Chinese adults | Inappropriate study design |
| 5522 | The calcium channel blocker felodipine attenuates the positive hemodynamic effects of the beta-blocker metoprolol in severe dilated cardiomyopathy--a prospective, randomized, double-blind and placebo-controlled study with invasive hemodynamic assessment | Inappropriate population |
| 5523 | Aortic stiffness correlates with an increased extracellular matrix turnover in patients with dilated cardiomyopathy | Inappropriate population |
| 5524 | Bilateral internal thoracic artery T grafting for coronary artery revascularization. Angiographic assessment and mid-term outcome | Inappropriate population |
| 5525 | Evaluation of a temperate environment test to predict heat tolerance | Inappropriate outcome |
| 5526 | Increased risk and worse prognosis of myocardial infarction in patients with prior hospitalization for epilepsy--the Stockholm Heart Epidemiology Program | Inappropriate population |
| 5527 | Long-term (3-month) effects of a new beta-blocker (nebivolol) on cardiac performance in dilated cardiomyopathy | Inappropriate population |
| 5528 | Risk factors for falls in a longitudinal cohort study of Saudi postmenopausal women: the Center of Excellence for Osteoporosis Research Study | Inappropriate population |

| | | |
|---|---|---|
| 5529 | Influence of Lifestyle on Incident Cardiovascular Disease and Mortality in Patients With Diabetes Mellitus | Inappropriate study design |
| 5530 | Determining the validity of the AMA guide: A historical cohort analysis of the assessment of driving related skills and crash rate among older drivers | Inappropriate outcome |
| 5531 | Final-year-of life pacemaker recipients | Inappropriate study design |
| 5532 | Normative Values of Muscle Power using Force Plate Jump Tests in Men Aged 77-101 Years: The Osteoporotic Fractures in Men (MrOS) Study | Inappropriate study design |
| 5533 | Randomized controlled trial to investigate influence of the fluid challenge on duration of hospital stay and perioperative morbidity in patients with hip fractures | Inappropriate outcome |
| 5534 | The 2010 Canadian Hypertension Education Program recommendations for the management of hypertension: part 2 - therapy | Inappropriate population |
| 5535 | Exercise- and strategy-based physiotherapy-delivered intervention for preventing repeat falls in people with Parkinson's: the PDSAFE RCT | Inappropriate study design |
| 5536 | The effect of routine, early invasive management on outcome for elderly patients with non-ST-segment elevation acute coronary syndromes | Inappropriate study design |
| 5537 | Cross-sectional association between serum vitamin D concentration and walking speed measured at usual and fast pace among older women: the EPIDOS study | Inappropriate population |
| 5538 | The efficacy and safety of a chest pain protocol for short stay unit patients: A one year follow-up | Inappropriate population |
| 5539 | Preventable clinical and psychosocial factors predicted two out of three recurrent cardiovascular events in a coronary population | Inappropriate population |
| 5540 | Effectiveness of robot-assisted training added to conventional rehabilitation in patients with humeral fracture early after surgical treatment: protocol of a randomised, controlled, multicentre trial | Inappropriate population |
| 5541 | Transcatheter Valve Repair for Patients With Mitral Regurgitation: 30-Day Results of the CLASP Study | Inappropriate population |
| 5542 | Use of carbonated water in reduction of adjacent gastric activity in 456 consecutive technetium-99m myocardial perfusion imaging studies | Inappropriate outcome |
| 5543 | Effect of single-drug therapy on reduction of left atrial size in mild to moderate hypertension: comparison of six antihypertensive agents | Inappropriate population |
| 5544 | Surveillance for Certain Health Behaviors, Chronic Diseases, and Conditions, Access to Health Care, and Use of Preventive Health Services Among States and Selected Local Areas - Behavioral Risk Factor Surveillance System, United States, 2012 | Inappropriate study design |
| 5545 | Study rationale and design of the CIMT trial: the Copenhagen Insulin and Metformin Therapy trial | Inappropriate study design |
| 5546 | Detection of silent myocardial ischemia in asymptomatic patients with diabetes: results of a randomized trial and meta-analysis assessing the effectiveness of systematic screening | Inappropriate study design |
| 5547 | Surveillance for certain health behaviors among States and selected local areas - United States, 2010 | Inappropriate study design |
| 5548 | Obesity and loss of disease-free years owing to major non-communicable diseases: a multicohort study | Inappropriate study design |
| 5549 | Increased consumption of fruit and vegetables for the primary prevention of cardiovascular diseases | Inappropriate population |
| 5550 | Three weekly injections (TWI) of low-dose growth hormone (GH) restore low normal circulating IGF-I concentrations and reverse cardiac abnormalities associated with adult onset GH deficiency (GHD) | Inappropriate study design |
| 5551 | Modifiable risk factors, cardiovascular disease, and mortality in 155 722 individuals from 21 high-income, middle-income, and low-income countries (PURE): a prospective cohort study | Inappropriate population |
| 5552 | Determinants of cigarette smoking status in a national cohort of black and white adult ever smokers in the USA: a cross-sectional analysis of the REGARDS study | Inappropriate study design |
| 5553 | Sleep duration and mortality: a prospective study of 113 138 middle-aged and elderly Chinese men and women | Inappropriate population |
| 5554 | Association of Change in Cardiovascular Risk Factors With Incident Cardiovascular Events | Inappropriate population |
| 5555 | Description of 1,108 older patients referred by their physician to the "Geriatric Frailty Clinic (G.F.C) for Assessment of Frailty and Prevention of Disability" at the gerontopole | Inappropriate population |
| 5556 | Surgical versus non-surgical treatment for lumbar spinal stenosis | Inappropriate study design |
| 5557 | Cilostazol for intermittent claudication | Inappropriate study design |
| 5558 | Physiotherapy for pain and disability in adults with complex regional pain syndrome (CRPS) types I and II | Inappropriate study design |

| | | |
|---|---|---|
| 5559 | Interactive computer-based interventions for weight loss or weight maintenance in overweight or obese people | Inappropriate study design |
| 5560 | Cholesterol and atherosclerosis: a contemporary perspective | Inappropriate study design |
| 5561 | Effectiveness of polypill for primary and secondary prevention of cardiovascular diseases (PolyIran): a pragmatic, cluster-randomised trial | Inappropriate study design |
| 5562 | Effects of Tai Chi Exercise on Cognition and Serum Biomarkers of Individuals With MCI | Inappropriate study design |
| 5563 | Effectiveness of Tai Chi to Improve Cognitive Function in Older Adults With Mild Cognitive Impairment | Inappropriate study design |
| 5564 | Cognitive Activation Therapy for MCI: a Randomized Control Study | Inappropriate study design |
| 5565 | Pioglitazone Or Exercise to Treat Mild Cognitive Impairment (MCI) | Inappropriate study design |
| 5566 | SYNchronizing Exercises, Remedies in GaIt and Cognition | Inappropriate study design |
| 5567 | Effectiveness of Walking Exercise in Improving Cognitive Function in Older Adults With Mild Cognitive Impairment | Inappropriate intervention |
| 5568 | Is transcranial direct current stimulation (tDCS) an effective adjunct to cognitive training for older adults presenting with mild cognitive impairment (MCI)? | Inappropriate study design |
| 5569 | T and B cell subsets differentially correlate with amyloid deposition and neurocognitive function in patients with amnestic mild cognitive impairment after one year of physical activity | Inappropriate study design |
| 5570 | THE EFFECTS OF A MODERATE-INTENSITY AEROBIC STEPPING EXERCISE PROGRAM ON COGNITIVE AND PSYCHOLOGICAL OUTCOMES IN CHINESE OLDER ADULTS WITH MILD COGNITIVE IMPAIRMENT: a RANDOMIZED CONTROLLED TRIAL | Inappropriate study design |
| 5571 | Multi-component Exercise on Mild Cognitive Impairment In Elderly Population | Inappropriate study design |
| 5572 | Effects of Square-stepping Exercise on Frailty and Cognitive Function in Elderly With Frailty and MCI | Inappropriate study design |
| 5573 | Aerobic exercise effectsoncognition in at-risk adults: role of insulin sensitivity and APOE-epsilon4 | Inappropriate population |
| 5574 | Effect of Exercise on Cognition, Conditioning, Muscle Endurance, and Balance in Older Adults With Mild Cognitive Impairment: a Randomized Controlled Trial | Inappropriate population |
| 5575 | Effects of aerobic exercise on cognition and biomarkers in adults with a double-hit risk for dementia: mild cognitive impairment and prediabetes | Inappropriate study design |
| 5576 | THE EFFECT OF AEROBIC AND GAME BASED EXERCISES ON COGNITIVE FUNCTIONS IN DEMENTIA | Inappropriate study design |
| 5577 | The Study of Mental and Resistance Training (SMART) Study-Resistance Training and/or Cognitive Training in Mild Cognitive Impairment: a Randomized, Double-Blind, Double-Sham Controlled Trial | Inappropriate population |
| 5578 | Effects of Multicomponent Exercise on Cognitive Performance and Fall Risk in Older Women With Mild Cognitive Impairment | Inappropriate intervention |
| 5579 | Improving brain health in at risk populations with aerobic exercise | Inappropriate study design |
| 5580 | Physical activity for older Australians with mild cognitive impairment or subjective cognitive decline â€“ A narrative review to support guideline development | Inappropriate study design |
| 5581 | Maintaining Independence and Quality of Life in Aging Adults Through Targeted Cognitive Training | Inappropriate study design |
| 5582 | Mind-motor Exercise in Older Adults With Type 2 Diabetes and Self-reported Cognitive Complaints | Inappropriate study design |
| 5583 | Exercise and brain health in older adults with mild cognitive impairment | Inappropriate study design |
| 5584 | The ACT Trial: effects of Combined Aerobic Exercise and Cognitive Training in MCI | Inappropriate study design |
| 5585 | Effects of aerobic, resistance and combination training on cognitive function in older adults: study protocol for a randomized controlled trial | Inappropriate study design |
| 5586 | The effects of exercise on memory performance in older adults with probable mild cognitive impairment: a 6-month randomized controlled trial | Inappropriate intervention |
| 5587 | The StayFitLonger Study: an Innovative Computerized Home-based Training to Foster Independent Life at Home | Inappropriate study design |
| 5588 | The ADAS-cog-exec: a novel cognitive composite outcome to assess therapeutic effects of exercise in the exert trial for adults with mci | Inappropriate study design |
| 5589 | Promotion of the mind through exercise (PROMoTE): a proof-of-concept randomized controlled trial of aerobic exercise training in older adults with vascular cognitive impairment | Inappropriate study design |
| 5590 | Effect of progressive resistance training on motor performance in older adults with mild cognitive impairment | Inappropriate population |

| | | |
|---|---|---|
| 5591 | Impact of a 12-week exercise intervention on non-cognitive outcomes in sedentary elders with cognitive complaints or mild cognitive impairment: findings from the max trial | Inappropriate study design |
| 5592 | Cognitive Effects of Aerobic Exercise for MCI Adults | Inappropriate study design |
| 5593 | Prescribing aerobic exercise to older adults with Alzheimer's disease | Inappropriate intervention |
| 5594 | Exergaming Versus Aerobic Exercises on Mild Cognitive Impairment | Inappropriate study design |
| 5595 | TREATMENT OF MILD COGNITIVE IMPAIRMENT DUE TO ALZHEIMER'S DISEASE OR MILD DEMENTIA OF ALZHEIMER'S TYPE: DESIGN AND RATIONALE OF THE RANDOMIZED PIVOTAL STUDY OF RENEWTM NCP-5 | Inappropriate study design |
| 5596 | Computerized cognitive stimulation and engagement programs in older adults with mild cognitive impairment: comparing feasibility, acceptability, and cognitive and psychosocial effects | Inappropriate population |
| 5597 | Supplementary Material for: effects of Pioglitazone or Exercise in Older Adults with Mild Cognitive Impairment and Insulin Resistance: a Pilot Study | Inappropriate population |
| 5598 | EXCEL: exercise for Cognition and Everyday Living for Seniors With Memory Complaints | Inappropriate study design |
| 5599 | Does Computerized Cognitive Training Improve Executive Functioning in the Older Adult? | Inappropriate study design |
| 5600 | Effects of tACS Combined With Cognitive Training or Aerobic Exercise for Patients With MCI | Inappropriate study design |
| 5601 | U.S. pointer: study design and trial kickoff | Inappropriate study design |
| 5602 | Cognitive training and neuroplasticity in mild cognitive impairment (COG-IT): protocol for a two-site, blinded, randomised, controlled treatment trial | Inappropriate study design |
| 5603 | Exercise training-induced changes on positron emission tomography (PET) with 2-deoxy-2-fluoro-D-glucose (FDG) in mild cognitively impaired older African Americans: gems study | Inappropriate population |
| 5604 | Dual Task Aerobic Exercise for Older Adults With Cognitive Impairment (HM2) | Inappropriate study design |
| 5605 | Effect of Baduanjin exercise on cognitive function in older adults with mild cognitive impairment: study protocol for a randomised controlled trial | Inappropriate intervention |
| 5606 | Effects of multicomponent exercise in older adults with mild cognitive impairment | Inappropriate study design |
| 5607 | The SEcReT Study: supervised Early Resistance Training Following Median Sternotomy: protocol for a Randomised Controlled Pilot Study | Inappropriate study design |
| 5608 | Effects of multi-task exercise program on cognitive and physical function in patients with mild cognitive impairment: a randomized controlled trial | Inappropriate study design |
| 5609 | Exercise Training and Cognitive Function in Kidney Disease | Inappropriate study design |
| 5610 | The efficiency of group cognitive rehabilitation to patients with Parkinson's disease in comparison to relaxation therapy: pilot data | Inappropriate study design |
| 5611 | High intensityaerobic exercise improves performance on computer tests of executive function in adults with mild cognitive impairment: implications for cognitive assessment in clinical trials | Inappropriate study design |
| 5612 | The exert trial: testing a model for effective community-based exercise intervention delivery for adults with MCI | Inappropriate intervention |
| 5613 | Effects of aerobic exercise on cognition and hippocampal volume in Alzheimer's disease: study protocol of a randomized controlled trial (The FIT-AD trial) | Inappropriate study design |
| 5614 | Exercise or vitamin B for lowering homocysteine levels in cognitively impaired adults? | Inappropriate study design |
| 5615 | A physical activity intervention to delay the progression of cerebrovascular disease in older adults with mild memory problems | Inappropriate study design |
| 5616 | Cerebral White Matter Integrity in Amnestic Mild Cognitive Impairment: a 1-Year Randomized Controlled Trial of Aerobic Exercise Training | Inappropriate population |
| 5617 | Cognitive Impairment during High-Intensity Exercise: influence of Cerebral Blood Flow | Inappropriate study design |
| 5618 | Falls in cognitive impairment and dementia: identifying those at risk and developing novel interventions | Inappropriate population |
| 5619 | Impact of aerobic exercise training during chemotherapy on cancer related cognitive impairments in patients suffering from acute myeloid leukemia or myelodysplastic syndrome - Study protocol of a randomized placebo-controlled trial | Inappropriate population |
| 5620 | Improving the Recovery and Outcome Every Day After the ICU | Inappropriate population |
| 5621 | Aerobic exercise reduces phosphorylated tau protein in cerebrospinal fluid in older adults with mild cognitive impairment | Inappropriate population |

| | | |
|---|---|---|
| 5622 | Effects and mechanism of the HECT study (hybrid exercise-cognitive trainings) in mild ischemic stroke with cognitive decline: fMRI for brain plasticity, biomarker and behavioral analysis | Inappropriate study design |
| 5623 | Balance on the Brain: a multi-modal exercise program for people with mild cognitive impairment | Inappropriate study design |
| 5624 | Aerobic Exercise and Cognitive Training in Older Adults | Inappropriate study design |
| 5625 | THE EFFECTS OF PHYSICAL EXERCISE ON COGNITIVE FUNCTION AMONG OLDER ADULTS WITH MILD COGNITIVE IMPAIRMENT: THE MEDITATING ROLE OF DEPRESSIVE SYMPTOMS AND SLEEP QUALITY | Inappropriate study design |
| 5626 | Supplementing intensive targeted computerized cognitive training with social cognitive exercises for people with schizophrenia: an interim report | Inappropriate study design |
| 5627 | The effect of aerobic dance intervention on brain spontaneous activity in older adults with mild cognitive impairment: a resting-state functional MRI study | Inappropriate population |
| 5628 | The impact of aerobic exercise on cognitive functioning and biomarkers of cognitive change in individuals with schizophrenia | Inappropriate study design |
| 5629 | Resting state network adaptation in older adults with MCI in the SMART trial: unique effects of combined cognitive training and physical exercise | Inappropriate study design |
| 5630 | Effect of handball training on cognitive ability in elderly with mild cognitive impairment | Inappropriate study design |
| 5631 | Aerobic exercise training in people with Schizophrenia: neural, cognitive, and functional benefits | Inappropriate study design |
| 5632 | From Antarctica to Alzheimers â€“ Exercise helps to prevent cognitive decline | Inappropriate study design |
| 5633 | Can exercise improve cognitive function in older persons? | Inappropriate intervention |
| 5634 | International Ballroom Dancing Against Neurodegeneration: a Randomized Controlled Trial in Greek Community-Dwelling Elders With Mild Cognitive impairment | Inappropriate study design |
| 5635 | The impact of adherence to multidomain lifestyle intervention on cognition: the finnish geriatric intervention study to prevent cognitive impairment and disability (FINGER) | Inappropriate study design |
| 5636 | Does creatine enhance the effects of cognitive training in older adults with mild cognitive impairment? | Inappropriate study design |
| 5637 | "Simultaneous aerobic exercise and memory training program in older adults with subjective memory impairments": erratum | Inappropriate study design |
| 5638 | Aerobic exercise versus stretching and toning: changes in physical function and mediation of cognitive effects in the max trial | Inappropriate population |
| 5639 | The effectiveness of a moderate-intensity stepping exercise program on cognitive and psychological outcomes of Chinese elders with mild cognitive impairment : a randomized controlled trial | Inappropriate population |
| 5640 | Effects of exercise on plasma nitrite/nitrate levels, and nitric oxide synthase activity in elderly African Americans with mild cognitive impairment | Inappropriate population |
| 5641 | Protocol for Exercise Program in Cancer and Cognition (EPICC): a randomized controlled trial of the effects of aerobic exercise on cognitive function in postmenopausal women with breast cancer receiving aromatase inhibitor therapy | Inappropriate study design |
| 5642 | A potential molecular mechanism linking aerobic exercise to improvement in dentate gyrus-specific cognitive function | Inappropriate study design |
| 5643 | The INDIGO Study - A randomised controlled trial of physical activity with individual goal-setting and volunteer mentors to increase physical activity in insufficiently active older adults at risk of cognitive decline | Inappropriate study design |
| 5644 | INITIAL EXPERIENCE WITH A YOGA INTERVENTION FOR MILD COGNITIVE IMPAIRMENT | Inappropriate study design |
| 5645 | Study of Exercise on Impact of Cognitive Functioning in Multiple Sclerosis Patients | Inappropriate study design |
| 5646 | Comparison of Three Motor-cognitive Training Programs | Inappropriate population |
| 5647 | A Care Model for Elderly Hip-fractured Persons With Cognitive Impairment and Their Family Caregivers | Inappropriate study design |
| 5648 | Exercise Training and Walking Ability After Chronic Stroke | Inappropriate study design |
| 5649 | Tai chi qigong as a means to improve night-time sleep quality among older adults with cognitive impairment: a pilot randomized controlled trial | Inappropriate population |
| 5650 | The Effect of Exercise on Depressive Symptoms in Unmedicated Patients | Inappropriate study design |
| 5651 | SIESTA: sleep Intervention to Enhance Cognitive Status and Reduce Beta Amyloid | Inappropriate population |
| 5652 | Cognitive Effects of Aerobic Exercise for IGT Adults | Inappropriate study design |
| 5653 | Effects of group exercise programmes on quality of life in older adults with mild cognitive impairment: preliminary results from a randomized controlled trial | Inappropriate study design |

| | | |
|---|---|---|
| 5654 | COgnitive and Physical Exercise (COPE) Prehabilitation Pilot Feasibility Study | Inappropriate study design |
| 5655 | Effects of mediterranean diet and resistance training on mild cognitive impairment in patients with type 2 diabetes | Inappropriate study design |
| 5656 | Integrated physical and cognitive training intervention to reverse cognitive frailty among community dwelling older adults | Inappropriate study design |
| 5657 | Aerobic exercise in Alzheimer's disease: the FIT-AD trial | Inappropriate population |
| 5658 | Vascular cognitive impairment and aerobic exercise: a 6-month randomized controlled trial | Inappropriate study design |
| 5659 | The Heart & Mind Study | Inappropriate study design |
| 5660 | Designing a randomized, sham-controlled, parallel-design trial to investigate the effects of a combined exercise priming and transcranial direct current stimulation intervention in mild cognitive impairment and alzheimer's disease | Inappropriate study design |
| 5661 | A randomised controlled trial of aerobic exercise after transient ischaemic attack or minor stroke to prevent cognitive decline: the moveit study | Inappropriate study design |
| 5662 | A Precision Rehabilitation Approach to Counteract Age-Related Cognitive Declines | Inappropriate study design |
| 5663 | DESIGNING A RANDOMIZED, DOUBLE-BLIND, PLACEBO-CONTROLLED PARALLEL GROUP STUDY ON THE EFFICACY AND SAFETY OF N-ACETYLCYSTEINE (NAC) IN PATIENTS WITH MILD VASCULAR COGNITIVE IMPAIRMENT | Inappropriate study design |
| 5664 | Effects of aerobic, resistance, or combined training on cognitive function in older adults: a randomized controlled trial | Inappropriate study design |
| 5665 | HOPE (Healthy Older People Everyday) To Age in Place | Inappropriate population |
| 5666 | Effects of functional tasks exercise on cognitive functions of older adults with mild cognitive impairment: a randomized controlled pilot trial | Inappropriate population |
| 5667 | Prefrontal cortex in aging and effect of aquatic therapy-from ANS and executive function perspective (PART II) | Inappropriate study design |
| 5668 | High-intensity aerobic exercise does not improve cognitive performance in people with multiple sclerosis: a randomised controlled trial | Inappropriate population |
| 5669 | The effects of exercise on cognitive function in older adults with Mild Cognitive Impairment | Inappropriate population |
| 5670 | Social Cognitive Training Improves Measures of Reward Processing in Schizophrenia | Inappropriate population |
| 5671 | Exercise to enhance hippocampal function and memory in older adults: proof of concept findings from a 12-week exercise intervention trial | Inappropriate study design |
| 5672 | Effects of different exercise models on pain, functionality, balance, proprioception and cognition features of patients diagnosed with fibromyalgia | Inappropriate study design |
| 5673 | Influence of aerobic exercise on improvement of topographic memory in patients with traumatic brain injury | Inappropriate study design |
| 5674 | Exploring the Long-term Effects of Cognitive Exercise on Cognition After Stroke | Inappropriate study design |
| 5675 | A customized home-based computerized cognitive rehabilitation platform for patients with chronic-stage stroke: study protocol for a randomized controlled trial | Inappropriate population |
| 5676 | The Aerobic & Cognitive Exercise Study | Inappropriate study design |
| 5677 | A multiple-modality exercise program plus dual-task training improved mobility but did not impact vascular health in active older adults without dementia | Inappropriate population |
| 5678 | Tai chi-based exergaming program for older adults at risk of cognitive impairment | Inappropriate study design |
| 5679 | Effects of Aerobic Exercise Intensity on Clinical & Neural Outcomes in Depressed Youth | Inappropriate study design |
| 5680 | Exercise and cognitive function in people living with HIV: a scoping review | Inappropriate outcome |
| 5681 | Cognitive Training Therapy Improves the Effect of Hypocaloric Treatment on Subjects with Overweight/Obesity: a Randomised Clinical Trial | Inappropriate study design |
| 5682 | The Influence of Tai Chi Practice on Blood Pressure and Brain Health Among Older Adults With Hypertension (ACTION) | Inappropriate study design |
| 5683 | Acute exercise-related cognitive effects are not attributable to changes in end-tidal CO2 or cerebral blood velocity | Inappropriate population |
| 5684 | Cognitive behavioral therapy for insomnia to enhance cognitive function and reduce the rate of Aβ deposition in older adults with symptoms of insomnia: a single-site randomized pilot clinical trial protocol | Inappropriate study design |
| 5685 | Lifestyle interventiontoprevent cognitive impairment | Inappropriate study design |
| 5686 | Refining exercise prescription to promote executive functions in older adults using multistate transition modeling | Inappropriate study design |

| | | |
|---|---|---|
| 5687 | Brain training with mobile application and effect of it on quality of life of older adults | Inappropriate population |
| 5688 | mSIM: mobile Simultaneous Aerobic Exercise and Memory Training Intervention for Older Adults With Amnestic Mild Cognitive Impairment | Inappropriate population |
| 5689 | Yoga and Aerobic Exercise in Psychosis | Inappropriate study design |
| 5690 | A precision medicine multimodal lifestyle intervention for treating cognitive impairment: conceptual framework of the prevention trial | Inappropriate study design |
| 5691 | Aerobic Dance Training in Small Vessel Disease | Inappropriate study design |
| 5692 | Sex and BDNF polymorphism: impact on cognitive decline and white matter lesion load in older adults with subcortical vascular cognitive impairment | Inappropriate outcome |
| 5693 | The Effects of High-Intensity Interval Exercise and Hypoxia on Cognition in Sedentary Young Adults | Inappropriate population |
| 5694 | Activities for Cognitive Enhancement of Seniors | Inappropriate study design |
| 5695 | Memory improvement after a cognitive rehabilitation program in multiple sclerosis | Inappropriate study design |
| 5696 | Aerobic Exercise Training in Mild Cognitive Impairment Study | Inappropriate study design |
| 5697 | Exercise-induced effects on serum and CSF BDNF levels in elderly african americans with mild cognitive impairment: gene, exercise, and memory study (GEMS) | Inappropriate outcome |
| 5698 | Standing Tall - Cognition and Gait: a clinical trial of balance exercises delivered by an app to improve cognition and mobility in people with cognitive impairment | Inappropriate study design |
| 5699 | The effects of a single night of complete and partial sleep deprivation on physical and cognitive performance: a Bayesian analysis | Inappropriate population |
| 5700 | Pre-FRONTal Brain STability, Key for Action Against Disability in AGing | Inappropriate population |
| 5701 | Intensive Auditory Cognitive Training Improves Verbal Memory in Adolescents and Young Adults at Clinical High Risk for Psychosis | Inappropriate population |
| 5702 | The influence of an aerobic exercise intervention on brain volume in late adulthood | Inappropriate study design |
| 5703 | Effects of combined physical and cognitive training on fall prevention and risk reduction in older persons with mild cognitive impairment: a randomized controlled study | Inappropriate population |
| 5704 | Session 320 | Inappropriate outcome |
| 5705 | The mechanism of Tai Chi Chuan improving cognitive function in patients with mild cognitive impairment -multiple model MRI study | Inappropriate population |
| 5706 | Decreasing Alzheimer's disease and related dementias after delirium: exercise and cognitive training | Inappropriate population |
| 5707 | Impact of aerobic exercise on cognitive impairment and oxidative stress markers in methamphetamine-dependent patients | Inappropriate study design |
| 5708 | Effects of Aerobic Exercise on Cognitive Impairment in Patients with Alzheimer's Disease | Inappropriate study design |
| 5709 | A Randomized Controlled Trial of Adherence to a 24-Month Home-Based Physical Activity Program and the Health Benefits for Older Adults at Risk of Alzheimer's Disease: the AIBL Active-Study | Inappropriate study design |
| 5710 | Piedmont Aging, Cognition & Exercise Study-2 | Inappropriate study design |
| 5711 | Effect of a long-term intensive lifestyle intervention on prevalence of cognitive impairment | Inappropriate study design |
| 5712 | Multifactorial intervention in older adults with mild cognitive impairment | Inappropriate study design |
| 5713 | Effects of a Single Bout of Aerobic Exercise Versus Resistance Training on Cognitive Vulnerabilities for Anxiety Disorders | Inappropriate population |
| 5714 | MOTIVATION FOR EXERCISE AND PHYSICAL ACTIVITY ENGAGEMENT IN PEOPLE WITH MILD COGNITIVE IMPAIRMENT OR MILD DEMENTIA: RESULTS FROM THE PRAISED FEASIBILITY STUDY | Inappropriate population |
| 5715 | Does walking protect against decline in cognitive functioning among breast cancer patients undergoing chemotherapy? Results from a small randomised controlled trial | Inappropriate study design |
| 5716 | Association between aerobic capacity and cognitive performance in people with multiple sclerosis-a cross-sectional study | Inappropriate population |
| 5717 | Apolipoprotein E-34 modulates effects of aerobic exercise on brain-derived neurotrophic factor levels in nondemented older adults | Inappropriate study design |
| 5718 | The effects of exercise on cognitive function in older adults with subjective cognitive impairment (SCI) | Inappropriate population |
| 5719 | Validation of Oxygen Uptake Efficiency Slope in Patients With Stroke | Inappropriate study design |

| | | |
|---|---|---|
| 5720 | Effect of elastic band-based high-speed power training on cognitive function, physical performance and muscle strength in older women with mild cognitive impairment | Inappropriate study design |
| 5721 | Exergaming Improves Executive Functions in Patients With Metabolic Syndrome | Inappropriate study design |
| 5722 | Aerobic exercise: a possible therapy for vascular cognitive impairment | Inappropriate study design |
| 5723 | Effects of Aerobic Exercise Training With and Without Blood Flow Restriction in Fibromyalgia | Inappropriate study design |
| 5724 | Improving cognition in people with progressive multiple sclerosis: a multi-arm, randomized, blinded, sham-controlled trial of cognitive rehabilitation and aerobic exercise (COGEx) | Inappropriate study design |
| 5725 | Reducing Sedentary Behaviour and Cognition in Older People | Inappropriate population |
| 5726 | An acute bout of dynamic sitting exercises improves stroop performance and quality of sleep in older adults with cognitive impairment | Inappropriate intervention |
| 5727 | Strategy training to ameliorate the effects of age-related executive dysfunction: a pilot randomized controlled trial | Inappropriate study design |
| 5728 | Multicomponent physical exercise with simultaneous cognitive training to enhance dual-task walking of older adults: a secondary analysis of a 6-month randomized controlled trial with 1-year follow-up | Inappropriate study design |
| 5729 | Cognitive and physical rehabilitation of intensive care unit survivors: results of the RETURN randomized controlled pilot investigation | Inappropriate population |
| 5730 | Effect of cognitive-only and cognitive-motor training on preventing falls in community-dwelling older people: protocol for the smartÂ±step randomised controlled trial | Inappropriate study design |
| 5731 | Optimizing Cognitive, Environmental, and Neuromotor Stimulation in Traumatic Brain Injury | Inappropriate study design |
| 5732 | Effects of multicomponent exercise including home exercise program on cerebral blood flow in older adults with mild cognitive impairment: a single-center, non-randomized controlled trial | Inappropriate study design |
| 5733 | Randomized controlled study with multifactorial intervention to prevent progression in elderly people at risk for dementia | Inappropriate study design |
| 5734 | Acute hypoxic gas breathing severely impairs cognition and task learning in humans | Inappropriate study design |
| 5735 | Preliminary findings from a 6-month randomized controlled trial of combined dual-task gait training and aerobic exercise in older adults with cognitive impairment but no dementia | Inappropriate intervention |
| 5736 | Kickboxing and cognition in parkinson's disease | Inappropriate population |
| 5737 | A Single Bout of High-Intensity Interval Training Reduces Awareness of Subsequent Hypoglycemia in Patients With Type 1 Diabetes | Inappropriate study design |
| 5738 | Cognitive remediation for treatment-resistant depression: effects on cognition and functioning and the role of online homework | Inappropriate study design |
| 5739 | The effect of aerobic exercise on fall risk reduction among older adults with mild vascular cognitive impairment | Inappropriate study design |
| 5740 | Cognitive remediation therapy (CRT) for young early onset patients with schizophrenia: an exploratory randomized controlled trial | Inappropriate study design |
| 5741 | Aerobic exercise and cognitive remediation in multiepisode schizophrenia and schizoaffective disorder: preliminary results: from a randomized clinical trial | Inappropriate outcome |
| 5742 | Functional and cognitive impairment prevention through early physical activity for geriatric hospitalized patients: study protocol for a randomized controlled trial | Inappropriate study design |
| 5743 | Home-based exercise training following ward-based rehabilitation in geriatric patients with cognitive impairment | Inappropriate study design |
| 5744 | Effect of different types of physical exercise in SUS patients with depression | Inappropriate study design |
| 5745 | Japan-multimodal intervention Trial for prevention of dementia (J-MINT) | Inappropriate study design |
| 5746 | Physical Fitness, Cardiovascular and Brain Health | Inappropriate study design |
| 5747 | Cognitive remediation of working memory deficits: durability of training effects in severely impaired and less severely impaired schizophrenia | Inappropriate population |
| 5748 | V-TIME: a treadmill training program augmented by virtual reality to decrease fall risk in older adults: study design of a randomized controlled trial | Inappropriate study design |
| 5749 | E4 cancer survivors show better fall and functional status outcomes after receiving exercise interventions than non-E4 cancer survivors | Inappropriate population |
| 5750 | Memory loss: five new things | Inappropriate study design |
| 5751 | Bicycle ergometry does not improve verbal memory in mild relapsing remitting MS - Results of a RCT | Inappropriate study design |

| | | |
|---|---|---|
| 5752 | A Controlled Trial of Daily Cognitive-Behavioral Therapy for Pediatric Obsessive-Compulsive Disorder | Inappropriate study design |
| 5753 | A comparison of cognitive performance decreases during acute, progressive fatigue arising from different concurrent stressors | Inappropriate study design |
| 5754 | Feasibility and Pilot Testing of a Mindfulness Intervention for Frail Older Adults and Individuals With Dementia | Inappropriate study design |
| 5755 | Sex differences in aerobic exercise efficacy on cognitive health: possible role of APOE-E4 | Inappropriate population |
| 5756 | Exploring synergistic effects of aerobic exercise and mindfulness training on cognitive function in older adults: protocol for a pilot randomized controlled trial | Inappropriate outcome |
| 5757 | Design of the WHIP-PD study: a phase II, twelve-month, dual-site, randomized controlled trial evaluating the effects of a cognitive-behavioral approach for promoting enhanced walking activity using mobile health technology in people with Parkinson-disease | Inappropriate population |
| 5758 | The interacting effects of cognitive demand and recovery of postural stability in balance-impaired elderly persons | Inappropriate study design |
| 5759 | CogStep: evaluation of a 12-week combined home-based exercise and psychoeducation program on dementia risk in older adults aged >60 years | Inappropriate study design |
| 5760 | Aerobic Exercise to Improve Executive Language Function In Older Adults | Inappropriate study design |
| 5761 | Impact of increasing physical activity on cognitive functioning in breast cancer survivors: rationale and study design of Memory & Motion | Inappropriate study design |
| 5762 | Effects of yoga, aerobic, and stretching and toning exercises on cognition in adult cancer survivors: protocol of the STAY Fit pilot randomized controlled trial | Inappropriate study design |
| 5763 | Cognitive Interventions in Older People following Traumatic Brain Injury | Inappropriate study design |
| 5764 | Effects of bronchodilator therapy and exercise with self-management behaviourmodification on psychological and cognitive outcomes in COPD | Inappropriate study design |
| 5765 | "Effect of a muscle strengthening program in neural functions and in emotional state of elderly with mild cognitive impairment" | Inappropriate study design |
| 5766 | Benefits of exercise in patients with rheumatoid arthritis: a randomized controlled trial of a patient-specific exercise programme | Inappropriate study design |
| 5767 | Visual versus auditory cognitive training for schizophrenia: effects on cognition, symptoms and quality of life | Inappropriate population |
| 5768 | CogXergaming to Promote Physical Activity and Cognitive Function in Frail Older Adults | Inappropriate study design |
| 5769 | Cognitive effects of aerobic exercise for adults with mild cognitive impairment: a controlled trial | Inappropriate outcome |
| 5770 | Cognitive remediation in schizophrenia | Inappropriate study design |
| 5771 | Computerized cognitive remediation training for schizophrenia: an open label, multi-site, multinational methodology study | Inappropriate study design |
| 5772 | Transforming dementia care and quality of life using innovative touch screen computer engagement: a research study-the birdsong initiative | Inappropriate study design |
| 5773 | Efficacy Study of GAÏA Program Cognitive Remediation of Facial Affects Processing in Schizophrenia | Inappropriate study design |
| 5774 | Training With Cognitive Aerobic Exercise in Older Adults at Risk of Cognitive Decline | Inappropriate population |
| 5775 | Interactive Cognitive-Motor Step Training Improves Cognitive Risk Factors of Falling in Older Adults - A Randomized Controlled Trial | Inappropriate study design |
| 5776 | Generations on the Move (GIB-Study): intergenerational Exercise and Health Promotion | Inappropriate outcome |
| 5777 | Effects of a traditional Chinese mind-body exercise, Baduanjin, on the physical and cognitive functions in the community of older adults with cognitive frailty: study protocol for a randomised controlled trial | Inappropriate study design |
| 5778 | Impact of exercise on cognition in multiple sclerosis | Inappropriate population |
| 5779 | Brain structure changes in nondemented seniors after six-month dance-exercise intervention | Inappropriate study design |
| 5780 | Innovative approach using neuromodulation techniques as a treatment for TBI related cognitive impairments | Inappropriate study design |
| 5781 | A Problem Solving Intervention for Post-ICU Cognitive Impairment in Older Adults | Inappropriate study design |
| 5782 | Effect of group-based dual-tasks exercise on physical, cognitive and psychosocial function in older adults with type 2 diabetes with balance impairment | Inappropriate outcome |

| | | |
|---|---|---|
| 5783 | A randomised trial comparing low-fat diets differing in carbohydrate and protein ratio, combined with regular moderate intensity exercise, on glycaemic control, cardiometabolic risk factors, food cravings, cognitive function and psychological wellbeing in adults with type 2 diabetes: study protocol | Inappropriate study design |
| 5784 | Building Better Caregivers: a Pragmatic 12-Month Trial of a Community-Based Workshop for Caregivers of Cognitively Impaired Adults | Inappropriate study design |
| 5785 | Amphetamine enhances gains in auditory discrimination training in adult schizophrenia patients: assessing tolerability and predictive biomarkers | Inappropriate study design |
| 5786 | EXERCISE INTERVENTIONS MAY PREVENT HIPPOCAMPAL ATROPHY IN THE FIRST YEAR AFTER STROKE: BLINDED ANALYSIS OF PISCES PILOT DATA | Inappropriate population |
| 5787 | The Watermemories swimming club for people with dementia: cluster randomised controlled trial | Inappropriate study design |
| 5788 | 2017 Canadian Association of Cardiovascular Prevention and Rehabilitation Annual Scientific Meeting | Inappropriate population |
| 5789 | The efficacy of a multi-modality exercise program combined with mind motortask training for older adults at risk of cognitive impairment on usual and dual-task gait: a randomized controlled trial | Inappropriate study design |
| 5790 | Cognitive behavioural therapy with optional graded exercise therapy in patients with severe fatigue with myotonic dystrophy type 1: a multicentre, single-blind, randomised trial | Inappropriate study design |
| 5791 | Sit Less or Exercise More: impact on Cardiometabolic Health in MS | Inappropriate study design |
| 5792 | Exercise for diagnosis and treatment of heart dysfunction among breast cancer patients | Inappropriate study design |
| 5793 | Feasibility of an RCT on exercise interventions in early-onset dementia: let's move on! | Inappropriate population |
| 5794 | Study protocol of the Aerobic exercise and CogniTIVe functioning in women with breAsT cancEr (ACTIVATE) trial: a two-arm, two-centre randomized controlled trial | Inappropriate study design |
| 5795 | Cerebrovascular Outcomes in Ischemic Heart Disease | Inappropriate study design |
| 5796 | Cognitive remediation training improves performance in patients with chronic fatigue syndrome | Inappropriate study design |
| 5797 | Neural Changes of Exercise: a Functional MRI Study | Inappropriate study design |
| 5798 | Efficacy of health promotion interventions in adults with lifestyle and health risk factors for dementia | Inappropriate study design |
| 5799 | Ba Duan Jin in Treatment of Attention Deficit Hyperactivity Disorder | Inappropriate study design |
| 5800 | Tyrosine Ingestion and Its Effects on Cognitive and Physical Performance in the Heat | Inappropriate study design |
| 5801 | HOMeCare: caring for the Dementia Caregiver and their Loved One via the HOMeCare Exercise and Mindfulness for Health Program | Inappropriate study design |
| 5802 | Influence of sports climbing and yoga on spasticity, cognitive function, mood and fatigue in patients with multiple sclerosis | Inappropriate population |
| 5803 | Comparison of dance Aerobic and Strength Training on brain function and Stress in Adults | Inappropriate study design |
| 5804 | Multimodal Training Reduces Fall Frequency as Physical Activity Increases in Individuals With Parkinson's Disease | Inappropriate study design |
| 5805 | Enrolling older adults with cognitive impairment in research: lessons from a study of Tai Chi for osteoarthritis knee pain | Inappropriate study design |
| 5806 | Effects of multicomponent exercises on the elderly health | Inappropriate population |
| 5807 | Feasibility of a symptom-adapted physical activity intervention during induction chemotherapy for older adults with acute myeloid leukemia (AML) | Inappropriate study design |
| 5808 | Examining glucose control and working memory as a function of age in older type 2 diabetics: results from the Look AHEAD Brain MRI ancillary study | Inappropriate population |
| 5809 | Testing the "pACT" strategy: amphetamine (AMPH) enhances gains in auditory discrimination training in adult schizophrenia (SZ) patients | Inappropriate study design |
| 5810 | Diet, physical exercise, and metabolic control intervention to reduce the incidence of major neurocognitive disorders among individuals with type 2 diabetes combined with mild neurocognitive impairment â€“ a pilot study | Inappropriate intervention |
| 5811 | The effect of aerobic exercise training on arterial stiffness and sleep impairment in persons with traumatic brain injury | Inappropriate population |
| 5812 | Acute Aerobic Exercise Effect | Inappropriate study design |
| 5813 | High-intensity interval exercise impairs neuroelectric indices of reinforcement-learning | Inappropriate study design |
| 5814 | Exercise Intervention to Reverse Frailty | Inappropriate study design |

| | | |
|---|---|---|
| 5815 | Effects of personalized exercise training on physical activity, health-related fitness, sleep quality, and quality of life among middle-aged and older adults with multiple health problems | Inappropriate study design |
| 5816 | Trauma-focused cognitive behaviour therapy and exercise for chronic whiplash: protocol of a randomised, controlled trial | Inappropriate study design |
| 5817 | The tired study: two interventions for fatigued advanced cancer patients, a randomised controlled trial | Inappropriate study design |
| 5818 | Effects of periodic carbohydrate ingestion on endurance and cognitive performances during a 40-km cycling time-trial under normobaric hypoxia in well-trained triathletes | Inappropriate study design |
| 5819 | A multi-modal virtual reality treadmill intervention for enhancing mobility and cognitive function in people with multiple sclerosis: protocol for a randomized controlled trial | Inappropriate study design |
| 5820 | The effects of 23-weeks golf training on cognitive performance, brain network activity and neurotrophic markers in older adults with subjective memory complaints: a randomized controlled pilot study | Inappropriate study design |
| 5821 | Effect of mild dehydration on cognitive function, mood and exercise performance | Inappropriate population |
| 5822 | Virtual reality for improving social activities and participation (VR-SOAP): development of a new treatment for young people with psychosis | Inappropriate study design |
| 5823 | Comparison of the effects on dynamic balance and aerobic capacity between objective and subjective methods of high-intensity robot-assisted gait training in chronic stroke patients: a randomized controlled trial | Inappropriate intervention |
| 5824 | Intermittent Running and Cognitive Performance after Ketone Ester Ingestion | Inappropriate study design |
| 5825 | Effects of active computer gaming and aerobic exercise on memory, attention and physical performance in older adults | Inappropriate population |
| 5826 | Combined methazolamide and theophylline improves oxygen saturation but not exercise performance or altitude illness in acute hypobaric hypoxia | Inappropriate study design |
| 5827 | Effect of an acute bout of exercise on cognition | Inappropriate study design |
| 5828 | E-BRAIN Project: investigating the effect of different forms of physical exercises on the cognition of the elderly | Inappropriate study design |
| 5829 | Mindful Meditation for Chronic Stroke | Inappropriate study design |
| 5830 | Improving rehabilitation in sarcopenia: a randomized-controlled trial utilizing a muscle-targeted food for special medical purposes | Inappropriate study design |
| 5831 | Using neuroplasticity-based auditory training to improve verbal memory in schizophrenia | Inappropriate study design |
| 5832 | The Effects of a 16-week Aerobic Exercise and Mindfulness-based Intervention on Chronic Psychosocial Stress: a Nonrandomized Pilot and Feasibility Trial | Inappropriate study design |
| 5833 | Effect of current stimulation on memory, attention in diabetes | Inappropriate population |
| 5834 | Effects of increased physical activity to prevent chronic pain in community-dwelling older adults: a randomized controlled trial | Inappropriate population |
| 5835 | Effectiveness of a targeted lifestyle intervention in primary care on diet and physical activity among South Asians at risk for diabetes: 2-year results of a randomised controlled trial in the Netherlands | Inappropriate outcome |
| 5836 | Acoustic, respiratory, cognitive and wellbeing comparisons of two groups of people with Parkinson's disease participating in voice and choral singing group therapy (VCST) versus a music appreciation activity | Inappropriate study design |
| 5837 | The Potential of Carnosine Supplementation in Optimising Cardiometabolic Health | Inappropriate study design |
| 5838 | Dual-Task Obstacle Crossing Training Could Immediately Improve Ability to Control a Complex Motor Task and Cognitive Activity in Chronic Ambulatory Individuals With Spinal Cord Injury | Inappropriate population |
| 5839 | Mild dehydration impairs cognitive performance and mood of men | Inappropriate study design |
| 5840 | A best practice fall prevention exercise program to improve balance, strength / power, and psychosocial health in older adults: study protocol for a randomized controlled trial | Inappropriate population |
| 5841 | Cognitive behaviour therapy plus aerobic exercise training to increase activity in patients with myotonic dystrophy type 1 (DM1) compared to usual care (OPTIMISTIC): study protocol for randomised controlled trial | Inappropriate study design |
| 5842 | Addition of a non-immersive virtual reality component to treadmill training to reduce fall risk in older adults (V-TIME): a randomised controlled trial | Inappropriate study design |
| 5843 | Effects of Walking and mHealth Activities for People With Parkinson Disease | Inappropriate study design |
| 5844 | Comparing the efficacy of cognitive-behavioral guided self-help and health at every size guided self-help for disordered eating | Inappropriate study design |

| | | |
|---|---|---|
| 5845 | Functional high-intensity exercise is more effective in acutely increasing working memory than aerobic walking: an exploratory randomized, controlled trial | Inappropriate study design |
| 5846 | Effects of exercise on disease progression and cognition in the marmoset EAE model | Inappropriate study design |
| 5847 | Effects of acute aerobic exercise on a task-switching protocol and brain-derived neurotrophic factor concentrations in young adults with different levels of cardiorespiratory fitness | Inappropriate study design |
| 5848 | IMPROVING REHABILITATION IN SARCOPENIA (IRIS) BY MUSCLE-TARGETED NUTRITIONAL SUPPORT: a RANDOMIZED, DOUBLE-BLIND, PLACEBO-CONTROLLED TRIAL | Inappropriate study design |
| 5849 | Cognitive Emotional Stimulation Program (CESP) for memory improvement in older community-dwelling adults (Programa de Estimulación Cognitiva Emocional [PECE] para la Mejora de la Memoria de Personas Mayores que Viven en la Comunidad) | Inappropriate study design |
| 5850 | Perioperative Enhancement of Cognitive Trajectory (The PROTECT trial) | Inappropriate outcome |
| 5851 | Medial temporal lobe cortical changes in response to exercise interventions in people with early psychosis: a randomized controlled trial | Inappropriate population |
| 5852 | CyberCycling to Reduce Therapeutic Side Effects and Improve Quality of Life in AYA Cancers | Inappropriate study design |
| 5853 | Correlation between exercise and motor skills with elderly patients cognition | Inappropriate study design |
| 5854 | The impact of endurance training on brain structure and function in schizophrenia patients | Inappropriate study design |
| 5855 | The Effect of Active Plus, a Computer-Tailored Physical Activity Intervention, on the Physical Activity of Older Adults with Chronic Illness(es)-A Cluster Randomized Controlled Trial | Inappropriate study design |
| 5856 | The effect of alternate-day caloric restriction on the metabolic consequences of 8 days of bed rest in healthy lean men: a randomized trial | Inappropriate study design |
| 5857 | Intravenous Iron Supplement for Iron Deficiency in Cardiac Transplant Recipients (IronIC): rationale and Design of a Randomized, Controlled Trial | Inappropriate population |
| 5858 | Exercise Effects on Word Learning | Inappropriate study design |
| 5859 | A randomized controlled trial of Cognitive Sensory Motor Training Therapy on the recovery of arm function in acute stroke patients | Inappropriate study design |
| 5860 | Aging Brain Structure and Memory in Response to Exercise | Inappropriate study design |
| 5861 | A non-inferiority randomized controlled trial comparing a home-based aerobic exercise program to a self-administered cognitive-behavioral therapy for insomnia in cancer patients | Inappropriate study design |
| 5862 | Omega-3 Fatty Acids and Exercise on Mobility and Cognition in Older Women | Inappropriate study design |
| 5863 | Forced and Voluntary Aerobic Cycling Interventions Improve Walking Capacity in Individuals With Chronic Stroke | Inappropriate population |
| 5864 | Feasibility Study of an Individualized Exergame Training for Older Adults With MI and/or UI (VITAAL) | Inappropriate study design |
| 5865 | Changes in daytime functioning following exercise training in adults with untreated obstructive sleep apnea: a randomized controlled trial | Inappropriate study design |
| 5866 | Associations of leucocyte telomere length with brain MRI and PIB-pet measures: the finger study | Inappropriate study design |
| 5867 | Effects of exergame and resistance training on reaction time, functional fitness and quality of life in older adults | Inappropriate study design |
| 5868 | Aerobic exercise increases hippocampal volume and improves memory in multiple sclerosis: preliminary findings | Inappropriate study design |
| 5869 | Early Intervention Programme for Young Patients (Aged 15 - 30 Years) With Symptoms Following Concussion | Inappropriate study design |
| 5870 | Enhancement of Physical Fitness in Older Adults | Inappropriate population |
| 5871 | Standardized Ambulatory Neurorehabilitation Program for Patients With Multiple Sclerosis | Inappropriate study design |
| 5872 | Forced Aerobic Exercise Preceding Task Practice Improves Motor Recovery Poststroke | Inappropriate population |
| 5873 | Acute aerobic exercise enhances pleasant compared to unpleasant visual scene processing | Inappropriate population |
| 5874 | Improving community walking after a stroke, a new approach | Inappropriate study design |
| 5875 | Holograms for individualised multi-factorial tele-rehabilitation vs standard care in an ageing population with balance disorders â€“ a proof of concept study | Inappropriate study design |
| 5876 | The effect of resistance training on white matter lesion progression: proof-of-concept results from a 12-month RCT | Inappropriate study design |

| | | |
|---|---|---|
| 5877 | Peer Mentorship: an Intervention To Promote Effective Pain Self-Management In Adolescents | Inappropriate study design |
| 5878 | Branched-chain amino acids, arginine, citrulline alleviate central fatigue after 3 simulated matches in taekwondo athletes: a randomized controlled trial | Inappropriate study design |
| 5879 | Coaching and Exercise for Better Walking (ComeBACK) trial | Inappropriate study design |
| 5880 | Muscle fatigability and control of force in men with posttraumatic stress disorder | Inappropriate population |
| 5881 | Evaluation of the effect of structured exercise therapy on neuro physiological and cognitive functions of young adults with type 2 diabetes mellitus | Inappropriate study design |
| 5882 | Thinking, Walking Outcomes by Type of Exercise | Inappropriate intervention |
| 5883 | Exercise reduces apnea-hypopnea index in cognitively impaired older adults | Inappropriate outcome |
| 5884 | Cardiac rehabilitation adapted to transient ischaemic attack and stroke (CRAFTS): a randomised controlled trial | Inappropriate population |
| 5885 | Improving outcomes for COPD patients with mild-to-moderate anxiety and depression: a systematic review of cognitive behavioural therapy | Inappropriate population |
| 5886 | ES15.03 Managing Sleep Difficulties and Cancer | Inappropriate study design |
| 5887 | Does mental fatigue impair physical performance? A replication study | Inappropriate study design |
| 5888 | Adjunctive tai chi in geriatric depression with comorbid arthritis: a randomized, controlled trial | Inappropriate study design |
| 5889 | A Home-based Intervention to Reduce Sedentary Behaviour and Improve Function After Stroke | Inappropriate study design |
| 5890 | A paradigm shift: rehabilitation robotics, cognitive skills training, and function after stroke | Inappropriate population |
| 5891 | Establishing the optimum TDCS application on dual task gait performance-exploration of peoplewith chronic stroke | Inappropriate study design |
| 5892 | Effects of physical activity on inflammation, skeletal muscle strength/function (sarcopenia) and fat infiltration (sarcopenic obesity) in older adults with knee osteoarthritis: a randomized controlled trial | Inappropriate study design |
| 5893 | The Influence of Regular Beef Consumption and Protein Density of the Diet on Training-induced Gains in Muscle Strength and Performance in Healthy Adults | Inappropriate study design |
| 5894 | FV 44 Interindividual differences in cognitive response to acute exercise â€“ A randomized controlled cross-over study | Inappropriate study design |
| 5895 | COVID-19, Chronic Adaptation and Response to Exercise (COVID-CARE) | Inappropriate population |
| 5896 | Influence of different rehabilitative aerobic exercise programs on (anti-) inflammatory immune signalling, cognitive and functional capacity in persons with MS - study protocol of a randomized controlled trial | Inappropriate study design |
| 5897 | Effect of exercise on quality of life (QoL) in 198 older patients with cancer: a URCC NCORP nationwide RCT | Inappropriate population |
| 5898 | Trial Investigating the Effect of Different Exercise Forms on Depression | Inappropriate study design |
| 5899 | Effects of aerobic exercise intensity on novel blood biomarkers of neuroplasticity after stroke | Inappropriate population |
| 5900 | Can exercise improve heart and lung function in people with Parkinsonâ€™s? The EXoCARP study | Inappropriate population |
| 5901 | Observational Prolonged Trial in Myotonic Dystrophy Type 1 | Inappropriate study design |
| 5902 | A risk model for falls in older patients after hospitalization for acute myocardial infarction: the SILVER-AMI study | Inappropriate study design |
| 5903 | Dopamine gene profiling to predict impulse control and effects of dopamine agonist ropinirole | Inappropriate population |
| 5904 | Multimorbidity rehabilitation in chronic disease: general rehabilitation compared to usual care | Inappropriate study design |
| 5905 | Efficacy of Tai Chi Training on Insomnia in Older Adults | Inappropriate study design |
| 5906 | Acute high-intensity interval training improves motor skill acquisition | Inappropriate population |
| 5907 | Recovery Improved in Covert Stroke With Exercise | Inappropriate study design |
| 5908 | Combining d-cycloserine with motor training does not result in improved general motor learning in neurologically intact people or in people with stroke | Inappropriate population |
| 5909 | Novel assessment of acute sport-related concussion sensory, motor, and neurocognitive impairment using robotics: a prospective validation study | Inappropriate study design |
| 5910 | Effects of a Home-Based and Volunteer-Administered Physical Training, Nutritional, and Social Support Program on Malnutrition and Frailty in Older Persons: a Randomized Controlled Trial | Inappropriate outcome |
| 5911 | Occupational Therapy in Patients With Multiple Sclerosis | Inappropriate study design |
| 5912 | The Post-Operative Pain in Cerebral Palsy (POPPIES) Trial | Inappropriate study design |

| | | |
|---|---|---|
| 5913 | Effect of Additional Rehabilitation After Botulinum Toxin-A on Upper Limb Activity in Chronic Stroke: the InTENSE Trial | Inappropriate study design |
| 5914 | Dishabituation as a treatment for impaired awareness of hypoglycaemia in type 1 diabetes | Inappropriate study design |
| 5915 | The effect of dual task and executive training on pattern of gait in older adults with balance impairment: a Randomized controlled trial | Inappropriate study design |
| 5916 | The resist diabetes trial: rationale, design, and methods of a hybrid efficacy/effectiveness intervention trial for resistance training maintenance to improve glucose homeostasis in older prediabetic adults | Inappropriate study design |
| 5917 | SPRINT HEART: study of Progressive Resistance and INterval Training in HEART disease | Inappropriate study design |
| 5918 | Comparison of the effects of clinical pilates and exercise treatment in multiple sclerosis patients: a single blind randomised study | Inappropriate study design |
| 5919 | The Effects of Thoracic Joint Mobilization on Pulmonary Functions of Patients With Stroke | Inappropriate study design |
| 5920 | Feasibility and preliminary efficacy of digital cognitive behavioural therapy for anxiety | Inappropriate population |
| 5921 | A pilot study of combined endurance and resistance exercise rehabilitation for verbal memory and functional connectivity improvement in epilepsy | Inappropriate study design |
| 5922 | Motor imagery of walking and walking while talking: a pilot randomized-controlled trial protocol for older adults | Inappropriate population |
| 5923 | Impact of low electrical stimulation on the scalp and physical training on the adaptations of the system that regulates body functions in patients with weakness on one side of the body resulting from a stroke | Inappropriate study design |
| 5924 | Intellectual disability, exercise and aging: the IDEA study: study protocol for a randomized controlled trial | Inappropriate population |
| 5925 | The Role of Methylphenidate on Performance in the Cold | Inappropriate study design |
| 5926 | Improving cognitive performance via high intensity aerobic exercise: a randomized controlled trial | Inappropriate study design |
| 5927 | Effectiveness of a computerized decision support system in decreasing the anticholinergics use in intensive care unit | Inappropriate study design |
| 5928 | No influence of noradrenaline manipulation on acute exercise-induced increase of brain-derived neurotrophic factor | Inappropriate study design |
| 5929 | Dysphagia therapy with surface electromyographic biofeedback: a feasibility randomised controlled trial in acute stroke | Inappropriate study design |
| 5930 | The Effect of a 2-Year Intervention Consisting of Diet, Physical Exercise, Cognitive Training, and Monitoring of Vascular Risk on Chronic Morbidity-the FINGER Randomized Controlled Trial | Inappropriate study design |
| 5931 | Effects of protein supplementation on aerobic training-induced gains in cardiopulmonary fitness, muscle mass, and functional performance in chronic stroke: a randomized controlled pilot study | Inappropriate study design |
| 5932 | Effects of a Group-Mediated Exercise and Dietary Intervention in the Treatment of Prostate Cancer Patients Undergoing Androgen Deprivation Therapy: results From the IDEA-P Trial | Inappropriate study design |
| 5933 | High intensity interval training may be superior to moderate intensity continuous exercise in chronic stroke | Inappropriate study design |
| 5934 | Cognitive enhancement by transcranial laser stimulation and acute aerobic exercise | Inappropriate study design |
| 5935 | Long Term Outcome on Brain and Lung of Different Oxygen Strategies in ARDS Patients | Inappropriate study design |
| 5936 | Effects of Slow-speed Traditional Resistance Training, High-speed Resistance Training and Multicomponent Training With Variable Resistances on Molecular, Body Composition, Neuromuscular, Physical Function and Quality of Life Variables in Older Adults | Inappropriate intervention |
| 5937 | Augmentation of Cognitive Training in Children With TBI With D-Cyloserine | Inappropriate study design |
| 5938 | Two-year follow-up of the OptiTrain randomised controlled exercise trial | Inappropriate study design |
| 5939 | Exercise Intensity Does not Modulate the Effect of Acute Exercise on Learning a Complex Whole-Body Task | Inappropriate study design |
| 5940 | High intensity exercise as a possible treatment for loss of symptoms of low blood sugar in people with type 1 diabetes | Inappropriate study design |
| 5941 | Study protocol of the TIRED study: a randomised controlled trial comparing either graded exercise therapy for severe fatigue or cognitive behaviour therapy with usual care in patients with incurable cancer | Inappropriate study design |
| 5942 | The impact of aerobic exercise training with vascular occlusion in patients with chronic heart failure | Inappropriate study design |

| | | |
|---|---|---|
| 5943 | Training intensity: a barrier or facilitator to engagement in rehabilitation following stroke? | Inappropriate outcome |
| 5944 | A Two-Part Approach to Examine the Effects of Theacrine (TeaCrine®) Supplementation on Oxygen Consumption, Hemodynamic Responses, and Subjective Measures of Cognitive and Psychometric Parameters | Inappropriate study design |
| 5945 | The effect of a 12-week aerobic exercise intervention on neurometabolites in young healthy adults using 7T magnetic resonance spectroscopy | Inappropriate study design |
| 5946 | Post-concussion and Transcranial Direct Current Stimulation | Inappropriate study design |
| 5947 | Mediterranean Lifestyle Intervention in Patients With Obstructive Sleep Apnea | Inappropriate study design |
| 5948 | Glucose and Counterregulatory Responses to Exercise in Adults With Type 1 Diabetes and Impaired Awareness of Hypoglycemia Using Closed-Loop Insulin Delivery: a Randomized Crossover Study | Inappropriate population |
| 5949 | PBWST (Partial Body-Weight Supported Treadmill Training) and Muscle Power Training After Sub-acute Stroke | Inappropriate study design |
| 5950 | Enhanced Medical Rehabilitation for Spinal Cord Injury | Inappropriate study design |
| 5951 | Predictors of outcome following treatment for chronic fatigue | Inappropriate outcome |
| 5952 | The effect of a single aerobic training session on cognitive flexibility in late middle-aged adults | Inappropriate study design |
| 5953 | A prospective test of the late effects of potentially antineuroplastic drugs in a stroke rehabilitation study | Inappropriate study design |
| 5954 | A study protocol of a randomised controlled trial to investigate if a community based strength training programme improves work task performance in young adults with Down syndrome | Inappropriate study design |
| 5955 | Repeated Sprints in Fasted State Impair Reaction Time Performance | Inappropriate study design |
| 5956 | Training fast or slow? Exercise for depression: a randomized controlled trial | Inappropriate population |
| 5957 | Exercise and Neurofeedback Intervention for improving postural stability and reducing risk of fall in elderly women | Inappropriate study design |
| 5958 | Alcohol pharmacodynamics in young-elderly adults contrasted with young and middle aged subjects | Inappropriate population |
| 5959 | Individualized counselling for active aging: protocol of a single-blinded, randomized controlled trial among older people (the AGNES intervention study) | Inappropriate study design |
| 5960 | Yoga for Pediatric Tension-Type Headache | Inappropriate study design |
| 5961 | Hypertension and exercise: an intervention study in older patients | Inappropriate study design |
| 5962 | Effectiveness of home-based and remotely supervised aerobic exercise in Parkinson's disease: a double-blind, randomised controlled trial | Inappropriate study design |
| 5963 | Train the brain! Immediate sensorimotor effects of mentally-performed flexor exercises in patients with neck pain. A pilot study | Inappropriate study design |
| 5964 | An integrative cognitive behavioral therapy program for adults with migraine: a pilot study | Inappropriate study design |
| 5965 | Is graded exercise better than cognitive behaviour therapy for fatigue? A UK randomized trial in primary care | Inappropriate study design |
| 5966 | Effect of combination therapy with drug, device, and exercise for improving post stroke gait: a randomised controlled trial | Inappropriate study design |
| 5967 | Randomized Trial of a Dissonance-Based Group Treatment for Eating Disorders Versus a Supportive Mindfulness Group Treatment | Inappropriate intervention |
| 5968 | Cardiorespiratory effects of forced-and voluntary-rate aerobic exercise in individuals with chronic stroke | Inappropriate population |
| 5969 | RESULTS FROM THE INTENSE PHYSICAL ACTIVITY AND COGNITION (IPAC) STUDY: a RANDOMIZED CONTROLLED TRIAL OF PHYSICAL ACTIVITY INTERVENTION AND EXECUTIVE FUNCTION IN COMMUNITY DWELLING OLDER ADULTS | Inappropriate study design |
| 5970 | Weight loss intervention for people with serious mental illness: a randomized controlled trial of the RENEW program | Inappropriate outcome |
| 5971 | Exercise as a diagnostic and therapeutic tool for preventing cardiovascular morbidity in breast cancer patients- the BReast cancer EXercise InTervention (BREXIT) trial protocol | Inappropriate population |
| 5972 | Concurrent Aerobic Exercise and Cognitive Training to Prevent Alzheimer's in At-risk Older Adults | Inappropriate study design |
| 5973 | Protein Intake and Step Reduction in Older Adults | Inappropriate study design |
| 5974 | A Pilot Study of the Wii Fit as a Low-Cost Virtual Reality System to Evaluate Balance Ability in Older Adults | Inappropriate study design |
| 5975 | Health promotion program through telemedicine for improving physical fitness among the pre-frail elderly in an urban area: a randomized controlled trial | Inappropriate population |

| | | |
|---|---|---|
| 5976 | Interval Running Training Improves Cognitive Flexibility and Aerobic Power of Young Healthy Adults | Inappropriate study design |
| 5977 | Applying tDCS during aerobic exercise: acute online effects on executive functions and perceived exertion | Inappropriate population |
| 5978 | Impact of a collaborative multidisciplinary team on ICU delirium | Inappropriate study design |
| 5979 | An evaluation of the effect of inositol-stabilized arginine silicate on heart rate and blood pressure | Inappropriate outcome |
| 5980 | An exaggerated blood pressure response to exercise is common in the subacute phase after stroke, but is not affected by randomization to 12 weeks of intensive aerobic exercise | Inappropriate study design |
| 5981 | Schizophrenia and Physical Exercise | Inappropriate study design |
| 5982 | Moderate cardiovascular exercise speeds up neural markers of stimulus evaluation during attentional control processes | Inappropriate study design |
| 5983 | Adding high-intensity interval training to conventional training modalities: optimizing health-related outcomes during chemotherapy for breast cancer: the OptiTrain randomized controlled trial | Inappropriate intervention |
| 5984 | MODERATE-INTENSITY AEROBIC EXERCISE IMPROVES COGNITIVE FUNCTION IN PATIENTS WITH ALZHEIMER'S DISEASE | Inappropriate population |
| 5985 | Effect of active versus passive recovery on metabolism and performance during subsequent exercise | Inappropriate study design |
| 5986 | Clinical Symptoms of Schizophrenia and Physical Exercise | Inappropriate study design |
| 5987 | Feasibility and impact of a brief cognitive-behavioral intervention in conjunction with an aerobic exercise program for managing non-cardiac chest pain: a pilot study | Inappropriate study design |
| 5988 | Effects of Aerobic Exercise in Parkinson's Disease | Inappropriate study design |
| 5989 | Combined endurance/resistance training early on, after a first myocardial infarction, does not induce negative left ventricular remodelling | Inappropriate study design |
| 5990 | Efficacy of a multi-faceted vestibular rehabilitation program: balance and eye-movement exercises for persons with multiple sclerosis (BEEMS) | Inappropriate study design |
| 5991 | Rehabilitation via home based gaming exercise for the upper-limb post stroke (RHOMBUS): results of an intervention feasibility trial | Inappropriate study design |
| 5992 | Effect of Action Observation and Imitation on Improving the Functional Activities Indices in Hemiplegic Patients Based on Mirror Neurons Theory | Inappropriate study design |
| 5993 | Feasibility of high intensity interval training for patients with cerebrovascular disease | Inappropriate study design |
| 5994 | Mindfulness vs. Support Groups for Irritable Bowel Syndrome | Inappropriate population |
| 5995 | Effects of Exercise and Nicotine on Cognition in Smokers | Inappropriate study design |
| 5996 | The effectiveness of aerobic training, cognitive behavioural therapy, and energy conservation management in treating MS-related fatigue: the design of the TREFAMS-ACE programme | Inappropriate study design |
| 5997 | The effect of counselling, graded exercise and usual care for people with chronic fatigue in primary care: a randomized trial | Inappropriate study design |
| 5998 | SKY vs. HEP in PTSD | Inappropriate study design |
| 5999 | Evaluation of lifestyle interventions to treat elevated cardiometabolic risk in primary care (E-LITE): a randomized controlled trial | Inappropriate population |
| 6000 | Effects of an Individualized Comprehensive Rehabilitation Program on Impaired Postural Control in Women With Chronic Pelvic Pain: a Randomized Controlled Trial | Inappropriate population |
| 6001 | Comparing Effects and Neural Mechanisms of Tai Chi and Light-to-Moderate Intensity Aerobic Exercises | Inappropriate study design |
| 6002 | Ventricular-arterial coupling and acute levels of DHEA-S in breast cancer survivors after a short term physical exercise protocol | Inappropriate study design |
| 6003 | Effects of Pilates exercises on sensory interaction, postural control and fatigue in patients with multiple sclerosis | Inappropriate population |
| 6004 | Healthy Older People Everyday (HOPE) - Role of Healthy Diet on Muscle and Bone Health | Inappropriate study design |
| 6005 | Physical Activity as Intervention Against Age-related Loss of Muscle Mass and Function | Inappropriate study design |
| 6006 | Cytokine inhibition in chronic fatigue syndrome patients: study protocol for a randomized controlled trial | Inappropriate population |
| 6007 | Effect of 10-Week Supervised Moderate-Intensity Intermittent vs. Continuous Aerobic Exercise Programs on Vascular Adhesion Molecules in Patients with Heart Failure | Inappropriate population |

| | | |
|---|---|---|
| 6008 | Randomized controlled trial to evaluate the effects of progressive resistance training compared to progressive muscle relaxation in breast cancer patients undergoing adjuvant radiotherapy: the BEST study | Inappropriate study design |
| 6009 | Effect of high-intensity interval training combined with l-citrulline supplementation on functional capacities and muscle function in dynapenic-obese older adults | Inappropriate study design |
| 6010 | Exercise, brain plasticity, and depression | Inappropriate population |
| 6011 | Physical activity level ideal for independent functional status at discharge in intensive care patients | Inappropriate study design |
| 6012 | A multifactorial interdisciplinary intervention reduces frailty, increases function and is cost-effective in older adults who are frail: randomised controlled trial | Inappropriate study design |
| 6013 | Acute low-intensity cycling with blood-flow restriction has no effect on metabolic signaling in human skeletal muscle compared to traditional exercise | Inappropriate study design |
| 6014 | Evaluation of a Treatment Algorithm for Patients With Patellofemoral Pain Syndrome | Inappropriate study design |
| 6015 | Physical fitness and psychological benefits of strength training in community dwelling older adults | Inappropriate study design |
| 6016 | Comparison of strength training, aerobic training, and additional physical therapy as supplementary treatments for Parkinson's disease: pilot study | Inappropriate population |
| 6017 | Remotely supervised home-based aerobic exercise for Parkinson patients | Inappropriate intervention |
| 6018 | High-Intensity Interval Training and Moderate-Intensity Continuous Training in Ambulatory Chronic Stroke: feasibility Study | Inappropriate population |
| 6019 | Does the application of constraint-induced movement therapy during acute rehabilitation reduce arm impairment after ischemic stroke? | Inappropriate study design |
| 6020 | Recovering functional independence after a stroke through Modified Constraint-Induced Therapy | Inappropriate study design |
| 6021 | Safety and Efficacy of Xenical in Children and Adolescents With Obesity-Related Diseases | Inappropriate study design |
| 6022 | Short- and long-term impact of adapted physical activity and diet counseling during adjuvant breast cancer therapy: the "APAD1" randomized controlled trial | Inappropriate population |
| 6023 | Aerobic Training Efficacy in Inflammation, Neurotrophins, and Function in Chronic Stroke Persons: a Randomized Controlled Trial Protocol | Inappropriate outcome |
| 6024 | The effect of high intensity aerobic exercise training over two years on biologic aortic age | Inappropriate study design |
| 6025 | Walking adaptability therapy after stroke: study protocol for a randomized controlled trial | Inappropriate study design |
| 6026 | SaveMySkin: a pilot study about an internet-based self-help intervention for skin picking | Inappropriate population |
| 6027 | An evaluation of an online mindfulness programme for people with multiple sclerosis | Inappropriate population |
| 6028 | Sensory integration balance training in patients with multiple sclerosis: a randomized, controlled trial | Inappropriate study design |
| 6029 | Beneficial Effects of Acute Exercise on Executive Function in Adolescents | Inappropriate study design |
| 6030 | Impact of a combined multimodal-aerobic and multimodal intervention compared to standard aerobic treatment in breast cancer survivors with chronic cancer-related fatigue - results of a three-armed pragmatic trial in a comprehensive cohort design | Inappropriate study design |
| 6031 | Exploring the dose-response relationship between resistance exercise intensity and cognitive function | Inappropriate population |
| 6032 | Effects of a multicomponent exercise program in physical function and muscle mass in sarcopenic/pre-sarcopenic adults | Inappropriate study design |
| 6033 | A Multicenter, Randomized, Double-blind, Placebo-controlled Trial to Assess the Efficacy and Safety of Subcutaneous Histamine Dihydrochloride for Migraine Prophylaxis | Inappropriate study design |
| 6034 | Adaptive physical activity and back pain: a non-randomised community-based intervention trial | Inappropriate outcome |
| 6035 | The Effect of Very Low Calorie Diet With and Without Exercise on Muscle Synthesis in Middle-aged Overweight Male | Inappropriate population |
| 6036 | A Randomized Controlled Trial of High-Intensity Exercise and Executive Functioning in Cognitively Normal Older Adults | Inappropriate population |
| 6037 | The feasibility of a home-based, subject-controlled, interactive physical exercise program to promote cognitive health in older adults | Inappropriate population |
| 6038 | A perturbation-based balance training program for older adults: study protocol for a randomised controlled trial | Inappropriate population |
| 6039 | Influence of Fitness on Brain and Cognition | Inappropriate study design |

| | | |
|---|---|---|
| 6040 | Shared and differential neurocognitive effects comparing high-intensity interval training and active video-gaming in schizophrenia-results from an RCT | Inappropriate study design |
| 6041 | Short-term high-Intensity interval training increases systemic brain-derived neurotrophic factor (BDNF) in healthy women | Inappropriate population |
| 6042 | Short-term effects of a 3-week interval training program on heart rate variability in chronic heart failure. A randomised controlled trial | Inappropriate study design |
| 6043 | Physical activity promotion in the primary care setting in pre- and type 2 diabetes - the Sophia step study, an RCT | Inappropriate study design |
| 6044 | Combined effects of cerebellar transcranial direct current stimulation and transcutaneous spinal direct current stimulation on robot-assisted gait training in patients with chronic brain stroke: a pilot, single blind, randomized controlled trial | Inappropriate study design |
| 6045 | Effect of community based practice of Baduanjin on self-efficacy of adults with cardiovascular diseases | Inappropriate population |
| 6046 | A randomised, double-blind, placebo-controlled study to assess cardiac effects of single-dose BI 409306 in healthy male volunteers | Inappropriate study design |
| 6047 | Auricular Vagus Nerve Stimulation For Fibromyalgia Syndrome | Inappropriate study design |
| 6048 | Effect of Nutritional Supplementation and Pulmonary Rehabilitation on the Clinical Status of HF and COPD | Inappropriate population |
| 6049 | Glucose tolerance following a 6-week sleep extension protocol in overweight short sleepers | Inappropriate study design |
| 6050 | Recruitment strategies, design, and participant characteristics in a trial of weight-loss and metformin in breast cancer survivors | Inappropriate study design |
| 6051 | A randomized trial of aerobic exercise for major depression: examining neural indicators of reward and cognitive control as predictors and treatment targets | Inappropriate population |
| 6052 | A randomized trial of aerobic exercise on cognitive control in major depression | Inappropriate outcome |
| 6053 | Aerobic exercise enhances cognitive training effects in first episode schizophrenia: cognitive and functional gains and promising biological mechanisms of action | Inappropriate population |
| 6054 | Exercise, Cognitive Function and Neuroplasticity in Healthy Adults | Inappropriate study design |
| 6055 | Balance, dizziness and proprioception in patients with chronic whiplash associated disorders complaining of dizziness: a prospective randomized study comparing three exercise programs | Inappropriate study design |
| 6056 | Comparison of high- and low-intensity exercise training early after acute myocardial infarction | Inappropriate population |
| 6057 | Balance rehabilitation using custom-made Wii Balance Board exercises: clinical effectiveness and maintenance of gains in an acquired brain injury population | Inappropriate study design |
| 6058 | Change in health-related quality of life and social cognitive outcomes in obese, older adults in a randomized controlled weight loss trial: does physical activity behavior matter? | Inappropriate study design |
| 6059 | HABIT-ILE: a randomised trial of hand arm bimanual intensive training including lower extremity training for children with bilateral cerebral palsy | Inappropriate study design |
| 6060 | Effect of 24 sessions of high-intensity aerobic interval training carried out at either high or moderate frequency, a randomized trial | Inappropriate study design |
| 6061 | Impact of Carbohydrate Ingestion on Cognitive Flexibility and Cerebral Oxygenation during High-Intensity Intermittent Exercise: a Comparison between Maple Products and Usual Carbohydrate Solutions | Inappropriate study design |
| 6062 | Effects of a high-intensity progressive-cycle program on quality of life and motor symptomatology in a Parkinson's disease population: a pilot randomized controlled trial | Inappropriate study design |
| 6063 | The effect of trunk rehabilitation on gait performance after stroke-preliminary results | Inappropriate study design |
| 6064 | A psychological intervention to promote acceptance and adherence to non-invasive ventilation in people with chronic obstructive pulmonary disease: study protocol of a randomised controlled trial | Inappropriate population |
| 6065 | Balance Training on Vestibular Function and Proprioception Feedback in Stroke Patient | Inappropriate population |
| 6066 | The cyclical lower extremity exercise for Parkinson's trial (CYCLE): methodology for a randomized controlled trial | Inappropriate outcome |
| 6067 | The Effect of "Chair Yoga" on knee Joints pain &physical Function, Balance and Fear of Falling among Older Adults with knee Osteoarthritis | Inappropriate study design |
| 6068 | Feasibility and Preliminary Efficacy of Visual Cue Training to Improve Adaptability of Walking after Stroke: multi-Centre, Single-Blind Randomised Control Pilot Trial | Inappropriate study design |

| | | |
|---|---|---|
| 6069 | Cardiopulmonary response in spinal cord injury patients: effect of pneumatic compressive devices. Effect of methylphenidate on brain injury-related anger. The combined effects of clonidine and cyproheptadine with interactive training on the modulation of locomotion in spinal cord injured subjects | Inappropriate population |
| 6070 | YMCA commit to quit: randomized trial outcomes | Inappropriate study design |
| 6071 | Effect of High Protein Weight Loss for Seniors | Inappropriate study design |
| 6072 | FitForLife: study protocol for a randomized controlled trial | Inappropriate population |
| 6073 | Walk2Bactive: a randomized controlled trial of a physical activity-focused behavioural intervention beyond pulmonary rehabilitation in chronic obstructive pulmonary disease | Inappropriate population |
| 6074 | Experimental Effects of Acute Exercise on Prospective Memory and False Memory | Inappropriate study design |
| 6075 | Effects of Aerobic Exercise on Cognitive Function and Brain-derived Neurotrophic Factor in Young Adults | Inappropriate study design |
| 6076 | Aerobic exercise for cognitive deficits in schizophrenia - The impact of frequency, duration, and fidelity with target training intensity | Inappropriate study design |
| 6077 | Evaluation of the ACT on Health Program | Inappropriate study design |
| 6078 | Maraviroc to Augment Rehabilitation Outcomes After Stroke | Inappropriate study design |
| 6079 | The Effectiveness of Aerobic Exercise in the Treatment of Adults With Attention Deficit Hyperactivity Disorder (ADHD) | Inappropriate study design |
| 6080 | High-Intensity Interval Versus Moderate-Intensity Continuous Training in Individuals With Parkinson's Disease: hemodynamic and Functional Adaptation | Inappropriate study design |
| 6081 | Effect of disease duration in a randomized Phase III trial of rintatolimod, an immune modulator for Myalgic Encephalomyelitis/Chronic Fatigue Syndrome | Inappropriate population |
| 6082 | The effects of 8 weeks of inspiratory muscle training on the balance of healthy older adults: a randomized, double-blind, placebo-controlled study | Inappropriate study design |
| 6083 | Effectsofmoderate alcohol consumption and age on driving performance | Inappropriate study design |
| 6084 | Circuit Training on Vertigo, Oscillopsia and Dizziness in Vestibular Hypo Function Hypo- Functions" | Inappropriate study design |
| 6085 | Aerobic exercise training facilitates the effectiveness of cognitive behavioral therapy in panic disorder | Inappropriate study design |
| 6086 | Chronic low back pain: physical training, graded activity with problem solving training, or both? The one-year post-treatment results of a randomized controlled trial | Inappropriate population |
| 6087 | Dose-response study of aerobic exercise in older adults | Inappropriate study design |
| 6088 | Improvement of cognitive function by mental and/or individualized aerobic training in healthy elderly subjects | Inappropriate study design |
| 6089 | Multimodal exercise improves fitness and motor function in people with Huntington disease: results from a randomized, feasibility trial | Inappropriate population |
| 6090 | A Single Bout of Aerobic Exercise Reduces Anxiety Sensitivity But Not Intolerance of Uncertainty or Distress Tolerance: a Randomized Controlled Trial | Inappropriate study design |
| 6091 | Effects of moderate and high-intensity exercise on P300 latency and reaction time in athletes and nonathletes - An interim analysis | Inappropriate study design |
| 6092 | Management of persistent postconcussion symptoms in youth: a randomised control trial protocol | Inappropriate study design |
| 6093 | Aerobic training reduces stroke risk in patients with atrial fibrillation | Inappropriate population |
| 6094 | Exercise and Markers of Medial Temporal Health in Youth At-risk for Psychosis | Inappropriate study design |
| 6095 | Aerobic Exercise and Cerebrovascular Function | Inappropriate population |
| 6096 | Exercise for Brain Health in the Fight Against Alzheimer's Disease | Inappropriate study design |
| 6097 | Effect of high-intensity interval training and moderate-intensity continuous training on circulating micrornas in sedentary males | Inappropriate study design |
| 6098 | Dose Response Study of Aerobic Exercise in Older Adults | Inappropriate study design |
| 6099 | Pulmonary Rehabilitation During Acute Exacerbations of Chronic Obstructive Pulmonary Disease: a Mixed-methods Approach | Inappropriate study design |
| 6100 | Aerobic but not thermoregulatory gains following a 10-day moderate-intensity training protocol are fitness level dependent: a cross-adaptation perspective | Inappropriate outcome |
| 6101 | DEcision-making Capacity: intervention Development & Evaluation in Schizophrenia-spectrum Disorder | Inappropriate study design |
| 6102 | Aerobic exercise dose determines the cognitive gain in first-episode schizophrenia, so how do we encourage sufficient exercise? | Inappropriate population |
| 6103 | Effects of High Intensity }Training on health of younger with Obesity | Inappropriate population |

| | | |
|---|---|---|
| 6104 | Combined effects of remote ischemic preconditioning and aerobic exercise on sympathetic and cerebrovascular responses: a novel adaptation of blood flow restriction exercise | Inappropriate outcome |
| 6105 | Mindfulness, Education, and Exercise for age-related cognitive decline: study protocol, pilot study results, and description of the baseline sample | Inappropriate study design |
| 6106 | Improving Function in Older Veterans With Hospital-Associated Deconditioning | Inappropriate study design |
| 6107 | Lokomat's augmented performance feedback activities result in moderate intensity cardiovascular exercise in sub-acute stroke patients | Inappropriate study design |
| 6108 | Physiological effects of exercise on stroke survivors | Inappropriate population |
| 6109 | Effects of aerobic conditioning on cardiovascular sympathetic response to and recovery from challenge | Inappropriate study design |
| 6110 | HIIT enhances endurance performance and aerobic characteristics more than high-volume training in trained rowers | Inappropriate study design |
| 6111 | Lessons Learned: the Difficulties of Incorporating Intensity Principles Into Inpatient Stroke Rehabilitation | Inappropriate population |
| 6112 | High-Intensity Interval Training in Patients With Heart Failure With Reduced Ejection Fraction | Inappropriate study design |
| 6113 | Compare the Effectiveness of Modified Toy Cars Training With Various Intensity of Postural Combinations: a Randomized Controlled Trial | Inappropriate study design |
| 6114 | Eight Weeks of Aerobic Interval Training Improves Psychomotor Function in Patients with Parkinson's Disease-Randomized Controlled Trial | Inappropriate study design |
| 6115 | Short term aerobic exercise alters the reinforcing value of food in inactive adults | Inappropriate outcome |
| 6116 | Walking and balance outcomes for stroke survivors: a randomized clinical trial comparing body-weight-supported treadmill training with versus without challenging mobility skills | Inappropriate study design |
| 6117 | Stair walking is more energizing than low dose caffeine in sleep deprived young women | Inappropriate population |
| 6118 | Aerobic high-intensity intervals improve VO2max more than moderate training | Inappropriate study design |
| 6119 | Managing Chronic Obstructive Pulmonary Disease (COPD)Together: participant-CENTREd Continuum of Care (PCCC). A comparison of telephone health-mentoring, home-based walking and rehabilitation with rehabilitation only | Inappropriate study design |
| 6120 | Comparison of time-matched aerobic, resistance, or concurrent exercise training in older adults | Inappropriate outcome |
| 6121 | Trial of exercise on aging and memory (team study) | Inappropriate study design |
| 6122 | A RANDOMIZED CONTROLLED TRIAL ON THE EFFECT OF EXERGAMING AND SINGLE AEROBIC TRAINING ON COGNITIVE FUNCTIONING IN OLDER ADULTS WITH DEMENTIA | Inappropriate population |
| 6123 | A phase II, pragmatic, randomized clinical trial on a high-intensity exercise and fall prevention boot camp for Parkinson's disease: feasibility and safety | Inappropriate population |
| 6124 | Effects of Exercise Training on Cognitive Function and Neurotrophic Factors in Overweight Adults | Inappropriate study design |
| 6125 | Capturing the perspectives of women with coronary artery disease regarding interval training or continuous exercise in cardiac rehabilitation | Inappropriate study design |
| 6126 | The PERFORM-TAVR Trial | Inappropriate study design |
| 6127 | Efficacy of Lifestyle Changes in Modifying Practical Markers of Wellness and Aging | Inappropriate study design |
| 6128 | Effects of the six-month training on cognitive, physical performance, and daily physical activity in older adults | Inappropriate population |
| 6129 | Effects Resistance Training on Motor Control and Cognition | Inappropriate study design |
| 6130 | Moderate-intensity exercise boosts the N2 neural inhibition marker: a randomized and counterbalanced ERP study with precisely controlled exercise intensity | Inappropriate study design |
| 6131 | LONGITUDINAL ANALYSIS OF MICROSTRUCTURAL WHITE MATTER CHANGE IN MCI FOLLOWING A 6-MONTH AEROBIC EXERCISE INTERVENTION | Inappropriate study design |
| 6132 | Alterations in Aerobic Exercise Performance and Gait Economy Following High-Intensity Dynamic Stepping Training in Persons With Subacute Stroke | Inappropriate study design |
| 6133 | Dietary nitrate improves cerebral perfusion, in young adults during exercise: relationship to cognitive performance | Inappropriate population |
| 6134 | Sex differences in exercise associated changes on cognition post-stroke | Inappropriate study design |
| 6135 | A pain neuroscience education program for fibromyalgia patients with cognitive deficits: a case series | Inappropriate study design |
| 6136 | A Randomised Trial of Exercise Therapy for Parkinson’s Disease | Inappropriate population |

| | | |
|---|---|---|
| 6137 | Exercise Enhances: a randomized controlled trial on aerobic exercise as depression treatment augmentation strategy | Inappropriate study design |
| 6138 | Tai Chi Training in Stroke Survivors | Inappropriate study design |
| 6139 | Exercise training improves vascular function in patients with Alzheimer's disease | Inappropriate population |
| 6140 | Does a bout of strength training affect 2,000 m rowing ergometer performance and rowing-specific maximal power 24 h later? | Inappropriate study design |
| 6141 | Influence of combined functional resistance and endurance exercise over 12 weeks on matrix metalloproteinase-2 serum concentration in persons with relapsing-remitting multiple sclerosis - a community-based randomized controlled trial | Inappropriate outcome |
| 6142 | The practice of active rest by workplace units improves personal relationships, mental health, and physical activity among workers | Inappropriate population |
| 6143 | Effects of treatment intensity in upper limb robot-assisted therapy for chronic stroke: a pilot randomized controlled trial | Inappropriate study design |
| 6144 | Arm crank ergometry with blood flow restriction technique as a feasible strategy for improving hand function in chronic stroke survivors-A randomized controlled study | Inappropriate study design |
| 6145 | Gender disparities in cognitive, motor and mood outcomes: preliminary data from the bugher foundation's combined aerobic and resistance exercise training (CARET) program and cognitive training intervention (CTI) study | Inappropriate study design |
| 6146 | Workers Compensation Board: rotator Cuff Tear Management | Inappropriate study design |
| 6147 | Exercise Trial in Chronic Kidney Disease | Inappropriate study design |
| 6148 | The REACT (retirement in action) study | Inappropriate population |
| 6149 | Protocol for a systematically-developed, phase I/II, single-blind randomized controlled trial of treadmill walking exercise training effects on cognition and brain function in persons with multiple sclerosis | Inappropriate study design |
| 6150 | Effect of Individually Tailored Biopsychosocial Workplace Interventions on Chronic Musculoskeletal Pain and Stress Among Laboratory Technicians: randomized Controlled Trial | Inappropriate population |
| 6151 | Effects of carbohydrate and protein co-ingestion during short-term moderate-intensity exercise on cognitive function | Inappropriate outcome |
| 6152 | Heat & Aerobic Training (HEAT) Study | Inappropriate study design |
| 6153 | Acute emotional and psychophysiological effects of aerobic exercise | Inappropriate study design |
| 6154 | Trial of Exercise to Prevent HypeRtension in Young Adults | Inappropriate study design |
| 6155 | Running for extinction? Aerobic exercise as an augmentation of exposure therapy in panic disorder with agoraphobia | Inappropriate population |
| 6156 | The effects of acute exercise intensity on episodic and false memory among young adult college students | Inappropriate population |
| 6157 | Cognitive and Aerobic Resilience for the Brain | Inappropriate study design |
| 6158 | An evaluation of training responses using self-regulation in a residential rehabilitation program | Inappropriate population |
| 6159 | Motor Learning-based Wheelchair Propulsion Training for Older Adults | Inappropriate study design |
| 6160 | Vitamin E and aerobic exercise: effects on physical performance in older adults | Inappropriate study design |
| 6161 | The efficacy of vigorous-intensity exercise as an aid to smoking cessation in adults with elevated anxiety sensitivity: study protocol for a randomized controlled trial | Inappropriate study design |
| 6162 | Fit-Joint: getting Fit for Hip or Knee Replacement | Inappropriate study design |
| 6163 | Aortic and Systemic Arterial Stiffness Responses to Acute Exercise in Patients With Small Abdominal Aortic Aneurysms | Inappropriate population |
| 6164 | Heart rate regulation during cycle-ergometer exercise via event-driven biofeedback | Inappropriate population |
| 6165 | Efficacy of a mobile technology-based brisk walking program in improving dynamic balance and motor performance in people with Parkinson's disease-a randomized controlled trial | Inappropriate population |
| 6166 | Efficacy of classification-based cognitive functional therapy in patients with non-specific chronic low back pain: a randomized controlled trial | Inappropriate study design |
| 6167 | Combining aerobic exercise and cognitive training in chronic stroke; enhanced cognition predicted by IGF-1 | Inappropriate study design |
| 6168 | The impact of dietary restraint and moderate-intensity exercise on post-exercise energy intake in sedentary males | Inappropriate study design |
| 6169 | Preventive efforts in the aftermath of analogue trauma: the effects of Tetris and exercise on intrusive images | Inappropriate study design |

| | | |
|---|---|---|
| 6170 | A single bout of either sprint interval training or moderate intensity continuous training reduces anxiety sensitivity: a randomized controlled trial | Inappropriate population |
| 6171 | The effect of a physiotherapy exercise program with a self-management approach v's usual care on physical activity in people with mild-moderate Parkinson's disease: a Randomised Controlled Trial | Inappropriate study design |
| 6172 | Response to Endurance Exercise Training in Older Adults with Heart Failure with Preserved or Reduced Ejection Fraction | Inappropriate population |
| 6173 | Acute effects on cognitive performance following bouts of standing and light-intensity physical activity in a simulated workplace environment | Inappropriate study design |
| 6174 | Improving wellbeing and promoting healthy ageing among older adults: a randomized controlled trial comparing yoga and aerobic exercise | Inappropriate population |
| 6175 | The DEMO trial: a randomized, parallel-group, observer-blinded clinical trial of strength versus aerobic versus relaxation training for patients with mild to moderate depression | Inappropriate study design |
| 6176 | A comparison of the impact of physical exercise, cognitive training and combined intervention on spontaneous walking speed in older adults | Inappropriate study design |
| 6177 | Effect of reactive balance training on physical fitness poststroke: study protocol for a randomised non-inferiority trial | Inappropriate population |
| 6178 | Physical Activity Intervention for Patients With Reduced Physical Performance After Acute Coronary Syndrome | Inappropriate study design |
| 6179 | Investigating the role of dopamine on exercise and motor learning in healthy young adults | Inappropriate population |
| 6180 | Motor-Driven (Passive) Cycling: a Potential Physical Inactivity Countermeasure? | Inappropriate outcome |
| 6181 | CHEST 2019 Regional Congress Abstracts | Inappropriate outcome |
| 6182 | Cardiovascular rehabilitation soon after stroke using feedback-controlled robotics-assisted treadmill exercise: study protocol of a randomised controlled pilot trial | Inappropriate outcome |
| 6183 | Intensity-dependent impacts of exercise on cognitive functions in multiple sclerosis - preliminary results of a randomized controlled trial | Inappropriate study design |
| 6184 | Randomized, Controlled Trial of Exercise on Objective and Subjective Sleep in Parkinson's Disease | Inappropriate population |
| 6185 | Effects of resistance and all-round, functional training on quality of life, vitality and depression of older adults living in long-term care facilities: a 'randomized' controlled trial | Inappropriate population |
| 6186 | Impact of aerobic training on cardiovascular reactivity to and recovery from challenge | Inappropriate study design |
| 6187 | Yoga, Aerobic and Stretching Exercise Effects on Neurocognition | Inappropriate study design |
| 6188 | Rehabilitation in progressive supranuclear palsy: effectiveness of two multidisciplinary treatments | Inappropriate study design |
| 6189 | Self-paced resistance training and walking exercise in community-dwelling older adults: effects on neuromotor performance | Inappropriate study design |
| 6190 | Does exercise evoke neurological symptoms in healthy subjects? | Inappropriate intervention |
| 6191 | Maximal strength training enhances strength and functional performance in chronic stroke survivors | Inappropriate study design |
| 6192 | Effects of aerobic exercise on brain metabolism and greymatter volume in older adults: results of the smart trial | Inappropriate population |
| 6193 | Intravenous nonopioid analgesic drugs in chronic low back pain patients on chronic opioid treatment: a crossover, randomised, double-blinded, placebo-controlled study | Inappropriate study design |
| 6194 | Aerobic exercise promotes executive functioning and associated functional neuroplasticity | Inappropriate population |
| 6195 | Exercise training and cerebral blood flow in preclinical Alzheimer's disease: results from the aerobic exercise and cognitive health (REACH) study | Inappropriate population |
| 6196 | A pilot randomized controlled trial of exercise to improve cognitive performance in patients with stable glioma: a proof of concept | Inappropriate study design |
| 6197 | Effects of elastic band resistance training and nutritional supplementation on physical performance of institutionalised elderly--A randomized controlled trial | Inappropriate population |
| 6198 | Resistance training is associated with spontaneous changes in aerobic physical activity but not overall diet quality in adults with prediabetes | Inappropriate population |
| 6199 | THE EFFECT OF AEROBIC EXERCISE TRAINING ON SERUM BDNF IN PRECLINICAL ALZHEIMER'S DISEASE | Inappropriate study design |
| 6200 | Exercise improves reaction time without compromising accuracy in a novel easy-to-administer tablet-based cognitive task | Inappropriate population |
| 6201 | Virtual training leads to physical, cognitive and neural benefits in healthy adults | Inappropriate study design |

| | | |
|---|---|---|
| 6202 | Is sleep disturbance in patients with chronic pain affected by physical exercise or ACT-based stress management? - A randomized controlled study | Inappropriate outcome |
| 6203 | A six-month, supervised, aerobic and resistance exercise program improves self-efficacy in people with human immunodeficiency virus: a randomised controlled trial | Inappropriate intervention |
| 6204 | Effects of Simultaneous Balance and Resistance Training in Older Adults | Inappropriate study design |
| 6205 | Low-volume resistance training improves the functional capacity of older individuals with Parkinson's disease | Inappropriate study design |
| 6206 | Effects of a low-volume aerobic-type interval exercise on VO2max and cardiac mass | Inappropriate population |
| 6207 | Accelerometry measuring the outcome of robot-supported upper limb training in chronic stroke: a randomized controlled trial | Inappropriate study design |
| 6208 | The effects of cognitive and exercise training on cognitive and physical functions in older adults with different types of frailty - Exploration of the elderly with physical frailty alone and cognitive frailty | Inappropriate study design |
| 6209 | High intensity interval training and its effects on short-term memory in first year doctor of physical therapy students | Inappropriate study design |
| 6210 | Work volume is an important variable in determining the degree of inhibitory control improvements following resistance exercise | Inappropriate population |
| 6211 | Exercise and Cognitive Functioning in People With Chronic Whiplash-Associated Disorders: a Controlled Laboratory Study | Inappropriate study design |
| 6212 | Effect of single physical load of different duration and intensity on cognitive function | Inappropriate population |
| 6213 | Effect of Moderate Aerobic Exercise Training on Endothelial Function and Arterial Stiffness in CKD Stages 3-4: a Randomized Controlled Trial | Inappropriate population |
| 6214 | Effects of Acute Exercise and Learning Strategy Implementation on Memory Function | Inappropriate population |
| 6215 | Maintain Your Brain: protocol of a 3-Year Randomized Controlled Trial of a Personalized Multi-Modal Digital Health Intervention to Prevent Cognitive Decline among Community Dwelling 55 to 77 Year Olds | Inappropriate study design |
| 6216 | Exercise for methamphetamine dependence: rationale, design, and methodology | Inappropriate population |
| 6217 | Interaction of physical training with white matter lesion evolution: the hir-naktivitaet study | Inappropriate study design |
| 6218 | High speed resistance training and balance training for people with knee osteoarthritis to reduce falls risk | Inappropriate study design |
| 6219 | A randomised, controlled trial of a 12 week multi-modal exercise intervention in huntington's disease | Inappropriate population |
| 6220 | Active Class Space Metabolic Benefits Study | Inappropriate study design |
| 6221 | Enhancing consolidation of a rotational visuomotor adaptation task through acute exercise | Inappropriate population |
| 6222 | Postexercise Hypotension and Exercise Adherence | Inappropriate study design |
| 6223 | Four weeks of combination exercise training improved executive functions, episodic memory, and processing speed in healthy elderly people: evidence from a randomized controlled trial | Inappropriate population |
| 6224 | "The Effects of Clinical Pilates Exercises on Cognitive Functions and Speed of Movement in Healthy Young Adults" | Inappropriate study design |
| 6225 | Acute effects of exercise on cognitive functions and mental health | Inappropriate study design |
| 6226 | The impact of acute exercise on cognitive function in healthy older adults | Inappropriate population |
| 6227 | Aerobic Exercise Combined With Noninvasive Positive Pressure Ventilation Increases Serum Brain-Derived Neurotrophic Factor in Healthy Males | Inappropriate intervention |
| 6228 | Cognitive dysfunction in prefrailty baseline analysis of toyota preventional intervention for cognitive decline and sarcopenia (TOPICS) study | Inappropriate population |
| 6229 | A randomized controlled trial of exercise in spinal and bulbar muscular atrophy | Inappropriate study design |
| 6230 | The Efficacy of Vigorous-Intensity Exercise as an Aid to Smoking Cessation in Adults With High Anxiety Sensitivity: a Randomized Controlled Trial | Inappropriate population |
| 6231 | Health-Promotion Intervention Increases Self-Reported Physical Activity in Sub-Saharan African University Students: a Randomized Controlled Pilot Study | Inappropriate study design |
| 6232 | Comparisons of exercise dose and symptom severity between exercisers and nonexercisers in women during and after cancer treatment | Inappropriate population |
| 6233 | Cognition and Exercise Training | Inappropriate study design |
| 6234 | Cognitive remediation and physical exercise in multi-episode schizophrenia: study protocol for a randomised controlled trial | Inappropriate study design |

| | | |
|---|---|---|
| 6235 | HOW MANY PEOPLE AGED 55-77 YEARS ARE ELIGIBLE FOR LIFESTYLE INTERVENTIONS TO PREVENT COGNITIVE DECLINE? | Inappropriate study design |
| 6236 | Critical Care Cycling Study (CYCLIST) trial protocol: a randomised controlled trial of usual care plus additional in-bed cycling sessions versus usual care in the critically ill | Inappropriate outcome |
| 6237 | Early physical rehabilitation in intensive care patients with sepsis syndromes-a randomised controlled trial | Inappropriate study design |
| 6238 | Expanding treatment of chronic pain in older adults: community- based exercise + CBT | Inappropriate outcome |
| 6239 | Combined lumbar stabilization exercise with cognitive behavioral therapy gives no additional benefit for individuals with non-specific chronic low back pain | Inappropriate study design |
| 6240 | Effects of a psychosocial intervention programme combined with exercise in community-dwelling older adults with chronic pain: a randomized controlled trial | Inappropriate study design |
| 6241 | Effects of Exercise on Endothelial Function in Stroke Patients | Inappropriate study design |
| 6242 | Warm-up for Sprint Swimming: race-Pace or Aerobic Stimulation? A Randomized Study | Inappropriate study design |
| 6243 | Exploring the uptake and implementation of tele-monitored home-exercise programmes in adults with Parkinson's disease: a mixed-methods pilot study | Inappropriate population |
| 6244 | Validity and Reliability of an On-Court Fitness Test for Assessing and Monitoring Aerobic Fitness in Squash | Inappropriate study design |
| 6245 | Combined Effects of Isokinetic Training and Botulinum Toxin Type A on Spastic Equinus Foot in Patients with Chronic Stroke: a Pilot, Single-blind, Randomized Controlled Trial | Inappropriate population |
| 6246 | Effect of Aerobic Exercise Training on Balance, Walking Capacity, and Quality of Life in Sub-acute Stroke | Inappropriate population |
| 6247 | Mindfulness-Based Stress Reduction, Health Education and Exercise | Inappropriate study design |
| 6248 | Effects of resistance and functional-skills training on habitual activity and constipation among older adults living in long-term care facilities: a randomized controlled trial | Inappropriate population |
| 6249 | Effect of a 10-week MS exercise programme on muscle strength and aerobic capacity in inactive people with MS | Inappropriate study design |
| 6250 | Suicide Reduction in Schizophrenia via Exercise (SUnRISE): study protocol for a multi-site, single-blind, randomized clinical trial of aerobic exercise for suicide risk reduction in individuals with schizophrenia | Inappropriate population |
| 6251 | Cognitive functional therapy in patients with non-specific chronic low back pain-a randomized controlled trial 3-year follow-up | Inappropriate study design |
| 6252 | The effect of Taichi for college students on sleep disorders: a randomized control | Inappropriate study design |
| 6253 | Beneficial effects of short-term combination exercise training on diverse cognitive functions in healthy older people: study protocol for a randomized controlled trial | Inappropriate population |
| 6254 | Metabolic Reponses to a physical exercise session in women with excess body mass: randomized clinical trial | Inappropriate population |
| 6255 | Chronic Pain Self-Management Support With Pain Science Education and Exercise (COMMENCE) for People With Chronic Pain and Multiple Comorbidities: a Randomized Controlled Trial | Inappropriate study design |
| 6256 | Multimodal interventions to prevent and manage cognitive frailty | Inappropriate study design |
| 6257 | INSIGHT, A Comprehensive, Multidisciplinary Brain Training System | Inappropriate study design |
| 6258 | A pilot randomised controlled trial to assess the feasibility and acceptability of high intensity interval training (HIIT) among inpatients with severe mental illness (SMI) | Inappropriate study design |
| 6259 | The contributions of diet and exercise to improving knee osteoarthritis in overweight adults | Inappropriate population |
| 6260 | Participants' experiences of multidomain lifestyle-based intervention (finger) to prevent cognitive decline | Inappropriate study design |
| 6261 | Effects of exercise on cognition in people of various age groups | Inappropriate study design |
| 6262 | Virtual reality distraction induces hypoalgesia in patients with chronic low back pain: a randomized controlled trial | Inappropriate study design |
| 6263 | Is tailored strength training combined with anodal transcranial direct current stimulation effective in restoring paretic arm function in chronic stroke survivors? A randomized controlled trial | Inappropriate population |
| 6264 | Attention to breath sensations does not engage endogenous opioids to reduce pain | Inappropriate population |
| 6265 | No Sex Difference in Mental Fatigue Effect on High-Level Runners' Aerobic Performance | Inappropriate study design |

| | | |
|---|---|---|
| 6266 | Effect of various exercises on frailty among older adults with subjective cognitive concerns: a randomised controlled trial | Inappropriate study design |
| 6267 | Biobehavioral Bases & Management of Type 2 Diabetes | Inappropriate study design |
| 6268 | Rationale and study protocol for the 'eCoFit' randomized controlled trial: integrating smartphone technology, social support and the outdoor physical environment to improve health-related fitness among adults at risk of, or diagnosed with, Type 2 Diabetes | Inappropriate population |
| 6269 | Reducing risk factors for cognitive decline through psychological interventions: a pilot randomized controlled trial | Inappropriate study design |
| 6270 | A Network Approach to Understanding the Emotion Regulation Benefits of Aerobic Exercise | Inappropriate study design |
| 6271 | Does integrated cognitive and balance (dual-task) training improve balance and reduce falls risk in individuals with cerebellar ataxia? | Inappropriate study design |
| 6272 | Whole body vibration and treadmill training in Parkinson's disease rehabilitation: effects on energy cost and recovery phases | Inappropriate population |
| 6273 | Exercise Therapy for Parkinson's Disease: pedaling Rate Is Related to Changes in Motor Connectivity | Inappropriate study design |
| 6274 | Long-term effects of two psychological interventions on physical exercise and self-regulation following coronary rehabilitation | Inappropriate population |
| 6275 | Do Trials of Resistance Training to Improve Mobility After Stroke Adhere to the American College of Sports Medicine Guidelines? A Systematic Review | Inappropriate study design |
| 6276 | Maximizing the efficacy of interoceptive exposure by optimizing inhibitory learning: a randomized controlled trial | Inappropriate study design |
| 6277 | Disability in acute low back pain: the role of pain severity, pain catastrophizing, depression, and exercise | Inappropriate study design |
| 6278 | Influence of aerobic training on neurohormonal and hemodynamic responses to head-up tilt test and on autonomic nervous activity at rest and after exercise in patients after bypass surgery | Inappropriate population |
| 6279 | Effects of exercise training in water and on land in patients with COPD: a randomised clinical trial | Inappropriate population |
| 6280 | The effect of exercise in reducing desire to smoke and cigarette withdrawal symptoms is not caused by distraction | Inappropriate population |
| 6281 | High-Intensity Acute Exercise and Directed Forgetting on Memory Function | Inappropriate study design |
| 6282 | The HIMALAYAS Pilot Study | Inappropriate study design |
| 6283 | Task-specific training in Huntington disease: a randomized controlled feasibility trial | Inappropriate study design |
| 6284 | Work-HIIT: a time efficient work-place physical activity program | Inappropriate study design |
| 6285 | Locomotor training improves daily stepping activity and gait efficiency in individuals poststroke who have reached a "plateau" in recovery | Inappropriate intervention |
| 6286 | The effects of multidisciplinary rehabilitation on neuroimaging, biological, cognitive and motor outcomes in individuals with premanifest Huntington's disease | Inappropriate study design |
| 6287 | Effect on Falls Reduction of a Multimodal Intervention in Frail and Pre-frail Elderly Community-dwelling People | Inappropriate study design |
| 6288 | The management of chronic pain in older adults: randomized controlled trial of a physical and cognitive-behavioral intervention program | Inappropriate outcome |
| 6289 | Beneficial effects of YOGA and aerobic exercise on cognitive functions among person with post-stroke | Inappropriate population |
| 6290 | Comparative Effectiveness of mHealth-Supported Exercise Compared With Exercise Alone for People With Parkinson Disease: randomized Controlled Pilot Study | Inappropriate population |
| 6291 | Supervised and home-based exercise in patients newly diagnosed with multiple myeloma-a randomized controlled feasibility study | Inappropriate population |
| 6292 | Effects of traumatic brain injury on exercise capacity | Inappropriate study design |
| 6293 | A systematic review on the effectiveness of physical and rehabilitation interventions for chronic non-specific low back pain | Inappropriate outcome |
| 6294 | Adapted Yoga for Inactive Older Adults | Inappropriate study design |
| 6295 | Both aerobic exercise and cognitive-behavioral therapy reduce chronic fatigue in FSHD | Inappropriate study design |
| 6296 | The short-term effects of graded activity versus physiotherapy in patients with chronic low back pain: a randomized controlled trial | Inappropriate population |
| 6297 | Home-Based Exercise in Older Adults Recently Discharged From the Hospital for Cardiovascular Disease in China: randomized Clinical Trial | Inappropriate study design |
| 6298 | Pilot randomized-controlled phase-IIa trial on the prevention of comorbid depression and obesity in attention-deficit/ hyperactivity disorder | Inappropriate population |

| | | |
|---|---|---|
| 6299 | Effect of combined cognitive-behavioural therapy and endurance training on cortisol and salivary alpha-amylase in panic disorder | Inappropriate study design |
| 6300 | The effects of task-oriented circuit training and treadmill aerobic exercise training on cardiac risk and metabolic outcomes in chronic stroke survivors | Inappropriate study design |
| 6301 | Randomized trial of trunk training on trunk control and community participation in chronic stroke | Inappropriate study design |
| 6302 | A multicomponent intervention for the management of chronic pain in older adults: study protocol for a randomized controlled trial | Inappropriate study design |
| 6303 | STRATEGIES OF BRAIN MRI DATA ACQUISITION, QUALITY CONTROL AND ANALYSIS FOR THE MULTICENTER RISK REDUCTION FOR ALZHEIMER'S DISEASE (RRAD) CLINICAL TRIAL | Inappropriate study design |
| 6304 | Randomized Trial of Combined Aerobic, Resistance, and Cognitive Training to Improve Recovery From Stroke: feasibility and Safety | Inappropriate population |
| 6305 | Aerobic fitness: a target for intervention in Alzheimer's disease | Inappropriate study design |
| 6306 | Nurse practitioners can effectively deliver pain coping skills training to osteoarthritis patients with chronic pain: a randomized, controlled trial | Inappropriate population |
| 6307 | Stride management assist exoskeleton vs functional gait training in stroke: a randomized trial | Inappropriate population |
| 6308 | A Randomized Controlled Trial of the Effect of Early Upper-Limb Training on Stroke Recovery and Brain Activation | Inappropriate study design |
| 6309 | Cognitive functional therapy compared with a group-based exercise and education intervention for chronic low back pain: a multicentre randomised controlled trial (RCT) | Inappropriate outcome |
| 6310 | Comparison of Three Balance Training Protocols for Individuals Post Stroke | Inappropriate study design |
| 6311 | Examining what factors mediate treatment effect in chronic low back pain: a mediation analysis of a Cognitive Functional Therapy clinical trial | Inappropriate outcome |
| 6312 | High-Intensity Interval Training Increases Cardiac Output and VO2max | Inappropriate study design |
| 6313 | Efficacy of high-intensity exercise training on left ventricular ejection fraction in men with coronary artery disease (the Training Level Comparison Study) | Inappropriate study design |
| 6314 | Exercise Training Effects on Metabolic Syndrome: interactions With Medication | Inappropriate study design |
| 6315 | Effects of a high DHA multi-nutrient supplement and exercise on mobility and cognition in older women (MOBILE): a randomised semi-blinded placebo controlled study | Inappropriate study design |
| 6316 | Influence of exercise duration and hydration status on cognitive function during prolonged cycling exercise | Inappropriate population |
| 6317 | Effects of exercise training on myocardial blood flow reserve in patients with heart failure and left ventricular systolic dysfunction | Inappropriate study design |
| 6318 | Efficacy of an exercise intervention for employees with work-related fatigue: study protocol of a two-arm randomized controlled trial | Inappropriate population |
| 6319 | Reduction in cardiovascular disease risk factors: 6-month results from Project Active | Inappropriate intervention |
| 6320 | Effects of low-level laser therapy (LLLT 808 nm) on lower limb spastic muscle activity in chronic stroke patients | Inappropriate study design |
| 6321 | Influence of a Multimodal and Multimodal-Aerobic Therapy Concept on Health-Related Quality of Life in Breast Cancer Survivors | Inappropriate study design |
| 6322 | AML in the elderly patient | Inappropriate outcome |
| 6323 | Fit & Active Seniors Trial | Inappropriate study design |
| 6324 | Physiotherapy in Order to Improve Walking Capacity and Participation in Chronic Stroke Subjects | Inappropriate study design |
| 6325 | Effects of the HEART Camp Trial on Adherence to Exercise in Patients With Heart Failure | Inappropriate study design |
| 6326 | Exercise intensity and self-efficacy effects on anxiety reduction in healthy, older adults | Inappropriate study design |
| 6327 | Body weight-supported treadmill training is no better than overground training for individuals with chronic stroke: a randomized controlled trial | Inappropriate study design |
| 6328 | A randomised-controlled trial of the effects of very low-carbohydrate and high-carbohydrate diets on cognitive performance in patients with type 2 diabetes | Inappropriate study design |
| 6329 | Acceptance and commitment therapy improves exercise tolerance in sedentary women | Inappropriate population |
| 6330 | Preventing weight gain: the baseline weight related behaviors and delivery of a randomized controlled intervention in community based women | Inappropriate population |
| 6331 | Exercise training reduces the acute physiological severity of post-menopausal hot flushes | Inappropriate study design |

| | | |
|---|---|---|
| 6332 | Acute Effects of Aerobic Exercise and Active Videogames on Cognitive Flexibility, Reaction Time, and Perceived Exertion in Older Adults | Inappropriate study design |
| 6333 | The effect of dietary inorganic nitrate supplementation on cardiac function during submaximal exercise in men with heart failure with reduced ejection fraction (Hfref): a pilot study | Inappropriate outcome |
| 6334 | Effect of eccentric-based rehabilitation on hand tremor intensity in Parkinson disease | Inappropriate population |
| 6335 | EFFECTS OF SQUARE DANCE ON EPISODIC MEMORY AND HIPPOCAMPUS IN CHINESE OLDER ADULTS | Inappropriate study design |
| 6336 | Differential influences of exercise intensity on information processing in the central nervous system | Inappropriate study design |
| 6337 | Comparison of exercise strategies for motor symptom improvement in Parkinsons disease | Inappropriate population |
| 6338 | The effect of virtual reality exercises on pain, functionality, cardiopulmonary capacity and quality of life in fibromyalgia syndrome: a randomized, single-blind, controlled study | Inappropriate study design |
| 6339 | Refinement and clinical evaluation of the H-man arm robot for stroke rehabilitation: results of a feasibility clinical trial in post-stroke hemiplegia | Inappropriate study design |
| 6340 | Physical activity as intervention for age-related loss of muscle mass and function: protocol for a randomised controlled trial (the LISA study) | Inappropriate study design |
| 6341 | Effectiveness of Aquatic Exercises for Improving Mental and Physical Health in Adults | Inappropriate study design |
| 6342 | Effect of Various Exercises on Intrinsic Capacity in Older Adults With Subjective Cognitive Concerns | Inappropriate population |
| 6343 | Effects of Physical Exercise on Blood Glucose and Pain From the Musculoskeletal System in Patients With Type 2 Diabetes | Inappropriate study design |
| 6344 | Exercise, Smoking Cessation and Adolescents | Inappropriate study design |
| 6345 | Effect of Intensive Blood Pressure Control on Gait Speed and Mobility Limitation in Adults 75 Years or Older: a Randomized Clinical Trial | Inappropriate study design |
| 6346 | Exercise for Adults With Parkinson Disease | Inappropriate study design |
| 6347 | Aquatic Sequential Preparatory Approach and Severe Traumatic Brain Injury | Inappropriate study design |
| 6348 | exercise-induced muscle damage; prophylactic and therapeutic countermeasures | Inappropriate population |
| 6349 | Acute Exercise, Psychological Stress Induction, and Episodic Memory | Inappropriate population |
| 6350 | Experimental effects of acute exercise on episodic memory acquisition: decomposition of multi-trial gains and losses | Inappropriate study design |
| 6351 | The effects of aerobic exercise in Alzheimer's disease thalamo-cortical connections using diffusion tensor imaging | Inappropriate study design |
| 6352 | Acute aerobic exercise increases implicit approach motivation for dessert images | Inappropriate study design |
| 6353 | Pragmatic cluster randomised double-blind pilot and feasibility trial of an active behavioural physiotherapy intervention for acute non-specific neck pain: a mixed-methods protocol | Inappropriate outcome |
| 6354 | Does exercise improve progression free survival and quality of life in patients with glioblastoma? a trial in progress | Inappropriate study design |
| 6355 | Is it possible to enhance intensity in guided imagery exercises? An experimental study | Inappropriate study design |
| 6356 | Integrating virtual reality with activity management for the treatment of fibromyalgia: acceptability and preliminary efficacy | Inappropriate study design |
| 6357 | Effectiveness of a multicomponent treatment versus conventional treatment in patients with fibromyalgia: study protocol | Inappropriate population |
| 6358 | Exploring the effectiveness of an integrated exercise/CBT intervention for young men's mental health | Inappropriate study design |
| 6359 | Physical exercise interventions have no effect on serum BDNF concentration in older adults living in long-term nursing homes | Inappropriate study design |
| 6360 | Effect of intensive training with a spring-assisted hand orthosis on movement smoothness in upper extremity following stroke: a pilot clinical trial | Inappropriate study design |
| 6361 | Weight-loss and exercise for communities with arthritis in North Carolina (we-can): design and rationale of a pragmatic, assessor-blinded, randomized controlled trial | Inappropriate study design |
| 6362 | Effects of aerobic exercise on the cerebral white matter in Parkinson's disease determined by diffusion tensor imaging | Inappropriate study design |
| 6363 | The Limits of Cognitive Reappraisal: changing Pain Valence, but not Persistence, during a Resistance Exercise Task | Inappropriate study design |

| | | |
|---|---|---|
| 6364 | FCET2EC (From controlled experimental trial to = 2 everyday communication): how effective is intensive integrative therapy for stroke-induced chronic aphasia under routine clinical conditions? A study protocol for a randomized controlled trial | Inappropriate study design |
| 6365 | Multidisciplinary group rehabilitation versus individual physiotherapy for chronic nonspecific low back pain: a randomized trial | Inappropriate study design |
| 6366 | Effects of mental fatigue on exercise decision-making | Inappropriate study design |
| 6367 | Sitting Balance Exercise Performed Using Virtual Reality Training on a Stroke Rehabilitation Inpatient Service: a Randomized Controlled Study | Inappropriate study design |
| 6368 | Exercise, weight management, and DASH diet are superior to DASH diet alone for lowering blood pressure | Inappropriate population |
| 6369 | Results from 10 years of a CBT pain self-management outpatient program for complex chronic conditions | Inappropriate population |
| 6370 | Relearning of Writing Skills in Parkinson's Disease After Intensive Amplitude Training | Inappropriate study design |
| 6371 | Comparative Efficacy and Mechanisms of a Single-Session Pain Psychology Class in Chronic Low Back Pain: study Protocol for a Randomized Controlled Trial | Inappropriate study design |
| 6372 | Effect of a home based, low intensity, physical exercise program in older adults dialysis patients: a secondary analysis of the EXCITE trial | Inappropriate outcome |
| 6373 | Effect of Vaporized Cannabis on Exertional Breathlessness and Exercise Endurance in Advanced Chronic Obstructive Pulmonary Disease. A Randomized Controlled Trial | Inappropriate population |
| 6374 | Predictors and brain connectivity changes associated with arm motor function improvement from intensive robotic practice in chronic stroke | Inappropriate population |
| 6375 | mACTonHEALTH: psychological Flexibility and Activity Tracker - Protocol | Inappropriate study design |
| 6376 | Effect of Standing or Walking at a Workstation on Cognitive Function: a Randomized Counterbalanced Trial | Inappropriate study design |
| 6377 | Effect of exercise on neurodegenerative parkinsonism unmasked by dopamine blocking drugs (DRBD) | Inappropriate intervention |
| 6378 | Progressive mobility program and technology to increase the level of physical activity and its benefits in respiratory, muscular system, and functionality of ICU patients: study protocol for a randomized controlled trial | Inappropriate population |
| 6379 | Home-based versus center-based aerobic exercise on cardiopulmonary performance, physical function, quality of life and quality of sleep of overweight patients with chronic kidney disease | Inappropriate population |
| 6380 | Acute Exercise Improves Mood and Motivation in Young Men with ADHD Symptoms | Inappropriate study design |
| 6381 | Randomised controlled trial using a theory-based m-health intervention to improve physical activity and sleep health in adults: the Synergy Study protocol | Inappropriate study design |
| 6382 | High-intensity resistance training amplifies muscle hypertrophy and functional gains in persons with Parkinson's disease | Inappropriate study design |
| 6383 | Effects of robot-assisted gait training in patients with Parkinson's disease: study protocol for a randomized controlled trial 11 Medical and Health Sciences 1103 Clinical Sciences 11 Medical and Health Sciences 1109 Neurosciences | Inappropriate study design |
| 6384 | Multi-domain Versus Uni-Domain Training on Executive Control and Memory Functions of Older Adults | Inappropriate study design |
| 6385 | Acute cardiac and vascular effects of ozone inhalation in genetically susceptible subjects | Inappropriate study design |
| 6386 | Comparison of the effects of open vs. closed skill exercise on the acute and chronic BDNF, IGF-1 and IL-6 response in older healthy adults | Inappropriate population |
| 6387 | MoCog - influence of physical activity and training on motor and cognitive performance in older people | Inappropriate population |
| 6388 | Ischemic preconditioning does not improve peak exercise capacity at sea level or simulated high altitude in trained male cyclists | Inappropriate population |
| 6389 | Effects of treadmill training on the balance, functional capacity and quality of life in Parkinson's disease: a randomized clinical trial | Inappropriate population |
| 6390 | Multidisciplinary Program of Treatment of Obesity: evaluation of Cardiometabolic Risk Factors | Inappropriate study design |
| 6391 | Effects of aerobic dance on memory and hippocampus in older adults | Inappropriate study design |
| 6392 | Abstracts of the 46th annual meeting of Japanese Society of Clinical Neurophysiology | Inappropriate outcome |
| 6393 | Cognitive training combined with aerobic exercises in multiple sclerosis patients: a pilot study | Inappropriate population |

| | | |
|---|---|---|
| 6394 | Improving maintenance of physical activity in older, knee osteoarthritis patients trial-pilot (IMPACT-P): design and methods | Inappropriate population |
| 6395 | Lifestyle Intervention Study for Dementia Risk Reduction in Healthy Adults Aged Over 50 Years - The LEISURE study | Inappropriate study design |
| 6396 | Efficacy of two brief cognitive-behavioral rehabilitation programs for chronic neck pain: results of a randomized controlled pilot study | Inappropriate study design |
| 6397 | Effects of enjoyable exercise on cognitive function in older adults | Inappropriate study design |
| 6398 | Does adding cognitive-behavioural physiotherapy to exercise improve outcome in patients with chronic neck pain? A randomised controlled trial | Inappropriate study design |
| 6399 | Effects of aerobic exercise therapy and cognitive behavioural therapy on functioning and quality of life in amyotrophic lateral sclerosis: protocol of the FACTS-2-ALS trial | Inappropriate population |
| 6400 | Feasibility & Effect of a Tele-rehabilitation Program for Chronic Obstructive Pulmonary Disease vs. Standard Rehabilitation | Inappropriate study design |
| 6401 | Testing a path-analytic mediation model of how motivational enhancement physiotherapy improves physical functioning in pain patients | Inappropriate study design |
| 6402 | Individualised cognitive functional therapy compared with a combined exercise and pain education class for patients with non-specific chronic low back pain: a multicentre randomised controlled trial | Inappropriate study design |
| 6403 | Inspiratory Muscle Training On The Activities Of Daily Living And Quality Of Live After Stroke | Inappropriate study design |
| 6404 | Effects of Inspiratory Muscle Training On The Functional Gait Performance After Stroke | Inappropriate study design |
| 6405 | Influence of acute sprint interval training on cognitive performance in younger adults | Inappropriate population |
| 6406 | Physical capacity tasks in chronic low back pain: what is the contributing role of cardiovascular capacity, pain and psychological factors? | Inappropriate outcome |
| 6407 | Robot-Assisted Arm Training in Chronic Stroke: addition of Transition-to-Task Practice | Inappropriate study design |
| 6408 | Efficacy of phosphodiesterase type 5 inhibitors in univentricular congenital heart disease: the SV-INHIBITION study design | Inappropriate population |
| 6409 | Rationale and design of REWARD (revving-up exercise for sustained weight loss by altering neurological reward and drive): a randomized trial in obese endometrial cancer survivors | Inappropriate study design |
| 6410 | Specialized Pacing for Patients With Congenital Heart Disease | Inappropriate study design |
| 6411 | Cognitive function and blood-brain barrier permeability during exercise in the heat: effect of fitness and bovine colostrum supplementation | Inappropriate intervention |
| 6412 | Dietary nitrate supplementation improves sprint and high-intensity intermittent running performance | Inappropriate population |
| 6413 | Combining motivational and volitional strategies to promote unsupervised walking in patients with fibromyalgia: study protocol for a randomized controlled trial | Inappropriate study design |
| 6414 | Examining evidence based resistance plus balance training in community-dwelling older adults with complex health care needs: trial protocol for the Muscling Up Against Disability project | Inappropriate population |
| 6415 | Diurnal Cortisol Secretion Is Not Related to Multiple Sclerosis-Related Fatigue | Inappropriate population |
| 6416 | Mood and selective attention in the cold: the effect of interval versus continuous exercise | Inappropriate study design |
| 6417 | AEROBIC TRAINING IS RELATED TO COGNITIVE HEALTH IN ADULTS AT RISK FOR ALZHEIMER'S DISEASE: RESULTS FROM THE AEROBIC EXERCISE AND COGNITIVE HEALTH (REACH) PILOT STUDY | Inappropriate study design |
| 6418 | Intensive isotonic training modifies basal and exercise Doppler indexes of systolic function: a comparative study of athletes and sedentary men | Inappropriate population |
| 6419 | Preliminary efficacy and feasibility of referral to exercise specialists, psychologists and provision of a technology-based behavior change support package to promote physical activity in school teachers 'at risk' of, or diagnosed with, type 2 diabetes: the 'SMART Health' Pilot Study Protocol | Inappropriate study design |
| 6420 | RESTORE-Cognitive functional therapy with or without movement sensor biofeedback versus usual care for chronic, disabling low back pain: study protocol for a randomised controlled trial | Inappropriate population |
| 6421 | Aerobic exercise and cognitive behavior therapy reduce fatigue and slow progression of muscle MR fatty infiltration in FSHD | Inappropriate population |
| 6422 | The effects of neuromuscular electrical stimulation during repetitive transcranial magnetic stimulation before repetitive facilitation exercise on the hemiparetic hand in chronic stroke patients | Inappropriate intervention |

| | | |
|---|---|---|
| 6423 | Reduced-intensity modified constraint-induced movement therapy versus conventional therapy for upper extremity rehabilitation after stroke: a multicenter trial | Inappropriate study design |
| 6424 | The effect of graded activity and pain education (GAPE): an early post-surgical rehabilitation programme after lumbar spinal fusion - Study protocol for a randomized controlled trial | Inappropriate study design |
| 6425 | A randomised controlled trial (RCT) of telephone delivered cognitive behaviour therapy (TCBT) and exercise in the management of chronic widespread pain (CWP): identifying long-term outcome and who benefits from which treatment | Inappropriate population |
| 6426 | Does a 12Â- Week Exercise Program Improve Cognitive Function in Young Adults with Down Syndrome?: a randomised feasibility trial | Inappropriate study design |
| 6427 | Mental and physical benefits of yoga and tai chi compared to aerobic exercise | Inappropriate study design |
| 6428 | Electrical stimulation of skeletal muscles. An alternative to aerobic exercise training in patients with chronic heart failure? | Inappropriate study design |
| 6429 | Does exercise training improve cardiopulmonary fitness and daily physical activity in children and young adults with corrected tetralogy of Fallot or Fontan circulation? A randomized controlled trial | Inappropriate study design |
| 6430 | Does Task-Specific Training Improve Upper Limb Performance in Daily Life Poststroke? | Inappropriate study design |
| 6431 | Effects of a Robot-Assisted Arm Training Plus Hand Functional Electrical Stimulation on Recovery After Stroke: a Randomized Clinical Trial | Inappropriate study design |
| 6432 | Effects of a Community-based Group Rehabilitation Program for Dynamic Balance and Mobility Post Stroke | Inappropriate study design |
| 6433 | Handwriting training in Parkinson's disease: a trade-off between size, speed and fluency | Inappropriate study design |
| 6434 | Cardiopulmonary exercise testing and aerobic treadmill training after stroke: feasibility of a controlled trial | Inappropriate population |
| 6435 | Effects of interleukin-1 blockade with anakinra on aerobic exercise capacity in patients with heart failure and preserved ejection fraction (from the D-HART pilot study) | Inappropriate outcome |
| 6436 | Medium-term effects of a multimodal therapy on patients with fibromyalgia. Results of a controlled efficacy study | Inappropriate study design |
| 6437 | Unfavorable effects of resistance training on central arterial compliance: a randomized intervention study | Inappropriate study design |
| 6438 | Physiological and cognitive responses to caffeine during repeated, high-intensity exercise | Inappropriate study design |
| 6439 | Effectiveness of dual-task functional power training for preventing falls in older people: study protocol for a cluster randomised controlled trial | Inappropriate study design |
| 6440 | Effects of task-oriented robot training on arm function, activity, and quality of life in chronic stroke patients: a randomized controlled trial | Inappropriate population |
| 6441 | Do cardiorespiratory variables predict the antinociceptive effects of deep and slow breathing? | Inappropriate intervention |
| 6442 | Constraint-induced movement therapy during early stroke rehabilitation | Inappropriate outcome |
| 6443 | Exercise to prevent disability in older adults-the life study | Inappropriate study design |
| 6444 | Intense and unpredictable perturbations during gait training improve dynamic balance abilities in chronic hemiparetic individuals: a randomized controlled pilot trial | Inappropriate population |
| 6445 | Cognitive effects of adding caloric restriction to aerobic exercise training in older adults with obesity | Inappropriate study design |
| 6446 | Gait parameters associated with responsiveness to treadmill training with body-weight support after stroke: an exploratory study | Inappropriate study design |
| 6447 | Multimodal physical activity increases brain-derived neurotrophic factor levels and improves cognition in institutionalized older women | Inappropriate outcome |
| 6448 | A dissociation between propriospinal facilitation and inhibition after bilateral transcranial direct current stimulation | Inappropriate study design |
| 6449 | Video-guided exercise after stroke: a feasibility randomised controlled trial | Inappropriate population |
| 6450 | Erratum: physical exercise augmented cognitive behaviour therapy for older adults with generalised anxiety disorder (PEXACOG): study protocol for a randomized controlled trial (Trials (2019) 20 (174) DOI: 10.1186/s13063-019-3268-9) | Inappropriate study design |
| 6451 | Efficacy of modified constraint-induced movement therapy in chronic stroke: a single-blinded randomized controlled trial | Inappropriate study design |
| 6452 | Effect of Brain Stimulation on Stepping Performance in Stroke Survivors and Healthy Adults | Inappropriate intervention |

| | | |
|---|---|---|
| 6453 | The effects of an acute bout of exercise on neural activity in alcohol and cocaine craving: study protocol for a randomised controlled trial | Inappropriate population |
| 6454 | Testing a non-surgical physiotherapy-led alternative (Cognitive Functional Therapy) for people with knee osteoarthritis | Inappropriate population |
| 6455 | Group aerobic and strength training combined with social cognitive theory based education, results of the step it up RCT | Inappropriate population |
| 6456 | Exercise combined with Acceptance and Commitment Therapy (ExACT) compared to a supervised exercise programme for adults with chronic pain: study protocol for a randomised controlled trial | Inappropriate study design |
| 6457 | Physical Activity in Multiple Sclerosis (MS): a Novel Approach to Study Outcomes | Inappropriate study design |
| 6458 | Intensive Balance Training for Adults With Incomplete Spinal Cord Injuries: protocol for an Assessor-Blinded Randomized Clinical Trial | Inappropriate study design |
| 6459 | Impact of a Weight Management Intervention on Eating Competence: importance of Measurement Interval in Protocol Design | Inappropriate population |
| 6460 | Addition of intensive repetition of facilitation exercise to multidisciplinary rehabilitation promotes motor functional recovery of the hemiplegic lower limb | Inappropriate study design |
| 6461 | Transferability and sustainability of effects of dual-task training in patients with dementia | Inappropriate study design |
| 6462 | Montmorency Tart cherries (Prunus cerasus L.) modulate vascular function acutely, in the absence of improvement in cognitive performance | Inappropriate population |
| 6463 | Effect of Early and Intensive Rehabilitation after Ischemic Stroke on Functional Recovery of the Lower Limbs: a Pilot, Randomized Trial | Inappropriate population |
| 6464 | The predimed studies: a leap forward in nutrition | Inappropriate population |
| 6465 | Yoga reduces the brain's amplitude of low-frequency fluctuations in patients with early psychosis | Inappropriate study design |
| 6466 | Predictors of intensive care unit admission and mortality in patients with ischemic stroke: investigating the effects of a pulmonary rehabilitation program | Inappropriate study design |
| 6467 | Land Plus Aquatic Therapy Versus Land-Based Rehabilitation Alone for the Treatment of Freezing of Gait in Parkinson Disease: a Randomized Controlled Trial | Inappropriate study design |
| 6468 | Effects of whole-body vibration on body functions and structures, activity and participation in individuals with stroke: a randomized controlled trial | Inappropriate population |
| 6469 | Passive leg raising predicts volume responsiveness in patients with severe sepsis and septic shock | Inappropriate study design |
| 6470 | Experimental study on the learning effect of Yoga based on motion sensor | Inappropriate intervention |
| 6471 | Effectiveness of Multicomponent Physical Exercise in old adults after hospitalization: short vs long supervised programs | Inappropriate study design |
| 6472 | Longitudinal social cognitive influences on physical activity and sedentary time in Hispanic breast cancer survivors | Inappropriate study design |
| 6473 | The efficacy of moderate-intensity exercise as an aid for smoking cessation in women: a randomized controlled trial | Inappropriate study design |
| 6474 | Mental practice-based rehabilitation training to improve arm function and daily activity performance in stroke patients: a randomized clinical trial | Inappropriate study design |
| 6475 | Effect of exercise on cigarette cravings and ad libitum smoking following concurrent stressors | Inappropriate population |
| 6476 | Effects of board games combined with aerobic exercise on cognitive and physical functions in older adults with frailty | Inappropriate study design |
| 6477 | The influence of physical activity on mental well-being | Inappropriate population |
| 6478 | Strength training improves performance and pedaling characteristics in elite cyclists | Inappropriate study design |
| 6479 | Clinical assessment of the HELLODOC tele-rehabilitation service | Inappropriate study design |
| 6480 | Rehabilitation of older people with Parkinson's disease: an innovative protocol for RCT study to evaluate the potential of robotic-based technologies | Inappropriate population |
| 6481 | COACHING FOR COGNITION IN ALZHEIMER'S (COCOA) TRIAL | Inappropriate study design |
| 6482 | An educational approach based on a non-injury model compared with individual symptom-based physical training in chronic LBP. A pragmatic, randomised trial with a one-year follow-up | Inappropriate population |
| 6483 | A Dyadic Approach for a Remote Physical Activity Intervention in Adults With AD and Their Caregivers | Inappropriate population |
| 6484 | Cognitive Control and Physical Exercise | Inappropriate study design |
| 6485 | Early physical and occupational therapy in mechanically ventilated, critically ill patients: a randomised controlled trial | Inappropriate study design |

| | | |
|---|---|---|
| 6486 | Does Manipulation of Arterial Shear Stress Enhance Cerebrovascular Function and Cognition in the Aging Brain? | Inappropriate study design |
| 6487 | The effects of exercise on falls in elderly patients. A preplanned meta-analysis of the FICSIT Trials. Frailty and Injuries: cooperative Studies of Intervention Techniques | Inappropriate population |
| 6488 | Long-term multivitamin supplementation and cognitive function in men: a randomized trial | Inappropriate study design |
| 6489 | aeRobic Exercise and Cognitive Health | Inappropriate study design |
| 6490 | Web/Internet-based telemonitoring of a randomized controlled trial evaluating the time-integrated effects of a 24-week multicomponent intervention on key health outcomes in patients with fibromyalgia | Inappropriate population |
| 6491 | Concurrent Aerobic Exercise and Virtual Reality Cognitive Training | Inappropriate study design |
| 6492 | Effects ofaerobic exercise on the aging brain | Inappropriate population |
| 6493 | Maximal oxygen uptake after attenuation of cardiovascular drift during heat stress | Inappropriate population |
| 6494 | Watermelon Focused Dietary Inflammatory Index Intervention | Inappropriate study design |
| 6495 | DEMO-II trial. Aerobic exercise versus stretching exercise in patients with major depression-a randomised clinical trial | Inappropriate population |
| 6496 | Comparison in muscle damage between maximal voluntary and electrically evoked isometric contractions of the elbow flexors | Inappropriate study design |
| 6497 | Effectiveness of Physical Therapy- and Occupational Therapy-Based Rehabilitation in People Who Have Glioma and Are Undergoing Active Anticancer Treatment: single-Blind, Randomized Controlled Trial | Inappropriate study design |
| 6498 | Aerobic exercise alters brain structure and function in Parkinson's disease - A randomized controlled trial | Inappropriate population |
| 6499 | Singapore Physical Activity and Nutrition Study: a community-based physical and nutrition intervention for women aged 50 years and above in recreational settings | Inappropriate study design |
| 6500 | Impact of intensive physiotherapy on cognitive function after coronary artery bypass graft surgery | Inappropriate study design |
| 6501 | Exercise for Swallowing Problems After Stroke | Inappropriate study design |
| 6502 | Comparative efficacy of eight-week continuous, interval and combined aerobic training modes on selected haematological and biochemical variables of stroke survivors-a randomized clinical trial | Inappropriate population |
| 6503 | Sildenafil does not improve VO2max in subjects with diabetes despite augmenting non-invasively assessed central haemodynamics | Inappropriate study design |
| 6504 | Intensive group training versus cognitive intervention in sub-acute low back pain: short-term results of a single-blind randomized controlled trial | Inappropriate study design |
| 6505 | Effect of high-intensity interval training on cardiorespiratory fitness, physical activity and body composition in people with schizophrenia: a randomized controlled trial | Inappropriate population |
| 6506 | Effect of pedaling technique on muscle activity and cycling efficiency | Inappropriate population |
| 6507 | Maximal oxygen uptake (VO2) is markedly decreased in prader-willi syndrome (PWS) | Inappropriate study design |
| 6508 | The LIFE Study: lifestyle Interventions and Independence for Elders, Pilot | Inappropriate study design |
| 6509 | Effectiveness of Rotigotine plus intensive and goal-based rehabilitation versus Rotigotine alone in "de-novo" Parkinsonian subjects: a randomized controlled trial with 18-month follow-up | Inappropriate study design |
| 6510 | The effectiveness of a cognitive behavioral exercise approach (CBEA) compared to usual care in patients with a Whiplash Associated Disorder: a quasi-experimental clinical trial | Inappropriate population |
| 6511 | ES15.01 Management of Cognitive Changes in Oncology Patients | Inappropriate study design |
| 6512 | A randomized controlled trial to assess the central hemodynamic response to exercise in patients with transient ischaemic attack and minor stroke | Inappropriate study design |
| 6513 | Neuroplasticity benefits of adding aerobic exercise to cognitive training in first-episode Schizophrenia patients | Inappropriate study design |
| 6514 | Effects of aerobic interval training and continuous training on cellular markers of endothelial integrity in coronary artery disease: a SAINTEX-CAD substudy | Inappropriate population |
| 6515 | Internet-delivered computerized cognitive & affective remediation training for the treatment of acute and chronic posttraumatic stress disorder: two randomized clinical trials | Inappropriate study design |
| 6516 | Design of the PERSPECTIVE study: personalized speech therapy for actIVE conversation | Inappropriate population |
| 6517 | The effect of cognitive and cognitive-motor training on falls in older people | Inappropriate population |

| | | |
|---|---|---|
| 6518 | Cognitive control therapy and transcranial direct current stimulation for depression: a randomized, double-blinded, controlled trial | Inappropriate study design |
| 6519 | Effects of land-and water-based exercise interventions on pain in people with fibromyalgia: a preliminary report from the Al-Ã¡ndalus randomised controlled trial | Inappropriate study design |
| 6520 | Home-Based Exercise Program Improves Balance and Fear of Falling in Community-Dwelling Older Adults with Mild Alzheimer's Disease: a Pilot Study | Inappropriate study design |
| 6521 | Implementation of a stroke self-management program: a randomized controlled pilot study of veterans with stroke: a randomized controlled pilot study of veterans with stroke | Inappropriate study design |
| 6522 | Acute and long-term effects of pimobendan (UD-CG 115) in patients with heart failure (NYHA classes II and III). Results of a randomized multicenter double-blind study. AKUT- UND LANGZEITEFFEKTE VON PIMOBENDAN (UD-CG 115) BEI HERZINSUFFIZIENZ NYHA II UND III. ERGEBNISSE EINER RANDOMISIERTEN MULTIZENTRISCHEN DOPPELBLINDSTUDIE | Inappropriate outcome |
| 6523 | A cluster-randomised controlled trial of a physical activity and nutrition programme in retirement villages: a study protocol | Inappropriate study design |
| 6524 | Influence of a physical exercise program on cardiac remodeling and on functional capacity of patients with stroke | Inappropriate study design |
| 6525 | Exercise-based videogames for stroke rehabilitation at home: a single subject randomised trial | Inappropriate study design |
| 6526 | Cognitive Effects of Tai Chi Chuan in Healthy Older Adults and Older Adults | Inappropriate study design |
| 6527 | A Pilot RCT to Investigate the Use of a Home-based Nintendo Wii Program for Rehabilitation in Older Adults With Lower Limb Amputation | Inappropriate study design |
| 6528 | Analysis of fMRI and finger tracking training in subjects with chronic stroke | Inappropriate population |
| 6529 | Exercise increases caudate dopamine release and ventral striatal activation in Parkinson's disease | Inappropriate study design |
| 6530 | Effectiveness of a Multicomponent Exercise Program in the Attenuation of Frailty in Long-Term Nursing Frail Elders: a Randomised Clinical Controlled Trial | Inappropriate study design |
| 6531 | Acute Soy Supplementation Improves 20-km Time Trial Performance, Power, and Speed | Inappropriate study design |
| 6532 | Long-term myocardial adaptations after cardiac rehabilitation in heart failure: a randomized six-year evaluation using magnetic resonance imaging | Inappropriate study design |
| 6533 | Changes in physiological and stroking parameters during interval swims at the slope of the d-t relationship | Inappropriate population |
| 6534 | New Zealand blackcurrant extract attenuates enterocyte damage following exertional heat stress | Inappropriate study design |
| 6535 | Psychological interventions to reduce suicidality in high-risk patients with major depression: a randomized controlled trial | Inappropriate study design |
| 6536 | The muscle mass, omega-3, diet, exercise and lifestyle (MODEL) study - a randomised controlled trial for women who have completed breast cancer treatment | Inappropriate study design |
| 6537 | Changes in the Brain-Derived Neurotrophic Factor Are Associated with Improvements in Diabetes Risk Factors after Exercise Training in Adolescents with Obesity: the HEARTY Randomized Controlled Trial | Inappropriate study design |
| 6538 | Effect of aerobic exercise on blood lipid level &sleep in older adult people | Inappropriate population |
| 6539 | Endurance exercise training in older patients with heart failure: results from a randomized, controlled, single-blind trial | Inappropriate study design |
| 6540 | Aerobic exercise and cognitive functioning in schizophrenia: results of a 1-year follow-up from a randomized controlled trial | Inappropriate outcome |
| 6541 | The IDEAL Randomised Controlled Trial | Inappropriate study design |
| 6542 | Nordic Walking and Walking in Parkinson's disease: a randomized single-blind controlled trial | Inappropriate study design |
| 6543 | Transcranial magnetic stimulation and aerobic exercise increase BDNF-TRKB signaling in Parkinson's disease | Inappropriate study design |
| 6544 | Culturally sensitive group therapy for Turkish patients suffering from chronic pain: a randomised controlled intervention trial | Inappropriate outcome |
| 6545 | The effect of exercise, cognitive therapy, and nutritional counseling in treating bulimia nervosa | Inappropriate study design |
| 6546 | Effects of 16-week Bikram yoga program on heart rate variability and cardiovascular disease risk factors in sedentary, stressed adults | Inappropriate study design |
| 6547 | Intervention to Improve Quality of Sleep of Palliative Patient Carers in the Community: clinical Trial | Inappropriate population |
| 6548 | Music meets robotics: a prospective randomized study on motivation during robot aided therapy | Inappropriate study design |

| | | |
|---|---|---|
| 6549 | The impact of 2 years of high-intensity exercise training on a model of integrated cardiovascular regulation | Inappropriate population |
| 6550 | Dance program for older adults | Inappropriate population |
| 6551 | Older adultsâ€™ perceptions of the usefulness of technologies for engaging in physical activity: using focus groups to explore physical literacy | Inappropriate study design |
| 6552 | Particulate Matter, Blood Pressure, and the Sympathetic Nervous System | Inappropriate population |
| 6553 | A heart for running-safety considerations on aerobic exercise in the subacute phase of stroke-data from the phys-stroke trial | Inappropriate study design |
| 6554 | Can we modify the functional and cognitive trajectory in a hospital with exercise? A randomized clinical trial | Inappropriate study design |
| 6555 | Efficacy of Brisk Walking in Parkinson's Disease | Inappropriate population |
| 6556 | Aerobic Exercise for Older Adults at Increased Risk of Alzheimer's Disease and Related Dementias | Inappropriate study design |
| 6557 | The effectiveness of a stratified group intervention using the STarTBack screening tool in patients with LBP--a non randomised controlled trial | Inappropriate study design |
| 6558 | Psychological features and outcomes of the Back School treatment in patients with chronic non-specific low back pain. A randomized controlled study | Inappropriate study design |
| 6559 | Individualised cognitive functional therapy compared with a combined exercise and pain education class for patients with non-specific chronic low back pain: study protocol for a multicentre randomised controlled trial | Inappropriate study design |
| 6560 | Functional outcomes can vary by dose: learning-based sensorimotor training for patients stable poststroke | Inappropriate study design |
| 6561 | How low is too low? Analysis of systolic and diastolic blood pressure in patients with and without prior cardiovascular disease in the sprint trial | Inappropriate population |
| 6562 | The efficacy of SMART Arm training early after stroke for stroke survivors with severe upper limb disability: a protocol for a randomised controlled trial | Inappropriate study design |
| 6563 | Effects of constraint-induced therapy versus bilateral arm training on motor performance, daily functions, and quality of life in stroke survivors | Inappropriate study design |
| 6564 | Sensory Attention Focused Exercise in Parkinson's Disease | Inappropriate study design |
| 6565 | Comparing the Effectiveness of Cognitive Functional Treatment and Lumbar Stabilization Treatment on Pain and Movement Control in Patients With Low Back Pain | Inappropriate population |
| 6566 | Usefulness of C-Reactive Protein Plasma Levels to Predict Exercise Intolerance in Patients With Chronic Systolic Heart Failure | Inappropriate population |
| 6567 | A tailored exercise program versus general exercise for a subgroup of patients with low back pain and movement control impairment: a randomised controlled trial with one-year follow-up | Inappropriate population |
| 6568 | The elucidation of the brain networks for the effects of high-intensity interval training on cognitive function | Inappropriate study design |
| 6569 | Manual and manipulative therapy in addition to rehabilitation for osteoarthritis of the knee: assessor-blind randomized pilot trial | Inappropriate study design |
| 6570 | The Effects of Two Different Types of Exercise Programs to Increase Physical Activity in Patients With MS | Inappropriate study design |
| 6571 | Protocol for a pragmatic randomised controlled trial of Body Brain Life-General Practice and a Lifestyle Modification Programme to decrease dementia risk exposure in a primary care setting | Inappropriate study design |
| 6572 | Preventing chronic back pain: evaluation of a theory-based cognitive-behavioural training programme for patients with subacute back pain | Inappropriate study design |
| 6573 | Chronic fatigue syndrome: an evaluation of an occupational therapy inpatient intervention | Inappropriate study design |
| 6574 | Effects of sling and voluntary constraint during constraint-induced movement therapy for the arm after stroke: a randomized, prospective, single-centre, blinded observer rated study | Inappropriate intervention |
| 6575 | Assessment of the effects of aquatic therapy on global symptomatology in patients with fibromyalgia syndrome: a randomized controlled trial | Inappropriate study design |
| 6576 | Body weight supported treadmill training versus traditional training in patients dependent on walking assistance after stroke: a randomized controlled trial | Inappropriate population |
| 6577 | Exercise, Age-Related Memory Decline, And Hippocampal Function | Inappropriate study design |
| 6578 | Effects of Moderate Aerobic Exercise on Cognitive Abilities and Redox State Biomarkers in Older Adults | Inappropriate population |
| 6579 | Constraint-induced therapy of chronic aphasia after stroke | Inappropriate population |
| 6580 | Dynamic Lycra Orthosis as an Adjunct to Botulinum Toxin-A Injection for Post-stroke Spasticity | Inappropriate study design |
| 6581 | The effects of a worksite chronic disease prevention program | Inappropriate population |

| | | |
|---|---|---|
| 6582 | The BDNF val66met polymorphism and changes in rumination and negative affect after a 6-month aerobic exercise randomized controlled intervention: a secondary analysis | Inappropriate study design |
| 6583 | Virtual Reality Social Prediction Improvement and Rehabilitation Intensive Training (VR-SPIRIT) for paediatric patients with congenital cerebellar diseases: study protocol of a randomised controlled trial | Inappropriate study design |
| 6584 | The relationship among exercise habit, cognitive functions, and mental health | Inappropriate study design |
| 6585 | ACTIVE: advanced Cognitive Training for Independent and Vital Elderly | Inappropriate study design |
| 6586 | Treatment expectancy and credibility are associated with the outcome of both physical and cognitive-behavioral treatment in chronic low back pain | Inappropriate study design |
| 6587 | Measurement of physical performance and objective fatigability in people with mild-to-moderate traumatic brain injury | Inappropriate population |
| 6588 | The Effect of a Novel Dual-Task Exercise Program for Balance, Mobility and Cognition in Community Dwelling Older Adults | Inappropriate study design |
| 6589 | Study protocol of a randomized intervention study to explore effects of a pure physical training and a mind–body exercise on cognitive executive function in independent living adults age 65–85 | Inappropriate population |
| 6590 | Preliminary effects of exposure-based cognitive behavior therapy for children with pain predominant functional gastrointestinal disorders | Inappropriate population |
| 6591 | Physiological and Performance Effects of Caffeine Gum Consumed During a Simulated Half-Time by Professional Academy Rugby Union Players | Inappropriate study design |
| 6592 | Cost-effectiveness of "Tele-Square Step Exercise" for falls prevention in fibromyalgia patients | Inappropriate population |
| 6593 | The SHED-IT community trial study protocol: a randomised controlled trial of weight loss programs for overweight and obese men | Inappropriate study design |
| 6594 | The effectiveness of multi-component interventions targeting physical activity or sedentary behaviour amongst office workers: a three-arm cluster randomised controlled trial | Inappropriate study design |
| 6595 | Could transcranial direct current stimulation potentiate the effect of a strenghthening program in chronic stroke survivors? preliminary results | Inappropriate study design |
| 6596 | Oxygen consumption, oxygen cost, heart rate, and perceived effort during split-belt treadmill walking in young healthy adults | Inappropriate population |
| 6597 | A randomised controlled cross-over trial of aerobic training versus Qigong in advanced Parkinson's disease | Inappropriate study design |
| 6598 | Modulating bdnf activity in Parkinson's disease: the impact of aerobic exercise and transcranial magnetic stimulation | Inappropriate population |
| 6599 | Stroke patients' and non-professional coaches' experiences with home-based constraint-induced movement therapy: a qualitative study | Inappropriate population |
| 6600 | Low frequency electrical muscle stimulation and endothelial function in advanced heart failure patients | Inappropriate study design |
| 6601 | Comparison of spinal fusion and nonoperative treatment in patients with chronic low back pain: long-term follow-up of three randomized controlled trials | Inappropriate study design |
| 6602 | Sustaining self-regulatory efficacy and psychological outcome expectations for postnatal exercise: effects of a group-mediated cognitive behavioural intervention | Inappropriate study design |
| 6603 | Enhancing Function in Later Life: exercise and Functional Network Connectivity | Inappropriate study design |
| 6604 | Enhanced Exercise Therapy in Parkinson's disease: a comparative effectiveness trial | Inappropriate study design |
| 6605 | Exercise in Patients on Dialysis: a Multicenter, Randomized Clinical Trial | Inappropriate population |
| 6606 | Exercise response in Parkinson's disease: insights from a cross-sectional comparison with sedentary controls and a per-protocol analysis of a randomised controlled trial | Inappropriate population |
| 6607 | Effects of a Home-Based Telephone-Supported Physical Activity Program for Older Adult Veterans With Chronic Low Back Pain | Inappropriate population |
| 6608 | Interim analysis of randomized controlled trial to maximize motor scores of the functional independence measure among patients following stroke in acute rehabilitation | Inappropriate study design |
| 6609 | Effect of virtual reality training on walking distance and physical fitness in individuals with Parkinson's disease | Inappropriate population |
| 6610 | Combined intervention approaches for initiating and maintaining physical activity in depressed individuals: design and rationale of the project MOVE randomized clinical trial | Inappropriate study design |

| | | |
|---|---|---|
| 6611 | Corrigendum: the impact of aerobic exercise on fronto-parietal network connectivity and its relation to mobility: an exploratory analysis of a 6-month randomized controlled trial (Front. Hum. Neurosci, 2017, 11, 344, 10.3389/fnhum.2017.00344) | Inappropriate study design |
| 6612 | Impact of mHealth technology on adherence to healthy PA after stroke: a randomized study | Inappropriate study design |
| 6613 | Determining the Feasibility of Implementing a Combined Cognitive and Exercise Training Program for Older Adults in a Community Setting | Inappropriate study design |
| 6614 | Results of a long-term training program after acute myocardial infarction | Inappropriate study design |
| 6615 | Occupation-based intervention versus rote exercise in modified constraint-induced movement therapy for patients with median and ulnar nerve injuries: a randomized controlled trial | Inappropriate population |
| 6616 | Optimizing a Positive Psychology Intervention to Promote Health Behaviors After an Acute Coronary Syndrome: the Positive Emotions After Acute Coronary Events III (PEACE-III) Randomized Factorial Trial | Inappropriate study design |
| 6617 | The efficacy of Psycho-Physical Intervention for obesity and weight-reduction among women- A randomized controlled trial | Inappropriate study design |
| 6618 | Fibromyalgia Integrative Training for Adolescents With Juvenile Fibromyalgia | Inappropriate population |
| 6619 | Immunoadsorption therapy for dilated cardiomyopathy using tryptophan column-A prospective, multicenter, randomized, within-patient and parallel-group comparative study to evaluate efficacy and safety | Inappropriate population |
| 6620 | Twelve weeks of nightly stretch does not reduce thumb web-space contractures in people with a neurological condition: a randomised controlled trial | Inappropriate population |
| 6621 | Low-intensity internet-delivered treatment for generalized anxiety symptoms in routine care: protocol for a randomized controlled trial | Inappropriate outcome |
| 6622 | Assessing the feasibility of a supervised exercise rehabilitation intervention with behavioural and motivational support, for people with postural orthostatic tachycardia syndrome | Inappropriate population |
| 6623 | A culturally adapted physical activity intervention for Latinas: a randomized controlled trial | Inappropriate population |
| 6624 | Intervention-related cognitive versus social mediators of exercise adherence in the elderly | Inappropriate study design |
| 6625 | Aerobic Exercise Reduces Symptoms of Posttraumatic Stress Disorder: a Randomized Controlled Trial | Inappropriate study design |
| 6626 | Effects of Yoga, Strength Training and Aerobic Exercise on Cognition in Adult Cancer Survivors | Inappropriate study design |
| 6627 | Yoga and Aerobic Exercise Effects on Cognition | Inappropriate study design |
| 6628 | Combining physical exercise and cognitive training in old age | Inappropriate study design |
| 6629 | Comparing stroke rehabilitation outcomes between acute inpatient and nonintense home settings | Inappropriate study design |
| 6630 | Neuroplasticity-based cognitive training in ultra-high risk adolescents and recent onset patients with schizophrenia | Inappropriate study design |
| 6631 | Cardiovascular rehabilitation adapted to transient ischaemic attack and stroke | Inappropriate population |
| 6632 | Correction: effect of aerobic exercise on cognition in younger adults: a randomized clinical trial: a randomized clinical trial (Neurology (2019) 92 (e905-e916) DOI: 10.1212/WNL.0000000000007003) | Inappropriate population |
| 6633 | Individualized Cognition-Action intervention to prevent behavioral disturbances and functional decline in institutionalized older adults: a randomized pilot trial | Inappropriate study design |
| 6634 | Aerobic exercise and cognitive therapy in the treatment of dysphoric moods | Inappropriate study design |
| 6635 | The effects of aerobic exercise and stretching on self-reported sleep quality in community dwelling older adults with cognitive complaints: a randomized, controlled trial | Inappropriate population |
| 6636 | Effect of Taichi softball on function-related outcomes in older adults: a randomized control trial | Inappropriate study design |
| 6637 | COMPUTERIZED COGNITIVE TRAINING IMPROVES PERFORMANCE ON EXECUTIVE FUNCTION IN COMMUNITY-DWELLING OLDER ADULTS: AN 8-WEEK RANDOMIZED CONTROLLED TRIAL | Inappropriate study design |
| 6638 | The effect of high-intensity exercise on objective sleep outcomes in parkinson's disease | Inappropriate population |
| 6639 | Virtual reality based rehabilitation speeds up functional recovery of the upper extremities after stroke: a randomized controlled pilot study in the acute phase of stroke using the rehabilitation gaming system | Inappropriate study design |
| 6640 | Cybercycling Effects on Classroom Behavior in Children With Behavioral Health Disorders: an RCT | Inappropriate population |

| | | |
|---|---|---|
| 6641 | High Frequency and Intensity Rehabilitation in 641 Subacute Ischemic Stroke Patients | Inappropriate population |
| 6642 | Evaluating an Online Cognitive Training Platform for Older Adults: user Experience and Implementation Requirements | Inappropriate intervention |
| 6643 | Comparing unilateral and bilateral upper limb training: the ULTRA-stroke program design | Inappropriate population |
| 6644 | Manual treatment for cervicogenic headache and active trigger point in the sternocleidomastoid muscle: a pilot randomized clinical trial | Inappropriate population |
| 6645 | The effects of different gait training techniques for improving walking performance in patients in the early stages of Parkinson's Disease: a pilot randomized clinical trial | Inappropriate population |
| 6646 | EXPLORING INDIVIDUAL VARIABILITY IN EXERCISE-INDUCED COGNITIVE CHANGE: THE ROLE OF CARDIORESPIRATORY FITNESS AND APOLIPOPROTEIN Îµ4 ALLELE CARRIAGE | Inappropriate population |
| 6647 | Effects of a 6-month self-rehabilitation programme in addition to botulinum toxin injections and conventional physiotherapy on limitations of patients with spastic hemiparesis following stroke (ADJU-TOX): protocol study for a randomised controlled, investigator blinded study | Inappropriate study design |
| 6648 | THE PROMOTING ACTIVITY, INDEPENDENCE AND STABILITY IN EARLY DEMENTIA (PRAISED) RESEARCH PROGRAMME | Inappropriate population |
| 6649 | Locomotor Adaptation Training to Prevent Mobility Disability | Inappropriate study design |
| 6650 | Delirium, sub-syndromal delirium, antipsychotic use, and associated clinical outcomes in critically ill mechanically ventilated patients enrolled in the sleap multicenter randomized trial | Inappropriate study design |
| 6651 | Evaluation of quality of life in elderly healthy subjects after aerobic and/or mental training | Inappropriate study design |
| 6652 | Physical activity in early-onset dementia | Inappropriate study design |
| 6653 | Exogenous Ketone Salts Do Not Improve Cognitive Performance During a Dual-Stress Challenge | Inappropriate population |
| 6654 | The Aerobic and Cognitive Training System | Inappropriate population |
| 6655 | The Effects of Computerized Cognitive Training with and without Physical Exercise on Cognitive Function in Older Adults: an 8-week Randomized Controlled Trial | Inappropriate study design |
| 6656 | Linking neuroplastic effects to behavioral changes after balance training in Parkinson's disease: a study protocol of a randomized controlled trial | Inappropriate study design |
| 6657 | Aerobic exercise as add-on treatment in depressed inpatients improves cognitive symptoms but not symptom severity | Inappropriate study design |
| 6658 | Randomised control trial to evaluating the effectiveness of the Hello Sunday Morning "Daybreak" program in reducing alcohol use by adults | Inappropriate study design |
| 6659 | Exploring effects of aerobic exercise and mindfulness training on cognitive function in older adults at risk of dementia: the active minds study | Inappropriate population |
| 6660 | Changes in brain inflammation are related to symptoms and aerobic exercise in first episode schizophrenia | Inappropriate outcome |
| 6661 | ENHANCING RECRUITMENT SUCCESS FOR AEROBIC EXERCISE TRIALS IN ALZHEIMER'S DEMENTIA | Inappropriate study design |
| 6662 | A feasibility study of a randomised controlled trial to examine the impact of the ABCDE bundle on quality of life in ICU survivors | Inappropriate population |
| 6663 | PEAK CARDIORESPIRATORY RESPONSES TO SYMPTOM-LIMITED CARDIOPULMONARY EXERCISE TESTING IN PERSONS WITH ALZHEIMER'S DISEASE | Inappropriate outcome |
| 6664 | Long-term Exercise After Pulmonary Rehabilitation (LEAP): design and rationale of a randomized controlled trial of Tai Chi | Inappropriate study design |
| 6665 | Exercise and weight loss in obese older adults with knee osteoarthritis: a preliminary study | Inappropriate study design |
| 6666 | Individual responsiveness to physical exercise intervention in acutely hospitalized older adults | Inappropriate population |
| 6667 | What are the clinical, methodological, logistical and operational challenges when designing and conducting a randomised controlled trial of a group based intervention for the treatment of osteoarthritis and how can they be managed? | Inappropriate study design |
| 6668 | Resistance versus Balance Training to Improve Postural Control in Parkinson's Disease: a Randomized Rater Blinded Controlled Study | Inappropriate study design |
| 6669 | Feasibility and Effectiveness of Remote Virtual Reality-Based Cardiac Rehabilitation | Inappropriate study design |

| | | |
|---|---|---|
| 6670 | Auditory-based targeted cognitive training improves verbal learning and reduces auditory hallucinations in chronic schizophrenia patients in locked residential care | Inappropriate study design |
| 6671 | Functional Engagement in Assisted Therapy Through Exercise Robotics | Inappropriate study design |
| 6672 | Pain education and exercise program supported by cell phone for older adults with low back pain | Inappropriate population |
| 6673 | Neck exercises, physical and cognitive behavioural-graded activity as a treatment for adult whiplash patients with chronic neck pain: design of a randomised controlled trial | Inappropriate study design |
| 6674 | Intense lower limb training enhances lower limb function and intense upper limb training enhances upper limb function following stroke | Inappropriate population |
| 6675 | Tinnitus: evaluation of biofeedback and stomatognathic treatment | Inappropriate population |
| 6676 | RISK REDUCTION IN ALZHEIMER'S DISEASE (RRAD): RATIONALE AND METHODS FOR A MULTICENTER CLINICAL TRIAL ASSESSING EXERCISE AND INTENSIVE VASCULAR RISK REDUCTION FOR THE PREVENTION OF DEMENTIA | Inappropriate study design |
| 6677 | Rationale and methods for a multicenter clinical trial assessing exercise and intensive vascular risk reduction in preventing dementia (rrAD Study) | Inappropriate study design |
| 6678 | Active mobility early after stroke (AMOBES), 1 year follow-up. A randomised controlled trial | Inappropriate population |
| 6679 | Effects of multidomain versus single-domain training on executive control and memory in older adults: study protocol for a randomized controlled trial | Inappropriate study design |
| 6680 | Comparison Between Multicomponent Exercise and Walking Interventions in Long-Term Nursing Homes: a Randomized Controlled Trial | Inappropriate outcome |
| 6681 | Reduction in body fat mass is independentlyassociatedwith improved executive function | Inappropriate study design |
| 6682 | Baseline aerobic fitness and cortical volume atrophy across 5 years in older adults from the general population | Inappropriate population |
| 6683 | Efficacy and safety of non-immersive virtual reality exercising in stroke rehabilitation (EVREST): a randomised, multicentre, single-blind, controlled trial | Inappropriate study design |
| 6684 | Efficacy of guided self-help behavioural activation and physical activity for depression: a randomized controlled trial | Inappropriate outcome |
| 6685 | Randomized controlled pilot study testing use of smartphone technology for obesity treatment | Inappropriate population |
| 6686 | Short-term Effects of High-Dose Caffeine on Cardiac Arrhythmias in Patients With Heart Failure: a Randomized Clinical Trial | Inappropriate population |
| 6687 | Effectiveness of energy conservation management on fatigue and participation in multiple sclerosis: a randomized controlled trial | Inappropriate study design |
| 6688 | Effects of a power strength training using elastic resistance exercises on the motor and non-motor symptoms in patients with Parkinson's disease H&Y 1-3: study protocol for a randomised controlled trial (PARK-BAND Study) | Inappropriate population |
| 6689 | Aortic stiffness, central pulse pressure and cognitive function following acute resistance exercise | Inappropriate study design |
| 6690 | Pulmonary Vascular Distensibility Predicts Pulmonary Hypertension Severity, Exercise Capacity, and Survival in Heart Failure | Inappropriate study design |
| 6691 | Effects of Exercise on Cognition: the Finnish Alzheimer Disease Exercise Trial: a Randomized, Controlled Trial | Inappropriate population |
| 6692 | Exercise alters response of reward anticipation in the ventral striatum of subjects with Parkinson's disease | Inappropriate outcome |
| 6693 | Combined Therapies–rTMS meets CBT. Novel approach for the treatment of Derealization/Depersonalization Syndrome | Inappropriate population |
| 6694 | Intensive Stuttering Group Therapy for School-age Children and the Effect of Emotional Reactivity on Therapy Outcomes | Inappropriate study design |
| 6695 | Physical ACtivity and Exercise Outcomes in Huntington Disease (PACE-HD): protocol for a 12-Month Trial Within Cohort Evaluation of a Physical Activity Intervention in People With Huntington Disease | Inappropriate population |
| 6696 | Robotic support for cognitive therapy of hand function | Inappropriate study design |
| 6697 | Normobaric hypoxic conditioning in men with metabolic syndrome | Inappropriate study design |
| 6698 | A pilot randomized controlled trial of D-cycloserine and distributed practice as adjuvants to constraint-induced movement therapy after stroke | Inappropriate study design |
| 6699 | Efficacy of intermittent aerobic exercise on sleep efficiency in patients with rheumatoid arthritis. A randomized controlled trial | Inappropriate study design |
| 6700 | Aerobic treadmill training plus physiotherapy improves walking speed and capacity in subacute, moderately affected patients after stroke | Inappropriate study design |

| | | |
|---|---|---|
| 6701 | Physical exercise training in older adults diagnosed with mild to moderate dementia | Inappropriate intervention |
| 6702 | Effect of Low-Intensity, Kinect™-Based Kaimai-Style Qigong Exercise in Older Adults With Type 2 Diabetes | Inappropriate study design |
| 6703 | Internet-delivered exposure-based cognitive behaviour therapy for adolescents with functional abdominal pain or functional dyspepsia: a feasibility study | Inappropriate outcome |
| 6704 | Can high-intensity interval/circuit training improve the cognitive function and quality of life of patients in recovery from substance use disorder? | Inappropriate study design |
| 6705 | Physical Fitness as Klotho Protein Stimulator | Inappropriate population |
| 6706 | Exercise training as S-Klotho protein stimulator in sedentary healthy adults: rationale, design, and methodology | Inappropriate outcome |
| 6707 | Exercise-induced changes in brain activity during memory encoding and retrieval after long-term bed rest | Inappropriate study design |
| 6708 | Immunological effects of behavioral activation with exercise in major depression: an exploratory randomized controlled trial | Inappropriate study design |
| 6709 | Is upper limb virtual reality training more intensive than conventional training for patients in the subacute phase after stroke? An analysis of treatment intensity and content | Inappropriate study design |
| 6710 | Healthy Lifestyles Program for Adults with an Acquired Brain Injury | Inappropriate study design |
| 6711 | Effects of exercise training on insulin-like growth factor-I expression in the skeletal muscle of non-cachectic patients with chronic heart failure | Inappropriate study design |
| 6712 | Physical activity in subacute stroke -phys-stroke | Inappropriate study design |
| 6713 | Feasibility and Outcomes of an Exercise Intervention for Chemotherapy-Induced Heart Failure | Inappropriate study design |
| 6714 | Post-exercise reduction in blood pressure in hypertensive subjects: effects of angiotensin converting enzyme inhibition | Inappropriate study design |
| 6715 | Clinical and cost effectiveness of computer treatment for aphasia post stroke (Big CACTUS): study protocol for a randomised controlled trial | Inappropriate study design |
| 6716 | Adherence to moderate-intensity exercise during breast cancer therapy | Inappropriate population |
| 6717 | Exercise Effects on Cognition in School-Aged Children | Inappropriate study design |
| 6718 | Effects of information processing load in abstinent and nonabstinent smokers' psychomotor task performance | Inappropriate study design |
| 6719 | Mechanisms for reducing low back pain: a mediation analysis of a multifaceted intervention in workers in elderly care | Inappropriate study design |
| 6720 | Rationale, design, and baseline data for Commit to Quit II: an evaluation of the efficacy of moderate-intensity physical activity as an aid to smoking cessation in women | Inappropriate study design |
| 6721 | The effect of a multimodal intervention on treatment-related symptoms in patients undergoing hematopoietic stem cell transplantation: a randomized controlled trial | Inappropriate study design |
| 6722 | A comparison of the effects of mirror therapy and phantom exercises on phantom limb pain | Inappropriate population |
| 6723 | Randomised controlled trial of an app-based intervention, Anchored, to support the mental health of Australians recently unemployed due to COVID-19 | Inappropriate population |
| 6724 | IMPACT OF A MOBILE HEALTH-INFUSED LATIN DANCE PROGRAM ON PHYSICAL ACTIVITY, FITNESS, AND EXECUTIVE FUNCTION IN OLDER LATINOS | Inappropriate outcome |
| 6725 | Effects of a supervised exercise program on cardiac structure during pregnancy. Randomized clinical trial | Inappropriate study design |
| 6726 | Aerobic exercise modulates transfer and brain signal complexity following cognitive training | Inappropriate study design |
| 6727 | Guided graded exercise self-help plus specialist medical care versus specialist medical care alone for chronic fatigue syndrome (GETSET): a pragmatic randomised controlled trial | Inappropriate population |
| 6728 | Individual differences in the neurobiology of fluid intelligence predict responsiveness to training: evidence from a comprehensive cognitive, mindfulness meditation, and aerobic exercise intervention | Inappropriate population |
| 6729 | A pilot study of respiratory muscle training to improve cough effectiveness and reduce the incidence of pneumonia in acute stroke: study protocol for a randomized controlled trial | Inappropriate study design |
| 6730 | Mode of physical activity and self-efficacy in older adults: a latent growth curve analysis | Inappropriate study design |
| 6731 | Perceived change among participants in an exercise program for older adults | Inappropriate study design |

| | | |
|---|---|---|
| 6732 | Influence of an exercise program on cardiac remodeling and functional capacity in patients with stroke (CRONuS trial): study protocol for a randomized controlled trial | Inappropriate population |
| 6733 | Yoga for Smoking Cessation Feasibility Study | Inappropriate study design |
| 6734 | Improving physical functional status in patients with fibromyalgia: a brief cognitive behavioral intervention | Inappropriate study design |
| 6735 | Hydration Does Not Change Postexercise Hypotension and Its Mechanisms | Inappropriate population |
| 6736 | Exercise fails to improve neurocognition in depressed middle-aged and older adults | Inappropriate population |
| 6737 | Effects of a home-based telephone-supported physical activity program on physical function among older adults with chronic low back pain | Inappropriate study design |
| 6738 | Depressive mood symptoms and fatigue after exercise withdrawal: the potential role of decreased fitness | Inappropriate study design |
| 6739 | The effect of transcranial direct stimulation(TDCS) and exercise of cognition and motor aspect in patients with chronic stroke | Inappropriate population |
| 6740 | Evidence of a shift in the excitability of the glutamatergic neurotransmission in the anterior cingulate during appraisal of emotional stimuli that is influenced by aerobic fitness | Inappropriate population |
| 6741 | Robot-assisted reaching exercise promotes arm movement recovery in chronic hemiparetic stroke: a randomized controlled pilot study | Inappropriate population |
| 6742 | A comparison of cycle exercise, functional training and Xbox Kinect® in cardiorespiratory fitness of elderly with Parkinson's disease: a randomized controlled trial | Inappropriate study design |
| 6743 | Cardiovascular effects of intensive lifestyle intervention in type 2 diabetes | Inappropriate study design |
| 6744 | A randomized controlled trial to examine combined multiple-modality and mind-motor exercise on cognitive functioning in community-dwelling older adults: a pilot study | Inappropriate study design |
| 6745 | Smart-sport and cerebral metabolism in old age: AN MRS/MRI study | Inappropriate study design |
| 6746 | Erratum: effect of aerobic exercise on cognition in younger adults: a randomized clinical trial (Neurology (2019) 92 9 (e905-e916)) | Inappropriate population |
| 6747 | Exercise on prescription for women aged 40-74 recruited through primary care: two year randomised controlled trial | Inappropriate study design |
| 6748 | Randomised controlled trial examining the effect of an outpatient exercise training programme on haemodynamics and cardiac MR parameters of right ventricular function in patients with pulmonary arterial hypertension: the ExPAH study protocol | Inappropriate study design |
| 6749 | Exercise training increases physical fitness for children with cerebral palsy | Inappropriate population |
| 6750 | Tendinopathy Education on the Achilles | Inappropriate study design |
| 6751 | Likelihood of myocardial infarction during stroke rehabilitation preceded by cardiovascular screening and an exercise tolerance test: the Locomotor Experience Applied Post-Stroke (LEAPS) trial | Inappropriate study design |
| 6752 | study for healthy ageing | Inappropriate study design |
| 6753 | An Intervention to Increase Physical Activity Among African American Women | Inappropriate study design |
| 6754 | Pyridostigmine, but not ivabradine, improves autonomic function after dynamic exercise in patients with heart failure | Inappropriate study design |
| 6755 | Aerobic exercise or stretching as add-on to inpatient treatment of depression: similar antidepressant effects on depressive symptoms and larger effects on working memory for aerobic exercise alone | Inappropriate population |
| 6756 | Pilot study of Lokomat versus manual-assisted treadmill training for locomotor recovery post-stroke | Inappropriate population |
| 6757 | Protocol for Fit Bodies, Fine Minds: a randomized controlled trial on the affect of exercise and cognitive training on cognitive functioning in older adults | Inappropriate outcome |
| 6758 | Robotic resistance/assistance training improves locomotor function in individuals poststroke: a randomized controlled study | Inappropriate study design |
| 6759 | Beta-blockers withdrawal in patients with heart failure with preserved ejection fraction and chronotropic incompetence: effect on functional capacity rationale and study design of a prospective, randomized, controlled trial (The Preserve-HR trial) | Inappropriate study design |
| 6760 | The psychophysiological effects of Tai-chi and exercise in residential schizophrenic patients: a 3-arm randomized controlled trial | Inappropriate population |
| 6761 | Differential exercise effects on quality of life and health-related quality of life in older adults: a randomized controlled trial | Inappropriate population |

| | | |
|---|---|---|
| 6762 | Effect of memory training allied to physical exercise in adult and elderly health parameters: association with genetic variables | Inappropriate study design |
| 6763 | Left ventricular mechanical limitations to stroke volume in healthy humans during incremental exercise | Inappropriate study design |
| 6764 | A randomized controlled trial assessing the efficacy of an upper limb self-rehabilitation programme among chronic Beninese stroke patients | Inappropriate study design |
| 6765 | Direct and long term influence of cardiovascular training on cognition in subacute stroke patients | Inappropriate study design |
| 6766 | Neuroplasticity-based cognitive training in early psychosis | Inappropriate study design |
| 6767 | Persistent improvements in arm use in stroke survivors with plegic hands following expanded ci therapy | Inappropriate study design |
| 6768 | Effect of a Long-Term Intensive Lifestyle Intervention on Cognitive Function: action for Health in Diabetes Study | Inappropriate outcome |
| 6769 | Determinants of Dropout and Nonadherence in a Dementia Prevention Randomized Controlled Trial: the Prevention of Dementia by Intensive Vascular Care Trial | Inappropriate study design |
| 6770 | Comparison of Two Music Training Approaches on Music and Speech Perception in Cochlear Implant Users | Inappropriate population |
| 6771 | Functional recovery and rehabilitation of postural impairment and gait ataxia in patients with acute cerebellar stroke | Inappropriate population |
| 6772 | Using a novel lever-drive wheelchair to increase arm movement practice early after stroke: preliminary results of a randomized controlled trial | Inappropriate study design |
| 6773 | The impact of three distinct exercise types on fatigue, anxiety, and depression in parkinson's disease | Inappropriate study design |
| 6774 | Aerobic exercise decreases cortisol awakening reaction in depressed inpatients | Inappropriate population |
| 6775 | Effects of Electromechanical Gait Trainer in Patients With Multiple Sclerosis | Inappropriate study design |
| 6776 | Effectiveness of Virtual Reality Exercises in STroke Rehabilitation (EVREST): rationale, design, and protocol of a pilot randomized clinical trial assessing the Wii gaming system | Inappropriate population |
| 6777 | Physical Fitness and Brain - Interventional Study | Inappropriate population |
| 6778 | Resistance training improves hyperglycemia and dyslipidemia, highly prevalent among nonelderly, nondiabetic, chronically disabled stroke patients | Inappropriate study design |
| 6779 | The effect of physical fitness (VO2) on white matter hyperintensity volume and cognition in older adults from the generation100 study | Inappropriate population |
| 6780 | Effect of aerobic exercise and relaxation training on fatigue and physical performance of cancer patients after surgery. A randomised controlled trial | Inappropriate study design |
| 6781 | Reaching Training Based on Robotic Hybrid Assistance for Stroke Patients ( RETRAINER-S2 ) | Inappropriate study design |
| 6782 | Effects of Rice Wine Lees on Cognitive Function in Community-Dwelling Physically Active Older Adults: a Pilot Randomized Controlled Trial | Inappropriate study design |
| 6783 | A 12-week physical and cognitive exercise program can improve cognitive function and neural efficiency in community-dwelling older adults: a randomized controlled trial | Inappropriate population |
| 6784 | Pilates vs. muscular training in older women. Effects in functional factors and the cognitive interaction: a randomized controlled trial | Inappropriate population |
| 6785 | Effects of short-term dietary nitrate supplementation on blood pressure, O2 uptake kinetics, and muscle and cognitive function in older adults | Inappropriate study design |
| 6786 | Treating depressed Australian workers using an app-based intervention: a randomised controlled trial (Anchored) | Inappropriate population |
| 6787 | Preventing loss of independence through exercise in persons with dementia in the VA (plie-va) | Inappropriate population |
| 6788 | Functional training for elderly with Alzheimer's disease | Inappropriate study design |
| 6789 | Cognitive Remediation Therapy in Anorexia Nervosa | Inappropriate study design |
| 6790 | Effects of short-term and prolonged supplementation of beetroot juice on cardiovascular and cognitive function and mood of healthy older compared to younger adults | Inappropriate population |
| 6791 | Does squatting need attention?-A dual-task study on cognitive resources in resistance exercise | Inappropriate study design |
| 6792 | Stroke volume during submaximal exercise in endurance-trained normotensive subjects and in untrained hypertensive subjects with beta blockade (propranolol and pindolol) | Inappropriate study design |
| 6793 | Should physical activity programs be tailored when older adults have compromised function? | Inappropriate population |

| | | |
|---|---|---|
| 6794 | Examining the Therapeutic Effects of Bacopa monnieri Supplementation and Cognitive Training on Brain Health, Cognition and Everyday Function in Healthy Older Adults | Inappropriate study design |
| 6795 | Moms in motion: a group-mediated cognitive-behavioral physical activity intervention | Inappropriate study design |
| 6796 | The association between comorbidity and physical activity levels in people with osteoarthritis: secondary analysis from two randomised controlled trials | Inappropriate study design |
| 6797 | Feasibility and Acceptability of a Web-based Multidomain Intervention to Maintain Cognitive Function in Older Adults | Inappropriate study design |
| 6798 | The Effect of an Expiratory Resistance Mask with Dead Space on Sleep, Acute Mountain Sickness, Cognition, and Ventilatory Acclimatization in Normobaric Hypoxia | Inappropriate outcome |
| 6799 | Adherence to multidomain interventions for the prevention of Alzheimer's disease: data from the MAPT and finger trials | Inappropriate population |
| 6800 | The effect of Cognitive behaviour therapy on adult mental health during the COVID-19 pandemic | Inappropriate population |
| 6801 | Optimization of Pulmonary Rehabilitation Programmes: the OPTION Study | Inappropriate study design |
| 6802 | Reciprocal effects of treatment-induced increases in exercise and improved eating, and their psychosocial correlates, in obese adults seeking weight loss: a field-based trial | Inappropriate population |
| 6803 | Translating the lifestyle interventions and independence for elders clinical trial to mobility-limited older adults in the community: the engage pilot study | Inappropriate population |
| 6804 | MBSR vs aerobic exercise in social anxiety: fMRI of emotion regulation of negative self-beliefs | Inappropriate study design |
| 6805 | The Management of Diabetes in Everyday Life Program | Inappropriate study design |
| 6806 | Arterial stiffness is associated with lower cerebral blood perfusion in older adults | Inappropriate population |
| 6807 | Feasibility of music-assisted treadmill training in parkinson's disease patients with and without deep brain stimulation: insights from an ongoing pilot randomized controlled trial | Inappropriate study design |
| 6808 | Qualitative assessment of verbal and non-verbal psychosocial interventions for people with severe mental illness | Inappropriate intervention |
| 6809 | Physical fitness training in Subacute Stroke (PHYS-STROKE) - study protocol for a randomised controlled trial | Inappropriate study design |
| 6810 | Exercise effects on night-to-night fluctuations in self-rated sleep among older adults with sleep complaints | Inappropriate study design |
| 6811 | A pilot RCT on the efficacy of home-based exercise to improve cognitive functioning in grade II and III glioma patients | Inappropriate population |
| 6812 | The effect of probiotics and resistance exercise on cognitive function, physical performance, and intestinal flora in the elderly: a randomized, double-blind, placebo-controlled trial | Inappropriate study design |
| 6813 | What Is the Best Treatment before Bariatric Surgery? Exercise, Exercise and Group Therapy, or Conventional Waiting: a Randomized Controlled Trial | Inappropriate population |
| 6814 | Physical activity and cognition in young-onset Parkinson's disease | Inappropriate intervention |
| 6815 | Effect of finger tracking combined with electrical stimulation on brain reorganization and hand function in subjects with stroke | Inappropriate study design |
| 6816 | Effects of intensity of arm training on hemiplegic upper extremity motor recovery in stroke patients: a randomized controlled trial | Inappropriate study design |
| 6817 | Evaluation of cognitive behaviour combined with biofeedback therapy based on the fusion of normed space and banach space on the emotional state of patients with coronary artery bypass grafting | Inappropriate outcome |
| 6818 | Sildenafil in severe pulmonary hypertension associated with chronic obstructive pulmonary disease: a randomized controlled multicenter clinical trial | Inappropriate study design |
| 6819 | High intensity intermittent training improves cardiac function and reduces liver fat in adults with Type 2 diabetes: an MRI/S study | Inappropriate study design |
| 6820 | Effects of exercise programs on falls and mobility in frail and pre-frail older adults: a multicenter randomized controlled trial | Inappropriate study design |
| 6821 | Home-based exercise improves functional performance and quality of life in women with diastolic heart failure | Inappropriate study design |
| 6822 | Multiple Joint, Structural Barbell Resistance Exercise Improves Information Processing Speed | Inappropriate study design |
| 6823 | Exercise as an Intervention to Reduce Study-Related Fatigue among University Students: a Two-Arm Parallel Randomized Controlled Trial | Inappropriate population |
| 6824 | Carbohydrate feedings during team sport exercise preserve physical and CNS function | Inappropriate population |

| | | |
|---|---|---|
| 6825 | Beer or Ethanol Effects on the Response to High Intensity Interval Training: a Controlled Study in Healthy Individuals | Inappropriate population |
| 6826 | Home-based, square-stepping exercise program among older adults with multiple sclerosis: results of a feasibility randomized controlled study | Inappropriate outcome |
| 6827 | Process evaluation of a culturally-tailored physical activity intervention in African-American mother-daughter dyads | Inappropriate study design |
| 6828 | The effect of a multifaceted efficacy intervention on exercise behavior in relatives of colon cancer patients | Inappropriate study design |
| 6829 | Effects of exercise training on cognitive functioning among depressed older men and women | Inappropriate study design |
| 6830 | A preliminary study on the performance of one-leg versus two-leg symptom-limited exercise test in individuals with subacute stroke | Inappropriate study design |
| 6831 | A comparative evaluation of cognitive-behavioral therapy (CBT) versus exercise therapy (ET) for the treatment of body image disturbance. Preliminary findings | Inappropriate study design |
| 6832 | Exercise Induced Improvement of the Venous Reserve Capacity in Formerly Pre-eclamptic Women | Inappropriate study design |
| 6833 | Abstract #106: aerobic Exercise and repetitive Transcranial Magnetic Stimulation in Parkinson's disease: exploring Potential Plasticity Biomarkers | Inappropriate population |
| 6834 | Controlled Frequency Breathing Reduces Inspiratory Muscle Fatigue | Inappropriate population |
| 6835 | Effects of aerobic exercise versus stress management treatment in fibromyalgia. A 4.5 year prospective study | Inappropriate population |
| 6836 | Creatine supplementation in fibromyalgia: a randomized, double-blind, placebo-controlled trial | Inappropriate population |
| 6837 | Effects of Six-Week Ginkgo biloba Supplementation on Aerobic Performance, Blood Pro/Antioxidant Balance, and Serum Brain-Derived Neurotrophic Factor in Physically Active Men | Inappropriate study design |
| 6838 | Influence of dietary nitrate supplementation on physiological and cognitive responses to incremental cycle exercise | Inappropriate study design |
| 6839 | Effect of mindfulness yoga on psychological distress in people with Parkinson's disease (PD): a randomized controlled trial | Inappropriate study design |
| 6840 | Effect of perceived choice of music on exercise intrinsic motivation | Inappropriate study design |
| 6841 | Shear mediated dilation of the internal carotid artery during exercise and hypercapnia in humans | Inappropriate study design |
| 6842 | Improved frontoparietal white matter integrity in overweight children is associated with attendance at an after-school exercise program | Inappropriate study design |
| 6843 | Diet, physical exercise and cognitive behavioral training as a combined workplace based intervention to reduce body weight and increase physical capacity in health care workers - a randomized controlled trial | Inappropriate study design |
| 6844 | Effects of lifestyle interventions on quality of life in breast cancer patients | Inappropriate study design |
| 6845 | Short-term effects of aerobic exercise on functional capacity, fatigue, and quality of life in patients with post-polio syndrome | Inappropriate outcome |
| 6846 | BOUNCE: a community-based mother-daughter healthy lifestyle intervention for low-income Latino families | Inappropriate population |
| 6847 | Novel Tools for the Delivery and Assessment of Exercise Programs Adapted to Individuals With Parkinson's Disease | Inappropriate study design |
| 6848 | Think Dental, Be Active!: a randomised control trial using psycho-education, physical activity and oral health interventions in older adults aged >50 years old, residing in Royal Freemason Benevolent Institute residential aged care facilities | Inappropriate study design |
| 6849 | Effect of a dual-task net-step exercise on cognitive and gait function in older adults | Inappropriate study design |
| 6850 | A technology-assisted, high intensive, task-oriented exercise program to improve arm and hand functions in stroke patients: study protocol for the techito feasibility and pilot trial | Inappropriate population |
| 6851 | 'PDSAFE'-A multi-dimensional fall rehabilitation intervention for people with Parkinson's with specialist physiotherapy training: qualitative exploration of physio's experiences | Inappropriate study design |
| 6852 | Blind randomized controlled study of the efficacy of cognitive training in Parkinson's disease | Inappropriate study design |
| 6853 | Brain reorganization after bilateral arm training and distributed constraint-induced therapy in stroke patients: a preliminary functional magnetic resonance imaging study | Inappropriate intervention |
| 6854 | Effects of exercise on brain activity during walking in older adults: a randomized controlled trial | Inappropriate study design |

| | | |
|---|---|---|
| 6855 | Increasing self-directed training in neurorehabilitation patients through competition | Inappropriate study design |
| 6856 | Contribution of the shaping and restraint components of Constraint-Induced Movement therapy to Treatment Outcome | Inappropriate study design |
| 6857 | Building resilience to body image triggers using brief cognitive training on a mobile application: a randomized controlled trial | Inappropriate population |
| 6858 | The β-Adrenergic Agonist Albuterol Improves Pulmonary Vascular Reserve in Heart Failure With Preserved Ejection Fraction | Inappropriate population |
| 6859 | The effects of antiepileptic drugs on cognition in normal volunteers | Inappropriate study design |
| 6860 | Effects of Virtual Reality Simulation on Worker Emergency Evacuation of Neonates | Inappropriate population |
| 6861 | Leisure sports and postural control: can a black belt protect your balance from aging? | Inappropriate study design |
| 6862 | The role of attributions of responsibility in the facilitation of unusual experiential states during alpha training: an analysis of the biofeedback placebo effect | Inappropriate population |
| 6863 | Implementing behaviour change theory and techniques to increase physical activity and prevent functional decline among adults aged 61–70: the PreventIT project | Inappropriate study design |
| 6864 | Heart rate acceleration with GLP-1 receptor agonists in type 2 diabetes patients: an acute and 12-week randomised, double-blind, placebo-controlled trial | Inappropriate study design |
| 6865 | Comparison of the effects of modified constraint-induced movement therapy and intensive conventional therapy with a botulinum-a toxin injection on upper limb motor function recovery in patients with stroke | Inappropriate study design |
| 6866 | RECRUITMENT STRATEGIES IN A MULTICENTER RCT FOR DEMENTIA PREVENTION - RRAD STUDY | Inappropriate population |
| 6867 | Effects of Early Bedside Cycle Exercise on Intracranial Pressure and Systemic Hemodynamics in Critically Ill Patients in a Neurointensive Care Unit | Inappropriate population |
| 6868 | Effects of lifestyle modification on metabolic parameters and carotid intima-media thickness in patients with type 2 diabetes mellitus | Inappropriate population |
| 6869 | The DIET study: long-term outcomes of a cognitive-behavioral weight-control intervention in independent-living elders. Dietary Intervention: evaluation of Technology | Inappropriate population |
| 6870 | Mind racing: the influence of exercise on long-term memory consolidation | Inappropriate study design |
| 6871 | Disease management of the metabolic syndrome in a community: study design and process analysis on baseline data | Inappropriate study design |
| 6872 | Mental techniques during manual stretching in spasticity--a pilot randomized controlled trial | Inappropriate study design |
| 6873 | Randomised Controlled Trial of Combining Cognitive Behaviour Therapy with either Exercise or Yoga for Adults with Posttraumatic Stress Disorder | Inappropriate study design |
| 6874 | Psychological modeling and adaptations in cognitive representations with increased resistance during motor skill acquisition | Inappropriate outcome |
| 6875 | The Dehydroepiandrosterone And WellNess (DAWN) study: research design and methods | Inappropriate study design |
| 6876 | Exercising before a nap benefits memory better than napping or exercising alone | Inappropriate intervention |
| 6877 | A new trial design for evaluating exercise outcomes in Huntington's disease | Inappropriate study design |
| 6878 | Carbohydrate ingestion improves performance of a new reliable test of soccer performance | Inappropriate population |
| 6879 | Functional electrical stimulation therapy for severe hemiplegia: randomized control trial revisited | Inappropriate population |
| 6880 | Effects of rTMS in rehabilitation after stroke | Inappropriate population |
| 6881 | Change of quality of life due to exercise training in knee osteoarthritis: sF-36 and Womac | Inappropriate study design |
| 6882 | Loss of effectiveness of dihydralazine in the long-term treatment of chronic heart failure | Inappropriate study design |
| 6883 | Cardiovascular and metabolic responses to electrical stimulation- induced leg exercise in spinal cord injury | Inappropriate outcome |
| 6884 | Hormonal responses to physical and cognitive stress in a school setting | Inappropriate study design |
| 6885 | Diet and exercise interventions improve health markers in individuals with an increased risk of disease | Inappropriate population |
| 6886 | Affect and exercise: positive affective expectations can increase post-exercise mood and exercise intentions | Inappropriate study design |
| 6887 | Prosocial Behavior and Exercise Among Older Adults | Inappropriate study design |
| 6888 | Step Monitoring to Improve ARTERial Health | Inappropriate study design |

| | | |
|---|---|---|
| 6889 | Overview of the Activity Counseling Trial (ACT) intervention for promoting physical activity in primary health care settings | Inappropriate study design |
| 6890 | The effect of low intensity exercise intervention with and without capsaicin analogues on physical activity, fitness, body composition and psychological factors in community dwelling older adults | Inappropriate study design |
| 6891 | Behavioral choice treatment promotes continuing weight loss: preliminary results of a cognitive-behavioral decision-based treatment for obesity | Inappropriate study design |
| 6892 | Does progressive resistance and balance exercise reduce falls in residential aged care? Randomized controlled trial protocol for the SUNBEAM program | Inappropriate population |
| 6893 | The effects of long- and short-term interdisciplinary treatment approaches in women with fibromyalgia: a randomized controlled trial | Inappropriate study design |
| 6894 | Health Benefits of a 6-month Brisk Walking Program in Sedentary Postmenopausal Women | Inappropriate population |
| 6895 | A randomized controlled study of brief family-based intervention in obsessive compulsive disorder | Inappropriate study design |
| 6896 | Reduction of Daily Sitting Time in Patients With Rheumatoid Arthritis | Inappropriate study design |
| 6897 | Robotic Pedaling Therapy for Targeted Neural Plasticity | Inappropriate study design |
| 6898 | Sustainability of health and lifestyle improvements following a non-dieting randomised trial in overweight women | Inappropriate study design |
| 6899 | Effects of cognitive behavioral therapy on sleep disturbances and quality of life among adults with type 2 diabetes mellitus: a randomized controlled trial | Inappropriate study design |
| 6900 | Enhancing cognitive functioning in the elderly: multicomponent vs resistance training | Inappropriate study design |
| 6901 | Combined acebutolol/nifedipine therapy in patients with chronic coronary artery disease: additional improvement of ischemia-induced left ventricular dysfunction | Inappropriate study design |
| 6902 | Phosphatidylserine and caffeine attenuate postexercise mood disturbance and perception of fatigue in humans | Inappropriate population |
| 6903 | Hemodialysis-based Interventions to Preserve Cognitive Function | Inappropriate study design |
| 6904 | Vitamin D and Resistance Exercise in Elderly (NUTRIAGINGVitD) | Inappropriate population |
| 6905 | Dietary Protein and Resistance Exercise in Elderly | Inappropriate study design |
| 6906 | Perioperative Cognitive Protection-Cognitive Exercise and Cognitive Reserve (The Neurobics Trial): a Single-blind Randomized Trial | Inappropriate population |
| 6907 | Carotid chemoreceptor control of muscle sympathetic nerve activity in hypobaric hypoxia | Inappropriate study design |
| 6908 | Does respiratory muscle training improve cough flow in acute stroke? Pilot randomized controlled trial | Inappropriate study design |
| 6909 | Supplemental therapeutic balance exercise with a robot in acute acquired brain injury rehabilitation a randomized, blinded study | Inappropriate study design |
| 6910 | Acute and chronic effects of molsidomine on gas exchange and hemodynamics in patients with primary vascular pulmonary hypertension | Inappropriate population |
| 6911 | ENAbLE: secondary prevention of stroke. A physical activity and diet trial protocol | Inappropriate population |
| 6912 | Impact of Exercise and Affirmations (IntenSati) on Addiction-related Cognitive and Psychosocial Deficits | Inappropriate study design |
| 6913 | Synchronized-dance on mood disorders, quality of life and functional independent performance in ambulatory stroke survivors: a clinical control study | Inappropriate population |
| 6914 | Effects of Resistance Exercise with Instability on Cognitive Function (REI Study): a Proof-Of-Concept Randomized Controlled Trial in Older Adults with Cognitive Complaints | Inappropriate study design |
| 6915 | Effects of acute ingestion of a pre-workout dietary supplement with and without p-synephrine on resting energy expenditure, cognitive function and exercise performance | Inappropriate intervention |
| 6916 | Effects of ingesting a pre-workout dietary supplement with and without synephrine for 8 weeks on training adaptations in resistance-trained males | Inappropriate study design |
| 6917 | Effect of combined physical training on cognitive function in people with epilepsy: results from a randomized controlled trial | Inappropriate population |
| 6918 | Effects of sleep restriction and exercise deprivation on somatic symptoms and mood in healthy adults | Inappropriate population |
| 6919 | Effects of an individual 12-week community-located “start-to-run” program on physical capacity, walking, fatigue, cognitive function, brain volumes, and structures in persons with multiple sclerosis | Inappropriate study design |

| | | |
|---|---|---|
| 6920 | Prevalent digoxin use and subsequent risk of death or hospitalization in ambulatory heart failure patients with a reduced ejection fraction-Findings from the Heart Failure: a Controlled Trial Investigating Outcomes of Exercise Training (HF-ACTION) randomized controlled trial | Inappropriate population |
| 6921 | Hypoxic exercise training improves cardiac/muscular hemodynamics and is associated with modulated circulating progenitor cells in sedentary men | Inappropriate study design |
| 6922 | A Study to Evaluate the Efficacy and Safety of BRL 49653C in Non-insulin Dependent Diabetes | Inappropriate study design |
| 6923 | The effect of self-care interventions on sleep quality in post coronary artery bypass graft surgery patients | Inappropriate study design |
| 6924 | Home-based physical activity intervention for colorectal cancer survivors | Inappropriate study design |
| 6925 | A randomized, controlled clinical trial of standard, group and brief cognitive-behavioral therapy for panic disorder with agoraphobia: a two-year follow-up | Inappropriate study design |
| 6926 | Rehabilitation program with or without a whey protein-based nutritional support enriched with essential aminoacids and vitamin D in patients with Parkinson's disease or parkinsonism: study design of a randomized, controlled trial | Inappropriate population |
| 6927 | Rehabilitation program with or without a muscle-targeted nutritional support in patients with Parkinson's disease or Parkinsonism: study design of a randomized, controlled trial | Inappropriate intervention |
| 6928 | Influences of nonselective, β1-selective and vasodilatory β1-selective β-blockers on arterial pulse wave velocity in normotensive subjects | Inappropriate study design |
| 6929 | Single Exercise Session or Meal vs Control in SCI: case Series | Inappropriate study design |
| 6930 | Specialized Addiction Treatment Versus Treatment as Usual for Young Patients With Cannabis Abuse and Psychosis | Inappropriate study design |
| 6931 | Kinematic and clinical analyses of upper-extremity movements after constraint-induced movement therapy in patients with stroke: a randomized controlled trial | Inappropriate study design |
| 6932 | Transcranial direct current stimulation for children with perinatal stroke and hemiparesis | Inappropriate study design |
| 6933 | Protein-enriched, milk-based supplement to counteract sarcopenia in acutely ill geriatric patients offered resistance exercise training during and after hospitalisation: study protocol for a randomised, double-blind, multicentre trial | Inappropriate population |
| 6934 | Divergence of subjective and performance-based cognitive gains following cognitive training in schizophrenia | Inappropriate study design |
| 6935 | Expectations, perceptions, and physiotherapy predict prolonged sick leave in subacute low back pain | Inappropriate outcome |
| 6936 | Randomized, multicenter, comparative study of NEURO versus CIMT in poststroke patients with upper limb hemiparesis: the NEURO-VERIFY Study | Inappropriate study design |
| 6937 | Habitual physical activity levels predict treatment outcomes in depressed adults: a prospective cohort study | Inappropriate population |
| 6938 | Psychomotor Performance Decrements following a Successful Physical Activity Intervention for Insomnia | Inappropriate study design |
| 6939 | Effect of carbohydrate intake during matches on the cognitive function of professional female football players | Inappropriate study design |
| 6940 | Reducing the fear of falling among community-dwelling elderly adults through cognitive-behavioural strategies and intense Tai Chi exercise: a randomized controlled trial | Inappropriate population |
| 6941 | Impact of a Multidomain Intensive Lifestyle Intervention on Complaints About Memory, Problem-Solving, and Decision-Making Abilities: the Action for Health in Diabetes Randomized Controlled Clinical Trial | Inappropriate study design |
| 6942 | A brief measure of perceived understanding of informed consent in a clinical trial was validated | Inappropriate study design |
| 6943 | Effects of memory training on cortical thickness in the elderly | Inappropriate population |
| 6944 | Progressive resistance training improves muscle strength of paretic lower limb in chronic stroke patients: a systematic review | Inappropriate study design |
| 6945 | Efficacy of a multi-component exercise programme and nutritional supplementation on musculoskeletal health in men treated with androgen deprivation therapy for prostate cancer (IMPACT): study protocol of a randomised controlled trial | Inappropriate population |
| 6946 | Changes in body composition, cardiovascular disease risk factors, and eating behavior after an intensive lifestyle intervention with high volume of physical activity in severely obese subjects: a prospective clinical controlled trial | Inappropriate study design |
| 6947 | Fostering adaptive stress management via technology-mediated mindfulness practice: self-report and psychophysiological evidence | Inappropriate study design |
| 6948 | Effective Weight Loss and Maintenance by Intensive Start with Diet and Exercise | Inappropriate population |

| | | |
|---|---|---|
| 6949 | Living Well with Diabetes: a randomized controlled trial of a telephone-delivered intervention for maintenance of weight loss, physical activity and glycaemic control in adults with type 2 diabetes | Inappropriate study design |
| 6950 | Tongue pressure training protocol for swallowing impairment post-stroke | Inappropriate study design |
| 6951 | Training and de-training effects: one year follow-up of a 3-month resistance exercise program in the pre-frail elderly | Inappropriate population |
| 6952 | BDNF Val66Met polymorphism as a moderator of exercise enhancement of smoking cessation treatment in anxiety vulnerable adults | Inappropriate population |
| 6953 | The Novel Single-Stroke Kayak Test: can It Discriminate Between 200-m and Longer-Distance (500- and 1000-m) Specialists in Canoe Sprint? | Inappropriate outcome |
| 6954 | Inspiratory muscular training in chronic stroke survivors: a randomized controlled trial | Inappropriate study design |
| 6955 | Telephone-administered cognitive behavioral therapy for depression in Parkinson's disease: a randomized controlled trial | Inappropriate study design |
| 6956 | Clinically significant improved fitness and weight loss among overweight persons with serious mental illness | Inappropriate study design |
| 6957 | The rehabilitation effect of gating-pad and healthy exercise on the balance and motor function of Parkinson's Disease | Inappropriate intervention |
| 6958 | Balance and gait training with augmented feedback improves balance confidence in people with parkinson's disease: a randomized controlled trial | Inappropriate study design |
| 6959 | Improving function after hospitalization in older adults with co-morbid conditions | Inappropriate study design |
| 6960 | Experiences of augmented arm rehabilitation including supported self-management after stroke: a qualitative investigation | Inappropriate study design |
| 6961 | Genome-wide sperm DNA methylation changes after 3 months of exercise training in humans | Inappropriate study design |
| 6962 | Strategies to increase amount and intensity of repetitive sit-to-stand training after stroke: the REPS trial | Inappropriate study design |
| 6963 | Leisure-time physical activity levels and risk of stroke. Contribution of the cohort Hermex | Inappropriate outcome |
| 6964 | Physical activity and health outcomes three months after completing a physical activity behavior change intervention: persistent and delayed effects | Inappropriate study design |
| 6965 | Can interactive, motion-capture-based rehabilitation in an inpatient stroke population increase physical activity levels for people undergoing rehabilitation for stroke? | Inappropriate study design |
| 6966 | Effects of amplitudes of whole-body vibration training on left ventricular stroke volume and ejection fraction in healthy young men | Inappropriate population |
| 6967 | Robot-assisted practice of gait and stair climbing in nonambulatory stroke patients | Inappropriate study design |
| 6968 | Caffeine improves exercise performance in a low carbohydrate, fasted state: a randomised, double-blind, placebo controlled trial | Inappropriate population |
| 6969 | How well do stroke survivors adhere to an 18-month physical activity and exercise programme? secondary results from a randomised controlled trial | Inappropriate population |
| 6970 | Flavonoid-Rich Mixed Berries Maintain and Improve Cognitive Function Over a 6 h Period in Young Healthy Adults | Inappropriate study design |
| 6971 | Insulin resistance correlations with executive functioning in sedentary older adults: the infinite study | Inappropriate study design |
| 6972 | Hemiparetic shoulder pain syndrome treated with deep dry needling during early rehabilitation: a prospective, open-label, randomized investigation | Inappropriate study design |
| 6973 | High prevalence of hypohydration in occupations with heat stress - Perspectives for performance in combined cognitive and motor tasks | Inappropriate outcome |
| 6974 | The effect of exercise on cognitive performance in soccer-specific tests | Inappropriate study design |
| 6975 | Effect of health education program on promoting physical activity among diabetic women in Mashhad, Iran: applying social cognitive theory | Inappropriate study design |
| 6976 | Physical Exercise and Health of Institutionalized Older Adults | Inappropriate study design |
| 6977 | Cognitive functional approach to manage low back pain in male adolescent rowers: a randomised controlled trial | Inappropriate study design |
| 6978 | Caffeine improves physical and cognitive performance during exhaustive exercise | Inappropriate study design |
| 6979 | Evaluating the Effectiveness of a Stepped-Care Approach to Long-Term Weight Loss (The Step-Up Study) | Inappropriate study design |
| 6980 | Short-term training effects on left ventricular diastolic function and oxygen uptake in older and younger men | Inappropriate study design |
| 6981 | Physical training as core component of multimodal treatment of older frail people-study protocol of a randomized controlled pilot study | Inappropriate study design |

| | | |
|---|---|---|
| 6982 | Physician-based activity counseling: intervention effects on mediators of motivational readiness for physical activity | Inappropriate study design |
| 6983 | Getting Physical on Cigarettes - Smoking Cessation & Relapse Prevention | Inappropriate study design |
| 6984 | Effects of a CHANGE intervention to increase exercise maintenance following cardiac events | Inappropriate study design |
| 6985 | The role of chair exercises for older adults following hip fracture | Inappropriate population |
| 6986 | Get in motion: an evaluation of the reach and effectiveness of a physical activity telephone counseling service for canadians living with spinalcord injury | Inappropriate population |
| 6987 | Pasos Hacia La Salud: a randomized controlled trial of an internet-delivered physical activity intervention for Latinas | Inappropriate population |
| 6988 | Carbohydrate ingestion before and during soccer match play and blood glucose and lactate concentrations | Inappropriate study design |
| 6989 | Effects of high-dose large neutral amino acid supplementation on exercise, motor skill, and mental performance in Australian Rules Football players | Inappropriate study design |
| 6990 | Exploring mediators of physical activity in young adult cancer survivors: evidence from a randomized trial of a Facebook-based physical activity intervention | Inappropriate outcome |
| 6991 | A randomized controlled trial of combined exercise and psycho-education for low-SES women: short- and long-term outcomes in the reduction of stress and depressive symptoms | Inappropriate study design |
| 6992 | Learning and developing individual exercise skills (L.A.D.I.E.S.) for a better life: a church-based physical activity intervention - Baseline participant characteristics | Inappropriate study design |
| 6993 | Effect of melatonin ingestion on physical performance, metabolic responses, and recovery after an intermittent training session | Inappropriate population |
| 6994 | Use of Wearable Technology and Social Media to Improve Physical Activity and Dietary Behaviors among College Students: a 12-Week Randomized Pilot Study | Inappropriate population |
| 6995 | High-volume exercise program in obese bariatric surgery patients: a randomized, controlled trial | Inappropriate study design |
| 6996 | Pragmatic exercise intervention in people with mild to moderate multiple sclerosis: a randomised controlled feasibility study | Inappropriate population |
| 6997 | A randomized prospective trial of a worksite intervention program to increase physical activity | Inappropriate study design |
| 6998 | Maintaining the outcomes of a successful weight gain prevention intervention in mid-age women: two year results from the 40-something randomized control trial | Inappropriate study design |
| 6999 | Multicomponent interdisciplinary group intervention for self-management of fibromyalgia: a mixed-methods randomized controlled trial | Inappropriate population |
| 7000 | Diabetes text study | Inappropriate study design |
| 7001 | Effects of a clinician referral and exercise program for men who have completed active treatment for prostate cancer: a multicenter cluster randomized controlled trial (ENGAGE) | Inappropriate study design |
| 7002 | Bronchial obstruction and exhaled nitric oxide response during exercise in cold air | Inappropriate study design |
| 7003 | Increasing physical activity in office workers--the Inphact Treadmill study; a study protocol for a 13-month randomized controlled trial of treadmill workstations | Inappropriate population |
| 7004 | Rationale, design, and baseline findings from HIPP: a randomized controlled trial testing a home-based, individually-tailored physical activity print intervention for African American women in the Deep South | Inappropriate study design |
| 7005 | Implementation-effectiveness trial of an ecological intervention for physical activity in ethnically diverse low income senior centers | Inappropriate study design |
| 7006 | Effect of intradialytic exercise on echocardiographic findings in hemodialysis patients | Inappropriate study design |
| 7007 | Home exercise in rheumatoid arthritis functional class II: goal setting versus pain attention | Inappropriate outcome |
| 7008 | Effects of music-based multitask exercise (Jaques-Dalcroze Eurhythmics) versus multicomponent exercise on physical function, falls and brain plasticity in older adults: a randomized controlled trial | Inappropriate study design |
| 7009 | Exercise in warm water decreases pain and improves cognitive function in middle-aged women with fibromyalgia | Inappropriate study design |
| 7010 | The family health project: cardiovascular risk reduction education for children and parents | Inappropriate study design |
| 7011 | Effects of a Kinect-based physical training program on body composition, functional fitness and depression in institutionalized older adults | Inappropriate study design |
| 7012 | Effect of transdermal nicotine administration on exercise endurance in men | Inappropriate study design |

| | | |
|---|---|---|
| 7013 | Preventing relapse in obesity through posttreatment maintenance systems: comparing the relative efficacy of two levels of therapist support | Inappropriate study design |
| 7014 | Acute mood but not cognitive improvements following administration of a single multivitamin and mineral supplement in healthy women aged 50 and above: a randomised controlled trial | Inappropriate population |
| 7015 | Yogic Breathing Changes Salivary Components | Inappropriate study design |
| 7016 | Cognitive awareness of carbohydrate intake does not alter exercise-induced lymphocyte apoptosis | Inappropriate study design |
| 7017 | Early effects of short-term aerobic training. Physiological responses to graded exercise | Inappropriate population |
| 7018 | Milk protein enriched beverage reduces post-exercise energy intakes in women with higher levels of cognitive dietary restraint | Inappropriate study design |
| 7019 | The effects of caffeine ingestion on the reaction time and short-term maximal performance after 36h of sleep deprivation | Inappropriate outcome |
| 7020 | In-patient treatment of fibromyalgia: a controlled nonrandomized comparison of conventional medicine versus integrative medicine including fasting therapy | Inappropriate population |
| 7021 | Effects of beta-blockade and exercise on cardiovascular and cognitive functioning | Inappropriate population |
| 7022 | Cost-effectiveness of a technology-supported multimodal prehabilitation program in moderate-to-high risk patients undergoing lung cancer resection: randomized controlled trial protocol | Inappropriate study design |
| 7023 | The PIT: sToPP trial- A feasibility randomised controlled trial of home-based physiotherapy for people with Parkinson's disease using video-based measures to preserve assessor blinding | Inappropriate study design |
| 7024 | Task-dependent modulation of corticospinal excitability and inhibition following strength training | Inappropriate population |
| 7025 | Effectiveness of a structured circuit class therapy model in stroke rehabilitation: a protocol for a randomised controlled trial | Inappropriate study design |
| 7026 | Effects of robot-assisted therapy on upper extremity function and activities of daily living in hemiplegic patients: a single-blinded, randomized, controlled trial | Inappropriate study design |
| 7027 | Exercise For Sub-acute Stroke Patients in Jamaica | Inappropriate study design |
| 7028 | Robot-aided neurorehabilitation: a robot for wrist rehabilitation | Inappropriate population |
| 7029 | The Effects of "Physical Activity on Prescription" in Persons With Transient Ischemic Attack: a Randomized Controlled Study | Inappropriate study design |
| 7030 | Effects of intensive repetition of a new facilitation technique on motor functional recovery of the hemiplegic upper limb and hand | Inappropriate population |
| 7031 | The synergistic benefits of physical and cognitive exercise in schizophrenia: promoting motivation to enhance community effectiveness | Inappropriate population |
| 7032 | Relaxation, reduction in angry articulated thoughts, and improvements in borderline hypertension and heart rate | Inappropriate study design |
| 7033 | Cardiorespiratory fitness moderates the effect of an affect-guided physical activity prescription: a pilot randomized controlled trial | Inappropriate population |
| 7034 | Constitutive and Stress-Induced Psychomotor Cortical Responses to Compound K Supplementation | Inappropriate population |
| 7035 | Community-based physical activity as adjunctive smoking cessation treatment: rationale, design, and baseline data for the Lifestyle Enhancement Program (LEAP) randomized controlled trial | Inappropriate study design |
| 7036 | Urge-specific and lifestyle coping strategies of alcoholics: relationships of specific strategies to treatment outcome | Inappropriate population |
| 7037 | Feasibility and acceptability of a self-managed exercise to rhythmic music intervention for ICU survivors | Inappropriate study design |
| 7038 | A Pilot Randomized Trial of an Intervention to Enhance the Health-Promoting Effects of Older Adults' Activity Portfolios: the Engaged4Life Program | Inappropriate population |
| 7039 | Exercise Trial for Alzheimer's Disease | Inappropriate study design |
| 7040 | Physical activity coaching of patients with rheumatoid arthritis in everyday practice: a long-term follow-up | Inappropriate population |
| 7041 | A tailored exercise program versus general exercise for a subgroup of patients with low back pain and movement control impairment: short-term results of a randomised controlled trial | Inappropriate outcome |
| 7042 | The effect of a cognitive-motor intervention on voluntary step execution under single and dual task conditions in older adults: a randomized controlled pilot study | Inappropriate study design |
| 7043 | Strength training does not influence serum brain-derived neurotrophic factor | Inappropriate study design |
| 7044 | Physical activityand cognitive stimulation improve cognition and alter levels of plasma beta-amyloid in healthy elderly | Inappropriate study design |

| | | |
|---|---|---|
| 7045 | Effect of acute modafinil ingestion on cognitive and physical performance following mental exertion | Inappropriate population |
| 7046 | Locomotor Training and Strength and Balance Exercises for Walking Recovery After Stroke: response to Number of Training Sessions | Inappropriate study design |
| 7047 | The central blood volume as measured by thoracic electrical impedance and plasma proANP is not compromised by donation of 900 mL of blood in men | Inappropriate study design |
| 7048 | Effect of cognitive remediation on gait in sedentary seniors | Inappropriate outcome |
| 7049 | Randomized controlled trial of yoga and exercise in multiple sclerosis | Inappropriate outcome |
| 7050 | Evaluation of a modified cognitive-behavioural programme for weight management | Inappropriate population |
| 7051 | Effectiveness of Senior Dance on risk factors for falls in older adults (DanSE): a study protocol for a randomised controlled trial | Inappropriate population |
| 7052 | Assessment of Sarcopenia Among Community-Dwelling At-Risk Frail Adults Aged 65 Years and Older Who Received Multidomain Lifestyle Interventions: a Secondary Analysis of a Randomized Clinical Trial | Inappropriate study design |
| 7053 | The PED-t trial protocol: the effect of physical exercise -and dietary therapy compared with cognitive behavior therapy in treatment of bulimia nervosa and binge eating disorder | Inappropriate population |
| 7054 | Physical Activity and Intermittent Exercise Training for Cardiovascular and Cognitive Gain in Obese Women | Inappropriate study design |
| 7055 | Twelve-Week Exercise Influences Memory Complaint but not Memory Performance in Older Adults: a Randomized Controlled Study | Inappropriate study design |
| 7056 | Assessment of an enhanced program for depression management in primary care: a cluster randomized controlled trial. The INDI project (Interventions for Depression Improvement) | Inappropriate study design |
| 7057 | Land- and water-based exercise intervention in women with fibromyalgia: the al-Andalus physical activity randomised controlled trial | Inappropriate population |
| 7058 | Cardiovascular Responses during and after Maximal Walking in Men and Women with Symptomatic Peripheral Artery Disease | Inappropriate population |
| 7059 | Randomized controlled trial of a culturally adapted, automated telephone exercise coach to improve physical activityamong hypertensive African-Americans | Inappropriate population |
| 7060 | The Self-Help, Exercise, Diet & Information Technology (SHED-IT) community trial of weight-loss programs for Australian men | Inappropriate population |
| 7061 | The role of self-efficacy, adaptive coping, and smoking urges in long-term cessation outcomes | Inappropriate population |
| 7062 | Yoga as a Complementary Therapy for Smoking Cessation | Inappropriate study design |
| 7063 | Effectiveness of a web- and mobile phone-based intervention to promote physical activity and healthy eating in middle-aged males: randomized controlled trial of the ManUp study | Inappropriate study design |
| 7064 | Comparison of human versus digital instructions for exercise in patients with hip osteoarthritis: results of a non-inferiority randomized cross-over trial | Inappropriate study design |
| 7065 | A Randomized, Controlled Trial of Exercise for Parkinsonian Individuals With Freezing of Gait | Inappropriate study design |
| 7066 | Exercise training by interval versus steady-state modus in chronic heart failure: improvement of functional capacity, hemodynamics and quality of life - A controlled study | Inappropriate study design |
| 7067 | A cognitive remediation programme for adults with Attention Deficit Hyperactivity Disorder | Inappropriate outcome |
| 7068 | Suppression of circulating free fatty acids with acipimox in chronic heart failure patients changes whole body metabolism but does not affect cardiac function | Inappropriate study design |
| 7069 | Once a week not enough, twice a week not feasible? A randomised controlled exercise trial in long-term care facilities | Inappropriate study design |
| 7070 | Exercise, Executive Processes and the Aging Brain | Inappropriate study design |
| 7071 | The EXPANd trial: effects of exercise and exploring neuroplastic changes in people with Parkinson's disease: a study protocol for a double-blinded randomized controlled trial | Inappropriate population |
| 7072 | Impact of Exercise on "Invisible" Symptoms and Quality of Life in Multiple Sclerosis Individuals | Inappropriate population |
| 7073 | The effectiveness of simple psychological and exercise interventions for high prevalence mental health problems in young people: a factorial randomised controlled trial | Inappropriate study design |
| 7074 | Effects of β-alanine supplementation on physical performance, cognition, endocrine function, and inflammation during a 24 h simulated military operation | Inappropriate study design |

| | | |
|---|---|---|
| 7075 | The effect of zolpidem on cognitive function and postural control at high altitude | Inappropriate population |
| 7076 | Strength-balance supplemented with computerized cognitive training to improve dual task gait and divided attention in older adults: a multicenter randomized-controlled trial | Inappropriate outcome |
| 7077 | Differences between energy-restricted high-protein versus high-carbohydrate, low-fat diet in weight loss and maintenance | Inappropriate study design |
| 7078 | Effects of intermittent hypoxia on cognitive performance and quality of life in elderly adults: a pilot study | Inappropriate study design |
| 7079 | Cognitive Function Improvements Mediate Exercise Intervention Effects on Physical Performance in Acutely Hospitalized Older Adults | Inappropriate population |
| 7080 | Park-ease trial: a randomized controlled trial to study the efficacy of exercises in early-stage Parkinson's disease | Inappropriate population |
| 7081 | Effect of exercise training on striatal dopamine D2/D3 receptors in methamphetamine users during behavioral treatment | Inappropriate study design |
| 7082 | Can Samba and Forró Brazilian rhythmic dance be more effective than walking in improving functional mobility and spatiotemporal gait parameters in patients with Parkinson's disease? | Inappropriate outcome |
| 7083 | Adolescent Mindfulness Mobile App Study (RCT) | Inappropriate study design |
| 7084 | Randomized trial of achieving healthy lifestyles in psychiatric rehabilitation: the ACHIEVE trial | Inappropriate population |
| 7085 | Better exercise adherence after treatment for cancer (BEAT Cancer) study: rationale, design, and methods | Inappropriate study design |
| 7086 | Two Resistance Training Protocols to Reduce the Risk of Falls in Parkinson's Disease | Inappropriate study design |
| 7087 | Efficacy of Repetitive Transcranial Magnetic Stimulation for Improvement of Memory in Older Adults With TBI | Inappropriate study design |
| 7088 | Effects of physical exercise following ischemic stroke: too early is not a good thing | Inappropriate study design |
| 7089 | Study to Validate the Effects of an Outpatient Secondary Prevention Program for Stroke Victims | Inappropriate study design |
| 7090 | Improving cardiac function and body composition through incentive-based smartphone application in sedentary overweight adults | Inappropriate study design |
| 7091 | Impact of a centre and home-based cardiac rehabilitation program on the quality of life of teenagers and young adults with congenital heart disease: the QUALI-REHAB study rationale, design and methods | Inappropriate population |
| 7092 | Identifying a low-flow phenotype in heart failure with preserved ejection fraction: a secondary analysis of the RELAX trial | Inappropriate outcome |
| 7093 | Living High-Training Low for 21 Days Enhances Exercise Economy, Hemodynamic Function, and Exercise Performance of Competitive Runners | Inappropriate population |
| 7094 | The effects of TeaCrine® and caffeine on endurance and cognitive performance during a simulated match in high-level soccer players | Inappropriate study design |
| 7095 | Sub-classification based specific movement control exercises are superior to general exercise in sub-acute low back pain when both are combined with manual therapy: a randomized controlled trial | Inappropriate study design |
| 7096 | Additive effects of exercise training upon cognitive function in regular red wine consumers compared to abstainers | Inappropriate population |
| 7097 | Cardiac Rehabilitation after Heart Valve Surgery: IMPROVEMENT in EXERCISE CAPACITY and MORBIDITY | Inappropriate study design |
| 7098 | A Randomized Trial Comparing Cardiac Rehabilitation to Standard of Care for Adults With Congenital Heart Disease | Inappropriate study design |
| 7099 | Effects of physical training on the recovery of the autonomic nervous activity during exercise after coronary artery bypass grafting: effects of physical training after CABG | Inappropriate study design |
| 7100 | Effects of Exercise Training and CPAP in Patients With Heart Failure and OSA: a Preliminary Study | Inappropriate outcome |
| 7101 | Comparative Effectiveness of Social Physical Play and Traditional Exercise Programming | Inappropriate study design |
| 7102 | Identifying responders to oral iron supplementation in heart failure with a reduced ejection fraction: a post-hoc analysis of the IRONOUT-HF trial | Inappropriate study design |
| 7103 | B.A.I.L.A. - a Latin dance randomized controlled trial for older Spanish-speaking Latinos: rationale, design, and methods | Inappropriate population |
| 7104 | Exercise videogames for physical activity and fitness: design and rationale of the Wii Heart Fitness trial | Inappropriate study design |
| 7105 | Influence of Exercise Time of Day on Salivary Melatonin Responses | Inappropriate population |

| | | |
|---|---|---|
| 7106 | The exercise-induced enhancement of influenza immunity is mediated in part by improvements in psychosocial factors in older adults | Inappropriate study design |
| 7107 | Implementation of comprehensive rehabilitation therapy in postoperative care of patients with cholangiocarcinoma and its impact on patients' quality of life | Inappropriate population |
| 7108 | Effect of voluntary hypocapnic hyperventilation on the metabolic response during Wingate anaerobic test | Inappropriate population |
| 7109 | Getting fit for hip and knee replacement: a protocol for the Fit-Joints pilot randomized controlled trial of a multi-modal intervention in frail patients with osteoarthritis | Inappropriate study design |
| 7110 | Yoga as a complementary treatment for smoking cessation in women | Inappropriate study design |
| 7111 | Effects of a lifestyle intervention program for treating obesity in lower socioeconomic status adults: a randomized controlled trial | Inappropriate population |
| 7112 | Oral Consumption of Vitamin K2 for 8 Weeks Associated With Increased Maximal Cardiac Output During Exercise | Inappropriate study design |
| 7113 | Low-dose verapamil in middle-aged and elderly patients with angina pectoris: no evidence of increased susceptibility to the cardiac effects | Inappropriate population |
| 7114 | Correlation Between Caregiver Reports of Physical Function and Performance-based Measures in a Cohort of Older Adults With Alzheimer Disease | Inappropriate study design |
| 7115 | The Arthritis, Diet and Activity Promotion Trial (ADAPT): design, rationale, and baseline results | Inappropriate study design |
| 7116 | Theory-based behavioral intervention increases self-reported physical activity in South African men: a cluster-randomized controlled trial | Inappropriate population |
| 7117 | Effects of a Multicomponent Life-Style Intervention on Weight, Glycemic Control, Depressive Symptoms, and Renal Function in Low-Income, Minority Patients With Type 2 Diabetes: results of the Community Approach to Lifestyle Modification for Diabetes Randomized Controlled Trial | Inappropriate population |
| 7118 | Patterns of exercise participation in individuals with traumatic brain injury (TBI) | Inappropriate study design |
| 7119 | Physical Activity during Cancer Treatment (PACT) Study: design of a randomised clinical trial | Inappropriate population |
| 7120 | A randomized trial to increase physical activity in breast cancer survivors | Inappropriate outcome |
| 7121 | The impact of an employee wellness programme in clothing/textile manufacturing companies: a randomised controlled trial | Inappropriate population |
| 7122 | Psychosocial modulation of antibody to Epstein-Barr viral capsid antigen and human herpesvirus type-6 in HIV-1-infected and at-risk gay men | Inappropriate population |
| 7123 | Men gain additional psychological benefits by adding exercise to a weight-loss program | Inappropriate study design |
| 7124 | Action seniors! - secondary falls prevention in community-dwelling senior fallers: study protocol for a randomized controlled trial | Inappropriate study design |
| 7125 | Upper extremity improvements in chronic stroke: coupled bilateral load training | Inappropriate study design |
| 7126 | Exercise and nutrition routine improving cancer health (ENRICH): the protocol for a randomized efficacy trial of a nutrition and physical activity program for adult cancer survivors and carers | Inappropriate population |
| 7127 | Effects of 6-month exercise training on quality of life, fatigue, and sleep problems in pancreatic cancer patients: results from a randomized controlled trial | Inappropriate study design |
| 7128 | Protein, leucine and vitamin D enhancing rehabilitation (PROLEADER) in patients with Parkinson's disease or parkinsonism: a RCT | Inappropriate study design |
| 7129 | Clinical effects of robot-assisted gait training and treadmill training for Parkinson's disease. A randomized controlled trial | Inappropriate study design |
| 7130 | A wearable proprioceptive stabilizer (Equistasi) for rehabilitation of postural instability in Parkinson's disease: a phase II randomized double-blind, double-dummy, controlled study | Inappropriate population |
| 7131 | Highly challenging balance and gait training for individuals with Parkinson's disease improves pace, rhythm and variability domains of gait - A secondary analysis from a randomized controlled trial | Inappropriate study design |
| 7132 | Motor training of upper extremity with functional electrical stimulation in early stroke rehabilitation | Inappropriate study design |
| 7133 | Cognitive and motor dual task gait training improve dual task gait performance after stroke - A randomized controlled pilot trial | Inappropriate study design |
| 7134 | ExStroke Pilot Trial of the effect of repeated instructions to improve physical activity after ischaemic stroke: a multinational randomised controlled clinical trial | Inappropriate study design |
| 7135 | Study protocol: home-based telehealth stroke care: a randomized trial for veterans | Inappropriate study design |
| 7136 | A multimodal training with visual biofeedback in subacute stroke survivors: a randomized controlled trial | Inappropriate population |

| | | |
|---|---|---|
| 7137 | Movement control exercise versus general exercise to reduce disability in patients with low back pain and movement control impairment. A randomised controlled trial | Inappropriate outcome |
| 7138 | Constraint-induced therapy versus dose-matched control intervention to improve motor ability, basic/extended daily functions, and quality of life in stroke | Inappropriate population |
| 7139 | Restitution of reaching and grasping promoted by functional electrical therapy | Inappropriate population |
| 7140 | The effects of two swallowing therapy approaches on swallowing in older adults | Inappropriate population |
| 7141 | The effect of visual scanning exercises integrated into physiotherapy in patients with unilateral spatial neglect poststroke: a matched-pair randomized control trial | Inappropriate study design |
| 7142 | Multicomponent, home-based resistance training for obese adults with type 2 diabetes: a randomized controlled trial | Inappropriate study design |
| 7143 | Efficacy of constraint-induced movement therapy for children with cerebral palsy with asymmetric motor impairment | Inappropriate study design |
| 7144 | Rationale and Design of Family-Based Approach in a Minority Community Integrating Systems-Biology for Promotion of Health (FAMILIA) | Inappropriate study design |
| 7145 | Fluvastatin and lifestyle modification for reduction of carotid intima-media thickness and left ventricular mass progression in drug-treated hypertensives | Inappropriate outcome |
| 7146 | Effects of intensive versus mild lipid lowering by statins in patients with ischemic congestive heart failure: korean Pitavastatin Heart Failure (SAPHIRE) study | Inappropriate population |
| 7147 | Experiences and perspectives of patients with post-polio syndrome and therapists with exercise and cognitive behavioural therapy | Inappropriate population |
| 7148 | Metabolic Predictors of Performance on Modified Mini Mental State (3MS) in a Randomized Trial | Inappropriate outcome |
| 7149 | Design and methods for the Ranger Resilience and Improved Performance on Phospholipid bound Omega-3's (RRIPP-3 study) | Inappropriate population |
| 7150 | The effects of exercise and nicotine replacement therapy on smoking rates in women | Inappropriate study design |
| 7151 | Impact of educational and patient decision aids on decisional conflict associated with total knee arthroplasty | Inappropriate study design |
| 7152 | Lifestyle counselling by persuasive information and communications technology reduces prevalence of metabolic syndrome in a dose–response manner: a randomized clinical trial (PrevMetSyn) | Inappropriate study design |
| 7153 | CYCLE: a Randomized Clinical Trial of Early In-bed Cycling for Mechanically Ventilated Patients | Inappropriate study design |
| 7154 | Minds "at attention": mindfulness training curbs attentional lapses in military cohorts | Inappropriate study design |
| 7155 | Independent Weight Loss Maintenance for Communities With Arthritis in North Carolina: the I-CAN Clinical Trial | Inappropriate study design |
| 7156 | Can physical therapists deliver a pain coping skills program? An examination of training processes and outcomes | Inappropriate study design |
| 7157 | The Lifestyle Intervention for the Treatment of Diabetes study (LIFT Diabetes): design and baseline characteristics for a randomized translational trial to improve control of cardiovascular disease risk factors | Inappropriate population |
| 7158 | Cardiovascular effects of the breathhold used in determining pulmonary diffusing capacity | Inappropriate study design |
| 7159 | Cognitive Behavioral Therapy and Ageism | Inappropriate study design |
| 7160 | Evaluation of Tipping Point | Inappropriate outcome |
| 7161 | Correlation of cardiac function indices and peripheral muscle mitochondrial changes in patients with severe adult growth hormone deficiency following growth hormone therapy | Inappropriate study design |
| 7162 | Effect of Senior Dance (DanSE) on Fall Risk Factors in Older Adults: a Randomized Controlled Trial | Inappropriate study design |
| 7163 | Cognitive activity for the treatment of older adults with mild Alzheimer's disease (AD)--PACE AD: study protocol for a randomised controlled trial | Inappropriate study design |
| 7164 | Preventing falls in mental health: a pilot RCT | Inappropriate study design |
| 7165 | Impacts of Omega-3-PUFA Enriched Chicken-meat and Eggs in Healthy Humans | Inappropriate intervention |
| 7166 | Breathing Muscle Training in Parkinson's Disease | Inappropriate study design |
| 7167 | Effectiveness evaluation of an integrated automatic thermomechanic massage system (SMATH® system) in non-specific sub-acute and chronic low back pain - a randomized double-blinded controlled trial, comparing SMATH therapy versus sham therapy: study protocol for a randomized controlled trial | Inappropriate population |
| 7168 | A cognitive-motor intervention using a dance video game to enhance foot placement accuracy and gait under dual task conditions in older adults: a randomized controlled trial | Inappropriate population |

| | | |
|---|---|---|
| 7169 | The relevance of dual tasking for improving trunk muscle endurance after back surgery | Inappropriate study design |
| 7170 | Randomized, controlled trial of resistance training in breast cancer patients receiving adjuvant radiotherapy: results on cancer-related fatigue and quality of life | Inappropriate study design |
| 7171 | Effects of game-based chin tuck against resistance exercise vs head-lift exercise in patients with dysphagia after stroke: an assessor-blind, randomized controlled trial | Inappropriate study design |
| 7172 | Effects of experimental emotional stress and physical exercise on the circulation in hypertensive patients and control subjects | Inappropriate study design |
| 7173 | Yearly periods of challenging balance training to prevent decline in gait and balance | Inappropriate population |
| 7174 | Benefits of a repetitive facilitative exercise program for the upper paretic extremity after subacute stroke: a randomized controlled trial | Inappropriate study design |
| 7175 | A single blind randomized controlled trial of global postural re-education: cognitive effects on Alzheimer disease patients | Inappropriate population |
| 7176 | Home based exercises to reduce falls in people with Parkinson's disease: a randomized trial | Inappropriate study design |
| 7177 | Effects of Rapid-Resisted Exercise and Bright Light Therapy on Ambulatory Adults With Traumatic Brain Injury | Inappropriate study design |
| 7178 | Does Cardiorespiratory Interval Training Improve Post-stroke Fatigue | Inappropriate study design |
| 7179 | Improving community ambulation after hip fracture: protocol for a randomised, controlled trial | Inappropriate study design |
| 7180 | Motor Imagery Exercise and Tongue Strength | Inappropriate study design |
| 7181 | The effect of muscular strenghtening exercise in motor function and balance in Parkinson's disease patients | Inappropriate study design |
| 7182 | Dual effects of body-weight supported treadmill training on cardiovascular fitness and walking ability early after stroke: a randomized controlled trial | Inappropriate study design |
| 7183 | The effects of cognitive remediation on functional outcomes among people with schizophrenia: a randomised controlled study | Inappropriate outcome |
| 7184 | Association Splitting: a randomized controlled trial of a new method to reduce craving among inpatients with alcohol dependence | Inappropriate study design |
| 7185 | How dowe measure resilience to brain aging, dementia, and Alzheimer's disease? | Inappropriate population |
| 7186 | Nutritional, Physical, Cognitive, and Combination Interventions and Frailty Reversal Among Older Adults: a Randomized Controlled Trial | Inappropriate study design |
| 7187 | Paired integrative exercise program for people with dementia and caregiviers (paired PLIÉ program) | Inappropriate study design |
| 7188 | The Healthy Hearts and Kidneys (HHK) study: design of a 2×2 RCT of technology-supported self-monitoring and social cognitive theory-based counseling to engage overweight people with diabetes and chronic kidney disease in multiple lifestyle changes | Inappropriate study design |
| 7189 | Improving cognitive function after brain injury: the use of exercise and virtual reality | Insufficient data available |
| 7190 | Transcranial direct current stimulation with halo sport enhances repeated sprint cycling and cognitive performance | Inappropriate population |
| 7191 | The effects of an 'exercise and education' programme on exercise self-efficacy and levels of independent activity in adults with acquired neurological pathologies: an exploratory, randomized study | Inappropriate study design |
| 7192 | 5-isosorbide mononitrate at rest and on exercise in coronary heart disease: acute and long-term effect | Inappropriate study design |
| 7193 | EXERCISE YOUR BRAIN NETWORKS TO PREVENT ALZHEIMER'S DISEASE | Inappropriate study design |
| 7194 | Changes in resting-state functional connectivity of the caudate following resistance exercise training in persons with MS: a preliminary report | Inappropriate study design |
| 7195 | beta-alanine supplementation improved workload capacity and cognitive function of middle age individuals | Inappropriate intervention |
| 7196 | Reduction of rise in blood pressure and cortisol release during stress by Ginkgo biloba extract (EGb 761) in healthy volunteers | Inappropriate study design |
| 7197 | Pilot Evaluation of "KeepWell" Using a Hybrid Effectiveness-Implementation Pragmatic Randomized Controlled Trial | Inappropriate intervention |
| 7198 | The effects of lower extremity cross-training on gait and balance in stroke patients: a double-blinded randomised controlled trial | Inappropriate outcome |
| 7199 | Feasibility, safety, acceptability, and functional outcomes of playing Nintendo Wii Fit PlusTM for frail older adults: a randomized feasibility clinical trial | Inappropriate study design |

| | | |
|---|---|---|
| 7200 | Improving Physical Health in Patients With Chronic Mental Disorders: twelve-Month Results From a Randomized Controlled Collaborative Care Trial | Inappropriate study design |
| 7201 | Vitamin D action in the treatment of Depression and reduction of Cardiovascular Risk | Inappropriate study design |
| 7202 | Effectiveness of constraint-induced movement therapy (CIMT) as home-based therapy on Barthel Index in patients with chronic stroke | Inappropriate outcome |
| 7203 | Virtual reality rehabilitation after stroke study (VRRASS): an inpatient blinded randomized control trial | Inappropriate study design |
| 7204 | Lack of anti-ischemic efficacy of the potassium channel opener bimakalim in patients with stable angina pectoris | Inappropriate population |
| 7205 | Efficacy of modified chin tuck against resistance exercise using hand-free device for dysphagia in stroke survivors: a randomised controlled trial | Inappropriate study design |
| 7206 | Physical Therapist-Delivered Pain Coping Skills Training and Exercise for Knee Osteoarthritis: randomized Controlled Trial | Inappropriate study design |
| 7207 | Use of nintendo wii and its effect on the balance of older adults at risk of falls: a pilot RCT | Inappropriate study design |
| 7208 | Omega 7 + 3 Combination and Systemic Inflammation | Inappropriate study design |
| 7209 | Does stress affect food preferences? - a randomized controlled trial investigating the effect of examination stress on measures of food preferences and obesogenic behavior | Inappropriate study design |
| 7210 | Balance and fear of falling in subjects with Parkinson's disease is improved after exercises with motor complexity | Inappropriate study design |
| 7211 | An integrative group movement program for people with dementia and care partners together (paired plié): initial qualitative feedback | Inappropriate study design |
| 7212 | A randomised controlled trial comparing the effectiveness of tai chi alongside usual care with usual care alone on the postural balance of community-dwelling people with dementia: protocol for the TACIT trial (TAi ChI for people with demenTia) | Inappropriate study design |
| 7213 | Cortical Î²-Amyloid in Older Adults Is Associated with Multidomain Interventions with and without Omega 3 Polyunsaturated Fatty Acid Supplementation | Inappropriate study design |
| 7214 | Preventing loss of independence through exercise in person with dementia in the va (plié-va): study protocol | Inappropriate study design |
| 7215 | FeSTivAPP - an innovative method for delivering functional strength training exercises for upper limb impairment after stroke | Inappropriate population |
| 7216 | Post-discharge management following hip fracture--get you back to B4: a parallel group, randomized controlled trial study protocol | Inappropriate study design |
| 7217 | Secondary mediation and regression analyses of the PTClinResNet database: determining causal relationships among the International Classification of Functioning, Disability and Health levels for four physical therapy intervention trials | Inappropriate study design |
| 7218 | Effects of eccentric and concentric resistance training on arterial stiffness | Inappropriate outcome |
| 7219 | DEMENTIA RISK REDUCTION IN PRIMARY CARE: RESEARCH DESIGN, PROGRESS AND CHALLENGES | Inappropriate outcome |
| 7220 | A Personalized Behavioral Intervention for Older Adults With Musculoskeletal Pain and Sleep Complaints | Inappropriate population |
| 7221 | The effect of bovine colostrum supplementation in older adults during resistance training | Inappropriate study design |
| 7222 | The Pacing Therapies for Congestive Heart Failure (PATH-CHF) study: rationale, design, and endpoints of a prospective randomized multicenter study | Inappropriate population |
| 7223 | A randomized comparative study of manually assisted versus robotic-assisted body weight supported treadmill training in persons with a traumatic brain injury | Inappropriate population |
| 7224 | The effects of a caffeine-like supplement, TeaCrine®, on muscular strength, endurance and power performance in resistance-trained men | Inappropriate population |
| 7225 | A structured physical activity and fitness programme for older adults with intellectual disabilities: results of a cluster-randomised clinical trial | Inappropriate population |
| 7226 | Sleep quality, depression state, and health status of older adults after silver yoga exercises: cluster randomized trial | Inappropriate study design |
| 7227 | Randomized Trial of Peripheral Nerve Stimulation to Enhance Modified Constraint-Induced Therapy After Stroke | Inappropriate population |
| 7228 | Unilateral strength training and mirror therapy for enhancing upper and lower limb motor function after stroke: a randomised controlled trial | Inappropriate population |
| 7229 | Effect of a six month walking intervention on platelet function in older sedentary adults: a randomised controlled trial | Inappropriate population |

| | | |
|---|---|---|
| 7230 | Cost-effectiveness of cognitive-behavioural therapy as an adjunct to pharmacotherapy for treatment-resistant depression in primary care: economic evaluation of the CoBalT Trial | Inappropriate study design |
| 7231 | A prospective study of the efficacy and safety of adjuvant metoprolol and xamoterol in combination with amiodarone for resistant ventricular tachycardia associated with impaired left ventricular function | Inappropriate study design |
| 7232 | Stepped care approach for medial tibiofemoral osteoarthritis | Inappropriate study design |
| 7233 | Functional electrical stimulation cycling does not improve mobility in people with acquired brain injury and its effects on strength are unclear: a randomised trial | Inappropriate population |
| 7234 | Power training induced change in bradykinesia and muscle power in Parkinson's disease | Inappropriate study design |
| 7235 | Fitness, Hearing and Quality of Life in Older Adults With Hearing Loss. Walk, Talk and Listen for Your Life | Inappropriate study design |
| 7236 | Intermittent hypobaric hypoxia exposure does not cause sustained alterations in autonomic control of blood pressure in young athletes | Inappropriate study design |
| 7237 | The Effect of Antioxidants on Skin Blood Flow During Local Heating | Inappropriate study design |
| 7238 | GMVs in Primary Care: an RCT of Group-Based Versus Individual Appointments to Reduce HbA1c in Older People | Inappropriate study design |
| 7239 | Promotion of healthy aging in Northern Tasmania: the benefits to older adults of resistance training | Inappropriate study design |
| 7240 | TEAMS: training and protEin recommendations for frAil older adultS (Part I: community-dwelling) (Dutch: training en Eiwit Aanbeveling op Maat voor kwetsbare Senioren - deel I: thuiswonende ouderen) | Inappropriate study design |
| 7241 | An elastic band exercise program for older adults using wheelchairs in Taiwan nursing homes: a cluster randomized trial | Inappropriate study design |
| 7242 | Cardiovascular function during supine rest in endurance-trained males with New Zealand blackcurrant: a dose-response study | Inappropriate study design |
| 7243 | Community Resilience Learning Collaborative and Research Network | Inappropriate study design |
| 7244 | Subcutaneous Median Nerve Neuromodulation For Drug-Treatment Resistant Hypertension | Inappropriate study design |
| 7245 | Can a text message a week improve breastfeeding? | Inappropriate study design |
| 7246 | Haemodynamic response to short-term daily cannabidiol | Inappropriate study design |
| 7247 | Student Wellness Workshop Study | Inappropriate intervention |
| 7248 | Sarpogrelate, a 5-hT2A receptor antagonist in intermittent claudication. A phase II European study | Inappropriate study design |
| 7249 | Shortened acquisition time or reduced-activity dose for gated myocardial perfusion SPECT with new reconstruction algorithm | Inappropriate study design |
| 7250 | The effect of exercise and induced expectations on visceral perception in asthmatic patients | Inappropriate population |
| 7251 | The effects of cognitive-behavioral and physical therapy preventive interventions on pain-related sick leave: a randomized controlled trial | Inappropriate study design |
| 7252 | Effect of an overground training session versus a treadmill training session on timed up and go in hemiparetic patients | Inappropriate population |
| 7253 | Immediate effects of blood donation on physical and cognitive performance-A randomized controlled double-blinded trial | Inappropriate study design |
| 7254 | FLY-Pilot Influence of chemotherapy, age an a single bout of endurance exercise on epigenetic modifications in tumor competetive immune cells after Non-Hodgkin Lymphoma | Inappropriate study design |
| 7255 | Comparisons between end-effector and exoskeleton rehabilitation robots regarding upper extremity function among chronic stroke patients with moderate-to-severe upper limb impairment | Inappropriate study design |
| 7256 | Effectiveness of a Combined Intervention on Psychological and Physical Capacities of Frail Older Adults: a Cluster Randomized Controlled Trial | Inappropriate population |
| 7257 | Mental Fatigue Reduces Training Volume in Resistance Exercise: a Cross-Over and Randomized Study | Inappropriate study design |
| 7258 | A comparison of three gastric lavage methods using the radionuclide gastric emptying study | Inappropriate study design |
| 7259 | Nitroglycerin-augmented 201Tl reinjection enhances detection of reversible myocardial hypoperfusion: a randomized, double-blind, parallel, placebo-controlled trial | Inappropriate population |
| 7260 | Impact of Trunk Resistance and Stretching Exercise on Fall-Related Factors in Patients with Parkinson's Disease: a Randomized Controlled Pilot Study | Inappropriate population |
| 7261 | Reduced isotope dose and imaging time with a high-efficiency CZT SPECT camera | Inappropriate population |

| | | |
|---|---|---|
| 7262 | Zinc carnosine works with bovine colostrum in truncating heavy exercise-induced increase in gut permeability in healthy volunteers | Inappropriate study design |
| 7263 | Resistance Training for Falls prevention in the Older Adults | Inappropriate study design |
| 7264 | Effects of exercise training started within 2 weeks after acute myocardial infarction on myocardial perfusion and left ventricular function: a gated SPECT imaging study | Inappropriate population |
| 7265 | Progressive resistance exercise training and changes in resting-state functional connectivity of the caudate in persons with multiple sclerosis and severe fatigue: a proof-of-concept study | Inappropriate population |
| 7266 | Outpatient cognitive behavioral pain management programs: a randomized comparison of a group-based multidisciplinary versus an individual therapy model | Inappropriate study design |
| 7267 | Effects of diltiazem i.v. on exercise induced myocardial ischaemia (a study of haemodynamics and reproducibility of haemodynamic findings | Inappropriate study design |
| 7268 | Unilateral bicep curl hemodynamics: low-pressure continuous vs high-pressure intermittent blood flow restriction | Inappropriate population |
| 7269 | Treatment outcome of chronic low back pain and radiographic lumbar disc degeneration are associated with inflammatory and matrix degrading gene variants: a prospective genetic association study | Inappropriate study design |
| 7270 | Testing mediator variables in a resistance training intervention for obese adults with type 2 diabetes | Inappropriate study design |
| 7271 | Fatigue After STroke Educational Recovery trial (FASTER) | Inappropriate study design |
| 7272 | A Digital Companion for Loneliness During the COVID-19 Pandemic: a Feasibility Study | Inappropriate population |
| 7273 | A randomized trial of a behavioral intervention for black MSM: the DiSH study | Inappropriate study design |
| 7274 | Dose reduction in half-time myocardial perfusion SPECT-CT with multifocal collimation | Inappropriate study design |
| 7275 | Internet-Based Vestibular Rehabilitation for Older Adults With Chronic Dizziness: a Randomized Controlled Trial in Primary Care | Inappropriate study design |
| 7276 | How could robotic training and botolinum toxin be combined in chronic post stroke upper limb spasticity? A pilot study | Inappropriate population |
| 7277 | Arm rehabilitation in post stroke subjects: a randomized controlled trial on the efficacy of myoelectrically driven FES applied in a task-oriented approach | Inappropriate study design |
| 7278 | A crossover pilot study evaluating the functional outcomes of two different types of robotic movement training in chronic stroke survivors using the arm exoskeleton BONES | Inappropriate study design |
| 7279 | Combined therapy with Ca-antagonists and beta-adrenergic receptor blocking agents in chronic stable angina | Inappropriate study design |
| 7280 | Power vs strength training to improve muscular strength, power, balance and functional movement in individuals diagnosed with Parkinson's disease | Inappropriate population |
| 7281 | A two-year randomized controlled trial of progressive resistance exercise for Parkinson's disease | Inappropriate study design |
| 7282 | An exercise program with patient's involvement and family support can modify the cognitive and affective trajectory of acutely hospitalized older medical patients: a pilot study | Inappropriate population |
| 7283 | Impact of limited treadmill exercise on adenosine Tc-99m sestamibi single-photon emission computed tomographic myocardial perfusion imaging in coronary artery disease | Inappropriate intervention |
| 7284 | Combined effects of robot-assisted gait training and botulinum toxin type A on spastic equinus foot in patients with chronic stroke: a pilot, single blind, randomized controlled trial | Inappropriate population |
| 7285 | The effect of high-load vs. high-repetition training on endurance performance | Inappropriate study design |
| 7286 | Metabolic responses to controlled frequency breathing in competitive swimmers | Inappropriate population |
| 7287 | Effects of sauna alone and postexercise sauna baths on blood pressure and hemodynamic variables in patients with untreated hypertension | Inappropriate population |
| 7288 | A preliminary examination of neurocognitive performance and symptoms following a bout of soccer heading in athletes wearing protective soccer headbands | Inappropriate population |
| 7289 | Ischemic Conditioning Chronic Stroke Study | Inappropriate intervention |
| 7290 | Clinical and cost effectiveness of booklet based vestibular rehabilitation for chronic dizziness in primary care: single blind, parallel group, pragmatic, randomised controlled trial | Inappropriate study design |
| 7291 | Effects of β-hydroxy-β-methylbutyrate (HMB) supplementation in addition to multicomponent exercise in adults older than 70 years living in nursing homes, a cluster randomized placebo-controlled trial: the HEAL study protocol | Inappropriate population |

| | | |
|---|---|---|
| 7292 | Comparison of Aquatic Therapy vs. Dry Land Therapy to Improve Mobility of Chronic Stroke Patients | Inappropriate study design |
| 7293 | Task-specific resistance training to improve the ability of activities of daily living-impaired older adults to rise from a bed and from a chair | Inappropriate population |
| 7294 | Impact of lowering pulmonary vascular resistance on right and left ventricular deformation in pulmonary arterial hypertension | Inappropriate study design |
| 7295 | Forearm vascular responses during semierect dynamic leg exercise in patients following myocardial infarction | Inappropriate outcome |
| 7296 | Factors influencing exercise performance in patients with left ventricular dysfunction. SOLVD Investigators. Studies of Left Ventricular Dysfunction | Inappropriate study design |
| 7297 | Effect of intravenous diltiazem on stress-induced myocardial ischemia (study of hemodynamics and reproducibility of repeated studies | Inappropriate population |
| 7298 | Microvascular response to metabolic and pressure challenge in the human coronary circulation | Inappropriate study design |
| 7299 | The psychological health benefits of mindfulness-based stress reduction in family caregivers of patients with dementia | Inappropriate population |
| 7300 | Effects of 6-month exercise training on quality of life in pancreatic cancer patients: results from a randomized controlled trial | Inappropriate population |
| 7301 | Unilateral vs Bilateral Hybrid Approaches for Upper Limb Rehabilitation in Chronic Stroke: a Randomized Controlled Trial | Inappropriate study design |
| 7302 | A randomised control trial of walking to ameliorate brain injury fatigue: a NIDRR TBI model system centre-based study | Inappropriate study design |
| 7303 | Improving balance control and self-reported lower extremity function in community-dwelling older adults: a randomized control trial | Inappropriate population |
| 7304 | The effects of virtual reality systems on balance impairments in patients with chronic stroke: a single blinded randomized controlled study | Inappropriate study design |
| 7305 | Telephone-linked Home-based Exercise Training in PD | Inappropriate study design |
| 7306 | Engaging men to maintain weight loss using innovative and cost-effective interventions: the SHED-IT Weight Loss Maintenance pilot study | Inappropriate study design |
| 7307 | Casting, taping or stretching after botulinum toxin type A for spastic equinus foot: a single-blind randomized trial on adult stroke patients | Inappropriate population |
| 7308 | Effects of Tai Chi on Multisite Pain and Falls in Older Adults | Inappropriate study design |
| 7309 | Dose response of task-specific upper limb training in people at least 6 months poststroke: a phase II, single-blind, randomized, controlled trial | Inappropriate population |
| 7310 | The effects of prior cognitive control task exposure on responses to emotional tasks in healthy participants | Inappropriate study design |
| 7311 | Spanish Diabetes Self-Management Program | Inappropriate study design |
| 7312 | Recovery of upper-limb function due to enhanced use-dependent plasticity in chronic stroke patients | Inappropriate study design |
| 7313 | Comparing the Effects of Mindfulness-Based Cognitive Therapy and Sleep Psycho-Education with Exercise on Chronic Insomnia: a Randomised Controlled Trial | Inappropriate population |
| 7314 | Sodium bicarbonate-augmented stress thallium myocardial scintigraphy | Inappropriate population |
| 7315 | Mirror Therapy Using Gesture Recognition for Upper Limb Function, Neck Discomfort, and Quality of Life After Chronic Stroke: a Single-Blind Randomized Controlled Trial | Inappropriate study design |
| 7316 | Robot-Assisted Reach Training With an Active Assistant Protocol for Long-Term Upper Extremity Impairment Poststroke: a Randomized Controlled Trial | Inappropriate study design |
| 7317 | Stroke volume measurement during supine and upright cycle exercise by impedance cardiography | Inappropriate intervention |
| 7318 | Improvements in manual dexterity relate to improvements in cognitive planning after assisted cycling therapy (ACT) in adolescents with down syndrome | Inappropriate study design |
| 7319 | Ultrasound Structural Changes in Triceps Surae After a 1-Year Daily Self-stretch Program: a Prospective Randomized Controlled Trial in Chronic Hemiparesis | Inappropriate study design |
| 7320 | Effects of toborinone on systemic circulation in patients under general anesthesia | Inappropriate study design |
| 7321 | The short-term effect of resisted grip activity on resting finger position | Inappropriate study design |
| 7322 | Lucid dreaming treatment for nightmares: a pilot study | Inappropriate study design |
| 7323 | 2013 SYR Accepted Poster Abstracts | Inappropriate study design |
| 7324 | Impact of a lifestyle intervention on metabolic pathways: results from the diet, exercise, emotional processing, and mindfulness (DEEM) intervention | Inappropriate study design |
| 7325 | Exercise imparts health benefits for CHF patients | Inappropriate population |

| | | |
|---|---|---|
| 7326 | Ischemic conditioning improves brachial artery flow mediated dilation in chronic stroke survivors | Inappropriate study design |
| 7327 | The effect of neck-specific exercise with, or without a behavioral approach, on pain, disability, and self-efficacy in chronic whiplash-associated disorders: a randomized clinical trial | Inappropriate study design |
| 7328 | Randomized clinical trial of lumbar instrumented fusion and cognitive intervention and exercises in patients with chronic low back pain and disc degeneration | Inappropriate study design |
| 7329 | Effects of training with a robot-virtual reality system compared with a robot alone on the gait of individuals after stroke | Inappropriate intervention |
| 7330 | A single-blinded, randomized pilot study of botulinum toxin type A combined with non-pharmacological treatment for spastic foot | Inappropriate population |
| 7331 | Alzheimer's disease multiple intervention trial (ADMIT): study protocol for a randomized controlled clinical trial | Inappropriate population |
| 7332 | Educating Dutch General Practitioners in Dementia Advance Care Planning: a Cluster Randomized Controlled Trial | Inappropriate study design |
| 7333 | Land-based versus pool-based exercise for people awaiting joint replacement surgery of the hip or knee: results of a randomized controlled trial | Inappropriate study design |
| 7334 | The influence of self-talk on challenge and threat states and performance | Inappropriate study design |
| 7335 | Influence of Two Different Exercise Programs on Physical Fitness and Cognitive Performance in Active Older Adults: functional Resistance-Band Exercises vs. Recreational Oriented Exercises | Inappropriate study design |
| 7336 | Similar effects of two modified constraint-induced therapy protocols on motor impairment, motor function and quality of life in patients with chronic stroke | Inappropriate population |
| 7337 | The protocol for a multisite, double blind, randomized, placebo-controlled trial of axillary nerve stimulation for chronic shoulder pain | Inappropriate study design |
| 7338 | Clinical assessment of Shenfu injection loading in the treatment of patients with exacerbation of chronic heart failure due to coronary heart disease: study protocol for a randomized controlled trial | Inappropriate study design |
| 7339 | Novel gait training alters functional brain connectivity during walking in chronic stroke patients: a randomized controlled pilot trial | Inappropriate population |
| 7340 | Genetic contribution of catechol-O-methyltransferase variants in treatment outcome of low back pain: a prospective genetic association study | Inappropriate study design |
| 7341 | Changes in cardiac function during a swallow exercise program in patients with coronary artery disease | Inappropriate outcome |
| 7342 | A randomized trial investigating the influence of strength training on quality of life in ischemic stroke | Inappropriate population |
| 7343 | Acute caffeine intake before and after fatiguing exercise improves target shooting engagement time | Inappropriate study design |
| 7344 | Adherence in positive airway pressure in naïve obstructive sleep apnea patients: the importance of the first month of treatment and the impact of telemedicine monitoring | Inappropriate population |
| 7345 | Arm weight support training improves functional motor outcome and movement smoothness after stroke | Inappropriate study design |
| 7346 | A water-based training program that includes perturbation exercises improves speed of voluntary stepping in older adults: a randomized controlled cross-over trial | Inappropriate population |
| 7347 | EFFECTIVENESS OF A DYADIC CHAIR YOGA INTERVENTION FOR REDUCING STRESS IN PATIENTS WITH DEMENTIA AND THEIR CAREGIVERS | Inappropriate study design |
| 7348 | Virtual reality-based therapy for the treatment of balance deficits in patients receiving inpatient rehabilitation for traumatic brain injury | Inappropriate study design |
| 7349 | Exercise intervention in brain injury: a pilot randomized study of Tai Chi Qigong | Inappropriate study design |
| 7350 | Effect of caffeine supplementation on repeated sprint running performance | Inappropriate study design |
| 7351 | Home-based hand rehabilitation after chronic stroke: randomized, controlled single-blind trial comparing the music glove with a conventional exercise program | Inappropriate population |
| 7352 | The effect of a wrist-hand stretching device for spasticity in chronic hemiparetic stroke patients | Inappropriate study design |
| 7353 | Voluntary postural sway discrimination sensitivity and mobility in older adults | Inappropriate study design |
| 7354 | Bosentan treatment is associated with improvement of right ventricular function and remodeling in chronic thromboembolic pulmonary hypertension | Inappropriate study design |
| 7355 | The effect of neck-specific exercise with or without a behavioral approach on psychological factors in chronic whiplash-associated disorders: a randomized controlled trial with a 2-year follow-up | Inappropriate study design |

| | | |
|---|---|---|
| 7356 | Individualized hydration plans improve performance outcomes for collegiate athletes engaging in in-season training | Inappropriate population |
| 7357 | Effect of expiratory muscle strength training intervention on the maximum expiratory pressure and quality of life of patients with Parkinson disease | Inappropriate study design |
| 7358 | Thermoregulation during prolonged exercise in heat: alterations with beta-adrenergic blockade | Inappropriate population |
| 7359 | Efficacy and Safety of Udenafil for the Treatment of Pulmonary Arterial Hypertension: a Placebo-controlled, Double-blind, Phase IIb Clinical Trial | Inappropriate population |
| 7360 | Influence of menopause status and age on integrated central and peripheral hemodynamic responses to subsystolic cuffing during submaximal exercise | Inappropriate population |
| 7361 | RAnoLazIne for the treatment of diastolic heart failure in patients with preserved ejection fraction: the RALI-DHF proof-of-concept study | Inappropriate population |
| 7362 | Physical Exercise Training versus Relaxation in Allogeneic stem cell transplantation (PETRA Study) - Rationale and design of a randomized trial to evaluate a yearlong exercise intervention on overall survival and side-effects after allogeneic stem cell transplantation | Inappropriate study design |
| 7363 | Feasibility and treatment effect of cognitive behavioral therapy for insomnia in individuals with multiple sclerosis: a pilot randomized controlled trial | Inappropriate population |
| 7364 | Mindfulness training to reduce anxiety in parkinson's disease: a cautionary tale about wearable technology | Inappropriate population |
| 7365 | Heart camp: long-term outcomes on physical function and symptoms in patients with heart failure | Inappropriate population |
| 7366 | Rehabilitation Enablement in Chronic Heart Failure-a facilitated self-care rehabilitation intervention in patients with heart failure with preserved ejection fraction (REACH-HFpEF) and their caregivers: rationale and protocol for a single-centre pilot randomised controlled trial | Inappropriate population |
| 7367 | The Effects of Pain Severity, Pain Catastrophizing, Depression, and Exercise on Perceived Disability in Acute Low Back Pain Patients | Inappropriate study design |
| 7368 | Effectiveness of upper-limb robotic-assisted therapy in the early rehabilitation phase after stroke: a single-blind, randomised, controlled trial | Inappropriate population |
| 7369 | Adherence to a depression self-care intervention among primary care patients with chronic physical conditions: a randomised controlled trial | Inappropriate intervention |
| 7370 | Effect of Add-On Yoga on Cognitive Functions among Substance Abusers in a Residential Therapeutic Center: randomized Comparative Study | Inappropriate population |
| 7371 | Japanese OMI treatment with Kit-positive cells for Enhanced Regeneration trial | Inappropriate population |
| 7372 | Assessment of a biofeedback program to treat chronic low back pain | Inappropriate study design |
| 7373 | Effects of real-time auditory stimulation feedback on balance and gait after stroke: a randomized controlled trial | Inappropriate study design |
| 7374 | Acute effect of single-session pilates based exercise intervention on cognitive function | Inappropriate population |
| 7375 | Effect of 12 weeks of once-daily tiotropium/olodaterol on exercise endurance during constant work-rate cycling and endurance shuttle walking in chronic obstructive pulmonary disease | Inappropriate population |
| 7376 | Hyperbaric Oxygen and Focused Rehabilitation Program: a Feasibility Study in Improving Upper Limb Motor Function After Stroke | Inappropriate study design |
| 7377 | Melatonin improves memory acquisition under stress independent of stress hormone release | Inappropriate study design |
| 7378 | Study protocol of a randomised controlled trial to examine the impact of a complex intervention in pre-frail older adults | Inappropriate study design |
| 7379 | 'We DECide optimized' - training nursing home staff in shared decision-making skills for advance care planning conversations in dementia care: protocol of a pretest-posttest cluster randomized trial | Inappropriate population |
| 7380 | Randomised factorial trial of falls prevention among older people living in their own homes | Inappropriate population |
| 7381 | Management of vascular and lifestyle-related risk factors for Alzheimer's disease and dementia in older adults: a european perspective | Inappropriate study design |
| 7382 | Dizziness and balance problems after traumatic brain injury (TBI): evaluation of an 8-week vestibular rehabilitation (VR) programme | Inappropriate population |
| 7383 | Cognitive Dual-Task training improves balance function in patients with stroke | Inappropriate study design |
| 7384 | Virtual Reality–Based Physical Exercise With Exergames (PhysEx) Improves Mental and Physical Health of Institutionalized Older Adults | Inappropriate population |
| 7385 | Early intervention in fatigue: a feasibility study | Inappropriate population |
| 7386 | Enhanced gait-related improvements after therapist- versus robotic-assisted locomotor training in subjects with chronic stroke: a randomized controlled study | Inappropriate study design |

| | | |
|---|---|---|
| 7387 | Training-induced changes in the pattern of triceps to biceps activation during reaching tasks after chronic and severe stroke | Inappropriate study design |
| 7388 | Balance training in virtual reality promotes superior generalization of gains in postural control to functionality in comparison conventional training-a randomized clinical trial | Inappropriate study design |
| 7389 | Dosage Matters | Inappropriate population |
| 7390 | Exercise Effectiveness on Fall Risk Factors | Inappropriate study design |
| 7391 | Randomized Controlled Pilot Trial of Behavioral Insomnia Treatment for Chronic Migraine With Comorbid Insomnia | Inappropriate study design |
| 7392 | Percutaneous renal artery denervation in patients with chronic systolic heart failure: a randomized controlled trial | Inappropriate study design |
| 7393 | Can Tai Chi training impact fractal stride time dynamics, an index of gait health, in older adults? Cross-sectional and randomized trial studies | Inappropriate population |
| 7394 | The prevalence of heart failure and asymptomatic left ventricular systolic dysfunction in a typical regional pacemaker population | Inappropriate study design |
| 7395 | The FeSTivaLS trial protocol: a randomized evaluation of the efficacy of functional strength training on enhancing walking and upper limb function later post stroke | Inappropriate study design |
| 7396 | Clinical evaluation of Functional Electrical Therapy in acute hemiplegic subjects | Inappropriate population |
| 7397 | Effects of resistive jaw opening exercise in stroke patients with dysphagia: a double- blind, randomized controlled study | Inappropriate study design |
| 7398 | Acute effects of transdermal administration of nitroglycerin on effort tolerance and myocardial perfusion, evaluated by Tl-201 scintigraphy, in patients with stable effort angina | Inappropriate study design |
| 7399 | A randomised controlled trial evaluating family mediated exercise (FAME) therapy following stroke | Inappropriate population |
| 7400 | The Effect of Acute and Chronic L-Citrulline Extract Supplementation on Vasodilatory, Nitric Oxide Bioavailability and Exercise Performance in Human Adults | Inappropriate study design |
| 7401 | Worksite interventions for preventing physical deterioration among employees in job-groups with high physical work demands: background, design and conceptual model of FINALE | Inappropriate study design |
| 7402 | A Comparison of the ARMEO to Tabletop Assisted Therapy Exercises as Supplemental Interventions in Acute Stroke Rehabilitation: a Randomized Single Blinded Study | Inappropriate intervention |
| 7403 | Dry-land strength training vs. electrical stimulation in sprint swimming performance | Inappropriate study design |
| 7404 | Impact of bed rest on conduit artery remodeling: effect of exercise countermeasures | Inappropriate population |
| 7405 | Lumbar instrumented fusion compared with cognitive intervention and exercises in patients with chronic back pain after previous surgery for disc herniation: a prospective randomized controlled study | Inappropriate study design |
| 7406 | Effect of a combined program of strength and dual cognitive-motor tasks in multiple sclerosis subjects | Inappropriate population |
| 7407 | Efficacy of low level laser therapy in lumbar discopathy pain (Randomized Controlled Study) | Inappropriate population |
| 7408 | Metabolic cost of walking with and without a shoe lift on the contralateral foot of an immobilised extended knee | Inappropriate population |
| 7409 | Feasibility of ballistic strengthening exercises in neurologic rehabilitation | Inappropriate study design |
| 7410 | Cognitive strategies and pain tolerance in subjects with muscle soreness | Inappropriate study design |
| 7411 | Exercise augmentation compared to usual care for post traumatic stress disorder: a randomised controlled trial (the REAP study: randomised Exercise Augmentation for PTSD) | Inappropriate outcome |
| 7412 | Examining the use of forceful language when designing exercise persuasive messages for adults: a test of conceptualizing reactance arousal as a two-step process | Inappropriate outcome |
| 7413 | A Structured Physical Exercise Program Reduces Professional Caregiver's Burden Caused by Neuropsychiatric Symptoms in Acute Dementia Care: randomized Controlled Trial Results | Inappropriate study design |
| 7414 | Investigation of a Lifestyle Intervention in Women at High Risk of Breast Cancer | Inappropriate study design |
| 7415 | The Effect of Bovine Colostrum Supplementation in Older Adults | Inappropriate population |
| 7416 | Combining Acceptance and Commitment Therapy With Exposure and Response Prevention to Enhance Treatment Engagement | Inappropriate study design |

| | | |
|---|---|---|
| 7417 | Study design for the fostering eating after stroke with transcranial direct current stimulation trial: a randomized controlled intervention for improving Dysphagia after acute ischemic stroke | Inappropriate intervention |
| 7418 | Cardio-respiratory fitness is associated with a verbal factor across cognitive domains in schizophrenia | Inappropriate study design |
| 7419 | Improving the effectiveness of psychological interventions for depression and anxiety in the cardiac rehabilitation pathway: a single-blind randomised controlled trial | Inappropriate population |
| 7420 | Alterations in salivary proteome following single twenty-minute session of yogic breathing | Inappropriate study design |
| 7421 | Action observation training for functional activities after stroke: a pilot randomized controlled trial | Inappropriate study design |
| 7422 | Effects of neuromuscular electrical stimulation on gait performance in chronic stroke with inadequate ankle control - A randomized controlled trial | Inappropriate intervention |
| 7423 | An active exercise and posture protocol reduced pain in acute whiplash injuries | Inappropriate outcome |
| 7424 | Task-specific ankle robotics gait training after stroke: a randomized pilot study | Inappropriate study design |
| 7425 | Changes in serum cellular adhesion molecule and matrix metalloproteinase-9 levels in patients with cerebral infarction following hyperbaric oxygen therapy: a case and intergroup control study | Inappropriate population |
| 7426 | Effectiveness of a self-rehabilitation program to improve upper-extremity function after stroke in developing countries: a randomized controlled trial | Inappropriate study design |
| 7427 | A Water-Based Sequential Preparatory Approach vs. Conventional Aquatic Training in Stroke Patients: a Randomized Controlled Trial With a 1-Month Follow-Up | Inappropriate outcome |
| 7428 | Developing a multidisciplinary rehabilitation package following hip fracture and testing in a randomised feasibility study: fracture in the Elderly Multidisciplinary Rehabilitation (FEMuR) | Inappropriate study design |
| 7429 | Effects of virtual reality training on gait biomechanics of individuals post-stroke | Inappropriate intervention |
| 7430 | Influence of an aquatic therapy program on perceived pain, stress, and quality of life in chronic stroke patients: a randomized trial | Inappropriate study design |
| 7431 | Home-based motor imagery training for gait rehabilitation of people with chronic poststroke hemiparesis | Inappropriate study design |
| 7432 | A single session of hatha yoga improves stress reactivity and recovery after an acute psychological stress task-A counterbalanced, randomized-crossover trial in healthy individuals | Inappropriate study design |
| 7433 | Effect of Ginko and Cistanche Against Fatigue Symptoms | Inappropriate study design |
| 7434 | Does self-efficacy mediate functional change in older adults participating in an exercise program after hip fracture? A randomized controlled trial | Inappropriate population |
| 7435 | Using Functional MRI to Evaluate Cognitive Behavioral Therapy Treatment Response in Adults With Chronic Pain | Inappropriate study design |
| 7436 | Acute and short-term effects of clonidine in heart failure | Inappropriate study design |
| 7437 | Rationale and design of the phase 2b clinical trials to study the effects of the partial adenosine A1-receptor agonist neladenoson bialanate in patients with chronic heart failure with reduced (PANTHEON) and preserved (PANACHE) ejection fraction | Inappropriate outcome |
| 7438 | Effectiveness of nocturnal home oxygen therapy to improve exercise capacity, cardiac function and cardiac sympathetic nerve activity in patients with chronic heart failure and central sleep apnea | Inappropriate study design |
| 7439 | Cardiac function improvement 24 hours after isradipine SRO in patients with chronic stable angina: a double-blind randomized study | Inappropriate population |
| 7440 | Clinical efficacy of functional strength training for upper limb motor recovery early after stroke: neural correlates and prognostic indicators | Inappropriate study design |
| 7441 | Body composition and training after stroke | Inappropriate study design |
| 7442 | The impact of cognitive remediation on psychiatric symptoms of schizophrenia | Inappropriate population |
| 7443 | The impact of submaximal exercise during heat and/or hypoxia on the cardiovascular and monocyte HSP72 responses to subsequent (post 24 h) exercise in hypoxia | Inappropriate study design |
| 7444 | The effects of mirror and non-mirror visual feedback conditions during unilateral exercise training on contralateral strength and passive joint position sense | Inappropriate study design |
| 7445 | Effectiveness of a Wii balance board-based system (eBaViR) for balance rehabilitation: a pilot randomized clinical trial in patients with acquired brain injury | Inappropriate study design |

| | | |
|---|---|---|
| 7446 | Effects of RM-493 on Energy Expenditure in Obese Individuals | Inappropriate study design |
| 7447 | Effects of a behavioral medicine intervention on pain, health, and behavior among community-dwelling older adults: a randomized controlled trial | Inappropriate study design |
| 7448 | The impact of a telemedicine monitoring on positive airway pressure in naïve obstructive sleep apnea patients' outcomes: a randomized controlled trial | Inappropriate population |
| 7449 | Effects of posture on cardiovascular responses to lower body positive pressure at rest and during dynamic exercise | Inappropriate study design |
| 7450 | Indenolol: a new antihypertensive agent: efficacy, toxicity, and hemodynamic effects in a crossover double-blind study with metoprolol | Inappropriate study design |
| 7451 | Training for muscle power in older adults: effects on functional abilities | Inappropriate population |
| 7452 | Mirror Training Augments the Cross-education of Strength and Affects Inhibitory Paths | Inappropriate population |
| 7453 | Protocol for promoting recovery optimization of walking activity in stroke (PROWALKS): a randomized controlled trial | Inappropriate population |
| 7454 | Unexpected effects of cognitive-behavioural therapy on self-reported exercise behaviour and functional outcomes in older adults | Inappropriate outcome |
| 7455 | Placebo Controlled Evaluation of Sedation and Physiological Response to Intranasal Dexmedetomidine in Severe COPD | Inappropriate study design |
| 7456 | Method for enhancing real-world use of a more affected arm in chronic stroke: transfer package of constraint-induced movement therapy | Inappropriate study design |
| 7457 | Community-based physical activity and nutrition programme for adults with metabolic syndrome in Vietnam: study protocol for a cluster-randomised controlled trial | Inappropriate outcome |
| 7458 | Otago home-based strength and balance retraining improves executive functioning in older fallers: a randomized controlled trial | Inappropriate study design |
| 7459 | Physical activity and nutrition program for seniors (PANS): protocol of a randomized controlled trial | Inappropriate population |
| 7460 | The efficacy of Wii-based Movement Therapy for upper limb rehabilitation in the chronic poststroke period: a randomized controlled trial | Inappropriate study design |
| 7461 | Priming the motor system enhances the effects of upper limb therapy in chronic stroke | Inappropriate outcome |
| 7462 | Activation and inhibition of the endogenous opioid system in human heart failure | Inappropriate study design |
| 7463 | Robotic-assisted rehabilitation of the upper limb after acute stroke | Inappropriate study design |
| 7464 | Quantitation of left ventricular volumes and ejection fraction in post-infarction patients from biplane and single plane two-dimensional echocardiograms. A prospective longitudinal study of 371 patients | Inappropriate study design |
| 7465 | Thermoregulatory responses to exercise in the heat: chronic caffeine intake has no effect | Inappropriate study design |
| 7466 | Comparative functional effects of chronic ventricular demand and atrial synchronous ventricular inhibited pacing | Inappropriate study design |
| 7467 | Comparative effects of propranolol and verapamil alone and in combination on left ventricular function and volumes in patients with chronic exertional angina: a double-blind, placebo-controlled, randomized, crossover study with radionuclide ventriculography | Inappropriate study design |
| 7468 | A randomized placebo-controlled trial of bupropion for Cancer-related fatigue: study design and procedures | Inappropriate study design |
| 7469 | Abstract edited-Supplemental therapeutic conventional vs. robotic upper limb exercise in acute stroke rehabilitation: a randomized, blinded assessor study | Inappropriate outcome |
| 7470 | Neurocognitive Driving Rehabilitation in Virtual Environments (NeuroDRIVE): a pilot clinical trial for chronic traumatic brain injury | Inappropriate study design |
| 7471 | Treadmill Walking Combined With Anodal Transcranial Direct Current Stimulation in Parkinson Disease: a Pilot Study of Kinematic and Neurophysiological Effects | Inappropriate study design |
| 7472 | Effects of a short message service-guided training after acute stroke or TIA (STROKEWALK): a randomized controlled trial | Inappropriate study design |
| 7473 | MBRP + Vortioxetine VS MBRP on Preventing Relapse in Chronic MA Users | Inappropriate study design |
| 7474 | Effectiveness of virtual reality devices in the rehabilitation of adults with stroke | Inappropriate study design |
| 7475 | Hemodynamic effects of medical and surgical therapy of hypertrophic obstructive cardiomyopathy (author's transl) | Inappropriate population |
| 7476 | Raised-heel stair-descent exercises as part of functional ambulation training post-stroke | Inappropriate study design |
| 7477 | Acute Whiplash Injury Study (AWIS): a protocol for a cluster randomised pilot and feasibility trial of an Active Behavioural Physiotherapy Intervention in an insurance private setting | Inappropriate population |

| | | |
|---|---|---|
| 7478 | Distinguishing active from passive components of ankle plantar flexor stiffness in stroke, spinal cord injury and multiple sclerosis | Inappropriate study design |
| 7479 | Cognitive Behavioural Therapy and Adjunctive Physical Activity for Functional Movement Disorders (Conversion Disorder): a Pilot, Single-Blinded, Randomized Study | Inappropriate study design |
| 7480 | Tai chi chuan effect on balance and fear of falling | Inappropriate study design |
| 7481 | Fluid replacement and heat stress during exercise alter post-exercise cardiac haemodynamics in endurance exercise-trained men | Inappropriate study design |
| 7482 | Randomized and comparative study between two intra-hospital exercise programs for heart transplant patients | Inappropriate intervention |
| 7483 | The effect of robot-assisted therapy and rehabilitative training on motor recovery following stroke | Inappropriate population |
| 7484 | Evaluation of individual activity descriptors of the MRC dyspnoea scale: do they add up? | Inappropriate intervention |
| 7485 | Peripheral chemosensitivity is not blunted during 2 h of thermoneutral head out water immersion in healthy men and women | Inappropriate outcome |
| 7486 | Effects of a one-year weight reduction program and physical activity on obesity and comorbid conditions | Inappropriate study design |
| 7487 | Comparison of a three-quarter electrode band configuration with a full electrode band configuration for impedance cardiography | Inappropriate study design |
| 7488 | Strength/endurance training versus endurance training in congestive heart failure | Inappropriate study design |
| 7489 | Effects of Massage on Muscular Strength and Proprioception After Exercise-Induced Muscle Damage | Inappropriate population |
| 7490 | Efficacy and cost-effectiveness of therapist-guided internet cognitive behavioural therapy for paediatric anxiety disorders: a single-centre, single-blind, randomised controlled trial | Inappropriate study design |
| 7491 | The PreFord Study. A prospective cohort study to evaluate the risk of a cardiovascular event (overall-collective) as well as a prospective, randomized, controlled, multicentre clinical intervention study (high-risk-collective) on primary prevention of cardiovascular diseases in the Ford Motor Company employees in Germany | Inappropriate population |
| 7492 | Endurance training in MS: short-term immune responses and their relation to cardiorespiratory fitness, health-related quality of life, and fatigue | Inappropriate study design |
| 7493 | Safety, Feasibility, and Efficacy of Vagus Nerve Stimulation Paired With Upper-Limb Rehabilitation After Ischemic Stroke | Inappropriate study design |
| 7494 | Proyecto Mamá: a lifestyle intervention in overweight and obese Hispanic women: a randomised controlled trial--study protocol | Inappropriate study design |
| 7495 | Effects of mental practice on affected limb use and function in chronic stroke | Inappropriate study design |
| 7496 | The Impact of Yoga-Based Physical Therapy On Heart Rate Variability After Traumatic Brain Injury | Inappropriate intervention |
| 7497 | Sickle Cell Disease and CardiovAscular Risk - Red Cell Exchange Trial (SCD-CARRE) | Inappropriate study design |
| 7498 | Managing chronic widespread pain in primary care: a qualitative study of patient perspectives and implications for treatment delivery | Inappropriate outcome |
| 7499 | Treatmentform with Mindfulness-Based Cognitive Therapy versus treatment as usual in a group of outpatients with chronic anxiety and / or mood disorder | Inappropriate study design |
| 7500 | Moderators of Response to Cognitive Behavior Therapy for Major Depression in Patients With Heart Failure | Inappropriate outcome |
| 7501 | Neurocognitive Training Integrated in OPUS Treatment Versus OPUS Treatment-as-usual | Inappropriate study design |
| 7502 | Three-dimensional, task-specific robot therapy of the arm after stroke: a multicentre, parallel-group randomised trial | Inappropriate population |
| 7503 | Recovery of left ventricular function following early reperfusion in acute myocardial infarction: a potential role for the calcium antagonist nisoldipine | Inappropriate study design |
| 7504 | Qigong intervention for breast cancer survivors with complaints of decreased cognitive function | Inappropriate study design |
| 7505 | Investigating the effects of cardio and non-cardio exercise on adults with Attention Deficit Hyperactivity Disorder (ADHD) | Inappropriate population |
| 7506 | Home-based Upper Extremity Stroke Therapy Using a Multiuser Virtual Reality Environment: a Randomized Trial | Inappropriate study design |
| 7507 | Increasing Physical Activity in Stroke Survivors Using STARFISH | Inappropriate study design |
| 7508 | Effectiveness of an Intervention Programme for Childhood Obesity: FAMILIACT | Inappropriate population |
| 7509 | Personal values and pain tolerance: does a values intervention add to acceptance? | Inappropriate population |

| | | |
|---|---|---|
| 7510 | Design considerations in a clinical trial of a cognitive behavioural intervention for the management of low back pain in primary care: back Skills Training Trial | Inappropriate population |
| 7511 | Effectiveness of the cognitive differentiation program of the integrated psychological therapy: group versus individual treatment | Inappropriate study design |
| 7512 | Dynamic post-activation potentiation protocol improves rowing performance in experienced female rowers | Inappropriate intervention |
| 7513 | Effectiveness of Suprascapular Nerve Pulsed Radiofrequency Treatment for Hemiplegic Shoulder Pain: a Randomized-Controlled Trial | Inappropriate outcome |
| 7514 | Development and evaluation of DASH diet tailored messages for hypertension treatment | Inappropriate outcome |
| 7515 | Clinical characteristics, myocardial perfusion deficits, and clinical outcomes of patients with non-specific chest pain hospitalized for suspected acute coronary syndrome: a 4-year prospective cohort study | Inappropriate study design |
| 7516 | Immediate and late modulation of interhemipheric imbalance with bilateral transcranial direct current stimulation in acute stroke | Inappropriate study design |
| 7517 | An approach in the assessment of pacing hemodynamics: a comparison of VVI and DDD | Inappropriate study design |
| 7518 | Effect of dual bronchodilation with umeclidinium vilanterol on patients with COPD, hyperinflation and heart failure | Inappropriate population |
| 7519 | IBDmindfulness-mindfulness for youth with Inflammatory Bowel Disease (IBD) and depression trial | Inappropriate study design |
| 7520 | Adolescent survivors: a secondary analysis of a clinical trial targeting behavior change | Inappropriate population |
| 7521 | Adverse events and deterioration reported by participants in the PACE trial of therapies for chronic fatigue syndrome | Inappropriate population |
| 7522 | Nocturnal non-invasive ventilation in addition to rehabilitation in hypercapnic patients with COPD | Inappropriate population |
| 7523 | Feasibility and safety of combining rtms with physical therapy: preliminary data in parkinson's disease | Inappropriate population |
| 7524 | Internet-enhanced management of fibromyalgia: a randomized controlled trial | Inappropriate study design |
| 7525 | Risedronate prevents exercise-induced hypercalcemia; a randomized controlled trial (RCT) | Inappropriate population |
| 7526 | Influence of naloxone on muscle sympathetic nerve activity, systemic and calf haemodynamics and ambulatory blood pressure after exercise in mild essential hypertension | Inappropriate study design |
| 7527 | Exercise effects on withdrawal and mood among women attempting smoking cessation | Inappropriate population |
| 7528 | Cognitive-behavioral treatment with behavioral activation for smokers with depressive symptomatology: study protocol of a randomized controlled trial | Inappropriate study design |
| 7529 | Cognitive enhancement therapy for schizophrenia: effects of a 2-year randomized trial on cognition and behavior | Inappropriate study design |
| 7530 | A cognitive remediation training for young adults with psychotic disorders to support their participation in education - Study protocol for a pilot randomized controlled trial | Inappropriate study design |
| 7531 | Acute Berry Intake on Metabolic Control and Cognitive Function | Inappropriate study design |
| 7532 | Influence of bone marrow stem cells on left ventricle perfusion and ejection fraction in patients with acute myocardial infarction of anterior wall: randomized clinical trial: impact of bone marrow stem cell intracoronary infusion on improvement of microcirculation | Inappropriate study design |
| 7533 | Addition of Supervised Exercise Training to a Post-Hospital Disease Management Program for Patients Recently Hospitalized With Acute Heart Failure: the EJECTION-HF Randomized Phase 4 Trial | Inappropriate study design |
| 7534 | Experimental acute hypoxia in healthy subjects: evaluation of systolic and diastolic function of the left ventricle at rest and during exercise using echocardiography | Inappropriate study design |
| 7535 | The access randomized clinical trial of public versus private physiotherapy for low back pain | Inappropriate study design |
| 7536 | Clinical Study of Inhaled GB002 for the Treatment of WHO Group 1 Pulmonary Arterial Hypertension (PAH) | Inappropriate study design |
| 7537 | Cost-effectiveness and safety of epidural steroids in the management of sciatica | Inappropriate study design |
| 7538 | Management of catastrophising and kinesiophobia improves rehabilitation after fusion for lumbar spondylolisthesis and stenosis. A randomised controlled trial | Inappropriate study design |
| 7539 | Central sleep apnea in patients with chronic heart failure and its treatment with acetazolamide | Inappropriate study design |
| 7540 | Exercise and cognitive performance in chronic fatigue syndrome | Inappropriate study design |

| | | |
|---|---|---|
| 7541 | Virtual Reality Rehabilitation Versus Conventional Physical Therapy for Improving Balance and Gait in Parkinson's Disease Patients: a Randomized Controlled Trial | Inappropriate study design |
| 7542 | The PACE trial of treatments for chronic fatigue syndrome: a response to WILSHIRE et al | Inappropriate population |
| 7543 | First in Human VisONE Heart Failure Study: asymptomatic Diaphragmatic Stimulation for Chronic Heart Failure: one Month Results | Inappropriate study design |
| 7544 | Effects of dual-task conditions on cervical spine movement variability | Inappropriate study design |
| 7545 | Randomised controlled trial of graded exercise in chronic fatigue syndrome | Inappropriate population |
| 7546 | Cardiopulmonary Responses to Exposure to Ozone and Diesel Exhaust | Inappropriate study design |
| 7547 | Single and dual task gait training in people with Parkinson's disease: a protocol for a randomised controlled trial | Inappropriate study design |
| 7548 | Functional walking ability 2 to 6 months after stroke: effects of progressive task-specific and impairment-based exercise rehabilitation programs compared to usual care | Inappropriate study design |
| 7549 | Robot-assisted vs. sensory integration training in treating gait and balance dysfunctions in patients with multiple sclerosis: a randomized controlled trial | Inappropriate population |
| 7550 | The Effects of Worry and Relaxation on Flexibility During Cognitive Restructuring | Inappropriate population |
| 7551 | Neck Exercises, Training and Pain Management as a Treatment for Whiplash Patients With Chronic Neck Pain | Inappropriate study design |
| 7552 | Mindfulness-based cognitive therapy v. treatment as usual in adults with ADHD: a multicentre, single-blind, randomised controlled trial | Inappropriate population |
| 7553 | High-frequency repetitive transcranial magnetic stimulation enhanced treadmill training effects on gait performance in individuals with chronic stroke: a double-blinded randomized controlled pilot trial | Inappropriate study design |
| 7554 | Brief intervention, physical exercise and cognitive behavioural group therapy for patients with chronic low back pain (The CINS trial) | Inappropriate study design |
| 7555 | Single session counselling for psychosomatic symptoms: fNdings from a pilot randomized control trial at a tertiary care center | Inappropriate study design |
| 7556 | The protocol for a randomised controlled trial comparing intermittent and graded exercise to usual care for chronic fatigue syndrome patients | Inappropriate population |
| 7557 | Qigong exercise for chronic fatigue syndrome | Inappropriate study design |
| 7558 | Treatment motivation in adolescents with psychosis or at high risk: determinants and impact on improvements in symptoms and cognitive functioning, preliminary results | Inappropriate population |
| 7559 | Cervico-vestibular Rehabilitation for Mild Traumatic Brain Injury | Inappropriate study design |
| 7560 | A classroom mind/body wellness intervention for older adults with chronic illness: comparing immediate and 1-year benefits | Inappropriate population |
| 7561 | Growth Hormone Alters Brain Morphometry, Connectivity and Behavior in Subjects with Fatigue after Mild Traumatic Brain Injury | Inappropriate outcome |
| 7562 | The role of auditory feedback in music-supported stroke rehabilitation: a single-blinded randomised controlled intervention | Inappropriate study design |
| 7563 | Assessing the impact of mindful meditation, combined with exercise, on mobility and cognitive function among older adults with chronic stroke: a proof-of-concept study | Inappropriate population |
| 7564 | Acute Effects of Assisted Cycling Therapy on Post-Stroke Motor Function: a Pilot Study | Inappropriate study design |
| 7565 | Comparing the Efficacy of CBASP with Two Versions of CBT for Depression in a Routine Care Center: a Randomized Clinical Trial | Inappropriate population |
| 7566 | Efficacy of prazosin in the management of chronic congestive heart failure: a 6-month randomized, double-blind, placebo-controlled study | Inappropriate study design |
| 7567 | Imagery-enhanced versus verbally-based group cognitive behavioural therapy for social anxiety disorder | Inappropriate study design |
| 7568 | Treatment of acute posttraumatic stress disorder with brief cognitive behavioral therapy: a randomized controlled trial | Inappropriate study design |
| 7569 | Study protocol for a non-inferiority randomised controlled trial of SKY breathing meditation versus cognitive processing therapy for PTSD among veterans | Inappropriate study design |
| 7570 | Neurocognitive responses to a single session of static squats with whole body vibration | Inappropriate population |
| 7571 | A tolerance study of single and multiple dosing of the selective dopamine uptake inhibitor GBR 12909 in healthy subjects | Inappropriate population |

| | | |
|---|---|---|
| 7572 | Influence of treadmill gait training with additional load on motor function, postural instability and history of falls for individuals with Parkinson's disease: a randomized clinical trial | Inappropriate study design |
| 7573 | The Chronic Conditions Course: a Phase III randomised controlled trial of an online psychological program for adults with chronic physical health conditions | Inappropriate study design |
| 7574 | Physiotherapy for sleep disturbance in chronic low back pain: a feasibility randomised controlled trial | Inappropriate study design |
| 7575 | Long-term effects of device-guided slow breathing in stable heart failure patients with reduced ejection fraction | Inappropriate intervention |
| 7576 | Effects of once weekly dual-task training in older adults: a pilot randomized controlled trial | Inappropriate population |
| 7577 | Does dual task training improve walking performance of older adults with concern of falling? | Inappropriate outcome |
| 7578 | Immunologic and psychologic therapy for patients with chronic fatigue syndrome: a double-blind, placebo-controlled trial | Inappropriate population |
| 7579 | Educational and Exercise Intervention to Prevent Falls and Improve Participation in Subjects With Neurological Conditions: the NEUROFALL Randomized Controlled Trial | Inappropriate study design |
| 7580 | Modeling and treating internalizing psychopathology in a clinical trial: a latent variable structural equation modeling approach | Inappropriate population |
| 7581 | A study to investigate the effects Aes-103 in patients with Sickle Cell Disease | Inappropriate study design |
| 7582 | Increased reward in ankle robotics training enhances motor control and cortical efficiency in stroke | Inappropriate study design |
| 7583 | Effect of circuit gait training vs traditional gait training on mobility performance in stroke | Inappropriate population |
| 7584 | The Effect of Occupation-Based Bilateral Upper Extremity Training in a Medical Setting for Stroke Patients: a Single-Blinded, Pilot Randomized Controlled Trial | Inappropriate study design |
| 7585 | The effects of dual-task balance training on gait in older women with osteoporosis: a randomized controlled trial | Inappropriate intervention |
| 7586 | Effects of dual-task balance training on postural performance in patients with Multiple Sclerosis: a double-blind, randomized controlled pilot trial | Inappropriate study design |
| 7587 | Treatment of traumatised refugees with basic body awareness therapy versus mixed physical activity as add-on treatment: study protocol of a randomised controlled trial | Inappropriate population |
| 7588 | A Pilot Clinical Trial to Objectively Assess the Efficacy of Electroacupuncture on Gait in Patients with Parkinson's Disease Using Body Worn Sensors | Inappropriate study design |
| 7589 | Clinical feasibility of Xbox KinectTM training for stroke rehabilitation: a single-blind randomized controlled pilot study | Inappropriate study design |
| 7590 | Advances in Vestibular Rehabilitation | Inappropriate population |
| 7591 | Multimodal treatment of persistent postural–perceptual dizziness | Inappropriate study design |
| 7592 | Hemodynamic effects of nifedipine given alone and in combination with atenolol in patients with impaired left ventricular function | Inappropriate study design |
| 7593 | Efficiency and safety of physical rehabilitation in patients with chronic heart failure III-IV class NYHA candidates for heart transplantation | Inappropriate study design |
| 7594 | Nonexcitatory, cardiac contractility modulation electrical impulses: feasibility study for advanced heart failure in patients with normal QRS duration | Inappropriate study design |
| 7595 | Effect of piboserod, a 5-HT4 serotonin receptor antagonist, on left ventricular function in patients with symptomatic heart failure | Inappropriate population |
| 7596 | Does a single gait training session performed either overground or on a treadmill induce specific short-term effects on gait parameters in patients with hemiparesis? A randomized controlled study | Inappropriate intervention |
| 7597 | Tailored, psychological intervention for anxiety or depression in people with chronic obstructive pulmonary disease (COPD), TANDEM (Tailored intervention for ANxiety and DEpression Management in COPD): protocol for a randomised controlled trial | Inappropriate population |
| 7598 | A Phase 2 Study to Investigate the Feasibility and Acceptability of the HOLOBalance System Compared to Standard Care in an Ageing Population With Balance Disorders | Inappropriate population |
| 7599 | Modulation of metoprolol pharmacokinetics and hemodynamics by diphenhydramine coadministration during exercise testing in healthy premenopausal women | Inappropriate population |
| 7600 | Structured cognitive-motor dual task training compared to single mobility training in persons with multiple sclerosis, a multicenter RCT | Inappropriate study design |

| | | |
|---|---|---|
| 7601 | Combination of Ivabradine With Low Dose of Bisoprolol Versus Optimized Dose of Bisoprolol Alone In Chronic Heart Failure Patients With Chronic Lung Disease and Risk Reduction of Stroke | Inappropriate outcome |
| 7602 | Effect of an office worksite-based yoga program on heart rate variability: a randomised controlled trial | Inappropriate study design |
| 7603 | Visual cue training to improve walking and turning after stroke: a study protocol for a multi-centre, single blind randomised pilot trial | Inappropriate study design |
| 7604 | Efficacy of graded activity versus supervised exercises in patients with chronic non-specific low back pain: protocol of a randomised controlled trial | Inappropriate study design |
| 7605 | Let's boogie: feasibility of a dance intervention in patients with chronic obstructive pulmonary disease | Inappropriate population |
| 7606 | Breathing biofeedback as an adjunct to exposure in cognitive behavioral therapy hastens the reduction of PTSD symptoms: a pilot study | Inappropriate study design |
| 7607 | Bilateral Arm Training vs Unilateral Arm Training for Severely Affected Patients With Stroke: exploratory Single-Blinded Randomized Controlled Trial | Inappropriate study design |
| 7608 | Treadmill training with music cueing: a new approach for Parkinson's gait facilitation | Inappropriate population |
| 7609 | Effects of Vestibular Rehabilitation on Fatigue and Activities of Daily Living in People with Parkinson's Disease: a Pilot Randomized Controlled Trial Study | Inappropriate population |
| 7610 | Effects of Treadmill Training with Load on Gait in Parkinson Disease: a Randomized Controlled Clinical Trial | Inappropriate study design |
| 7611 | Physical Exercise and Jaques-Dalcroze Eurhythmics: effects on Physical and Cognitive Functions, and Falls in Seniors | Inappropriate study design |
| 7612 | Simultaneous inhibition of catechol-O-methyltransferase and monoamine oxidase A: effects on hemodynamics and catecholamine metablism in healthy volunteers | Inappropriate study design |
| 7613 | Perceived Activities and Participation Outcomes of a Yoga Intervention for Individuals with Parkinson's Disease: a Mixed Methods Study | Inappropriate population |
| 7614 | Effects of handwriting exercise on functional outcome in Parkinson's disease: a randomized controlled trial | Inappropriate population |
| 7615 | Four-week trunk-specific exercise program decreases forward trunk flexion in Parkinson's disease: a single-blinded, randomized controlled trial | Inappropriate study design |
| 7616 | Postural rehabilitation and kinesio taping for axial postural disorders in Parkinson's disease | Inappropriate population |
| 7617 | Study protocol for a single-blind randomised controlled trial to evaluate the clinical effects of an Integrated Qigong exercise intervention on freezing of gait in Parkinson's disease | Inappropriate study design |
| 7618 | STABLE (Strength and Balance Leg Exercise) Study | Inappropriate population |
| 7619 | Walking after stroke: what does treadmill training with body weight support add to overground gait training in patients early after stroke?: a single-blind, randomized, controlled trial | Inappropriate population |
| 7620 | Robot Assisted Training for the Upper Limb after Stroke (RATULS): study protocol for a randomised controlled trial | Inappropriate population |
| 7621 | Anemia in acute heart failure | Inappropriate population |
| 7622 | Long-term effects of autologous bone marrow CD133+ cell transplantation on myocardial structure and function after acute myocardial infarction | Inappropriate population |
| 7623 | Early signaling, referral, and treatment of adolescent chronic pain: a study protocol | Inappropriate study design |
| 7624 | Investigation of the Effects of Dual Task Balance Training on Gait and Balance in Transfemoral Amputees: a Randomised Controlled Trial | Inappropriate study design |
| 7625 | Three-Dimensional Balance Training Using Visual Feedback on Balance and Walking Ability in Subacute Stroke Patients: a Single-Blinded Randomized Controlled Pilot Trial | Inappropriate population |
| 7626 | Therapist-aided exposure for women with lifelong vaginismus: a replicated single-case design | Inappropriate study design |
| 7627 | Topiramate for migraine prevention | Inappropriate outcome |
| 7628 | Efficacy of internet-delivered cognitive behavioural therapy following an acute coronary event: a randomized controlled trial | Inappropriate study design |
| 7629 | Aquatic Therapy Improves Outcomes for Subacute Stroke Patients by Enhancing Muscular Strength of Paretic Lower Limbs Without Increasing Spasticity: a Randomized Controlled Trial | Inappropriate study design |
| 7630 | A dual-task home-based rehabilitation programme for improving balance control in patients with acquired brain injury: a single-blind, randomized controlled pilot study | Inappropriate study design |

| | | |
|---|---|---|
| 7631 | Effects of constraint-induced movement therapy for lower limbs on measurements of functional mobility and postural balance in subjects with stroke: a randomized controlled trial | Inappropriate population |
| 7632 | Impact of mental fatigue on self-paced exercise | Inappropriate population |
| 7633 | Robotic-assisted gait training combined with transcranial direct current stimulation in chronic stroke patients: a pilot double-blind, randomized controlled trial | Inappropriate population |
| 7634 | Efficacy of gait training using a treadmill with and without visual biofeedback in patients after stroke: a randomized study | Inappropriate study design |
| 7635 | Effects of robot-assisted gait training on cardiopulmonary fitness in subacute stroke patients: a randomized controlled study | Inappropriate study design |
| 7636 | Exercises for paretic upper limb after stroke: a combined virtual-reality and telemedicine approach | Inappropriate study design |
| 7637 | 10% Higher Rowing Power Outputs After Flexion-Extension-Cycle Compared to an Isolated Concentric Contraction in Sub-Elite Rowers | Inappropriate population |
| 7638 | Evaluation of Tai Chi Yunshou exercises on community-based stroke patients with balance dysfunction: a study protocol of a cluster randomized controlled trial | Inappropriate study design |
| 7639 | Efficacy of a Virtual Reality Commercial Gaming Device in Upper Limb Recovery after Stroke: a Randomized, Controlled Study | Inappropriate study design |
| 7640 | Do Core Stability Exercise Classes Help Children With Cerebral Palsy Improve Their Balance | Inappropriate study design |
| 7641 | Improving balance with wobble board exercises in stroke patients: single-blind, randomized clinical trial | Inappropriate study design |
| 7642 | Clinical impact of gait training enhanced with visual kinematic biofeedback: patients with Parkinson's disease and patients stable post stroke | Inappropriate population |
| 7643 | Computerized arm training improves the motor control of the severely affected arm after stroke: a single-blinded randomized trial in two centers | Inappropriate study design |
| 7644 | Effects of ULEM apparatus on motor function of patients with stroke | Inappropriate study design |
| 7645 | Effectiveness of LiuZiJue Qigong versus traditional core stability training for poststroke patients complicated with abnormal trunk postural control: study protocol for a single-center randomized controlled trial | Inappropriate population |
| 7646 | A mechanical arm trainer for the treatment of the severely affected arm after stroke: a single-blinded randomized trial in two centres | Inappropriate study design |
| 7647 | Virtual Reality Therapy Can Improve Fatigue and Cognitive Function after Radiation and Chemotherapy | Inappropriate study design |
| 7648 | Improving neurocognitive functioning in schizophrenia by addition of cognitive remediation therapy to a standard treatment of metacognitive training | Inappropriate population |
| 7649 | Comparison between treadmill training with rhythmic auditory stimulation and ground walking with rhythmic auditory stimulation on gait ability in chronic stroke patients: a pilot study | Inappropriate population |
| 7650 | eHealth-supported case management for patients with panic disorder or depression in primary care: study protocol for a cRCT (PREMA) | Inappropriate study design |
| 7651 | Improving Health and Reducing Disability of Depressed Elderly With Chronic Conditions Through Qigong Exercise | Inappropriate study design |
| 7652 | Does rehabilitation improve walking performance? | Inappropriate study design |
| 7653 | Adhesive taping vs. daily manual muscle stretching and splinting after botulinum toxin type A injection for wrist and fingers spastic overactivity in stroke patients: a randomized controlled trial | Inappropriate study design |
| 7654 | A brief bout of exercise in hypoxia reduces ventricular filling rate and stroke volume response during muscle metaboreflex activation | Inappropriate study design |
| 7655 | International randomized clinical trial, stroke inpatient rehabilitation with reinforcement of walking speed (SIRROWS), improves outcomes | Inappropriate population |
| 7656 | The impact of a group exercise programme on usual walking performance in adults who are at least 6 months post stroke: a single blinded randomised controlled trial | Inappropriate population |
| 7657 | Can robotic gait rehabilitation plus Virtual Reality affect cognitive and behavioural outcomes in patients with chronic stroke? A randomized controlled trial involving three different protocols | Inappropriate intervention |
| 7658 | The effects of 1Hz low frequency transcranial magnetic stimulation on swallowing function in post-stroke dysphagia | Inappropriate study design |
| 7659 | Effects of treadmill inclination on hemiparetic gait: controlled and randomized clinical trial | Inappropriate study design |
| 7660 | Effects of Home-Based Versus Clinic-Based Rehabilitation Combining Mirror Therapy and Task-Specific Training for Patients With Stroke: a Randomized Crossover Trial | Inappropriate study design |

| | | |
|---|---|---|
| 7661 | Adaptive Physical Activity for Chronic Stroke | Inappropriate study design |
| 7662 | Bilateral and unilateral movement training on upper limb function in chronic stroke patients: a TMS study | Inappropriate population |
| 7663 | Efficacy of physician-delivered brief counselling intervention for binge drinkers: randomised controlled trial in primary care practice | Inappropriate population |
| 7664 | Prevent it: can internet-delivered psychotherapy reduce usage of online child sexual abuse material? | Inappropriate study design |
| 7665 | Combined Effects of Prolonged Sitting and Mental Stress on the Cardiovascular System | Inappropriate study design |
| 7666 | The effects of the Bali Yoga Program (BYP-BC) on reducing psychological symptoms in breast cancer patients receiving chemotherapy: results of a randomized, partially blinded, controlled trial | Inappropriate study design |
| 7667 | Primum non nocere: shared informed decision making in low back pain--a pilot cluster randomised trial | Inappropriate study design |
| 7668 | Identifying attendance patterns in a smoking cessation treatment and their relationships with quit success | Inappropriate population |
| 7669 | Functional electrical stimulation-assisted active cycling--therapeutic effects in patients with hemiparesis from 7 days to 6 months after stroke: a randomized controlled pilot study | Inappropriate study design |
| 7670 | Acute Supplementation with Nitrate-Rich Beetroot Juice Causes a Greater Increase in Plasma Nitrite and Reduction in Blood Pressure of Older Compared to Younger Adults | Inappropriate study design |
| 7671 | Effects of Simultaneous or Sequential Combination of Physical and Cognitive Training | Inappropriate population |
| 7672 | Treadmill walking with partial body weight support versus floor walking in hemiparetic subjects | Inappropriate population |
| 7673 | Unilateral and bilateral upper-limb training interventions after stroke have similar effects on bimanual coupling strength | Inappropriate study design |
| 7674 | Prescribed Light Exercise for ED Patients With MTBI | Inappropriate study design |
| 7675 | The effectiveness of Cognitive Behavioral Therapy (CBT) with general exercises versus general exercises alone in the management of chronic low back pain | Inappropriate intervention |
| 7676 | Comparing the effects of cognitive and social stress among individuals with headache: an experimental manipulation | Inappropriate study design |
| 7677 | Physiotherapy and Psychological Among Refugees From Syria | Inappropriate study design |
| 7678 | An integrated motivational interviewing and cognitive-behavioural intervention promoting physical activity maintenance for adults with chronic health conditions: a feasibility study | Inappropriate study design |
| 7679 | Effectiveness of repetitive transcranial magnetic stimulation and visual feedback exercise on neuroplasticity and lower limb function in person with chronic stroke | Inappropriate population |
| 7680 | Evaluation of a practice team-supported exposure training for patients with panic disorder with or without agoraphobia in primary care - study protocol of a cluster randomised controlled superiority trial | Inappropriate study design |
| 7681 | Delivering an Optimised Behavioural Intervention (OBI) to people with low back pain with high psychological risk; results and lessons learnt from a feasibility randomised controlled trial of Contextual Cognitive Behavioural Therapy (CCBT) vs. Physiotherapy | Inappropriate study design |
| 7682 | Technology-assisted balance and gait training rreduces falls in patients with Parkinson's disease: a randomized controlled trial with 12-month follow-up | Inappropriate study design |
| 7683 | Use of game console for rehabilitation of Parkinson's disease | Inappropriate population |
| 7684 | The application of mobile health technology to increase physical activity in persons with Parkinson's disease | Inappropriate population |
| 7685 | Effect of health Baduanjin Qigong for mild to moderate Parkinson's disease | Inappropriate intervention |
| 7686 | On-field testing environment and balance error scoring system performance during preseason screening of healthy collegiate baseball players | Inappropriate study design |
| 7687 | Immediate effects of perturbation treadmill training on gait and postural control in patients with Parkinson's disease | Inappropriate study design |
| 7688 | //Therapist-supervised compared to home-based balance training encourages a 'posture first' strategy during turn-to-sit transitions in individuals with Parkinson's disease | Inappropriate study design |
| 7689 | Dynamic soft tissue mobilisation increases hamstring flexibility in healthy male subjects | Inappropriate population |
| 7690 | Comparison of the effectiveness of two programmes on older adults at risk of falling: unsupervised home exercise and supervised group exercise | Inappropriate study design |

| | | |
|---|---|---|
| 7691 | Spinal use-dependent plasticity of synaptic transmission in humans after a single cycling session | Inappropriate population |
| 7692 | How relevant are exercise capacity measures for evaluating treatment effects in chronic fatigue syndrome? results from a prospective, multidisciplinary outcome study | Inappropriate outcome |
| 7693 | The Effects of Abdominal Massage on Functional (Primary) Chronic Constipation | Inappropriate study design |
| 7694 | Robotic-Assisted Shoulder Rehabilitation Therapy Effectively Improved Poststroke Hemiplegic Shoulder Pain: a Randomized Controlled Trial | Inappropriate study design |
| 7695 | Comparative effects of nitroglycerin, nifedipine and metoprolol on regional left ventricular function in patients with one-vessel coronary disease | Inappropriate study design |
| 7696 | The effects of repeated thermal therapy for patients with chronic pain | Inappropriate study design |
| 7697 | Cognitive behavioral stress management effects on injury and illness among competitive athletes: a randomized clinical trial | Inappropriate study design |
| 7698 | No Reduction of Severe Fatigue in Patients With Postpolio Syndrome by Exercise Therapy or Cognitive Behavioral Therapy: results of an RCT | Inappropriate study design |
| 7699 | Is increasing physical activity necessary to diminish fatigue during cancer treatment? Comparing cognitive behavior therapy and a brief nursing intervention with usual care in a multicenter randomized controlled trial | Inappropriate study design |
| 7700 | The effects of dual-tasking on temporal gait adaptation and de-adaptation to the split-belt treadmill in older adults | Inappropriate population |
| 7701 | Ankle flexors produce peak torque at longer muscle lengths after whole-body vibration | Inappropriate population |
| 7702 | Improving anxiety regulation in patients with breast cancer at the beginning of the survivorship period: a randomized clinical trial comparing the benefits of single-component and multiple-component group interventions | Inappropriate population |
| 7703 | Effects of whole body vibration training using side-alternating vibration platform with tilt table in hospitalized older adults with sarcopenia: a randomized controlled pilot study | Inappropriate population |
| 7704 | Efficacy of a child-friendly form of constraint-induced movement therapy in hemiplegic cerebral palsy: a randomized control trial | Inappropriate study design |
| 7705 | Quality of life and exercise capacity in patients with prolonged PQ interval and dual chamber pacemakers: a randomized comparison of permanent ventricular stimulation vs intrinsic AV conduction | Inappropriate study design |
| 7706 | Effects of gait and balance training on gait and balance in older adults | Inappropriate study design |
| 7707 | Attention training to reduce attention bias and social stressor reactivity: an attempt to replicate and extend previous findings | Inappropriate population |
| 7708 | Battlefield-like stress following simulated combat and suppression of attention bias to threat | Inappropriate study design |
| 7709 | Bodyweight-supported treadmill training for retraining gait among chronic stroke survivors: a randomized controlled study | Inappropriate population |
| 7710 | Comparison of Brunnstrom movement therapy and Motor Relearning Program in rehabilitation of post-stroke hemiparetic hand: a randomized trial | Inappropriate outcome |
| 7711 | Biological determinants of P300: the effects of a barbiturate on latency and amplitude | Inappropriate study design |
| 7712 | Effects of Pilates Standing Exercises on Walking Mobility and Postural Balance | Inappropriate study design |
| 7713 | Bio-psycho-social Pathways Underlying the Effects of Qigong Among Comorbid Depressed Elderly With Chronic Conditions | Inappropriate population |
| 7714 | Effect of switching to atypical antipsychotics on memory in patients with chronic schizophrenia | Inappropriate population |
| 7715 | Treatment of fibromyalgia differentiating profiles in adherence to walking behavior in women with fibromyalgia after the application of a brief motivational-volitional intervention | Inappropriate population |
| 7716 | Evaluating an internet-delivered cognitive behavioural therapy program for reducing depression and anxiety after a cardiac event | Inappropriate study design |
| 7717 | Backward walking observational training improves gait ability in patients with chronic stroke: randomised controlled pilot study | Inappropriate population |
| 7718 | Segmental muscle vibration improves walking in chronic stroke patients with foot drop: a randomized controlled trial | Inappropriate study design |
| 7719 | Is a behavioral graded activity program more effective than manual therapy in patients with subacute neck pain? Results of a randomized clinical trial | Inappropriate study design |
| 7720 | ICT-based system to predict and prevent falls (iStoppFalls): study protocol for an international multicenter randomized controlled trial | Inappropriate study design |
| 7721 | Size acceptance and intuitive eating improve health for obese, female chronic dieters | Inappropriate study design |

| | | |
|---|---|---|
| 7722 | The effects of clinical pilates training in patients with fibromyalgia: a randomized controlled trial | Inappropriate population |
| 7723 | CapaCiTY study 1-chronic constipation treatment pathway | Inappropriate study design |
| 7724 | Minimal effects of age and prolonged physical and mental exercise on healthy adultsâ€™ gait | Inappropriate study design |
| 7725 | Balance exercise program reduced falls in people with multiple sclerosis: a single-group, pretest-posttest trial | Inappropriate population |
| 7726 | Rationale, design and methodology for a Prospective Randomized Study of graft patency in Off-pump and On-pump MultI-Vessel coronary artery bypasS Surgery (PROMISS) using multidetector computed tomography | Inappropriate population |
| 7727 | Can 'strength snacking' offset losses of muscle mass and strength with physical inactivity in older adults? | Inappropriate study design |
| 7728 | Compare effect of PNF for trunk versus weight shift therapy on trunk control and dynamic balance in chronic hemiplegia | Inappropriate study design |
| 7729 | Chronicity Dependence of a Balance Training in Adults Post-stroke | Inappropriate study design |
| 7730 | A prospective, randomised, doubleblind study effect of levodopa in combination with physiotherapy on functional motor recovery after stroke | Inappropriate outcome |
| 7731 | Efficacy of Bobath versus orthopaedic approach on impairment and function at different motor recovery stages after stroke: a randomized controlled study | Inappropriate population |
| 7732 | Efficacy of the Nintendo Wii combination with Conventional Exercises in the rehabilitation of individuals with Parkinson's disease: a randomized clinical trial | Inappropriate study design |
| 7733 | Comparison of the effects of a physiotherapist-supervised exercise programme and a self-supervised exercise programme on quality of life in patients with Parkinson's disease | Inappropriate population |
| 7734 | Comparing computer-aided therapy with conventional physiotherapy in Parkinsonâ€™s disease: an equivalence study | Inappropriate study design |
| 7735 | Effect of balance training on postural instability in patients with idiopathic Parkinson's disease | Inappropriate population |
| 7736 | The Effect of Breathing Exercises on the Sleep Quality of Older Adults with Chronic Lung Disease | Inappropriate population |
| 7737 | The Maintaining Musculoskeletal Health Study | Inappropriate study design |
| 7738 | Fatigue Is Associated With Altered Monitoring and Preparation of Physical Effort in Patients With Chronic Fatigue Syndrome | Inappropriate population |
| 7739 | Effects of telerehabilitation on physical function and disability for stroke patients: a randomized, controlled trial | Inappropriate population |
| 7740 | Treadmill gait training combined with functional electrical stimulation on hip abductor and ankle dorsiflexor muscles for chronic hemiparesis | Inappropriate study design |
| 7741 | Ten weeks of physical-cognitive-mindfulness training reduces fear-avoidance beliefs about work-related activity: randomized controlled trial | Inappropriate population |
| 7742 | Embracing complexity in the development of a community-based exercise transition program for community-dwelling individuals with chronic stroke: results of a scoping review and focus group interviews | Inappropriate study design |
| 7743 | Sequential combination of robot-assisted therapy and constraint-induced therapy in stroke rehabilitation: a randomized controlled trial | Inappropriate population |
| 7744 | Does robot-assisted gait training improve mobility, activities of daily living and quality of life in stroke? A single-blinded, randomized controlled trial | Inappropriate population |
| 7745 | Interdisciplinary Comprehensive Arm Rehabilitation Evaluation (ICARE): a randomized controlled trial protocol | Inappropriate population |
| 7746 | Effect of Stride Management Assist Gait Training for Poststroke Hemiplegia: a Single Center, Open-Label, Randomized Controlled Trial | Inappropriate study design |
| 7747 | Pelvic floor muscle training is effective in women with urinary incontinence after stroke: a randomised, controlled and blinded study | Inappropriate population |
| 7748 | Varied overground walking-task practice versus body-weight-supported treadmill training in ambulatory adults within one year of stroke: a randomized controlled trial protocol | Inappropriate population |
| 7749 | Prediction of responders for outcome measures of locomotor Experience Applied Post Stroke trial | Inappropriate study design |
| 7750 | Effects of “Trunk Dissociation Retrainer” in Improving Trunk Performance and Functional Activities in Hemiplegia | Inappropriate population |
| 7751 | Comparative effects of oral verapamil and propranolol on exercise-induced myocardial ischemia and energetics in patients with coronary artery disease: single-blind placebo crossover evaluation using radionuclide ventriculography | Inappropriate study design |
| 7752 | Does Perturbation-Based Balance Training Improve Control of Reactive Stepping in Individuals with Chronic Stroke? | Inappropriate study design |

| | | |
|---|---|---|
| 7753 | The Virgin PulsE Global ChAllenge Study (VEGAS): a Single-Blind, Randomised Controlled Trial to Evaluate the Health Outcomes of the Virgin Pulse Global Challenge (VPGC) Programme | Inappropriate study design |
| 7754 | Community-based rehabilitation training after stroke: results of a pilot randomised controlled trial (ReTrain) investigating acceptability and feasibility | Inappropriate population |
| 7755 | The addition of virtual reality training for sitting balance to inpatient rehabilitation in the subacute stage post-stroke | Inappropriate study design |
| 7756 | Living better beyond pain: randomized trial of office-versus community-based self-management of chronic pain | Inappropriate study design |
| 7757 | Compelled body weight shift approach in rehabilitation of individuals with chronic stroke | Inappropriate population |
| 7758 | Does an online psychological intervention improve self-efficacy and disability in people also receiving Multimodal Manual Therapy for chronic low back pain compared to Multimodal Manual Therapy alone? Design of a randomized controlled trial | Inappropriate outcome |
| 7759 | Liuzijue Qigong vs traditional breathing training for patients with post-stroke dysarthria complicated with abnormal respiratory control: study protocol of a single center randomized controlled trial | Inappropriate study design |
| 7760 | Comparative Effect of Liuzijue Qigong and Conventional Respiratory Training on Trunk Control Ability and Respiratory Muscle Function in Patients at an Early Recovery Stage From Stroke: a Randomized Controlled Trial | Inappropriate population |
| 7761 | Effect of constraint-induced movement therapy with modified opposition restriction orthosis in chronic hemiparetic patients with stroke | Inappropriate population |
| 7762 | Efficacy of exercise rehabilitation program accompanied by experiential music for recovery of aphasia in single cerebrovascular accidents: a randomized controlled trial | Inappropriate population |
| 7763 | SMART Arm with outcome-triggered electrical stimulation: a pilot randomized clinical trial | Inappropriate study design |
| 7764 | The impact of transcranial direct current stimulation (tDCS) combined with modified constraint-induced movement therapy (mCIMT) on upper limb function in chronic stroke: a double-blind randomized controlled trial | Inappropriate outcome |
| 7765 | Effects of Kinect-based virtual reality game training on upper extremity motor recovery in chronic stroke | Inappropriate study design |
| 7766 | Twelve-month effect of chronic pain self-management intervention delivered in an easily accessible primary healthcare service - a randomised controlled trial | Inappropriate study design |
| 7767 | Physical exercise program to improve cognitive function in older adults | Inappropriate study design |
| 7768 | Can a cognitive therapy group intervention improve glycemia in patients with poorly controlled diabetes? | Inappropriate study design |
| 7769 | Effects of a 12-Week Digital Care Program for Chronic Knee Pain on Pain, Mobility, and Surgery Risk: randomized Controlled Trial | Inappropriate study design |
| 7770 | Right ventricular function and effort tolerance in patients with chronic congestive heart insufficiency. A cross-over double-blind study | Inappropriate study design |
| 7771 | The impact of stroke nurse specialist input on risk factor modification: a randomised controlled trial | Inappropriate study design |
| 7772 | Improving Quality of Life and Depression After Stroke Through Telerehabilitation | Inappropriate intervention |
| 7773 | Further effects of electromechanically assisted gait trainer (Exowalk®) in patients with chronic stroke: a randomized controlled trial | Inappropriate study design |
| 7774 | Effects of combining robot-assisted therapy with neuromuscular electrical stimulation on motor impairment, motor and daily function, and quality of life in patients with chronic stroke: a double-blinded randomized controlled trial | Inappropriate intervention |
| 7775 | Protocol for the Optimune trial: a randomized controlled trial evaluating a novel Internet intervention for breast cancer survivors | Inappropriate study design |
| 7776 | EFFICACY OF EGb 761® 120 mg bid VERSUS PLACEBO IN PATIENTS SUFFERING FROM FRIEDREICH ATAXIA A 3 months, phase II, randomised, double blind, placebo-controlled, parallel groups, clinical study. - EGB 761 in FRIEDREICH ATAXIA | Inappropriate study design |
| 7777 | Training Improves Balance Control | Inappropriate study design |
| 7778 | Home-based nerve stimulation to enhance effects of motor training in patients in the chronic phase after stroke: a proof-of-principle study | Inappropriate population |
| 7779 | Differential effects of water-based exercise on the cognitive function in independent elderly adults | Inappropriate study design |
| 7780 | Evaluation of the chronic disease self-management program in a Chinese population | Inappropriate population |

| | | |
|---|---|---|
| 7781 | The impact of remote ischemic preconditioning on cardiac biomarker and functional response to endurance exercise | Inappropriate population |
| 7782 | Does Change Occur for the Reasons We Think It Does? A Test of Specific Therapeutic Operations During Cognitive-Behavioral Treatment of Chronic Pain | Inappropriate study design |
| 7783 | Over-ground and robotic-assisted locomotor training in adults with chronic stroke: a blinded randomized clinical trial | Inappropriate population |
| 7784 | Evaluating a 'non-diet' wellness intervention for improvement of metabolic fitness, psychological well-being and eating and activity behaviors | Inappropriate population |
| 7785 | Adaptation of a Health Promotion Nutrition Program for Older Adults | Inappropriate study design |
| 7786 | Randomized controlled trial on the effectiveness of cognitive behavior group therapy in chronic back pain patients | Inappropriate population |
| 7787 | Pain in chronic fatigue syndrome: response to rehabilitative treatments in the PACE trial | Inappropriate population |
| 7788 | A short goal-pursuit intervention to improve physical capacity: a randomized clinical trial in chronic back pain patients | Inappropriate outcome |
| 7789 | The effects of a personal oxygen supplement on performance, recovery, and cognitive function during and after exhaustive exercise | Inappropriate population |
| 7790 | Effects of diagonally aligned sitting training with a tilted surface on sitting balance for low sitting performance in the early phase after stroke: a randomised controlled trial | Inappropriate study design |
| 7791 | Hand splinting for poststroke spasticity: a randomized controlled trial | Inappropriate study design |
| 7792 | Effects of exercises with action observation and aquatic physical therapy in arm recovery after stroke | Inappropriate outcome |
| 7793 | Randomized controlled trial of truncal exercises early after stroke to improve balance and mobility | Inappropriate study design |
| 7794 | Better balance: a randomised controlled trial of oculomotor and gaze stability exercises to reduce risk of falling after stroke | Inappropriate population |
| 7795 | Peer-delivered cognitive behavior intervention reduced depression and stress | Inappropriate population |
| 7796 | Effect of the Nursing Process on the Quality of Life of the Patient With Definitive Pacemaker Implantation | Inappropriate study design |
| 7797 | Differences in long-term physical activity trajectories among individuals with chronic widespread pain: a secondary analysis of a randomized controlled trial | Inappropriate population |
| 7798 | Intervening to reduce workplace sitting: mediating role of social-cognitive constructs during a cluster randomised controlled trial | Inappropriate study design |
| 7799 | A randomized-controlled pilot trial of an online compassionate mind training intervention to help people with chronic pain avoid analgesic misuse | Inappropriate study design |
| 7800 | Effects of oral amino acid supplementation on myocardial function in patients with type 2 diabetes mellitus | Inappropriate study design |
| 7801 | New Nutraceutical Combination Reduces Blood Pressure and Improves Exercise Capacity in Hypertensive Patients Via a Nitric Oxide-Dependent Mechanism | Inappropriate population |
| 7802 | Effect of pioglitazone compared with glimepiride on carotid intima-media thickness in type 2 diabetes: a randomized trial | Inappropriate study design |
| 7803 | The utility of traditional chinese medicine (SHENMAI) in the cardiac rehabilitation after coronary artery bypass grafting: a single-center randomized clinical trial | Inappropriate population |
| 7804 | Effect of plant sterols and endurance training on LDL particle size and distribution in previously sedentary hypercholesterolemic adults | Inappropriate study design |
| 7805 | Evaluation of myocardial perfusion and function by gated SPECT in patients with Behçet's disease | Inappropriate population |
| 7806 | Randomised double-blinded control adaptive trial to measure of the effectiveness of acipimox in patients with Mitochondrial Myopathy | Inappropriate study design |
| 7807 | Effect of muscle activity immediately after botulinum toxin injection for writer's cramp | Inappropriate study design |
| 7808 | The impact of a nurse-led care programme on events and physical and psychosocial parameters in patients with heart failure with preserved ejection fraction: a randomized clinical trial in primary care in Russia | Inappropriate population |
| 7809 | Comparison of adaptive pacing therapy, cognitive behaviour therapy, graded exercise therapy, and specialist medical care for chronic fatigue syndrome (PACE): a randomised trial | Inappropriate outcome |
| 7810 | The avian hippocampus is sensitive to chronic stress at the caudal pole | Inappropriate outcome |
| 7811 | Effects of trunk stabilization exercise while wearing a pelvic compression belt on walking and balancing abilities in patients whit stroke: an assessor blinded, preliminary, randomized, controlled study | Inappropriate study design |

| | | |
|---|---|---|
| 7812 | Maximizing upper-limb rehabilitation for chronic post-stroke patients using a remotely monitored virtual reality exergame: preliminary results of a randomized clinical trial | Inappropriate study design |
| 7813 | Cerebrolysin in post-stroke spasticity | Inappropriate study design |
| 7814 | Single and choice reaction time during prolonged exercise in trained subjects: influence of carbohydrate availability | Inappropriate population |
| 7815 | Home-based constraint-induced movement therapy for patients with upper limb dysfunction after stroke (HOMECIMT): a cluster-randomised, controlled trial | Inappropriate study design |
| 7816 | Testosterone dose-response relationships in healthy young men | Inappropriate study design |
| 7817 | Influence of body temperature on the development of fatigue during prolonged exercise in the heat | Inappropriate study design |
| 7818 | A theory-based online health behaviour intervention for new university students (U@Uni: lifeGuide): results from a repeat randomized controlled trial | Inappropriate study design |
| 7819 | Effects of tai chi on postural control during dual-task stair negotiation in knee osteoarthritis: a randomised controlled trial protocol | Inappropriate outcome |
| 7820 | Reboot Online: a Randomized Controlled Trial Comparing an Online Multidisciplinary Pain Management Program with Usual Care for Chronic Pain | Inappropriate outcome |
| 7821 | Immediate effects of speed-dependent treadmill training on gait parameters in early Parkinson's disease | Inappropriate study design |
| 7822 | Methylphenidate modified release as treatment of MS-associated fatigue | Inappropriate study design |
| 7823 | Therapeutic yoga improves balance and balance confidence in Parkinson's disease | Inappropriate study design |
| 7824 | Long-term effect of epoetin alfa on clinical and biochemical markers in friedreich ataxia | Inappropriate study design |
| 7825 | Randomized controlled trial of abdominal vibration stimulation and walking exercise for bowel cleansing prior to colonoscopy | Inappropriate study design |
| 7826 | Effects of home exercises on motor performance in patients with Parkinson's disease | Inappropriate study design |
| 7827 | A randomized controlled feasibility study comparing a multimodal prehabilitation protocol to normal care for women undergoing neo-adjuvant chemotherapy for breast cancer | Inappropriate study design |
| 7828 | Effects of a core stabilization training program on balance ability in persons with Parkinson's disease: a randomized controlled trial | Inappropriate study design |
| 7829 | Effects of curved-walking training on curved-walking performance and freezing of gait in individuals with Parkinson's disease: a randomized controlled trial | Inappropriate study design |
| 7830 | Effects of interactive video-game-based exercise on balance in older adults with mild-to-moderate Parkinson's disease | Inappropriate study design |
| 7831 | Do people with acquired brain impairment benefit from additional therapy specifically directed at the hand? A randomized controlled trial | Inappropriate study design |
| 7832 | The ENIGMA-II Trial. Nitrous oxide anaesthesia and cardiac morbidity after major surgery: a randomised controlled trial - Nitrous oxide anaesthesia and cardiac morbidity after major surgery | Inappropriate population |
| 7833 | Bilateral coordination and gait symmetry after body-weight supported treadmill training for persons with chronic stroke | Inappropriate study design |
| 7834 | Walking economy during cued versus non-cued treadmill walking in persons with Parkinson's disease | Inappropriate study design |
| 7835 | Telerehabilitation in multiple sclerosis: results of a randomized, 3-arm, rater blinded, feasibility and efficacy pilot study; patient-reported outcomes report | Inappropriate study design |
| 7836 | The effects of mindfulness-based yogic breathing on craving, affect, and smoking behavior | Inappropriate population |
| 7837 | Effect of Alternate Nostril Breathing Exercise on Experimentally Induced Anxiety in Healthy Volunteers Using the Simulated Public Speaking Model: a Randomized Controlled Pilot Study | Inappropriate study design |
| 7838 | Effect of early exercise therapy on the recovery of motor function in patients with cerebral infarction and the changes of somatosensory evoked potential | Inappropriate study design |
| 7839 | Video Game Rehabilitation for Outpatient Stroke (VIGoROUS): protocol for a multi-center comparative effectiveness trial of in-home gamified constraint-induced movement therapy for rehabilitation of chronic upper extremity hemiparesis | Inappropriate population |
| 7840 | Gait training induced change in corticomotor excitability in patients with chronic stroke | Inappropriate population |
| 7841 | The effect of a stretching device on hand spasticity in chronic hemiparetic stroke patients | Inappropriate study design |

| | | |
|---|---|---|
| 7842 | Robotic therapy provides a stimulus for upper limb motor recovery after stroke that is complementary to and distinct from conventional therapy | Inappropriate population |
| 7843 | Stroke volume decline during prolonged exercise is influenced by the increase in heart rate | Inappropriate population |
| 7844 | Improving Blood Pressure in Colorado | Inappropriate study design |
| 7845 | Use of exercise Doppler echocardiography to evaluate haemodynamic benefits of rate responsive pacing | Inappropriate population |
| 7846 | Daily exercise facilitates phase delays of circadian melatonin rhythm in very dim light | Inappropriate study design |
| 7847 | The effect of combined somatosensory stimulation and task-specific training on upper limb function in chronic stroke: a double-blind randomized controlled trial | Inappropriate study design |
| 7848 | Balance performance with a cognitive task: a dual-task testing paradigm | Inappropriate study design |
| 7849 | Cognitive Mediators of Change in Physical Functioning in Response to a Multifaceted Intervention for Managing Osteoarthritis | Inappropriate population |
| 7850 | Trunk muscle strength, cross-sectional area, and density in patients with chronic low back pain randomized to lumbar fusion or cognitive intervention and exercises | Inappropriate study design |
| 7851 | Delivering a back school programme with a cognitive behavioural modification: a randomised pilot trial on patients with chronic non-specific low back pain and functional disability | Inappropriate population |
| 7852 | Evaluating the effectiveness of a return to work programme for adults living in Waikato receiving a 'sickness' benefit | Inappropriate study design |
| 7853 | Dipyridamole in chronic stable angina pectoris; a randomized, double blind, placebo-controlled, parallel group study | Inappropriate study design |
| 7854 | How can cognitive remediation therapy modulate brain activations in schizophrenia? An fMRI study | Inappropriate study design |
| 7855 | Cognition and HPA axis reactivity in mildly to moderately depressed outpatients. A case-control study | Inappropriate study design |
| 7856 | EVALUATION OF THE REGULATORY-ADAPTIVE STATUS FOR PROGNOSTICATION IN SYSTOLIC CHRONIC HEART FAILURE | Inappropriate outcome |
| 7857 | Effect of prazosin vs placebo on chronic left ventricular heart failure | Inappropriate population |
| 7858 | Neurocognitive improvements after best-practice intervention for chronic fatigue syndrome: preliminary evidence of divergence between objective indices and subjective perceptions | Inappropriate study design |
| 7859 | A Community-Based, Bionic Leg Rehabilitation Program for Patients with Chronic Stroke: clinical Trial Protocol | Inappropriate study design |
| 7860 | Effects of long-term treatment with calcium antagonists on left ventricular diastolic function in stable angina and heart failure | Inappropriate study design |
| 7861 | Four-year follow-up of surgical versus non-surgical therapy for chronic low back pain | Inappropriate study design |
| 7862 | Using the wolf motor test to quantify stroke upper extremity function on an inpatient stroke unit can be problematic-a randomized controlled trial | Inappropriate study design |
| 7863 | Virtual reality exercise improves mobility after stroke: an inpatient randomized controlled trial | Inappropriate study design |
| 7864 | Qigong could be an effective and alternative complementary management for post-stroke rehabilitation: preliminary results | Inappropriate intervention |
| 7865 | Specific trunk and general exercise elicit similar changes in anticipatory postural adjustments in patients with chronic low back pain: a randomized controlled trial | Inappropriate population |
| 7866 | ENGAGE for Brain Health | Inappropriate study design |
| 7867 | A Comparative Study of Conventional Physiotherapy versus Robot-Assisted Gait Training Associated to Physiotherapy in Individuals with Ataxia after Stroke | Inappropriate study design |
| 7868 | Long term inotropic effects of flecainide and propafenone | Inappropriate study design |
| 7869 | Effectiveness of myofunctional exercises associated with vocal exercises in the rehabilitation of neurogenic oropharyngeal dysphagia: a randomized double blind clinical trial | Inappropriate study design |
| 7870 | Comparison of the subacute hemodynamic effects of atenolol, propranolol, pindolol, and nebivolol | Inappropriate population |
| 7871 | Physical work capacity with rate responsive ventricular pacing (VVIR) versus dual chamber pacing (DDD) in patients with normal and diminished left ventricular function | Inappropriate study design |
| 7872 | The Reconditioning Exercise and Chronic Obstructive Pulmonary Disease Trial II (REACT II): rationale and study design for a clinical trial of physical activity among individuals with chronic obstructive pulmonary disease | Inappropriate population |

| | | |
|---|---|---|
| 7873 | Augmented community telerehabilitation intervention to improve outcomes for people with stroke AKTIV-a randomised controlled trial | Inappropriate outcome |
| 7874 | A novel neurocognitive rehabilitation tool in the recovery of hemiplegic hand grip after stroke: a case report | Inappropriate study design |
| 7875 | Internet-based Treatment for Adults With Autism Spectrum Disorder (iALMA) | Inappropriate study design |
| 7876 | Effects of Training in a Virtual Environment in Healthy Elderly | Inappropriate study design |
| 7877 | Health-related quality of life in patients with chronic fatigue syndrome: group cognitive behavioural therapy and graded exercise versus usual treatment. A randomised controlled trial with 1 year of follow-up | Inappropriate population |
| 7878 | Does a standardised exercise protocol incorporating a cognitive task provoke postconcussion-like symptoms in healthy individuals? | Inappropriate study design |
| 7879 | The Effects of Exercise with Cycleergometer in Patients after Acute Stroke | Inappropriate study design |
| 7880 | Effect of motor control exercises versus graded activity in patients with chronic nonspecific low back pain: a randomized controlled trial | Inappropriate study design |
| 7881 | Effects on presenteeism and absenteeism from a 1-year workplace randomized controlled trial among health care workers | Inappropriate study design |
| 7882 | A theory-based exercise intervention in patients with heart failure: a protocol for randomized, controlled trial | Inappropriate outcome |
| 7883 | Older adults with chronic disease: benefits of group-mediated counseling in the promotion of physically active lifestyles | Inappropriate outcome |
| 7884 | Rethinking the treatment of chronic fatigue syndrome-a reanalysis and evaluation of findings from a recent major trial of graded exercise and CBT | Inappropriate study design |
| 7885 | Eliciting upper extremity purposeful movements using video games: a comparison with traditional therapy for stroke rehabilitation | Inappropriate study design |
| 7886 | Walking and balance ability gain from two types of gait intervention in adult patients with chronic hemiplegic stroke: a pilot study | Inappropriate population |
| 7887 | Clinical effectiveness of transcranial direct current stimulation and virtual reality on chronic individuals post-stroke with severe hemiparesis | Inappropriate study design |
| 7888 | Internet-based cognitive behavioural therapy (FITNET) is an effective treatment for adolescents with chronic fatigue syndrome | Inappropriate study design |
| 7889 | Tailored cognitive-behavioral therapy plus exercise training improved clinical and functional outcomes in fibromyalgia | Inappropriate study design |
| 7890 | Effects of joint mobilization and stretching on the range of motion for ankle joint and spatiotemporal gait variables in stroke patients | Inappropriate population |
| 7891 | Feasibility of post-stroke upper limb rehabilitation using embodied virtual reality | Inappropriate study design |
| 7892 | Effectiveness of a brief intervention using mental simulations in reducing alcohol consumption in corporate employees | Inappropriate study design |
| 7893 | Effect of a novel two-desk sit-to-stand workplace (ACTIVE OFFICE) on sitting time, performance and physiological parameters: protocol for a randomized control trial | Inappropriate population |
| 7894 | Improving insight and non-insight problem solving with brief interventions | Inappropriate study design |
| 7895 | Weight loss among female health care workers--a 1-year workplace based randomized controlled trial in the FINALE-health study | Inappropriate study design |
| 7896 | A randomized controlled evaluation of a psychosocial intervention in adults with chronic lung disease | Inappropriate study design |
| 7897 | Pain, coping, emotional state and physical function in patients with chronic radicular neck pain. A comparison between patients treated with surgery, physiotherapy or neck collar--a blinded, prospective randomized study | Inappropriate study design |
| 7898 | A randomized controlled graded exercise trial for chronic fatigue syndrome: outcomes and mechanisms of change | Inappropriate study design |
| 7899 | The effect of X box 360 kinect-virtual reality intervention on balance and gait training in stroke patient": an interventional study | Inappropriate study design |
| 7900 | Healthy living partnerships to prevent diabetes (help PD): a randomized controlled trialto prevent diabetes through diet and exercise: 2 year effects on the metabolic syndrome | Inappropriate study design |
| 7901 | Mental contrasting with implementation intentions (MCII) improves physical activity and weight loss among stroke survivors over one year | Inappropriate study design |
| 7902 | Feasibility, Safety and Efficacy of a Virtual Reality Exergame System to Supplement Upper Extremity Rehabilitation Post-Stroke: a Pilot Randomized Clinical Trial and Proof of Principle | Inappropriate study design |
| 7903 | The effects of Bobath-based trunk exercises on trunk control, functional capacity, balance, and gait: a pilot randomized controlled trial | Inappropriate population |
| 7904 | Camera-Based Mirror Visual Input for Priming Promotes Motor Recovery, Daily Function, and Brain Network Segregation in Subacute Stroke Patients | Inappropriate study design |

| | | |
|---|---|---|
| 7905 | Exercise therapy for arm function in stroke patients: a systematic review of randomized controlled trials | Inappropriate study design |
| 7906 | Effects of 8-week sensory electrical stimulation combined with motor training on EEG-EMG coherence and motor function in individuals with stroke | Inappropriate study design |
| 7907 | Transient impact of prolonged versus repetitive stretch on hand motor control in chronic stroke | Inappropriate study design |
| 7908 | Rehabilitative treatments for chronic fatigue syndrome: long-term follow-up from the PACE trial | Inappropriate outcome |
| 7909 | Self help approach to graded exercise may help chronic fatigue syndrome | Inappropriate study design |
| 7910 | Advances in Group-based Falls Rehabilitation | Inappropriate study design |
| 7911 | Feldenkrais method-based exercise improves quality of life in individuals with Parkinson's disease: a controlled, randomized clinical trial | Inappropriate study design |
| 7912 | The effects of a mindfulness-based lifestyle programme for adults with Parkinson's disease: protocol for a mixed methods, randomised two-group control study | Inappropriate outcome |
| 7913 | "Worth the Walk": culturally Tailored Stroke Risk Factor Reduction Intervention in Community Senior Centers | Inappropriate study design |
| 7914 | Assessment of the Efficacy of EEG-Based MI-BCI with Visual Feedback and EEG Correlates of Mental Fatigue for Upper-Limb Stroke Rehabilitation | Inappropriate population |
| 7915 | Effects of mindfulness based cognitive therapy on disability, thickness of the lumbar multifidus muscle, fear avoidance beliefs and metacognitive beliefs in the subjects with non-specific low back pain | Inappropriate population |
| 7916 | Enhanced motivational interviewing for reducing weight and increasing physical activity in adults with high cardiovascular risk: the MOVE IT three-arm RCT | Inappropriate study design |
| 7917 | Feasibility study of dual-task-managing training to improve gait performance of older adults | Inappropriate outcome |
| 7918 | Balance training with multi-task exercises improves fall-related self-efficacy, gait, balance performance and physical function in older adults with osteoporosis: a randomized controlled trial | Inappropriate study design |
| 7919 | Basal and dynamic relationships between implicit power motivation and estradiol in women | Inappropriate study design |
| 7920 | Enhancing Cardiac Rehabilitation With Stress Management Training: a Randomized, Clinical Efficacy Trial | Inappropriate outcome |
| 7921 | The effect of strengthening exercises on exaggerated muscle tonicity in chronic hemiparesis following stroke | Inappropriate population |
| 7922 | Comparative Assessment of Two Robot-Assisted Therapies for the Upper Extremity in People With Chronic Stroke | Inappropriate study design |
| 7923 | Biofeedback rehabilitation of posture and weightbearing distribution in stroke: a center of foot pressure analysis | Inappropriate study design |
| 7924 | Effects of task oriented exercises with altered sensory input on balance and functional mobility in chronic stroke: a pilot randomized controlled trial | Inappropriate population |
| 7925 | Effects of robot-assisted upper limb rehabilitation on daily function and real-world arm activity in patients with chronic stroke: a randomized controlled trial | Inappropriate population |
| 7926 | Short- and long-term effects of an inhibitor hand splint in poststroke patients: a randomized controlled trial | Inappropriate study design |
| 7927 | Neurocognitive performance and physical function do not change with physical-cognitive-mindfulness training in female laboratory technicians with chronic musculoskeletal pain: randomized controlled trial | Inappropriate population |
| 7928 | The Efficacy and Safety of Xinmailong Injection in Patients with Chronic Heart Failure: a Multicenter Randomized Double-Blind Placebo-Controlled Trial | Inappropriate study design |
| 7929 | The effect of multiple micronutrient supplementation on left ventricular ejection fraction in patients with chronic stable heart failure: a randomized, placebo-controlled trial | Inappropriate study design |
| 7930 | The effects of exercise on cardiovascular biomarkers in patients with chronic heart failure | Inappropriate population |
| 7931 | Study on propionyl-L-carnitine in chronic heart failure | Inappropriate population |
| 7932 | Addition of candesartan to angiotensin converting enzyme inhibitor therapy in patients with chronic heart failure does not reduce levels of oxidative stress | Inappropriate study design |
| 7933 | Is beta-blockade useful in heart failure patients with atrial fibrillation? An analysis of data from two previously completed prospective trials | Inappropriate study design |
| 7934 | Elamipretide Prevents the Natural History Progression of Cardiomyopathy-Related Disease Symptomatology in Patients with Barth Syndrome: a Tazpower Analysis | Inappropriate study design |
| 7935 | Balance outcomes after additional sit-to-stand training in subjects with stroke: a randomized controlled trial | Inappropriate study design |

| | | |
|---|---|---|
| 7936 | Effect of dual-task exercise in conjunction with fluoxetine & transcranial direct current stimulation on postural stability and gait in stroke patients | Inappropriate outcome |
| 7937 | Practice Structure and Locomotor Learning After Stroke | Inappropriate population |
| 7938 | Group-based multimodal exercises integrated with cognitive-behavioural therapy improve disability, pain and quality of life of subjects with chronic neck pain: a randomized controlled trial with one-year follow-up | Inappropriate study design |
| 7939 | Depression Outcomes Study of Exercise | Inappropriate study design |
| 7940 | Predictors of change in trunk muscle strength for patients with chronic low back pain randomized to lumbar fusion or cognitive intervention and exercises | Inappropriate study design |
| 7941 | Rehabilitative treatment for knee osteoarthritis in 28 hemiplegic patients after stroke | Inappropriate study design |
| 7942 | The role of technology in task-oriented training in persons with subacute stroke: a randomized controlled trial | Inappropriate study design |
| 7943 | Feasibility of a physiotherapist-led community-based self-management programme for reducing non-specific chronic low back pain disability in rural Nigeria | Inappropriate study design |
| 7944 | Effects of treadmill training with partial body weight support and the proprioceptive neuromuscular facilitation method on hemiparetic gait: a randomized controlled study | Inappropriate study design |
| 7945 | Comparison of dual task specific training and conventional physical therapy in ambulation of hemiplegic stroke patients: a randomized controlled trial | Inappropriate population |
| 7946 | Beta-alanine supplementation in patients with COPD receiving non-linear periodised exercise training or neuromuscular electrical stimulation: protocol of two randomised, double-blind, placebo-controlled trials | Inappropriate study design |
| 7947 | Gait training in subacute non-ambulatory stroke patients using a full weight-bearing gait-assistance robot: a prospective, randomized, open, blinded-endpoint trial | Inappropriate outcome |
| 7948 | Reducing Trunk Compensation in Stroke Survivors: a Randomized Crossover Trial Comparing Visual and Force Feedback Modalities | Inappropriate study design |
| 7949 | Essential amino acid supplementation in patients with severe COPD: a step towards home rehabilitation | Inappropriate study design |
| 7950 | Efficacy of Behavioural Graded Activity Compared with Conventional Exercise Therapy in Chronic Non-Specific Low Back Pain: implication for Direct Health Care Cost | Inappropriate population |
| 7951 | Return to work after comparing outpatient multidisciplinary treatment programs versus treatment in general practice for patients with chronic widespread pain | Inappropriate study design |
| 7952 | Daily factors driving daily substance use and chronic pain among older adults with HIV: an exploratory study using ecological momentary assessment | Inappropriate population |
| 7953 | Clinical Outcomes Following Language-Specific Attention Treatment Versus Direct Attention Training for Aphasia: a Comparative Effectiveness Study | Inappropriate outcome |
| 7954 | Effects of repetitive trascranial magnetic stimulation on repetitive facilitation exercises of the hemiplegic hand in chronic stroke patients | Inappropriate study design |
| 7955 | Combined therapeutic effect of probucol and cilostazol on endothelial function in patients with silent cerebral lacunar infarcts and hypercholesterolemia: a preliminary study | Inappropriate outcome |
| 7956 | Neuromuscular Electrical Stimulation in Acute Ischemic Stroke | Inappropriate population |
| 7957 | No difference in 9-year outcome in CLBP patients randomized to lumbar fusion versus cognitive intervention and exercises | Inappropriate study design |
| 7958 | OUTCOMES OF ERCP IN PATIENTS WITH ACUTE STROKE: a NATIONAL STUDY | Inappropriate population |
| 7959 | Correction to: why all randomised controlled trials produce biased results (Annals of Medicine, (2018), 50, 4, (312-322), 10.1080/07853890.2018.1453233) | Inappropriate outcome |
| 7960 | Cardiovascular effects of tricyclic antidepressants in depressed patients with chronic heart disease | Inappropriate outcome |
| 7961 | A dose response study with oral prenalterol in patients with chronic congestive cardiac failure | Inappropriate study design |
| 7962 | A role of activation of proinflammatory cytokines and production of autoimmune complexes in the pathogenesis of chronic heart failure in patients with postinfarct cardiac dysfunction | Inappropriate outcome |
| 7963 | Phase II Randomized Controlled Trial of Constraint-Induced Movement Therapy in Multiple Sclerosis. Part 2: effect on White Matter Integrity | Inappropriate study design |
| 7964 | Effects of Virtual Reality Training using Xbox Kinect on Motor Function in Stroke Survivors: a Preliminary Study | Inappropriate outcome |

| | | |
|---|---|---|
| 7965 | Effects of the K+ channel blocker tedisamil on hemodynamics, myocardial ischemia and neurohumoral system in patients with stable angina pectoris. A comparison with the beta blocker atenolol | Inappropriate study design |
| 7966 | Cathodal transcranial direct current stimulation of the primary motor cortex improves selective muscle activation in the ipsilateral arm | Inappropriate population |
| 7967 | Intrathecal fentanyl blockade of afferent neural feedback from skeletal muscle during exercise in heart failure patients: influence on circulatory power and pulmonary vascular capacitance | Inappropriate study design |
| 7968 | This is a single center study wherein the efficacy of SabeetÂ® in cardiovascular and exercise performance will be studied in 20 adult male healthy volunteers in comparison to Placebo using a crossover design | Inappropriate population |
| 7969 | Treatment of severe hand impairment following stroke by combining assisted movement, muscle vibration, and biofeedback | Inappropriate population |
| 7970 | The effects of functional action-observation training on gait function in patients with post-stroke hemiparesis: a randomized controlled trial | Inappropriate study design |
| 7971 | Treadmill training post stroke: are there any secondary benefits? A pilot study | Inappropriate study design |
| 7972 | Bicycle Simulator Training in Older Adults: a Randomized Controlled Trial | Inappropriate study design |
| 7973 | Healthy youth places promoting nutrition and physical activity | Inappropriate study design |
| 7974 | No difference in long-term trunk muscle strength, cross-sectional area, and density in patients with chronic low back pain 7 to 11 years after lumbar fusion versus cognitive intervention and exercises | Inappropriate study design |
| 7975 | A health promotion study for the effectiveness to strengthen mobility among users in residential homes in the framework of a pilot project for setting-oriented health promotion in residential long term care | Inappropriate study design |
| 7976 | Effect of biofeedback cycling training on functional recovery and walking ability of lower extremity in patients with stroke | Inappropriate study design |
| 7977 | Comparison of different methods of functional evaluation in patients with chronic heart failure | Inappropriate population |
| 7978 | Hawthorn Extract Randomized Blinded Chronic Heart Failure (HERB CHF) trial | Inappropriate study design |
| 7979 | A comparison of enalapril with hydralazine-isosorbide dinitrate in the treatment of chronic congestive heart failure | Inappropriate study design |
| 7980 | Controlled release metoprolol for aortic regurgitation: a randomised clinical trial | Inappropriate outcome |
| 7981 | Hydralazine in the long-term treatment of chronic heart failure: lack of difference from placebo | Inappropriate study design |
| 7982 | Hypertension Improvement Project (HIP): study protocol and implementation challenges | Inappropriate study design |
| 7983 | Effects of exercise training in patients with chronic heart failure and advanced left ventricular systolic dysfunction receiving β-blockers | Inappropriate study design |
| 7984 | Improved left ventricular performance during exercise with verapamil or nifedipine in patients with chronic stable angina | Inappropriate study design |
| 7985 | Influence of the beta-blocker nebivolol on left ventricular function in patients with chronic heart failure | Inappropriate study design |
| 7986 | A multicentre, double-blind, placebo-controlled trial of quinapril in mild, chronic heart failure | Inappropriate study design |
| 7987 | Functional electrical stimulation improves endothelial function and reduces peripheral immune responses in patients with chronic heart failure | Inappropriate population |
| 7988 | Effect of Rhythm-based Multitask Training on Falls Among Older Adults | Inappropriate population |
| 7989 | Does dual task placement and duration affect split-belt treadmill adaptation? | Inappropriate intervention |
| 7990 | Cost-effectiveness of cognitive behavioural therapy, graded exercise and usual care for patients with chronic fatigue in primary care | Inappropriate study design |
| 7991 | Chronic neck pain and treatment of cognitive and behavioural factors: results of a randomised controlled clinical trial | Inappropriate study design |
| 7992 | Effect of a long-lasting multidisciplinary program on disability and fear-avoidance behaviors in patients with chronic low back pain: results of a randomized controlled trial | Inappropriate study design |
| 7993 | Potential mechanisms involved in the effect of cognitive behavioral therapy on fatigue severity in Type 1 diabetes | Inappropriate population |
| 7994 | The effect of caffeine on cognitive performance is influenced by CYP1A2 but not ADORA2A genotype, yet neither genotype affects exercise performance in healthy adults | Inappropriate population |
| 7995 | Randomized Evaluation of Cognitive-Behavioral Therapy and Graded Exercise Therapy for Post-Cancer Fatigue | Inappropriate study design |

| | | |
|---|---|---|
| 7996 | Differential Effects of Physical Exercise, Cognitive Training, and Mindfulness Practice on Serum BDNF Levels in Healthy Older Adults: a Randomized Controlled Intervention Study | Inappropriate study design |
| 7997 | Gait outcomes after acute stroke rehabilitation with supported treadmill ambulation training: a randomized controlled pilot study | Inappropriate population |
| 7998 | Effects of robot-assisted gait training on spatiotemporal gait parameters and balance in patients with chronic stroke: a randomized controlled pilot trial | Inappropriate study design |
| 7999 | Bilateral upper limb training with functional electric stimulation in patients with chronic stroke | Inappropriate population |
| 8000 | Activity-sensing rate responsive versus conventional fixed-rate pacing: a comparison of rate behavior and patient well-being during routine daily exercise | Inappropriate study design |
| 8001 | Effects on abnormal gait pattern in hemiparetic patients using ankle foot orthoses | Inappropriate population |
| 8002 | Verification of the left ventricular ejection fraction from gated myocardial perfusion studies (GSPECT) | Inappropriate study design |
| 8003 | The Post-Myocardial Infarction Pacing Remodeling Prevention Therapy (PRomPT) Trial: design and Rationale | Inappropriate population |
| 8004 | Assessment of diastolic function using 16-frame 99mTc-sestamibi gated myocardial perfusion SPECT: normal values | Inappropriate population |
| 8005 | Multiparametric comparison of CARvedilol, vs. NEbivolol, vs. BIsoprolol in moderate heart failure: the CARNEBI trial | Inappropriate study design |
| 8006 | Left Ventricular Myocardial and Hemodynamic Response to Exercise in Young Patients after Endovascular Stenting for Aortic Coarctation | Inappropriate study design |
| 8007 | Simulated laughter, perceived stress, and discourse in adults with aphasia | Inappropriate intervention |
| 8008 | Home-based telemonitored Nordic walking training is well accepted, safe, effective and has high adherence among heart failure patients, including those with cardiovascular implantable electronic devices: a randomised controlled study | Inappropriate population |
| 8009 | Effect of ventricular function on the exercise hemodynamics of variable rate pacing | Inappropriate study design |
| 8010 | The Mind Body-Wellness in Supportive Housing (Mi-WiSH) study: design and rationale of a cluster randomized controlled trial of Tai Chi in senior housing | Inappropriate study design |
| 8011 | To Compare the Closed Kinetic Chain Exercises and Open Kinetic Chain Exercises on Gait Velocity, Cadence, Stride Length and Strength of Knee Flexors and Extensors in Chronic Stroke Patients | Inappropriate study design |
| 8012 | Heart rate variability biofeedback therapy and graded exercise training in management of chronic fatigue syndrome: an exploratory pilot study | Inappropriate population |
| 8013 | Peer-Delivered Cognitive Behavioral Training to Improve Functioning in Patients With Diabetes: a Cluster-Randomized Trial | Inappropriate study design |
| 8014 | A multimodal intervention approach for cross-sectoral care of degenerative diseases of the spine | Inappropriate population |
| 8015 | Cognitive behavior therapy, exercise, or both for treating chronic widespread pain | Inappropriate study design |
| 8016 | A multidisciplinary rehabilitation programme improves disability, kinesiophobia and walking ability in subjects with chronic low back pain: results of a randomised controlled pilot study | Inappropriate population |
| 8017 | Tailored cognitive-behavioral therapy and exercise training for high-risk patients with fibromyalgia | Inappropriate population |
| 8018 | Psychometric properties of the Multidimensional Pain Inventory, Dutch language version (MPI-DLV) | Inappropriate study design |
| 8019 | Comparing mind-body wellness interventions for older adults with chronic illness: classroom versus home instruction | Inappropriate study design |
| 8020 | A Cluster Randomized Trial of Tai Chi vs Health Education in Subsidized Housing: the MI-WiSH Study | Inappropriate population |
| 8021 | Trunk muscle stabilization training plus general exercise versus general exercise only: randomized controlled trial of patients with recurrent low back pain | Inappropriate study design |
| 8022 | Incorporating Behavioral Trigger Messages Into a Mobile Health App for Chronic Disease Management: randomized Clinical Feasibility Trial in Diabetes | Inappropriate study design |
| 8023 | Investigating generalizability of results from a randomized controlled trial of the management of chronic widespread pain: the MUSICIAN study | Inappropriate population |
| 8024 | Recovery from chronic fatigue syndrome after treatments given in the PACE trial | Inappropriate outcome |
| 8025 | Motor Performance in Chronic Stroke Patients | Inappropriate study design |
| 8026 | Effect of dual-task functional power and mobility training on falls and physical function in older people living in retirement villages: a cluster randomised controlled trial | Inappropriate study design |

| | | |
|---|---|---|
| 8027 | An online physical activity program for German and Dutch adults delivered by SMS | Inappropriate study design |
| 8028 | Precooling's Effect on American Football Skills | Inappropriate study design |
| 8029 | The American football uniform: uncompensable heat stress and hyperthermic exhaustion | Inappropriate population |
| 8030 | Counselling for physical activity, life-space mobility and falls prevention in old age (COSMOS): protocol of a randomised controlled trial | Inappropriate population |
| 8031 | Efficacy and Safety of the Physical Activity Program in Women With Post-menopausal Osteoporosis | Inappropriate study design |
| 8032 | The effect of circuit training in a modified constraint induced movement therapy program | Inappropriate population |
| 8033 | FOOD INTAKE RESTRICTION FOR HEALTH OUTCOME SUPPORT AND EDUCATION (FIREHOUSE) TRIAL: STUDY DESIGN | Inappropriate population |
| 8034 | Comparing the effectiveness of an enhanced MOtiVational intErviewing InTervention (MOVE IT) with usual care for reducing cardiovascular risk in high risk subjects: study protocol for a randomised controlled trial | Inappropriate study design |
| 8035 | Rationale, design, and baseline data for the Healthy Mom II Trial: a randomized trial examining the efficacy of exercise and wellness interventions for the prevention of postpartum depression | Inappropriate study design |
| 8036 | Efficacy of a behavior change program on cardiovascular parameters in patients with hypertension: a randomized controlled trial | Inappropriate study design |
| 8037 | Intravenous iron supplement for patients with severe aortic stenosis who have iron deficiency | Inappropriate study design |
| 8038 | Randomised controlled pilot trial of an exercise plus behaviour change intervention in people with multiple sclerosis: the Step it Up study | Inappropriate study design |
| 8039 | Influence of paravertebral muscles training on brain plasticity and postural control in chronic low back pain | Inappropriate study design |
| 8040 | Efficacy of GP referral of insufficiently active patients for expert physical activity counseling: protocol for a pragmatic randomized trial (The NewCOACH trial) | Inappropriate intervention |
| 8041 | A randomized control trial (RCT) of an online, symptom self-management curriculum (Reimagine) among breast cancer survivors | Inappropriate population |
| 8042 | Does adapted physical activity-based rehabilitation improve mental and physical functioning? A randomized trial | Inappropriate population |
| 8043 | Physiotherapy informed by Acceptance and Commitment Therapy (PACT) for people with chronic low back pain: a randomised controlled trial | Inappropriate study design |
| 8044 | The efficacy of a perceptive rehabilitation on postural control in patients with chronic nonspecific low back pain | Inappropriate population |
| 8045 | The effects of the three-dimensional active trunk training exercise on trunk control ability, trunk muscle strength, and balance ability in sub-acute stroke patients: a randomized controlled pilot study | Inappropriate study design |
| 8046 | Repetitive reaching training combined with transcranial Random Noise Stimulation in stroke survivors with chronic and severe arm paresis is feasible: a pilot, triple-blind, randomised case series | Inappropriate study design |
| 8047 | Total disc replacement versus multidisciplinary rehabilitation in patients with chronic low back pain and degenerative discs: 8-year follow-up of a randomized controlled multicenter trial | Inappropriate study design |
| 8048 | A randomized controlled clinical trial of the Seattle Protocol for Activity in older adults | Inappropriate outcome |
| 8049 | Anxiety, depressive symptoms, and cardiac autonomic function in perimenopausal and postmenopausal women with hot flashes: a brief report | Inappropriate population |
| 8050 | A qualitative study of older adults' perspectives on initiating exercise and mindfulness practice | Inappropriate outcome |
| 8051 | Dance therapy combined with patient education improves quality of life of persons with obesity: a pilot feasibility study for a randomised controlled trial | Inappropriate population |
| 8052 | A Systematic Approach for Prescribing Posthospitalization Home-Based Physical Activity for Mobility in Older Adults: the PATH Study | Inappropriate population |
| 8053 | Behavioral weight loss and physical activity intervention in obese adults with asthma. A randomized trial | Inappropriate population |
| 8054 | Treadmill training with tilt sensor functional electrical stimulation for improving balance, gait, and muscle architecture of tibialis anterior of survivors with chronic stroke: a randomized controlled trial | Inappropriate study design |
| 8055 | Longer versus shorter mental practice sessions for affected upper extremity movement after stroke: a randomized controlled trial | Inappropriate outcome |
| 8056 | Step training with body weight support: effect of treadmill speed and practice paradigms on poststroke locomotor recovery | Inappropriate study design |

| | | |
|---|---|---|
| 8057 | Effects of innovative hip-knee-ankle interlimb coordinated robot training on ambulation, cardiopulmonary function, depression, and fall confidence in acute hemiplegia | Inappropriate population |
| 8058 | Automated Constraint-Induced Therapy Extension (AutoCITE) for movement deficits after stroke | Inappropriate study design |
| 8059 | What is the long-term benefit of constraint-induced movement therapy? A four-year follow-up | Inappropriate study design |
| 8060 | Blood pressure changes in patients with chronic heart failure undergoing slow breathing training | Inappropriate population |
| 8061 | Endurance exercise modulates levodopa induced growth hormone release in patients with Parkinson's disease | Inappropriate study design |
| 8062 | Effect of a specific training focusing on the trophy position to improve the tennis serve in children | Inappropriate population |
| 8063 | Influence of earlier rehabilitative interventions on the emotions and the ability of daily living of patients following the first onset of acute stroke | Inappropriate study design |
| 8064 | Improvement of visual search after audiovisual exploration training in hemianopic patients | Inappropriate population |
| 8065 | An 8 session exercise programme was effective for subacute or chronic low back pain: commentary | Inappropriate intervention |
| 8066 | Neurophysiological and behavioral effects of tDCS combined with constraint-induced movement therapy in poststroke patients | Inappropriate study design |
| 8067 | Effects of an aquatic therapy approach (Halliwick-Therapy) on functional mobility in subacute stroke patients: a randomized controlled trial | Inappropriate study design |
| 8068 | Group cognitive behavioral treatment improves chronic low back pain in a cost-effective manner | Inappropriate population |
| 8069 | Telephone-based cognitive-behavioural therapy and a structured exercise programme are effective for chronic widespread pain (fibromyalgia) | Inappropriate study design |
| 8070 | Effects of the Novel Long-Acting GLP-1 Agonist, Albiglutide, on Cardiac Function, Cardiac Metabolism, and Exercise Capacity in Patients With Chronic Heart Failure and Reduced Ejection Fraction | Inappropriate study design |
| 8071 | Effects of Tai Chi on Multisite Pain and Brain Functions in Older Adults | Inappropriate study design |
| 8072 | An investigation of the effects of scapular exercises on the body control in stroke patients: double-blind randomized controlled pilot study | Inappropriate study design |
| 8073 | Autologous stem cell transplantation in acute myocardial infarction: the ASTAMI randomized controlled trial. Intracoronary transplantation of autologous mononuclear bone marrow cells, study design and safety aspects | Inappropriate study design |
| 8074 | Functional outcome in acute stroke patients with oropharyngeal Dysphagia after swallowing therapy | Inappropriate study design |
| 8075 | Is the clinical presentation of chronic heart failure different in elderly versus younger patients and those with preserved versus reduced ejection fraction? | Inappropriate study design |
| 8076 | Effect of the calcium antagonist felodipine as supplementary vasodilator therapy in patients with chronic heart failure treated with enalapril: v-HeFT III. Vasodilator-Heart Failure Trial (V-HeFT) Study Group | Inappropriate study design |
| 8077 | Control of rate versus rhythm in rheumatic atrial fibrillation: a randomized study | Inappropriate study design |
| 8078 | Beneficial effects of exercise training in heart failure patients with low cardiac output response to exercise - a comparison of two training models | Inappropriate outcome |
| 8079 | Left ventricular apical thrombus and myocardial viability: a dobutamine stress echocardiographic study | Inappropriate study design |
| 8080 | Molecular basis for the improvement in muscle metaboreflex and mechanoreflex control in exercise-trained humans with chronic heart failure | Inappropriate outcome |
| 8081 | A Comparison of 16Weeks of Continuous vs Intermittent Exercise Training in Chronic Heart Failure Patients | Inappropriate study design |
| 8082 | Prospective, randomized comparison of effect of long-term treatment with metoprolol or carvedilol on symptoms, exercise, ejection fraction, and oxidative stress in heart failure | Inappropriate intervention |
| 8083 | Comparative effects of oral acebutolol and propranolol at rest and during exercise in ischemic heart disease: double-blind placebo crossover study utilizing radionuclide ventriculography | Inappropriate study design |
| 8084 | A failed improvement in pulmonary function and exercise capacity with carvedilol in congestive heart failure despite an excellent effect on left ventricular function | Inappropriate study design |
| 8085 | Variables Measured During Cardiopulmonary Exercise Testing as Predictors of Mortality in Chronic Systolic Heart Failure | Inappropriate population |
| 8086 | Effects of n-3 polyunsaturated fatty acids on left ventricular function and functional capacity in patients with dilated cardiomyopathy | Inappropriate study design |

| | | |
|---|---|---|
| 8087 | Effects of Kanlijian on exercise tolerance, quality of life, and frequency of heart failure aggravation in patients with chronic heart failure | Inappropriate population |
| 8088 | Rationale and design of the BUDAPEST-CRT Upgrade Study: a prospective, randomized, multicentre clinical trial | Inappropriate population |
| 8089 | Testosterone therapy in men with moderate severity heart failure: a double-blind randomized placebo controlled trial | Inappropriate study design |
| 8090 | Stepped care approach for medial tibiofemoral osteoarthritis (streamline) | Inappropriate study design |
| 8091 | The impact of specialized physiotherapy methods on BODE index in COPD patients during hospitalization | Inappropriate study design |
| 8092 | Long-term effect of early exercise engagement on clinical outcomes and cost implications, in patients with TIA and non-disabling stroke | Inappropriate study design |
| 8093 | Efficacy and safety of long-term application of spironolactone in patients with moderate and severe chronic heart failure receiving optimal therapy | Inappropriate study design |
| 8094 | Group-based task-oriented exercises aimed at managing kinesiophobia improved disability in chronic low back pain | Inappropriate study design |
| 8095 | Effect of a supplemental robotic therapeutic walking exercise in acute stroke rehabilitation | Inappropriate population |
| 8096 | Core-stability exercises improve gait pattern after stroke. A randomized control trial protocol | Inappropriate population |
| 8097 | Long-Term Effect of Participation in an Early Exercise and Education Program on Clinical Outcomes and Cost Implications, in Patients with TIA and Minor, Non-Disabling Stroke | Inappropriate study design |
| 8098 | A pilot study exploring the effects of a 12-week t'ai chi intervention on somatic symptoms of depression in patients with heart failure | Inappropriate study design |
| 8099 | Cognitive Behavioral Therapy With Stabilization Exercises Affects Transverse Abdominis Muscle Thickness in Patients With Chronic Low Back Pain: a Double-Blinded Randomized Trial Study | Inappropriate study design |
| 8100 | Project Osteoarthritis: recovering Quality of Life Through Education | Inappropriate study design |
| 8101 | The Acute Effect of Mental Fatigue on Badminton Performance in Elite Players | Inappropriate intervention |
| 8102 | Morning bright light treatment improves function and reduces pain sensitivity in fibromyalgia | Inappropriate study design |
| 8103 | Non-pharmacologic interventions for CFS: a randomized trial | Inappropriate study design |
| 8104 | Feasibility of a randomized controlled trial conducted in belo horizonte/brazilwith post-stroke individuals | Inappropriate population |
| 8105 | Effectiveness of adaptive physical activity combined with therapeutic patient education in stroke survivors at twelve months: a non-randomized parallel group study | Inappropriate study design |
| 8106 | Referral for Expert Physical Activity Counseling: a Pragmatic RCT | Inappropriate study design |
| 8107 | DO-HEALTH: vitamin D3 - Omega3 - Home exercise - Healthy aging and longevity trial - Design of a multinational clinical trial on healthy aging among European seniors | Inappropriate study design |
| 8108 | Development of a service delivery intervention and implementation plan for optimising primary care management of knee osteoarthritis: the partner project | Inappropriate study design |
| 8109 | Effectiveness of a multifactorial context-enhancing functional therapy to promote functional arm use and recovery of stroke survivors: study protocol for a clinical trial | Inappropriate population |
| 8110 | A Web-based health promotion program for older workers: randomized controlled trial | Inappropriate study design |
| 8111 | Evaluation of the behavioral program in diabetes mellitus type 2: randomized clinical trial study protocol | Inappropriate study design |
| 8112 | Modified constraint-induced therapy in acute stroke: a randomized controlled pilot study | Inappropriate study design |
| 8113 | Design of a randomized, placebo-controlled multicenter trial on the long-term effects of intermittent transdermal nitroglycerin on left ventricular remodeling after acute myocardial infarction. Transdermal Nitroglycerin Investigators Group | Inappropriate study design |
| 8114 | Influence of phosphatidylserine on cognitive performance and cortical activity after induced stress | Inappropriate population |
| 8115 | Respiratory muscles stretching acutely increases expansion in hemiparetic chest wall | Inappropriate study design |
| 8116 | Sensory retraining of the lower limb after acute stroke: a randomized controlled pilot trial | Inappropriate study design |
| 8117 | The cardiac insufficiency talinolol study (CITAS) study design | Inappropriate study design |
| 8118 | Effects of xamoterol in moderate heart failure. EFFETS DU XAMOTEROL DANS L'INSUFFISANCE CARDIAQUE MODEREE | Inappropriate population |

| | | |
|---|---|---|
| 8119 | Hand rehabilitation following stroke: a pilot study of assisted finger extension training in a virtual environment | Inappropriate study design |
| 8120 | The effect of aquatic therapy on postural balance and muscle strength in stroke survivors--a randomized controlled pilot trial | Inappropriate study design |
| 8121 | Effect of training eye hand coordination using emerging user interface technologies in stroke survivors a randomized controlled study | Inappropriate population |
| 8122 | Tele-rehabilitation service at home for patients after stroke | Inappropriate study design |
| 8123 | Wireless, accelerometry-triggered functional electrical stimulation of the peroneal nerve in spastic paresis: a randomized, controlled pilot study | Inappropriate study design |
| 8124 | Effects of atenolol as add-on therapy to fosinopril in heart failure | Inappropriate study design |
| 8125 | Enalapril in patients with chronic heart failure: a placebo-controlled, randomized, double-blind study | Inappropriate outcome |
| 8126 | Is digoxin really important in treatment of compensated heart failure? A placebo-controlled crossover study in patients with sinus rhythm | Inappropriate study design |
| 8127 | Cognitive-behavioural health-promotion intervention increases fruit and vegetable consumption and physical activity among South African adolescents: a cluster-randomised controlled trial | Inappropriate population |
| 8128 | Error-augmented bimanual therapy for stroke survivors | Inappropriate population |
| 8129 | Effect of mirror therapy combined with somatosensory stimulation on motor recovery and daily function in stroke patients: a pilot study | Inappropriate population |
| 8130 | Effects of repetitive arm cycling exercise on post-stroke upper limb function and satisfaction: a preliminary study | Inappropriate population |
| 8131 | Mindfulness Attitude to Deliver Dietary Approach to Stop Hypertension | Inappropriate study design |
| 8132 | Early mobilization in neurocritical care patients | Inappropriate study design |
| 8133 | Influence of rehabilitation exercise on physiological characteristics and heart function in patients with acute myocardial infarction | Inappropriate study design |
| 8134 | Effects of robot-assisted and conventional exercise on muscle synergies in stroke patients | Inappropriate intervention |
| 8135 | Retraining and assessing hand movement after stroke using the MusicGlove: comparison with conventional hand therapy and isometric grip training | Inappropriate study design |
| 8136 | Comparing integrated training of the hand and arm with isolated training of the same effectors in persons with stroke using haptically rendered virtual environments, a randomized clinical trial | Inappropriate study design |
| 8137 | Developing a suite of motion-controlled games for upper extremities training in patients with chronic stroke-a randomised control trial | Inappropriate study design |
| 8138 | Robotic Assistance for Training Finger Movement Using a Hebbian Model: a Randomized Controlled Trial | Inappropriate outcome |
| 8139 | Creatine loading does not impact on stroke performance in tennis | Inappropriate population |
| 8140 | Efficacy of a self-directed behavioral health change program: weight, body composition, cardiovascular fitness, blood pressure, health risk, and psychosocial mediating variables | Inappropriate study design |
| 8141 | Innovative program to increase physical activity following an acute coronary syndrome: randomized controlled trial | Inappropriate population |
| 8142 | The assessment of cardiac rehabilitation on echocardiographic parameters of left ventricular systolic function in patients treated by primary percutaneous coronary intervention due to acute ST-Segment elevation myocardial infarction: a randomized clinical trial | Inappropriate study design |
| 8143 | The effect of music-movement therapy on physical and psychological states of stroke patients | Inappropriate study design |
| 8144 | Prevention of Genu Recurvatum in Poststroke Patients Using a Hinged Soft Knee Orthosis | Inappropriate population |
| 8145 | Using the manumeter for continuous feedback on hand activity after stroke: preliminary results | Inappropriate population |
| 8146 | Behavioral Profile Matching: a Precision Medicine Approach to Concussion Rehabilitation | Inappropriate population |
| 8147 | Exercise and Weight Loss for Improving Mobility in Older Adults Who Are Obese | Inappropriate study design |
| 8148 | Improving Physical and Psychosocial Well-being of African American Older Adults | Inappropriate study design |
| 8149 | Project SHINE: effects of a randomized family-based health promotion program on the physical activity of African American parents | Inappropriate population |
| 8150 | Web-based Support to Manage Arthritis Pain | Inappropriate study design |
| 8151 | Exercise on Multiple Sclerosis Patients | Inappropriate study design |

| | | |
|---|---|---|
| 8152 | Effectiveness of a web-based self-help smoking cessation intervention: protocol of a randomised controlled trial | Inappropriate study design |
| 8153 | Effectiveness of mental skills training in the military context | Inappropriate study design |
| 8154 | Program to break up long bouts of sitting among male office workers at a public university in Saudi Arabia: SLIM trial | Inappropriate study design |
| 8155 | Practice makes progress: simulation improves neurology resident attitudes and self-efficacy with leading stroke codes | Inappropriate study design |
| 8156 | A Backward Walking Training Program to Improve Balance and Mobility in Acute Stroke: a Pilot Randomized Controlled Trial | Inappropriate study design |
| 8157 | Dobutamine-induced increase of right ventricular contractility without increased stroke volume in adolescent patients with transposition of the great arteries: evaluation with magnetic resonance imaging | Inappropriate population |
| 8158 | Efficacy of early intervention of motor relearning program on post-stroke hemiplemia: a randomized controlled observation | Inappropriate intervention |
| 8159 | The Effect of Vestibulo-ocular Reflex Improving Exercise on Gait and Balance Among Post Stroke Sub Acute Patients | Inappropriate study design |
| 8160 | Effectiveness of mid-thoracic spine mobilization versus therapeutic exercise in patients with subacute stroke: a randomized clinical trial | Inappropriate population |
| 8161 | The effects of METhotrexate therapy on the physical capacity of patients with ISchemic heart failure: a randomized double-blind, placebo-controlled trial (METIS trial) | Inappropriate population |
| 8162 | Effect of preventive messages tailored to family history on health behaviors: the Family Healthware Impact Trial | Inappropriate population |
| 8163 | Dimensionality and Item-Difficulty Hierarchy of the Lower Extremity Fugl-Meyer Assessment in Individuals with Subacute and Chronic Stroke Presented in part as a poster to the American Congress of Rehabilitation Medicine, November 13, 2013, Orlando, FL | Inappropriate outcome |
| 8164 | Effects of early rehabilitation on activities of daily living and complications in acute stroke patients | Inappropriate population |
| 8165 | Effect of spironolactone on ability to exercise and heart stiffness in people with irregular heart beats (atrial fibrillation) and normal pumping capacity of the heart | Inappropriate study design |
| 8166 | The comparison of two physiotherapeutic approaches for gait improvement in sub-acute stroke patients | Inappropriate study design |
| 8167 | The effects of clinical Pilates training on walking, balance, fall risk, respiratory and cognitive functions in persons with multiple sclerosis: a randomized controlled trial | Inappropriate population |
| 8168 | Multifactorial cardiac rehabilitation did not reduce mortality or morbidity after acute myocardial infarction | Inappropriate intervention |
| 8169 | The effect of chair-based pedal exercises for older people admitted to hospital: a feasibility study | Inappropriate study design |
| 8170 | Effects of early enteral nutrition on sub-acute ischemic stroke via gut microbiota reconstruction: a randomized controlled trial | Inappropriate study design |
| 8171 | Pindolol and propranolol in patients with angina pectoris and normal or near-normal ventricular function. Lack of influence of intrinsic sympathomimetic activity on global and segmental left ventricular function assessed by radionuclide ventriculography | Inappropriate intervention |
| 8172 | Three-month treatment with adaptive servoventilation improves cardiac function and physical activity in patients with chronic heart failure and cheyne-stokes respiration: a prospective randomized controlled trial | Inappropriate population |
| 8173 | Telehealth weight management intervention for adults with physical disabilities: a randomized controlled trial | Inappropriate outcome |
| 8174 | Reduced ventricular flow propagation velocity in elite athletes is augmented with the resumption of exercise training | Inappropriate intervention |
| 8175 | Improvement of postreceptor events by metoprolol treatment in patients with chronic heart failure | Inappropriate study design |
| 8176 | The effects and costs of home-based rehabilitation for heart failure with reduced ejection fraction: the REACH-HF multicentre randomized controlled trial | Inappropriate population |
| 8177 | Progression of mild untreated heart failure during six months follow-up and clinical and neurohumoral effects of ibopamine and digoxin as monotherapy. DIMT Study Group. Dutch Ibopamine Multicenter Trial | Inappropriate study design |
| 8178 | NORDIC WALKING AND STANDARD EXERCISE THERAPY IN PATIENTS WITH CHRONIC HEART FAILURE: a RANDOMIZED, CONTROLLED-TRIAL COMPARISON | Inappropriate population |
| 8179 | Radiofrequency ablation for persistent atrial fibrillation in patients with advanced heart failure and severe left ventricular systolic dysfunction: a randomised controlled trial | Inappropriate study design |

| | | |
|---|---|---|
| 8180 | Impact of Hemodynamic Ramp Test-Guided HVAD RPM and Medication Adjustments on Exercise Tolerance and Quality of Life: the RAMP-IT-UP Multicenter Study | Inappropriate population |
| 8181 | A randomised controlled trial of a facilitated home-based rehabilitation intervention in patients with heart failure with preserved ejection fraction and their caregivers: the REACH-HFpEF Pilot Study | Inappropriate study design |
| 8182 | Grenada Heart Project-Community Health ActioN to EncouraGe healthy BEhaviors (GHP-CHANGE): a randomized control peer group-based lifestyle intervention | Inappropriate study design |
| 8183 | A placebo-controlled trial of captopril in refractory chronic congestive heart failure. Captopril Multicenter Research Group | Inappropriate population |
| 8184 | Splinting the hand in the functional position after brain impairment: a randomized, controlled trial | Inappropriate intervention |
| 8185 | Relationship between galectin-3 levels and mineralocorticoid receptor antagonist use in heart failure: analysis from HF-ACTION | Inappropriate population |
| 8186 | Staying Active With Arthritis: RCT of Physical Activity for Older Adults With Osteoarthritis and Hypertension | Inappropriate study design |
| 8187 | Baroreflex Activation Therapy for the Treatment of Heart Failure With a Reduced Ejection Fraction | Inappropriate study design |
| 8188 | Effects of Sacubitril/Valsartan on Physical and Social Activity Limitations in Patients With Heart Failure: a Secondary Analysis of the PARADIGM-HF Trial | Inappropriate study design |
| 8189 | Retro-walking in Knee Osteoarthritis | Inappropriate study design |
| 8190 | Study to assess the effectiveness of modified constraint-induced movement therapy in stroke subjects: a randomized controlled trial | Inappropriate outcome |
| 8191 | Effect of Gaming Assisted Visual Feedback on Functional Standing Balance among Acute Hemiparetic Stroke Patients | Inappropriate study design |
| 8192 | Impact of age, sex and heart rate variability on the acute cardiovascular response to isometric handgrip exercise | Inappropriate outcome |
| 8193 | Effects of treatment with trapidil and nifedipine on physical, emotional and cognitive exercise tolerance in patients with coronary heart disease | Inappropriate study design |
| 8194 | Radionuclide assessment of cardiac performance in cystic fibrosis. Reproducibility and effect of theophylline on cardiac function | Inappropriate population |
| 8195 | Behaviour change physiotherapy intervention to increase physical activity following hip and knee replacement (PEP-TALK): study protocol for a pragmatic randomised controlled trial | Inappropriate study design |
| 8196 | Nuclear magnetic resonance and radionuclide angiographic assessment of acute myocardial infarction in a randomized trial of intravenous streptokinase | Inappropriate study design |
| 8197 | Effects of left ventricular shape and captopril therapy on exercise capacity after anterior wall acute myocardial infarction | Inappropriate population |
| 8198 | Efficacy of an early home-based cardiac rehabilitation program for patients after acute myocardial infarction: a three-dimensional speckle tracking echocardiography randomized trial | Inappropriate intervention |
| 8199 | Determinants of left ventricular function two weeks and one year after an acute myocardial infarction | Inappropriate population |
| 8200 | Randomized double-blind trial of intravenous streptokinase for acute myocardial infarction | Inappropriate intervention |
| 8201 | Acute effects of dried apple consumption on metabolic and cognitive responses in healthy individuals | Inappropriate population |
| 8202 | Radial Extracorporeal Shock Wave Therapy for Spastic Plantar Flexor Muscles in Young Children With Cerebral Palsy | Inappropriate study design |
| 8203 | Family planning to promote physical activity: a randomized controlled trial protocol | Inappropriate population |
| 8204 | Effectiveness of behavioral graded activity after first-time lumbar disc surgery: short term results of a randomized controlled trial | Inappropriate study design |
| 8205 | "Active Team" a social and gamified app-based physical activity intervention: randomised controlled trial study protocol | Inappropriate population |
| 8206 | Stand out in Class: restructuring the classroom environment to reduce sedentary behaviour in 9-10-yearolds - Study protocol for a pilot cluster randomised controlled trial | Inappropriate study design |
| 8207 | Combining behavioural activation with physical activity promotion for adults with depression: findings of a parallel-group pilot randomised controlled trial (BAcPAc) | Inappropriate outcome |
| 8208 | Effect of Dapagliflozin on the Progression From Prediabetes to T2DM in Subjects With Myocardial Infarction | Inappropriate study design |
| 8209 | Trimetazidine attenuates high-altitude fatigue and cardiorespiratory fitness impairment: a randomized double-blinded placebo-controlled clinical trial | Inappropriate study design |

| | | |
|---|---|---|
| 8210 | Restoration of left ventricular synchronous contraction after acute myocardial infarction by stem cell therapy: new insights into the therapeutic implication of stem cell therapy for acute myocardial infarction | Inappropriate study design |
| 8211 | Comparison of strength training and task specific exercises to improve upper limb function in stroke patients | Inappropriate study design |
| 8212 | Upright pedalling is a potential intervention for driving lower limb recovery in people who cannot walk early after stroke | Inappropriate study design |
| 8213 | Lowering Blood Pressure by Changing Lifestyle | Inappropriate population |
| 8214 | Obesity Surgery, Counseling, and Psychological Well-Being | Inappropriate study design |
| 8215 | Effect of normobaric hypoxic exercise on dynamic cerebral autoregulation | Inappropriate study design |
| 8216 | Stroke treatment: comparison of integrated behavioral-physical therapy vs traditional physical therapy programs | Inappropriate outcome |
| 8217 | Comprehensive Use of Dynamic Electrical Neurostimulation and Botulinum Toxin Therapy in Patients with Post-Stroke Spasticity | Inappropriate study design |
| 8218 | Monitoring effects of acute hypoxia on brain cortical activity by using electromagnetic tomography | Inappropriate population |
| 8219 | Acute Effects of Fitness Drink Formulas on Energy Expenditure and Fat Metabolism | Inappropriate study design |
| 8220 | The effects of an acute dose of Rhodiola rosea on endurance exercise performance | Inappropriate study design |
| 8221 | Co-creating and evaluating an online self-help treatment (iSelf-help) for people with persistent pain | Inappropriate study design |
| 8222 | Multicenter randomized clinical trial evaluating the effectiveness of the Lokomat in subacute stroke | Inappropriate population |
| 8223 | History of intracranial hemorrhage is associated with in-hospital mortality in ischemic stroke patients treated with intravenous thrombolytics | Inappropriate study design |
| 8224 | Persistent effects of cognitive-behavioral stress management on cortisol responses to acute stress in healthy subjects--a randomized controlled trial | Inappropriate population |
| 8225 | Does cerebral oxygenation affect cognitive function during exercise? | Inappropriate study design |
| 8226 | Influence of sildenafil on lung diffusion during exposure to acute hypoxia at rest and during exercise in healthy humans | Inappropriate study design |
| 8227 | Effects of early captopril administration on infarct expansion, left ventricular remodeling and exercise capacity after acute myocardial infarction | Inappropriate study design |
| 8228 | Dietary Nitrate Increases VO2peak and Performance but Does Not Alter Ventilation or Efficiency in Patients With Heart Failure With Reduced Ejection Fraction | Inappropriate population |
| 8229 | Accuracy of new-generation handheld ecg devices compared to traditional 3-lead electrocardiogram | Inappropriate population |
| 8230 | Cycling induced by electrical stimulation improves muscle activation and symmetry during pedaling in hemiparetic patients | Inappropriate study design |
| 8231 | Lack of effect of low levels of carboxyhemoglobin on cardiovascular function in patients with ischemic heart disease | Inappropriate population |
| 8232 | Interaction between exercise training and ejection fraction in predicting prognosis after a first myocardial infarction | Inappropriate study design |
| 8233 | Usefulness of amiodarone for the Prevention of Atrial Fibrillation after implantation of a cardiac valve | Inappropriate study design |
| 8234 | Does Walking Mitigate Affective and Cognitive Responses to Social Exclusion? | Inappropriate study design |
| 8235 | Effect of transcranial direct current stimulation on sports performance for two profiles of athletes (power and endurance) (COMPETE): a protocol for a randomised, crossover, double blind, controlled exploratory trial | Inappropriate study design |
| 8236 | Effect of continuous cooling on inhibition and attention while wearing firefighter's PPE in a hot environment | Inappropriate population |
| 8237 | Psychological effects of a short behavior modification program in patients with acute myocardial infarction or coronary artery bypass grafting. A randomized controlled trial | Inappropriate outcome |
| 8238 | A randomised controlled trial of ramipril versus placebo when used in conjunction with angioplasty | Inappropriate study design |
| 8239 | Effect of a web site intervention on physical activity of college females | Inappropriate population |
| 8240 | Complete Decongestive Therapy on Head and Neck Lymphedema | Inappropriate population |
| 8241 | Danish trial of beta blocker treatment after myocardial infarction without signs of heart failure (DANBLOCK) | Inappropriate study design |
| 8242 | Rehabilitation Techniques on Survivors of Acute Respiratory Distress Syndrome | Inappropriate study design |

| | | |
|---|---|---|
| 8243 | Substrate Utilization, Exercise Performance, and Skeletal Muscle Response to Energy Deficit and Altitude Acclimatization | Inappropriate study design |
| 8244 | Effect of Home program Based activities of daily living therapy in individuals with vertigo to reduce symptoms and improve participation in routine tasks | Inappropriate population |
| 8245 | Caffeinated energy drink intake modulates motor circuits at rest, before and after a movement | Inappropriate study design |
| 8246 | Acute exercise induces cortical inhibition and reduces arousal in response to visual stimulation in young children | Inappropriate study design |
| 8247 | Modelflow underestimates cardiac output in heat-stressed individuals | Inappropriate study design |
| 8248 | Inhaled iloprost, exercise hemodynamics, and ventricular performance in heart failure with preserved ejection fraction | Inappropriate outcome |
| 8249 | Effects of exercise training on mobilization of BM-CPCs and migratory capacity as well as LVEF after AMI | Inappropriate study design |
| 8250 | Effects of perioperative administration of dexamethasone on postoperative complications and mortality after non-cardiac major surgery : a randomized, multicentre, double blind, study | Inappropriate study design |
| 8251 | Rumination induction after a TSST predicts non-habituation of salivary alpha-amylase responses to repeated stress | Inappropriate population |
| 8252 | A Multicenter, Randomized, Double-Blind Study to Evaluate the Safety and Efficacy of MK-0431 Monotherapy in Patients with Type 2 Diabetes Mellitus Who Have Inadequate Glycemic Control - MK-0431 Monotherapy Study in Patients with Type 2 DM | Inappropriate study design |
| 8253 | Elevated stress responses following sleep deprivation in healthy adults | Inappropriate intervention |
| 8254 | Examining the effectiveness of psychological strategies on physiologic markers: evidence-based suggestions for holistic care of the athlete | Inappropriate study design |
| 8255 | The influence of personal values and patient intoxication on nurses concerns about opioids: results of a prospective cross-sectional multi-centre study | Inappropriate study design |
| 8256 | A pilot randomized clinical trial of tDCS for increasing exercise engagement in individuals with elevated depressive symptoms: Rationale, design, and baseline characteristics | Inappropriate population |
| 8257 | Strengths-building through life purpose, self-care goal setting and social support: Study protocol for Caregiver Support | Inappropriate population |
| 8258 | The Effects of Low-intensity Resistance Training with Blood Flow Restriction Versus Traditional Resistance Exercise on Lower Extremity Muscle Strength, Walking Capacity, and Balance in Ischemic Stroke Survivors: A Study Protocol for the BFR-Stroke RESILIENCE Trial | Inappropriate population |
| 8259 | Comparison Between Effects of Functional Training Program and Conventional Therapy on Postural Control and Functional Mobility in Chronic Stroke | Inappropriate study design |
| 8260 | Effects of probiotic supplements on cognition, anxiety, and physical activity in subjects with mild and moderate Alzheimer’s disease: A randomized, double-blind, and placebo-controlled study | Inappropriate population |
| 8261 | Prescribing or co-designing exercise in healthy adults? Effects on mental health and interoceptive awareness | Inappropriate study design |
| 8262 | Association between trunk muscle endurance with change of direction, lower limb endurance and power performance in the special military police force of Rio de Janeiro (BOPE) | Inappropriate outcome |
| 8263 | Economic evaluation of the e-Health StandingTall balance exercise programme for fall prevention in people aged 70 years and over [with consumer summary] | Inappropriate population |
| 8264 | Protocol for a cluster randomised crossover pilot trial of Goal Management Training+ (GMT+) for methamphetamine use disorder | Inappropriate interention |
| 8265 | Does dual task merged in a mixed physical exercise protocol impact the mobility under dual task conditions in mild impaired stroke survivors? A feasibility, safety, randomized, and controlled pilot trial [with consumer summary] | Inappropriate outcome |
| 8266 | Effectiveness of Motor Imagery Training to Improve Gait Abilities of Patients with Sub-Acute Stroke | Inappropriate study design |
| 8267 | Efficacy of Robot-Assisted Gait Training Combined with Robotic Balance Training in Subacute Stroke Patients: A Randomized Clinical Trial | Inappropriate study design |
| 8268 | Approaches to Promote Reduction in Sedentary Behavior in Patients With Minor Ischemic Stroke: A Randomized Controlled Trial | Inappropriate population |
| 8269 | Impact of Activity Tracker Usage in Combination with a Physical Activity Intervention on Physical and Cognitive Parameters in Healthy Adults Aged 60+: A Randomized Controlled Trial | Inappropriate population |
| 8270 | Determinants of Health-Related Quality of Life Among Community-Dwelling Thai Older Adults in Chiang Mai, Northern Thailand | Inappropriate study design |

| | | |
|---|---|---|
| 8271 | EFFECT OF PERIPHERAL NEUROMODULATION ON HEADACHE IN POST COVID-19 SURVIVORS | Inappropriate population |
| 8272 | Efficacy of Body weight support treadmill as an accompaniment to conventional gait training Program in chronic stroke patients: An Experimental Study | Inappropriate population |
| 8273 | Aortic Valve Replacement Versus Conservative Treatment in Asymptomatic Severe Aortic Stenosis: The AVATAR Trial | Inappropriate population |
| 8274 | High-Volume Image-Guided Injections in Achilles and Patellar Tendinopathy in a Young Active Military Population: A Double-Blind Randomized Controlled Trial | Inappropriate population |
| 8275 | Examining the effect of increased aerobic exercise in moderately fit adults on psychological state and cognitive function | Inappropriate outcome |
| 8276 | Automatic topic selection for long-term interaction with embodied conversational agents in health coaching: A micro-randomized trial | Inappropriate study design |
| 8277 | Acute effects of beetroot juice and caffeine co-ingestion during a team-sport-specific intermittent exercise test in semi-professional soccer players: a randomized, double-blind, placebo-controlled study | Inappropriate interention |
| 8278 | Cortisol reactivity to a physical stressor in patients with depression and Alzheimer's disease | Inappropriate population |
| 8279 | Resistance Training Combined with Balance or Gait Training for Patients with Parkinson's Disease: A Randomized Controlled Pilot Study | Inappropriate outcome |
| 8280 | Effectiveness of the population-based a € check your health preventive programme' conducted in a primary care setting: A pragmatic randomised controlled trial | Inappropriate study design |
| 8281 | The effects of aerobic exercise training on cerebrovascular and cognitive function in sedentary, obese, older adults | Inappropriate population |
| 8282 | Exercise and Escitalopram in the Treatment of Anxiety in Patients with Coronary Heart Disease: One Year Follow-Up of the UNWIND Randomized Clinical Trial | Inappropriate population |
| 8283 | Young men's body hemodynamics variability in different states of motor activity | Inappropriate outcome |
| 8284 | An on-line school-based randomised controlled trial to prevent non-specific low back pain in children | Inappropriate study design |
| 8285 | Molecular Hydrogen Positively Affects Physical and Respiratory Function in Acute Post-COVID-19 Patients: A New Perspective in Rehabilitation | Inappropriate population |
| 8286 | Impact of combined versus single-mode training programs based on drop jump and specific multidirectional repeated sprint on bio-motor ability adaptations: a parallel study design in professional basketball players | Inappropriate population |
| 8287 | The Effects of 3D Custom Foot Orthotics with Mechanical Plantar Stimulation in Older Individuals with Cognitive Impairment: A Pilot Study | Inappropriate population |
| 8288 | Patterns and determinants of adherence to resistance and endurance training during cancer treatment in the Phys-Can RCT | Inappropriate study design |
| 8289 | Clinical efficacy of a personalized exercise program in the rehabilitation of patients with atrial fibrillation after radiofrequency ablation | Inappropriate study design |
| 8290 | Comparison of the Effect of Board Game and Tobacco Cessation Education on Nicotine Addiction in Adolescent Smokers | Inappropriate study design |
| 8291 | Effect of traditional Chinese manual therapy on alleviating pain and dysfunction of lumbar disc herniation: a randomized controlled pilot study | Inappropriate population |
| 8292 | Repetitive transcranial magnetic stimulation combined with respiratory muscle training for pulmonary rehabilitation after ischemic stroke—A randomized, case-control study | Inappropriate study design |
| 8293 | Previous functional social and behavioral rhythms affect resilience to COVID-19-related stress among old adults | Inappropriate population |
| 8294 | Effects of Vivifrail multicomponent intervention on functional capacity: a multicentre, randomized controlled trial | Inappropriate interention |
| 8295 | Molecular and Brain Volume Changes Following Aerobic Exercise, Cognitive and Combined Training in Physically Inactive Healthy Late-Middle-Aged Adults: The Projecte Moviment Randomized Controlled Trial | Inappropriate study design |
| 8296 | Effects of Resistance Exercise with Instability on Concerns about Falling and Depressive Symptoms in Cognitively Impaired Older Adults | Inappropriate interention |
| 8297 | The multi-domain exercise intervention for memory and brain function in late middle-aged and older adults at risk for Alzheimer's disease: A protocol for Western–Eastern Brain Fitness Integration Training trial | Inappropriate population |
| 8298 | The Patient Health Engagement Model in Cancer Management: Effect of Physical Activity, Distress Management, and Social Support Intervention to Improve the Quality of Life in Breast Cancer Patients | Inappropriate population |
| 8299 | Effect of Whole-body Vibration on Depression, Anxiety, Stress, and Quality of Life in College Students: A Randomized Controlled Trial | Inappropriate study design |

| | | |
|---|---|---|
| 8300 | Anterior or Posterior Ankle Foot Orthoses for Ankle Spasticity: Which One Is Better? | Inappropriate study design |
| 8301 | Isotemporal Substitution of Sedentary Behavior With Moderate to Vigorous Physical Activity Is Associated With Lower Risk of Disability: A Prospective Longitudinal Cohort Study | Inappropriate study design |
| 8302 | Efficacy of Fu's Subcutaneous Needling in Treating Soft Tissue Pain of Knee Osteoarthritis: A Randomized Clinical Trial | Inappropriate study design |
| 8303 | Six nights of sleep extension increases regional cerebral oxygenation without modifying cognitive performance at rest or following acute aerobic exercise | Inappropriate interention |
| 8304 | Benefits of meditation in reducing blood pressure and stress in patients with high blood pressure | Inappropriate study design |
| 8305 | A physiotherapy-led transition to home intervention for older adults following emergency department discharge: protocol for a pilot feasibility randomised controlled trial | Inappropriate study design |
| 8306 | IMPROvE-CED Trial: Intracoronary Autologous CD34+ Cell Therapy for Treatment of Coronary Endothelial Dysfunction in Patients With Angina and Nonobstructive Coronary Arteries | Inappropriate population |
| 8307 | Effects of game-based interventions on functional capacity in acutely hospitalised older adults: results of an open-label non-randomised clinical trial | Inappropriate population |
| 8308 | Randomised Controlled Trial of Fish Oil Supplementation on Responsiveness to Resistance Exercise Training in Sarcopenic Older Women; Pfizer(Brazil) | Inappropriate study design |
| 8309 | Chronic pain in the elderly: Mechanisms and perspectives | Inappropriate population |
| 8310 | Adjunctive Inspiratory Muscle Training During a Rehabilitation Program in Patients With Breast Cancer: An Exploratory Double-Blind, Randomized, Controlled Pilot Study | Inappropriate population |
| 8311 | CCRT and aerobic exercise: a randomised controlled study of processing speed, cognitive flexibility, and serum BDNF expression in schizophrenia | Inappropriate population |
| 8312 | Moderate-intensity continuous training and high-intensity interval training improve cognition, and BDNF levels of middle-aged overweight men | Inappropriate study design |
| 8313 | Effect of an exercise intervention on global cognition after transient ischemic attack or minor stroke: the MoveIT randomized controlled trial | Inappropriate population |
| 8314 | Mental and physical training with meditation and aerobic exercise improved mental health and well-being in teachers during the COVID-19 pandemic | Inappropriate study design |
| 8315 | Hippocampal maintenance after a 12-month physical activity intervention in older adults: the REACT MRI study [with consumer summary] | Inappropriate study design |
| 8316 | Assessment of Yogic Exercise Intervention on Aerobic Capacity (Vo2max) of Women Soccer Players | Inappropriate study design |
| 8317 | Blood withdrawal acutely impairs cardiac filling, output and aerobic capacity in proportion to induced hypovolemia in middle-aged and older women | Inappropriate study design |
| 8318 | Playfulness for Brain Health in Mid- and Late-Life: Scoping Review to Support Preventive Non-Pharmacological Interventions in Community Settings | Inappropriate study design |
| 8319 | Obesity, Psychological Distress, and Resting State Connectivity of the Hippocampus and Amygdala Among Women With Early-Stage Breast Cancer | Inappropriate interention |
| 8320 | Feasibility Randomized Controlled Trial of a Mind-Body Activity Program for Older Adults With Chronic Pain and Cognitive Decline: The Virtual "Active Brains" Study | Inappropriate population |
| 8321 | Multiple routes to help you roam: A comparison of training interventions to improve cognitive-motor dual-tasking in healthy older adults | Inappropriate study design |
| 8322 | PSYCHOPHYSIOLOGY OF BASKETBALL PLAYERS SUBMITTED TO HIGH INTENSITY EXERCISE | Inappropriate study design |
| 8323 | No evidence for a boost in psychosocial functioning in older age after a 6-months physical exercise intervention | Inappropriate outcome |
| 8324 | Benefits of aquatic exercise in adults with and without chronic disease—A systematic review with meta?analysis | Inappropriate population |
| 8325 | A 2x2 randomised factorial SWAT of the use of a pen and small, financial incentive to improve recruitment in a randomised controlled trial of yoga for older adults with multimorbidity | Inappropriate study design |
| 8326 | Acute Effects of Augmented Reality Exergames versus Cycle Ergometer on Reaction Time, Visual Attention, and Verbal Fluency in Community Older Adults | Inappropriate population |
| 8327 | Effects of social media on smartphone use before and during velocity-based resistance exercise on cognitive interference control and physiological measures in trained adults | Inappropriate outcome |
| 8328 | The Effect of Self-Paced Exercise Intensity and Cardiorespiratory Fitness on Frontal Grey Matter Volume in Cognitively Normal Older Adults: A Randomised Controlled Trial | Inappropriate study design |

| | | |
|---|---|---|
| 8329 | An integrated intervention of computerized cognitive training and physical exercise in virtual reality for people with Alzheimer's disease: The jDome study protocol | Inappropriate interention |
| 8330 | Imagery Rescripting for Reducing Body Image Dissatisfaction: A Randomized Controlled Trial | Inappropriate study design |
| 8331 | Efficacy of Occupational Performance Coaching with and without Four Quadrant Model of Facilitated Learning for mothers of children with specific learning disorder: Study protocol for a randomized controlled trial | Inappropriate outcome |
| 8332 | Sedentary Behavior and Physical Functioning in Middle-Age and Older Adults Living in the United States: The Reasons for Geographic and Racial Differences in Stroke Study | Inappropriate population |
| 8333 | A qualitative evaluation of participants experiences of living with back pain, lumbar fusion surgery, and post-operative rehabilitation | Inappropriate outcome |
| 8334 | Recruitment and Screening Methods in Alzheimer's Disease Research: The FIT-AD Trial | Inappropriate outcome |
| 8335 | Virtual reality for management of cancer pain: Study rationale and design | Inappropriate interention |
| 8336 | Using gait robotics to improve symptoms of Parkinson's disease: an open-label, pilot randomized controlled trial | Inappropriate study design |
| 8337 | Exercise Effects on Cognition in Older African Americans: A Pilot Randomized Trial | Inappropriate outcome |
| 8338 | What is real change in submaximal cardiorespiratory fitness in older adults? Retrospective analysis of a clinical trial | Inappropriate study design |
| 8339 | Evaluating the Management of chronic Pelvic girdle Pain following pregnancy (EMaPP): study protocol for a randomised controlled feasibility trial to compare a customised pelvic orthosis with standard care | Inappropriate study design |
| 8340 | A Tailored Music-Motor Therapy and Real-Time Biofeedback Mobile Phone App (‘GotRhythm’) to Promote Rehabilitation Following Stroke: A Pilot Study | Inappropriate population |
| 8341 | The Effect of Ginger (Zingiber officinale Rosc.) Consumption in Headache Prophylaxis in Patients with Migraine: A Randomized Placebo-Controlled Clinical Trial; dineh pharmaceutical supplements(Iran) | Inappropriate population |
| 8342 | Exercise volume load in women with breast cancer: Study protocol for the ABRACE randomized clinical trial | Inappropriate outcome |
| 8343 | Sex differences during a cold-stress test in normobaric and hypobaric hypoxia: A randomized controlled crossover study | Inappropriate population |
| 8344 | Three-month Chan-Chuang Qigong program improves physical performance and quality of life of patients with cognitive impairment: a randomized controlled trial | Inappropriate study design |
| 8345 | Sex differences in the association between different obesity parameters and cognitive function in older adults: A cross-sectional study in rural China | Inappropriate interention |
| 8346 | Effectiveness of using calligraphic activity to treat people with schizophrenia: a randomized controlled trial in Southern Taiwan | Inappropriate outcome |
| 8347 | Promoting independence in Lewy body dementia through exercise: the PRIDE study | Inappropriate study design |
| 8348 | Comparative Effects of Whole-Body Vibration and Squatting Training on Balance and Mobility in Chronic Stroke Patients. A Randomized Controlled Trial | Inappropriate population |
| 8349 | The effect of home-based cardiac rehabilitation on depression score in patients with ischemic heart disease: A longitudinal clinical trial study | Inappropriate population |
| 8350 | Feasibility of a home-based physiotherapy intervention to promote post-stroke mobility: A randomized controlled pilot study | Inappropriate population |
| 8351 | The effect of a masticatory muscle training program on chewing efficiency and bite force in people with dementia | Inappropriate population |
| 8352 | Transcranial Direct Current Stimulation May Reduce Prefrontal Recruitment During Dual Task Walking in Functionally Limited Older Adults – A Pilot Study | Inappropriate study design |
| 8353 | Degenerative Mitral Stenosis--Diagnostic Challenges and Future Directions | Inappropriate study design |
| 8354 | Feasibility and Pilot Testing of Mobile Health Apps to Supplement 2 Healthy Lifestyle Interventions in Chronic Traumatic Brain Injury | Inappropriate population |
| 8355 | Effects of a multidisciplinary intervention to promote physical activity in patients with stroke undergoing rehabilitation: Study protocol for the ActivePAS pilot randomised controlled trial | Inappropriate study design |
| 8356 | Targeting Adiposity and Inflammation With Movement to Improve Prognosis in Breast Cancer Survivors (The AIM Trial): Rationale, Design, and Methods | Inappropriate population |
| 8357 | Treatment of Oropharyngeal Symptoms: A Prospective, Single-Dose, Placebo-Controlled, Randomized Clinical Trial; bemis(United States) | Inappropriate outcome |
| 8358 | Efficacy of Adaptogens in Patients with Long COVID-19: A Randomized, Quadruple-Blind, Placebo-Controlled Trial | Inappropriate outcome |

| | | |
|---|---|---|
| 8359 | Efficacy of Wearable Device Gait Training on Parkinson's Disease: A Randomized Controlled Open-label Pilot Study | Inappropriate outcome |
| 8360 | Comparing the Cognitive Functioning Effects of Aerobic and Pilates Exercises for Inactive Young Adults: A Randomized Controlled Trial | Inappropriate interention |
| 8361 | Flexibility and resistance exercises versus usual care for improving pain and function after distal radius fracture in adults aged 50 years or over: protocol for the WISE randomised multicentre feasibility trial | Inappropriate interention |
| 8362 | Effect of aerobic exercise as adjunct therapy on the improvement of negative symptoms and cognitive impairment in patients with schizophrenia: a randomized, case-control clinical trial | Inappropriate outcome |
| 8363 | Is Swiss-ball-based exercise superior to plinth-based exercise in improving trunk motor control and balance in subjects with sub-acute stroke? A pilot randomized control trial | Inappropriate population |
| 8364 | MicroRNA Expression Variation in Female Dog (Canis familiaris) Reproductive Organs with Age and Presence of Uteropathy | Inappropriate interention |
| 8365 | The Efficacy of Pain Neuroscience Education on Active Rehabilitation following Arthroscopic Rotator Cuff Repair: A CONSORT?Compliant Prospective Randomized Single?Blind Controlled Trial | Inappropriate study design |
| 8366 | Effects of Immersive Virtual Therapy as a Method Supporting Recovery of Depressive Symptoms in Post-Stroke Rehabilitation: Randomized Controlled Trial | Inappropriate population |
| 8367 | The Effects of Standardised versus Individualised Aerobic Exercise Prescription on Fitness-Fatness Index in Sedentary Adults: A Randomised Controlled Trial | Inappropriate interention |
| 8368 | Efficacy of Cognitive-Behavioral Therapy for the Prophylaxis of Migraine in Adults: A Three-Armed Randomized Controlled Trial | Inappropriate interention |
| 8369 | Handgrip Strength Asymmetry and Weakness Are Associated With Future Morbidity Accumulation in Americans | Inappropriate study design |
| 8370 | Patients' and clinicians' experiences with stratified exercise therapy in knee osteoarthritis: a qualitative study | Inappropriate study design |
| 8371 | Exercise restores brain insulin sensitivity in sedentary adults who are overweight and obese | Inappropriate population |
| 8372 | A comparative study on effectiveness of aerobic exercise versus resistance exercise on improving cognitive function in older adults | Inappropriate study design |
| 8373 | The Impact of Health Promotion Activities on the Physiological, Psychological, and Social Functions of Inpatients With Chronic Mental Illness | Inappropriate outcome |
| 8374 | Attenuated inflammatory profile following single and repeated handgrip exercise and remote ischemic preconditioning in patients with cerebral small vessel disease | Inappropriate interention |
| 8375 | Long-term effect of additional rehabilitation following botulinum toxin-A on upper limb activity in chronic stroke: the InTENSE randomised trial | Inappropriate outcome |
| 8376 | Postexercise hypotensive response in stroke patients following acute moderate or high intensity cycling session | Inappropriate outcome |
| 8377 | Immersive Technology for Cognitive-Motor Training in Parkinson's Disease | Inappropriate population |
| 8378 | Effects of functional task exercise on everyday problem-solving ability and functional status in older adults with mild cognitive impairment -- a randomised controlled trial [with consumer summary] | Inappropriate outcome |
| 8379 | The effects of acute exercise on driving and executive functions in healthy older adults | Inappropriate study design |
| 8380 | Effects of Mindfulness Training and Exercise on Cognitive Function in Older Adults: A Randomized Clinical Trial | Inappropriate study design |
| 8381 | Exercise interveNtion outdoor proJect in the cOmmunitY—ENJOY program for independence in dementia: a feasibility pilot randomised controlled trial study protocol | Inappropriate study design |
| 8382 | Effect of Omecamtiv Mecarbil on Exercise Capacity in Chronic Heart Failure With Reduced Ejection Fraction: The METEORIC-HF Randomized Clinical Trial | Inappropriate study design |
| 8383 | A cognitively enhanced online Tai Ji Quan training intervention for community-dwelling older adults with mild cognitive impairment: a feasibility trial | Inappropriate population |
| 8384 | Effects of exergaming-based Tai Chi on cognitive function and dual-task gait performance in older adults with mild cognitive impairment: a randomized control trial | Inappropriate population |
| 8385 | Effects of Exercise Intervention on Type 2 Diabetes Patients With Abdominal Obesity and Low Thigh Circumference (EXTEND): Study Protocol for a Randomized Controlled Trial | Inappropriate population |
| 8386 | Motor Training After Stroke: A Novel Approach for Driving Rehabilitation | Inappropriate population |
| 8387 | IMPROVING PATIENT HEALTH BEHAVIOR IN PRIMARY CARE THROUGH SELF-REGULATION | Inappropriate population |

| | | |
|---|---|---|
| 8388 | Cognitive-motor multitasking in older adults: a randomized controlled study on the effects of individual differences on training success | Inappropriate study design |
| 8389 | Program of Rehabilitative Exercise and Education to Avert Vascular Events After Non-Disabling Stroke or Transient Ischemic Attack (PREVENT Trial): A Randomized Controlled Trial | Inappropriate study design |
| 8390 | Effects of a 6-month multifaceted diet and exercise intervention on cognition in older adults at risk of cognitive decline: the PONDER double-blind, placebo-controlled randomized trial | Inappropriate study design |
| 8391 | Effect of core stabilization exercises in addition to conventional therapy in improving trunk mobility, function, ambulation and quality of life in stroke patients: a randomized controlled trial | Inappropriate interention |
| 8392 | Sunbeam program reduces rate of falls in long-term care residents with mild to moderate cognitive impairment or dementia: subgroup analysis of a cluster randomized controlled trial | Inappropriate study design |
| 8393 | The Influence of a Competitive Field Hockey Match on Cognitive Function | Inappropriate study design |
| 8394 | Feasibility and efficacy of home-based neurofunctional exercise vs. resistance exercise programs for ambulatory disability of multiple sclerosis patients with cognitive impairment | Inappropriate population |
| 8395 | Feasibility and efficacy of home-based neurofunctional exercise versus resistance exercise programs for ambulatory disability of multiple sclerosis patients with cognitive impairment [with consumer summary] | Inappropriate study design |
| 8396 | Effects of self-selected or randomly selected music on performance and psychological responses during a sprint interval training session | Inappropriate population |
| 8397 | Comparison of Different Recovery Strategies After High-Intensity Functional Training: A Crossover Randomized Controlled Trial | Inappropriate population |
| 8398 | Effectiveness of Ultrasonography Visual Biofeedback of the Diaphragm in Conjunction with Inspiratory Muscle Training on Muscle Thickness, Respiratory Pressures, Pain, Disability, Quality of Life and Pulmonary Function in Athletes with Non-Specific Low Back Pain: A Randomized Clinical Trial | Inappropriate population |
| 8399 | The Effect of Physical Activity on Sleep Quality among Older Stroke Survivors: Secondary Analysis from a Randomized Controlled Trial | Inappropriate interention |
| 8400 | Does Cognitive Training Improve Mobility, Enhance Cognition, and Promote Neural Activation? | Inappropriate study design |
| 8401 | The Effect of Brief Stair-Climbing on Divergent and Convergent Thinking | Inappropriate population |
| 8402 | Do patients with fibromyalgia have body image and tactile acuity distortion? | Inappropriate study design |
| 8403 | Magnitude, timing and duration of mood state and cognitive effects of acute moderate exercise in major depressive disorder | Inappropriate study design |
| 8404 | Effects of Two Short-Term Aerobic Exercises on Cognitive Function in Healthy Older Adults during COVID-19 Confinement in Japan: A Pilot Randomized Controlled Trial | Inappropriate interention |
| 8405 | Executive functions in children engaging in open- and closed-skilled sports | Inappropriate population |
| 8406 | What are the effects of acute exercise and exercise training on cerebrovascular hemodynamics following stroke? A systematic review and meta-analysis | Inappropriate study design |
| 8407 | Interrelationships between exercise, functional connectivity, and cognition among healthy adults: A systematic review | Inappropriate population |
| 8408 | Sixteen weeks of aerobic exercise does not alter resting-state connectivity of the precuneus in patients with Alzheimer's disease | Inappropriate population |
| 8409 | Evaluation of quality of life and degree of autonomy among elderly subjects with type 2 diabetes | Inappropriate population |
| 8410 | Physical activity from young adulthood to middle age and premature cardiovascular disease events: A 30-year population-based cohort study | Inappropriate study design |
| 8411 | Effects of a Brief Stair-Climbing Intervention on Cognitive Functioning and Mood States in Older Adults: A Randomized Controlled Trial | Inappropriate study design |
| 8412 | Feasibility of context-specific activities for improving physical activity levels among Indian adults with stroke | Inappropriate population |
| 8413 | Group-led creative writing and behavioural health in cancer: A randomised clinical trial | Inappropriate population |
| 8414 | Effects of a taped filter mask on peak power, perceived breathlessness, heart rate, blood lactate and oxygen saturation during a graded exercise test in young healthy adults: a randomized controlled trial | Inappropriate population |
| 8415 | Immediate Effect of Ujjayi Pranayama on Attention and Anxiety among University Students: A Randomised Self-control Study | Inappropriate study design |
| 8416 | Aerobic exercise for HIV-associated neurocognitive disorders in individuals on antiretroviral therapy: a randomised controlled trial [with consumer summary] | Inappropriate study design |

| | | |
|---|---|---|
| 8417 | Testing the efficacy of a motor analogy designed to promote safe landing by older adults who fall accidentally: a study protocol for a randomised control study | Inappropriate population |
| 8418 | Effects of intranasal oxytocin on the self-perception and anxiety of singers during a simulated public singing performance: A randomized, placebo-controlled trial | Inappropriate study design |
| 8419 | Cerebrovascular response to exercise interacts with individual genotype and amyloid-beta deposition to influence response inhibition with aging | Inappropriate population |
| 8420 | Longitudinal study of the effect of a 5-year exercise intervention on structural brain complexity in older adults A generation 100 substudy | Inappropriate study design |
| 8421 | Dual-task Zumba Gold for improving the cognition of people with mild cognitive impairment: a pilot randomized controlled trial | Inappropriate study design |
| 8422 | Effects of acute moderate-intensity aerobic exercise on executive function and prefrontal cortex activity in community-dwelling older adults: A single-blind, randomized controlled trial | Inappropriate study design |
| 8423 | Sympathoexcitatory responses to isometric handgrip exercise are associated with white matter hyperintensities in middle-aged and older adults | Inappropriate study design |
| 8424 | Acute Effect of a Simultaneous Exercise and Cognitive Task on Executive Functions and Prefrontal Cortex Oxygenation in Healthy Older Adults | Inappropriate population |
| 8425 | Effects of 12-Week Supervised Early Resistance Training (SEcReT) Versus Aerobic-Based Rehabilitation on Cognitive Recovery Following Cardiac Surgery via Median Sternotomy: A Pilot Randomised Controlled Trial | Inappropriate population |
| 8426 | Effects of cycling dual-task on cognitive and physical function in Parkinson's disease: a randomized double-blind pilot study | Inappropriate study design |
| 8427 | Step Number and Aerobic Minute Exercise Prescription and Progression in Stroke: A Roadmap | Inappropriate study design |
| 8428 | Physical activity, memory function, and hippocampal volume in adults with Down syndrome | Inappropriate population |
| 8429 | Cognitive and psychosocial effects of an acute sun salutation intervention among adults with stress | Inappropriate outcome |
| 8430 | Feasibility of Online High-Intensity Interval Training (HIIT) on Psychological Symptoms in Students in Lockdown During the COVID-19 Pandemic: A Randomized Controlled Trial | Inappropriate study design |
| 8431 | High-intensity aerobic interval training and resistance training are feasible in rectal cancer patients undergoing chemoradiotherapy: a feasibility randomized controlled study | Inappropriate outcome |
| 8432 | Body mass index and cognitive decline among community-living older adults: the modifying effect of physical activity | Inappropriate study design |
| 8433 | TRACking health behaviors in people with Multiple Sclerosis (TRAC-MS): Study protocol and description of the study sample | Inappropriate interention |
| 8434 | The effects of blood flow restriction resistance training on indices of delayed onset muscle soreness and peak power | Inappropriate population |
| 8435 | The association between sleep deficits and sedentary behavior in people with mild Parkinson disease | Inappropriate outcome |
| 8436 | Early hip abductor training to improve balance and walking ability after total knee arthroplasty | Inappropriate outcome |
| 8437 | Association between ultra-processed food consumption and cognitive performance in US older adults: a cross-sectional analysis of the NHANES 2011–2014 | Inappropriate population |
| 8438 | Leisure-time sedentary behaviors are differentially associated with all-cause dementia regardless of engagement in physical activity | Inappropriate outcome |
| 8439 | A STUDY ON THE MANAGEMENT OF ANAEMIA IN CHRONIC KIDNEY DISEASE | Inappropriate population |
| 8440 | Whole-Body Vibration Associated with Strength Training on the Lower-Limb Blood Flow and Mobility in Older Adults with Type 2 Diabetes: A Study Protocol for a Randomized Controlled Trial | Inappropriate population |
| 8441 | Effectiveness of Negative Pulsed-Pressure Myofascial Vacuum Therapy and Therapeutic Exercise in Chronic Non-Specific Low Back Pain: A Single-Blind Randomized Controlled Trial | Inappropriate study design |
| 8442 | Short- and medium-term effects of manual therapy on the upper cervical spine combined with exercise vs isolated exercise in patients with cervicogenic headache. A randomized controlled trial | Inappropriate study design |
| 8443 | Multicomponent exercise program effects on fitness and cognitive function of elderlies with mild cognitive impairment: Involvement of oxidative stress and BDNF | Inappropriate population |

| | | |
|---|---|---|
| 8444 | Diagnostic Strategy for Suspected Pulmonary Embolism in Emergency Departments Based on the 4-Level Pulmonary Embolism Clinical Probability Score: Study Protocol of SPEED&PEPS Trial | Inappropriate population |
| 8445 | One night of total sleep deprivation impairs decision-making skills, anaerobic performances, mood states, and perceptual responses in elite Kung-Fu athletes | Inappropriate population |
| 8446 | The Impact of the Otago Exercise Program on Frailty and Empowerment in Older Nursing Home Residents: A Randomized Controlled Trial | Inappropriate study design |
| 8447 | Study Protocol for a Multicenter, Randomized Controlled Trial to Improve Upper Extremity Hemiparesis in Chronic Stroke Patients by One-to-One Training (NEURO®) with Repetitive Transcranial Magnetic Stimulation | Inappropriate study design |
| 8448 | Exercise Dose and Aerobic Fitness Response in Alzheimer?s Dementia: Findings from the FIT-AD Trial | Inappropriate population |
| 8449 | Increasing time spent engaging in moderate-to-vigorous physical activity by community-dwelling adults following a transient ischemic attack or non-disabling stroke: a systematic review | Inappropriate outcome |
| 8450 | Examining the cognitive benefits of probiotic supplementation in physically active older adults: A randomized clinical trial | Inappropriate population |
| 8451 | Valued living among veterans in breath-based meditation treatment or cognitive processing therapy for posttraumatic stress disorder: Exploratory outcome of a randomized controlled trial | Inappropriate population |
| 8452 | The impact of a multicomponent-functional training with postural correction on functional balance in the elderly with a history of falling | Inappropriate outcome |
| 8453 | Randomization to Treadmill Training Improves Physical and Metabolic Health in Association With Declines in Oxidative Stress in Stroke | Inappropriate outcome |
| 8454 | Effects of Retro-Walking on Pain, Functional Disability and Performance in Knee Osteoarthritis | Inappropriate population |
| 8455 | Auditory rhythmical cueing to improve gait in community-dwelling stroke survivors (ACTIVATE): a pilot randomised controlled trial | Inappropriate outcome |
| 8456 | Effects of multidisciplinary exercise management on patients after percutaneous coronary intervention: A randomized controlled study | Inappropriate population |
| 8457 | Equipment-free, unsupervised high intensity interval training elicits significant improvements in the physiological resilience of older adults | Inappropriate population |
| 8458 | Cardiovascular Mechanisms of Exercise Intolerance in Older Patients with Heart Failure | Inappropriate study design |
| 8459 | An Educational and Exercise Mobile Phone-Based Intervention to Elicit Electrophysiological Changes and to Improve Psychological Functioning in Adults With Nonspecific Chronic Low Back Pain (BackFit App): Nonrandomized Clinical Trial | Inappropriate population |
| 8460 | Effect of facemask use on cognitive function during a maximal running aerobic fitness test | Inappropriate study design |
| 8461 | Short-and Long-Term Effectiveness of Low-Level Laser Therapy Combined with Strength Training in Knee Osteoarthritis: A Randomized Placebo-Controlled Trial | Inappropriate population |
| 8462 | A randomized controlled trial for reduction of risk factors for cardiovascular disease in young adults: Methods of the Aspire study | Inappropriate outcome |
| 8463 | Deep Brain Stimulation in Early-Stage Parkinson’s Disease: Patient Experience after 11 Years | Inappropriate population |
| 8464 | The Salzburg 10/7 HIIT shock cycle study: the effects of a 7-day high-intensity interval training shock microcycle with or without additional low-intensity training on endurance performance, well-being, stress and recovery in endurance trained athletes—study protocol of a randomized controlled trial | Inappropriate population |
| 8465 | EFFICACY OF MANUAL THERAPY COMPARED WITH USUAL GENERAL PRACTITIONER CARE FOR CHRONIC TENSION HEADACHE | Inappropriate population |
| 8466 | The Efficacy of a Newly Developed Cueing Device for Gait Mobility in Parkinson's Disease | Inappropriate study design |
| 8467 | Efficacy and Safety of Acute Phase Intensive Electrical Muscle Stimulation in Frail Older Patients with Acute Heart Failure: Results from the ACTIVE-EMS Trial | Inappropriate population |
| 8468 | Physiological Adaptations to High-Intensity Interval Training Combined with Blood Flow Restriction in Masters Road Cyclists | Inappropriate population |
| 8469 | The power of instruction on retropulsion: A pilot randomized controlled trial of therapeutic exercise focused on ankle joint movement in Parkinson's disease | Inappropriate population |
| 8470 | Effects of task-oriented training on walking in children with cerebral palsy | Inappropriate study design |
| 8471 | Aerobic exercise training and neurocognitive function in cognitively normal older adults: A one-year randomized controlled trial | Inappropriate study design |

| | | |
|---|---|---|
| 8472 | Effectiveness of a complex intervention of group-based nutrition and physical activity to prevent frailty in pre-frail older adults (SUPER): a randomised controlled trial | Inappropriate study design |
| 8473 | Efficacy and Safety of TurmXTRA® 60N in Delayed Onset Muscle Soreness in Healthy, Recreationally Active Subjects: A Randomized, Double-Blind, Placebo-Controlled Trial | Inappropriate population |
| 8474 | Acute Effects of Exercise on Risk-Taking: Different Responses in Males and Females | Inappropriate study design |
| 8475 | The Relationship between Physical Fitness and Reasoning in Community-Dwelling Older Adults | Inappropriate study design |
| 8476 | Acute effects of mind-body practices and exercise in depressed inpatients: A randomized clinical trial | Inappropriate population |
| 8477 | Feasibility and usefulness of a consumer-grade device for physical activity monitoring for patients with Parkinson's disease and Parkinsonism | Inappropriate study design |
| 8478 | Acute and protocol-dependent effects of aerobic exercise on neurobiochemical indices and neuropsychological performance of working memory | Inappropriate population |
| 8479 | Effects of Combination of Functional Electric Stimulation and Robotic Leg Movement Using Dynamic Tilt Table on Walking Characteristics in Post-Stroke Patients with Spastic Hemiplegia: A Randomized Crossover-Controlled Trial | Inappropriate study design |
| 8480 | Effects of exercise training on brain metabolism and cognitive functioning in sleep apnea | Inappropriate population |
| 8481 | Regional cortical perfusion increases induced by a 6-month endurance training in young sedentary adults | Inappropriate outcome |
| 8482 | Self-Management Of Patients With Mild Copd In Primary Care: A Random Controlled Trial | Inappropriate outcome |
| 8483 | Study protocol for the BRAIN Training Trial: A randomised controlled trial of Balance, Resistance, and INterval training on cognitive function in older adults with mild cognitive impairment | Inappropriate interention |
| 8484 | Effect of high-intensity interval training on hippocampal metabolism in older adolescents | Inappropriate interention |
| 8485 | High-Intensity Inspiratory Muscle Training Improves Scalene and Sternocleidomastoid Muscle Oxygenation Parameters in Patients With Weaning Difficulties: A Randomized Controlled Trial | Inappropriate outcome |
| 8486 | Menstrual cycle-based undulating periodized program effects on body composition and strength in trained women: a pilot study | Inappropriate interention |
| 8487 | Dementia risk and dynamic response to exercise: A non-randomized clinical trial | Inappropriate population |
| 8488 | Combined Stimulation of the Substantia Nigra and the Subthalamic Nucleus for the Treatment of Refractory Gait Disturbances in Parkinson’s Disease: A Preliminary Study | Inappropriate interention |
| 8489 | Comparison of the Cognitive Benefits of Doing Aerobic Exercise versus Performing Theater in Healthy Older Adults | Inappropriate interention |
| 8490 | A Comparison of the Effect of Physical Activity and Cognitive Training on Dual-Task Performance in Older Adults | Inappropriate interention |
| 8491 | Protective effect of aerobic fitness on the detrimental influence of exhaustive exercise on information processing capacity | Inappropriate population |
| 8492 | Chronotype and cognitive function: Observational study and bidirectional Mendelian randomization | Inappropriate study design |
| 8493 | Effect of Tai Chi Quan on the Pressure Pain Thresholds of Lower Back Muscles in Healthy Women | Inappropriate population |
| 8494 | Tai Chi Quan Versus Physical Therapy on Pain and Cognitive Performance for Elderly People With Chronic Low Back Pain: Study Protocol for a Randomized Controlled Trial | Inappropriate population |
| 8495 | CORE STRENGTH TRAINING IN UNIVERSITY FEMALE TENNIS PLAYERS | Inappropriate interention |
| 8496 | Rationale and methodology for examining the acute effects of aerobic exercise combined with varying degrees of virtual reality immersion on cognition in persons with TBI | Inappropriate population |
| 8497 | Prolonged electrocardiography registration does not lead to increased diagnosis of atrial fibrillation in pulmonary embolism patients, but sex affects generic health-related quality of life: Findings from a randomized clinical trial | Inappropriate interention |
| 8498 | Use of real-time multimodal sensory feedback home program improved backward stride and retention for people with Parkinson Disease: A pilot study | Inappropriate population |
| 8499 | Transcranial Direct Current Stimulation on Different Targets to Modulate Cortical Activity and Dual-Task Walking in Individuals With Parkinson’s Disease: A Double Blinded Randomized Controlled Trial | Inappropriate population |

| | | |
|---|---|---|
| 8500 | EFFECTS OF EXERCISE INTERVENTIONS ON COGNITIVE FUNCTIONS IN HEALTHY POPULATIONS: A SYSTEMATIC REVIEW AND META-ANALYSIS | Inappropriate study design |
| 8501 | Exercise Improves Physical Capacity, Cognition, Quality of Life and Promotes Neurotrophic Factors in Patients with Multiple Sclerosis | Innapropriate intervention |
| 8502 | De-frailing intervention for hospitalized cardiovascular patients in the TARGET-EFT randomized clinical trial [with consumer summary] | Inappropriate population |
| 8503 | The effects of low-intensity resistance training with blood flow restriction versus traditional resistance exercise on lower extremity muscle strength and motor functionin ischemic stroke survivors: a randomized controlled trial | Inappropriate outcome |
| 8504 | Seated postural organization during bilateral upper limb symmetric and asymmetric pushing tasks in individuals after stroke compared to healthy controls | Inappropriate study design |
| 8505 | Yoga and aerobic exercise in epilepsy: study protocol for a randomized controlled trial | Inappropriate study design |
| 8506 | A Randomized Clinical Trial to Evaluate a Digital Therapeutic to Enhance Gait Function in Individuals With Parkinson's Disease | Inappropriate outcome |
| 8507 | Do executive function performance, gaze behavior, and pupil size change during incremental acute physical exercise? | Inappropriate study design |
| 8508 | Chronic exercise and neuropsychological function in healthy young adults: a randomised controlled trial investigating a running intervention | Inappropriate population |
| 8509 | Effects of Mental Fatigue Induced by Stroop Task and by Social Media Use on Resistance Training Performance, Movement Velocity, Perceived Exertion, and Repetitions in Reserve: a Randomized and Double-Blind Crossover Trial | Inappropriate population |
| 8510 | Mental fatigue impairs physical performance but not the neural drive to the muscle: a preliminary analysis | Inappropriate population |
| 8511 | Mobile health technology, exercise adherence and optimal nutrition post rehabilitation among people with Parkinson's Disease (mHEXANUT) - a randomized controlled trial protocol | Inappropriate study design |
| 8512 | Gamified family-based health exercise intervention to improve adherence to 24-h movement behaviors recommendations in children: 3, 2, 1 Move on Study"" | Inappropriate study design |
| 8513 | Home-based self-management for sedentary individuals with mild walking disability after stroke: protocol for a randomised pilot study | Inappropriate study design |
| 8514 | Effect of Sit-to-Stand Training on Balance, Muscle Strength, and Activities of Daily Living in Patients with Stroke: a Randomised Controlled Trial | Inappropriate population |
| 8515 | Effect of the association of continuous shortwave diathermy and Pilates-based exercises on pain, depression, and anxiety in chronic non-specific low back pain: a randomized clinical trial | Inappropriate population |
| 8516 | TPVB and general anesthesia affects postoperative functional recovery in elderly patients with thoracoscopic pulmonary resections based on ERAS pathway | Inappropriate population |
| 8517 | To combine or not physical therapy with tDCS for stroke with shoulder pain? Analysis for rehabilitation of painful shoulder stroke | Inappropriate study design |
| 8518 | Improved processing speed and decreased functional connectivity in individuals with chronic stroke after paired exercise and motor training | Innapropriate intervention |
| 8519 | Improved cognitive-motor processing speed is related to a decrease in functional connectivity between the dorsolateral prefrontal cortex and the sensorimotor network after aerobic exercise in chronic stroke | Inappropriate population |
| 8520 | Effects of Nordic walking in Alzheimer's disease: a single-blind randomized controlled clinical trial | Innapropriate intervention |
| 8521 | #REF! | Innapropriate intervention |
| 8522 | Effects of exercise training on cardiac toxicity markers in women with breast cancer undergoing chemotherapy with anthracyclines: a randomized controlled trial | Inappropriate population |
| 8523 | Acute exercise and children's cognitive functioning: What is the optimal dose of cognitive challenge? | Inappropriate study design |
| 8524 | Motor, cognitive, and combined rehabilitation approaches on MS patients' cognitive impairment | Innapropriate intervention |
| 8525 | Effect of exergames by using Xbox 360 Kinect on cognition of older adults with mild cognitive impairment | Innapropriate intervention |
| 8526 | A Randomized Controlled Trial of Whole-Body Vibration on Gait Ability and Balance among Older Hemodialysis Patients | Inappropriate population |
| 8527 | Effect of Pre-Hospitalization Fall History on Physical Activity and Sedentary Behavior After the Implementation of a Behavioral Change Approach in Patients with Minor Ischemic Stroke: a Secondary Analysis of a Randomized Controlled Trial | Inappropriate outcome |

| | | |
|---|---|---|
| 8528 | Medium chain triglycerides with a C8: C10 ratio of 30: 70 enhances cognitive performance and mitigates the cognitive decline associated with prolonged exercise in young and healthy adults | Inappropriate study design |
| 8529 | Rowing in Los Angeles: Performance Considerations for the Change to 1500 m at the 2028 Olympic Games | Inappropriate study design |
| 8530 | Effectiveness of Cognitive Functional Therapy Versus Core Exercises and Manual Therapy in Patients with Chronic Low Back Pain after Spinal Surgery: randomized Controlled Trial | Inappropriate population |
| 8531 | Comparison of dance and aerobic exercise on cognition and neuropsychiatric symptoms in sedentary older adults with cognitive impairment | Innapropriate intervention |
| 8532 | The remote exercise SWEDEHEART study-Rationale and design of a multicenter registry-based cluster randomized crossover clinical trial (RRCT) | Inappropriate study design |
| 8533 | Study design and methods: U.S. study to protect brain health through lifestyle intervention to reduce risk (U.S. POINTER) | Inappropriate study design |
| 8534 | Exercise counters the negative impact of bed rest on executive functions in middle-aged and older adults: a proof-of-concept randomized controlled trial | Inappropriate population |
| 8535 | Sustained increase in physical fitness independently predicts improvements in cardiometabolic risk profile in type 2 diabetes | Inappropriate population |
| 8536 | Reducing Pulmonary Capillary Wedge Pressure During Exercise Exacerbates Exertional Dyspnea in Patients With Heart Failure With Preserved Ejection Fraction: implications for V˙/Q˙ Mismatch | Inappropriate study design |
| 8537 | The relationship between the brain-derived neurotrophic factor and neurocognitive response to physical exercise in individuals with schizophrenia | Innapropriate intervention |
| 8538 | Aerobic exercise improves executive functions in females, but not males, without the BDNF Val66Met polymorphism | Innapropriate intervention |
| 8539 | Load carriage physiology in normoxia and hypoxia | Inappropriate population |
| 8540 | Running red lights: Risk recognition versus follow-up implementation in the case of Germany's pandemic preparation | Inappropriate study design |
| 8541 | Effects of aerobic exercise on neurocognitive function in postmenopausal women receiving endocrine therapy for breast cancer: the Exercise Program in Cancer and Cognition randomized controlled trial | Innapropriate intervention |
| 8542 | The impact of visuospatial and executive function on activity performance and outcome after robotic or conventional gait training, long-term after stroke-as part of a randomized controlled trial | Innapropriate intervention |
| 8543 | Effects of a remotely supervised physical training program combined with cognitive training for older individuals at increased risk of clinical-functional vulnerability: study protocol for a randomized clinical trial | Inappropriate study design |
| 8544 | CARDIOPULMONARY REHABILITATION IMPROVES CARDIORESPIRATORY FITNESS IN LONG-COVID-19 PATIENTS: a RANDOMIZED CONTROLLED TRIAL | Inappropriate population |
| 8545 | Impact of exercise on brain neurochemicals: a comprehensive review | Inappropriate study design |
| 8546 | Unraveling the unparalleled benefits of orienteering versus hiking on gait performance and cognition: a randomized clinical trial | Inappropriate population |
| 8547 | Therapeutic Lifestyle Intervention Targeting Enhanced Cardiometabolic Health and Function for Persons with Chronic Spinal Cord Injury in Caregiver/Care-Receiver Co-Treatment: a Study Protocol of a Multisite Randomized Controlled Trial | Inappropriate study design |
| 8548 | A long-duration race induces a decrease of left ventricular strains, twisting mechanics and myocardial work in trained adolescents | Inappropriate study design |
| 8549 | Relative Aerobic Load of Daily Activities After Stroke | Inappropriate study design |
| 8550 | Mind body exercise improves cognitive function more than aerobic- and resistance exercise in healthy adults aged 55 years and older – an umbrella review | Inappropriate study design |
| 8551 | Improved intraindividual variability in cognitive performance following cognitive and exercise training in older adults | Inappropriate study design |
| 8552 | Multidomain cognitive training increases physical activity in people with Parkinson's disease with mild cognitive impairment | Innapropriate intervention |
| 8553 | A pilot multiple-baseline study of a mobile cognitive behavioral therapy for the treatment of eating disorders in university students | Inappropriate study design |
| 8554 | Association of Device-Measured Physical Activity With Cardiovascular Outcomes in Individuals With Hypertension | Inappropriate study design |
| 8555 | Muscle metaboreflex control of left ventricular systolic function in heart failure with reduced ejection fraction | Inappropriate study design |
| 8556 | Optimal Intensity and Duration of Walking Rehabilitation in Patients With Chronic Stroke: a Randomized Clinical Trial | Inappropriate outcome |

| | | |
|---|---|---|
| 8557 | Combining exercise with cognitive training and vitamin D3 to improve functional brain connectivity (FBC) in older adults with mild cognitive impairment (MCI). Results from the SYNERGIC trial | Innapropriate intervention |
| 8558 | Effect of aquatic exercise programs according to the International Classification of Functionality, Disability and Health domains in individuals with Parkinson's disease: a systematic review and meta-analysis with GRADE quality assessment | Inappropriate study design |
| 8559 | Effects of high-intensity respiratory muscle training on respiratory muscle strength in individuals with Parkinson's disease: protocol of a randomized clinical trial | Inappropriate study design |
| 8560 | Train Smart Study: protocol for a randomised trial investigating the role of exercise training dose on markers of brain health in sedentary middle-aged adults | Inappropriate study design |
| 8561 | Adding cognitive remediation to employment support services: A randomized controlled trial | Inappropriate outcome |
| 8562 | The role of physical activity in long-term weight loss: 36-month results from a randomized controlled trial | Inappropriate population |
| 8563 | The effect of physical activity breaks, including motor-cognitive coordination exercises, on employees' cognitive functions in the workplace | Inappropriate population |
| 8564 | Physical Activity Trajectories and Functional Recovery After Acute Stroke Among Adults in Sweden | Inappropriate study design |
| 8565 | An investigation of the effects of dual-task balance exercises on balance, functional status and dual-task performance in children with Down syndrome | Inappropriate population |
| 8566 | The Additive Benefits of Aerobic Exercise and Cognitive Training Postconcussion: Current Clinical Concepts | Inappropriate study design |
| 8567 | The Improvement of Left Atrial Function after Twelve Weeks of Supervised Concurrent Training in Patients with Heart Failure with Mid-Range Ejection Fraction: a Pilot Study | Inappropriate population |
| 8568 | The importance of AMPK in obesity and chronic diseases and the relationship of AMPK with nutrition: a literature review | Inappropriate study design |
| 8569 | Rationale and Design of the PARTNER Trial: partnered Rhythmic Rehabilitation for Enhanced Motor-Cognition in Prodromal Alzheimer's Disease | Inappropriate study design |
| 8570 | Repeated Massage Improves Swimmers' Perceptions during Training Sessions but Not Sprint and Functional Performance: a Randomized Controlled Trial | Inappropriate population |
| 8571 | Study protocol of a clinical randomized controlled trial on the efficacy of an innovative Digital thErapy to proMote wEighT loss in patients with obesity by incReasing their Adherence to treatment: the DEMETRA study | Inappropriate study design |
| 8572 | Effect of Modified Constraint-Induced Movement Therapy on Upper Extremity Function for Stroke Patients with Right/Left Arm Paresis: a Single-Blind Randomized Controlled Trial | Inappropriate population |
| 8573 | Treadmill Walking Maintains Dual-task Gait Performance and Reduces Frontopolar Cortex Activation in Healthy Adults | Inappropriate population |
| 8574 | I am fatigued of being stigmatized": On the need to investigate stigma-related barriers to physical activity" | Inappropriate study design |
| 8575 | Influences of Aerobic Exercise on Motor Sequence Learning and Corticomotor Excitability in People With Parkinson's Disease | Inappropriate population |
| 8576 | Effects of physical activity breaks during prolonged sitting on vascular and executive function—A randomised cross-over trial | Inappropriate population |
| 8577 | The effect of exercise on cognition and clinical symptoms of patients with schizophrenia: a systematic review of randomized controlled trial | Inappropriate study design |
| 8578 | Blackcurrant Supplementation and Habitual Physical Activity Enhance Functional Capacity and Quality of Life in Previously Sedentary Older Women | Inappropriate population |
| 8579 | EFFECT OF A SENSORIMOTOR EXERCISE REGIME FOR UPPER LIMB DEFICITS IN THE IN-HOSPITAL PHASE AFTER A SUPRATENTORIAL STROKE (SMULS) - A RANDOMIZED CONTROLLED TRIAL | Inappropriate population |
| 8580 | Effects of acute moderate-intensity exercise on executive function in children with preterm birth: a randomized crossover study | Inappropriate population |
| 8581 | Effects of Resistance Exercise on Cognitive Performance and Depressive Symptoms in Community-Dwelling Older Chinese Americans: a Pilot Randomized Controlled Trial | Innapropriate intervention |
| 8582 | Effects of Tai Chi Chuan on cognitive function in adults 60 years or older with type 2 diabetes and mild cognitive impairment in China: a randomized clinical trial | Innapropriate intervention |
| 8583 | Associated Factors of Time Spent Walking for Community-Dwelling Stroke Survivors | Inappropriate study design |
| 8584 | Effects of cognitive demand during acute exercise on inhibitory control and its electrophysiological indices: a randomized crossover study | Inappropriate population |

| | | |
|---|---|---|
| 8585 | Cross-education in people with Parkinson's disease, a short-term randomized controlled trial | Inappropriate outcome |
| 8586 | Concurrent validity of walking speed measured by a wearable sensor and a stopwatch during the 10-meter walk test in individuals with stroke | Inappropriate study design |
| 8587 | Effects of Different Exercise Programs on Symptoms, Sleep, and Quality of Life in Patients with Primary Restless Legs Syndrome | Inappropriate population |
| 8588 | Sex differences in hemodynamic response to high-intensity interval exercise | Inappropriate study design |
| 8589 | The Effect of Different Types of Exercise on Sleep Quality and Architecture in Parkinson Disease: A Single-Blinded Randomized Clinical Trial Protocol | Inappropriate study design |
| 8590 | Online neuropilates classes in chronic stroke patients: protocol for a randomised controlled feasibility study | Inappropriate study design |
| 8591 | D2 receptor blockade eliminates exercise-induced changes in cortical inhibition and excitation | Inappropriate study design |
| 8592 | Adaptations to Optimized Interval Training in Soccer Players: a Comparative Analysis of Standardized Methods for Individualizing Interval Interventions | Inappropriate study design |
| 8593 | Prior brain endurance training improves endurance exercise performance | Inappropriate population |
| 8594 | A dissonance-based group approach to address the internalized weight stigma in patients with obesity: theory and general organization | Inappropriate study design |
| 8595 | Acute effect of high-intensity functional training (HIFT) using a benchmark on cognition and physiological parameters according to the competitive level | Inappropriate study design |
| 8596 | Multicomponent and mat Pilates training increased gait speed in individuals with Parkinson's disease when walking and carrying a load: a single-blinded randomized controlled trial | Innapropriate intervention |
| 8597 | POST-TRAUMATIC STRESS DISORDER IN INDIVIDUALS WHO REQUIRED HOSPITALIZATION FOR COVID-19: A CROSS-SECTIONAL STUDY | Inappropriate study design |
| 8598 | Effects of inspiratory muscle training on the severity of obstructive sleep apnea in individuals after stroke: a protocol for a randomized controlled trial | Inappropriate study design |
| 8599 | Effects of a physical exercise or motor activity protocol on cognitive function, lipid profile, and BDNF levels in older adults with mild cognitive impairment | Innapropriate intervention |
| 8600 | Effects of a 1 year aerobic and strength training on cognitive functioning after transient ischemic attack or minor stroke: a randomized controlled trial | Innapropriate intervention |
| 8601 | SHAREABLE RESOURCE: Exercise for Those Living with Parkinson's Disease: A Basic Guide | Inappropriate study design |
| 8602 | Graded repetitive arm supplementary programme (GRASP) and virtual reality to improve upper limb function in patients with acute stroke-An experimental study | Inappropriate outcome |
| 8603 | Dose–response relationship between leisure-time physical activity and biomarkers of inflammation and oxidative stress in overweight/obese populations | Inappropriate study design |
| 8604 | The Effects of Different Exercise Approaches on Attention Deficit Hyperactivity Disorder in Adults: A Randomised Controlled Trial | Inappropriate population |
| 8605 | The effects of an acute bout of ergometer cycling on young adults' executive function: A systematic review and meta-analysis | Inappropriate study design |
| 8606 | The association of increased body mass index on cardiorespiratory fitness, physical activity, and cognition in adults with down syndrome | Inappropriate study design |
| 8607 | Effect of resistance exercise on cardiac perturbations and systolic performance: a cross-over randomized trial comparing volumes and techniques | Inappropriate population |
| 8608 | Essential amino acid supplements ingestion has a positive effect on executive function after moderate-intensity aerobic exercise | Inappropriate study design |
| 8609 | At-home computerized executive-function training to improve cognition and mobility in normal-hearing adults and older hearing aid users: a multi-centre, single-blinded randomized controlled trial | Inappropriate study design |
| 8610 | Factors Associated With Physical Activity in Hospitalized Patients With Dementia | Inappropriate study design |
| 8611 | Effects of three physical exercise modalities on respiratory function of older adults with Parkinson's disease: a randomized clinical trial | Inappropriate outcome |
| 8612 | High-Intensity Interval Training and Cardiometabolic Health in the General Population: A Systematic Review and Meta-Analysis of Randomised Controlled Trials | Inappropriate study design |
| 8613 | Interpreting the CPASS Trial: do Not Shift Motor Therapy to the Subacute Phase | Inappropriate study design |
| 8614 | The effect of task-oriented circuit training-based telerehabilitation on upper extremity motor functions in patients with Parkinson's disease: A randomized controlled trial | Inappropriate outcome |
| 8615 | Secondary prevention of stroke. A telehealth-delivered physical activity and diet pilot randomized trial (ENAbLE-pilot) | Inappropriate population |
| 8616 | Effects of Melatonin Supplementation and Different Exercise Models on Cognitive Function in Long-Term Exposure to Constant Light in Rats | Inappropriate study design |

| | | |
|---|---|---|
| 8617 | Identity, social engagement and community participation impact physical activity levels of stroke survivors: A mixed-methods study | Inappropriate study design |
| 8618 | Anodal tDCS over the left DLPFC but not M1 increases muscle activity and improves psychophysiological responses, cognitive function, and endurance performance in normobaric hypoxia: A randomized controlled trial | Inappropriate population |
| 8619 | A digital exercise and augmented reality training system improved mobility among stroke patients: a randomized control trial | Inappropriate outcome |
| 8620 | Multimodal Exercise and Cognitive Training Program Improves Cognitive Function in Amnestic Mild Cognitive Impairment | Innapropriate intervention |
| 8621 | High intensity interval training as a novel treatment for impaired awareness of hypoglycaemia in people with type 1 diabetes (HIT4HYPOS): a randomised parallel-group study | Inappropriate population |
| 8622 | DETERMINANTS OF INDIVIDUAL DIFFERENCES IN THE EFFICACY OF AEROBIC EXERCISE TO IMPROVE BRAIN HEALTH AND REDUCE ALZHEIMER'S DISEASE RISK IN OLDER AFRICAN AMERICANS | Inappropriate study design |
| 8623 | Cognitive prescriptions for reducing dementia risk factors among Black/African Americans: Feasibility, acceptability, and preliminary efficacy | Inappropriate population |
| 8624 | A Multi-Arm, Randomized, Blinded, Sham- Controlled Trial of Cognitive Rehabilitation and Aerobic Exercise (The CogEx trial) for cognitive impairment in people with Progressive Multiple Sclerosis | Innapropriate intervention |
| 8625 | Resistance Exercise Program in Cognitively Normal Older Adults: cERT-Based Exercise Protocol of the AGUEDA Randomized Controlled Trial | Inappropriate study design |
| 8626 | THE ACUTE EFFECT OF TDCS COMBINED WITH PHYSIOTHERAPY ON GAIT TURNING IN INDIVIDUALS WITH PARKINSON'S DISEASE: A RANDOMIZED CONTROLLED TRIAL | Inappropriate outcome |
| 8627 | Effects of Multimodal Exercise With Augmented Reality on Cognition in Community-Dwelling Older Adults | Inappropriate study design |
| 8628 | Mental Fatigue Impairs Second Serve Accuracy in Tennis Players | Inappropriate population |
| 8629 | Is postexercise hypotension a method-dependent phenomenon in chronic stroke? A crossover randomized controlled trial | Inappropriate outcome |
| 8630 | Right ventricular performance during acute hypoxic exercise | Inappropriate study design |
| 8631 | Vagus Nerve Stimulation Paired With Upper-Limb Rehabilitation After Stroke: 2- and 3-Year Follow-up From the Pilot Study | Inappropriate study design |
| 8632 | With life there is motion. Activity biomarkers signal important health and performance outcomes | Inappropriate study design |
| 8633 | Acute Evening High-Intensity Interval Training may Attenuate the Detrimental Effects of Sleep Restriction on Long-Term Declarative Memory | Inappropriate population |
| 8634 | Changes in Cognition and Brain Function After 26 Weeks of Progressive Resistance Training in Older Adults at Risk for Diabetes: a Pilot Randomized Controlled Trial | Inappropriate population |
| 8635 | Effects of power training in older patients with multiple sclerosis on neurodegeneration, neuromuscular function, and physical function. A study protocol for the “power training in older multiple sclerosis patients (PoTOMS) randomized control trial | Inappropriate study design |
| 8636 | THE RECARDIO TRIAL PROTOCOL: COMPARING THE EFFECTS OF COGNITIVE TRAINING AND PHYSICAL EXERCISE ON COGNITION AND CEREBRAL AUTOREGULATION IN MEN AND WOMEN WITH HEART FAILURE | Inappropriate study design |
| 8637 | Effect of Continuous Training versus Interval Training on Functional Parameters of Stage-1 Hypertensive Patients: a Randomised Clinical Study | Inappropriate study design |
| 8638 | Effect of a Sensorimotor Exercise Regimefor Upper Limb Deficits in the In-HospitalPhase After a Supratentorial Stroke(SMULS)—Protocol for a RandomizedControlled Trial | Inappropriate study design |
| 8639 | A comparative open label study comparing the efficacy of structured physiotherapy versus nonstructured physiotherapy in reducing poststroke spasticity related shoulder pain in patients treated with botulinum toxin A | Inappropriate study design |
| 8640 | Virtual reality to improve low-back pain and pelvic pain during pregnancy: a pilot RCT for a multicenter randomized controlled trial | Inappropriate study design |
| 8641 | Equine-assisted Therapy (EAT) in Parkinson's disease - A Pilot Study | Innapropriate intervention |
| 8642 | Effect of Training on Vascular Function and Repair in Heart Failure With Preserved Ejection Fraction | Inappropriate population |
| 8643 | Wellness intervention improves quality of life and cognition in persons with early Multiple Sclerosis | Inappropriate study design |

| | | |
|---|---|---|
| 8644 | Hemodynamic Tolerance of New Resistance Training Methods in Patients With Heart Failure and Coronary Artery Disease: a RANDOMIZED CROSSOVER STUDY | Inappropriate population |
| 8645 | Examining the efficacy of a cardio-dance intervention on brain health and the moderating role of ABCA7 in older African Americans: a protocol for a randomized controlled trial | Inappropriate study design |
| 8646 | The impact of 12 weeks of resistance exercise on hippocampus structural and neurochemical characteristics in older adults: evidence from MRI and 1H-MRS findings | Innapropriate intervention |
| 8647 | (HIIT-The Track) High-Intensity Interval Training for People with Parkinson's Disease: individual Response Patterns of (Non-)Motor Symptoms and Blood-Based Biomarkers—A Crossover Single-Case Experimental Design | Inappropriate study design |
| 8648 | The personalised integrated COgnitive-somato Sensory-MOtor training protocol to improve upper limb function after stroke: a pilot randomized controlled protocol | Inappropriate study design |
| 8649 | Exercise induced plasma volume expansion lowers cardiovascular strain during 15-km cycling time-trial in acute normobaric hypoxia | Inappropriate population |
| 8650 | Yoga, aerobic and stretching exercise effects on neurocognition: randomized controlled trial protocol | Inappropriate study design |
| 8651 | The effect of acute exercise on attentional control and theta power in young adults | Inappropriate population |
| 8652 | A Randomized Controlled Trial of Argentine Tango to Improve Cancer-Related Fatigue and Quality of Life in Breast Cancer Survivors | Inappropriate outcome |
| 8653 | Effects of multicomponent training and HMB supplementation on disability, cognitive and physical function in institutionalized older adults aged over 70 years: a cluster-randomized controlled trial | Innapropriate intervention |
| 8654 | The effects of different exercise types on cognitive and physical functions in dementia patients: a randomized comparative study | Innapropriate intervention |
| 8655 | THE IMPACT OF A SUPERVISED BRACE (BALANCE, RESISTANCE, AEROBIC, COGNITIVE EXERCISES) PROTOCOL ON THE MOBILITY OF ELDERLY INDIVIDUALS AT RISK OF FALLING | Inappropriate population |
| 8656 | A qualitative study exploring experiences of female family caregivers of people with dementia in acceptance and commitment therapy | Inappropriate study design |
| 8657 | Individualized Hospital to Home, Exercise-Nutrition Self-Managed Intervention for Pre-Frail and Frail Hospitalized Older Adults: the INDEPENDENCE Randomized Controlled Pilot Trial | Innapropriate intervention |
| 8658 | Effect of brain-computer interface based action observation combined with peripheral electrical stimulation for inducing corticospinal plasticity in healthy and stroke patients | Innapropriate intervention |
| 8659 | Exploring How Active and Insufficiently Active Individuals Respond to Specific and Non-Specific Physical Activity Goals | Inappropriate study design |
| 8660 | The Independent Walking for Brain Health Intervention for Older Adults: Protocol for a Pilot Randomized Controlled Trial | Inappropriate study design |
| 8661 | EFFECTS OF HIGH-INTENSITY TRAINING ON BASKETBALL PLAYERS | Inappropriate population |
| 8662 | Telehealth multicomponent exercise and health education in breast cancer patients undergoing primary treatment: rationale and methodological protocol for a randomized clinical trial (ABRACE: telehealth) | Inappropriate study design |
| 8663 | The role of exerkines on brain mitochondria: a mini-review | Inappropriate study design |
| 8664 | Physical activity and brain health in patients with atrial fibrillation | Inappropriate study design |
| 8665 | Effects of intermittent exercise during prolonged sitting on executive function, cerebrovascular, and psychological response: a randomized crossover trial | Inappropriate population |
| 8666 | Effects of exergame and bicycle exercise intervention on blood pressure and executive function in older adults with hypertension: a three-group randomized controlled study | Inappropriate population |
| 8667 | BLOOD PRESSURE TRAJECTORY OF INPATIENT REHABILITATION STROKE PATIENTS FROM THE DETERMINING OPTIMAL POST-STROKE EXERCISE (DOSE) TRIAL OVER THE FIRST 12-MONTHS POSTSTROKE | Inappropriate study design |
| 8668 | Effects of home-based computerized cognitive training and Tai Chi exercise on cognitive functions in older adults with mild cognitive impairment | Innapropriate intervention |
| 8669 | Association between intensity or accumulating pattern of physical activity and executive function in community-dwelling older adults: A cross-sectional study with compositional data analysis | Inappropriate study design |
| 8670 | Effect of Personalized Accelerated Pacing on Quality of Life, Physical Activity, and Atrial Fibrillation in Patients With Preclinical and Overt Heart Failure With Preserved Ejection Fraction: the myPACE Randomized Clinical Trial | Inappropriate population |

| | | |
|---|---|---|
| 8671 | A randomized controlled trial of transdermal magnesium oil for chronic low back pain management | Inappropriate population |
| 8672 | Validity and reliability of VO2-max testing in persons with Parkinson's disease | Inappropriate study design |
| 8673 | Synergistic effects of aerobic exercise and transcranial direct current stimulation on executive function and biomarkers in healthy young adults | Inappropriate population |
| 8674 | Association between healthy lifestyle and memory decline in older adults: 10 year, population based, prospective cohort study | Inappropriate study design |
| 8675 | Application of transcranial alternating current stimulation to improve eSports-related cognitive performance | Inappropriate population |
| 8676 | Influence of High-Intensity Interval Training on IGF-1 Response, Brain Executive Function, Physical Fitness and Quality of Life in Sedentary Young University Women-Protocol for a Randomized Controlled Trial | Inappropriate study design |
| 8677 | Moderate-to-high Intensity Group Exercise Improves Function of Stroke Survivors | Inappropriate outcome |
| 8678 | Relationship of Functional Outcome With Sarcopenia and Objectively Measured Physical Activity in Patients With Stroke Undergoing Rehabilitation | Inappropriate study design |
| 8679 | Efficacy and safety of a combination of emotional freedom technique with acupuncture versus acupuncture alone to treat psychiatric symptoms in Parkinson's disease: a protocol for a randomized, assessor-blind, parallel-group clinical trial | Inappropriate study design |
| 8680 | Effects of high-intensity interval training with robotassist walking exercise on cardiopulmonary function and gait ability in chronic stroke patients | Inappropriate outcome |
| 8681 | Effectiveness of the physiotherapy interventions on complex regional pain syndrome in patients with stroke: A systematic review and meta-analysis | Inappropriate study design |
| 8682 | Acute effects of mindful interval exercise on cognitive performance in a higher education setting | Inappropriate population |
| 8683 | The effect of single and dual-task balance exercises on balance performance in older adult patients with degenerative lumbar spinal stenosis: a randomized controlled trial | Inappropriate population |
| 8684 | Osteocalcin: A Multifaceted Bone-Derived Hormone | Inappropriate study design |
| 8685 | Physical-cognitive training with dietary intervention to improve cognitive function and circulating biomarkers in postmenopausal women with obesity: study protocol for a randomized controlled trial | Inappropriate study design |
| 8686 | Use of a Robotic Walking Device for Home and Community Mobility in Parkinson Disease: a Randomized Controlled Trial | Inappropriate outcome |
| 8687 | The feasibility and acceptability of a home-based dual task exercise and cognition intervention after stroke | Innapropriate intervention |
| 8688 | THE EFFECT OF AEROBIC FITNESS TRAINING ON THE COURSE OF CEREBROVASCULAR RISK FACTORS OVER SIX MONTHS AFTER SUBACUTE STROKE - RESULTS FROM THE PHYS-STROKE STUDY | Inappropriate outcome |
| 8689 | EFFECT OF AEROBIC FITNESS TRAINING ON PERIPHERAL ENDOTHELIAL FUNCTION IN SUBACUTE STROKE PATIENTS - RESULTS FROM THE PHYS-STROKE STUDY | Inappropriate outcome |
| 8690 | Cognitive functional therapy with or without movement sensor biofeedback versus usual care for chronic, disabling low back pain (RESTORE): A randomised, controlled, three-arm, parallel group, phase 3, clinical trial | Inappropriate population |
| 8691 | Optimizing sprint interval exercise for post-exercise hypotension: A randomized crossover trial | Inappropriate population |
| 8692 | Cognitive and biomarker responses in healthy older adults to a 18-hole golf round and different walking types: a randomised cross-over study | Inappropriate population |
| 8693 | Effects of an acute bout of cycling on different domains of cognitive function | Inappropriate population |
| 8694 | Effects of Physical Exercise Program for Older Family Caregivers of Persons With Dementia | Inappropriate study design |
| 8695 | Effects of high-intensity interval training and moderate-intensity continuous training on sarcopenia-related parameters in participants with Parkinson's disease: a 24-week randomized pilot trial substudy | Inappropriate outcome |
| 8696 | Long-term environmental enrichment is associated with better fornix microstructure in older adults | Inappropriate study design |
| 8697 | Effect of physical exercise on the hippocampus and global grey matter volume in breast cancer patients: a randomized controlled trial (PAM study) | Innapropriate intervention |
| 8698 | Effect of Physical Exercise on MRI-Assessed Brain Perfusion in Chemotherapy-Treated Breast Cancer Patients: a Randomized Controlled Trial | Innapropriate intervention |
| 8699 | Investigation of the Protective Effect of Treadmill Exercise on Molecular Pathways and Cognitive Behaviors in Alzheimer Disease Model | Inappropriate study design |

| | | |
|---|---|---|
| 8700 | Isometric Resistance Training Is Recommended as a Safe and Effective Adjunct Therapy to Reduce Blood Pressure | Inappropriate study design |
| 8701 | High-intensity training in patients with lacunar stroke: a one-year follow-up | Inappropriate outcome |
| 8702 | Moderating effects of gender, age and cognitive reserve on combined cognitive training and transcranial electrical stimulation | Inappropriate population |
| 8703 | Systematic balance exercise affects the level of Sirt1 and Sirt3 in older adults and persons with Parkinson's disease - pilot study | Inappropriate outcome |
| 8704 | The effects of an exercise intervention on executive function among overweight adults with obstructive sleep apnea | Inappropriate study design |
| 8705 | Association of Malnutrition With Physical Activity Intensity in Patients With Subacute Stroke | Inappropriate study design |
| 8706 | Dry Needling and Neurodevelopmental Therapy versus Neurodevelopmental Therapy Alone on Spasticity and Functions in Patients with Stroke: a Randomized Controlled Trial | Inappropriate outcome |
| 8707 | AEROBIC EXERCISE TRAINING IN ACUTE STROKE: a FEASIBILITY STUDY | Innapropriate intervention |
| 8708 | Beta-band MEG signal power changes in older adults after physical exercise program with and without additional cognitive training | Inappropriate population |
| 8709 | The Japan-Multimodal Intervention Trial for Prevention of Dementia PRIME Tamba (J-MINT PRIME Tamba): Study protocol of a randomised controlled multi-domain intervention trial | Inappropriate study design |
| 8710 | Internet of Things (IoT) Enables Robot-Assisted Therapy as a Home Program for Training Upper Limb Functions in Chronic Stroke: A Randomized Control Crossover Study | Inappropriate outcome |
| 8711 | Improvement of the motor and functional capacity of post-cerebrovascular accident hemiplegics by exercise retraining combined with a varied exercises program | Innapropriate intervention |
| 8712 | Acceptability of a digital health application to empower persons with multiple sclerosis with moderate to severe disability: single-arm prospective pilot study | Inappropriate study design |
| 8713 | A randomized clinical trial of mindfulness meditation versus exercise in Parkinson’s disease during social unrest | Innapropriate intervention |
| 8714 | Stroke volume response during prolonged exercise depends on left ventricular filling: evidence from a β-blockade study | Inappropriate population |
| 8715 | Caring for Informal Dementia Caregivers and Their Loved Ones Via the HOMeCARE Exercise and Mindfulness for Health Program (HOMeCARE): a Randomized, Single-Blind, Controlled Trial | Inappropriate outcome |
| 8716 | Climb up! Head up! Climbing improves posture in Parkinson's disease. A secondary analysis from a randomized controlled trial | Inappropriate outcome |
| 8717 | ADDITION OF HIGH INTENSITY INTERVAL TRAINING TO A MODERATE INTENSITY CONTINUOUS TRAINING CARDIOVASCULAR REHABILITATION PROGRAM AFTER ISCHEMIC CEREBROVASCULAR DISEASE: a RANDOMIZED CONTROLLED TRIAL | Inappropriate population |
| 8718 | Does intensity matter? A randomized crossover study of the role of acute exercise intensity on cognitive performance and motor speed and accuracy | Inappropriate population |
| 8719 | Intensity-dependent cardiopulmonary response during and after strength training | Inappropriate population |
| 8720 | Exercise as a strategy to mitigate the cognitive effects of partial sleep deprivation | Inappropriate population |
| 8721 | Treatment engagement in first-episode schizophrenia: associations between intrinsic motivation and attendance during cognitive training and an aerobic exercise program | Inappropriate outcome |
| 8722 | Changes in surrogate markers of intestinal epithelial injury and microbial translocation in young and older men during prolonged occupational heat stress in temperate and hot conditions | Inappropriate study design |
| 8723 | Effects of the home-based exercise program with an augmented reality system on balance in patients with stroke: A randomized controlled trial | Inappropriate outcome |
| 8724 | A non-pharmacological multidomain intervention of dual-task exercise and social activity affects the cognitive function in community-dwelling older adults with mild to moderate cognitive decline: a randomized controlled trial | Innapropriate intervention |
| 8725 | Therapeutic Singing and Semi-Occluded Vocal Tract Exercises for Individuals with Parkinson's Disease: a Randomized Controlled Trial of a Single Session Intervention | Inappropriate outcome |
| 8726 | A bibliometric analysis of physical activity interventions and cognition in children and adolescents | Inappropriate study design |
| 8727 | Exercise interveNtion outdoor proJect in the cOmmunitY -- results from the ENJOY program for independence in dementia: a feasibility pilot randomised controlled trial | Innapropriate intervention |

| | | |
|---|---|---|
| 8728 | Clinical Effectiveness of Cognitively Enhanced Tai Ji Quan Training on Global Cognition and Dual-Task Performance During Walking in Older Adults With Mild Cognitive Impairment or Self-Reported Memory Concerns : a Randomized Controlled Trial | Innapropriate intervention |
| 8729 | Acute aerobic exercise benefits allocation of neural resources related to selective attention | Inappropriate population |
| 8730 | Neurogranin as an important regulator in swimming training to improve the spatial memory dysfunction of mice with chronic cerebral hypoperfusion | Inappropriate population |
| 8731 | The Utilization of Forced-Rate Cycling to Facilitate Motor Recovery Following Stroke: a Randomized Clinical Trial | Innapropriate intervention |
| 8732 | Increased Comfortable Gait Speed Is Associated With Improved Gait Biomechanics in Persons With Chronic Stroke Completing an 8-Week Forced-Rate Aerobic Cycling Intervention: A Preliminary Study | Inappropriate study design |
| 8733 | An 8-week Forced-rate Aerobic Cycling Program Improves Cardiorespiratory Fitness in Persons With Chronic Stroke: a Randomized Controlled Trial | Inappropriate outcome |
| 8734 | Exercise-Induced Left Atrial Hypertension in Heart Failure With Preserved Ejection Fraction | Inappropriate population |
| 8735 | Runners with better cardiorespiratory fitness had higher prefrontal cortex activity during both single and exercise-executive function dual tasks: an fNIRS study | Inappropriate study design |
| 8736 | The joint effect and hemodynamic mechanism of PA and PM2.5 exposure on cognitive function: a randomized controlled trial study | Inappropriate population |
| 8737 | Effects of Cognitive Training in Flanker Task on Mobility of Older Adults: a Randomized Controlled Trail | Inappropriate population |
| 8738 | The effects of a 15-week physical exercise intervention on pain modulation in fibromyalgia: increased pain-related processing within the cortico-striatal- occipital networks, but no improvement of exercise-induced hypoalgesia | Inappropriate study design |
| 8739 | Trained athletes and cognitive function: a systematic review and meta-analysis | Inappropriate study design |
| 8740 | The complex associations between adiposity, fitness, mental wellbeing and neurocognitive function after exercise: a randomized crossover trial in preadolescent children | Inappropriate population |
| 8741 | Exercise and epigenetic ages in older adults with myeloid malignancies | Inappropriate study design |
| 8742 | EXPRESS: lack of Effects of Acute Exercise Intensity on Mnemonic Discrimination | Inappropriate study design |
| 8743 | Application of cross-migration theory in limb rehabilitation of stroke patients with hemiplegiaz | Inappropriate outcome |
| 8744 | Effects of Cognitive and Physical Load of Acute Exercise on Inhibitory Control and Prefrontal Cortex Hemodynamics in Children | Inappropriate population |
| 8745 | Neurorehabilitation of the upper extremity – immersive virtual reality vs. electromechanically assisted training. A comparative study | Inappropriate study design |
| 8746 | Physical Activity Behavior and Its Association With Global Cognitive Function Three Months After Stroke: The Nor-COAST Study† | Inappropriate study design |
| 8747 | Effect of a monitored exercise protocol in cerebral and hepatic hemodynamics in patients with cirrhosis and portal hypertension | Inappropriate population |
| 8748 | Physiological Variables that Contribute to Aerobic Fitness in Boys during Early Adolescence in the Context of Basketball Training and the Maturity Level | Inappropriate study design |
| 8749 | Aerobic physical exercise versus dual-task cognitive walking in cognitive rehabilitation of people with stroke: a randomized clinical trial | Innapropriate intervention |
| 8750 | Walking endurance with the Nordic walking modality in people with hemiparesis due to stroke | Inappropriate study design |
| 8751 | Investigating the impact of physical activity on mitochondrial function in Parkinson's disease (PARKEX): study protocol for A randomized controlled clinical trial | Inappropriate study design |
| 8752 | Effects of moderate-intensity aerobic exercise on serum BDNF and motor learning in the upper-limb in patients after chronic-stroke: a randomized, controlled feasibility study with embedded health economic evaluation | Innapropriate intervention |
| 8753 | Design and methods of a randomized telehealth-based intervention to improve fitness in survivors of childhood Cancer with exercise intolerance | Inappropriate population |
| 8754 | Arm swing deviations in patients with Parkinson's disease at different gait velocities | Inappropriate study design |
| 8755 | Long-term effects of a 6-month brisk walking and balance program on physical performance and health-related quality of life in people with Parkinson disease: a randomized controlled trial | Innapropriate intervention |
| 8756 | Physiological Responses to Exercise in Hypoxia in Preterm Adults: Convective and Diffusive Limitations in the O 2 Transport | Inappropriate study design |

| | | |
|---|---|---|
| 8757 | Feasibility of a Modified Otago Exercise Program for Older Adults With Cognitive Vulnerability | Innapropriate intervention |
| 8758 | Water-based therapeutic exercise in stroke: a scoping review | Inappropriate study design |
| 8759 | Blood flow restriction training activates the muscle metaboreflex during lowintensity sustained exercise | Inappropriate study design |
| 8760 | Feasibility, usability, and acceptance of "Brain-IT"—A newly developed exergame-based training concept for the secondary prevention of mild neurocognitive disorder: a pilot randomized controlled trial | Inappropriate study design |
| 8761 | The effects of nordic walking, high-intensity interval training, and moderate-to-vigorous intensity continuous training on arterial stiffness in patients with coronary artery disease | Inappropriate population |
| 8762 | Effect of an Artificial Intelligence-Based Self-Management App on Musculoskeletal Health in Patients With Neck and/or Low Back Pain Referred to Specialist Care: a Randomized Clinical Trial | Inappropriate population |
| 8763 | Cycling: how can we activate care-dependent older adults with a mild cognitive impairment? | Inappropriate outcome |
| 8764 | Acute interleukin-6 modulates key enzymes involved in prefrontal cortex and hippocampal amyloid precursor protein processing | Inappropriate study design |
| 8765 | Distinct Impact of Noncardiac Comorbidities on Exercise Capacity and Functional Status in Chronic Heart Failure | Inappropriate study design |
| 8766 | Could a Tailored Exercise Intervention for Hospitalised Older Adults Have a Role in the Resolution of Delirium? Secondary Analysis of a Randomised Clinical Trial | Inappropriate outcome |
| 8767 | Tailored Prevention of Functional Decline through a Multicomponent Exercise Program in Hospitalized Oncogeriatric Patients: study Protocol for a Randomized Clinical Trial | Inappropriate study design |
| 8768 | Effect of High-Intensity Interval Training and Moderate-Intensity Continuous Training in People With Poststroke Gait Dysfunction: a Randomized Clinical Trial | Inappropriate outcome |
| 8769 | Cognitive behavioural techniques reduce exercise anxiety and improve adherence to a resistance training program for people with anxiety-related disorders: a randomized controlled trial | Inappropriate population |
| 8770 | Multimodal physical training combined with tDCS improves physical fitness components in people after stroke: a double-blind randomized controlled trial | Inappropriate outcome |
| 8771 | Effects of Functional Electrical Stimulation Cycling Combined With Arm Cranking Exercise on Cardiorespiratory Fitness in People With Central Nervous System Disorders: A Systematic Review and Meta-analysis | Inappropriate study design |
| 8772 | Influence of Supervised Maternal Aerobic Exercise during Pregnancy on 1-Month-Old Neonatal Cardiac Function and Outflow: A Pilot Study | Inappropriate population |
| 8773 | Inpatient stroke survivors with low gait functioning benefit from music interventions during cardiorespiratory exercise: A randomized cross-over trial | Innapropriate intervention |
| 8774 | A combined exercise and cognitive training intervention induces fronto-cingulate cortical plasticity in first-episode psychosis patients | Innapropriate intervention |
| 8775 | Exercising with a robotic exoskeleton can improve memory and gait in people with Parkinson's disease by facilitating progressive exercise intensity | Innapropriate intervention |
| 8776 | The feasibility of a multidomain dementia risk reduction randomised controlled trial for people experiencing cognitive decline: The Body, Brain, Life for Cognitive Decline (BBL-CD) | Innapropriate intervention |
| 8777 | Mobilisation in the EveNing to TreAt deLirium (MENTAL): protocol for a mixed-methods feasibility randomised controlled trial | Inappropriate study design |
| 8778 | Exercise effects on motor function, manual dexterity, and brain oscillatory activity in individuals with Parkinson's disease: randomized controlled trial protocol | Inappropriate study design |
| 8779 | The Effects of Active Breaks on Primary School Students' Attentional Processes and Motivational Regulation | Inappropriate study design |
| 8780 | Cognitive work on a walking desk does not lead to compensatory appetitive responses in healthy young adults | Inappropriate population |
| 8781 | Mixed circuit training acutely reduces arterial stiffness in patients with chronic stroke: a crossover randomized controlled trial | Inappropriate outcome |
| 8782 | Immediate Effects of High-Cadence Cycling on Core Outcomes in Individuals With Parkinson's Disease | Inappropriate study design |
| 8783 | The effects of acute exercise and a nap on heart rate variability and memory in young sedentary adults | Inappropriate study design |
| 8784 | The Effect of Wii Training on Neurocognitive Function in Athletes with Functional Ankle Instability: matched Randomized Clinical Trial | Inappropriate population |

| | | |
|---|---|---|
| 8785 | Do they align? Congruence between patient preferences of people living with cognitive impairments and physicians' judgements for person-centered care: An analytic hierarchy process study | Inappropriate study design |
| 8786 | Voice improvement following conventional speech therapy combined with singing intervention in people with Parkinson's disease: a three-arm randomised controlled trial | Inappropriate outcome |
| 8787 | Effects of exergaming on cognition, lower limb functional coordination, and stepping time in people with multiple sclerosis: A randomized controlled trial | Inappropriate population |
| 8788 | Effects of a laboratory-based aerobic exercise intervention on brain volume and cardiovascular health markers: protocol for a randomised clinical trial | Inappropriate study design |
| 8789 | Physiological and cognitive responses to hyperoxic exercise in full water submersion | Inappropriate study design |
| 8790 | Is high intensity Pilates exercise treatment beneficial for people with Parkinson's disease? | Inappropriate outcome |
| 8791 | Self-reported physical activity is associated with cardiorespiratory fitness but not walking capacity among community-dwelling individuals poststroke | Inappropriate study design |
| 8792 | Effects of exercise alone or combined with cognitive training and vitamin D supplementation to improve cognition in adults with mild cognitive impairment: a randomized clinical trial [with consumer summary] | Innapropriate intervention |
| 8793 | A combined heart rate variability biofeedback and progressive aerobic exercise intervention for remediating persistent post-concussive symptoms | Inappropriate population |
| 8794 | ACUTE EFFECTS OF MODERATE-INTENSITY CONTINUOUS PHYSICAL EXERCISE PERFORMED WITH DIFFERENT AMOUNTS OF MUSCLE MASS ON EXECUTIVE FUNCTION IN HEALTHY YOUNG ADULTS: a RANDOMIZED TRIAL | Inappropriate population |
| 8795 | The Effects of Polyphenol Supplementation on BDNF, Cytokines and Cognition in Trained Male Cyclists following Acute Ozone Exposure during High-Intensity Cycling | Inappropriate study design |
| 8796 | Targeted exercise for African-Americans with multiple sclerosis: project TEAAMS | Inappropriate study design |
| 8797 | Effects of Resistive Diaphragmatic Training on the Pulmonary Functions in patients with Chronic Stroke | Inappropriate outcome |
| 8798 | The Effects of Physical Activity on Cognitive Function in Older Adults: Evidence From Randomized Controlled Trials | Inappropriate study design |
| 8799 | Effects of instability resistance training on static postural control of cognitively impaired older adults | Inappropriate outcome |
| 8800 | What baseline and intervention characteristics predict walking speed six months after stroke? | Inappropriate outcome |
| 8801 | Physical activity and relationship to physical function, quality of life, and cognitive function in older patients with acute decompensated heart failure | Inappropriate study design |
| 8802 | The effect of acute exercise on cognitive and motor inhibition - Does fitness moderate this effect? | Inappropriate population |
| 8803 | The impact of a single in-person dance class on social connectedness, mood, and stress post-stroke | Inappropriate outcome |
| 8804 | A Multidomain Intervention Program for Older People with Dementia: a Pilot Study | Inappropriate study design |
| 8805 | The PROMOTe study: prebiotic supplementation improves cognition versus placebo in healthy older twins | Inappropriate population |
| 8806 | Compositional association of 24-h movement behavior with incident major adverse cardiac events and all-cause mortality | Inappropriate study design |
| 8807 | Effectiveness of a multicomponent exercise intervention in community-dwelling older Chinese people with cognitive frailty: protocol for a mixed-methods research | Inappropriate study design |
| 8808 | Exercise program to reduce the risk of cognitive decline and physical frailty in older adults: study protocol for an open label double-arm clinical trial | Inappropriate study design |
| 8809 | What do we know about the effects of physical exercise on dementia, without alzheimer's? systematic review of international clinical trials | Inappropriate study design |
| 8810 | Exercised blood plasma promotes hippocampal neurogenesis in the Alzheimer's disease rat brain | Inappropriate study design |
| 8811 | Effect of different types of aerobic training on peak VO2 and ejection fraction for diastolic heart failure patients; a comparative randomized control trial | Inappropriate population |
| 8812 | The Effectiveness of a Multimodal Brain Empowerment Program in Mild Cognitive Impairment: a Single-Blind, Quasi-Randomized Experimental Study | Inappropriate study design |

| | | |
|---|---|---|
| 8813 | EFFECTS OF OVERGROUND WALKING EXERCISE AND COGNITIVE REHABILITATION ON BRAIN-DERIVED NEUROTROPHIC FACTOR, COGNITION AND QUALITY OF LIFE AMONG STROKE SURVIVORS IN KANO, NIGERIA | Innapropriate intervention |
| 8814 | Interventions to Change Movement Behaviors After Stroke: A Systematic Review and Meta-analysis | Inappropriate study design |
| 8815 | Effects of scapulo-humeral training on ultrasonographic and clinical evaluations in stroke: a randomized controlled trial | Inappropriate outcome |
| 8816 | THE EFFECTS OF YOGA AND AEROBIC EXERCISE ON PAIN, FUNCTION, METABOLIC CAPACITY, QUALITY OF LIFE AND COGNITIVE VARIABLES IN INDIVIDUALS WITH CHRONIC LOW BACK PAIN: PRELIMINARY RESULTS | Inappropriate population |
| 8817 | Effects of scapular exercises on trunk control ın patients with acute stroke: a double-blind randomized controlled study | Inappropriate outcome |
| 8818 | Effects of multi-task training on motor and cognitive performances in multiple sclerosis patients without clinical disability: a single-blinded randomized controlled trial | Innapropriate intervention |
| 8819 | Effect of mHealth-based relaxation program on stress coping and anxiety levels in mothers of children with cancer: a randomized controlled study | Inappropriate population |
| 8820 | Efficacy of hippotherapy simulator exercise program in patients with stroke: a randomized single-blind clinical trial | Inappropriate outcome |
| 8821 | PULSE OXIMETRY SELF-MONITORING DURING EXERCISE ENHANCES PERCEIVED SAFETY, AUTONOMY, AND USABILITY IN IDIOPATHIC PULMONARY FIBROSIS | Inappropriate study design |
| 8822 | Safety and effectiveness of standardized exercise training in patients with pulmonary hypertension associated with heart failure with preserved ejection fraction (TRAIN-HFpEF-PH): study protocol for a randomized controlled multicenter trial | Inappropriate study design |
| 8823 | The Effect of DeskCycle Use in a College Classroom | Inappropriate study design |
| 8824 | THE EFFECTIVENESS OF MULTIMODAL PROGRAM BRAIN FIT | Inappropriate study design |
| 8825 | Efficacy of virtually-delivered Qigong/Tai Chi for depression in middle- and older-age adults with bipolar disorder (QT-BD): a pilot randomized controlled trial during the COVID-19 pandemic | Inappropriate population |
| 8826 | Low-intensity resistance exercise with blood flow restriction for patients with claudication: a randomized controlled feasibility trial | Inappropriate population |
| 8827 | Soccer heading immediately alters brain function and brain-muscle communication | Inappropriate study design |
| 8828 | Active passive trainer and bilateral upper limb exercise improve outcomes in sub-acute stroke patients: a pilot study | Inappropriate outcome |
| 8829 | Effects of inpatient energy management education and high-intensity interval training on health-related quality of life in persons with multiple sclerosis: a randomized controlled superiority trial with six-month follow-up | Inappropriate outcome |
| 8830 | Effects of Exercise Training on Cognitive Function in Individuals With Heart Failure: A Meta-Analysis | Inappropriate study design |
| 8831 | The Effect of Supervised Early Resistance Training Versus Aerobic-Based Rehabilitation on Cognitive Recovery Following Cardiac Surgery Via Median Sternotomy (SECRET): a Pilot Randomised Controlled Trial | Inappropriate population |
| 8832 | An Equation With Clinical Applicability and Adequate Validity to Predict the Maximum Oxygen Consumption of Individuals Post-stroke | Inappropriate study design |
| 8833 | Impact of multidomain preventive strategies on functional brain connectivity in older adults with cognitive complaint: Subset from the Montpellier center of the ancillary MAPT-MRI study | Inappropriate study design |
| 8834 | Effects of chronic physical activity on cognition across the lifespan: a systematic meta-review of randomized controlled trials and realist synthesis of contextualized mechanisms | Inappropriate study design |
| 8835 | Sex moderates the associations between physical activity intensity and attentional control in older adolescents | Inappropriate study design |
| 8836 | The Impact of Acute Mild Normobaric Hypoxia and a Single Bout of Exercise to Volitional Exhaustion on Cognitive Performance in Endurance and Strength-Trained Athletes: The role of BDNF, EP-1, Catecholamines and Lactate | Inappropriate study design |
| 8837 | Implementation of a Falls Prevention Program in Guyana: an International Collaboration to Improve Balance Confidence | Inappropriate outcome |
| 8838 | Comparative Efficacy of High-Intensity Training Versus Conventional Training in Individuals With Chronic Traumatic Brain Injury: a Pilot Randomized Controlled Study | Inappropriate outcome |

| | | |
|---|---|---|
| 8839 | mHealth to Support Outdoor Gym Resistance Training: the ecofit Effectiveness RCT | Inappropriate population |
| 8840 | The PD-Ballet study: study protocol for a randomised controlled single-blind hybrid type 2 clinical trial evaluating the effects of ballet dancing on motor and non-motor symptoms in Parkinson's disease | Inappropriate study design |
| 8841 | "InMotion"—Mixed physical exercise program with creative movement as an intervention for adults with schizophrenia: study protocol for a randomized controlled trial | Inappropriate study design |
| 8842 | Habitual walking speed and fatigue explain self-reported functional capacity after stroke | Inappropriate study design |
| 8843 | Clinical and neurophysiologic effects of upper limb robot-assisted rehabilitation on motor recovery in patients with subacute stroke: preliminary results | Inappropriate population |
| 8844 | Neurophysiological and Clinical Effects of Upper Limb Robot-Assisted Rehabilitation on Motor Recovery in Patients with Subacute Stroke: a Multicenter Randomized Controlled Trial Study Protocol | Inappropriate study design |
| 8845 | Immersive virtual reality exergame for patients with Parkinson's disease: feasibility and potential benefits | Inappropriate study design |
| 8846 | Increased neural differentiation after a single session of aerobic exercise in older adults | Inappropriate population |
| 8847 | Positive effects of brisk walking and Tai Chi on cognitive function in older adults: an fNIRS study | Inappropriate population |
| 8848 | Metabolic, cognitive and neuromuscular responses to different multidirectional agility-like sprint protocols in elite female soccer players – a randomised crossover study | Inappropriate population |
| 8849 | Safety and feasibility of split-belt treadmill virtual-reality rehabilitation in sub acute stroke patients | Inappropriate study design |
| 8850 | HomeStyles-2 Nutrition Education Curriculum Positively Impacts Physical Activity Behaviors of SNAP-Eligible Adults | Inappropriate population |
| 8851 | Hippocampal neurochemicals are associated with exercise group and intensity, psychological health, and general cognition in older adults | Inappropriate population |
| 8852 | Consequences of sarcolemma fatigue on maximal muscle strength production in patients with myalgic encephalomyelitis/chronic fatigue syndrome | Inappropriate study design |
| 8853 | Perturbation-based balance training leads to improved reactive postural responses in individuals with Parkinson's disease and freezing of gait | Inappropriate outcome |
| 8854 | Exercise Therapy in Early Multiple Sclerosis Improves Physical Function But Not Cognition: Secondary Analyses From a Randomized Controlled Trial | Innapropriate intervention |
| 8855 | Assisted Cycle Therapy (ACT) Improved Self-Efficacy and Exercise Perception in Middle-Age Adults with Down Syndrome | Inappropriate study design |
| 8856 | Effects of high-intensity functional training (HIFT) on the functional capacity, frailty, and physical condition of older adults with mild cognitive impairment: a blind randomized controlled clinical trial | Inappropriate outcome |
| 8857 | Enhancing cognition in older adults with mild cognitive impairment through high-intensity functional training: a single-blind randomized controlled trial | Innapropriate intervention |
| 8858 | Return-to-Competition Progression After Exertional Heat Stroke in an Adolescent Runner: A Case Report | Inappropriate study design |
| 8859 | Effects of Resistance Training on Academic Outcomes in School-Aged Youth: A Systematic Review and Meta-Analysis | Inappropriate study design |
| 8860 | Effects of cognitive rehabilitation on brain grey matter volume in progressive multiple sclerosis: results from the CogEx study | Innapropriate intervention |
| 8861 | DOES INTENSITY MATTER IN PSYCHOSOCIAL RESPONSES TO EXERCISE IN CHRONIC STROKE? | Innapropriate intervention |
| 8862 | Changes in cardiovascular health and white matter integrity with aerobic exercise, cognitive and combined training in physically inactive healthy late-middle-aged adults: the Projecte Moviment" randomized controlled trial" | Inappropriate population |
| 8863 | Use of a Home-Based, Commercial Exercise Platform to Remotely Monitor Aerobic Exercise Adherence and Intensity in People With Parkinson Disease | Inappropriate outcome |
| 8864 | Feasibility of performing a multi-arm clinical trial examining the novel combination of repetitive transcranial magnetic stimulation and aerobic exercise for post-stroke depression | Inappropriate population |
| 8865 | Effect of an intensive lifestyle intervention on the structural and functional substrate for atrial fibrillation in people with metabolic syndrome | Inappropriate population |
| 8866 | Lifestyle intervention of food and exercise for lymphoma survivors (LIFE-L): a pilot intervention protocol for adults | Innapropriate intervention |
| 8867 | FEASIBILITY OF A SELF-MANAGEMENT INTERVENTION FOR IMPROVING MOBILITY FOR PATIENTS FOLLOWING STROKE IN THE COMMUNITY | Inappropriate outcome |

| | | |
|---|---|---|
| 8868 | Assessment of memory recognition using a smartphone-based test system: a pilot study | Inappropriate population |
| 8869 | TeleRehabilitation with Aims to Improve Lower extremity recovery in community-dwelling individuals who have had a stroke: protocol for a multisite, parallel group, assessor-blinded, randomised attention-controlled trial | Inappropriate study design |
| 8870 | rTMS for post-covid-19 condition: a sham-controlled case series involving iTBS-300 and iTBS-600 | Inappropriate study design |
| 8871 | The Effects of Exergame Telerehabilitation in Persons with Subjective Cognitive Decline | Innapropriate intervention |
| 8872 | Randomized, double-blind, sham-controlled study to evaluate effectiveness of photobiomodulation plus exercise, to enhance motor, cognition and quality of life in those with Parkinson's disease | Inappropriate study design |
| 8873 | Effects of plyometric jump training on measures of physical fitness and lower-limb asymmetries in prepubertal male soccer players: a randomized controlled trial | Inappropriate population |
| 8874 | The Effects Of Resistance And Balance Exercise Program In Combination With Beta-Hydroxy-Methyl Butyrate In The Older Adult With Sarcopenia: preliminary Results Of A Double-Blind, Randomized, Placebo-Controlled Trial | Inappropriate population |
| 8875 | The effects of cognitive rehabilitation combined with aerobic exercise or stretching-and-toning on new learning and memory in persons with moderate-to-severe TBI: protocol for a randomized controlled trial | Inappropriate study design |
| 8876 | Feasibility of remotely delivered and supported aerobic walking exercise training for cognitive processing speed impairment in fully-ambulatory persons with multiple sclerosis | Innapropriate intervention |
| 8877 | Effectiveness of neurobic exercise program on memory performance in community-dwelling older adults with mild cognitive impairment: a randomized controlled crossover trial | Innapropriate intervention |
| 8878 | SERCA2a Agonist Effects on Cardiac Performance During Exercise in Heart Failure With Preserved Ejection Fraction | Inappropriate study design |
| 8879 | A Prospective Randomised Control Study to Check Best Time to Start Pulmonary Rehabilitation in Intensive Care Unit at a Tertiary Referral Centre | Inappropriate population |
| 8880 | Benefits of a Dual-Task Training on Motor and Cognitive Functions in Community-Dwelling Older Adults: a Controlled Clinical Trial | Inappropriate study design |
| 8881 | Feasibility and cost description of highly intensive rehabilitation involving new technologies in patients with post-acute stroke-A trial of the Swiss RehabTech Initiative | Inappropriate study design |
| 8882 | Relief of post-stroke spasticity with acute vibrotactile stimulation: controlled crossover study of muscle and skin stimulus methods | Inappropriate population |
| 8883 | The impact of aerobic endurance training on cognitive performance in schizophrenic inpatients in a clinical routine setting | Innapropriate intervention |
| 8884 | Effectiveness of an online multicomponent program (FATIGUEWALK) for chronic fatigue syndrome: a randomized controlled trial | Inappropriate outcome |
| 8885 | The influence of baseline sleep on exercise-induced cognitive change in cognitively unimpaired older adults: a randomised clinical trial | Inappropriate population |
| 8886 | Non-therapeutic strategies to promote physical activity of stroke survivors in hospital settings: A systematic review | Inappropriate study design |
| 8887 | Effects of Virtual Reality–Based Exercise on Balance in Patients With Stroke: A Systematic Review and Meta-analysis | Inappropriate study design |
| 8888 | Home-based mobile-guided exercise-based cardiac rehabilitation among patients undergoing transcatheter aortic valve replacement (REHAB-TAVR): protocol for a randomised clinical trial | Inappropriate study design |
| 8889 | Effects of remote combine exercise-music training on physical and cognitive performance in patients with Alzheimer’s disease: A randomized controlled trial | Innapropriate intervention |
| 8890 | Balance telerehabilitation and wearable technology for people with Parkinson's disease (TelePD trial) | Inappropriate study design |
| 8891 | Acute bouts of aerobic and resistance exercise similarly alter inhibitory control and response time while inversely modifying plasma BDNF concentrations in middle-aged and older adults with type 2 diabetes | Inappropriate study design |
| 8892 | Effects of Controlled Supramaximal High-Intensity Interval Training on Cardiorespiratory Fitness and Global Cognitive Function in Older Adults: the Umeå HIT Study-A Randomized Controlled Trial | Inappropriate population |
| 8893 | Home-Based Buddhist Walking Meditation Mitigates Cardiotoxicity of Anthracycline Chemotherapy in Breast Cancer Patients: a Randomized Controlled Trial | Inappropriate population |

| | | |
|---|---|---|
| 8894 | Factors That Influence Meeting the Recommended Weekly Physical Activity Target Among Older People With Physical Multimorbidity: Evidence From 6 Low- and Middle-Income Countries | Inappropriate study design |
| 8895 | Getting Recovery Right After Neck Dissection (GRRAND-F): mixed-methods feasibility study to design a pragmatic randomised controlled trial | Inappropriate population |
| 8896 | Active Gains in brain Using Exercise During Aging (AGUEDA): protocol for a randomized controlled trial | Inappropriate study design |
| 8897 | Effect of an Aerobic Dancing Program on Sleep Quality for Older Adults With Mild Cognitive Impairment and Poor Sleep: a Randomized Controlled Trial | Innapropriate intervention |
| 8898 | Adaptive yoga versus low-impact exercise for adults with chronic acquired brain injury: a pilot randomized control trial protocol | Inappropriate study design |
| 8899 | EXERTION: a pilot trial on the effect of aerobic, smartwatch-controlled exercise on stroke recovery: effects on motor function, structural repair, cognition, mental well-being, and the immune system | Innapropriate intervention |
| 8900 | Effect of a customized safety harness on movement kinematics in a dance-based exergaming among people with chronic stroke | Inappropriate study design |
| 8901 | Effects of resistance exercise in elderly cancer patients | Inappropriate population |
| 8902 | Replacement of leisure-time sedentary behavior with various physical activities and the risk of dementia incidence and mortality: A prospective cohort study | Inappropriate study design |
| 8903 | Bobath vs Task-Oriented taining after stroke: An assessor-blind randomized controlled trial | Inappropriate population |
| 8904 | Optimized Home Rehabilitation Technology Reduces Upper Extremity Impairment Compared to a Conventional Home Exercise Program: a Randomized, Controlled, Single-Blind Trial in Subacute Stroke | Inappropriate population |
| 8905 | Efficacy of exercises in early-stage Parkinson's disease (PARK-EASE trial): single-blind, randomised, controlled trial | Inappropriate outcome |
| 8906 | The feasibility of a stepping exergame prototype for older adults with major neurocognitive disorder residing in a long-term care facility: A mixed methods pilot study | Innapropriate intervention |
| 8907 | Effect of a pulmonary rehabilitation program combined with cognitive training on exercise tolerance and cognitive functions among Tunisian male patients with chronic obstructive pulmonary disease: a randomized controlled trial | Inappropriate population |
| 8908 | EFFECTS OF ROUTINE PHYSICAL THERAPY WITH AND WITHOUT HIGH INTENSITY INTERVAL TRAINING ON BALANCE, QUALITY OF LIFE AND FUNCTION IN PARKINSON’S DISEASE PATIENTS | Inappropriate outcome |
| 8909 | Elevated brain temperature under severe heat exposure impairs cortical motor activity and executive function | Inappropriate study design |
| 8910 | Utility of paced breathing tablet guidance apparatus with real-time feedback on autonomic function for individuals with mild cognitive impairment: a pilot study | Inappropriate outcome |
| 8911 | Effects of high intensity interval-based inspiratory muscle training in patients with heart failure: a single-blind randomized controlled trial | Inappropriate population |
| 8912 | Dual-task versus single-task gait rehabilitation after stroke: the protocol of the cognitive-motor synergy multicenter, randomized, controlled superiority trial (SYNCOMOT) | Inappropriate study design |
| 8913 | Nordic Walking in Water on Cerebrovascular Reactivity and Cognitive Function in Elderly Patients with Type 2 Diabetes | Innapropriate intervention |
| 8914 | Effects of an Exercise Program to Reduce Falls in Older People Living in Long-Term Care: a Randomized Controlled Trial | Inappropriate population |
| 8915 | Improving Sleep Quality, Daytime Sleepiness, and Cognitive Function in Patients with Dementia by Therapeutic Exercise and NESA Neuromodulation: a Multicenter Clinical Trial | Innapropriate intervention |
| 8916 | Increasing Activity after Stroke: a Randomized Controlled Trial of High-Intensity Walking and Step Activity Intervention | Inappropriate population |
| 8917 | Mobile health to promote physical activity in people post stroke or transient ischemic attack – study protocol for a feasibility randomised controlled trial | Inappropriate study design |
| 8918 | 'Smart reminder': a feasibility pilot study on the effects of a wearable device treatment on the hemiplegic upper limb in persons with stroke | Inappropriate population |
| 8919 | Exercise Prescription for Older Adults | Inappropriate study design |
| 8920 | Effects of 2-Year-Long Maintenance Training and Detraining on 558 Subacute Ischemic Stroke Patients' Clinical–Motor Symptoms | Inappropriate population |
| 8921 | Does multiple-component intensive pelvic floor muscle training decrease muscle fatigue and symptoms in women with urinary incontinence? | Inappropriate population |
| 8922 | Cognitive-motor dual-task training improves dynamic stability during straight and curved gait in patients with multiple sclerosis: a randomized controlled trial | Inappropriate outcome |

| | | |
|---|---|---|
| 8923 | Combining specific task-oriented training with manual therapy to improve balance and mobility in patients after stroke: a mixed methods pilot randomised controlled trial | Inappropriate population |
| 8924 | Effects of two different dual-task training protocols on gait, balance, and cognitive function in community-dwelling older adults: a 24-week randomized controlled trial | Inappropriate population |
| 8925 | Multi-disciplinary community-based group intervention for fibromyalgia: a pilot randomized controlled trial | Inappropriate population |
| 8926 | Effects of Motor-Cognitive Dual-Task Standing Balance Exergaming Training on Healthy Older Adults' Standing Balance and Walking Performance | Inappropriate study design |
| 8927 | Aerobic exercise and dual-task training combination is the best combination for improving cognitive status, mobility and physical performance in older adults with mild cognitive impairment | Innapropriate intervention |
| 8928 | Trimetazidine in heart failure with preserved ejection fraction: a randomized controlled cross-over trial | Inappropriate population |
| 8929 | Implementation of exergaming for people with dementia: Facilitators, barriers, and recommendations | Inappropriate study design |
| 8930 | The short and long-term effects of a lifestyle intervention in children with mental illnesses: A randomized controlled trial (Movementss study) | Inappropriate population |
| 8931 | Telerehabilitation-based trunk exercise training for motor symptoms of individuals with Parkinson’s disease: A randomized controlled clinical trial | Inappropriate outcome |
| 8932 | A new set of estimated cardiorespiratory fitness equations are associated with cognitive performance in older adults | Inappropriate study design |
| 8933 | A pilot study of the effects of individualized home dual task training by mobile health technology in people with dementia | Inappropriate study design |
| 8934 | Resistance exercise effects on hippocampus subfield volumes and biomarkers of neuroplasticity and neuroinflammation in older adults with low and high risk of mild cognitive impairment: a randomized controlled trial | Innapropriate intervention |
| 8935 | The effect of chrono-nutritional manipulation of carbohydrate intake on sleep macrostructure: a randomized controlled trial | Inappropriate population |
| 8936 | EFFects of Exposure and Cognitive behavioral Therapy for chronic BACK pain (“EFFECT-BACK”): study protocol for a randomized controlled trial | Inappropriate study design |
| 8937 | It Is Smart to Set Treatment Goals, But Are Set Treatment Goals SMART? A Qualitative Assessment of Goals Described in the Assessment of the Burden of COPD Tool | Inappropriate study design |
| 8938 | CycLing and EducATion (CLEAT): protocol for a single centre randomised controlled trial of a cycling and education intervention versus standard physiotherapy care for the treatment of hip osteoarthritis | Inappropriate study design |
| 8939 | A Ketone Monoester with Carbohydrate Improves Cognitive Measures Post-Exercise, but not Performance in Trained Females | Inappropriate population |
| 8940 | A randomised clinical trial comparing outcomes of a single digit volar plate injury - Buddy loops versus dorsal thermoplastic orthosis in a neutral position: study protocol | Inappropriate study design |
| 8941 | Individualizing Basketball-Specific Interval Training Using Anaerobic Speed Reserve: effects on Physiological and Hormonal Adaptations | Inappropriate study design |
| 8942 | Comparison of the Effects of Constraint-Induced Movement Therapy and Unconstraint Exercise on Oxidative Stress and Limb Function—A Study on Human Patients and Rats with Cerebral Infarction | Inappropriate outcome |
| 8943 | Exergames improves cognitive functions in adolescents with depression: Study protocol of a prospective, assessor-blind, randomized controlled trial | Inappropriate population |
| 8944 | RESEARCH ON SPORTS INJURY PREVENTION AND REGIONAL SPORTS FITNESS | Inappropriate study design |
| 8945 | Effects of square dance exercise on cognitive function in elderly individuals with mild cognitive impairment: the mediating role of balance ability and executive function | Innapropriate intervention |
| 8946 | The effect of aerobic dancing on physical fitness and cognitive function in older adults during the COVID-19 pandemic-a natural experiment | Inappropriate population |
| 8947 | Effectiveness and Electrophysiological Mechanisms of Focal Vibration on Upper Limb Motor Dysfunction in Patients with Subacute Stroke: a Randomized Controlled Trial | Inappropriate outcome |
| 8948 | Aerobic training increases hippocampal volume and protects cognitive function for type 2 diabetes patients with normal cognition | Inappropriate population |
| 8949 | Effects of PROtein enriched MEDiterranean Diet and EXercise on nutritional status and cognition in adults at risk of undernutrition and cognitive decline: the PROMED-EX Randomised Controlled Trial | Inappropriate study design |

| | | |
|---|---|---|
| 8950 | Longitudinal changes in physical activity accumulation patterns during 1-year follow-up in stroke survivors | Inappropriate study design |
| 8951 | Effects of Yoga and Aerobic Exercise on Verbal Fluency in Physically Inactive Older Adults: randomized Controlled Trial (FitForAge) | Inappropriate population |
| 8952 | Acute changes in affective valence and perceived distress predict reductions in PTSD symptom severity | Inappropriate population |
| 8953 | Relationship of Changes in Circulating BDNF and Motor Impairment Following a Stroke Rehabilitation Intervention | Inappropriate study design |
| 8954 | The Effects of Sleep Deprivation, Acute Hypoxia, and Exercise on Cognitive Performance: a Multi-Experiment Combined Stressors Study | Inappropriate population |
| 8955 | Effectiveness and cost of integrated cognitive and balance training for balance and falls in cerebellar ataxia: a blinded two-arm parallel group RCT | Inappropriate outcome |
| 8956 | The Characteristics of the Reduction of Interference Effect during Dual-Task Cognitive-Motor Training Compared to a Single Task Cognitive and Motor Training in Elderly: a Randomized Controlled Trial | Inappropriate population |
| 8957 | Randomized, controlled trial of karate for Parkinson's disease yields improvements in quality of life | Inappropriate outcome |
| 8958 | Virtual Reality-Assisted Exercise Therapy during Inpatient Treatment after Intensive Chemotherapy for Acute Myeloid Leukemia (AML) and after Autologous or Allogeneic Stem Cell Transplantation | Inappropriate study design |
| 8959 | Effects of a combination therapy of music-based movement, neuromodulation, cognitive training, and emotion regulation on cognition, physical and mental fitness in community older adults: a three-arm randomized controlled trial | Inappropriate population |
| 8960 | Functional neuroplasticity of facilitation and interference effects on inhibitory control following 3-month physical exercise in aging | Inappropriate study design |
| 8961 | The effects of exergaming on executive and physical functions in older adults with dementia: Randomized controlled trial | Innapropriate intervention |
| 8962 | Multidomain interventions for non-pharmacological enhancement (MINE) program in Chinese older adults with mild cognitive impairment: a multicenter randomized controlled trial protocol | Inappropriate study design |
| 8963 | Effects of breaking up sitting with light-intensity physical activity on cognition and mood in university students | Inappropriate population |
| 8964 | Effects of mind-body exercise Baduanjin on cognition in community-dwelling older people with mild cognitive impairment: a randomized controlled trial | Innapropriate intervention |
| 8965 | Neurobiomechanical mechanism of Tai Chi to improve upper limb coordination function in post-stroke patients: a study protocol for a randomized controlled trial | Inappropriate study design |
| 8966 | Effects of Tai Chi combined with tDCS on cognitive function in patients with MCI: a randomized controlled trial | Innapropriate intervention |
| 8967 | Improved interference control after exercise with blood flow restriction and cooling is associated with but not mediated by increased lactate | Inappropriate population |
| 8968 | Effect of Multicomponent Exercise on Cognition, Physical Function and Activities of Daily Life in Older Adults With Dementia or Mild Cognitive Impairment: A Systematic Review and Meta-analysis | Innapropriate intervention |
| 8969 | Community-based group rehabilitation program for stroke patients with dysphagia on quality of life, depression symptoms, and swallowing function: a randomized controlled trial | Inappropriate population |
| 8970 | Effects of a multicomponent physical activity programme, Mobility-Fit, compared with a standard care lower limb strengthening programme, to promote safe mobility among older adults in care facilities: protocol for a cluster randomised controlled trial | Inappropriate study design |
| 8971 | Effect of cag regimen chemotherapy combined with empowerment theory on anxiety and depression and cancer-related fatigue in leukemia patients | Inappropriate population |
| 8972 | ID: 219084 The Effects of Transcranial Direct Current Stimulation and Rehab on CVA Patients: preliminary Results | Inappropriate population |
| 8973 | Evaluating the effectiveness of video-game based swallowing function training in patients with dysphagia: study protocol for a randomized controlled trial | Inappropriate study design |
| 8974 | Effects of Moderate-to-Vigorous Acute Exercise on Conscious Perception and Visual Awareness | Inappropriate study design |
| 8975 | Alterations in functional connectivity in patients with non-specific chronic low back pain after motor control exercise: a randomized trial | Inappropriate population |
| 8976 | Effects of Multicomponent Exercise on Community-Dwelling Older Adults With Mild Cognitive Impairment | Innapropriate intervention |
| 8977 | The proGnostic role of caRdiac rehAbilitation in patients with left ventriCular anEurysm formation after anterior myocardial infarction (the GRACE study): study rationale and design of a prospective randomized controlled trial | Inappropriate study design |

| | | |
|---|---|---|
| 8978 | Ankle rehabilitation robot training for stroke patients with foot drop: optimizing intensity and frequency | Inappropriate population |
| 8979 | A study on the effect of traditional Chinese exercise combined with rhythm training on the intervention of older adults with mild cognitive impairment | Innapropriate intervention |
| 8980 | Effects of Different Exercise Modes on Gait Performance of Parkinson's Disease Patients: A Systematic Review and Network Meta-Analysis | Inappropriate study design |
| 8981 | Immediate and short-term effects of single-task and motor-cognitive dual-task on executive function | Inappropriate population |
| 8982 | Effects of acute moderate-intensity aerobic exercise on cognitive function in E-athletes: a randomized controlled trial | Inappropriate population |
| 8983 | The impact of aerobic exercise training on cognitive function and gut microbiota in methamphetamine-dependent individuals in the community | Inappropriate population |
| 8984 | Look into my eyes: What can eye-based measures tell us about the relationship between physical activity and cognitive performance? | Inappropriate study design |
| 8985 | Larger splenic emptying correlate with slower EPOC kinetics in healthy men and women during supine cycling | Inappropriate study design |
| 8986 | High intensity interval training as a novel treatment for impaired awareness of hypoglycaemia in people with type 1 diabetes (HIT4HYPOS): a randomised parallel-group study [with consumer summary] | Inappropriate population |
| 8987 | Effects of combined dietary intervention and physical-cognitive exercise on cognitive function and cardiometabolic health of postmenopausal women with obesity: a randomized controlled trial | Inappropriate population |
| 8988 | An Investigation into the effects of acute exercise on activities of daily living and cognition in older adults | Inappropriate population |
| 8989 | Association Between Subjective Cognitive Decline and Twice-Weekly Muscle-Strengthening Activities in Middle-Aged and Older US Adults: An Analysis of the 2019 Behavioral Risk Factor Surveillance System | Inappropriate study design |
| 8990 | Clinical pharmacology and tolerability of REC-994, a redox-cycling nitroxide compound, in randomized phase 1 dose-finding studies | Inappropriate population |
| 8991 | Use of athletic tape in the speech-language-hearing treatment of post-stroke facial paralysis in the acute phase | Inappropriate population |
| 8992 | Effectiveness of Immersive Virtual Reality-Based Hand Rehabilitation Games for Improving Hand Motor Functions in Subacute Stroke Patients | Inappropriate population |
| 8993 | The effect of training to target cognitive biases towards social rejection in eating disorders | Inappropriate population |
| 8994 | Cardiovascular Safety of Ozanimod in Patients With Ulcerative Colitis: true North and Open-Label Extension Analyses | Inappropriate study design |
| 8995 | Effects of dual-task resistance exercise on cognition, mood, depression, functional fitness, and activities of daily living in older adults with cognitive impairment: a single-blinded, randomized controlled trial | Innapropriate intervention |
| 8996 | Using 30-day modified rankin scale score to predict 90-day score in patients with intracranial hemorrhage: derivation and validation of prediction model | Inappropriate study design |
| 8997 | Protocol for a randomised controlled trial of ketamine versus ketamine and behavioural activation therapy for adults with treatment-resistant depression in the community | Inappropriate study design |
| 8998 | Decompressive craniectomy plus best medical treatment versus best medical treatment alone for spontaneous severe deep supratentorial intracerebral haemorrhage: a randomised controlled clinical trial | Inappropriate population |
| 8999 | Results of a cohort single-center randomized study of the modulating effect of the drug Mexidol in the rehabilitation of patients who suffered acute cerebral insufficiency | Inappropriate population |
| 9000 | Beyond social engagement: cognitive training leads to greater cognitive improvement in older adults | Inappropriate study design |
| 9001 | A single-center, assessor-blinded, randomized controlled clinical trial to test the safety and efficacy of a novel brain-computer interface controlled functional electrical stimulation (BCI-FES) intervention for gait rehabilitation in the chronic stroke population | Inappropriate study design |
| 9002 | The effectiveness of reminiscence therapy on the symptom management, the life satisfaction, and the self-transcendence in palliative care patients: a randomized controlled trial | Inappropriate population |
| 9003 | Effects of Physical Therapy and Dalfampridine on Function and Quality of Life in Nonambulatory Individuals With Multiple Sclerosis: a Randomized Controlled Trial | Innapropriate intervention |
| 9004 | Tailored videoconferencing counselling program to support family carers of people living with dementia during the transition to permanent residential care: a pilot and feasibility randomised trial | Inappropriate population |

| | | |
|---|---|---|
| 9005 | Hearing Impairment and Physical Activity and Physical Functioning in Older Adults: baseline Results From the ACHIEVE Trial | Inappropriate study design |
| 9006 | Changes in cognitive function after a 12-week POWER rehabilitation in older adults with schizophrenia and frailty | Innappropriate intervention |
| 9007 | The Effects of a Nonpharmacological Intervention Practice for Older Adults with Mild Cognitive Impairment and Their Family Caregivers in China | Innappropriate intervention |
| 9008 | Bilateral effect of acupuncture on cerebrum and cerebellum in ischaemic stroke patients with hemiparesis: a randomised clinical and neuroimaging trial | Inappropriate outcome |
| 9009 | Wild Blueberry Extract Intervention in Healthy Older Adults: a Multi-Study, Randomised, Controlled Investigation of Acute Cognitive and Cardiovascular Effects | Inappropriate population |
| 9010 | Chanwuyi Lifestyle Medicine Program Improves Memory and Executive Functions of Older Adults With Mild Cognitive Impairment | Innappropriate intervention |
| 9011 | Information technology-supported integrated health service for older adults in long-term care settings | Inappropriate outcome |
| 9012 | Findings from a pragmatic cluster randomised controlled feasibility trial of a music and dance programme for community dwelling older adults | Inappropriate population |
| 9013 | Prismatic adaptation coupled with cognitive training as novel treatment for developmental dyslexia: a randomized controlled trial | Innappropriate intervention |
| 9014 | Effects of robot-assisted upper limb training combined with intermittent theta burst stimulation (iTBS) on cortical activation in stroke patients: a functional near-infrared spectroscopy study | Innappropriate intervention |
| 9015 | Performance in myoelectric pattern recognition improves with transcranial direct current stimulation | Inappropriate study design |
| 9016 | Hearing Loss and Dementia: where to From Here? | Inappropriate study design |
| 9017 | A multidomain lifestyle intervention to maintain optimal cognitive functioning in Dutch older adults-study design and baseline characteristics of the FINGER-NL randomized controlled trial | Innappropriate intervention |
| 9018 | Design and Methods of the Early Age-Related Hearing Loss Investigation Randomized Controlled Trial | Inappropriate study design |
| 9019 | Video clips for patient comprehension of atrial fibrillation and deep vein thrombosis in emergency care. A randomised clinical trial | Inappropriate population |
| 9020 | A feasibility randomised waitlist-controlled trial of a personalised multi-level language treatment for people with aphasia: the remote LUNA study | Innappropriate intervention |
| 9021 | Effects of Neuropilates on Functional Outcomes in Chronic Stroke: a Randomized Clinical Trial | Inappropriate outcome |
| 9022 | Embedding Caregiver Support Within Adult Day Services: outcomes of a Multisite Trial | Inappropriate population |
| 9023 | Early tirofiban administration after intravenous thrombolysis in acute ischemic stroke (ADVENT): study protocol of a multicenter, randomized, double-blind, placebo-controlled clinical trial | Inappropriate study design |
| 9024 | A randomized clinical trial: efficacy of group-based acceptance and commitment therapy program for breast cancer patients with high fear of progression | Inappropriate population |
| 9025 | Increased trailing limb angle in hemiplegic patients after training with a knee orthosis: a randomized controlled trial | Inappropriate population |
| 9026 | Randomized controlled trials of non-pharmacological interventions for healthy seniors: effects on cognitive decline, brain plasticity and activities of daily living-A 23-year scoping review | Inappropriate study design |
| 9027 | Ketamine vs Electroconvulsive Therapy for Treatment-Resistant Depression: a Secondary Analysis of a Randomized Clinical Trial | Inappropriate population |
| 9028 | Patient-reported outcome measures for monitoring primary care patients with depression: the PROMDEP cluster RCT and economic evaluation | Inappropriate population |
| 9029 | Web-Based Therapist-Guided Mindfulness-Based Cognitive Behavioral Therapy for Body Dysmorphic Disorder: pilot Randomized Controlled Trial | Inappropriate population |
| 9030 | A pilot randomized controlled trial of a telemedicine psychosocial intervention to improve symptom management in adults with long COVID: the COPE study protocol | Inappropriate study design |
| 9031 | Inhaled iloprost in off-pump coronary artery bypass surgery: a randomized controlled trial | Inappropriate population |
| 9032 | Protocol for a double-blind placebo-controlled randomised controlled trial assessing the impact of oral semaglutide in amyloid positivity (ISAP) in community dwelling UK adults | Inappropriate study design |
| 9033 | Greater Role of Cognitive Impairment Over Fatigue in Post-COVID-19 Quality of Life: a Post-Hoc Analysis of a Randomized Controlled Trial | Innappropriate intervention |

| | | |
|---|---|---|
| 9034 | Effectiveness of Extracorporeal Shock Wave Therapy after Botulinum Toxin Injection for Post-Stroke Upper Extremity Spasticity: a Randomized Controlled Study | Inappropriate population |
| 9035 | Hemodynamic changes in the prone position according to fluid loading after anaesthesia induction in patients undergoing lumbar spine surgery: a randomized, assessor-blind, prospective study | Inappropriate population |
| 9036 | Cognitive enhancing effect of rTMS combined with tDCS in patients with major depressive disorder: a double-blind, randomized, sham-controlled study | Innapropriate intervention |
| 9037 | Effect of task-oriented training assisted by force feedback hand rehabilitation robot on finger grasping function in stroke patients with hemiplegia: a randomised controlled trial | Inappropriate population |
| 9038 | Effects of Maitland mobilization technique on upper extremity function in stroke survivors with spasticity: an experimental study | Inappropriate population |
| 9039 | An Examination into the Effects of a Nutraceutical Supplement on Cognition, Stress, Eye Health, and Skin Satisfaction in Adults with Self-Reported Cognitive Complaints: a Randomized, Double-Blind, Placebo-Controlled Trial | Innapropriate intervention |
| 9040 | Motor imagery-based brain-computer interface rehabilitation programs enhance upper extremity performance and cortical activation in stroke patients | Inappropriate population |
| 9041 | Effects of mindfulness-based cognitive therapy for Chinese adults with PTSD symptoms: protocol for a randomised controlled trial | Inappropriate study design |
| 9042 | Exploring the effects of cannabis health warnings on protective health intentions among US adults in legal recreational states | Inappropriate study design |
| 9043 | Associations between brachial-ankle pulse wave velocity and hypertensive retinopathy in treated hypertensive adults: results from the China Stroke Primary Prevention Trial (CSPPT) | Inappropriate study design |
| 9044 | Effectiveness of multi-modal home-based videoconference interventions on sleep in older adults: study protocol for a randomized controlled trial | Inappropriate study design |
| 9045 | The Effects of High-Intensity Interval Training and Moderate Alcohol Consumption on Cognitive Performance-A Multidisciplinary Intervention in Young Healthy Adults | Inappropriate population |
| 9046 | Australian trial of behavioural activation for people with schizophrenia experiencing negative symptoms: a feasibility randomised controlled trial protocol | Inappropriate study design |
| 9047 | Emerging From the Haze™: pilot Feasibility Study Comparing Two Virtual Formats of a Cognitive Rehabilitation Intervention | Innapropriate intervention |
| 9048 | Effects of Withania somnifera Extract in Chronically Stressed Adults: a Randomized Controlled Trial | Inappropriate population |
| 9049 | Once-daily upadacitinib versus placebo in adults with extensive non-segmental vitiligo: a phase 2, multicentre, randomised, double-blind, placebo-controlled, dose-ranging study | Inappropriate population |
| 9050 | Targeting GPVI with glenzocimab in COVID-19 patients: results from a randomized clinical trial | Inappropriate population |
| 9051 | Effectiveness of a Web-Based Cognitive Behavioral Self-Help Intervention for Binge Eating Disorder: a Randomized Clinical Trial | Inappropriate population |
| 9052 | Oral phytate supplementation on the progression of mild cognitive impairment, brain iron deposition and diabetic retinopathy in patients with type 2 diabetes: a concept paper for a randomized double blind placebo controlled trial (the PHYND trial) | Inappropriate study design |
| 9053 | Preoperative walking exercise to improve prognosis in patients with supratentorial brain tumours after craniotomy: protocol for a randomised controlled trial | Inappropriate study design |
| 9054 | Adherence to the Mediterranean Diet, Sodium and Potassium Intake in People at a High Risk of Dementia | Inappropriate study design |
| 9055 | A Self-Guided Internet-Based Intervention for the Reduction of Gambling Symptoms: a Randomized Clinical Trial | Inappropriate population |
| 9056 | Prognostic Value and Determinants of High-Sensitivity Cardiac Troponin T in Patients With a Systemic Right Ventricle: insights From the SERVE Trial | Inappropriate population |
| 9057 | Multicomponent Exercise Intervention for Preventing Falls and Improving Physical Functioning in Older Nursing Home Residents: a Single-Blinded Pilot Randomised Controlled Trial | Inappropriate outcome |
| 9058 | Effects of modified-constraint induced movement therapy based telerehabilitation on upper extremity motor functions in stroke patients | Inappropriate population |
| 9059 | Evaluation of the internet-based intervention Selfapy" in participants with unipolar depression and the impact on quality of life: a randomized, parallel group study" | Inappropriate population |
| 9060 | Disruption of dopamine D2/D3 system function impairs the human ability to understand the mental states of other people | Inappropriate population |

| | | |
|---|---|---|
| 9061 | Effects of flexor reflex stimulation on gait aspects in stroke patients: randomized clinical trial | Inappropriate population |
| 9062 | Re-Imagining Hospital Patient Room Design for People After stroke: a Randomized Controlled Study Using Virtual Reality | Inappropriate population |
| 9063 | Optimizing the physical activity intervention for older adults with mild cognitive impairment: a factorial randomized trial | Innapropriate intervention |
| 9064 | Strength training improves functional capacity of individuals with chronic heart failure: randomized clinical trial | Innapropriate intervention |
| 9065 | Musical and electrical stimulation as intervention in disorder of consciousness (DOC) patients: a randomised cross-over trial | Inappropriate outcome |
| 9066 | Virtual reality-based training may improve visual memory and some aspects of sustained attention among healthy older adults - preliminary results of a randomized controlled study | Inappropriate population |
| 9067 | The Effectiveness of Virtual Reality-Based Training on Cognitive, Social, and Physical Functioning in High-Functioning Older Adults (CoSoPhy FX): 2-Arm, Parallel-Group Randomized Controlled Trial | Inappropriate population |
| 9068 | Impact of KRASG12 mutations on survival with trifluridine/tipiracil plus bevacizumab in patients with refractory metastatic colorectal cancer: post hoc analysis of the phase III SUNLIGHT trial | Inappropriate population |
| 9069 | Cognition after a 4-week high phenylalanine intake in adults with phenylketonuria - a randomized controlled trial | Inappropriate population |
| 9070 | Functional Limitations and Exercise Intolerance in Patients With Post-COVID Condition: a Randomized Crossover Clinical Trial | Inappropriate outcome |
| 9071 | Government Direct-to-Consumer Education to Reduce Prescription Opioid Use: a Cluster Randomized Clinical Trial | Inappropriate population |
| 9072 | Multidomain Dementia Risk Reduction in Primary Care is Feasible: a Proof-of-concept study | Innapropriate intervention |
| 9073 | Post-Stroke Impairments of Manual Dexterity and Finger Proprioception: their Contribution to Upper Limb Activity Capacity | Inappropriate study design |
| 9074 | Intraoral Acupuncture for Sialorrhea in Stroke Patients: a Study Protocol for a Randomized Controlled Trial | Inappropriate study design |
| 9075 | Effects of a Multicomponent Intervention With Cognitive Training and Lifestyle Guidance for Older Adults at Risk of Dementia: A Randomized Controlled Trial | Innapropriate intervention |
| 9076 | Effect of Metformin on Plasma and Cerebrospinal Fluid Biomarkers in Non-Diabetic Older Adults with Mild Cognitive Impairment Related to Alzheimer's Disease | Inappropriate outcome |
| 9077 | The Effect of Acupuncture on Brain Iron Deposition and Body Iron Metabolism in Vascular Cognitive Impairment: protocol for a Randomized Controlled Trial | Inappropriate study design |
| 9078 | Functional near-infrared spectroscopy-based neurofeedback training regulates time-on-task effects and enhances sustained cognitive performance | Inappropriate population |
| 9079 | Minimally invasive surgeries for spontaneous hypertensive intracerebral hemorrhage (MISICH): a multicenter randomized controlled trial | Inappropriate population |
| 9080 | The safety and efficacy of escitalopram and sertraline in post-stroke depression: a randomized controlled trial | Inappropriate population |
| 9081 | Effects of Baduanjin exercise on cognitive frailty, oxidative stress, and chronic inflammation in older adults with cognitive frailty: a randomized controlled trial | Innapropriate intervention |
| 9082 | Effectiveness of Shoulder Taping in Treating Hemiplegic Shoulder Subluxation: a Randomized Controlled Study of 35 Patients | Inappropriate population |
| 9083 | Paraxanthine provides greater improvement in cognitive function than caffeine after performing a 10-km run | Inappropriate population |
| 9084 | Multi-night acoustic stimulation is associated with better sleep, amyloid dynamics, and memory in older adults with cognitive impairment | Inappropriate study design |
| 9085 | Combining muscle-computer interface guided training with bihemispheric tDCS improves upper limb function in patients with chronic stroke | Inappropriate population |
| 9086 | Exoskeleton rehabilitation robot training for balance and lower limb function in sub-acute stroke patients: a pilot, randomized controlled trial | Inappropriate population |
| 9087 | Antiplatelet therapy after coronary artery bypass surgery: five year follow-up of randomised DACAB trial | Inappropriate study design |
| 9088 | Erratum: sublingual edaravone dexborneol for the treatment of acute ischemic stroke: the TASTE-SL randomized clinical trial (JAMA Neurol. (2024) DOI: 10.1001/jamaneurol.2023.5716) | Inappropriate study design |
| 9089 | CSF proteomic analysis of semorinemab Ph2 trials in prodromal-to-mild (Tauriel) and mild-to-moderate (Lauriet) Alzheimer’s disease identifies distinct trial cell-type specific proteomic signatures | Inappropriate study design |

| | | |
|---|---|---|
| 9090 | Manual Perturbation and Proprioceptive Neuromuscular Facilitation for Trunk Stability & Lower Extremity Function in subjects with Stroke: a Randomized Clinical Trial | Inappropriate population |
| 9091 | Investigation of the effects of cocoa powder on fatigue severity in patients with relapsing-remitting multiple sclerosis: a randomized controlled trial | Inappropriate outcome |
| 9092 | Feasibility and efficacy of an online cognitive behavioral therapy program for insomnia and anxiety in older adults | Inappropriate population |
| 9093 | Effect of acceptance and commitment therapy-based interventions on mental health and cognitive impairment for patients with advanced colorectal cancer | Innapropriate intervention |
| 9094 | Effect of Early Coma Arousal Therapy on Conscious Level Recovery and Cognition in Traumatic Brain Injury | Innapropriate intervention |
| 9095 | Schema therapy versus treatment as usual for outpatients with difficult-to-treat depression: study protocol for a parallel group randomized clinical trial (DEPRE-ST) | Inappropriate study design |
| 9096 | Outcomes of Exposure Based Treatment for Hoarding Disorder | Inappropriate population |
| 9097 | 104. Modafinil Exerts Dose-Dependent Effects on Risk-Taking as Measured by the Human Iowa Gambling Task | Inappropriate population |
| 9098 | 242. Adjunctive Treatment With Semaglutide for Cognitive Dysfunction in Major Depressive Disorder: reduction in Depression Scores Associated With Increased HbA1c Function and Glucose Levels | Innapropriate intervention |
| 9099 | Safety of Dabigatran in Patients With Acute Minor Non-Cardioembolic Ischemic Stroke or Transient Ischemic Attack and Cerebral Microbleeds: a Post Hoc Analysis of the DATAS-II Randomized Trial | Inappropriate study design |
| 9100 | CUMULATIVE EFFECTS OF MILD SLEEP RESTRICTION ON LAPSES OF ATTENTION | Inappropriate study design |
| 9101 | PULLING AN ALL-NIGHTER IMPAIRS ORGANIC CHEMISTRY LEARNING: AN AT-HOME, ZOOM-BASED SLEEP DEPRIVATION EXPERIMENT | Inappropriate population |
| 9102 | The robot assisted upper-limb rehabilitation program on functional recovery in persons with stroke: results from a randomized clinical trial | Inappropriate population |
| 9103 | Evaluation of effectiveness and acceptability of a psychological treatment for smoking cessation combined with a smartphone App: a pilot study | Inappropriate study design |
| 9104 | THE COMPARISON OF THE CAREN VIRTUAL REALITY SYSTEM-BASED PROTOCOL AND ROUTINE PHYSICAL THERAPY ON BALANCE AND POSTURAL CONTROL IN PATIENTS WITH STROKE: a RANDOMIZED CONTROLLED TRIAL | Inappropriate outcome |
| 9105 | Effectiveness of Simultaneous Combined Intervention for Enhancing Cognitive Function in Patients with Moderate Alzheimer's Disease | Innapropriate intervention |
| 9106 | 464. Ketamine Disrupts Consciousness in Healthy Controls in Relation With Psychotic-Like Symptoms | Inappropriate population |
| 9107 | EVALUATION OF EFFICACY OF SEXUAL NOVELTY AS A NOVEL THERAPEUTIC STRATEGY TO TREAT HYPOACTIVE SEXUAL DESIRE DYSFUNCTION IN WOMEN IN MONOGAMOUS HETEROSEXUAL RELATIONSHIPS ATTRIBUTING REDUCED SEXUAL DESIRE TO SEXUAL BOREDOM: a PROSPECTIVE RANDOMIZED STUDY | Inappropriate population |
| 9108 | RE-OPEN: randomised trial of biosimilar TNK versus TPA during endovascular therapy for acute ischaemic stroke due to large vessel occlusions | Inappropriate study design |
| 9109 | EFFICACY OFMINDFULNESS BASED COGNITIVE THERAPY IN MANAGEMENT OF PROVOKED VULVODYNIA: a FEASIBILITY PILOT STUDY | Inappropriate population |
| 9110 | Accelerated Theta Burst Transcranial Magnetic Stimulation for Refractory Depression in Autism Spectrum Disorder | Inappropriate study design |
| 9111 | The Effect of Remote Ischemic Conditioning in Stroke Subtypes: a Subgroup Analysis From the RESIST Trial | Inappropriate study design |
| 9112 | Trial in Progress: the REJOIN Trial to Use Exercise and Education to Relieve Joint Pain in Older Breast Cancer Survivors Taking Aromatase Inhibitors | Inappropriate study design |
| 9113 | Randomized Controlled Trial of Intradialytic Cognitive and Physical Training to Enhance Functional Capacity | Innapropriate intervention |
| 9114 | Cognitive Effects of Reducing First-Generation Antipsychotic Dose Compared to Switching to Ziprasidone in Long-Stay Patients with Schizophrenia | Innapropriate intervention |
| 9115 | The effectiveness of cognitive training in people in psychiatry with depressive symptoms – A randomized controlled study | Inappropriate population |
| 9116 | Treatment and Clinical Outcome of Patients Excluded From ACST-2 in a Trial Collaborating Hospital | Inappropriate population |
| 9117 | Adverse Events During and After Early Mobilization: a Post-hoc Analysis of the TEAM Trial | Inappropriate study design |

| | | |
|---|---|---|
| 9118 | Evaluating the Implementation Fidelity of a Pilot Pragmatic Randomized Clinical Trial Comparing Daily-Delivered Meals to Mailed Frozen Meals | Inappropriate population |
| 9119 | Feasibility and Efficacy of a Virtual Reality Social Prediction Training in Children and Young Adults with Congenital Cerebellar Malformations | Innapropriate intervention |
| 9120 | Rapid Intravenous Glyceryl Trinitrate in Ischemic Damage (RIGID): a potential neuroprotection strategy for acute ischemic stroke (AIS) patients | Inappropriate population |
| 9121 | Post-traumatic Growth, Anxiety, Depression, and Post-traumatic Stress Disorder Among Survivors of Critical Illness Six Months Post-discharge | Inappropriate population |
| 9122 | Scrambler Therapy for Treatment of Post Stroke Pain | Inappropriate study design |
| 9123 | A preventive school-based paradigm using virtual reality technologies for improving emotional regulation, depressive and anxiety symptoms in children and adolescents (e-Emotio project): a randomized controlled pilot trial | Inappropriate study design |
| 9124 | Efficacy of internet-based self-help cognitive behavioural therapy for insomnia (CBT-I) in reducing insomnia symptoms among adults with sub-threshold insomnia symptoms: a randomized controlled trial | Inappropriate population |
| 9125 | Synergistic efficacy of repetitive peripheral magnetic stimulation on central intermittent theta burst stimulation for upper limb function in patients with stroke: a double-blinded, randomized controlled trial | Inappropriate population |
| 9126 | A 4-year Follow-up of Brain-computer Interface on Upper Limb Motor Recovery of Aging Post-stroke Patients | Inappropriate study design |
| 9127 | Effectiveness of e-based cognitive behavioral therapy for insomnia on enhancing depression and insomnia outcome in Chinese youth with both diagnoses | Inappropriate population |
| 9128 | A Novel Adjuvant Strategy After Endovascular Therapy: arterial Glyceryl Trinitrate in Acute Ischemic Stroke for Neuroprotection (AGAIN) | Inappropriate population |
| 9129 | Group-based cognitive behavioural therapy and bright light therapy in youths with insomnia and evening chronotype: interim analysis of a randomised controlled trial | Inappropriate population |
| 9130 | The Effect of Task-Oriented Training and Progressive Resisted Exercise on Upper Limb Function and Quality of Life in Chronic Stroke Patients | Inappropriate population |
| 9131 | The effect of self-focused attention during mirror gazing on body image evaluations, appearance-related imagery, and urges to mirror gaze | Inappropriate study design |
| 9132 | 435. Improvement of Auditory Steady State Response (ASSR) and Auditory Hallucinations in First-Episode Schizophrenia: preliminary Findings From a Single-Blind RCT of Auditory Control Enhancement (ACE) Therapy | Inappropriate outcome |
| 9133 | Patients Randomised to Glenzocimab Suffered Less Haemorrhagic Transformation at 24 Hours Compared to Placebo: aI-Imaging Sub-Analysis of the ACTIMIS Trial | Inappropriate population |
| 9134 | Think Drink: feasibility trial of a multicomponent hydration care intervention for people living with dementia in care homes | Inappropriate outcome |
| 9135 | 248MITICA - A PROSPECTIVE STUDY OF ACTIVE SURVEILLANCE (AS) VS IMMEDIATE SURGERY (IS) FOR PAPILLARY THYROID MICROCARCINOMA (PTMC) | Inappropriate study design |
| 9136 | Theta-burst rTMS in schizophrenia to ameliorate negative and cognitive symptoms: study protocol for a double-blind, sham-controlled, randomized clinical trial | Inappropriate study design |
| 9137 | A digital behavioral therapy for fibromyalgia progressively improves clinical outcomes: results of the PROSPER-FM randomized clinical trial | Inappropriate population |
| 9138 | Long-term Support Following Bariatric Surgery: 18-month outcomes of a Stepped Care Intervention Program using social media | Inappropriate population |
| 9139 | A multidomain lifestyle intervention to maintain optimal cognitive functioning in Dutch older adults—study design and baseline characteristics of the FINGER-NL randomized controlled trial | Innapropriate intervention |
| 9140 | Evaluating the effects of Bacopa monnieri on cognitive performance and sleep quality of patients with mild cognitive impairment: a triple-blinded, randomized, placebo-controlled trial | Innapropriate intervention |
| 9141 | Surgery compared to fibrinolytic therapy for symptomatic left-sided prosthetic heart valve thrombosis (SAFE-PVT): rationale and design of a randomized controlled trial | Inappropriate study design |
| 9142 | Influence of Living Arrangements and Within Household Communications on Blood Pressure Control Among Stroke Patients | Inappropriate population |
| 9143 | Evidence of anchoring bias in novice (first year) osteopathic French students in the context of the primary respiratory mechanism: a randomized-experimental study | Inappropriate study design |
| 9144 | A Randomized Controlled Trial of Palliative Care to Improve Health Related Quality of Life in Pulmonary Arterial Hypertension | Inappropriate population |

| | | |
|---|---|---|
| 9145 | Aquatic Cognitive-Motor Exercise Training on Cognition and Neurotrophic Factors in Older Adults | Inappropriate population |
| 9146 | The effectiveness of cognitive behavioral therapy for insomnia on sleep outcomes in the context of pain among older adult veterans | Inappropriate population |
| 9147 | Patient acceptability and adherence to the COMPASS trial drug recommendations following symptomatic carotid endarterectomy (CEA) | Inappropriate study design |
| 9148 | Hormone Replacement and Alternatives for Relief of Vasomotor Symptoms in Women at Increased Risk for Breast Cancer | Inappropriate population |
| 9149 | Rehabilitation of Patients with Post-Stroke Cognitive Impairments Using a P300-Based Brain–Computer Interface: results of a Randomized Controlled Trial | Innapropriate intervention |
| 9150 | A Randomized Clinical Trial Evaluating the Impact of a Personalized Video-Based Educational Platform After Discharge | Inappropriate population |
| 9151 | Efficacy of cognitive training on cognition, symptoms and functioning: impact of motivation and attendance | Innapropriate intervention |
| 9152 | Effect of the Multicontext Approach on Functional Cognition in Parkinson Disease: pilot Randomized Controlled Trial | Innapropriate intervention |
| 9153 | Gamma oscillations induced by 40-Hz visual-auditory stimulation for the treatment of acute-phase limb motor rehabilitation after stroke: study protocol for a prospective randomized controlled trial | Inappropriate study design |
| 9154 | A multicenter, randomized, double-blind, placebo-controlled ascending dose study to evaluate the safety, tolerability, pharmacokinetics (PK) and pharmacodynamic (PD) effects of Posiphen in subjects with Early Alzheimer's Disease | Innapropriate intervention |
| 9155 | Immediate- or Delayed-Intensive Statin in Acute Cerebral Ischemia: the INSPIRES Randomized Clinical Trial | Inappropriate population |
| 9156 | Bimanual coordinated motor skill learning in patients with a chronic cerebellar stroke | Inappropriate study design |
| 9157 | Implementation of CF-CBT in US and Canadian CF centers: adoption, effectiveness and reach in phase one | Inappropriate population |
| 9158 | Shape configuration of mental target representation as a holistic measure in a 3D real world pointing test for spatial orientation | Inappropriate study design |
| 9159 | STENT OMISSION AFTER URETEROSCOPY AND LITHOTRIPSY: a PATIENT-CENTERED COMPARATIVE EFFECTIVENESS CLINICAL TRIAL | Inappropriate population |
| 9160 | The role of nutritional supplement on post-stroke fatigue: a pilot randomized controlled trial | Innapropriate intervention |
| 9161 | EXPERIMENTAL SLEEP RESTRICTION AFFECTS ARTWORK PERCEPTIONS | Inappropriate population |
| 9162 | REDUCED-DOSE ONABOTULINUMTOXINA FOR URGENCY INCONTINENCE AMONG OLDER FEMALES (RELIEF): a RANDOMIZED CONTROLLED COMPARATIVE EFFECTIVENESS TRIAL WITH EMBEDDED QUALITATIVE AND COSTING ANALYSES | Inappropriate population |
| 9163 | Results of a randomized, multicenter trial to determine the efficacy of sovateltide in treating acute cerebral ischemic stroke patients | Inappropriate population |
| 9164 | Endocrine response assessment in HR+ HER2- early breast cancer: concordance of central versus local Ki67 measurements in the ADAPTcycle trial | Inappropriate population |
| 9165 | Different Response to Blood Pressure Reduction in Lobar and Deep Intracerebral Hemorrhage | Inappropriate study design |
| 9166 | Efficacy of Dual Antiplatelet Therapy After Lacunar versus Non-Lacunar Syndromes in the POINT Trial | Inappropriate study design |
| 9167 | Tackle your Tics, a brief intensive group-based exposure treatment for young people with tics: results of a randomised controlled trial | Inappropriate population |
| 9168 | A Randomized Controlled Trial to Assess the Benefit of Art & Music Therapy on Quality of Life in Patient with Breast Cancer | Inappropriate population |
| 9169 | The Effect of Brainwave Synchronization Using Binaural Beats on Improving Working Memory and Reducing the Severity of Symptoms in Women With Obsessive-compulsive Disorder | Inappropriate population |
| 9170 | EMDR therapy vs. supportive therapy as adjunctive treatment in trauma-exposed bipolar patients: a randomised controlled trial | Inappropriate population |
| 9171 | The Effects of Affective Expectations on Willingness to Expend Cognitive Effort in Dysphoric and Non-Dysphoric Individuals | Inappropriate population |
| 9172 | Transcatheter or surgical aortic valve implantation: 10-year outcomes of the NOTION trial | Inappropriate study design |

| | | |
|---|---|---|
| 9173 | PO-02-117 INTRACARDIAC ECHOCARDIOGRAPHY VERSUS TRANSESOPHAGEAL ECHOCARDIOGRAPHY FOR ATRIAL FIBRILLATION ABLATION: AN INTERIM ANALYSIS OF A MULTICENTER RANDOMIZED CONTROLLED TRIAL (THE ICE VS TEE STUDY) | Inappropriate population |
| 9174 | Vision and hearing difficulty and effects of cognitive training in older adults | Innapropriate intervention |
| 9175 | Personalised functional imaging-guided multitarget continuous theta burst stimulation for post-stroke aphasia: study protocol for a randomised controlled trial | Inappropriate study design |
| 9176 | A Pilot Case Study: diverse-Equitable-Inclusive-Empathetic Practitioners Share a Common Language | Inappropriate population |
| 9177 | Multidomain Intervention for the Reversal of Cognitive Frailty Using a Personalized Approach (AGELESS Trial): recruitment and Baseline Characteristics of Participants | Innapropriate intervention |
| 9178 | Efficacy of prasugrel versus clopidogrel for early neurological deterioration in branch atheromatous disease: an exploratory study | Inappropriate population |
| 9179 | A Nationwide Pilot Study Testing a Remotely-Delivered Prolonged Nightly Fasting Intervention in Stressed Midlife Adults Living with Obesity and Memory Decline | Inappropriate population |
| 9180 | Attentional state-synchronous peripheral electrical stimulation during action observation induced distinct modulation of corticospinal plasticity after stroke | Inappropriate study design |
| 9181 | 730 LACTIPLANTIBACILLUS PLANTARUM HEAL9 REDUCES FECAL CALPROTECTIN LEVELS IN ELDERLY INDIVIDUALS WITH LOW-GRADE INFLAMMATION | Inappropriate population |
| 9182 | Cognitive training reshapes functional lateralization of frontoparietal network in patients with vascular cognitive impairment no dementia | Innapropriate intervention |
| 9183 | Telehealth sleep intervention for young children with autism: recent findings from a randomized clinical trial | Inappropriate population |
| 9184 | Beneficial effects of mindfulness-based intervention on hippocampal volumes and episodic memory for childhood adversity survivors | Inappropriate population |
| 9185 | Event-Related Potential Changes Following 12-week Yoga Practice in T2DM Patients: a Randomized Controlled Trial | Innapropriate intervention |
| 9186 | Intoxication due to Δ9-tetrahydrocannabinol is characterized by disrupted prefrontal cortex activity | Inappropriate population |
| 9187 | Critical Care Decisions After Large Core Cerebral Infarctions: a Secondary Analysis From the SELECT2 Trial | Inappropriate population |
| 9188 | Preliminary Evidence to Support the use of Startle Adjuvant Rehabilitation Therapy (START) via tele-health sessions to promote accessibility and reduce socioeconomic disparities in post-stroke care | Inappropriate population |
| 9189 | EFFICACY OF YWTL EXERCISES ON POSTURE IN MOTOR AND COGNITIVE DUAL TASK DURING SMARTPHONE USE | Inappropriate study design |
| 9190 | Rates of Perceived Cognitive Exertion are Elevated in Older Adults with Mild Cognitive Impairment during Rhythmic Motor-Cognitive Tasks | Inappropriate outcome |
| 9191 | Rhythmic Movement Training Improves Spatial and Temporal Modulation of Gait in Older Adults with and Without Mild Cognitive Impairment | Inappropriate outcome |
| 9192 | Effects of Phytoncide Fragrance on Resting-State Brain Activity in Mild Cognitive Impairment: a Randomized Double-Blind Controlled Study | Innapropriate intervention |
| 9193 | Effects of Brain Computer Interface (BCI) rehabilitation robot on upper limb function in chronic stroke patients - A randomized controlled trial | Inappropriate population |
| 9194 | Vagus Nerve Stimulation Paired With Rehabilitation for Post-Stroke Arm Impairment: one Year Follow-Up of the VNSREHAB Pivotal Trial | Inappropriate population |
| 9195 | Addressing Pain Using a Mediterranean Ketogenic Nutrition Program in Older Adults with Mild Cognitive Impairment | Inappropriate outcome |
| 9196 | Novel computer-based trainings for patients with aphasia | Innapropriate intervention |
| 9197 | Reliability Of B-Mode Ultrasound Carotid Plaque Characterization in the CREST-2 Trial | Inappropriate population |
| 9198 | The role of the anterior cingulate cortex in inhibitory control - The ACDC TMS study with patients with obsessive-compulsive disorder | Inappropriate population |
| 9199 | ProstACT GLOBAL: a phase 3 study best standard of care with and without 177Lu-DOTA-rosopatamab (TLX591) for patients with PSMA expressing metastatic castration-resistant prostate cancer progressing despite prior treatment with a novel androgen axis drug | Inappropriate population |

| | | |
|---|---|---|
| 9200 | Preliminary Study on Application of Combined Monitoring of Neuroelectrophysiology-Intracranial Pressure-Cerebral Perfusion Pressure in Craniotomy for Intracranial Aneurysm Clipping: an Anatomical and Clinical Study | Inappropriate population |
| 9201 | Construction and empirical of ICU patient follow-up model based on symptom management theory: a quasi-randomized controlled trial study protocol | Inappropriate study design |
| 9202 | An assessment of the mediating role of hypertension in the effect of long-term air pollution exposure on dementia | Inappropriate study design |
| 9203 | Tai Chi-Induced Exosomal LRP1 is Associated With Memory Function and Hippocampus Plasticity in aMCI Patients | Innapropriate intervention |
| 9204 | Electroacupuncture improves vascular cognitive impairment no dementia: a Randomized Clinical Trial | Innapropriate intervention |
| 9205 | Improvements in Cardiovascular Fitness Following Forced-Rate Aerobic Cycling in Individuals With Chronic Stroke | Inappropriate population |
| 9206 | Forced-Rate Aerobic Cycling Enhances Motor Recovery in Persons With Chronic Stroke: a Randomized Clinical Trial | Inappropriate population |
| 9207 | Effect of transcranial direct current stimulation combined with transcutaneous auricular vagus nerve stimulation on poststroke cognitive impairment: a study protocol for a randomised controlled trial | Inappropriate study design |
| 9208 | Group-Based Acceptance and Commitment Therapy for PTSD in a HMO Psychiatry Clinic: an Open Trial | Inappropriate study design |
| 9209 | LBA1 Denosumab added to 2 different nab-paclitaxel regimens as neoadjuvant therapy in patients with primary breast cancer: time to event analysis from the GeparX 2 × 2 randomized clinical trial | Inappropriate population |
| 9210 | The effect of mindfulness-based cognitive therapy on the Clinical efficacy and psychological state in patients with functional dyspepsia | Inappropriate population |
| 9211 | WCN24-1376 EFFECT OF BRIGHT LIGHT THERAPY IN THE TREATMENT OF SLEEP DISORDERS IN PATIENTS WITH CHRONIC KIDNEY DISEASE UNDERGOING HEMODIALYSIS | Inappropriate population |
| 9212 | Effects of Shuxuetong combined with butylphthalide on neurological function and inflammatory response in patients with acute ischemic stroke | Innapropriate intervention |
| 9213 | Effectiveness of Yijinjing on cognitive and motor functions in patients with Parkinson’s disease: study protocol for a randomized controlled trial | Inappropriate study design |
| 9214 | Remote ischemic conditioning in acute ischemic stroke and small vessel disease - a feasibility study | Innapropriate intervention |
| 9215 | Health behavior outcomes in stroke survivors prescribed wearables for atrial fibrillation detection stratified by age | Inappropriate population |
| 9216 | Comparison of the Effects of Cognitive Dual-Task and Single-Task Balance Exercises on Static Balance among People with Anterior Cruciate Ligament Reconstruction: a Randomized Controlled Trial | Inappropriate population |
| 9217 | Construct and Criterion Validity Testing of GOT-Cog, a Novel Cognitive Screen to Better Inform Occupational Therapy Treatment Planning | Inappropriate population |
| 9218 | A network analysis exploration of PTSD, depression, somatic complaints, and anxiety disorders in a Colombian sample of victims of the Armed conflict | Inappropriate study design |
| 9219 | Improved hand function in individuals with chronic stroke: preliminary evidence from a Novel Hand Therapy Device | Inappropriate study design |
| 9220 | The effects of cannabis abstinence on cognition and resting state network activity in people with multiple sclerosis: a preliminary study | Inappropriate study design |
| 9221 | The Effect of Upright Stance and Vision on a Cognitive Task in Elderly Subjects and Patients with Parkinson’s Disease | Inappropriate population |
| 9222 | MYOCARDIAL STRAIN GUIDED CARDIOPROTECTION DURING CHEMOTHERAPY WITH CARDIAC MRI IMPROVES CARDIAC HEALTH | Inappropriate population |
| 9223 | 863 PROTON PUMP INHIBITOR THERAPY DOES NOT INCREASE THE RISK OF VASCULAR LESIONS ON MRI OR COGNITIVE DECLINE: RESULTS OF A DOUBLE BLIND RANDOMIZED, PLACEBO CONTROLLED COMPASS MIND SUB-STUDY | Innapropriate intervention |
| 9224 | Relationship Between Change in Sleep Impairments and Burden of Post- Intensive Care Syndrome Among Intensive Care Unit Survivors | Inappropriate study design |
| 9225 | Value of Automatically Derived Full Thrombus Characteristics: an Explorative Study of Their Associations with Outcomes in Ischemic Stroke Patients | Inappropriate study design |
| 9226 | The Effects of High-Intensity Interval Training and Moderate Alcohol Consumption on Cognitive Performance—A Multidisciplinary Intervention in Young Healthy Adults | Inappropriate population |
| 9227 | No Effect of New Zealand Blackcurrant Extract on Physiological and Cardiovascular Responses During Low Intensity Sustained Intermittent Isometric Contractions | Inappropriate study design |

| | | |
|---|---|---|
| 9228 | APOE genotype, hippocampal volume, and cognitive reserve predict improvement by cognitive training in older adults without dementia: a randomized controlled trial | Innapropriate intervention |
| 9229 | Noninvasive spinal stimulation improves walking in chronic stroke survivors: a proof-of-concept case series | Inappropriate population |
| 9230 | Radial versus Femoral Access for Mechanical Thrombectomy in Patients With Large Vessel Occlusion: a Randomized Clinical Trial | Inappropriate population |
| 9231 | Mental Health Resource Utilization in Individuals with Acute Traumatic and Nontraumatic Spinal Cord Injury with Mild or Moderate Depressive Symptoms | Inappropriate population |
| 9232 | 2. Investigating the Effects of Real-Time Biofeedback Using 7-Tesla MRI for Depression | Inappropriate population |
| 9233 | Submaximal, Low-Dose Eccentric vs Traditional Cycling Exercise: reduced Oxygen Uptake and Pulmonary Artery Pressure Assessed by Echocardiography in Healthy Middle-aged Adults. A Randomized Controlled, Crossover Trial | Inappropriate population |
| 9234 | Effect of Taichi on cognitive function in Breast cancer patients undergoing radiotherapy: a randomized controlled trial | Innapropriate intervention |
| 9235 | Digital therapeutics using virtual reality-based visual perceptual learning for visual field defects in stroke: a double-blind randomized trial | Inappropriate population |
| 9236 | EMPATHY AND PUNISHMENT RATINGS FOR COPYRIGHT INFRINGEMENT IN RESTED AND SLEEP RESTRICTED INDIVIDUALS | Inappropriate population |
| 9237 | The effect of intense endurance training with supplementation of egg white and wheat germ powder on cardiopulmonary function of endurance runners | Inappropriate population |
| 9238 | The feasibility of exercise to improve cerebrovascular health in older adults | Innapropriate intervention |
| 9239 | Effect of Qualifying Atherosclerotic Cardiovascular Disease Diagnosis Proximity on Cardiovascular Risk and Benefit of Empagliflozin in the EMPA-REG OUTCOME Trial | Inappropriate study design |
| 9240 | 436. Effects of Personalized Transcranial Magnetic Stimulation on Social Cognitive Network Functional Connectivity in Schizophrenia Spectrum Disorders | Inappropriate population |
| 9241 | Protective effects of Panax Ginseng against 131I-induced genotoxicity in patients with differentiated thyroid cancer | Inappropriate population |
| 9242 | Effect of furosemide combined with continuous renal replacement therapy on cardiorenal function and inflammatory response in patients with chronic renal and heart failure after hemodialysis | Inappropriate population |
| 9243 | Breathlessness Relieved by Employing Medical Air Titrated by Hospitalized Patients (BREATH) Randomized Clinical Trial | Inappropriate population |
| 9244 | 267PROPHYLACTIC CENTRAL COMPARTMENT LYMPH NODE DISSECTION IN CN0 PAPILLARY THYROID CANCER: NO IMPACT ON SURGICAL AND ONCOLOGICAL OUTCOMES AFTER 10 YEARS OF FOLLOW-UP - FINDINGS FROM A PROSPECTIVE RANDOMIZED CONTROLLED SINGLE INSTITUTION STUDY | Inappropriate population |
| 9245 | Effectiveness of proprioceptive and exteroceptive stimulation to improve motor performance in stroke | Inappropriate population |
| 9246 | 7P Patient-relevant endpoints from PAPILLON: amivantamab plus chemotherapy vs chemotherapy as first-line treatment of EGFR exon 20 insertion-mutated (Ex20ins) advanced NSCLC | Inappropriate population |
| 9247 | Bilateral ultrasound-guided erector spinae plane block for postoperative pain relief in major traumatic spine surgery: a randomized controlled trial | Inappropriate population |
| 9248 | Effect of timed dosing of usual antihypertensives according to patient chronotype on cardiovascular outcomes: the Chronotype sub-study cohort of the Treatment in Morning versus Evening (TIME) study | Inappropriate population |
| 9249 | Online dance versus online therapeutic exercise on quality of life - a study protocol for a randomized controlled trial investigating social telerehabilitation efficacy in Parkinson disease | Inappropriate study design |
| 9250 | Linguistic-based Mild Cognitive Impairment detection using Informative Loss | Inappropriate study design |
| 9251 | Effect of Marine Omega-3 Fatty Acids on Risk of Incident Cardiovascular Disease Among Individuals With Non- Alcoholic Fatty Liver Disease: secondary Analysis of a Randomized Clinical Trial | Inappropriate population |
| 9252 | LACROSSE Trial: lumbar Drainage Compared to External Ventricular Drainage in Aneurysmal Subarachnoid Hemorrhage, a Randomized Control Trial | Inappropriate population |
| 9253 | 437. Identification of Brain/Behavior-Based Pro-Cognitive Pharmacodynamic Effects for ALTO-101 in Healthy Volunteers: results From a Randomized, Double-Blind Phase 1 Study | Inappropriate population |

| | | |
|---|---|---|
| 9254 | ASSESSMENT OF COGNITIVE FUNCTION IN A RANDOMIZED, PLACEBO-CONTROLLED, INTERVENTIONAL TRIAL CONDUCTED IN PATIENTS WITH MODERATE-TO-SEVERE SYSTEMIC LUPUS ERYTHEMATOSUS (SLE) | Inappropriate population |
| 9255 | Olfactory Epithelium Stimulation Using Rhythmic Nasal Air-Puffs Improves the Cognitive Performance of Individuals with Acute Sleep Deprivation | Inappropriate study design |
| 9256 | Feasibility of a Home-based Cognitive-physical Exercise Program in Patients With Chronic Obstructive Pulmonary Disease: feasibility and Pilot Randomized Controlled Trial | Inappropriate population |
| 9257 | A Preliminary Study on the Pharmacodynamics of Oral Cannabis Ingestion in Older Adults | Inappropriate population |
| 9258 | Preliminary results from the ongoing aphenity extension study: sepiapterin reduces blood PHE with improved dietary PHE tolerance in participants with phenylketonuria | Inappropriate population |
| 9259 | Study protocol for a randomized controlled trial: effect of an everyday cognition training program on cognitive function, emotional state, frailty and functioning in older adults without cognitive impairment | Inappropriate study design |
| 9260 | Glucose Color Index: development and Validation of a Novel Measure of the Shape of Glycemic Variability | Inappropriate study design |
| 9261 | PROSTACT GLOBAL: a PHASE 3 STUDY OF BEST STANDARD OF CARE WITH AND WITHOUT 177LU-DOTA-ROSOPATAMAB (TLX591) FOR PATIENTS WITH PSMA EXPRESSING METASTATIC CASTRATION-RESISTANT PROSTATE CANCER PROGRESSING DESPITE PRIOR TREATMENT WITH A NOVEL ANDROGEN AXIS DRUG | Inappropriate study design |
| 9262 | Efficacy of Dashmool Vasti as an adjuvant therapy with the standard of care (modern + physiotherapy) in the rehabilitation of stroke as compared to standard of care – a clinical trial protocol | Inappropriate study design |
| 9263 | The potential of infra-low frequency neurofeedback training in peak performance: the first double-blinded placebo-controlled longitudinal study in healthy adults | Inappropriate population |
| 9264 | 637 LONG-TERM EFFECTS OF COGNITIVE-BEHAVIORAL AND MINDFULNESS INTERVENTION (COBMINDEX) IN ADULT PATIENTS WITH CROHN'S DISEASE (CD) | Inappropriate population |
| 9265 | Comparative effectiveness of the FIBROWALK multicomponent therapy in online and outdoor formats for individuals with fibromyalgia: the On&Out study | Inappropriate population |
| 9266 | Immune-inflammatory pathways of the FIBROWALK multicomponent program in fibromyalgia | Inappropriate population |
| 9267 | Constraint-induced movement therapy through telerehabilitation for upper extremity function in stroke | Inappropriate study design |
| 9268 | Optimal homeostatic stress to maximize the homogeneity of adaptations to interval interventions in soccer players | Inappropriate population |
| 9269 | Patient-reported outcomes for the phase 3 FURLONG study of furmonertinib versus gefitinib as first-line therapy for Chinese patients with locally advanced or metastatic EGFR mutation-positive non-small cell lung cancer | Inappropriate population |
| 9270 | Analysis of risk factors for post-stroke anxiety and depression and the therapeutic effect of Jiangqi Dayu Decoction | Inappropriate population |
| 9271 | Immersive virtual reality as a complement to physical therapy for Parkinson's disease – A randomized controlled trial | Innapropriate intervention |
| 9272 | Safety and efficacy of InTandemTM, a neurorehabilitation system to improve walking in chronic stroke | Inappropriate population |
| 9273 | The Association of CHADS-P2A2RC Risk Score With Clinical Outcomes in Patients Taking P2Y12 Inhibitor Monotherapy After 3 Months of Dual Antiplatelet Therapy Following Percutaneous Coronary Intervention | Inappropriate population |
| 9274 | Acute management of cerebral venous thrombosis: indications, technique, and outcome of endovascular treatment in two high-volume centers | Inappropriate study design |
| 9275 | Neural Outcomes and Predictors of Cognitive-Behavioral Treatment for Hoarding Disorder | Inappropriate population |
| 9276 | Effect of Exergaming-Based Dance Training Paradigm in Improving Cardiovascular Function and Functional Mobility Among People With Chronic Stroke | Inappropriate study design |
| 9277 | 73. Preliminary Efficacy Findings for a Placebo Controlled Randomized Clinical Trial of a Novel Executive Working Memory Training Intervention for Adolescents With ADHD | Innapropriate intervention |
| 9278 | Effects of alteplase and aspirin on functional outcomes in patients with acute ischemic stroke and mild non-disabling neurological deficit | Innapropriate intervention |

| | | |
|---|---|---|
| 9279 | Moral decision-making at night and the impact of night work with blue-enriched white light or warm white light: a counterbalanced crossover study | Inappropriate population |
| 9280 | Longitudinal Biomarker Analysis in Patients With Ischemic Stroke: exploratory Targeted and Omics Assessments From a Phase 2a Clinical Trial | Inappropriate population |
| 9281 | Virtual reality-based training may improve visual memory and some aspects of sustained attention among healthy older adults – preliminary results of a randomized controlled study | Inappropriate population |
| 9282 | The value of consciousness coaching in Parkinson's disease: experiences and possible impact of holistic coaching | Inappropriate population |
| 9283 | Role of oral melatonin in prevention of postoperative delirium in patients undergoing elective surgery under general anesthesia: a Randomized controlled trial | Inappropriate population |
| 9284 | Efficacy of Daoyin combined with lower limb robot as a comprehensive rehabilitation intervention for stroke patients: a randomized controlled trial | Inappropriate population |
| 9285 | The effect of transcranial direct current stimulation on balance, gait function and quality of life in patients with stroke | Inappropriate population |
| 9286 | Dynamic Cognitive–Motor Training versus Cognitive Computer-Based Training in People with Multiple Sclerosis: a Preliminary Randomized Controlled Trial with 2-Month Follow-Up | Innapropriate intervention |
| 9287 | Cognition after a 4-week high phenylalanine intake in adults with phenylketonuria – a randomized controlled trial | Innapropriate intervention |
| 9288 | Acute resistance exercise combined with whole body vibration and blood flow restriction: molecular and neurocognitive effects in late-middle-aged and older adults | Inappropriate population |
| 9289 | 105. Enhancing Affective Cognitive Control in Bipolar Disorder Using Transcranial Alternating Current Stimulation (tACS): preliminary Evidence From a Randomized, Sham-Controlled Clinical Trial | Inappropriate population |
| 9290 | The impact of regular exercise on cardiometabolic fitness and memory is related to reduced levels of residual platinum in testicular germ cell cancer survivors | Inappropriate study design |
| 9291 | Effects of solriamfetol on cognition in participants with cognitive impairment associated with excessive daytime sleepiness in obstructive sleep apnea: SHARP study results | Innapropriate intervention |
| 9292 | Neurocognitive Moderators of Group Psychotherapy Augmented by Individual Uncluttering Practice (BIT+) vs Waitlist Control | Inappropriate population |
| 9293 | Breathing Right… or Left! The Effects of Unilateral Nostril Breathing on Psychological and Cognitive Wellbeing: a Pilot Study | Inappropriate study design |
| 9294 | Effects of cognitive training and behavior modification on aggressive behavior and sleep quality in schizophrenia | Inappropriate study design |
| 9295 | Food-related inhibitory control deficits in young male adults with obesity: behavioral and ERP evidence from a food-related go/no-go task | Inappropriate study design |
| 9296 | Clinical effect of Suhexiang Pills in treatment of hemiplegia patients in convalescent period of cerebral infarction | Innapropriate intervention |
| 9297 | Transcutaneous auricular vagus nerve stimulation with task-oriented training improves upper extremity function in patients with subacute stroke: a randomized clinical trial | Inappropriate population |
| 9298 | Cognitive and Hippocampal Changes in Older Adults With Subjective Cognitive Decline After Acupuncture Intervention | Innapropriate intervention |
| 9299 | EFFECTS OF LIGHT EXPOSURE ON SLEEP, RESTACTIVITY RHYTHMS, AND COGNITIVE FUNCTION IN ADULTS WITH HEART FAILURE | Inappropriate population |
| 9300 | Local and global effects of sedation in resting-state fMRI: a randomized, placebo-controlled comparison between etifoxine and alprazolam | Inappropriate population |
| 9301 | DIFFERENTIAL BRAIN ACTIVATION TO A FINE MOTOR TASK IN INDIVIDUALS WITH NONALCOHOLIC FATTY LIVER DISEASE (NAFLD) COMPARED TO THOSE WITHOUT | Inappropriate study design |
| 9302 | Pimavanserin for Psychosis in Parkinson's Disease Dementia: subgroup Analysis of the HARMONY Trial | Inappropriate study design |
| 9303 | Behavioral avoidance as a factor in concentrated exposure and response prevention for obsessive-compulsive disorder | Inappropriate population |
| 9304 | Randomized waitlist-controlled trial of buried in treasures facilitated support groups and in-home uncluttering (BIT+) for hoarding disorder | Inappropriate population |
| 9305 | Feasibility, Acceptability, and Preliminary Efficacy of a Smartphone App–Led Cognitive Behavioral Therapy for Depression Under Therapist Supervision: open Trial | Inappropriate study design |
| 9306 | Effects of DLPFC tDCS followed by treadmill training on dual-task gait and cortical excitability in Parkinson's disease: a randomized controlled trial | Innapropriate intervention |

| | | |
|---|---|---|
| 9307 | Effects of ankle–foot orthoses on gait parameters in post-stroke patients with different Brunnstrom stages of the lower limb: a single-center crossover trial | Inappropriate study design |
| 9308 | Capturing the Effects of Non-Body Weight Supported Backward and Forward Treadmill Training on Chronic Post-Stroke Lower Extremity Spasticity and Walking Speed | Inappropriate study design |
| 9309 | Closed-loop transcutaneous auricular vagus nerve stimulation for the improvement of upper extremity motor function in stroke patients: a study protocol | Inappropriate study design |
| 9310 | Synergistic effects of neuroendoscopic minimally invasive surgery with mannitol and furosemide in managing hypertensive intracerebral hemorrhage | Inappropriate study design |
| 9311 | The Effects of Traditional Chinese Herbal Dietary Formula on the Ability of Daily Life and Physical Function in Elderly Patients with Mild Cognitive Impairment | Innapropriate intervention |
| 9312 | 330. Assessing the Combined Effects of Repetitive Transcranial Magnetic Stimulation and Cognitive Training for Improving Response Inhibition | Inappropriate population |
| 9313 | Therapeutic sleep deprivation for major depressive disorder: a randomized controlled trial | Inappropriate population |
| 9314 | Targeted High-Definition Transcranial Direction Current Stimulation Improves Upper Extremity Rehabilitation in Stroke Individuals | Inappropriate population |
| 9315 | Application of integrated Chinese and western medicine nursing based on the individual and family self-management in post-stroke urinary incontinence patients | Inappropriate population |
| 9316 | Therapeutic singing-induced swallowing exercise for dysphagia in advanced-stage Parkinson's disease | Inappropriate study design |
| 9317 | Structured different exercise protocols improve lung function, respiratory muscle strength, and thickness in stroke patients. A randomized controlled trial | Inappropriate population |
| 9318 | Effect of Extended Nursing on Quality of Life and Clinical Treatment in Patients with Chronic Renal Failure Undergoing Hemodialysis with Non Drug Therapy | Inappropriate population |
| 9319 | A Randomized, Double-Blind, PlaceboControlled, Phase II Trial in Acute Non-Cardioembolic Ischemic Stroke With Adrenomedullin | Inappropriate population |
| 9320 | Comparison of Task-Oriented and Bobath-Based Trunk Training on Balance, Functional Abilities and Walking in Ischemic Stroke Patients | Inappropriate study design |
| 9321 | Healthcare Students' Perceptions and Attitudes Towards Peers with Autism Spectrum Disorders | Inappropriate study design |
| 9322 | Effect of transcranial direct current stimulation with different current density on upper limb motor disorder after stroke | Inappropriate study design |
| 9323 | Impact of Radiotherapy on Radiation-Induced Brain Injury in Patients with Nasopharyngeal Carcinoma Using Nano-Magnetic Resonance Imaging | Inappropriate study design |
| 9324 | Endovascular therapy in acute ischaemic stroke with large infarction with matched or mismatched clinical-radiological severities: a post-hoc analysis of the ANGEL-ASPECT trial | Inappropriate population |
| 9325 | Association of RASGRP1 polymorphism with vascular complications in Chinese diabetic patients with glycemic control and antihypertensive treatment | Inappropriate population |
| 9326 | Effect of proprotein convertase subtilisin/kexin type 9 inhibitor combined with rosuvastatin on lipid levels and short-term prognosis in patients with acute ischemic stroke | Inappropriate study design |
| 9327 | CARDIOVASCULAR BENEFIT OF EVOLOCUMAB IN 27,564 PATIENTS WITH AND WITHOUT AUTOIMMUNE OR INFLAMMATORY DISEASES: AN ANALYSIS OF THE FOURIER TRIAL | Inappropriate study design |
| 9328 | Efficacy and safety of oral sepiapterin in participants with phenylketonuria on sapropterin dihydrochloride at time of Phase 3 aphenity study entry | Inappropriate study design |
| 9329 | EFFICACY OF ICOSAPENT ETHYL FOR REDUCING CARDIOVASCULAR OUTCOMES BY BASELINE LOW DENSITY LIPOPROTEIN CHOLESTEROL LEVEL | Inappropriate population |
| 9330 | Neuromodulation of Executive Dysfunction in Patients with Acute Stroke Using Transcranial Direct Current Stimulation: study Protocol for a Triple-Blind, Randomized Sham-Controlled Trial | Inappropriate population |
| 9331 | BEMPEDOIC ACID AND CARDIOVASCULAR DISEASE OUTCOMES IN PATIENTS WITH OBESITY: a CLEAR OUTCOMES SUBSET ANALYSIS | Inappropriate study design |
| 9332 | ELECTRONIC NUDGES TO INCREASE INFLUENZA VACCINATION UPTAKE AMONG PARTICIPANTS WITH HISTORY OF ACUTE MYOCARDIAL INFARCTION: INSIGHTS FROM NUDGE-FLU | Inappropriate population |
| 9333 | Angong niuhuang pill improves the neurological function of younger patients with basal ganglia and cerebral lobe intracerebral hemorrhage: a randomized controlled trial | Innapropriate intervention |

| | | |
|---|---|---|
| 9334 | A comparative study using gastric ultrasound to evaluate the safety of shortening the fasting time before pediatric echocardiography: a randomized controlled non-inferiority study | Inappropriate study design |
| 9335 | Individualized Treatment Effects of CPAP in Obstructive Sleep Apnea: a Post-hoc Machine Learning Analysis in Save | Inappropriate population |
| 9336 | Predictors of bleeding and thrombotic events among patients admitted to the hospital with COVID-19 and elevated D-dimer: insights from the ACTION randomized clinical trial | Inappropriate population |
| 9337 | CE-482906-005 YIELD OF GENETIC TESTING IN EARLY-ONSET ATRIAL FIBRILLATION: a SUBSTUDY OF THE BRAIN-AF RANDOMIZED CLINICAL TRIAL | Inappropriate population |
| 9338 | Intensive Blood Pressure Lowering Improves Left Ventricular Geometry in Older Hypertensive Patients: the STEP Trial | Inappropriate population |
| 9339 | REAL-WORLD ANALYSES OF MACE AND MORTALITY RISK IN PATIENTS WITH PROSTATE CANCER AFTER INITIATION OF LHRH AGONISTS VS. GNRH ANTAGONIST | Inappropriate population |
| 9340 | Preliminary Results of a Pilot Clinical Trial Using a Reimbursable Form of Constraint-induced Movement Therapy: the Keys Treatment protocol | Inappropriate study design |
| 9341 | Time from symptom onset and number of health care encounters prior to diagnosis of cerebral venous thrombosis | Inappropriate study design |
| 9342 | Percutaneous Coronary Intervention Versus Coronary Artery Bypass Grafting in Patients with Left Main Disease with and Without Diabetes: findings from a Pooled Analysis of 4 Randomized Clinical Trials | Inappropriate population |
| 9343 | REMNANT LIPOPROTEIN CHOLESTEROL LEVELS, INTENSIVE BLOOD PRESSURE TREATMENT, AND CARDIOVASCULAR OUTCOMES: INSIGHTS FROM THE SPRINT TRIAL | Inappropriate study design |
| 9344 | BIOEQUIVALENCE OF RIVAROXABAN HARD CAPSULES VS. FILM-COATED TABLETS IN HEALTHY CAUCASIAN VOLUNTEERS | Inappropriate population |
| 9345 | EFFICACY AND SAFETY OF DUAL ANTIPLATELET THERAPY AFTER ERIPHERAL ARTERY REVASCULARIZATION: INSIGHTS FROM VOYAGER PAD | Inappropriate population |
| 9346 | Effect of Tongnaoyin on Cerebral Hemodynamics in Patients with Acute Cerebral Infarction of Phlegm and Blood Stasis Syndrome Based on CTA/CTP | Inappropriate population |
| 9347 | Cerebellar tDCS enhances functional communication skills in chronic Aphasia | Innapropriate intervention |
| 9348 | Aspirin for Primary Prevention of Cardiovascular Events in Relation to Lipoprotein(a) Genotypes | Inappropriate population |
| 9349 | Prevention of Tuberculosis in people living with Diabetes Mellitus - the PROTID project | Inappropriate population |
| 9350 | A RANDOMIZED CONTROLLED TRIAL TO EXAMINE THE UTILITY OF TADALAFIL (5MG) FOR TREATMENT OF EARLY STORAGE SYMPTOMS AND ERECTILE DYSFUNCTION AFTER ANATOMICAL ENDOSCOPIC ENUCLEATION OF PROSTATE (AEEP) | Inappropriate study design |
| 9351 | EFFICACY OF INTRAVENOUS FERRIC DERISOMALTOSE IN HOSPITALISED AND OUTPATIENT SETTINGS: INSIGHTS FROM IRONMAN TRIAL | Inappropriate population |
| 9352 | Impact of Intravascular Imaging-Guided Stent Optimization According to Clinical Presentation in Patients Undergoing Complex PCI | Inappropriate population |
| 9353 | Should renin–angiotensin system inhibitors be held prior to major surgery? | Inappropriate population |
| 9354 | AB-482902-003 THE IMPACT OF ATRIAL REMODELING ON THE EFFICACY OF CATHETER ABLATION IN REDUCING CARDIOVASCULAR RISK IN ATRIAL FIBRILLATION: RESULTS FROM THE CABANA TRIAL | Inappropriate population |
| 9355 | WCN24-432 FINERENONE IN CHINESE PATIENTS WITH TYPE 2 DIABETES AND CHRONIC KIDNEY DISEASE: a FIDELITY ANALYSIS | Inappropriate population |
| 9356 | MULTICENTER STUDY TO EVALUATE THE EFFICACY OF 1-L ASCORBIC ACID PEG PLUS LINACLOTIDE FOR BOWEL PREPARATION: a RANDOMIZED, ENDOSCOPIST-BLINDED, CONTROLLED TRIAL (APPLE TRIAL) | Inappropriate population |
| 9357 | EFFECTIVENESS OF PATIENT'S EDUCATION IN SELF-MANAGEMENT OF BLOOD PRESSURE IN HYPERTENSIVE PATIENTS | Inappropriate population |
| 9358 | Coherent activity within and between hemispheres: cortico-cortical connectivity revealed by rTMS of the right posterior parietal cortex | Inappropriate study design |
| 9359 | Inhaled argon as a neuroprotective agent after an ischemic event – a fully translational story from animal studies to a human trial | Inappropriate study design |

| | | |
|---|---|---|
| 9360 | Immediate Effects of Calf Muscle Release via Foam Rolling and Active Stretching on Blood Pressure, Heart Rate and the Rate Pressure Product in People with High-Normal Blood Pressure – A Comparative Study | Inappropriate study design |
| 9361 | REGRESSION OF INFARCT SCARS IN COMET-AMI (CONCOMITANT MILRINONE AND ESMOLOL TREATMENT IN PATIENTS WITH ACUTE MYOCARDIAL INFARCTION) PILOT RANDOMISEDCONTROLLED STUDY | Inappropriate study design |
| 9362 | REAL-WORLD APPLICATION OF SELECT TRIAL CRITERIA IN A LARGE URBAN AND SUBURBAN HEALTH SYSTEM | Inappropriate population |
| 9363 | Hypoxic Burden but Not AHI Predicts Risk of Cardiovascular Events: a Secondary Analysis of the RICCADSA Clinical Trial | Inappropriate population |
| 9364 | ASSOCIATION OF URINARY DICKKOPF-3 LEVELS WITH CARDIOVASCULAR EVENTS AND KIDNEY DISEASE PROGRESSION IN THE SYSTOLIC BLOOD PRESSURE INTERVENTION TRIAL PARTICIPANTS | Inappropriate study design |
| 9365 | Implications of trial eligibility in patients with heart failure with mildly reduced or preserved ejection fraction | Inappropriate study design |
| 9366 | Comparison of Fibrinogen Concentrate and Cryoprecipitate in Children Undergoing Cardiac Surgery: final Results of a Pilot Feasibility Trial | Inappropriate population |
| 9367 | 759 THE EFFICACY AND SAFETY OF GUSELKUMAB AS MAINTENANCE THERAPY IN PATIENTS WITH MODERATELY TO SEVERELY ACTIVE ULCERATIVE COLITIS: RESULTS FROM THE PHASE 3 QUASAR MAINTENANCE STUDY | Inappropriate population |
| 9368 | Sildenafil Modifies Platelet Activation and Reduces Mediators of Vascular Remodeling During Durable Left Ventricular Assist Device Support: a Double-Blind, Placebo-Controlled, Randomized Study | Inappropriate population |
| 9369 | Michelangelo Effect in Cognitive Rehabilitation: using Art in a Digital Visuospatial Memory Task | Inappropriate study design |
| 9370 | Evaluation of the Hemocompatibility of the Direct Oral Anticoagulant Apixaban in Left Ventricular Assist Devices: the DOAC LVAD Study | Inappropriate population |
| 9371 | Sex-based outcomes on unguided de-escalation from ticagrelor to clopidogrel in stabilized patients with acute myocardial infarction undergoing percutaneous coronary intervention: a post-hoc analysis of the TALOS-AMI | Inappropriate population |
| 9372 | Psilocybin Desynchronizes Brain Networks | Inappropriate population |
| 9373 | Benefits of using a support bra in women undergoing coronary artery bypass graft surgery: a randomized trial | Inappropriate population |
| 9374 | The Effects of Alpha Linolenic Acid (Omega-3 Fatty Acid) on Tobacco Craving Intensity, Withdrawal Symptoms and Biochemical Markers in Tobacco Users | Inappropriate study design |
| 9375 | ICOSAPENT ETHYL REDUCES MACE IN PATIENTS WITH ELEVATED TRIGLYCERIDES AND HIGH OR LOW LIPOPROTEIN(A) CONCENTRATIONS: a REDUCE-IT SUBANALYSIS | Inappropriate population |
| 9376 | Does Platelet-Rich Fibrin Enhance Recovery From Neurosensory Disturbance Following Mandibular Fractures? A Double-Blind, Split-Mouth Randomized Clinical Trial | Inappropriate population |
| 9377 | OUTCOMES OF PATIENTS WITH DIABETES AFTER FFR-GUIDED PCI OR CABG | Inappropriate study design |
| 9378 | An Aspirin-Free Strategy for Immediate Treatment Following Complex Percutaneous Coronary Intervention | Inappropriate population |
| 9379 | Rationale and Design of the IMPROVE Trial: a Multicenter, Randomized, Controlled, Open-label, Blinded-endpoint Trial Assessing the Efficacy of Remote Ischemic Conditioning in Patients Undergoing Off-Pump Coronary Artery Bypass Grafting | Inappropriate study design |
| 9380 | Efficacy of early administration of sacubitril/valsartan after coronary artery revascularization in patients with acute myocardial infarction complicated by moderate-to-severe mitral regurgitation: a randomized controlled trial | Inappropriate population |
| 9381 | Exploring the multifaceted potential of (R)-ketamine beyond antidepressant applications | Inappropriate study design |
| 9382 | Effect of remote ischemic preconditioning intervention on serum levels of microRNA-582–5p/HMGB1 in patients with acute cerebral infarction | Inappropriate population |
| 9383 | EFFECT OF SEMAGLUTIDE ON CARDIORENAL OUTCOMES BY BASELINE BODY MASS INDEX AND PRIOR CARDIOVASCULAR DISEASE: POOLED ANALYSIS OF SUSTAIN 6 AND PIONEER 6 | Inappropriate population |
