## Supplementary File 4 for "Effects of an acute bout of exercise on cognitive function in adults with cognitive impairment: A systematic review with meta-analysis of randomised controlled trials"

*Study characteristics (N=15)*

| Study | Year | Country | N | Age (yrs.) Mean (SD) | Sex | Cognitive impairment |
| --- | --- | --- | --- | --- | --- | --- |
| Amjad et al. | 2019 | Pakistan | EX = 21  CON = 19 | EX = 58.00 (2.00)  CON = 60.00 (3.00) | ND | MoCA <25 |
| Amjad et al. | 2020 | Pakistan | EX = 14  CON = 14 | ND | ND | MMSE <25 |
| Barcelos et al. | 2015 | United States & Ireland | EX (game) = 27  EX (tour) = 30 | EX (game) = 84.10 (10.60)  EX (tour) = 80.60 (8.60) | MF | EX (game) MoCA = 21.60 (SD = 4.30)  EX (tour) MoCA = 20.60 (SD = 5.60) |
| Bonuzzi et al. | 2023 | Brazil | EX (before) = 9  CON = 11 | EX (before) = 51.66 (9.18)  CON = 53.40 (15.10) | MF | EX (before) MoCA = 23.13 (SD = 1.99)  CON MoCA = 23.4 (SD=2.5) |
| Chen et al. | 2015 | United States | EX = 10  CON = 10 | EX = 23.45 (4.86)  CON = 20.58 (5.74) | MF | Down’s Syndrome |
| Devenney et al. | 2019 | Ireland | EX = 35  CON = 29 | EX = 71.70 (5.30)  CON = 69.00 (7.20) | MF | MoCA = 22.00 (SD = 2.50) |
| Molloy et al. | 1988 | Canada | EX = 15  CON = 15 | EX = 66.00 (ND)  CON = 66.00 (ND) | MF | MMSE = 23.50 (SD = 1.50) |
| Lefferts et al. | 2021 | United States | EX = 16  CON = 16 | EX = 42.00 (8.00)  CON = 42.00 (8.00) | ND | Multiple Sclerosis |
| Salerno et al. | 2019 | United States | EX = 27  CON = 27 | EX = 49.11 (8.05)  CON = 49.11 (8.05) | F | Cancer related cognitive impairment |
| Salerno et al. | 2020 | United States | EX (10 min) = 15  EX (20 min) = 16  EX (30 min) = 17  CON (10 min) = 15  CON (20 min) = 16  CON (30 min) = 17 | EX (10 min) = 56.80 (10.67)  EX (20 min) = 55.13 (13.26)  EX (30 min) = 56.18 (9.44)  CON (10 min) = 56.80 (10.67)  CON (20 min) = 55.13 (13.26)  CON (30 min) = 56.18 (9.44) | F | Cancer related cognitive impairment |
| Subramaniapillai et al. | 2016 | Canada | EX = 17  CON = 16 | EX = 45.00 (12.87)  CON = 45.00 (12.87) | MF | Schizophrenia (n=22)  Schizoaffective (n=13)  Psychosis (n=1) |
| Swatridge et al. | 2017 | Canada | EX = 9  CON = 9 | EX = 58.80 (11.40)  CON = 58.80 (11.40) | MF | MoCA = 25.30 (SD=4.90) |
| Tsai et al. | 2018 | Taiwan | EX (Aerobic) = 25  EX (Resistance) = 21  CON = 20 | EX (Aerobic) = 65.48 (7.53)  EX (Resistance) = 66.05 (6.64)  CON = 64.50 (6.95) | MF | EX (Aerobic) MMSE = 26.96 (SD = 1.21)  EX (Resistance) MMSE = 26.76 (SD = 1.38)  CON MMSE = 27.00 (SD = 1.59) |
| Venturelli et al. | 2012 | Italy | EX = 20  CON = 20 | EX = 83.00 (4.00)  CON = 83.00 (4.00) | MF | MMSE = 11.00 (SD = 3.00) |
| Wanner et al. | 2021 | Germany | EX = 8  CON = 9 | EX = 59.70 (7.40)  CON = 60.80 (8.80) | MF | EX MoCA = 25.90 (SD = 2.50)  CON MoCA = 26.20 (SD = 1.00) |
