## Supplementary File 5 for "Effects of an acute bout of exercise on cognitive function in adults with cognitive impairment: A systematic review with meta-analysis of randomised controlled trials"

*Study level risk of bias assessment results for studies included in the meta-analysis (N=10)*

| Study | Randomisation process | Bias arising from period and carry over effects | Deviations from intended interventions | Missing outcome data | Measurement of the outcome | Selection of the reported result | Overall bias |
| --- | --- | --- | --- | --- | --- | --- | --- |
| Chen et al. (2015) | Some concerns | N/A | High | High | Low | High | High |
| Devenney et al. (2019) | Low | N/A | Some concerns | Low | Low | High | High |
| Molloy et al. (1988) | Low | Low | High | High | Low | Some concerns | High |
| Lefferts et al. (2021) | Some Concerns | Low | Low | Low | Low | Some concerns | Some concerns |
| Salerno et al. (2019) | Low | High | High | Low | Low | Some concerns | High |
| Salerno et al. (2020) | Low | Some concerns | Some concerns | Low | Low | Some concerns | Some concerns |
| Subramaniapillai et al. (2016) | Low | Low | Low | Low | Low | Some concerns | Some concerns |
| Swatridge et al. (2017) | Some concerns | Some concerns | Low | Low | Low | Some concerns | Some concerns |
| Tsai et al. (2018) | Low | N/A | High | High | Low | Some concerns | High |
| Venturelli et al. (2012) | Low | Some concerns | High | Low | Low | Some concerns | High |
