## Supplementary File 6 for "Effects of an acute bout of exercise on cognitive function in adults with cognitive impairment: A systematic review with meta-analysis of randomised controlled trials"

**Question:** Acute exercise compared to no exercise for improving cognitive function in those with cognitive impairment

| **Certainty assessment** | | | | | | | **№ of patients** | | **Effect** | **Certainty** | **Importance** |
| --- | --- | --- | --- | --- | --- | --- | --- | --- | --- | --- | --- |
| **№ of studies** | **Study design** | **Risk of bias** | **Inconsistency** | **Indirectness** | **Imprecision** | **Other considerations** | **Acute exercise** | **no exercise** | **Absolute (95% CI)** |  |  |
| **Executive Function before and after acute exercise (assessed with: Executive Function Tests)** | | | | | | | | | | | |
| 9 | randomised trials | very serious^a^ | not serious | not serious | not serious | all plausible residual confounding would reduce the demonstrated effect | 216 | 203 | SMD **0.35 SD higher** (0.05 higher to 0.64 higher) | ⨁⨁⨁◯ Moderate^a,b^ | CRITICAL |
| **Reaction Time before and after acute exercise (assessed with: Reaction time tests)** | | | | | | | | | | | |
| 7 | randomised trials | very serious^a^ | serious^b^ | not serious | serious^c^ | all plausible residual confounding would reduce the demonstrated effect | 191 | 179 | SMD **0.23 SD higher** (0.08 lower to 0.55 higher) | ⨁◯◯◯ Very low^a,b,c^ | CRITICAL |
| **Memory before and after acute exercise (assessed with: Memory tests)** | | | | | | | | | | | |
| 4 | randomised trials | very serious^a^ | serious^d^ | not serious | serious^d^ | strong association all plausible residual confounding would reduce the demonstrated effect | 97 | 91 | see comment | ⨁⨁◯◯ Low^a,d^ | CRITICAL |
| **Motor Memory before and after acute exercise (assessed with: Tests of motor memory consolidation)** | | | | | | | | | | | |
| 3 | randomised trials | very serious^a^ | serious^d^ | not serious | serious^d^ | all plausible residual confounding would reduce the demonstrated effect | 43 | 44 | see comment | ⨁◯◯◯ Very low^a,d^ | CRITICAL |
| **Attention before and after acute exercise (assessed with: Attention tests)** | | | | | | | | | | | |
| 2 | randomised trials | very serious^a^ | serious^e^ | not serious | serious^e^ | all plausible residual confounding would reduce the demonstrated effect | 45 | 39 | see comment | ⨁◯◯◯ Very low^a,e^ | CRITICAL |
| **Information processing before and after acute exercise (assessed with: Information processing tests)** | | | | | | | | | | | |
| 2 | randomised trials | very serious^a^ | serious^e^ | not serious | serious^e^ | all plausible residual confounding would reduce the demonstrated effect | 42 | 42 | see comment | ⨁◯◯◯ Very low^a,e,f^ | CRITICAL |
| **Crystallised Intelligence before and after acute exercise (assessed with: Test of severe impairment - general knowledge)** | | | | | | | | | | | |
| 1 | randomised trials | very serious^a^ | not serious | not serious | very serious^f^ | strong association all plausible residual confounding would reduce the demonstrated effect | 20 | 20 | MD **0.4 TSI score lower** (0 to 0 ) | ⨁⨁◯◯ Low^a,g^ | IMPORTANT |
| **Global cognitive function before and after acute exercise (assessed with: MMSE)** | | | | | | | | | | | |
| 1 | randomised trials | very serious^a^ | not serious | not serious | very serious^f^ | all plausible residual confounding would reduce the demonstrated effect | 15 | 15 | MD **1.2 MMSE score higher** (0 to 0 ) | ⨁◯◯◯ Very low^a,g^ | CRITICAL |

**CI:** confidence interval; **MD:** mean difference; **SMD:** standardised mean difference

#### Explanations

a. More than 50% of studies at an overall high risk of bias based on the Cochrane Risk of Bias Assessment Instrument (version 2.0).

b. Overall I-squared statistic >50% but <75%.

c. Overall mean effect in the direction of benefit but 95% confidence included zero (0).

d. Most studies found effects in the direction of benefit, however the magnitude of effect sizes ranged widely (indicating inconsistency and imprecision)

e. One study found effect in the direction of benefit while the other found an effect in the direction away from benefit (indicating inconsistency and imprecision)

f. Small sample size (N<20) reduces statistical power and lack of confidence intervals reduces precision
