## Supplementary File 7 for "Effects of an acute bout of exercise on cognitive function in adults with cognitive impairment: A systematic review with meta-analysis of randomised controlled trials"

**Supplementary File 7 – Executive function and Reaction time meta-analyses**

1.
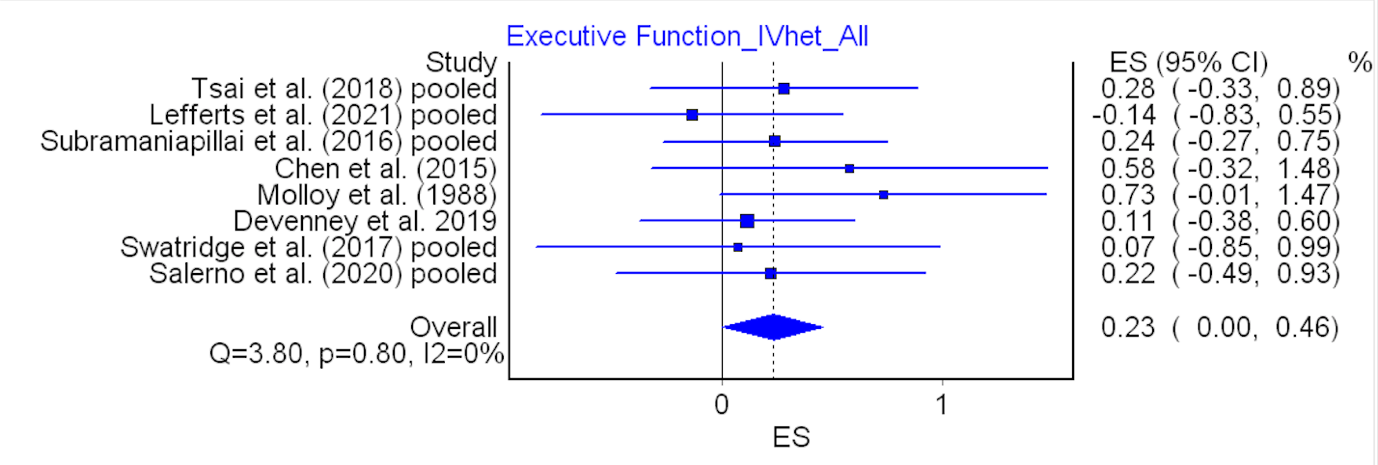

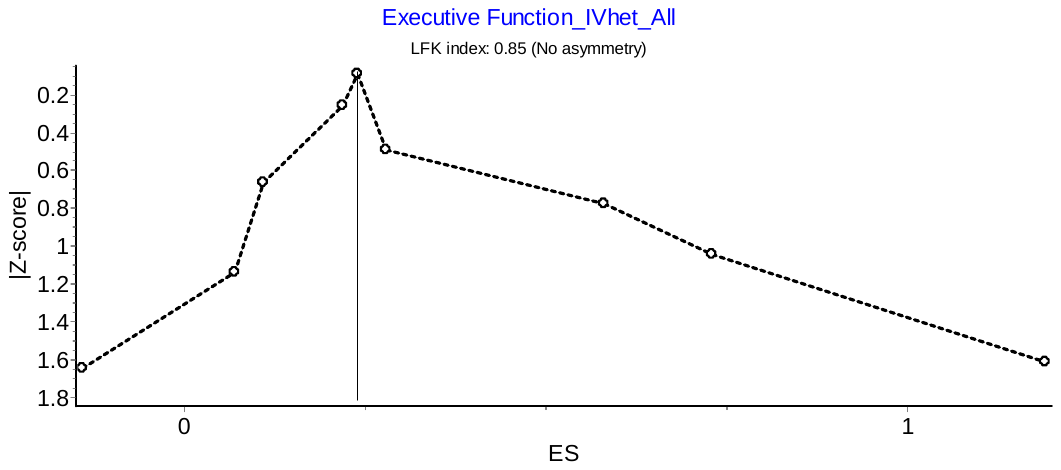
**The impact of acute exercise on executive function (N=9)**

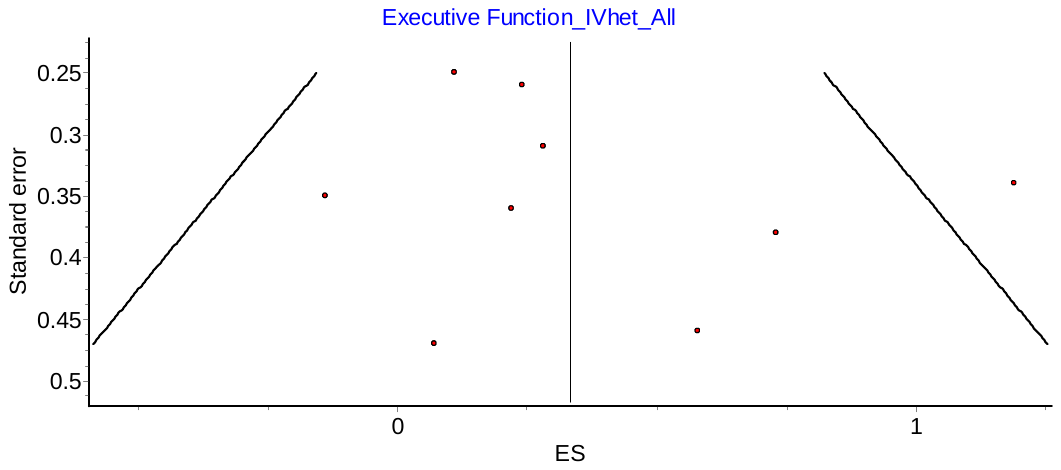

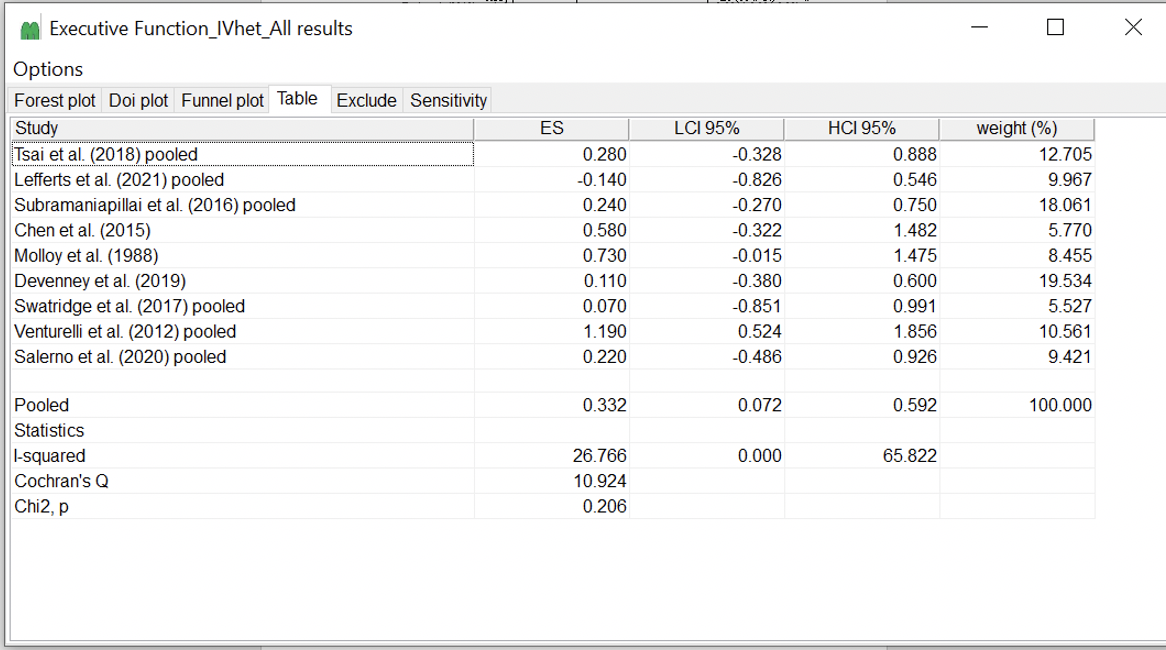

2. **The impact of acute exercise on reaction time (N=7)**

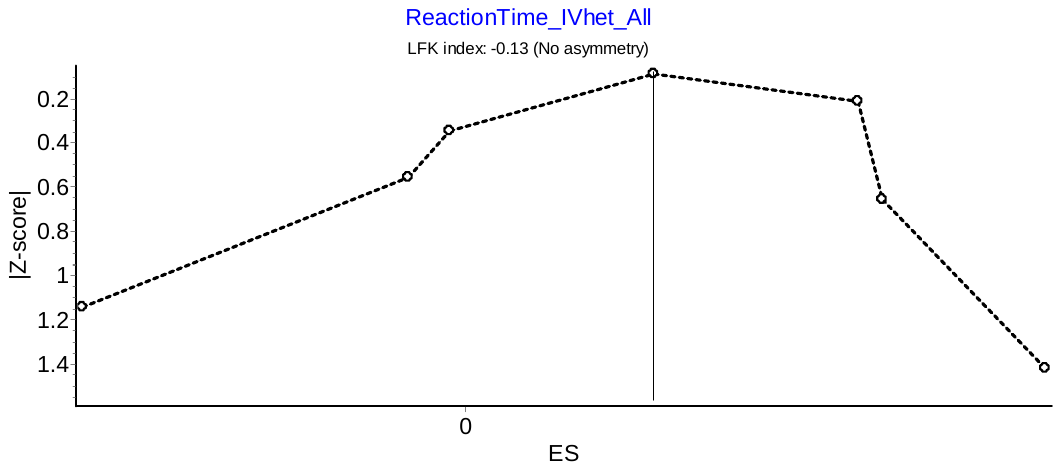

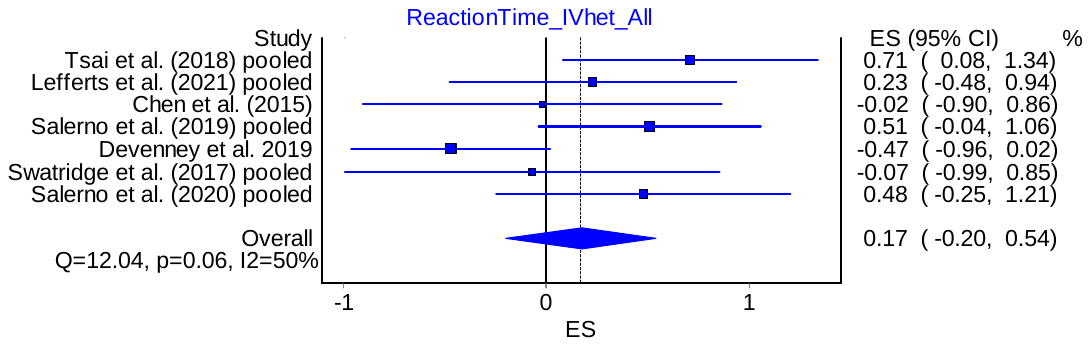

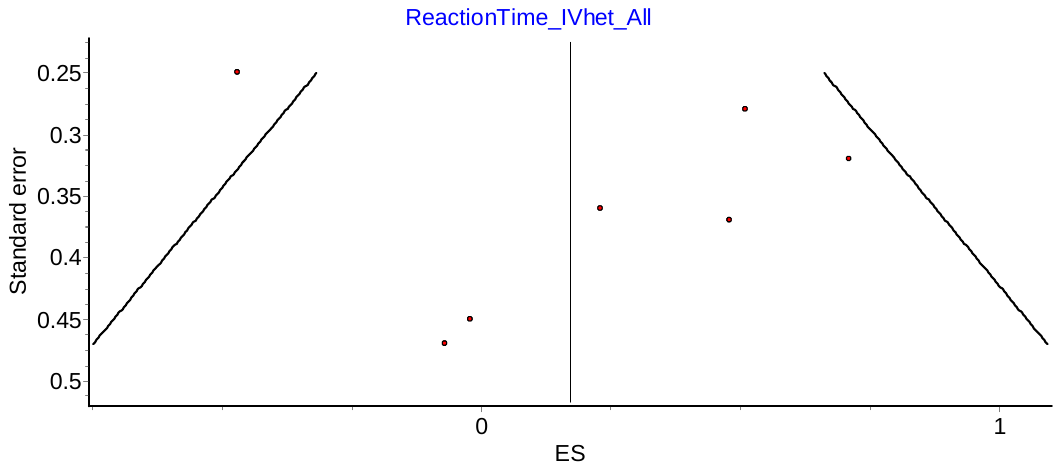

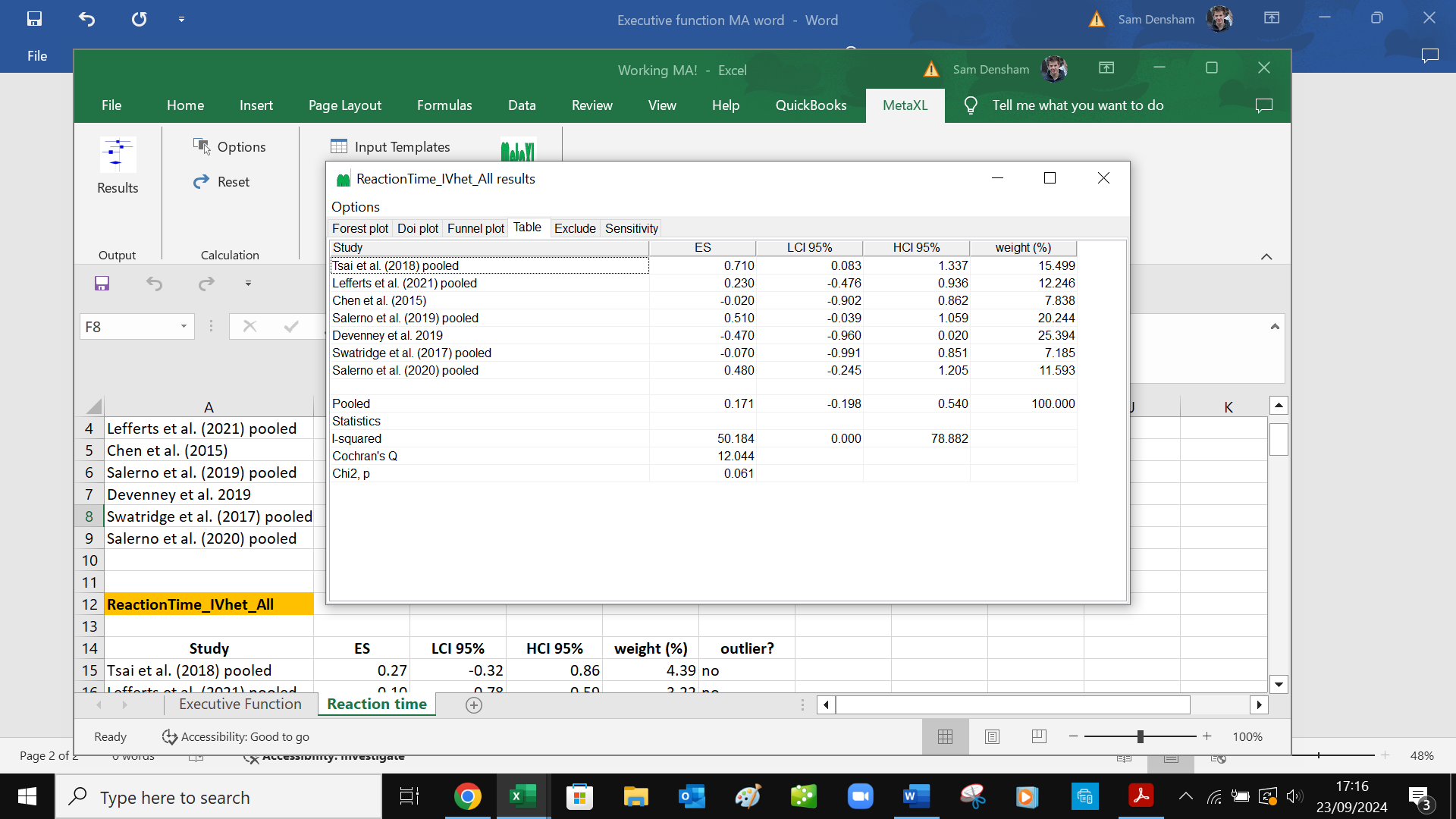

3. **Executive function sensitivity analyses**

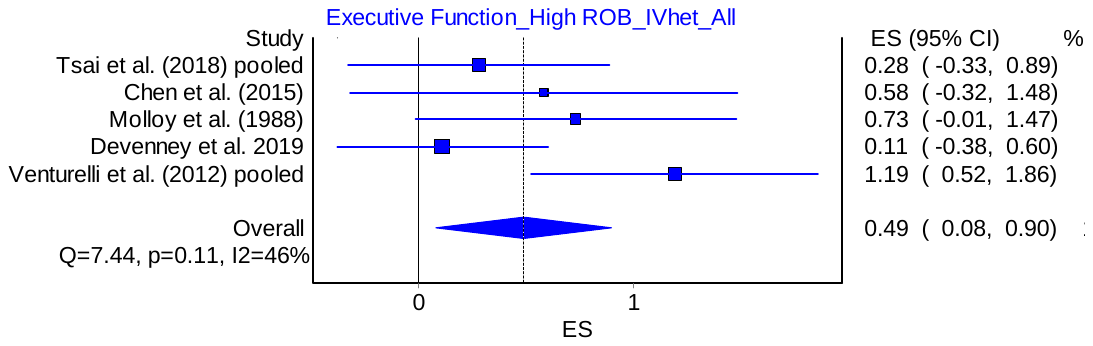
3a. The effect of acute exercise on executive function – high risk of bias studies only (N=5)

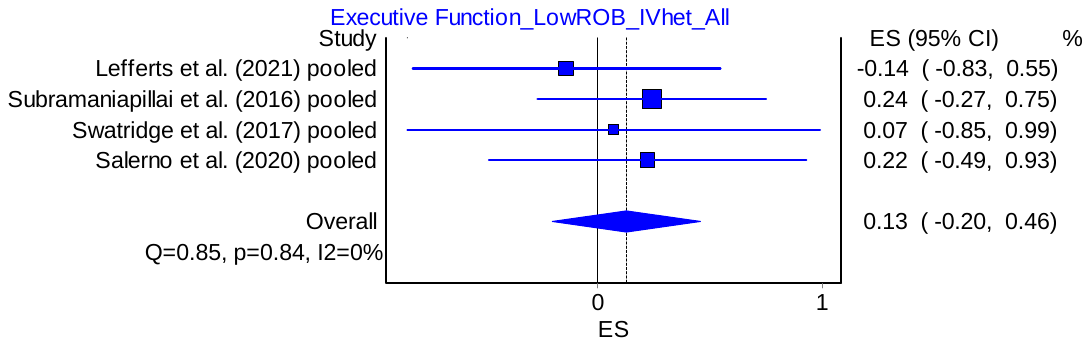
3b. The effect of acute exercise on executive function – low risk of bias studies only (N=4)

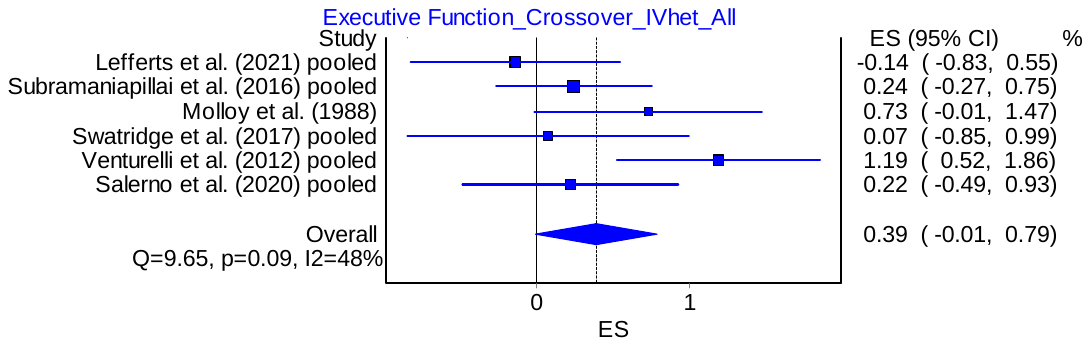
3c. The effect of acute exercise on executive function – crossover design studies only (N=6)

3d. The effect of acute exercise on executive function – parallel groups design studies only (N=3)

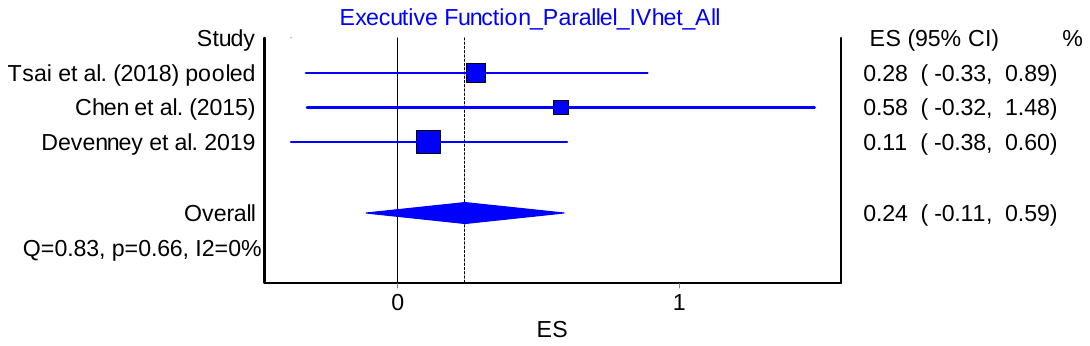
